# Sturgeon V2: a clinically applicable CNS tumor classifier trained on relabeled data

**DOI:** 10.64898/2026.10.19.26365910

**Authors:** C. Vermeulen, H. van der Ent, S.L. Maas, E.M. Bos, K. Cammel, A.D. Maier, T. Mathiesen, M.E.G. Kranendonk, P. Wesseling, D. Seinstra, EJ. Kooi, B. Stikker, H.J. Dubbink, M. Sie, O.H.J. Eelkman Rooda, E.W. Hoving, N. Verburg, M. Pagès-Gallego, J. Hench, S. Frank, K. Aldape, M. Quezado, Z. Abdullaev, O. Singh, L. Kester, B.B.J. Tops, J. de Ridder

**Affiliations:** Princess Máxima Center for Pediatric Oncology, Utrecht, The Netherlands; Oncode Institute, Utrecht, The Netherlands; Center for Molecular Medicine, UMC Utrecht, Utrecht, The Netherlands; Department of Pathology, Brain Tumor Center, Erasmus MC Cancer Institute, University Medical Center Rotterdam, the Netherlands; Department of Neurosurgery, Copenhagen University Hospital, Copenhagen, Denmark; Department of pathology, Amsterdam University Medical Centers, Amsterdam, The Netherlands; Cyclomics BV, Amsterdam, The Netherlands; Institut für Medizinische Genetik und Pathologie, Basel, Switzerland; Mayo Clinic, Rochester, USA; National Cancer Institute, Bethesda, USA

## Abstract

DNA methylation-based classification is increasingly a cornerstone of CNS tumor diagnostics. Recently,nanopore sequencing enables very rapid methylation profiling to support surgical decision making. Time-constrained sequencing yields sparse methylation profiles, necessitating specialized classifiers for accurate predictions. All publicly available sparse-profile classifiers rely on a single reference set of only 2,801 samples published in 2018 and its accompanying classification system. Here we present Sturgeon V2, trained on nearly 10,000 methylation profiles assembled from a range of public repositories and collaborations. To harmonize differences in tumor class labels, we developed an iterative semi-supervised relabeling strategy with a two-tiered concordance filter. We demonstrate that this filtering is essential for optimal performance. Compared to Sturgeon V1, we further redesigned the classifier architecture to provide more stable, and consistent confidence estimates while accumulating sequencing data and we introduce an updated class system including new classes, such as meningioma subtypes, PATZ-fusion neuroepithelial tumors and PLAG1/2-amplified embryonal tumors. In a retrospective intraoperative cohort of 94 pediatric cases, Sturgeon V2 was confidently correct in 85 cases (Sturgeon V1: 78) with one confident misclassification, and in simulated sequencing runs (n=1880) reduced the error rate among confident calls from 7.1% to 2.8%, outperforming publicly available alternatives. Sturgeon V2 is the first openly available, locally executable classifier for sparse methylation profiles trained beyond the 2018 reference era.

## Introduction

Central nervous system (CNS) tumors encompass some of the most lethal types of cancer in both adults and children^1^. Classification of CNS tumors increasingly relies on DNA methylation profiling, and advancements in methylation-based classification have given rise to two main classifiers, the Heidelberg classifier ^2^ (184 methylation classes) and the Bethesda classifier (https://methylscape.ccr.cancer.gov/) (216 methylation classes), to classify tumors based on profiles generated with genome-wide methylation arrays. Surgical resection is a critical step in the treatment ^3^ of CNS tumors, but requires the neurosurgeon to balance the risk of relapse against the risk of neurological damage and comorbidity ^4^. Precise methylation-based classification of the tumor, before or during surgery, can indicate whether an extensive resection is warranted ^5–7^.

To provide such a molecular diagnosis during surgery, intraoperative nanopore sequencing was devised ^8^ and further developed ^9–13^. The intraoperative time window precludes whole-genome coverage, and is instead limited to sparse methylation profiles. This gives rise to a unique challenge: classifiers must reliably classify a tumor from a random, very sparse sampling of its methylation profile. We previously developed Sturgeon for this purpose ^9^, a deep-learned neural network classifier (henceforth Sturgeon V1) trained on millions of simulated nanopore sequencing runs. Sturgeon has been employed successfully in both research and clinical settings ^14–17^. Alternatives include an ad-hoc random forest ^8^, CrossNN ^8,11^, a Bayesian inference network MethyLYZR ^10^, an unsupervised clustering approach NanoDiP ^10,13^, and an XGBoost-based algorithm ^12^. All these tools rely on methylation arrays (450K, EPIC, EPIC V2) as reference data, either simulating sparse nanopore profiles from arrays for training, or downsampling the reference to match the intraoperative profile.

Three limitations constrain the current generation of sparse-profile classifiers. First, all publicly available tools rely on the reference set of Capper et al. ^18^, consisting of only 2,801 samples and spanning 82 tumor classes. Therefore, although array-based classifiers moved on from the 2018 classification system, most sparse-profile classifiers still rely on the 2018 definitions. Second, the one classifier trained on a larger, up-to-date dataset (MNP-FLEX) is available only through a web interface ^12^, precluding intraoperative deployment and independent benchmarking. Third, as we and others have observed, the original Sturgeon architecture can exhibit class ‘flipping’ in challenging samples ^16,17^.

Here we present an iterative semi-supervised relabeling strategy to harmonize data from different sources and with incompatible labeling systems, while filtering out low-quality samples. With the resulting high-quality dataset consisting of 9907 samples we developed Sturgeon V2, a substantially improved, publicly available and locally executable nanopore-compatible classifier.

The Sturgeon V2 class system adheres to the latest WHO guidelines for tumor classification ^19^ where possible. As intraoperative sequencing does not require the finest level subclassification as available in the Bethesda and Heidelberg classifiers, we opted for a class system that prioritizes intraoperative usability without having too fine-grained subclasses, which may compromise classifier performance. Improving Sturgeon V2’s architecture and applying coverage-dependent calibration stabilize confidence estimation and prevent class-flipping. We further introduce clinically oriented classes, including meningioma subtypes, PATZ-fusion neuroepithelial tumors and PLAGL1/2-amplified embryonal tumors. We benchmark Sturgeon V2 against publicly available sparse-profile classifiers. Sturgeon V2 confidently and correctly classified 90% (85/94) of retrospective intraoperative pediatric cases outperforming other classifiers available for benchmarking, and is freely available for non-commercial, local use.

## Results

### Developing a classification system for intraoperative use

*V2 class label definition -* We define classes at the granularity at which they alter intraoperative decision-making. The most recent Heidelberg 12.8 system, spanning 184 tumor subtypes, was considered to be too fine-grained for effective intraoperative use as many tumor subclasses are highly similar with an (yet) unclear significance for intraoperative decision-making. Highly similar subclasses can adversely affect classifier confidence and increase the demand for training samples. After deliberation with adult and pediatric neurosurgeons and pathologists, taking into consideration the clinical implications for intraoperative (sub)classification, and experience from clinical use of Sturgeon V1, we developed a labeling system consisting of 102 classes (see **Supplementary Table 1**). The most notable changes in comparison to the Heidelberg 11b4 system are a revision of IDH-wildtype high grade glioma subclasses, addition of germ cell and pituitary adenoma subclasses, addition of PATZ-fusion neuro-epithelial tumors and PLAGL1/2-amplified tumors and a split of meningiomas into subtypes.

#### Creating and expanding the training dataset by iterative self-training

*–* We assembled a training set of ∼10,000 samples from public repositories and through collaborations. The majority, however, are labeled according to the v11b4 classifier ^18^, or in broader diagnostic categories, and are therefore incompatible with the V2 class system. To harmonize these data we applied a form of iterative self-training, an established semi-supervised technique in which a classifier trained on labeled data (a ‘relabeling classifier’) assigns labels to unlabeled data that are then added to the training set of the next generation classifier ^20^.

To benchmark intermediate classifier performance, an external nanopore sequencing dataset was assembled consisting of 416 samples published by Kuschel et al. ^21^ and 61 samples published previously ^9^, all labeled with the Heidelberg 11b4 system. Importantly, these benchmark data carry labels established independently (integrated histomolecular diagnosis or Heidelberg classification of the source studies), and no benchmark sample entered training or relabeling.

#### Relabeling initiation (V2.1)

*-* To initiate the iterative relabeling, the largest and most homogeneously labeled dataset was selected, consisting of array-based methylation profiles of 7,494 CNS tumor samples, provided by the National Institutes of Health (NIH). These samples were classified using the Heidelberg 12b6 classifier (www.molecularneuropathology.org/mnp/, since retired), and only those with a high confidence score (>0.84) were retained (N=5,855). These samples were converted to the Sturgeon V2 class system by merging classes (for example by merging Benign Meningioma subclasses 1, 2 and 3 into Benign Meningioma, and merging all medulloblastoma group 3 subtypes into MB_G3 see Methods and **Supplementary Table 2**).

We also included control samples from the Capper et al. dataset, bringing the total number of samples to 6173. Performance of the V2.2 classifier on the external nanopore benchmark cohort is reported in **Figure 1e**.

**Figure 1:**
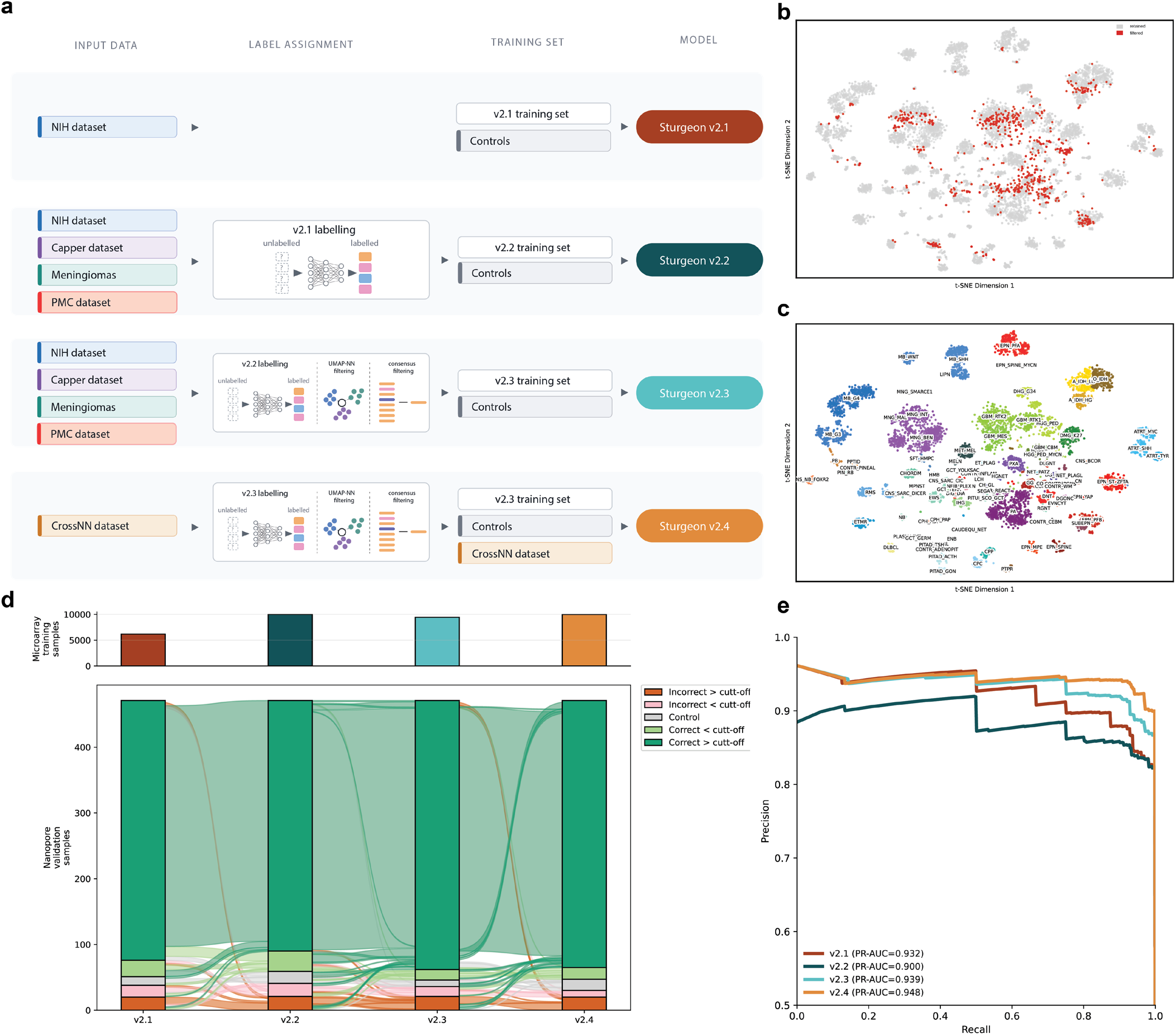
Relabeling strategy and performance on nanopore validation cohort. **a** shows a schematic overview of the relabeling schema. **b** shows a t-SNE plot of all data points that were considered, where the red data points were discarded in one of the filtering steps. **c** shows a t-SNE plot of all the labeled datapoints that were used to train Sturgeon v2.4. **d** shows an alluvial plot with the predictions of each of the Sturgeon versions on two external nanopore validation datasets ^9,21^. The cut-off was determined by maximizing the recall for a precision of 0.95. Additionally, it shows the number of samples that were used to train the respective Sturgeon version. **e** shows a precision-recall curve for each of the Sturgeon versions for the two external validation datasets.

#### Downsampled relabeling (V2.2)

*-* The V2.1 classifier was subsequently used for the first iteration of relabeling. To enhance robustness, we performed classifications on resampled data (see Methods, **Figure 1a**). To this end, per sample, 20 *in-silico* downsamplings of 50,000 CpG sites each were subjected to V2.1 classification. Samples were relabeled with the most frequent highest confidence class. We also aimed to further expand the ability to subclassify meningiomas. To this end, we included a dataset published by Choudhury et al. comprising 281 meningiomas ^22^. We additionally included 18 more clinically relevant clear cell meningioma samples defined by SMARCE1 alterations and a distinct methylation profile ^23^. Finally, 13 plexus neurofibroma samples with confirmed histopathological classification were included to support this distinct tumor subclass. We again retained all healthy control samples from the Capper 2018 dataset with unchanged labels, as well as a set of healthy meninges profiles reported in Maier *et al*. as a meninges control class ^24^. These data (N=10,012) were used to train the V2.2 classifier (**Figure 1a**). However, when benchmarked on the external nanopore data we found that naive self-training at scale underperforms and drops PR-AUC from 0.932 to 0.900 (**Figure 1e**, V2.2). The number of confidently classified samples also decreased when training on the relabeled dataset (**Figure 1d, Supplementary Figure 1a**). This is possibly due to confirmation bias, where existing label errors are propagated to the classifier ^25^, and due to the hard-labeling of low-quality samples (for example due to very low purity or technical failures). This demonstrates that simple re-training is insufficient, and more stringent filtering and additional relabeling iterations are required.

#### Filtered relabeling (V2.3)

To improve training data quality, we implemented two orthogonal concordance filters. In the first filter, we only retained samples for which at least 10 out of 20 V2.2 downsampled classifications agreed. The second filter is based on UMAP-neighborhood, where samples were removed when the most frequent of the 10 UMAP nearest neighbors was discordant with the Sturgeon V2.2 classification (**Figure 1a**). Several classes with a small sample size would be lost due to filtering; we therefore retained these classes if the classifier identified them correctly despite discordant UMAP neighborhoods (PITAD_PRL, GCT_TERA and GCT_GERM) and removed PLNTY and MYXGNT entirely due to their similarity to other low-grade glioneuronal tumors (**Supplementary Figure 2**). With this set of filters, 725 samples were removed (7.2%) and the training dataset consisted of 9,287 samples, which were used to train the next generation classifier (**Figure 1a**, V2.3). Sturgeon V2.3 showed robust improvement over the unfiltered classifiers (**Figure 1d,e**).

#### Improved classifier architecture to prevent label flips

As noted by us, as well as others ^16,17^, the original Sturgeon implementation can exhibit “class flipping”, in which the predicted class flips over time, usually between two classes. This is a real problem in intraoperative practice, where sequencing is stopped once the operator deems the classification complete and the last prediction becomes the final call: in the presence of flipping, one never knows whether the class would have flipped again had sequencing continued, rendering the final call ambiguous. The flipping effect arises because the Sturgeon V1 architecture reports the outcome of the most confident of four subclassifiers, each trained on different folds of the training data. To reduce the occurrence of label flips, we adjusted the classifier architecture. Instead of reporting the most confident of four subclassifiers, we now report the class with the highest mean score of 10 classifiers, again trained on different folds of the data (see Methods). Calibration of the 10-classifier mean score was achieved based on a hold-out set consisting of 25% of the data. We noted that calibration error correlates to the sparsity, specifically, the ensemble was underconfident at low coverage and overconfident at high coverage (Supplementary Figure 3). We adjusted the calibration accordingly using temperature scaling per coverage bin (see Methods). The most performant classifier with 10 subclassifiers and coverage bin-wise calibration was finalized as Sturgeon V2.4b and applied for further validation and implementation, and is henceforth referred to as Sturgeon V2.

#### Relabeling strategy validation and final relabeling (V2.4)

To test the efficacy of the relabeling strategy, we used 687 CNS tumor samples recently published in the context of the CrossNN classifier ^11^(GSE289137). Because the labels in this dataset were not compatible with the V2 class system, these data offered an opportunity to test if relabeling and adding these samples can improve classifier performance. Using Sturgeon V2.3 as a relabeling classifier, followed by UMAP filtering, added 620 out of 687 CrossNN samples to the training data, bringing the total number of training samples to 9,907 (**Figure 1a**). These samples were used to train Sturgeon V2.4. We found that adding this newly labeled data increased the classification performance (PR-AUC) on the external benchmark dataset from 0.939 to 0.948 (**Figure 1e**). Relabeling results are shown in **Supplementary Table 3**. These results illustrate how iterative relabeling of newly available data is a viable method that can be applied for future development of methylation-based classifiers.

### Sturgeon V2 validation

#### Performance on in-silico downsampled microarray data

To prepare for clinical implementation, we designed a suite of validation tests to be executed and logged prior to deploying the classifier as an in-house developed test. The first test evaluates performance on the hold-out fold of the crossvalidation. Overall, classification performance was high, with minor misclassifications limited to closely related subclasses of neuroepithelial tumors (NET) and pituitary adenomas (PITAD) (**Supplementary Figure 4a,b**). Subsequently, we test the classifiers on a series of 177 Illumina array methylation profiles generated at the Princess Máxima Center covering a wide range of primary CNS tumor resections.

Importantly this series was not used as training data. To evaluate performance, we simulated 400 sparse profiles for each sample, distributed across sequencing depths of 1000, 2500, 5000, and 15000 CpG sites (drawn from the 357,738 common probes), and subsequently applied the classifier to each individual sampling. Applying Sturgeon V2 to this set results in 50,332 out of 70,800 confident (>0.95) classifications, out of which 47,895 (95.1%) were correct, 1,978 (3.9)% were incorrect and 459 (0.9%) were confidently classified as a control class (**Supplementary Figure 5 and 6**). It should be noted that misclassifications are not evenly distributed over the samples; in 163 samples 74% of calls were confident, and only 0.5% of confident calls were incorrect, while in the 14 challenging samples only 32% of calls were confident, but 94% of confident calls were incorrect. Importantly among the 14 difficult samples, four Diffuse midline glioma, H3 K27-altered (which were also challenging to classify using the Heidelberg and Bethesda classifier) were misclassified as MYB altered diffuse gliomas.

To further investigate these challenging samples, we also performed nanopore sequencing on DNA extracted from the three samples for which this was available. In the nanopore sequencing derived data two of these were classified at low confidence and one as control tissue **(Supplementary Figure 7a)**.

We conclude that these samples were challenging due to a low tumor fraction, which is corroborated by histological assessment. While unlikely to result in misclassifications on nanopore data, caution is advised when running Sturgeon V2 on methylation array data of low purity samples.

#### Performance on retrospective nanopore data

We next compared Sturgeon V2 to Sturgeon V1 in a series of 24 samples sequenced deeply with nanopore sequencing. This amount of data enabled *in-silico* downsampling each sequencing run to a range of 663 to 25,000 CpG sites, representing virtual sequencing times. At each “timepoint” we collected performance metrics. Strikingly, Sturgeon V2 shows a higher recall rate (87.9% over all timepoints), whilst making only very few (<0.1%) errors (**Supplementary Figure 7b**).

#### Performance on low-purity samples

We note that the retrospective nanopore samples were retrieved from a biobank and were selected for high sample purity. True intraoperative samples, on the other hand, undergo limited selection based on frozen section microscopy. Therefore, we also tested robustness against sample impurity by artificially mixing data between a control class and a tumor class. We find that mixed samples are unlikely to be misclassified, and that tumor fractions <50% are increasingly less likely to result in high confidence classifications (**Supplementary Figure 7c)**.

#### Performance on intraoperative data

We retrospectively assessed the ability of Sturgeon V2 to provide intraoperative diagnoses in a clinical setting. For this we leveraged our recently published cohort of 94 intraoperative cases from the Princess Máxima center ^17^. In each case we compared the performance of Sturgeon V1 and Sturgeon V2 at each timepoint (all classifier output is shown in **Supplementary Data 1**, for all patients, two examples are shown in **Figure 2 a** and **b** and an overview shown in **Figure 2c**). In 76 cases, both classifiers yielded a confidently correct final diagnosis. In 12 cases it was not possible to provide a diagnosis using Sturgeon V1. Strikingly, in 9 out of these 11 cases (Patient 16, 26, 38, 50, 52, 58, 61, 72, 80) Sturgeon V2 achieved a correct diagnosis. In the remaining 3 cases the classification of Sturgeon V2 remained unclear (Patient 27, 42, 94). Sturgeon V1 produced one error, classifying Patient 51 as MNG instead of SFT. Sturgeon V2, on the other hand, did not reach high confidence for this sample. The 3 cases where Sturgeon V1 classified as a “broadly correct” outcome (Patient 2: EPN PF, B instead of EPN PF, A, Patient 34: MB, Group 3 instead of MB, Group 4 and Patient 49: ATRT, TYR in an ATRT with SMARCB1 somatic mutations that is classified ATRT, SHH using RNA classifiers and ATRT, TYR by methylation-based techniques), Sturgeon V2 predicted the correct class (EPN_PFB) in patient 2, Healthy Cerebellum in Patient 34 and ATRT-TYR in Patient 49. One case (Patient 54) was classified correctly as MB, Group 3 by Sturgeon V1 and in early timepoints as MB_G4 by Sturgeon V2, and unclear at later timepoints. Patient 94, a PA was confidently misclassified as DNT by Sturgeon V2 (but only for three successive timepoints). Two cases were correctly classified by Sturgeon V1 and unconfident by Sturgeon V2 (Patient 59 and 68). The inclusion of additional classes correctly classified one sample in this cohort (NET_PATZ Patient 16) that was not classifiable in the 11b4 classification system. Choroid plexus tumors were revised to Choroid Plexus Carcinoma and Choroid Plexus Papilloma (CPC and CPP, respectively), rather than the previously used Type A and B pediatric choroid plexus tumors. The five Choroid plexus cases (Patient 8, 40, 60, 71, 90) present in this cohort were correctly classified.

**Figure 2:**
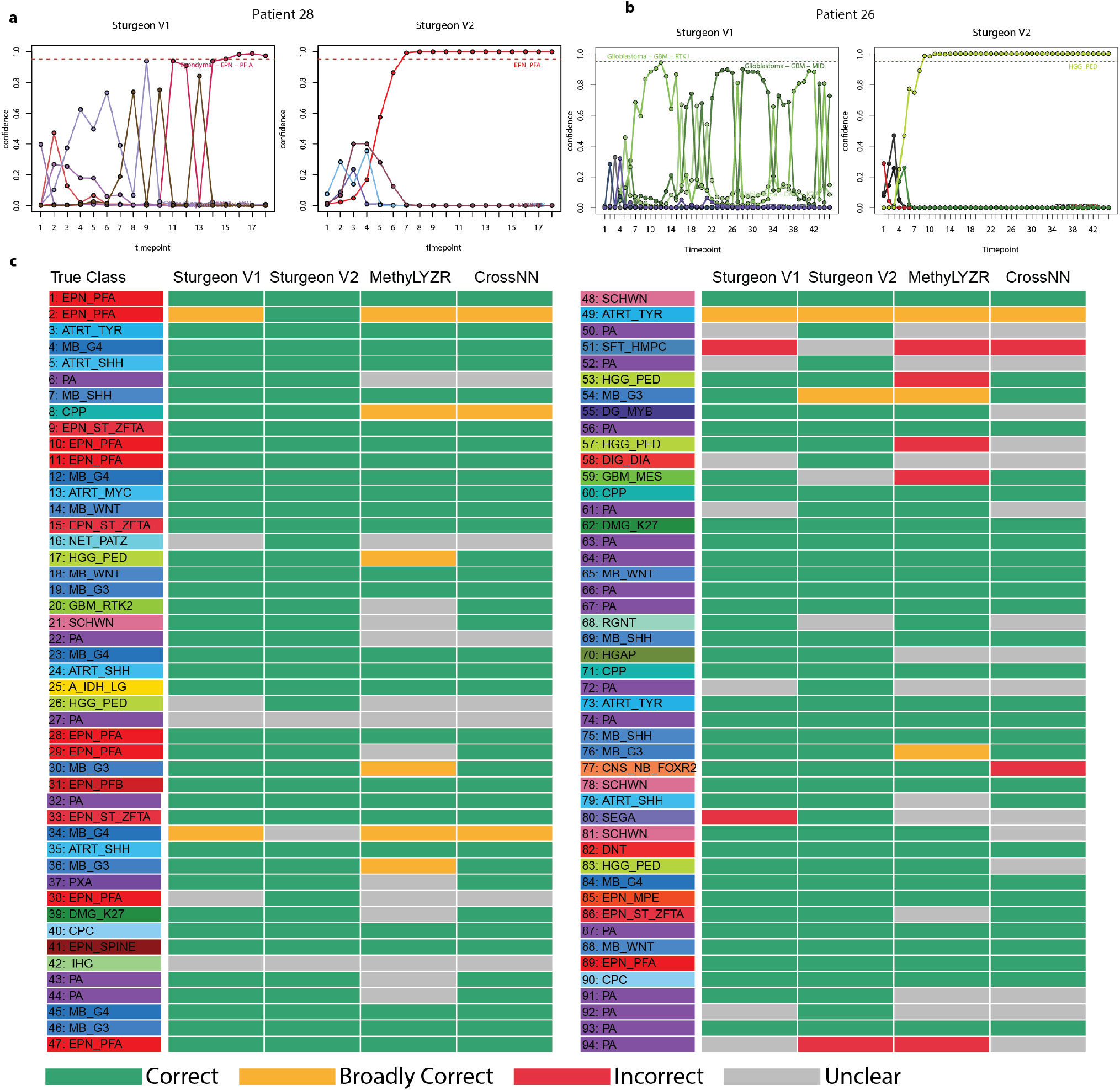
Sturgeon V2 performance in a retrospective cohort. **a** and **b** show reproduced classifier confidence traces from intraoperative data for two cases where Sturgeon V1 showed “label flipping” behavior. The updated architecture of Sturgeon V2 results in traces where the confidence increases almost monotonically. **c.** Results of the 94 reproduced intraoperative sequence runs, rows indicate patients and columns indicate classifier performance, green indicates confidently correct classifications, yellow indicates broadly correct classifications (ie. a wrong subclass), red indicates incorrect classifications and grey indicates unclear results. We applied a 2-iteration decision window for these results: the classifier was required to be confident for at least two successive timepoints.

Overall Sturgeon V2 was confidently correct in 85 out of 94 cases, broadly correct in 2 out of 94 cases and incorrect in one case. This is a marked improvement over Sturgeon V1, which was confidently correct in 78 cases, broadly correct in 3 cases and incorrect in two cases. **Benchmarking to other available methylation-based CNS classifiers**

In addition to the Sturgeon classifiers, MethyLYZR and CrossNN are publicly available and can thus be systematically benchmarked. We compared the performance of all four classifiers on the same intraoperative cohort used above ^17^. It should be noted that comparison between classifiers on intraoperative data is challenging, as in practice the final classification is made by considering the whole ‘classifier trace’, i.e. the classifier confidence over sequencing time. Consider for instance the trace of Patient 22. Here, MethyLYZR provides a stable confidence for the correct class, yet slightly below the confidence threshold. Strictly speaking this represents an unclear outcome, but in practice the operator may still make a (in this case correct) call. We provide all classifier traces in **Supplementary Data 1**, to facilitate manual inspection.

We moreover note that the intraoperative cohort was generated in an intraoperative setting where Sturgeon V1 was used for real-time classification. Sequencing durations were therefore influenced by the Sturgeon V1 confidence. This provides a slight advantage relative to the other classifiers, which were only run post-hoc; possibly some classifiers would have reached confidence with longer sequencing. For the benchmark, we used the advised confidence cutoffs of 0.6 and 0.2 for MethyLYZR and CrossNN, respectively, with a decision window of two consecutive timepoints (**Figure 2c**).

In the majority of cases, all four classifiers are in agreement and correct, with minor differences in speed, where MethyLYZR often provides a high-confidence classification in the very early timepoints (**Figure 2c, Supplementary Data 1**). A striking and relevant difference between classifiers is the recall rate (i.e. the number of cases where the confidence threshold is reached for the correct class); Sturgeon V2: 85, Sturgeon: 79, MethyLYZR: 56 and CrossNN: 65. The number of cases where no confident diagnosis is reached varied, Sturgeon V2: 7, Sturgeon: 11, MethyLYZR: 24 and CrossNN: 22. Sturgeon V2 made one confident misclassification, Sturgeon made two confident misclassifications, while MethyLYZR and CrossNN made 5 and 3 misclassifications, respectively (**Figure 2c**). We also applied MethyLYZR and CrossNN to the 1,880 simulated sequencing runs, to more extensively compare classifier performance (**Supplementary Data 2**). Using this benchmark we also compared the effect of different windowing strategies. Also for MethyLYZR and CrossNN, using a 2-timepoint window reduces the error rate while only modestly affecting the recall (**Figure 3e and f**). When comparing performance between all four classifiers in this dataset, we note that MethyLYZR is faster than the others, but with a substantially lower recall and higher error rate, Sturgeon and CrossNN perform similarly, with Sturgeon reaching a slightly higher recall at later timepoints. Notably, as sequencing duration is influenced by the Sturgeon V1 confidence used in the intraoperative setting in which these data were collected, the higher recall may be caused by this slight advantage of Sturgeon V1. Sturgeon V2 is slower than the other classifiers (at 2,000 CpG sites, Sturgeon V2 accurately classified 288 out of 1,880 simulations, versus 276, 902 and 764 in Sturgeon V1, Methylyzr and crossNN respectively), but in this cohort reaches the highest recall and lowest error rates at the later timepoints: at 6,000 covered sites, Sturgeon V2 correctly classifies

**Figure 3:**
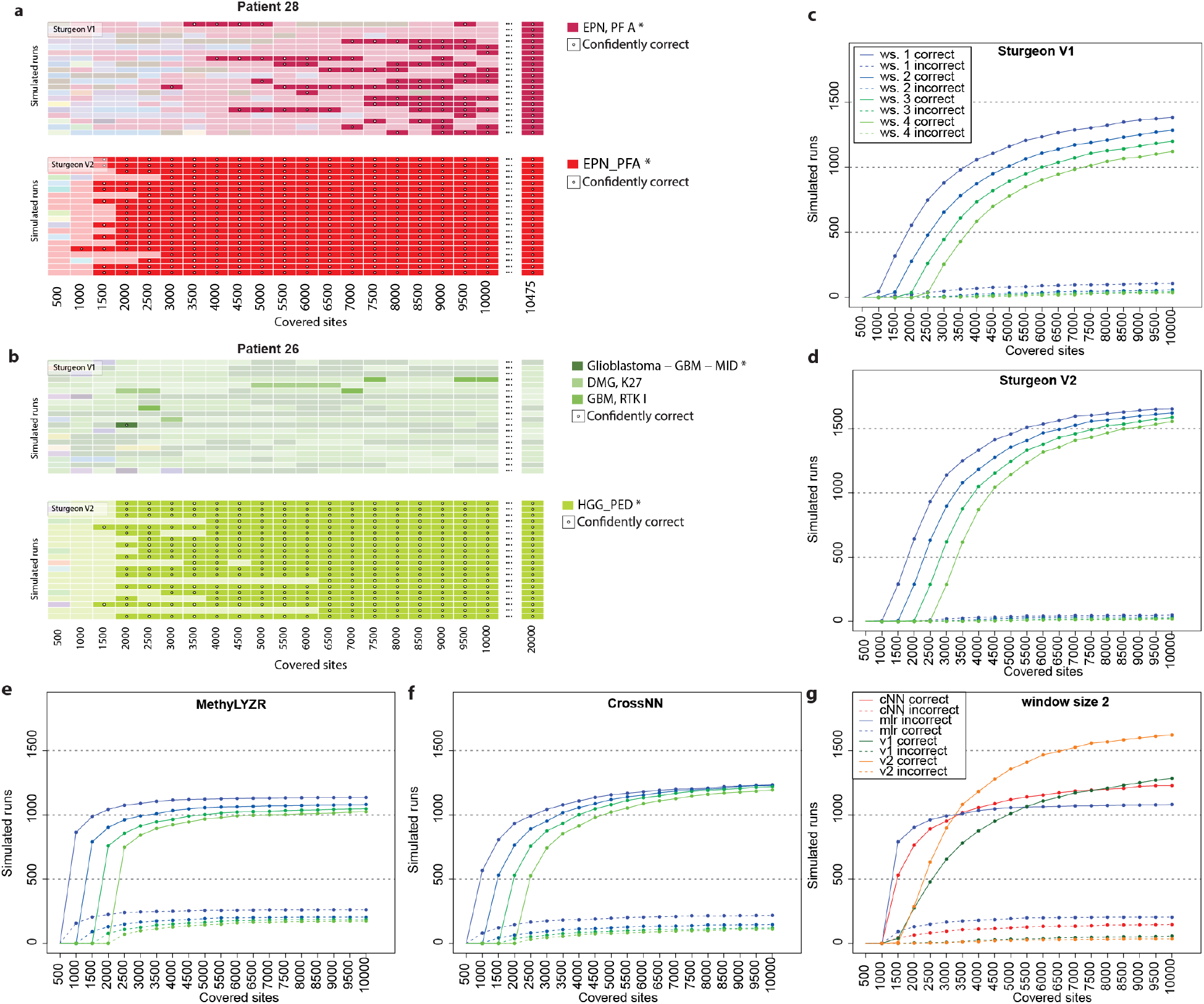
Benchmarking results. **a** and **b:** the resampled sequence runs in two samples, where each row represents a simulated sequence run, and each column represents a sequence depth. Saturated colors represent a class with high confidence (>0.95), desaturated colors represent underconfident classifications, dots represent confident correct calls. **c-f** The effect of using different decision windows for each classifier on the 1,880 in silico resampled sequence runs, solid lines indicate the number of correct classifications and dashed lines indicate the number of misclassifications when strictly adhering to the indicated decision windows and confidence thresholds (Sturgeon V1 and V2: 0.95, MethyLYZR 0.6 and CrossNN 0.2). Colors indicate different decision window sizes, blue to green: window size 1 to 4) **g.** Comparison of classifier performance at decision window size of 2 iterations (red: CrossNN, blue: MethyLYZR, green: Sturgeon V1, yellow: Sturgeon V2, solid lines: Correctly concluded simulated runs, dashed lines: Incorrectly concluded simulated runs). 1467 simulations, versus 1,108, 1,063 and 1,154 in Sturgeon V1, MethyLYZR and CrossNN respectively. (5000 CpG sites are sequenced in approximately 10-15 minutes on a promethION and 20-30 minutes on a minION) (**Figure 3g)**.

### Intraoperative Meningioma classification

In the previously used Heidelberg V11b4 class system, Meningiomas were collectively classified as a single class. However, meningiomas can usually already be diagnosed before surgery based on MRI. For intraoperative decision-making, it is imperative to identify subclasses according to the more recent molecular classification systems. In current practice multiple classification systems exist with different levels of granularity and overlap. In general, three main molecular groups are defined: *NF2-*wildtype low-risk, *NF2-*altered low-risk and *NF2-*altered high-risk meningioma ^26^. Based on the methodology used, different models have distinct levels of granularity and may add an intermediate risk group ^22,27–29^.

Currently no definitive consensus class system for meningiomas is established and clinical use is mostly defined by local preferences ^30^. Subclasses may not be as distinct and may reflect a risk continuum ^26^, which leaves the subclass definitions up for debate. In the Heidelberg/DKFZ classification systems MNG was first split into benign, intermediate and malignant meningioma classes, and into subclasses: benign 1 (*NF2-*altered low-risk), benign 2 (*NF2-*wildtype low-risk), benign 3 (*NF2-*mixed low-risk), intermediate A and B (both mostly *NF2-*altered) and malignant (*NF2-*altered high-risk) ^27^. The UCSF system splits MNG into Merlin (the protein encoded by *NF2*) Intact, Immune enriched (*NF2-* altered low-risk) and Hypermitotic (*NF2-*altered high-risk) ^22^.

For the Sturgeon V2 classification system we did not seek to incorporate and support one single system. While we opt to use the “simplified Heidelberg system” with benign, intermediate and malignant meningiomas by default, we developed and evaluated four different classifiers (available through Zenodo), identical in training and architecture, with the exception of the meningioma subclass definitions (**Figure 4a**). Three of these follow established entity-based subtyping: the full 6-subclass Heidelberg system, the simplified 3-class Heidelberg system (Benign, Intermediate and Malignant), and the UCSF system. For the fourth, we explored whether prognostic risk can be inferred from methylation data alone, by training directly on the integrated risk score (IntS: Low, Medium or High). The IntS is a prognostic model that combines histological WHO grade, methylation family and copy number variations (losses of 1p, 6q and 14q) into a three-tiered risk score, and outperforms purely molecular models in predicting progression^31,32^. For the development of Sturgeon V2 meningioma classifiers, we labeled the Choudhury dataset, consisting of 281 meningioma samples in all four classification systems. We found the expected differences and overlaps between class systems (**Figure 4a**)^22,27,32^.

**Figure 4:**
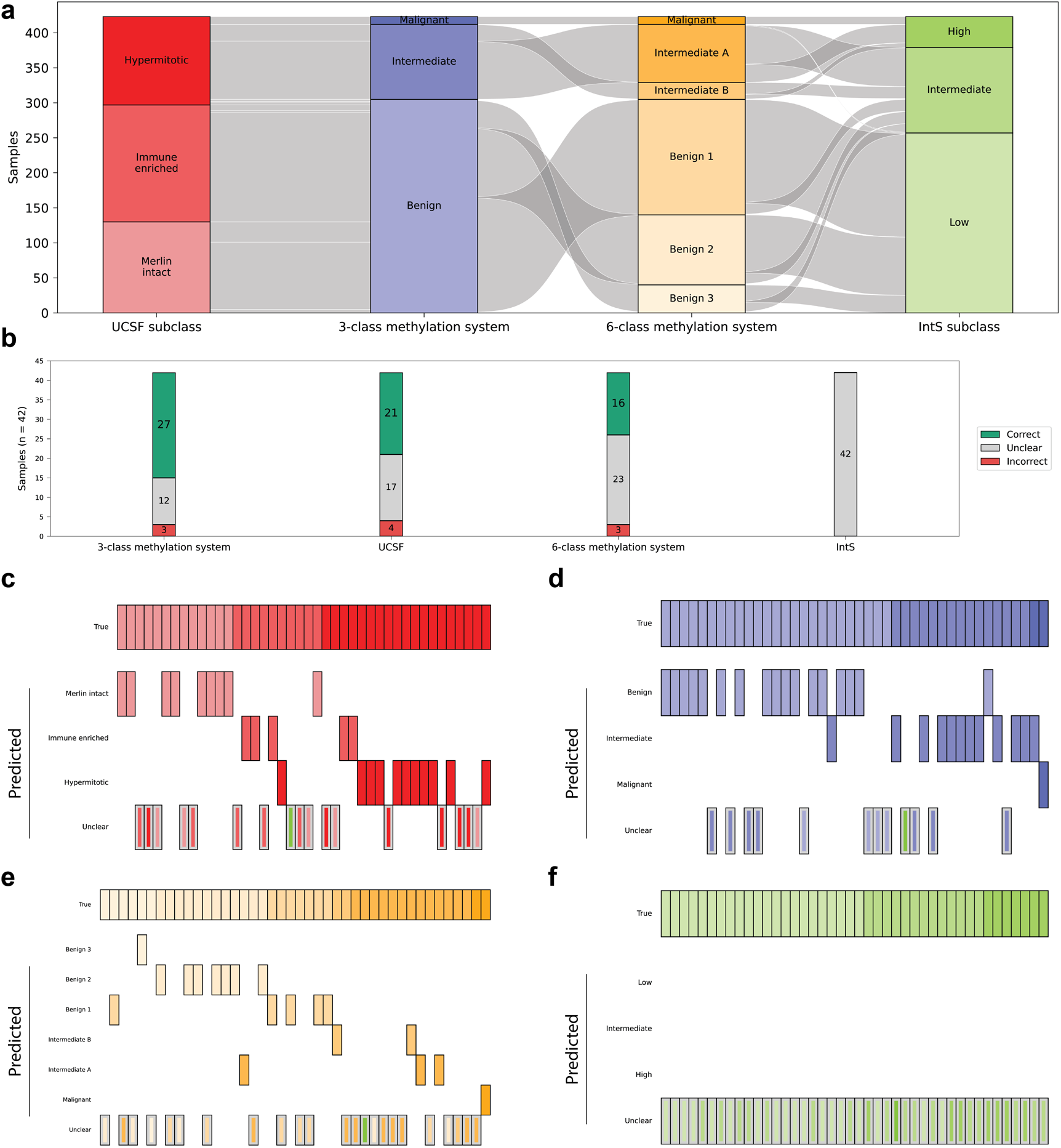
Performance with different meningioma classification systems. **a.** Class overlap in the reference cohort of 423 meningioma array samples which were labeled in the UCSF, IntS, 3-class and 6-class Heidelberg systems. **b**. shows the performance of Sturgeon V2 trained with different meningioma classifications systems on an intraoperative nanopore validation set. **c** shows a sample by class confusion matrix of Sturgeon V2 trained with the UCSF system on an intraoperative nanopore validation set. **d**. shows a sample by class confusion matrix of Sturgeon V2 trained with the 3-class Heidelberg system on an intraoperative nanopore validation set. **e**. A sample by class confusion matrix of Sturgeon V2 trained with the 6-class Heidelberg system on an intraoperative nanopore validation set. **f**. A sample by class confusion matrix of Sturgeon V2 trained with the IntS system on an intraoperative nanopore validation set.

We evaluated the ability of the four classifiers to differentiate meningioma subtypes in a cohort of 43 intraoperatively sequenced meningiomas. Outcomes were compared against the integrated histomolecular diagnosis, supplemented with Heidelberg V12.8 classifications on Illumina EPIC arrays performed post-operatively where available. Performance was generally high for the 3-class Heidelberg and UCSF systems. The 6-class Heidelberg system showed occasional confusion between subclasses within the same class (**Figure 4b,c,d,e**), that is, errors remained within the clinically relevant risk category. Performance was markedly lower for IntS prediction, with few samples reaching the confidence threshold and misclassifications largely restricted to adjacent tiers (low versus intermediate, intermediate versus high) rather than between extremes (**Figure 4b,f**). This is consistent with the composite nature of the IntS, which incorporates histological grade and copy number status alongside methylation family; the methylome alone appears to carry enough signal to place samples in the approximate risk range, but not to reproduce the integrated score with the confidence required for intraoperative use.

## Discussion

Here we present Sturgeon V2, a classifier for sparse methylation profiles that combines a substantially expanded training set, an updated and clinically oriented class system, and a redesigned architecture with coverage-dependent calibration. In a retrospective pediatric intraoperative cohort, Sturgeon V2 provided a confident and correct diagnosis in 85 of 94 cases, including 9 cases that could not be diagnosed with Sturgeon V1, and reduced the error rate among confident calls in simulated sequence runs from 7.1% to 2.8%. Benchmarking against the publicly available alternatives showed the highest recall and lowest error rate at clinically realistic sequencing depths.

The required level of diagnostic granularity depends strongly on the intended clinical use. Sturgeon V2 is designed for intraoperative diagnostics, prioritizing rapid and robust classification from limited sequencing data within a clinically actionable timeframe. For this purpose, classification at the level of the major CNS tumor type or molecular group is often sufficient to inform surgical decision-making. Sturgeon V2 substantially broadens the range of tumors that can be recognized, including recently described pediatric entities such as PATZ1 fusion-positive and PLAGL1/2-amplified tumors. However, Sturgeon V2 does not resolve all finer molecular subtypes, including selected subtypes of pediatric-type diffuse high-grade glioma, medulloblastoma groups 3/4 and pineoblastoma. Such granularity may be important for definitive diagnosis, prognostication or clinical trial stratification, for which comprehensive methylation classifiers such as Heidelberg or Bethesda remain complementary. For the large proportion of CNS tumors in which further subclassification does not provide additional clinically relevant information, however, Sturgeon V2 already offers classification at a diagnostically sufficient level and may provide an efficient alternative to comprehensive array-based methylation profiling. This may be particularly valuable in low-resource settings, including LMICs, where access to comprehensive methylation-array diagnostics and the required laboratory infrastructure is limited.

Analysis of the classification of meningiomas demonstrates why limiting subclassification level is important for intraoperative performance: Classifiers with a three-class meningioma definition performed well, whereas the six-class Heidelberg system confused subclasses within the same risk category. Direct prediction of the integrated risk score performed markedly worse, indicating that multi-modal classification that is based on histological information and copy number profiles may be required for intraoperative prediction of prognostic risk.

The central methodological contribution is the iterative relabeling strategy. Assembling reference data across sources is limited less by data availability than by label compatibility: methylation profiles are annotated in different class systems, at different levels of granularity, and with varying reliability. To address this, our strategy leverages the ability of Sturgeon-like classifiers to classify sparse data: by repeatedly sparsifying a sample and requiring concordance across these subsamples, labels can be assigned to samples that resemble the training distribution only partially. We find, however, that naive self-training at scale degrades performance, because label errors and low-quality samples are propagated into the next generation of the classifier. Two orthogonal concordance filters, one based on the classifier itself, one on the geometry of a UMAP projection, were sufficient to reverse this. We conclude that data harmonization efforts must strike a balance between too lenient inclusion, which leads to underperformance, and too strict inclusion, which leads to underconfident classifiers that only classify the clearest of cases.

Given the existence of multiple publicly available sparse methylation classifiers, running them in parallel is a viable option to generate a consensus call ^16^. However disagreement between classifiers poses a challenge that requires clear guidelines to prevent operator bias. In pediatric intraoperative setting in the princess Máxima center we therefore use a single classifier with a defined confidence threshold.

All classifiers are constrained by the coverage of their training data. A remaining challenge for methylation-based classification are rare tumors with mixed or atypical phenotypes and/or atypical epigenetic profiles (i.e. “exotic” cases) which are absent from the reference set. Reference data spanning diverse, including rare, tumor types are therefore essential. Increasingly, these reference data will be generated on nanopore and Illumina sequencing platforms rather than arrays, and classification systems will continue to be improved. The strategy presented here accommodates both: retraining a revised class system requires no manual re-annotations, and newly published datasets can be incorporated by re-running the relabeling and filtering procedure, as demonstrated with the CrossNN cohort. Sturgeon V2 thus provides not only an improved classifier, but a framework for keeping intraoperative methylation-based diagnostics current as reference data and classification systems evolve. Sturgeon V2 is now deployed in our intraoperative diagnostic workflow, where fewer unclear cases directly translates into better-informed surgical decisions.

## Data Availability

All data produced in the present study are available upon reasonable request to the authors

## Author declarations

J.d.R. is co-founder of Cyclomics BV, a genomics company. C.V., M.P.G and J.d.R. are named inventors on a patent application describing the Sturgeon technology, licenced to Cyclomics BV.

## Data availability

Capper et al. dataset: GEO GSE109381

CrossNN dataset: GEO GSE289137

Kuschel et al. dataset: GEO GSE209865

Choudhury et al.: GEO GSE183647

Sie et al. cohort: ArrayExpress E-MTAB-16333

Methylation array data from Vermeulen, Pages-Gallego et al.: EGAD00010002599

Data available on request:

NIH data: Kenneth Aldape, NIH methylation unit, Martha Quezado

Meninges control data: Andrea Maier and Tiit Mathiesen Máxima validation arrays: Carlo Vermeulen Plexus Neurofibroma: Juergen Hench

## Software availability

The Sturgeon V2 inference software is available via github (https://github.com/deRidderLab/sturgeon-v2) for non-commercial use, and the model weights are avaible via Zenodo (https://doi.org/10.5281/zenodo.22899108). Sturgeon V2 is also compatible with the Epinn software available via Cyclomics bv. and an R-shiny app available through github: https://github.com/bstikker/Shiny_Sturgeon.

## Methods

### Methylation array processing

Raw IDAT files were processed with minfi ^33^ to obtain beta values for each CpG site. Analysis was restricted to the 357,738 probes common to the Illumina HumanMethylation450 (450K), Infinium MethylationEPIC v1.0, and Infinium MethylationEPIC v2.0 arrays. To train the classifier on sparse inputs, we simulated low-coverage profiles like in Vermeulen *et al*. ^9^. For each sample, we sampled a random subset of CpGs without replacement, with the subset size drawn uniformly from 2,500 to 26,500. Each retained beta value was then binarized by drawing from a Bernoulli distribution with the beta value as its success probability. Which diverges from the approach presented in Vermeulen et al. where reads were simulated uniformly across the genome instead.

### Self-training relabeling classifier

An initial relabeling classifier (Sturgeon V2.1) was trained on 10 million sparse nanopore samples simulated from 5,855 CNS tumor samples used in the Bethesda classifier from the NIH and control samples from the Capper et al. dataset. This dataset was labeled with the Heidelberg v12b6 classifier, which was mapped to our new V2 class system (see Supplementary Table 2). Only samples with a high (>0.84) confidence classification by the Heidelberg V12b6 classifier were retained. The Sturgeon V2.1 model is a 1-fold version of the architecture that was published by Vermeulen et al. 2023.

### Two-tier concordance filtering

The V2.2 classifier was trained on the full 10,012 sample trainingset, where every sample was retained. The labels for each sample were based on the classification of the V2.1 classifier in 20 *in-silico* downsamplings of 50,000 sites, with the most frequent highest confidence classification accepted as the label. The only exception were the control classes which were retained with their original labels regardless of V2.1 classification. Results from V2.2 benchmarking revealed additional filtering of training samples was required, prompting us to implement two orthogonal filtering steps. In the first filter, samples were retained only if the majority class was called in at least 10 of 20 subsamples.

The second filter removes samples that are discordant with the majority of the sample’s 10-nearest neighbors in the 2D UMAP projection (computed on 50,000 randomly sampled CpG sites). The filtered dataset (N=9,287) was used to train the Sturgeon V2.3 classifier. The final Sturgeon V2.4 classifier was obtained by training once more on a dataset appended with the CrossNN samples (N=9,907), relabeled using the Sturgeon V2.3 classifier .

### Concordance filtering exceptions

After UMAP-projection-based filtering, five classes (PLNTY, MYXGNT, PITAD_PRL, GCT_GERM, GCT_TERA) retained fewer than 8 samples, the minimum we required for classifier training. For PITAD_PRL, GCT_TERA and GCT_GERM, the downsampled classifications were consistently correct despite discordant UMAP neighborhoods; for these classes we therefore waived the UMAP filter and relied on the downsampled-classification filter alone. PLNTY and MYXGNT were difficult to classify robustly by both the downsampled classifications and the UMAP nearest neighbors, with which they clustered with GG and DNT, respectively (**Supplementary Figure 2e**), and were thus removed from the dataset.

### Classifier self-training external benchmarking

Each Sturgeon V2.(1-4) classifier was benchmarked in an external nanopore sequencing dataset consisting of 416 samples published by Kuschel et al.^21^ and 61 samples published previously ^9^, all labeled with the Heidelberg 11.4 system (**Figure 1e**). For each sample we calculated the average softmax probability across 10 subsamples of 20,000 random CpG sites. The precision-recall curve was calculated as the macro-average of the one-vs-rest curves for each subclass.

### Classifier training and calibration

The dataset was first split into 4 folds.One fold was held out for testing, and the remaining 3 folds were used for training and validation.From these 3 folds, we generated 10 different splits, each using 2 folds for training and 1 fold for validation. We applied the same architecture that was proposed by Vermeulen et al. with SiLU activation. Each submodel was trained for 100 epochs of 100,000 simulation sparse nanopore samples (2,500-26,500 CpG sites). We applied a learning rate of 0.0001 and a batch size of 256. The resulting 10 models were used as an ensemble with average pooling on the output softmax layer.

### Classifier calibration

The ensemble logit layers are calibrated with a bin-based temperature scaling approach. Specifically, the CpG site coverage range was subdivided into bins of 1,000. Each of these bins was calibrated on the held out set as proposed by Guo et al. using 10,240 simulated samples using the LBFGS algorithm for a max 1,000 iterations. Coverage-based bin calibration was applied to ensure equal calibration even at longer sequencing times.

### Meningioma validation

Four specialized Sturgeon V2 models were trained on different meningioma classification systems (3-class methylation, 6-class methylation, UCSF, IntS). For this purpose, the 423 meningioma methylation array samples from Choudhury et al. were labeled with all four systems. A cohort of 43 intraoperatively nanopore sequenced meningioma samples was used to test the performance of these models. Each sample was classified by each model using 10 subsamples of 20,000 random CpG sites, and the consensus class was compared against the post-operative integrated molecular diagnosis.

### Benchmarking on simulated sequence runs

To benchmark classifier performance at different sequence depths we simulated sequence runs. The simulation algorithm runs for 20 iterations, and at each iteration samples 500 new CpG sites from the total number of sequenced sites up to a total of 10,000 sites. These 500 sites are combined with the sites selected at earlier iterations, and presented to each classifier for classification. Finally, a classification is performed on 20,000 random CpG sites, or the maximum available. We used the recommended confidence cutoffs for High (>0.95) and Medium (0.8-0.95) confidence for Sturgeon V1 and Sturgeon V2, >0.6 for MethyLYZR and >0.2 for CrossNN. For fair comparison of performance metrics and windowing strategy we used a cutoff of >0.9 for Sturgeon V1 and 0.95 for Sturgeon V2 to call a confident classification.

## Table legends

**Supplementary Table 1: Conversion of Heidelberg 12.6 class system to the Sturgeon V2 class system.**

*Column 1: Sturgeon V2 classes, Column 2: Heidelberg 12.6 classes.*

**Supplementary Table 2: NIH dataset conversion for Sturgeon V2.1 training**. Column 1: IDAT identifier, column 2 Heidelberg V12.6 class, Column 3: Heidelberg V12.6 confidence score, column 4: True/ False indicating whether a sample was used for Sturgeon V2.1 training, column 5: Label used for Sturgeon V2.1 training.

**Supplementary Table 3: Relabeling result for Sturgeon V2.4.** Final_label: Label in the Sturgeon V2 class system, Description: Class description, Original label: Original labels of samples that constitute the class as provided by the available metadata. N_retained: Number of samples retained for training, n_discarded: Number of samples removed by filtering. Class_total_retained: Total number of samples that constitute the class.

## Supplementary Figures

**Supplementary Figure 1:**
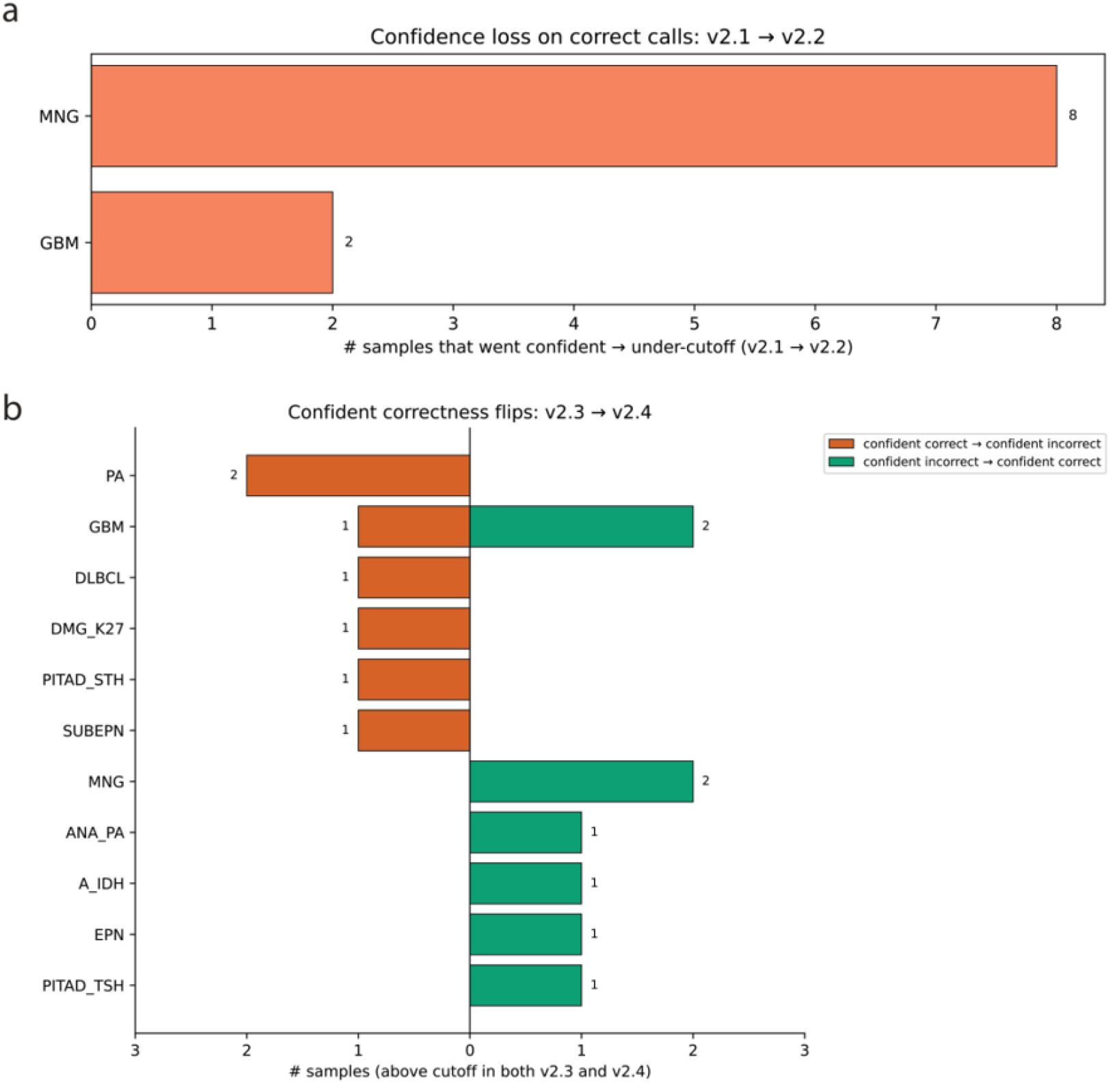
Relabeling effect on model errors. **a**. Barplot with samples that were confidently predicted as a specific subclass in Sturgeon v2.1 and became underconfident in Sturgeon v2.2 **b**. Barplot showing the change in correctness for confidently predicted samples between Sturgeon v2.3 and Sturgeon v2.4.

**Supplementary Figure 2:**
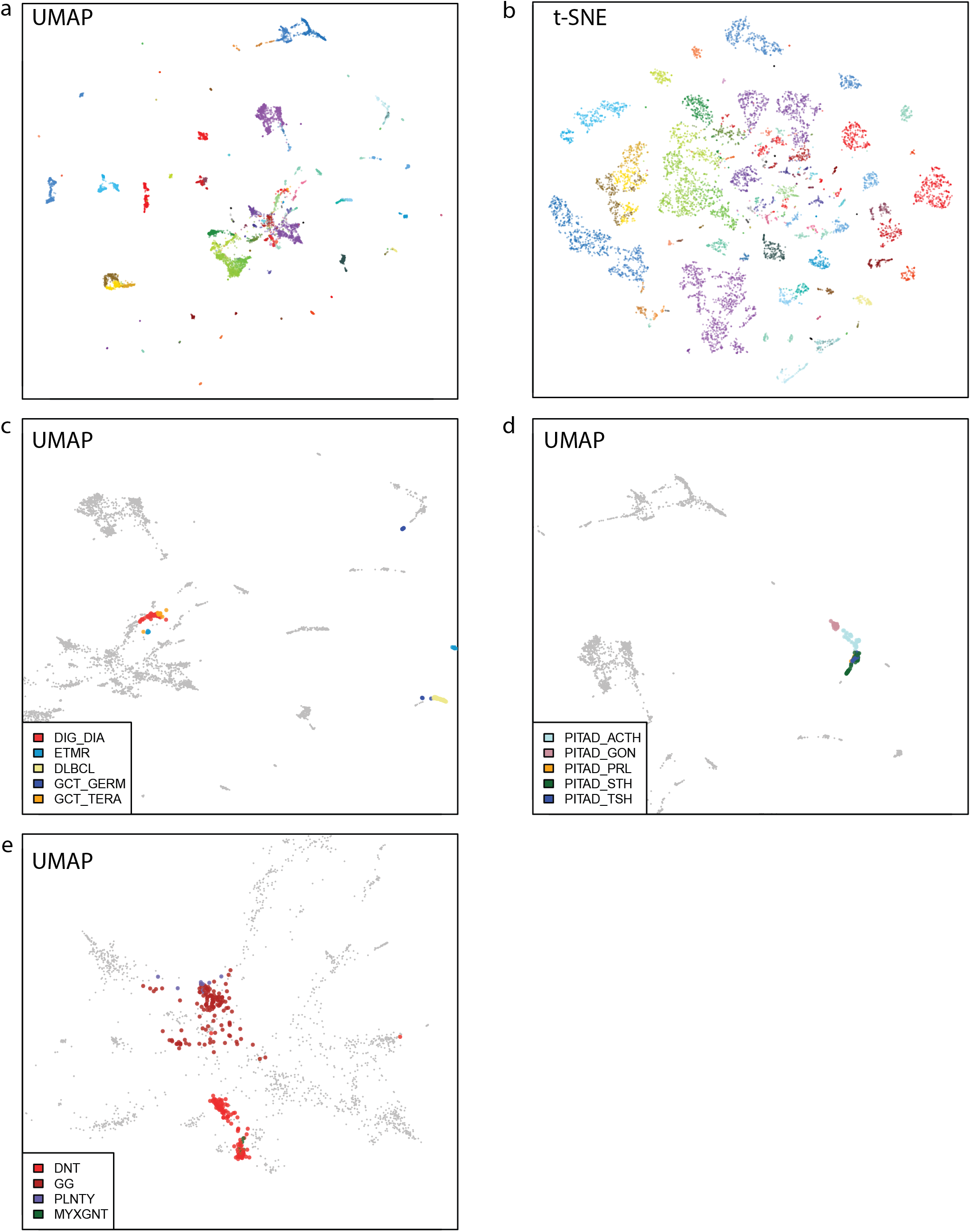
low-dimensional representations of dataset filtering. Umap (**a**) and t-SNE (**b**) projections of the Sturgeon V2 dataset, using 50,000 randomly selected probes. Zoomed-in views of the UMAP projection of **c**. Germ-cell tumors. **d**. Pituitary Adenomas **e**. PLNTY and MYXGNT tumors within the cluster of low-grade gliomas.

**Supplementary Figure 3:**
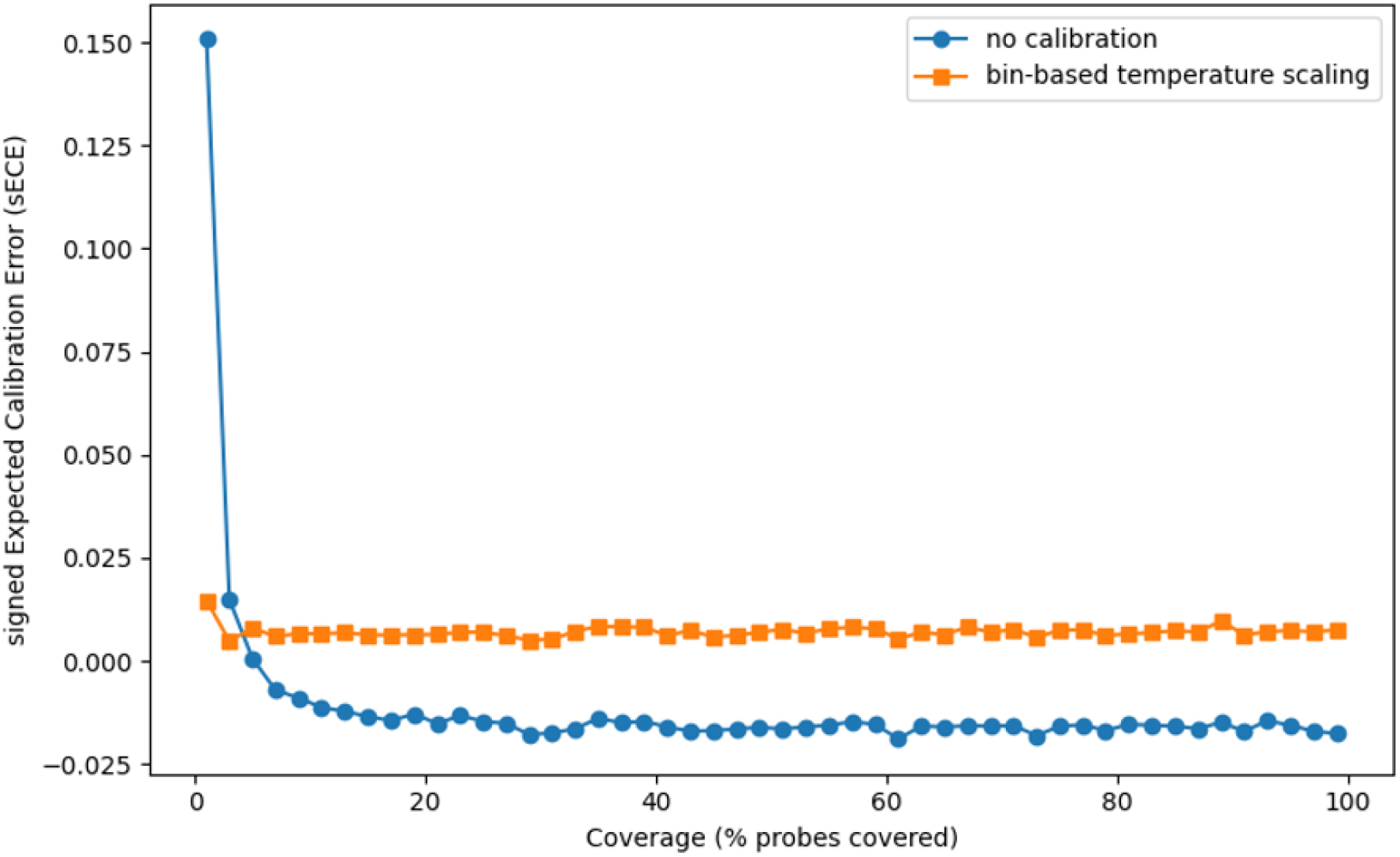
Effect of bin-based calibration across coverage range on hold-out test set. Signed expected calibration error (sECE) shows the non-absolute version of the more commonly used ECE. Positive sECE values denote underconfidence. Negative sECE values denote overconfidence.

**Supplementary Figure 4:**
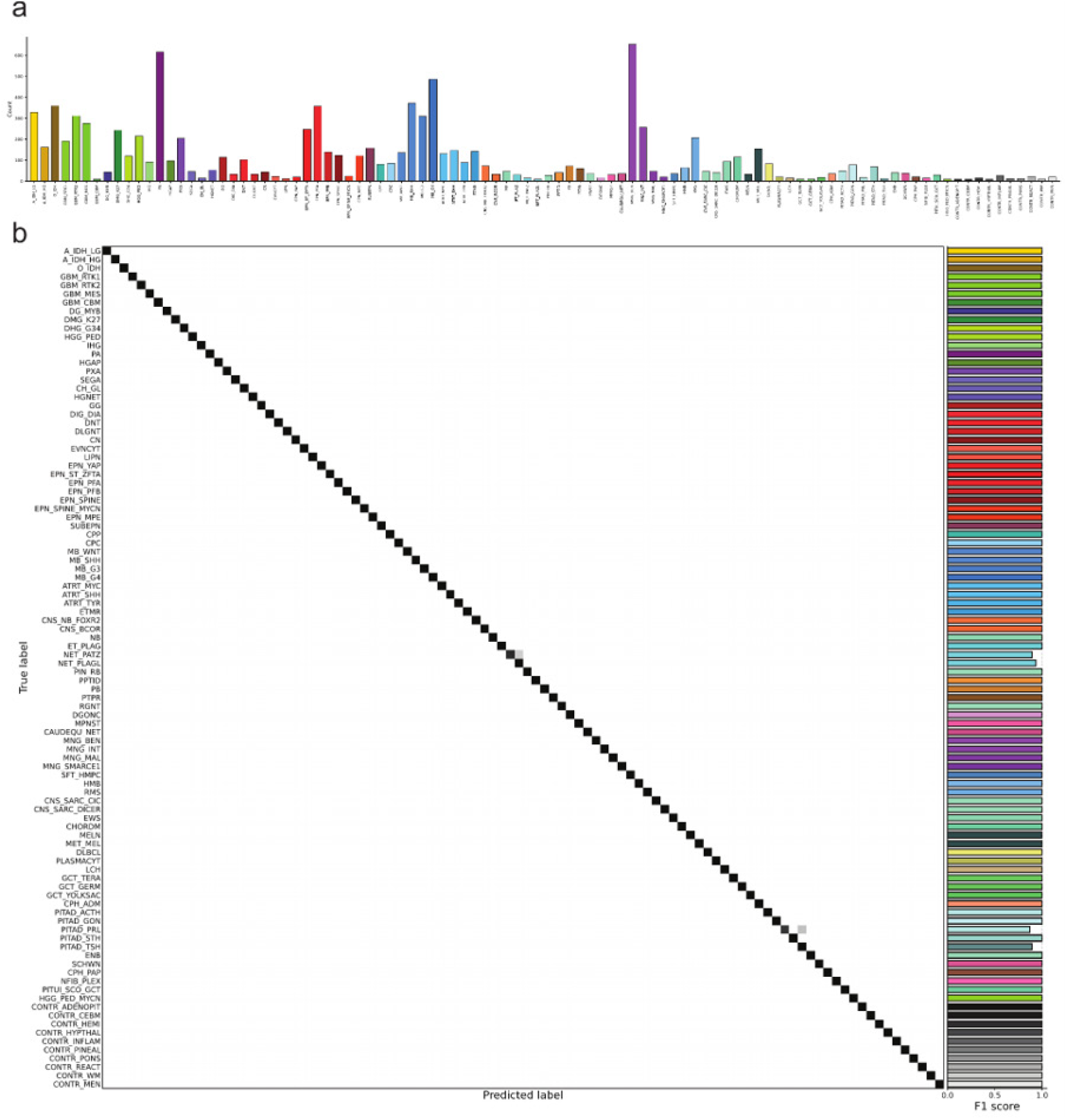
Sturgeon V2.4 class distribution and performance on hold-out test set. **a**. Methylation array subclass counts in the dataset used to train Sturgeon V2. **b**. Confusion matrix and F1 scores for subclasses in Sturgeon V2 based on hold-out test set.

**Supplementary Figure 5:**
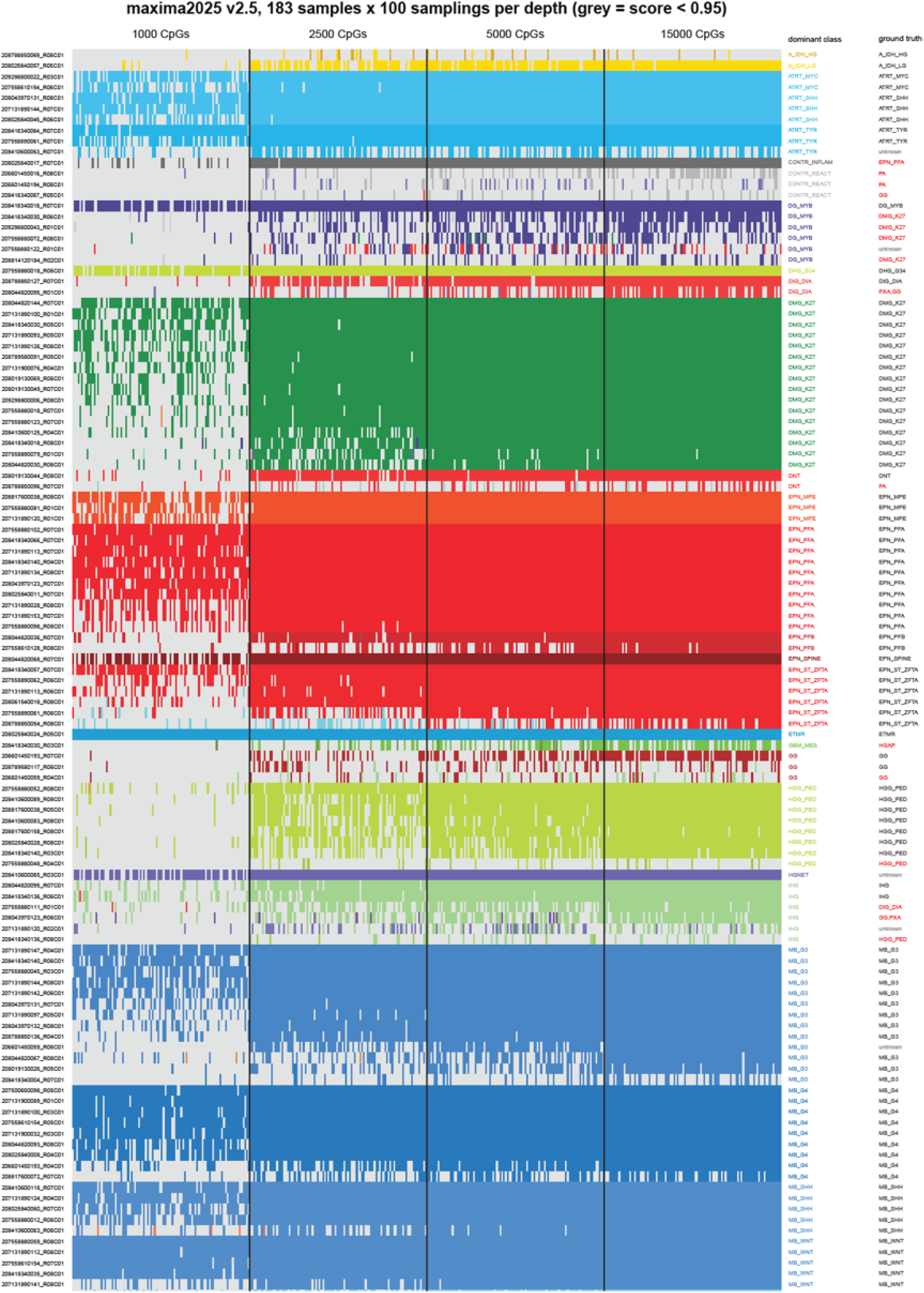
Retrospective performance on array data. Each row represents a sample, per sample 100 random samplings of 1,000, 2,500, 5,000 and 15,000 CpG sites (out of the 356K total CpG sites), shown per column. The dominant class is highlighted and the True label, if available, is shown on the right. Discrepancies between the True label and the dominant label are highlighted in red. Classifications are shown in their respective class label color if confident (>0.95 confidence) or grey when not confident.

**Supplementary figure 6:**
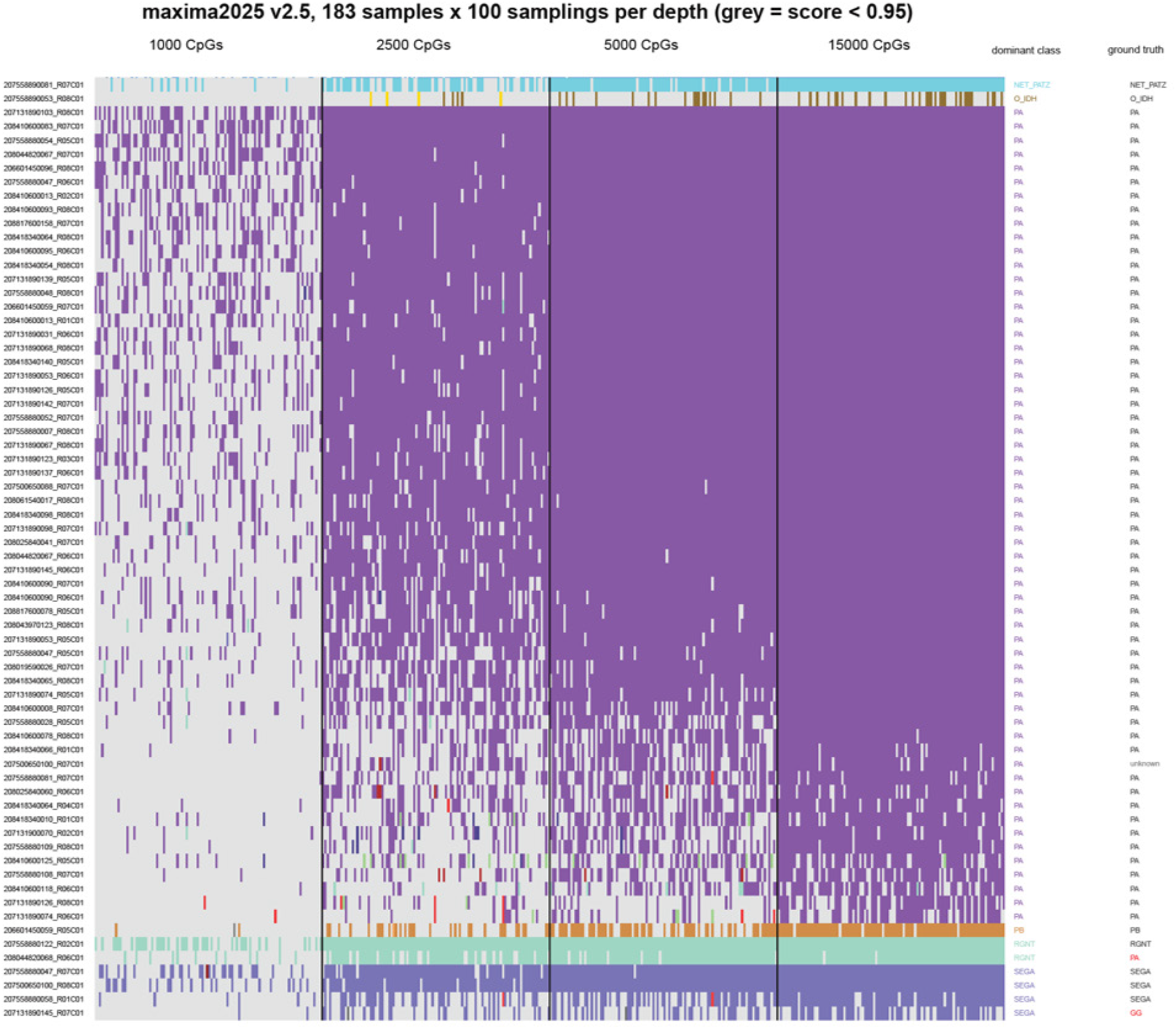
Retrospective performance on array data (continued). Each row represents a sample, per sample 100 random samplings of the data were taken of 1,000, 2,500, 5,000 and 15,000 CpG sites (out of the 356K total CpG sites), shown per column. The dominant class is highlighted and the True label, if available, is shown on the right. Classifications are shown in their respective class label color if confident (>0.95 confidence) or grey when not confident.

**Supplementary Figure 7:**
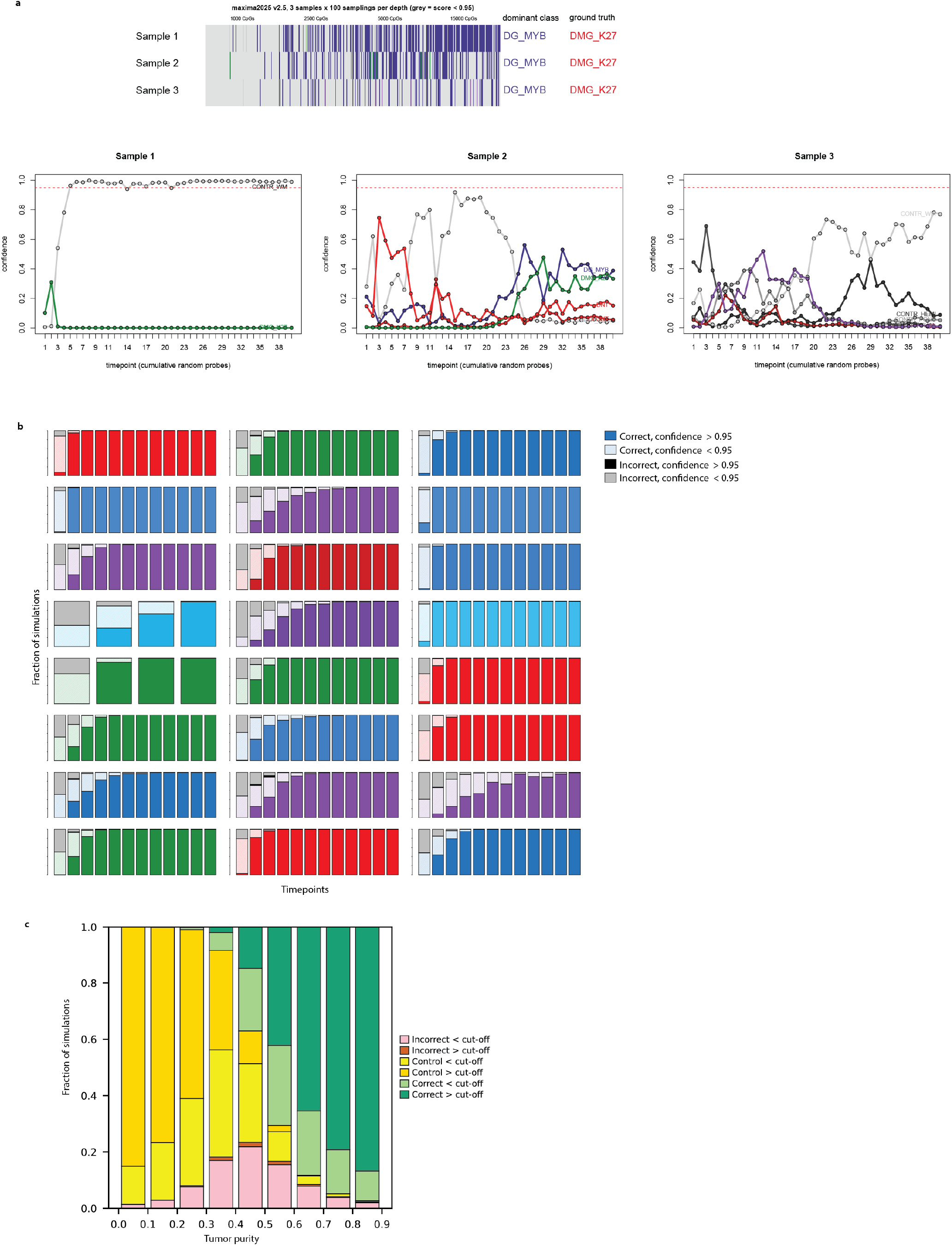
Validation tests for Sturgeon V2. **a**. Several IDH-wildtype high grade gliomas, K27 altered, were misclassified as MYB altered Diffuse glioma in the Array-downsampling experiments (Supplementary Figure 4). We obtained DNA from these samples and performed nanopore sequencing. **b**. Robustness testing on 24 biobanked samples, at different simulated timepoints 100 sequence runs are simulated and the performance of the classifier is evaluated. Bars are colored in their respective class colors. **c**. Stress-testing is performed by mixing samples with an increasing fraction of control signal, we then evaluate the occurrence of correct and incorrect classifications, as well as control-class classifications.

## Notes

### Author Declarations

The research was approved by the Biobank and Data access committee (BDAC, PMCLAB2023.0429) of the Princess Maxima Center for pediatric oncology. The biobank collection is registered as a clinical trial in the Netherlands and has been approved by the Dutch Medical Ethical Commission. All included patients provided written informed consent for participation in the biobank (International Clinical Trials Registry Platform: NL7744; https://onderzoekmetmensen.nl/en/trial/21619). I confirm that all necessary patient/participant consent has been obtained and the appropriate institutional forms have been archived, and that any patient/participant/sample identifiers included were not known to anyone (e.g., hospital staff, patients or participants themselves) outside the research group so cannot be used to identify individuals.

### Funding

- Hanarth fonds
- Foundation KiKa
- Health Holland

