## Supplementary data 1 for "Sturgeon V2: a clinically applicable CNS tumor classifier trained on relabeled data"

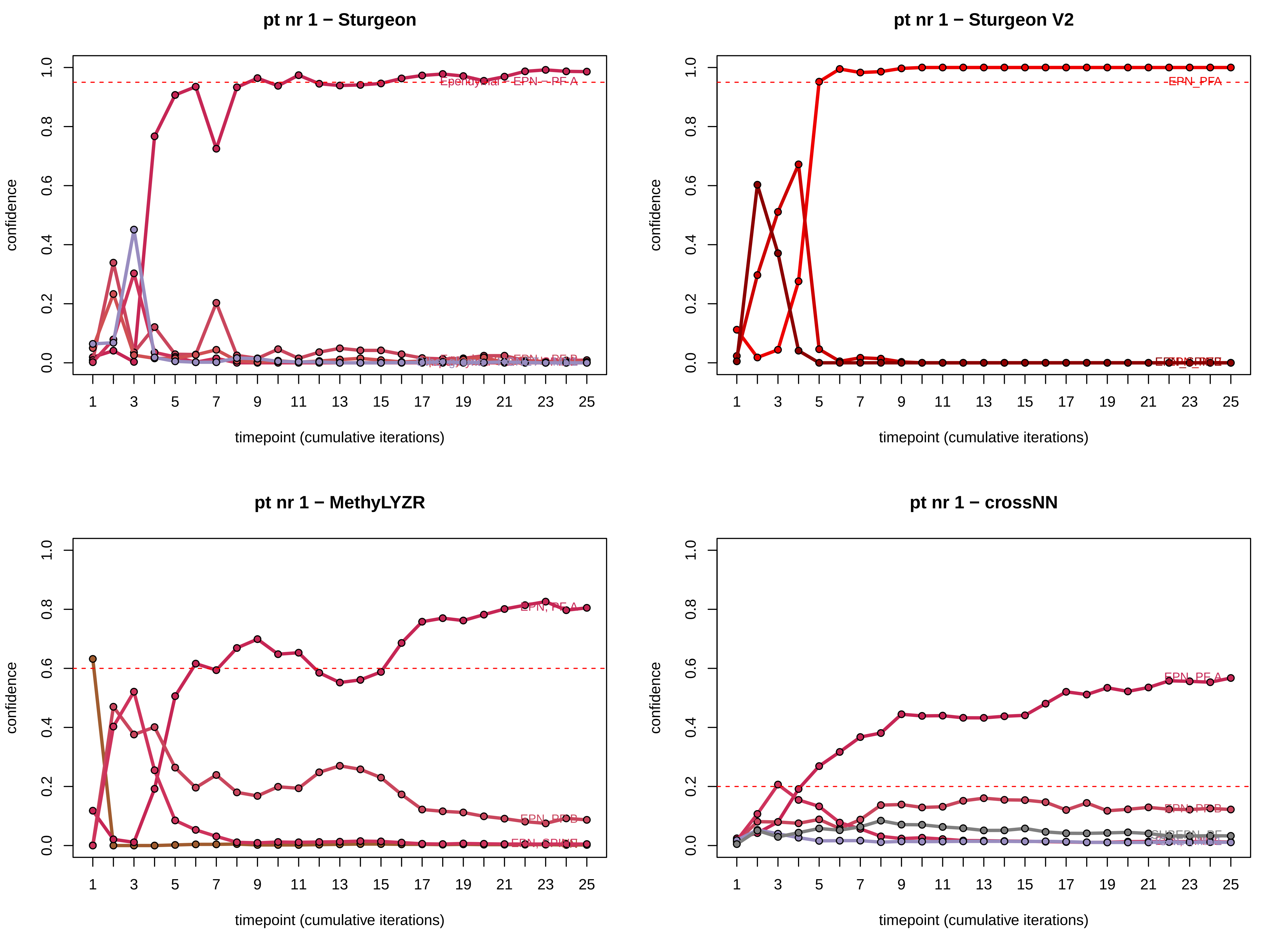
pt nr 1 − Sturgeon
pt nr 1 − Sturgeon V2
pt nr 1 − MethyLYZR
pt nr 1 − crossNN
confidence
timepoint (cumulative iterations)
Ependymal - EPN - PF A
EPN_PFA
EPN, PF A
EPN, PFB
EPN_PF A
EPN_PFB

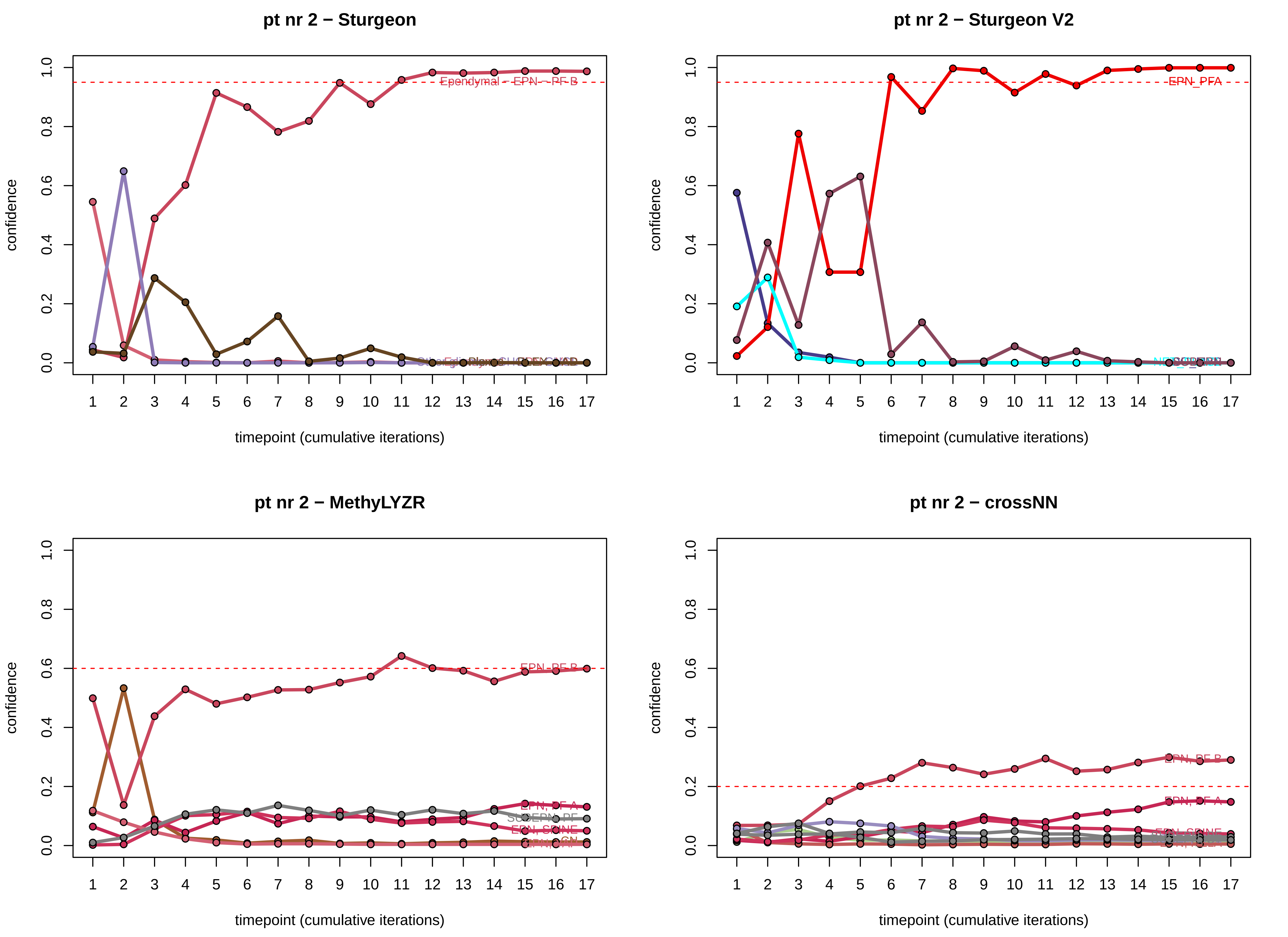

pt nr 2 − Sturgeon
pt nr 2 − Sturgeon V2
pt nr 2 − MethyLYZR
pt nr 2 − crossNN
confidence
timepoint (cumulative iterations)
0.0
0.2
0.4
0.6
0.8
1.0
Ependymal - EPN - PF B
EPN_PFA
EPN_PF B
EPN, PF A
EPN, SPINE
EPN, PF B

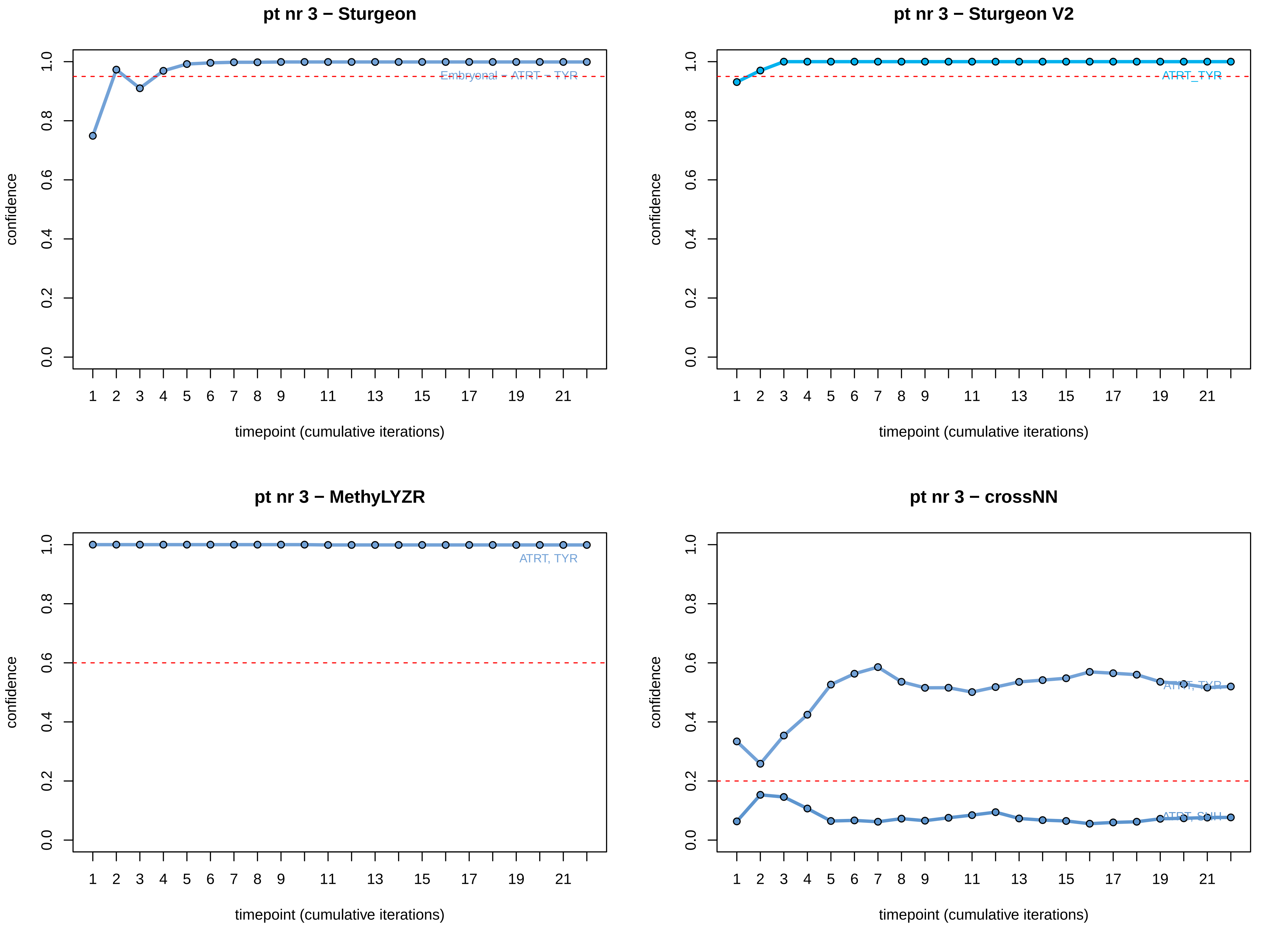
pt nr 3 − Sturgeon
Embryonal − ATRT − TYR
pt nr 3 − Sturgeon V2
ATRT_TYR
pt nr 3 − MethyLYZR
ATRT, TYR
pt nr 3 − crossNN
ATRT, TYR
ATRT, SHH
confidence
0.0
0.2
0.4
0.6
0.8
1.0
timepoint (cumulative iterations)
1
2
3
4
5
6
7
8
9
11
13
15
17
19
21

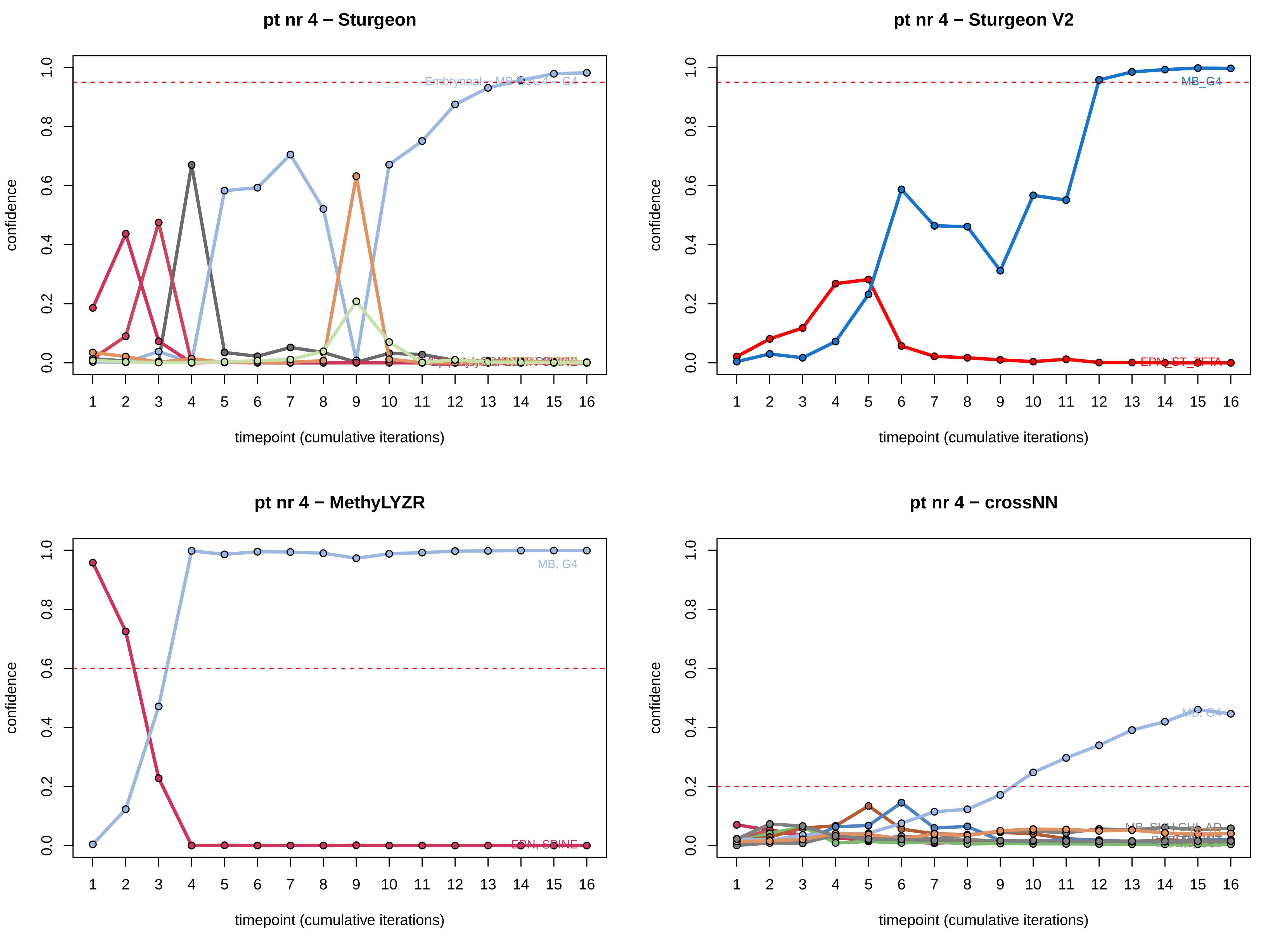

pt nr 4 − Sturgeon
confidence
timepoint (cumulative iterations)
0.0
0.2
0.4
0.6
0.8
1.0
1 2 3 4 5 6 7 8 9 10 11 12 13 14 15 16
pt nr 4 − Sturgeon V2
MB_G4
EPN_ST_ZFTA
pt nr 4 − MethyLYZR
MB, G4
EPN, SPINE
pt nr 4 − crossNN
MB, G4

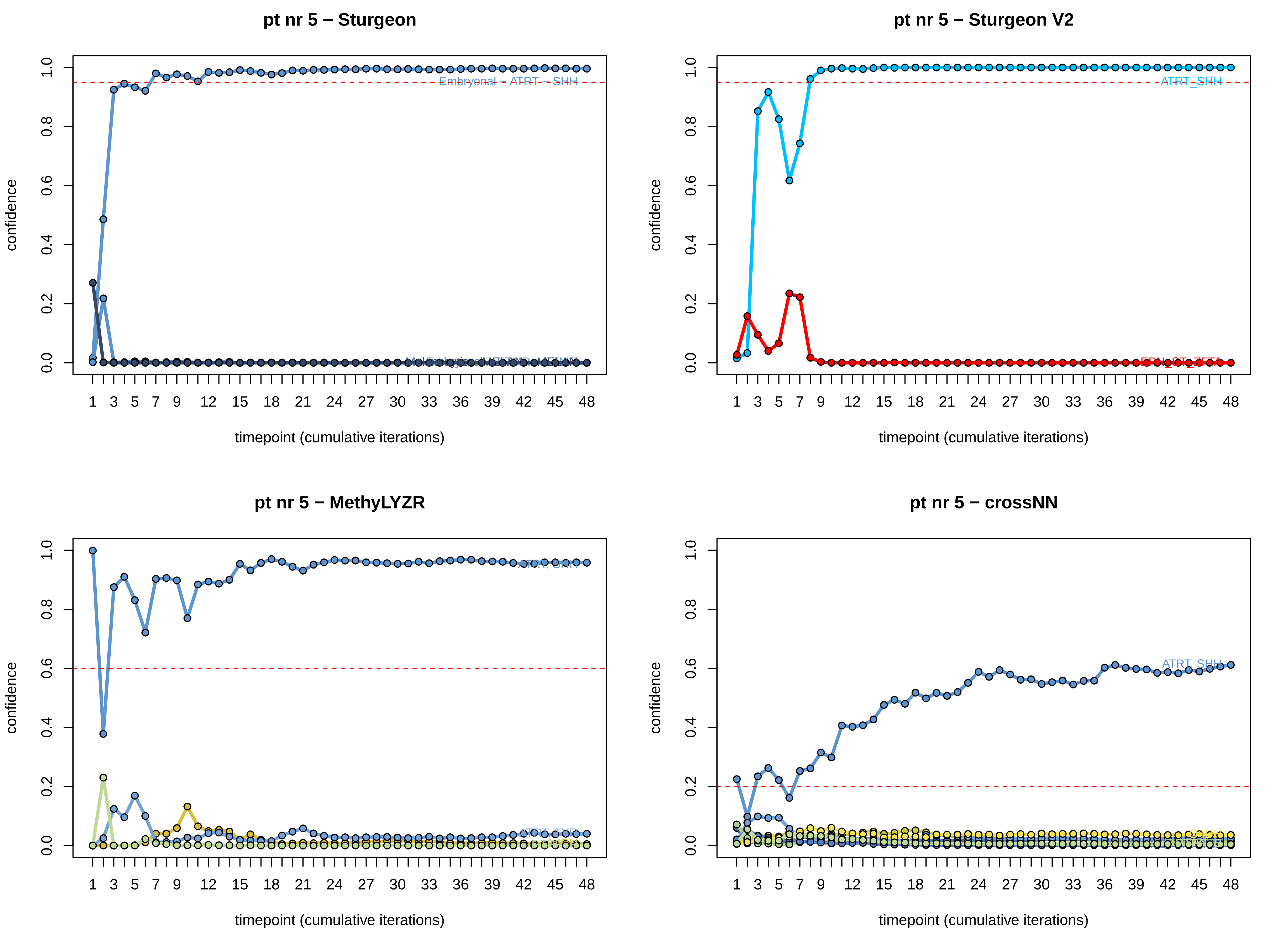
pt nr 5 − Sturgeon
Embryonal − ATRT − SHH
confidence
timepoint (cumulative iterations)
pt nr 5 − Sturgeon V2
ATRT_SHH
confidence
timepoint (cumulative iterations)
pt nr 5 − MethyLYZR
ATRT, SHH
ATRT, TYR
confidence
timepoint (cumulative iterations)
pt nr 5 − crossNN
ATRT_SHH
confidence
timepoint (cumulative iterations)

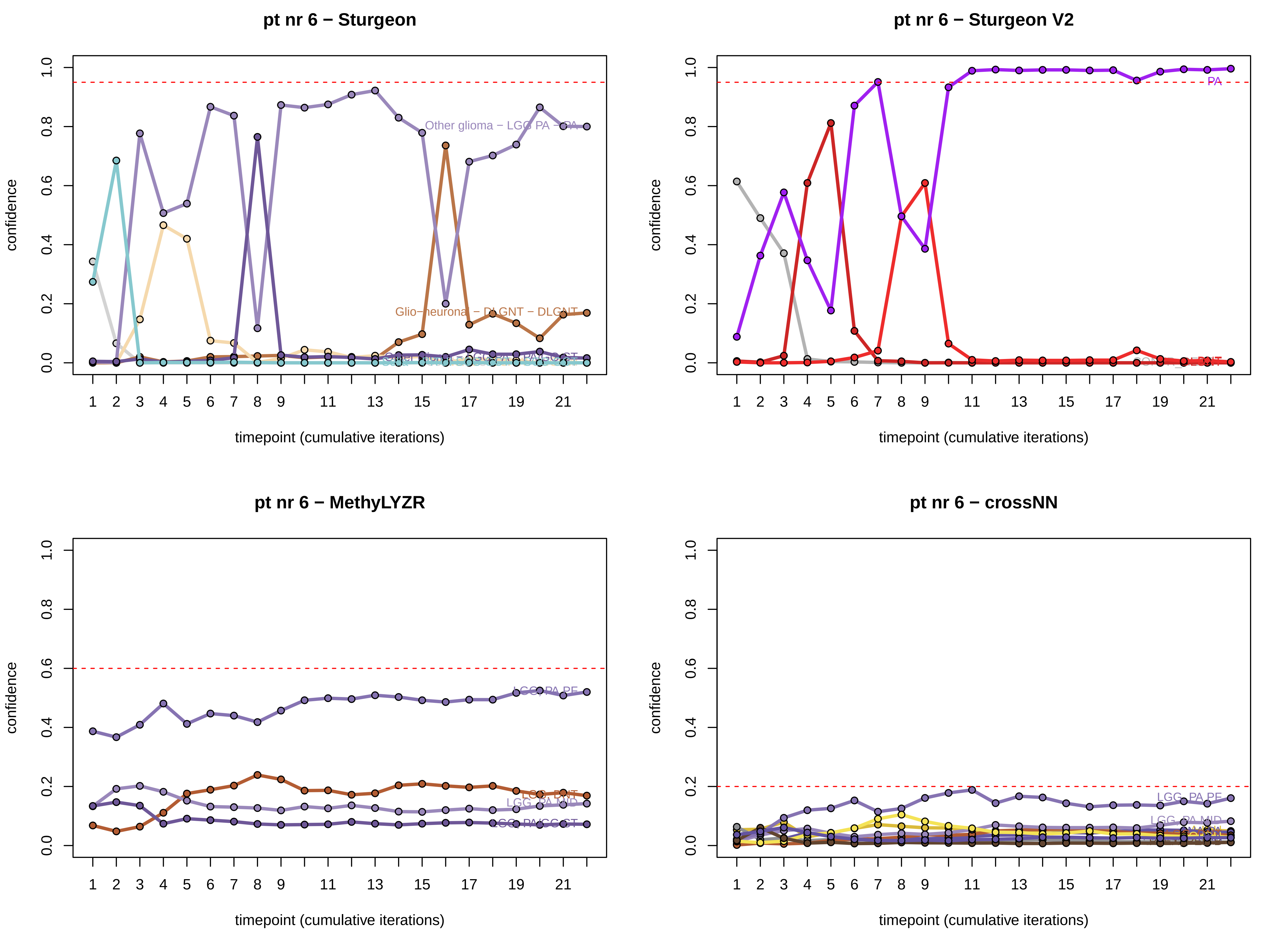
pt nr 6 − Sturgeon
confidence
timepoint (cumulative iterations)
Other glioma − LGG PA − PA
Glio−neuronal − DLGNT − DLGNT
pt nr 6 − Sturgeon V2
PA
pt nr 6 − MethyLYZR
LGG_PA_PF
LGG_DNT
LGG_PA_MID
LGG_PA/GG_ST
pt nr 6 − crossNN
LGG_PA_PF
LGG_PA_MID
0.0
0.2
0.4
0.6
0.8
1.0
1
2
3
4
5
6
7
8
9
11
13
15
17
19
21

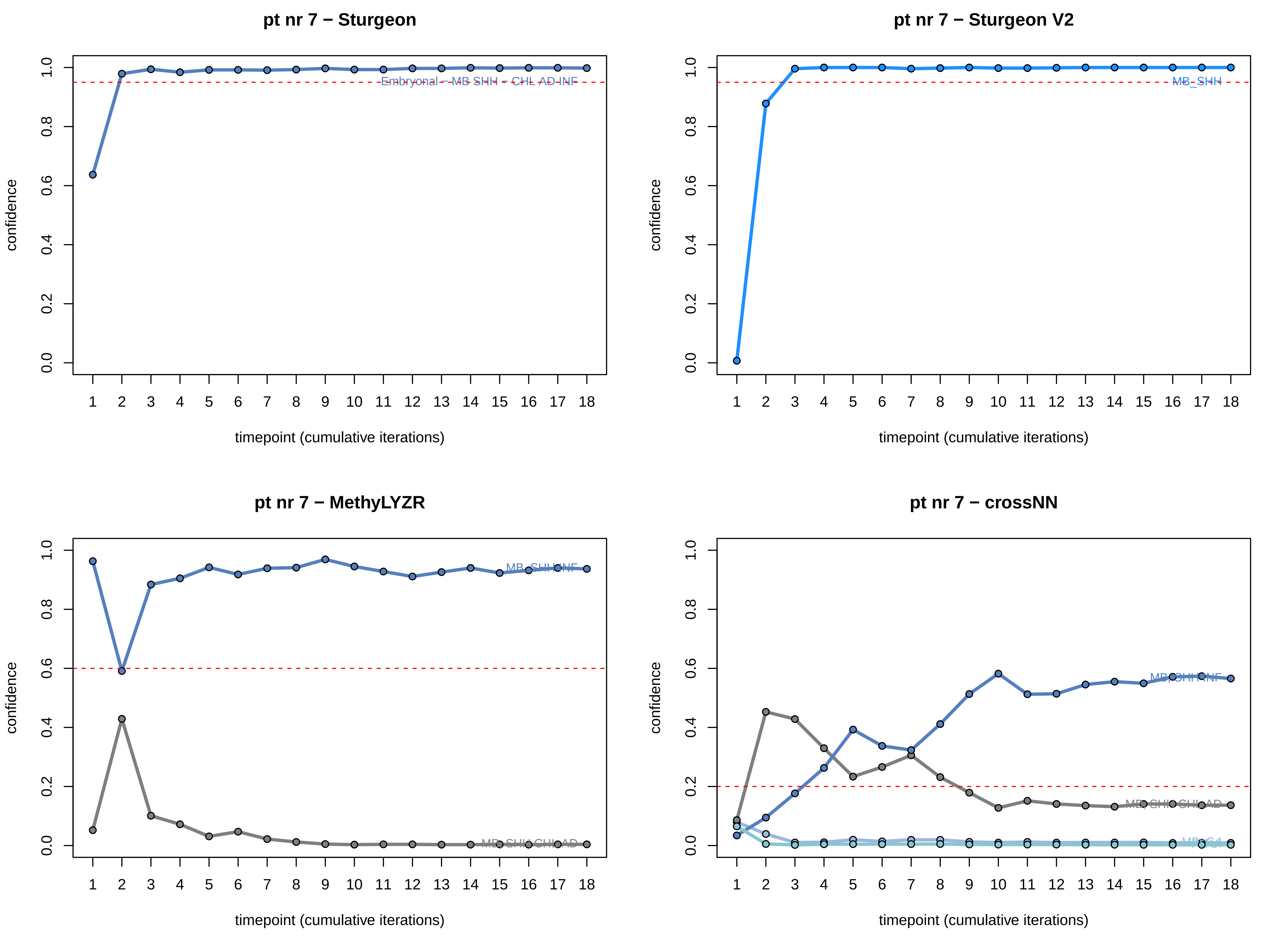
pt nr 7 − Sturgeon
Embryonal – MB SHH – CHL AD INF
pt nr 7 − Sturgeon V2
MB_SHH
pt nr 7 − MethyLYZR
pt nr 7 − crossNN
confidence
0.0
0.2
0.4
0.6
0.8
1.0
1 2 3 4 5 6 7 8 9 10 11 12 13 14 15 16 17 18
timepoint (cumulative iterations)

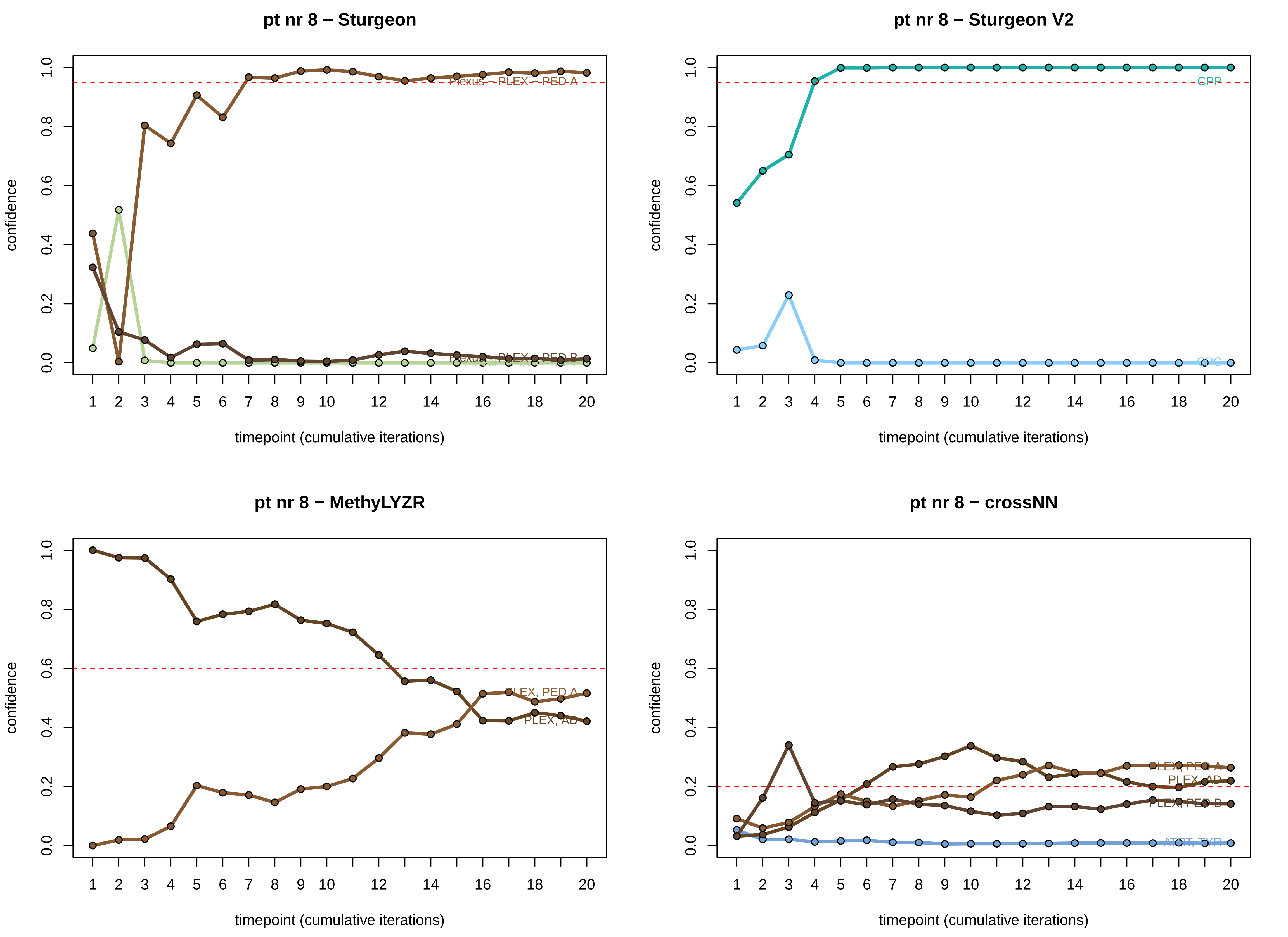
pt nr 8 − Sturgeon
confidence
timepoint (cumulative iterations)
Plexus − PLEX − PED A
pt nr 8 − Sturgeon V2
confidence
timepoint (cumulative iterations)
CPP
pt nr 8 − MethyLYZR
confidence
timepoint (cumulative iterations)
PLEX, PED A
PLEX, AD
pt nr 8 − crossNN
confidence
timepoint (cumulative iterations)
PLEX, PED A
PLEX, AD
PLEX, PED B
ATRT, TYR

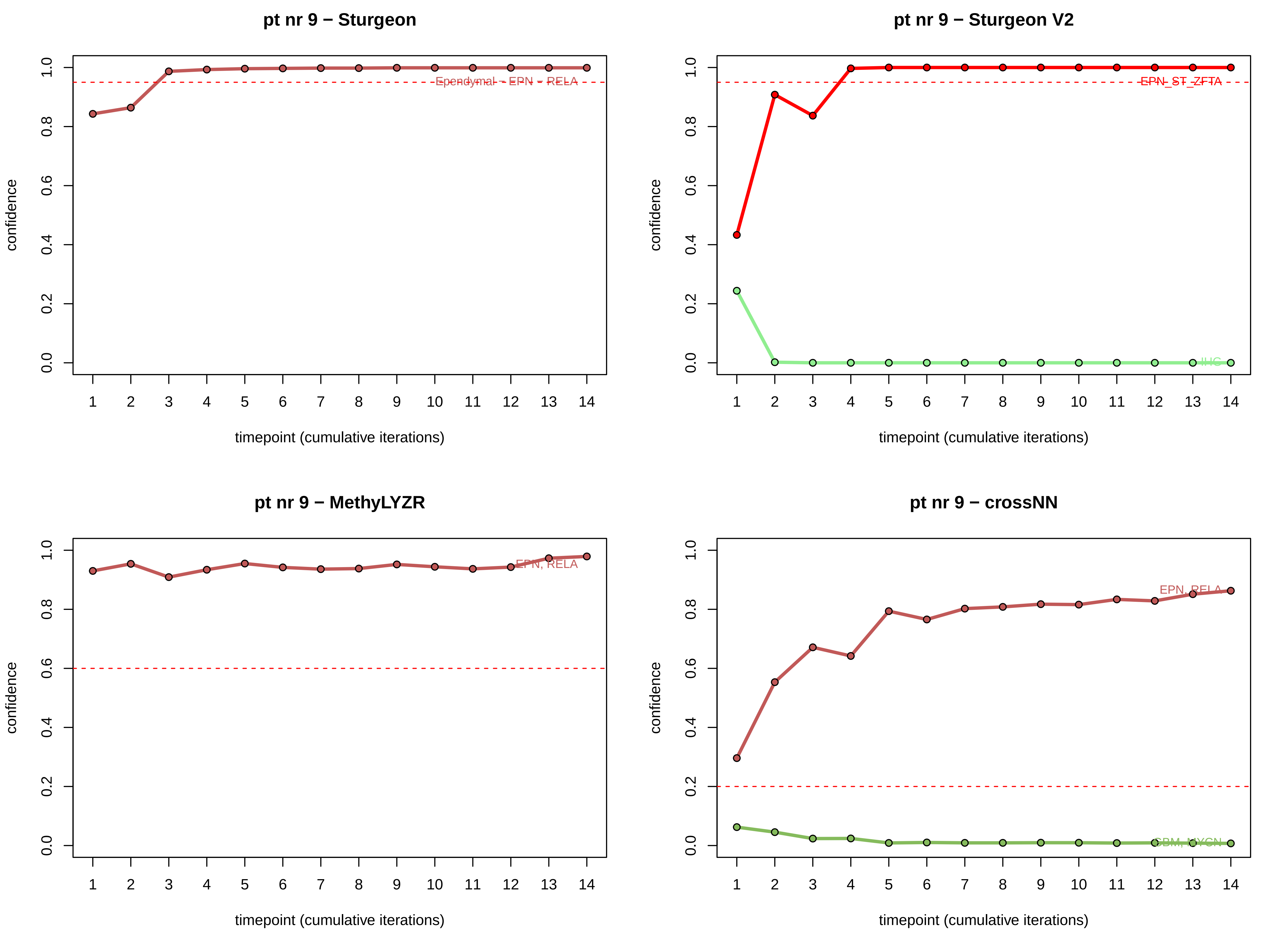
pt nr 9 − Sturgeon
confidence
timepoint (cumulative iterations)
Ependymal - EPN - RELA
pt nr 9 − Sturgeon V2
confidence
timepoint (cumulative iterations)
EPN_ST_ZFTA
IHG
pt nr 9 − MethyLYZR
confidence
timepoint (cumulative iterations)
EPN, RELA
pt nr 9 − crossNN
confidence
timepoint (cumulative iterations)
EPN, RELA
GBM, MYCN

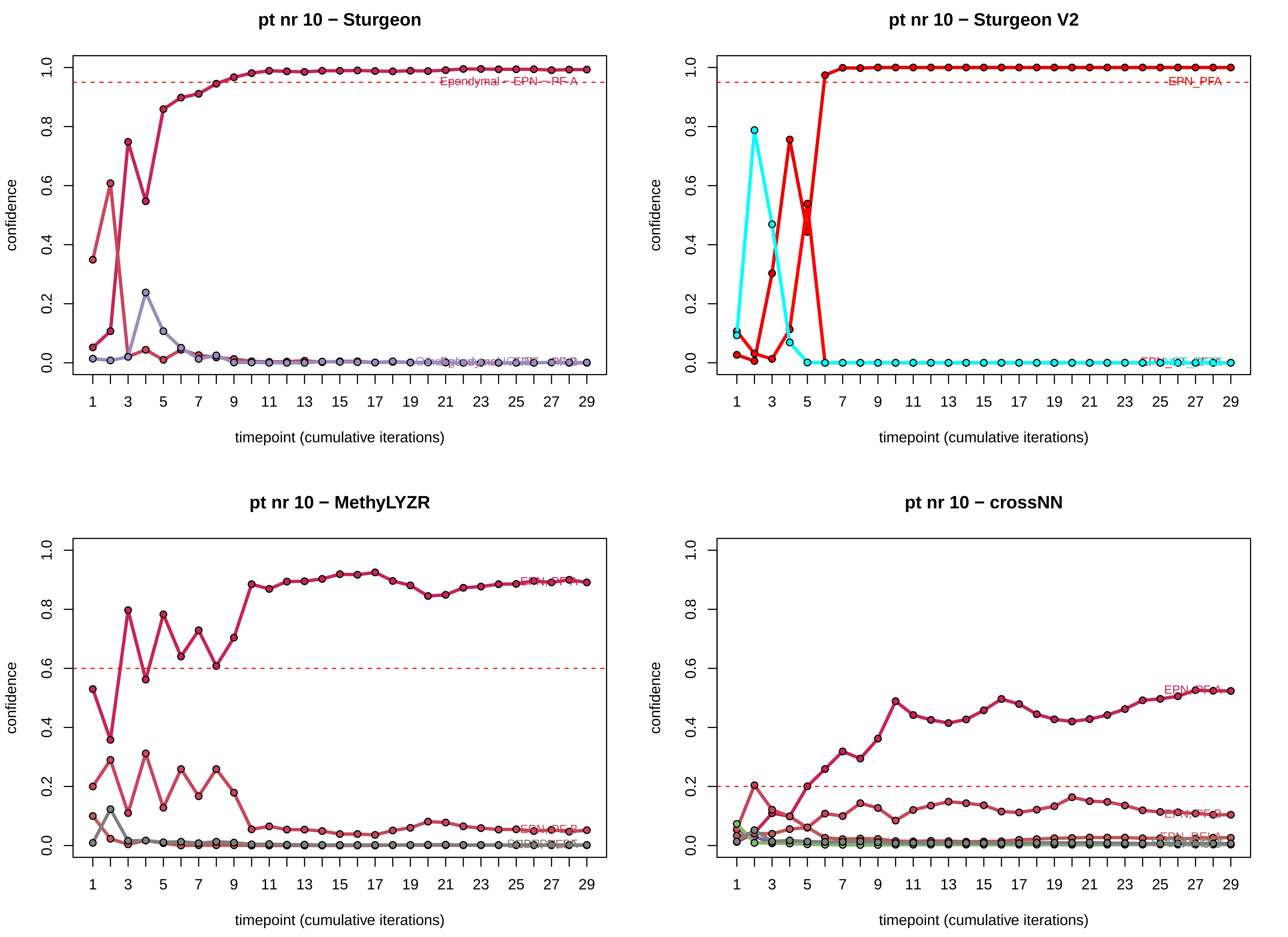
pt nr 10 − Sturgeon
Ependymal - EPN - PF A
confidence
timepoint (cumulative iterations)
pt nr 10 − Sturgeon V2
EPN_PFA
confidence
timepoint (cumulative iterations)
pt nr 10 − MethyLYZR
EPN_PFA
EPN_PFB
confidence
timepoint (cumulative iterations)
pt nr 10 − crossNN
EPN_PFA
EPN_PFB
confidence
timepoint (cumulative iterations)

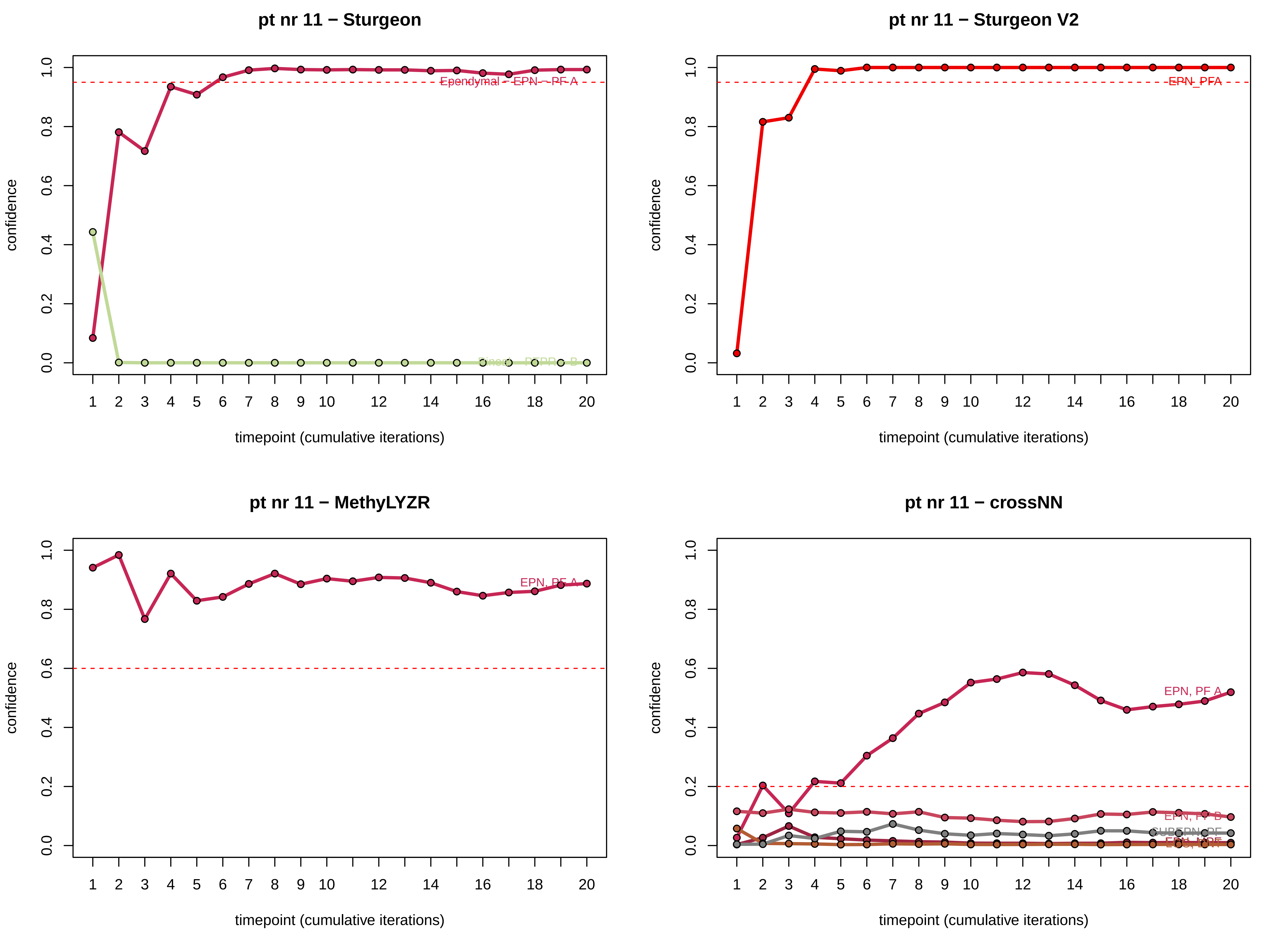
pt nr 11 − Sturgeon
pt nr 11 − Sturgeon V2
pt nr 11 − MethyLYZR
pt nr 11 − crossNN
confidence
timepoint (cumulative iterations)
EPN_PFA
EPN, PF A
EPN, PF B

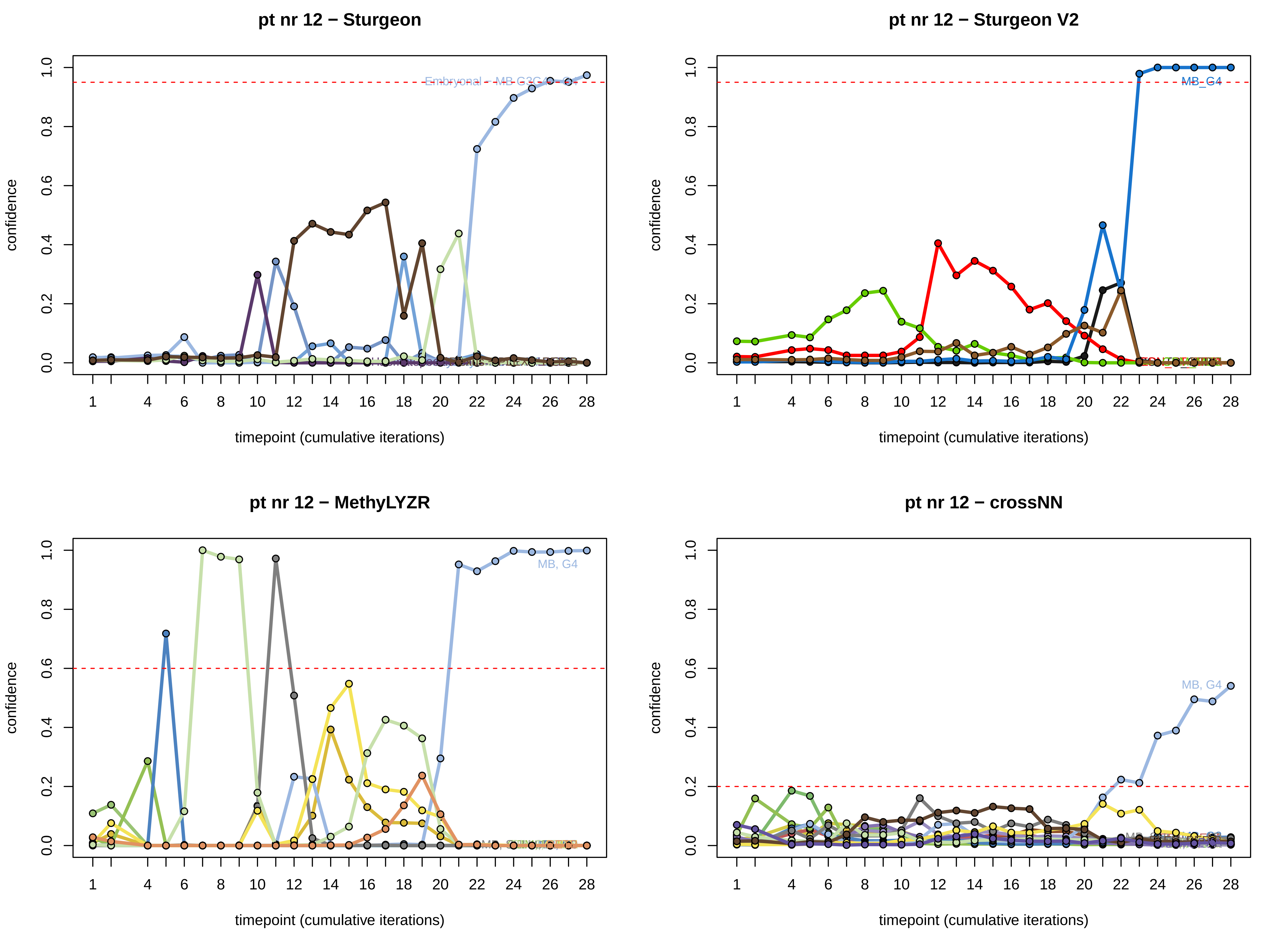
pt nr 12 − Sturgeon
confidence
timepoint (cumulative iterations)
0.0
0.2
0.4
0.6
0.8
1.0
1
4
6
8
10
12
14
16
18
20
22
24
26
28
pt nr 12 − Sturgeon V2
confidence
timepoint (cumulative iterations)
MB_G4
pt nr 12 − MethyLYZR
confidence
timepoint (cumulative iterations)
MB, G4
pt nr 12 − crossNN
confidence
timepoint (cumulative iterations)
MB, G4

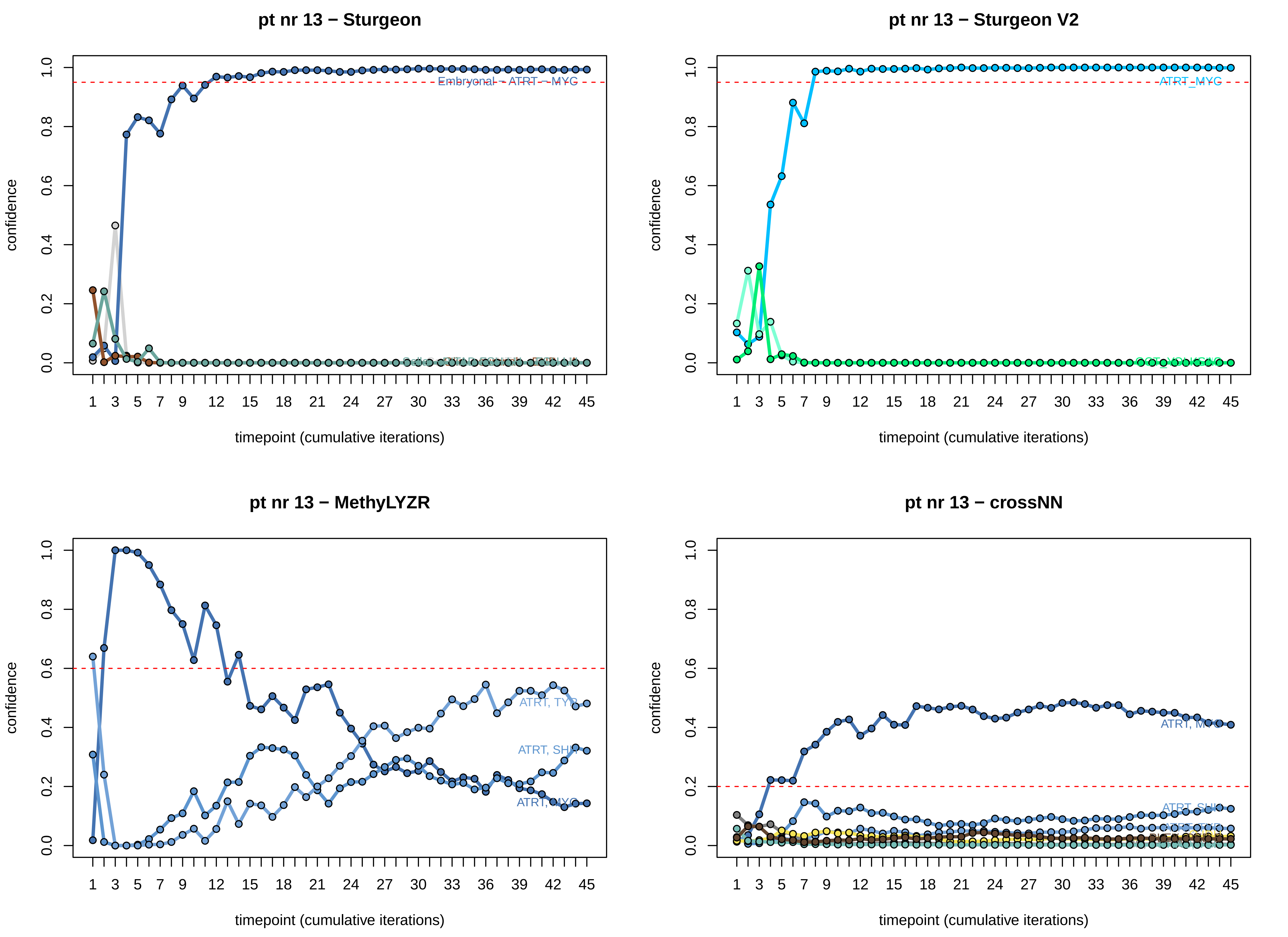

pt nr 13 − Sturgeon
Embryonal − ATRT − MYC
pt nr 13 − Sturgeon V2
ATRT_MYC
pt nr 13 − MethyLYZR
ATRT, TYR
ATRT, SHH
ATRT, MYC
pt nr 13 − crossNN
ATRT, MYC
ATRT, SHH
confidence
timepoint (cumulative iterations)

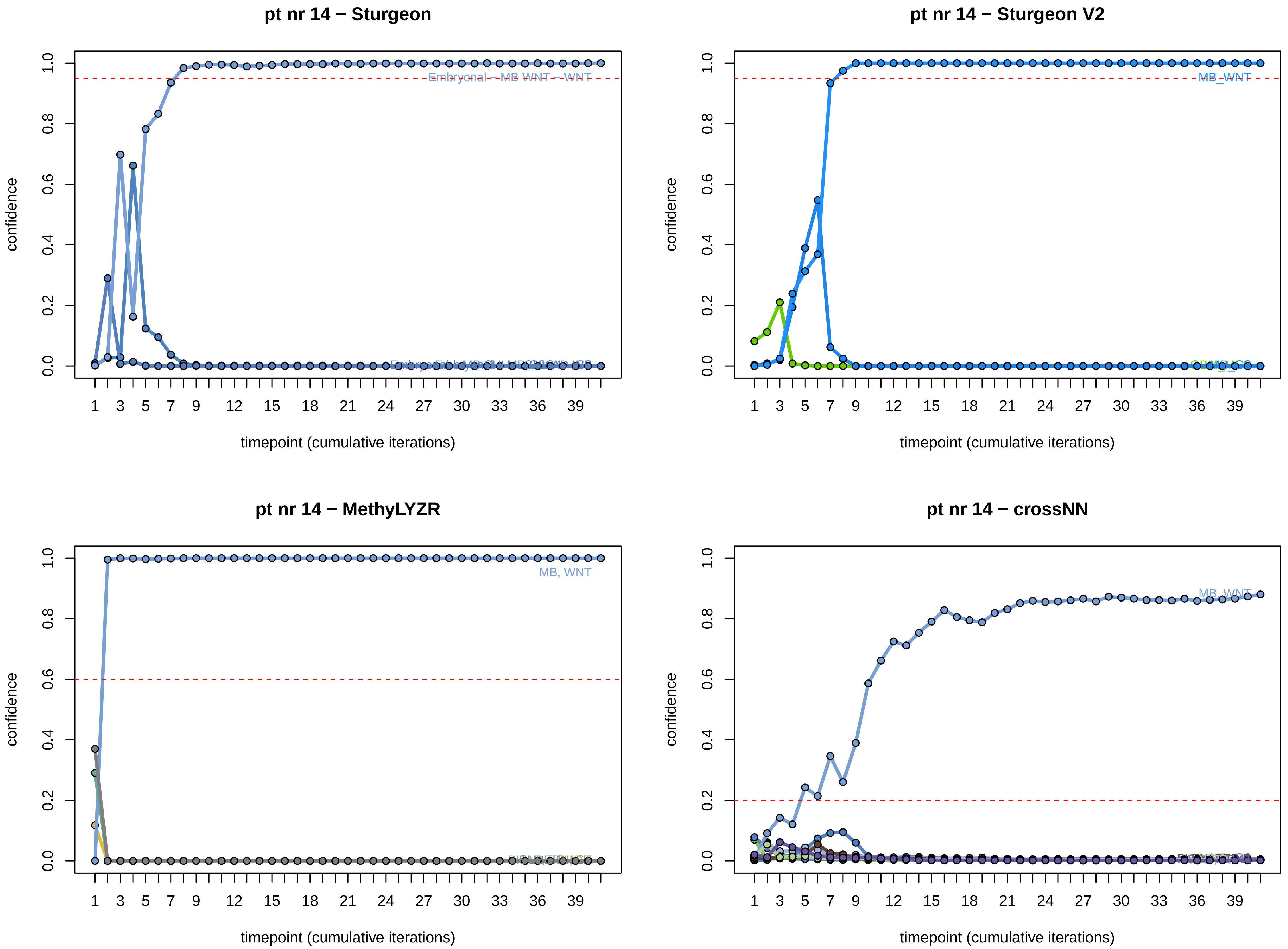
pt nr 14 − Sturgeon
confidence
timepoint (cumulative iterations)
Embryonal – MB WNT – WNT
pt nr 14 − Sturgeon V2
confidence
timepoint (cumulative iterations)
MB_WNT
pt nr 14 − MethyLYZR
confidence
timepoint (cumulative iterations)
MB, WNT
pt nr 14 − crossNN
confidence
timepoint (cumulative iterations)
MB_WNT

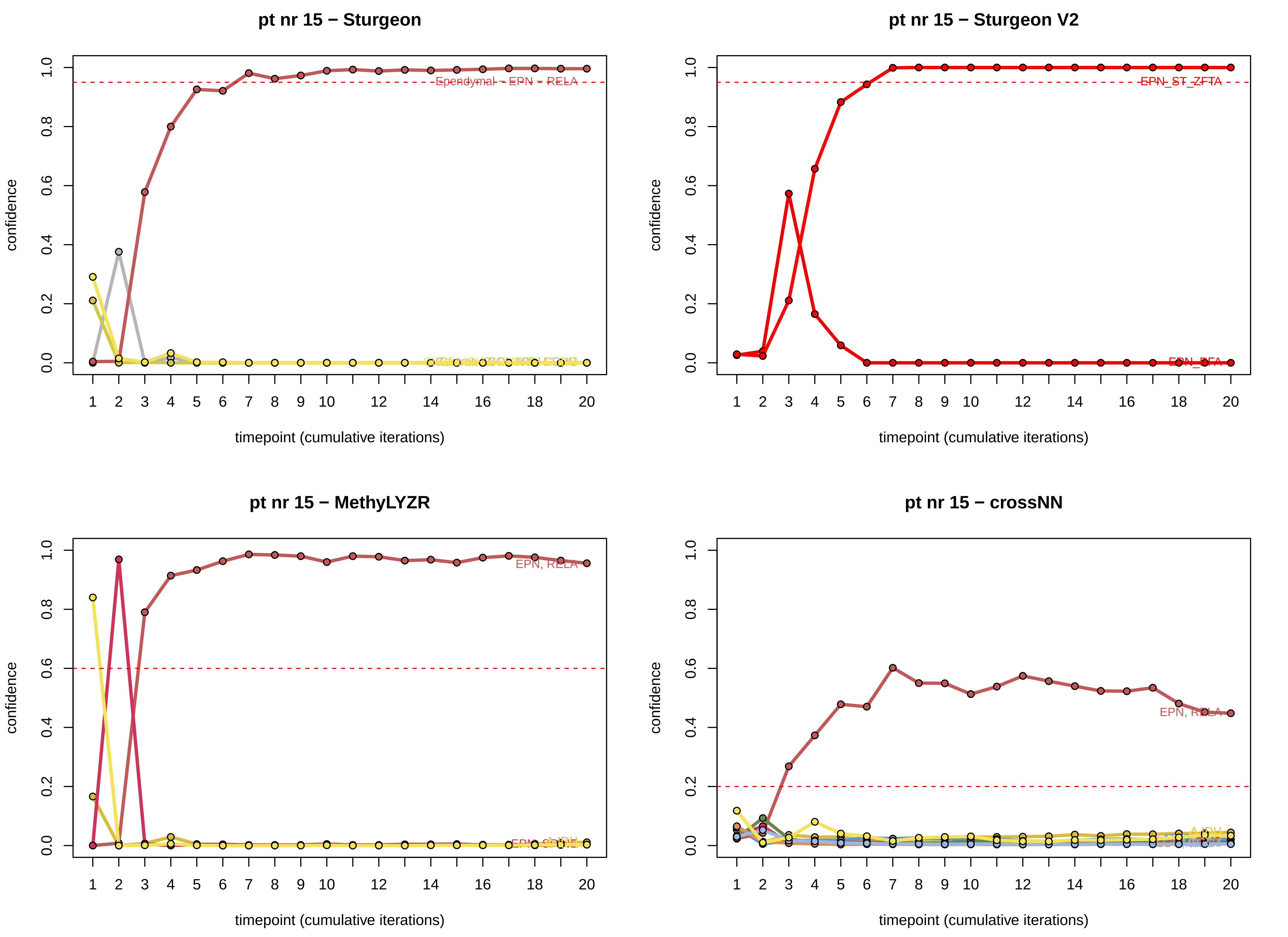
pt nr 15 − Sturgeon
Ependymal - EPN - RELA
confidence
timepoint (cumulative iterations)
pt nr 15 − Sturgeon V2
EPN_ST_ZFTA
EPN_PFA
confidence
timepoint (cumulative iterations)
pt nr 15 − MethyLYZR
EPN, RELA
A IDH
confidence
timepoint (cumulative iterations)
pt nr 15 − crossNN
EPN, RELA
A IDH
confidence
timepoint (cumulative iterations)

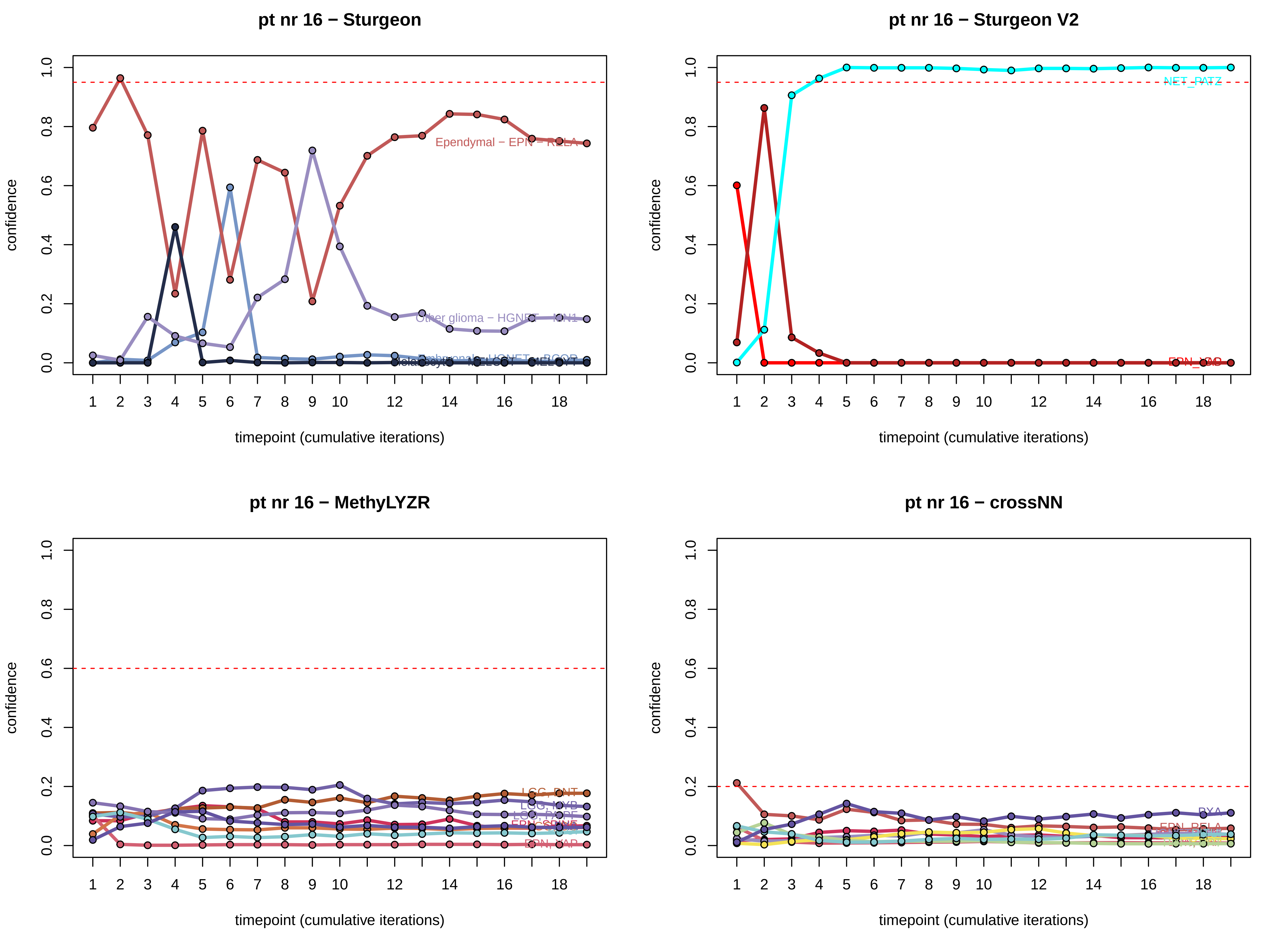
pt nr 16 − Sturgeon
Ependymal − EPN − RELA
Other glioma − HGNET − MN1
confidence
timepoint (cumulative iterations)
pt nr 16 − Sturgeon V2
NET_PATZ
confidence
timepoint (cumulative iterations)
pt nr 16 − MethyLYZR
LGG, DNT
LGG, MYB
EPN, YAP
confidence
timepoint (cumulative iterations)
pt nr 16 − crossNN
PXA
EPN_RELA
confidence
timepoint (cumulative iterations)

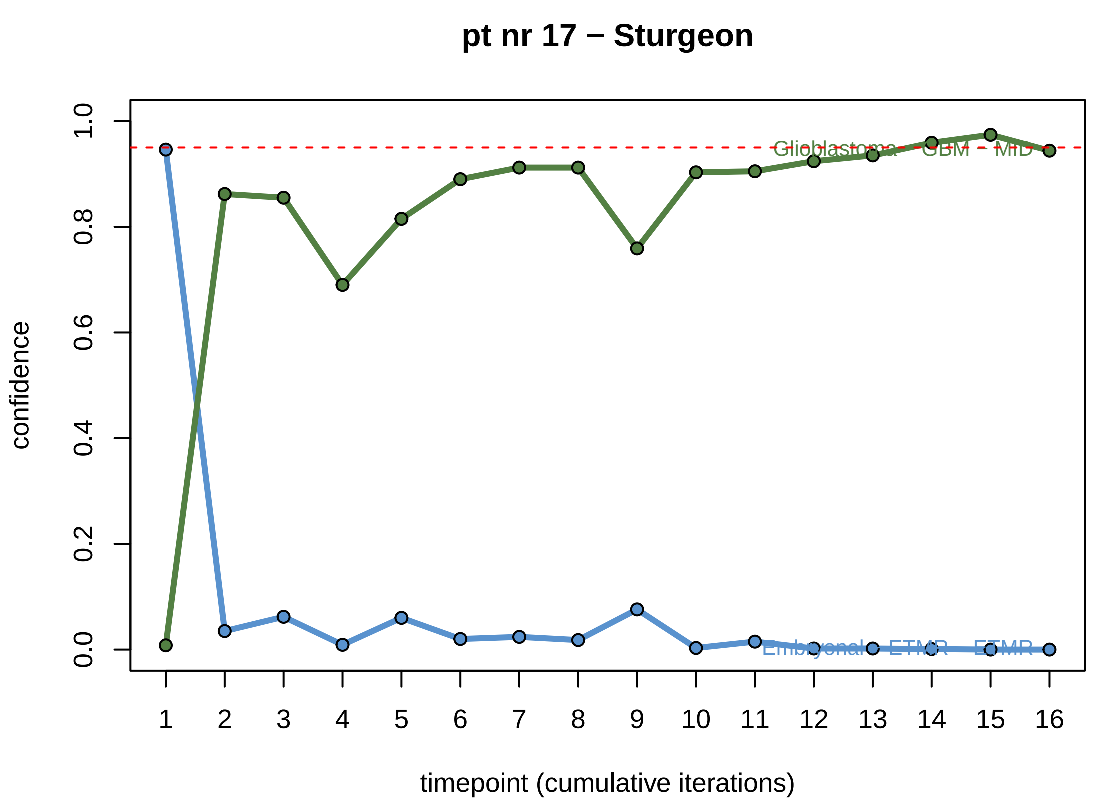
pt nr 17 − Sturgeon
confidence
timepoint (cumulative iterations)
Glioblastoma - GBM - MID
Embryonal - ETMR - ETMR

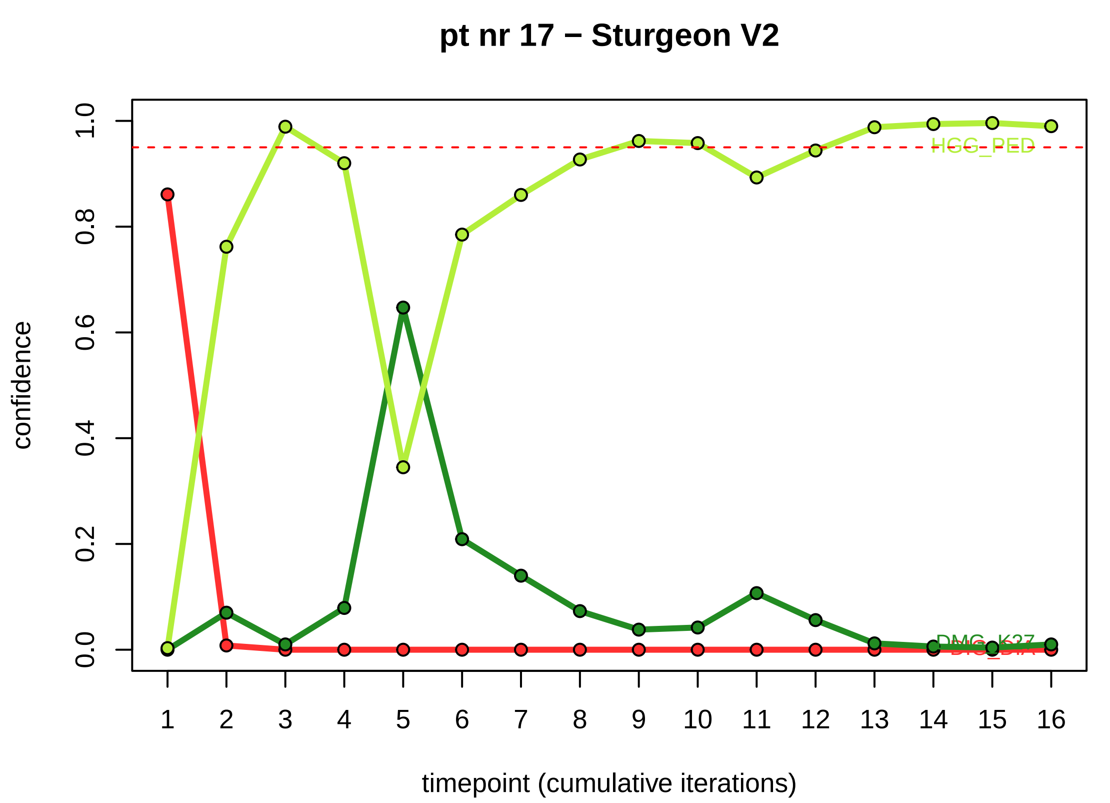
pt nr 17 − Sturgeon V2
confidence
timepoint (cumulative iterations)
HGG_PED
DMG_K27

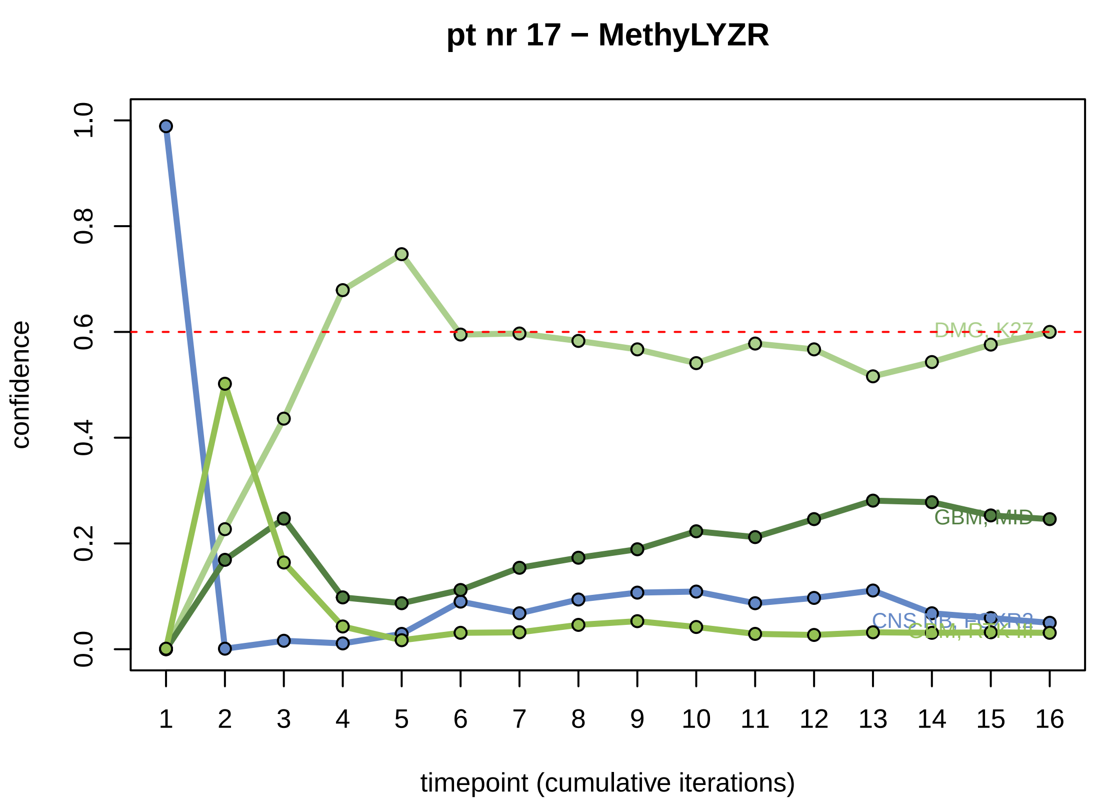
pt nr 17 − MethyLYZR
confidence
timepoint (cumulative iterations)
DMG_K27
GBM, MID

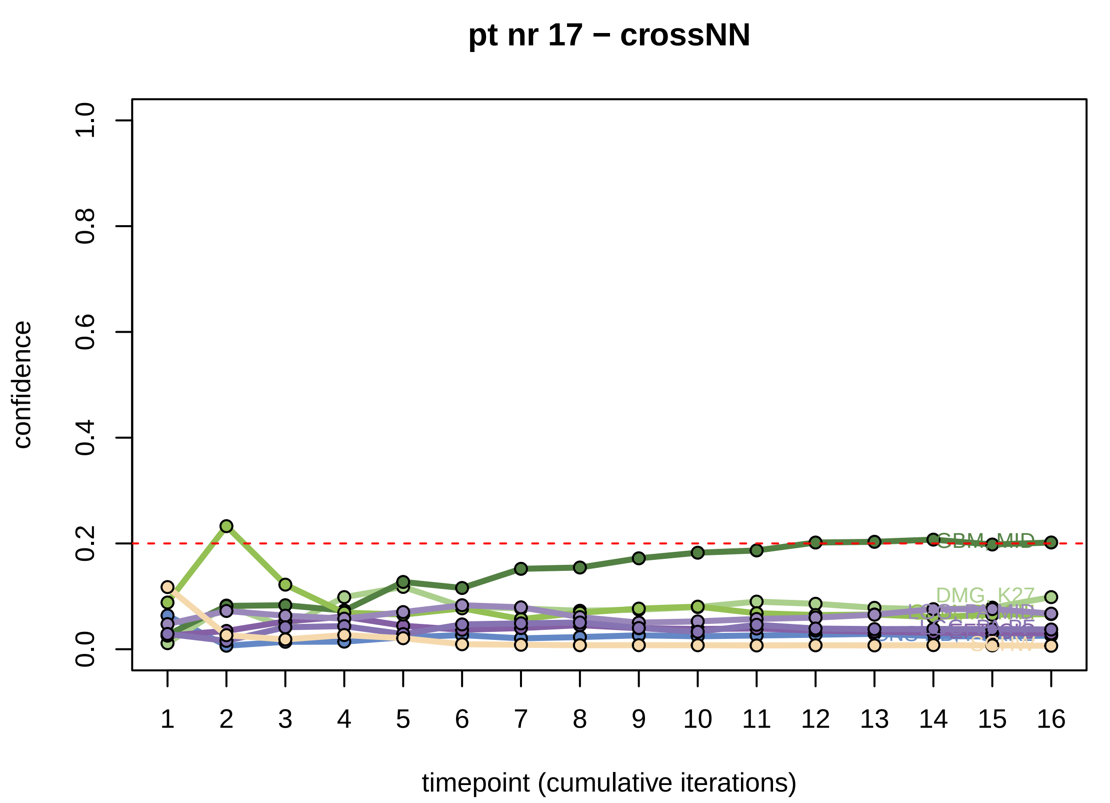
pt nr 17 − crossNN
confidence
timepoint (cumulative iterations)
GBM_MID
DMG_K27

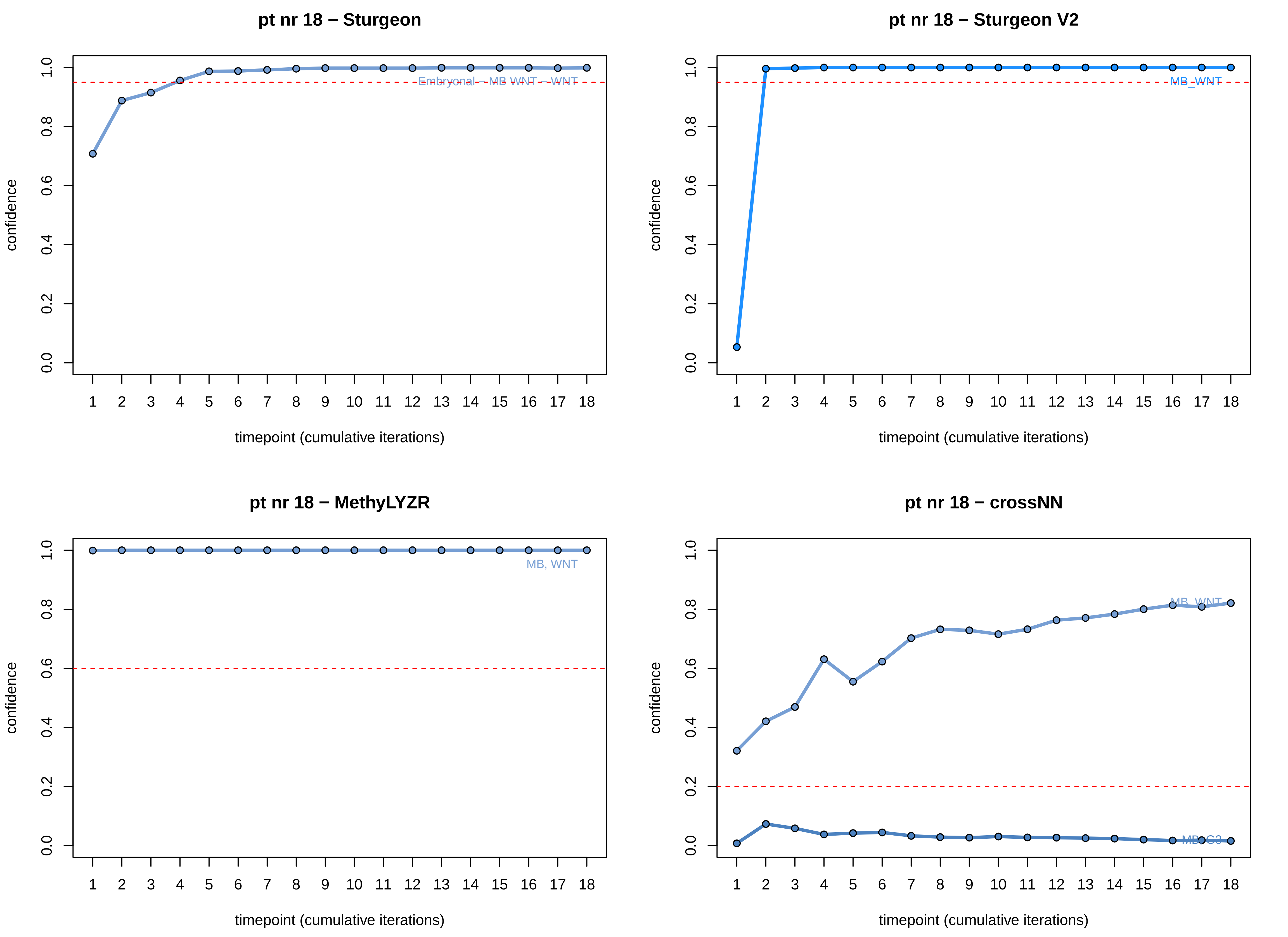

pt nr 18 − Sturgeon
Embryonal − MB WNT − WNT
pt nr 18 − Sturgeon V2
MB_WNT
pt nr 18 − MethyLYZR
MB, WNT
pt nr 18 − crossNN
MB_WNT
MB_G3
confidence
timepoint (cumulative iterations)

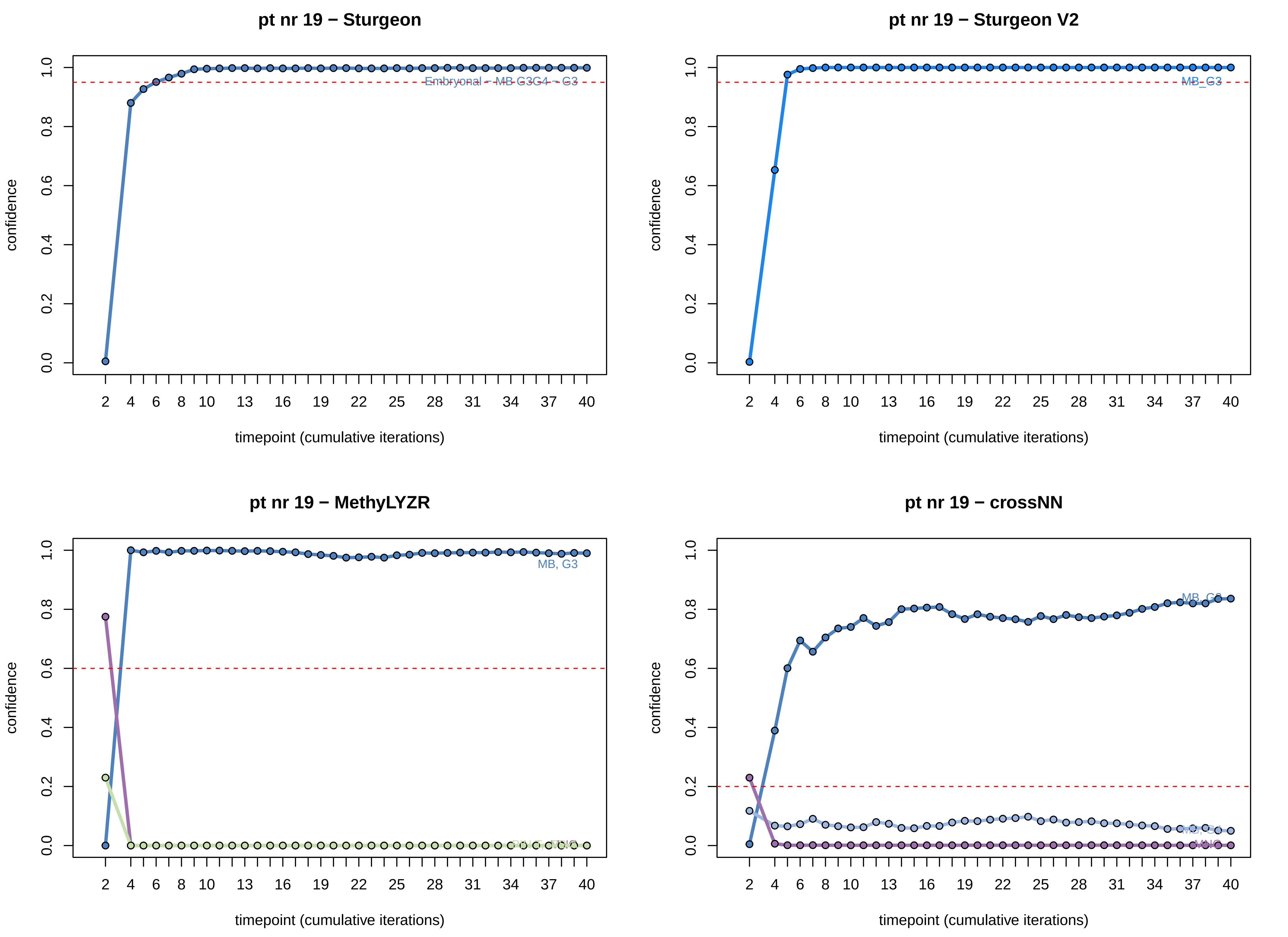

pt nr 19 − Sturgeon
confidence
0.0
0.2
0.4
0.6
0.8
1.0
Embryonal – MB G3G4 – G3
2
4
6
8
10
13
16
19
22
25
28
31
34
37
40
timepoint (cumulative iterations)
pt nr 19 − Sturgeon V2
confidence
MB_G3
timepoint (cumulative iterations)
pt nr 19 − MethyLYZR
confidence
MB, G3
timepoint (cumulative iterations)
pt nr 19 − crossNN
confidence
MB, G3
MB, G4
MNG
timepoint (cumulative iterations)

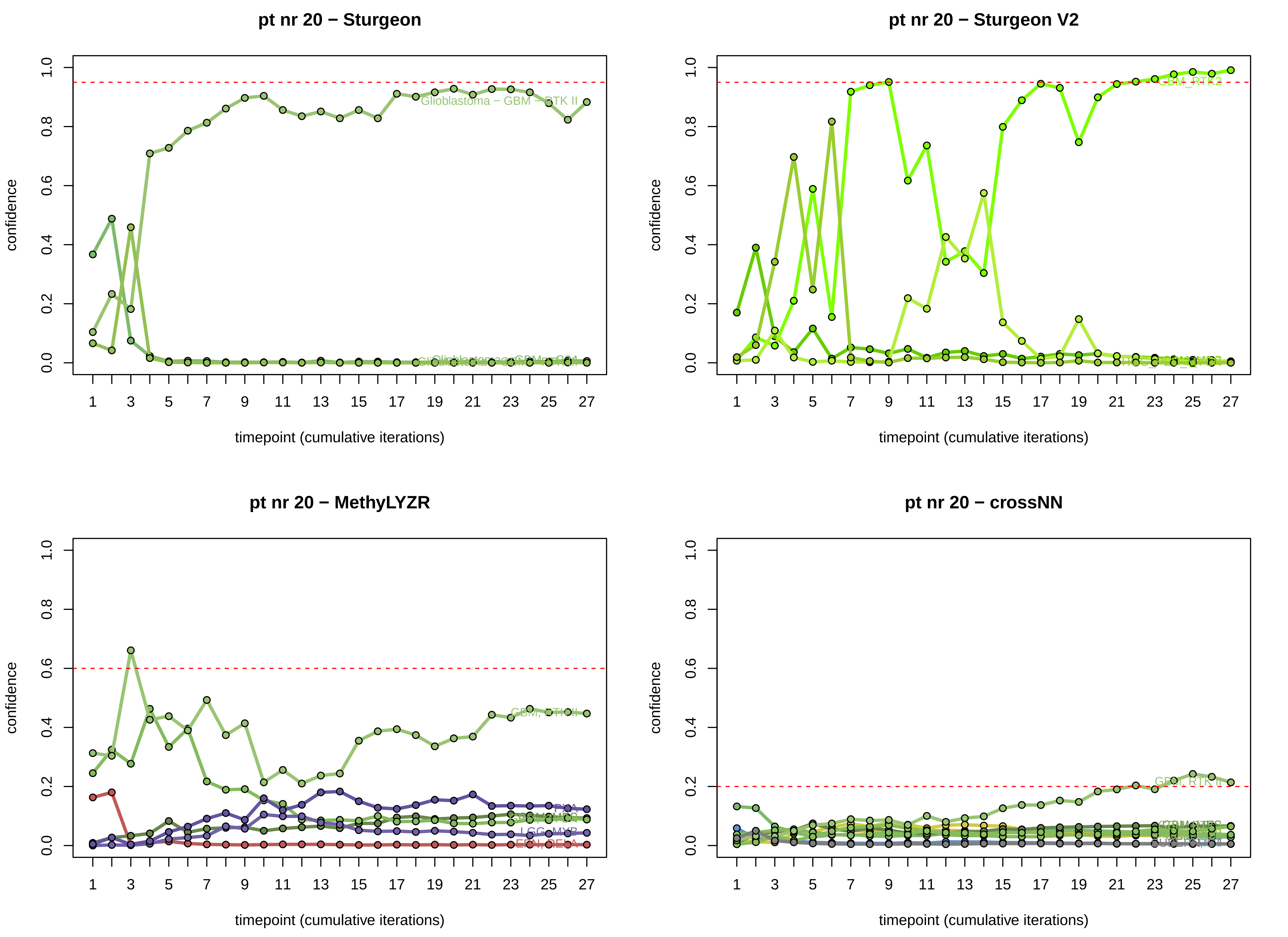
pt nr 20 − Sturgeon
confidence
timepoint (cumulative iterations)
Glioblastoma – GBM – RTK II
pt nr 20 − Sturgeon V2
confidence
timepoint (cumulative iterations)
GBM_RTK2
pt nr 20 − MethyLYZR
confidence
timepoint (cumulative iterations)
GBM, RTK II
PXA
LGG, MYB
EPN, RELA
pt nr 20 − crossNN
confidence
timepoint (cumulative iterations)
GBM, RTK II

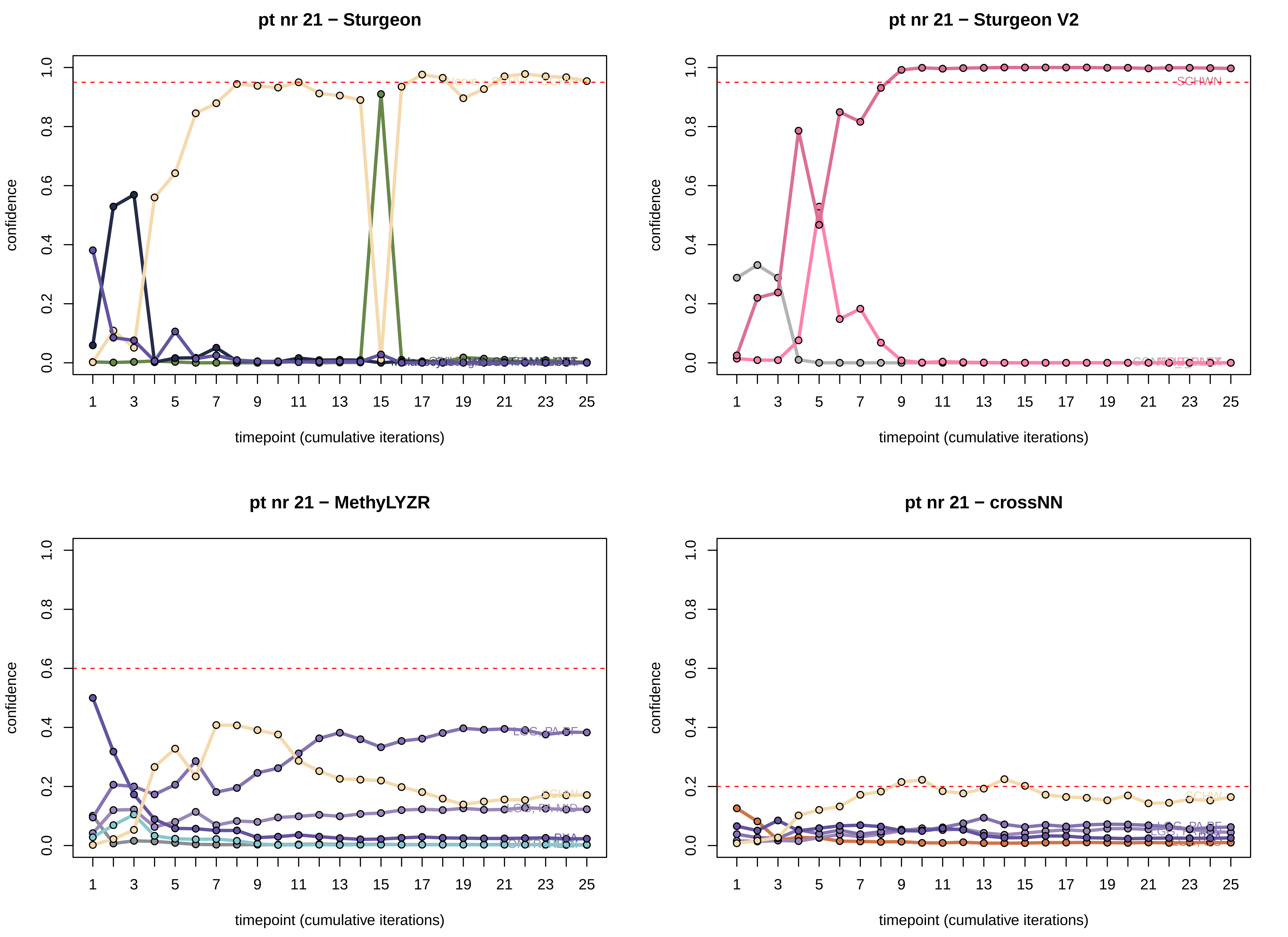
pt nr 21 − Sturgeon
pt nr 21 − Sturgeon V2
pt nr 21 − MethyLYZR
pt nr 21 − crossNN
confidence
timepoint (cumulative iterations)
0.0
0.2
0.4
0.6
0.8
1.0
1 3 5 7 9 11 13 15 17 19 21 23 25
SCHWN
LGG, PA PF
SCHW
LGG, PA MID

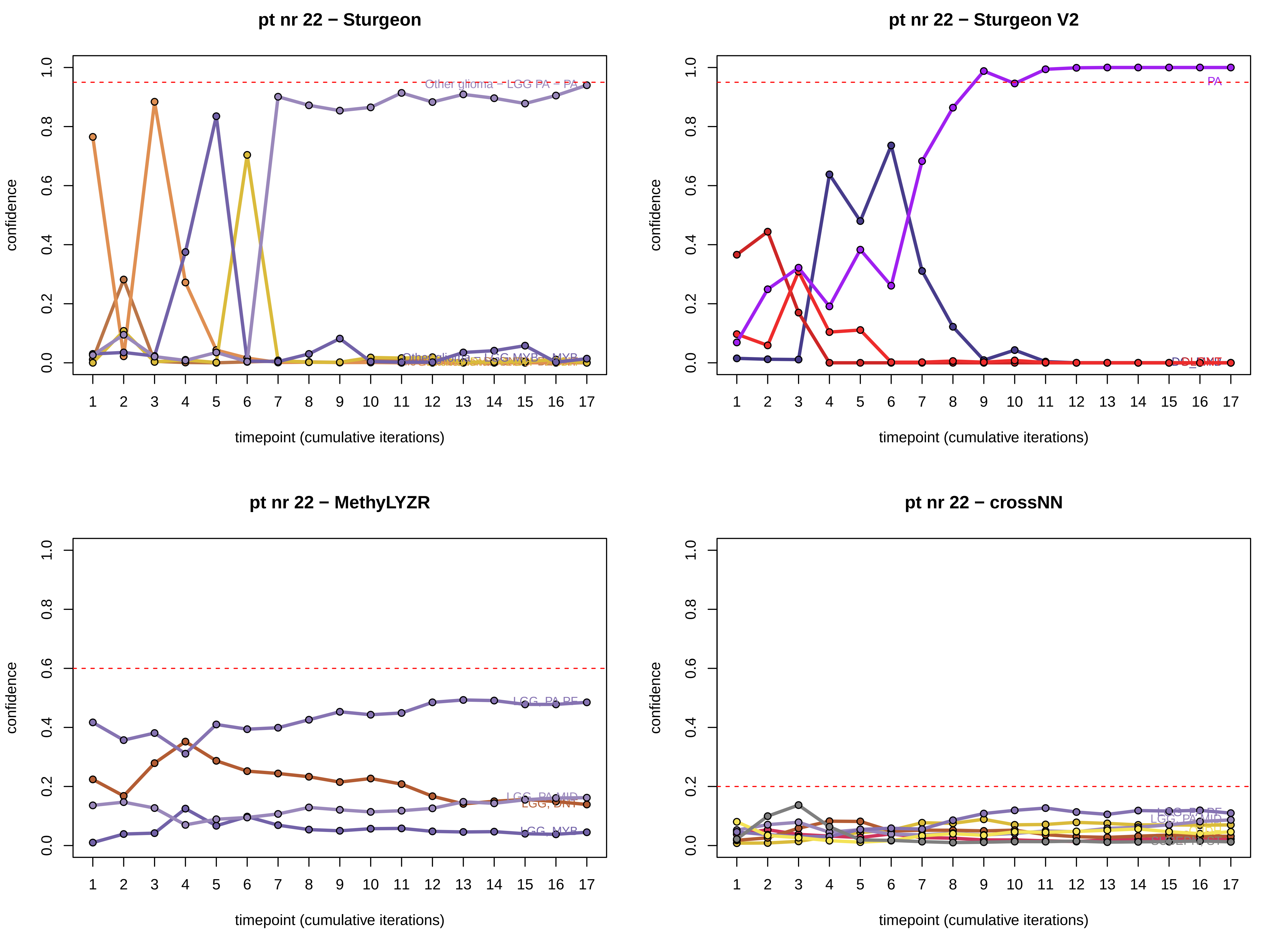

pt nr 22 − Sturgeon
Other glioma − LGG PA − PA
Other glioma − LGG MYB − MYB
pt nr 22 − Sturgeon V2
PA
pt nr 22 − MethyLYZR
LGG, PA PF
LGG, PA MID
LGG, DNT
LGG, MYB
pt nr 22 − crossNN
LGG, PA PF
LGG, PA MID
confidence
timepoint (cumulative iterations)
0.0
0.2
0.4
0.6
0.8
1.0

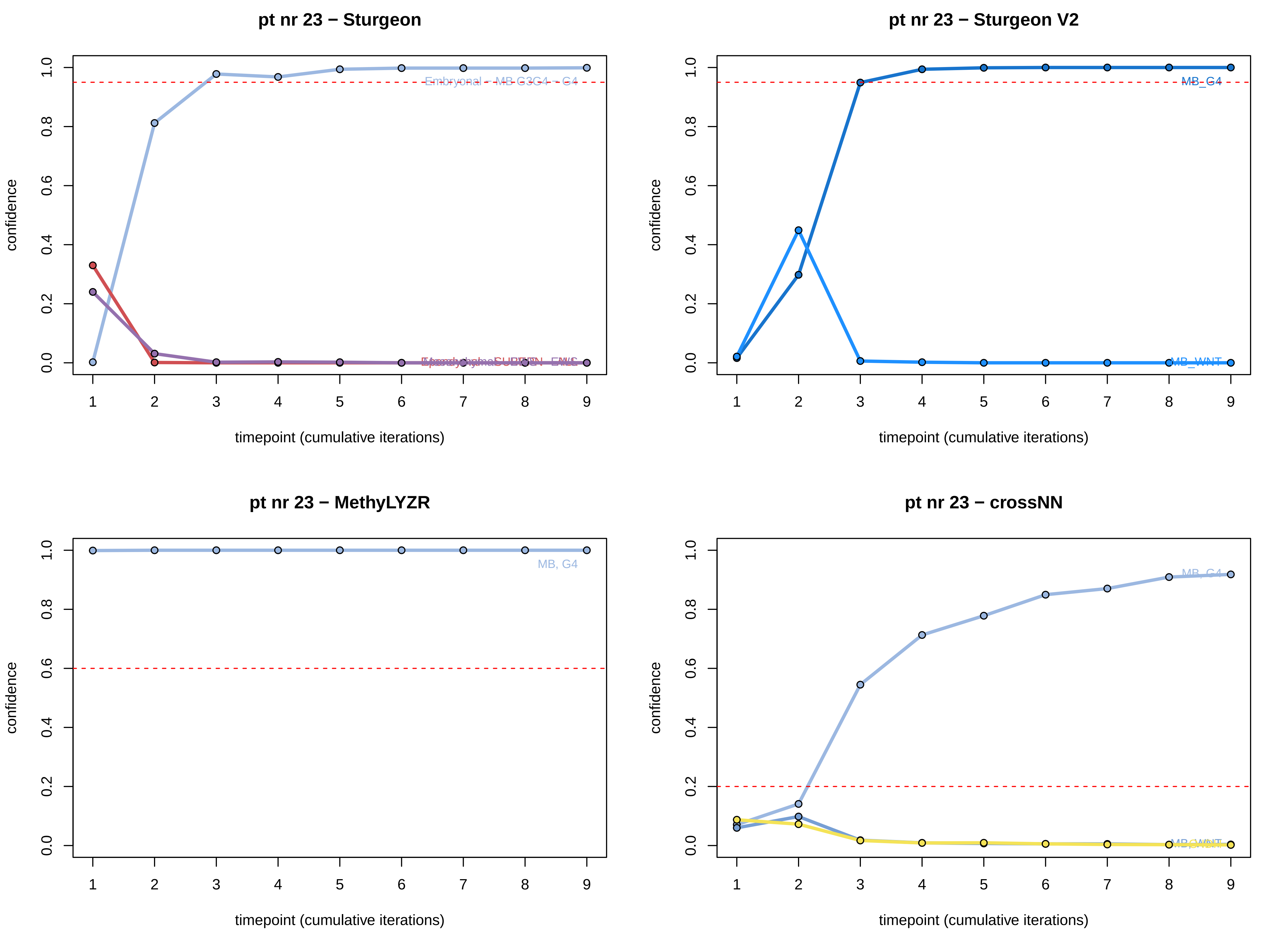
pt nr 23 − Sturgeon
Embryonal - MB G3G4 - G4
pt nr 23 − Sturgeon V2
MB_G4
MB_WNT
pt nr 23 − MethyLYZR
MB, G4
pt nr 23 − crossNN
MB, G4
confidence
timepoint (cumulative iterations)
0.0
0.2
0.4
0.6
0.8
1.0
1
2
3
4
5
6
7
8
9

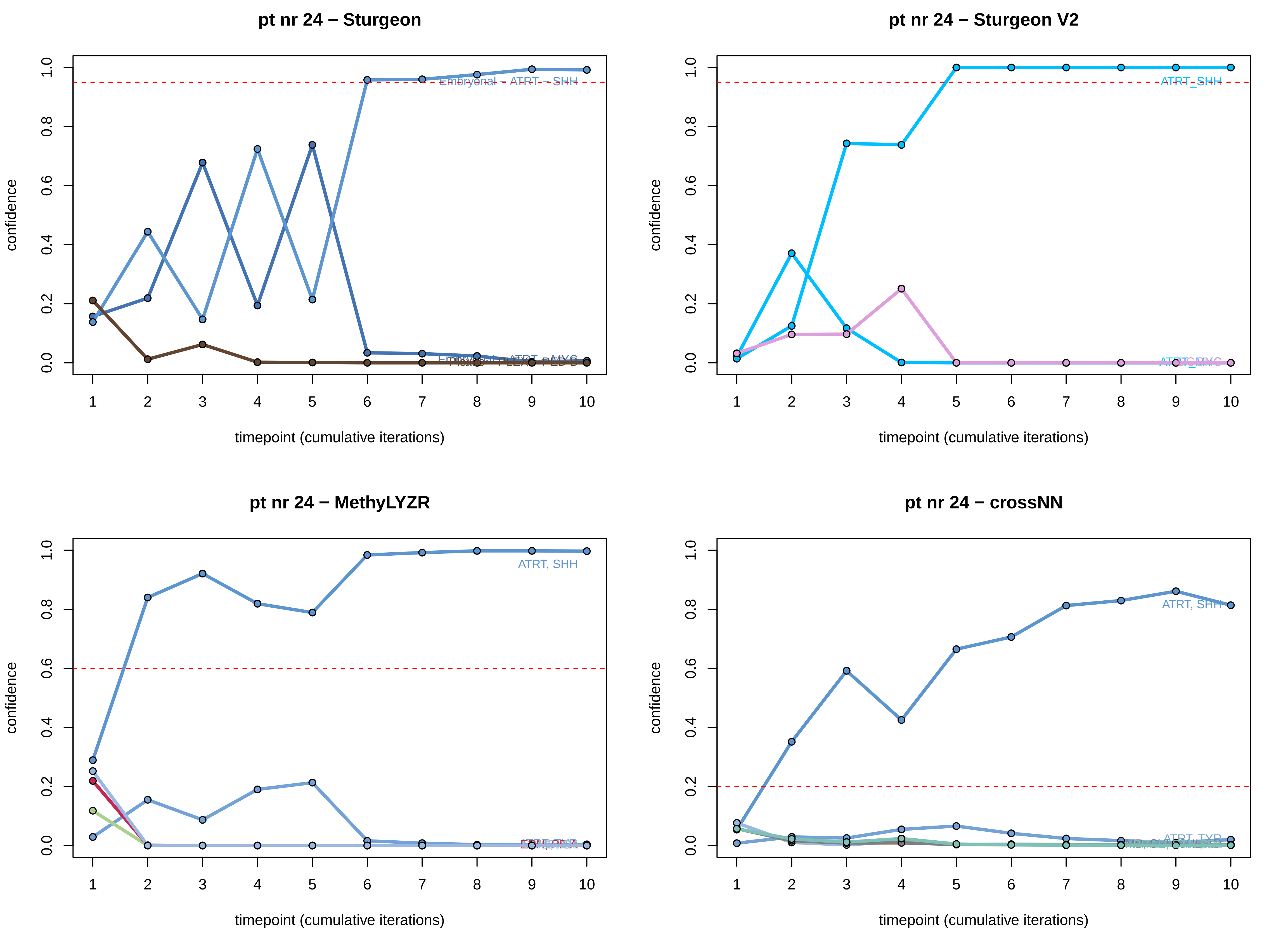

pt nr 24 − Sturgeon
Embryonal − ATRT − SHH
confidence
timepoint (cumulative iterations)
pt nr 24 − Sturgeon V2
ATRT_SHH
confidence
timepoint (cumulative iterations)
pt nr 24 − MethyLYZR
ATRT, SHH
confidence
timepoint (cumulative iterations)
pt nr 24 − crossNN
ATRT, SHH
ATRT, TYR
confidence
timepoint (cumulative iterations)

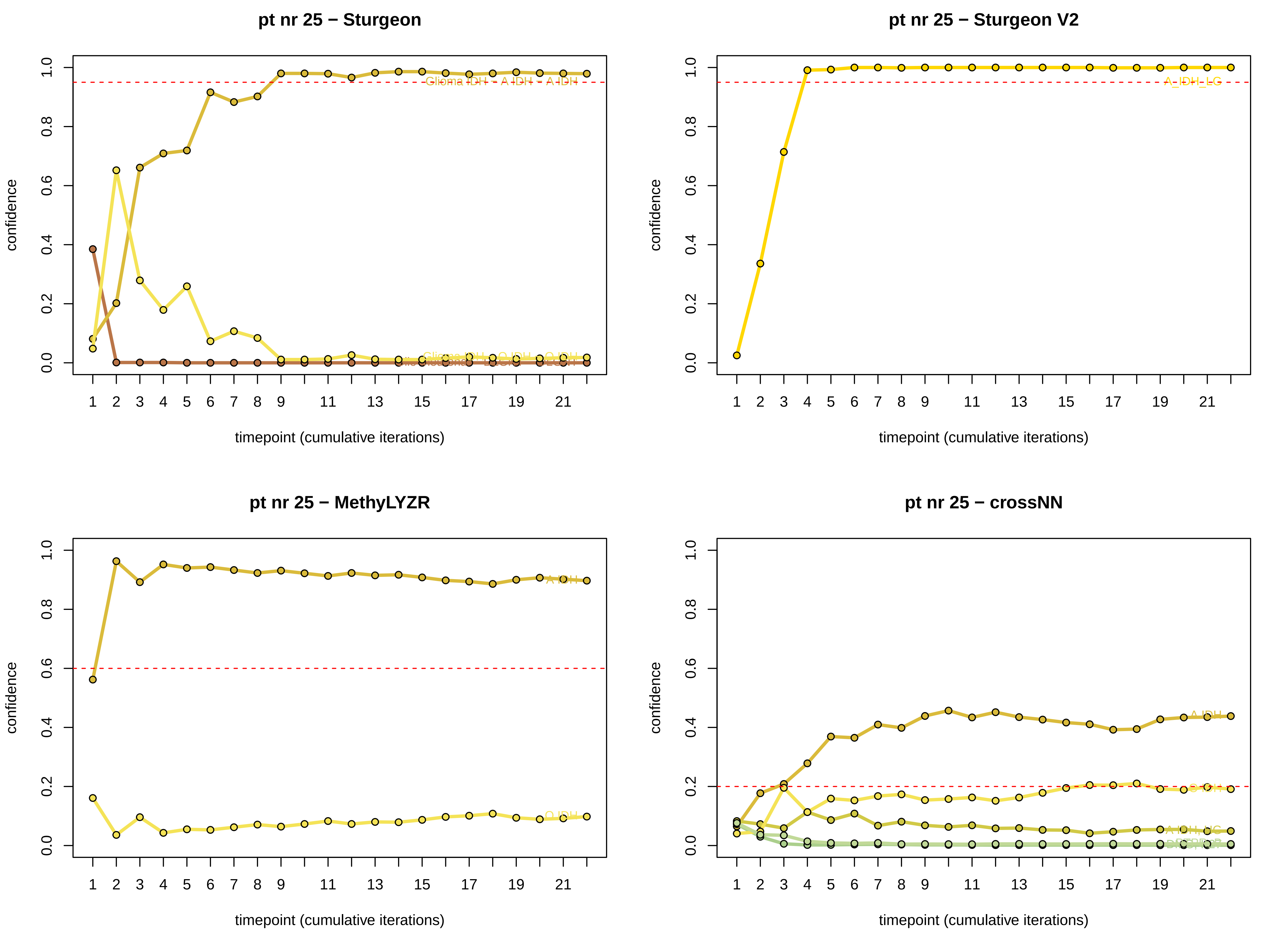

pt nr 25 − Sturgeon
pt nr 25 − Sturgeon V2
pt nr 25 − MethyLYZR
pt nr 25 − crossNN
confidence
timepoint (cumulative iterations)
Glioma IDH − A IDH − A IDH
A_IDH_LG
A IDH
O IDH

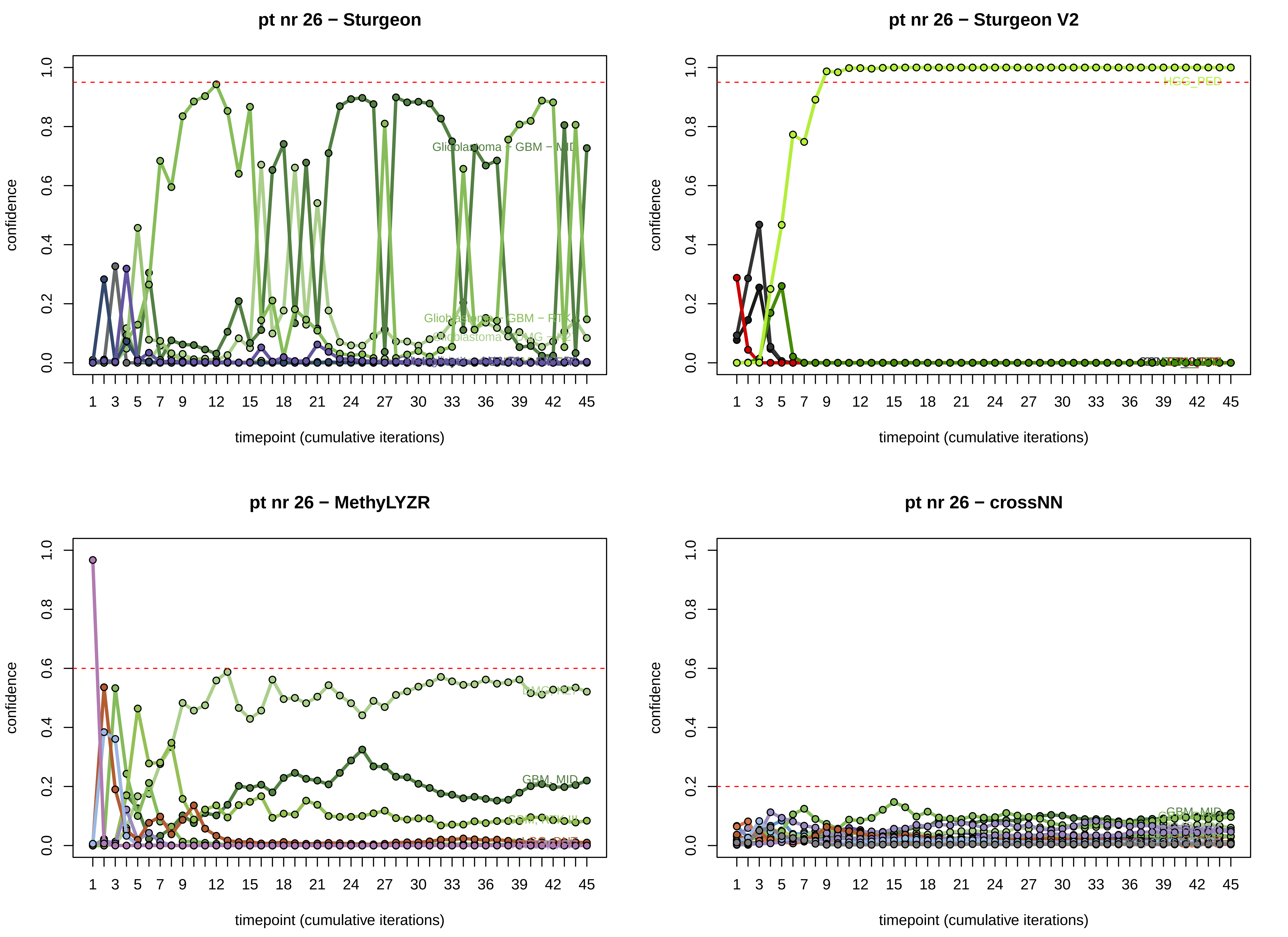
pt nr 26 − Sturgeon
pt nr 26 − Sturgeon V2
pt nr 26 − MethyLYZR
pt nr 26 − crossNN
confidence
timepoint (cumulative iterations)
Glioblastoma − GBM − MID
HGG_PED
DMG_K27
GBM, MID
LGG, DNT

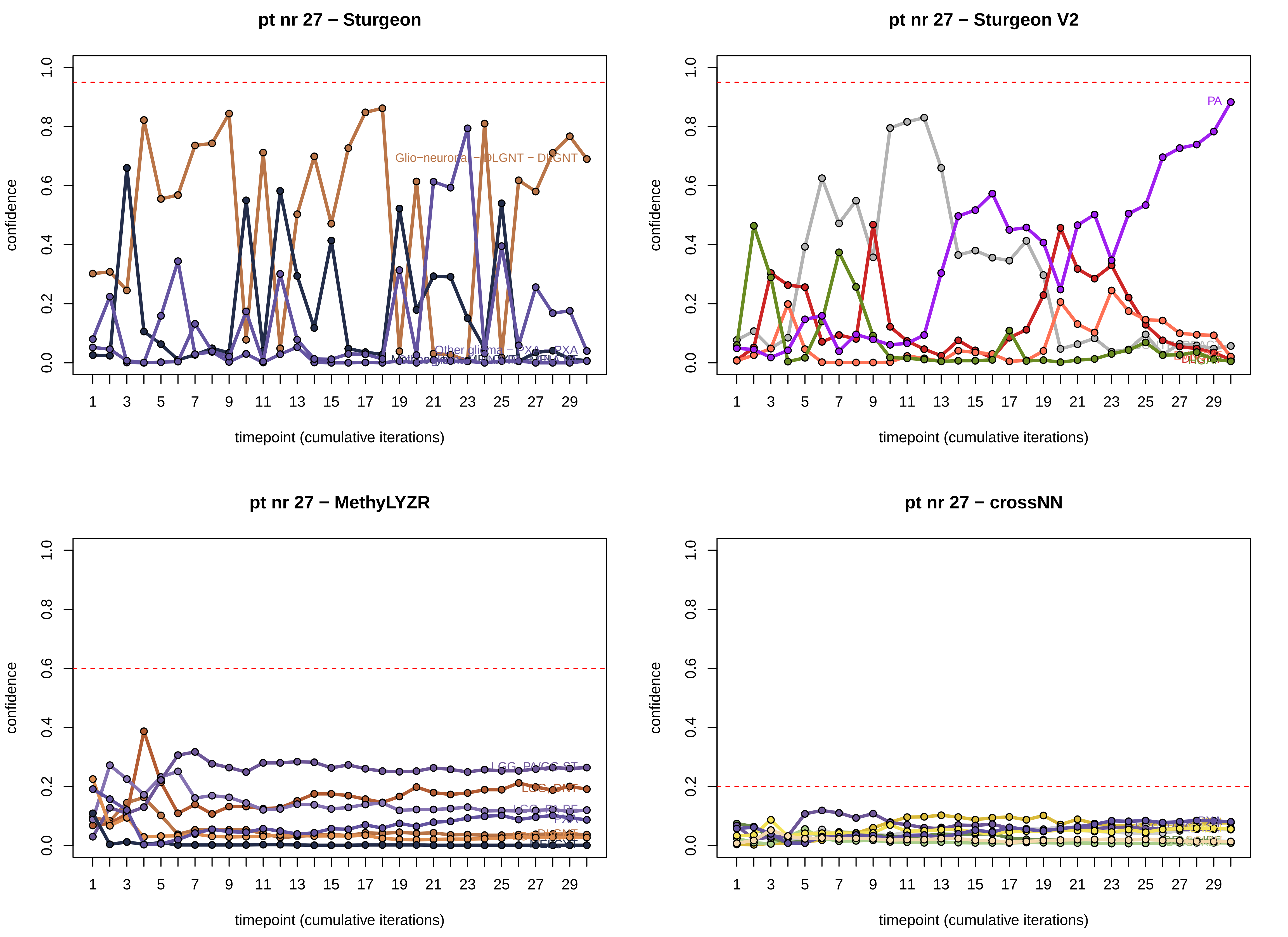
pt nr 27 − Sturgeon
Glio−neuronal − DLGNT − DLGNT
Other glioma − PXA − PXA
pt nr 27 − Sturgeon V2
PA
pt nr 27 − MethyLYZR
LGG, PA/GG ST
LGG, DNT
LGG, PA PF
pt nr 27 − crossNN
confidence
timepoint (cumulative iterations)

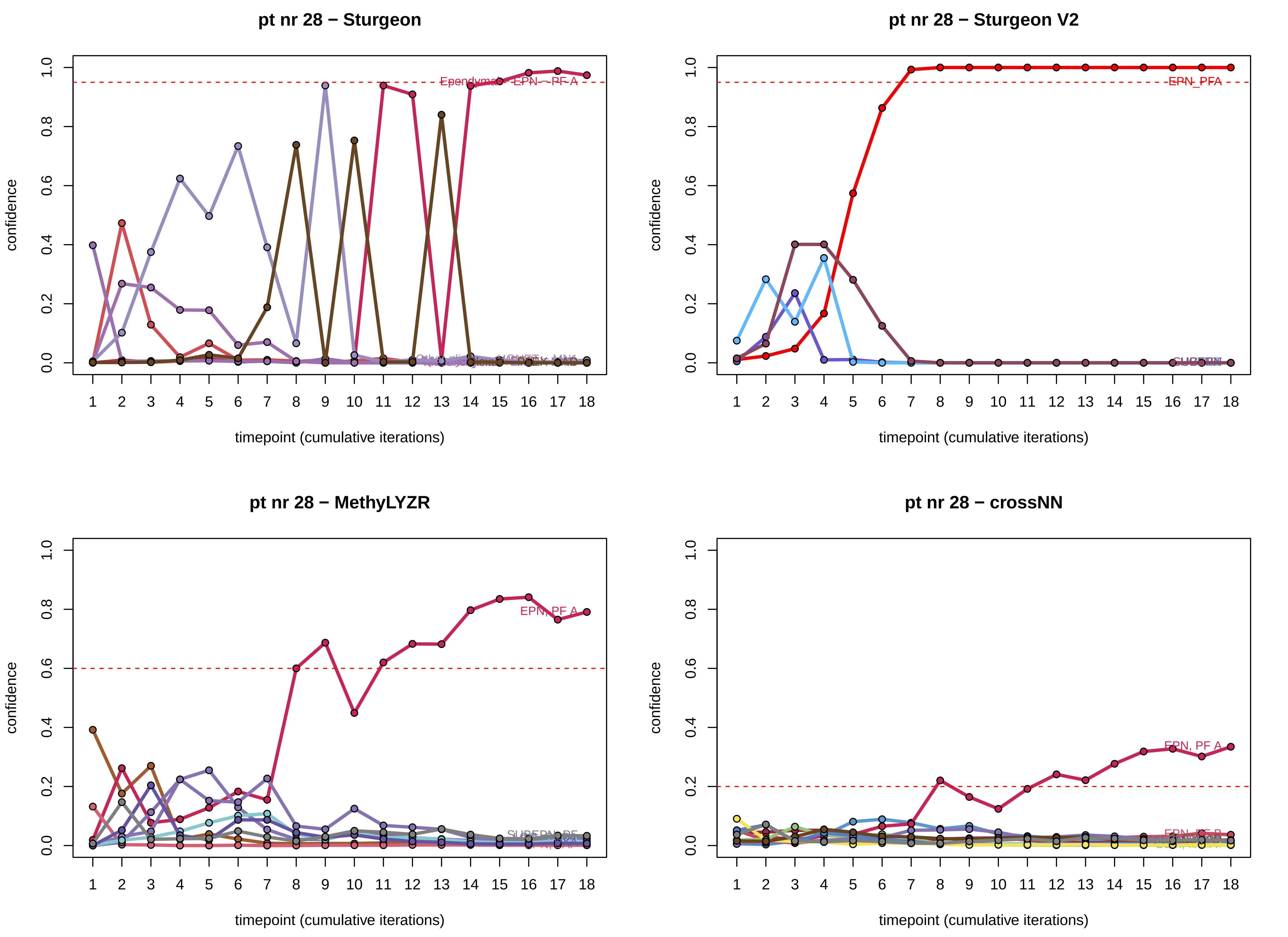
pt nr 28 − Sturgeon
pt nr 28 − Sturgeon V2
pt nr 28 − MethyLYZR
pt nr 28 − crossNN
confidence
timepoint (cumulative iterations)
0.0
0.2
0.4
0.6
0.8
1.0
1 2 3 4 5 6 7 8 9 10 11 12 13 14 15 16 17 18
Ependymal - EPN - PF A
EPN_PFA
EPN, PF A
EPN, PF B

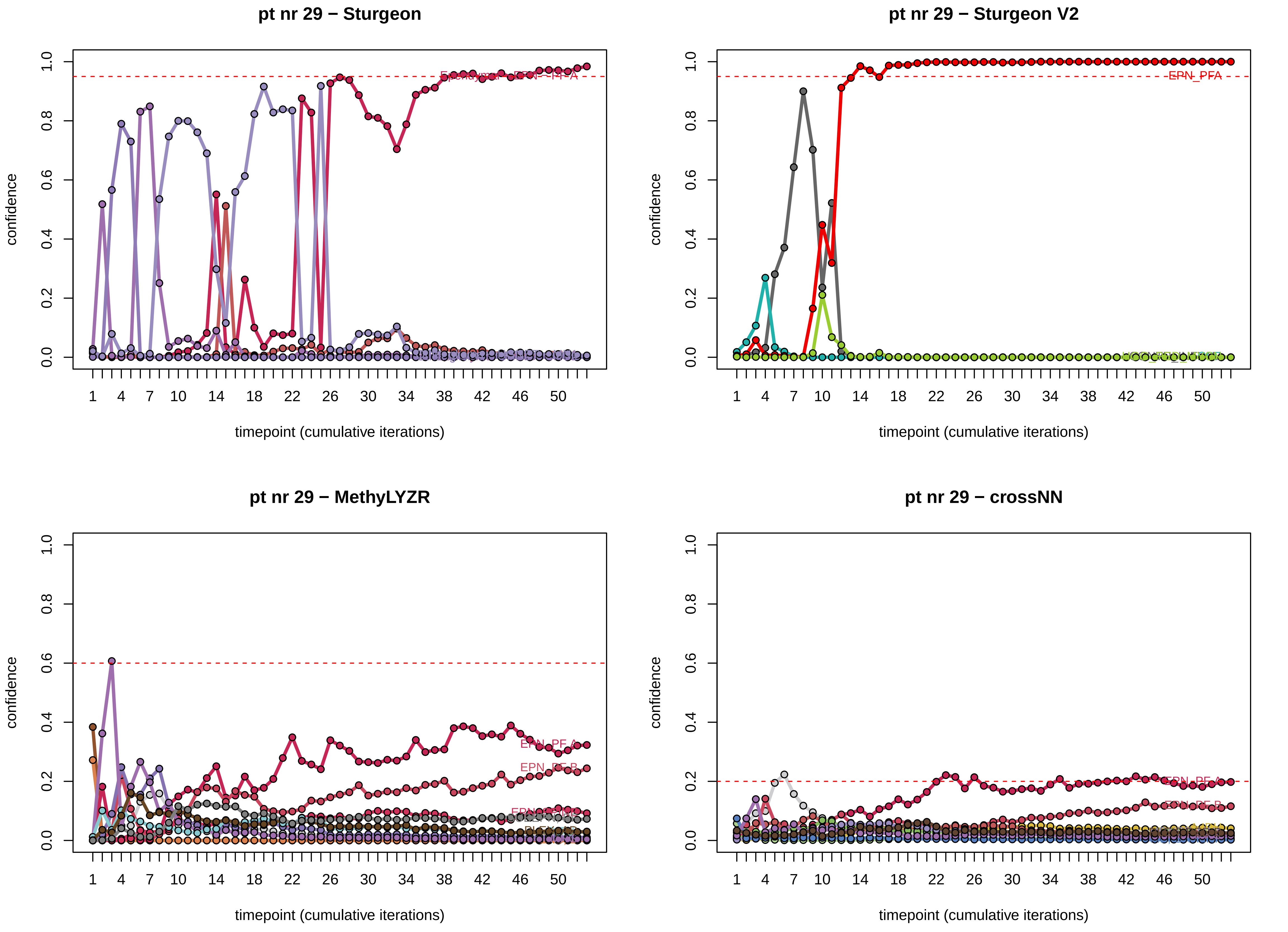
pt nr 29 − Sturgeon
pt nr 29 − Sturgeon V2
pt nr 29 − MethyLYZR
pt nr 29 − crossNN
confidence
timepoint (cumulative iterations)
EPN_PFA
EPN_PF_A
EPN_PF_B

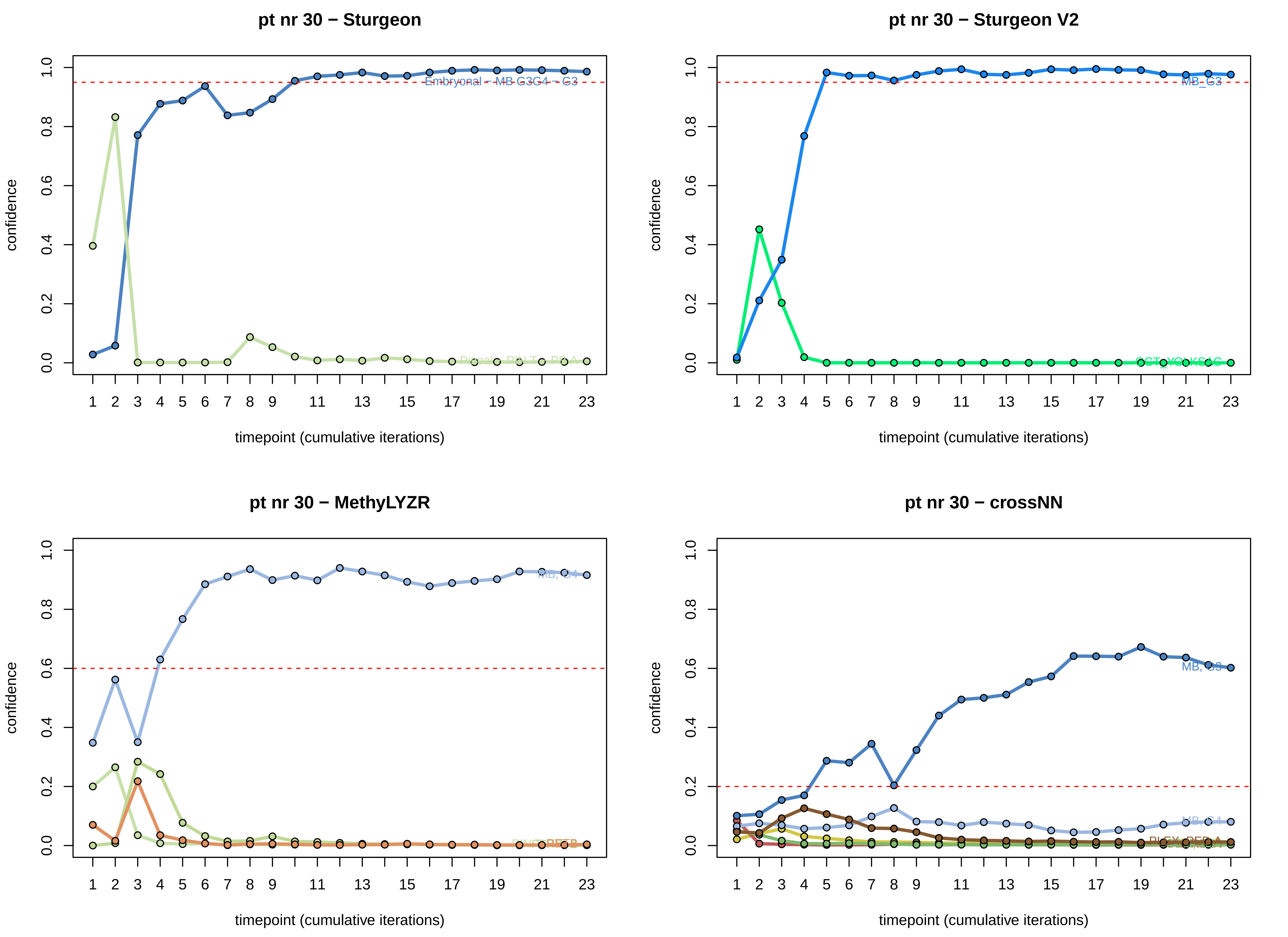

pt nr 30 − Sturgeon
pt nr 30 − Sturgeon V2
pt nr 30 − MethyLYZR
pt nr 30 − crossNN
confidence
timepoint (cumulative iterations)
Embryonal − MB G3G4 − G3
MB_G3
MB, G4
MB, G3

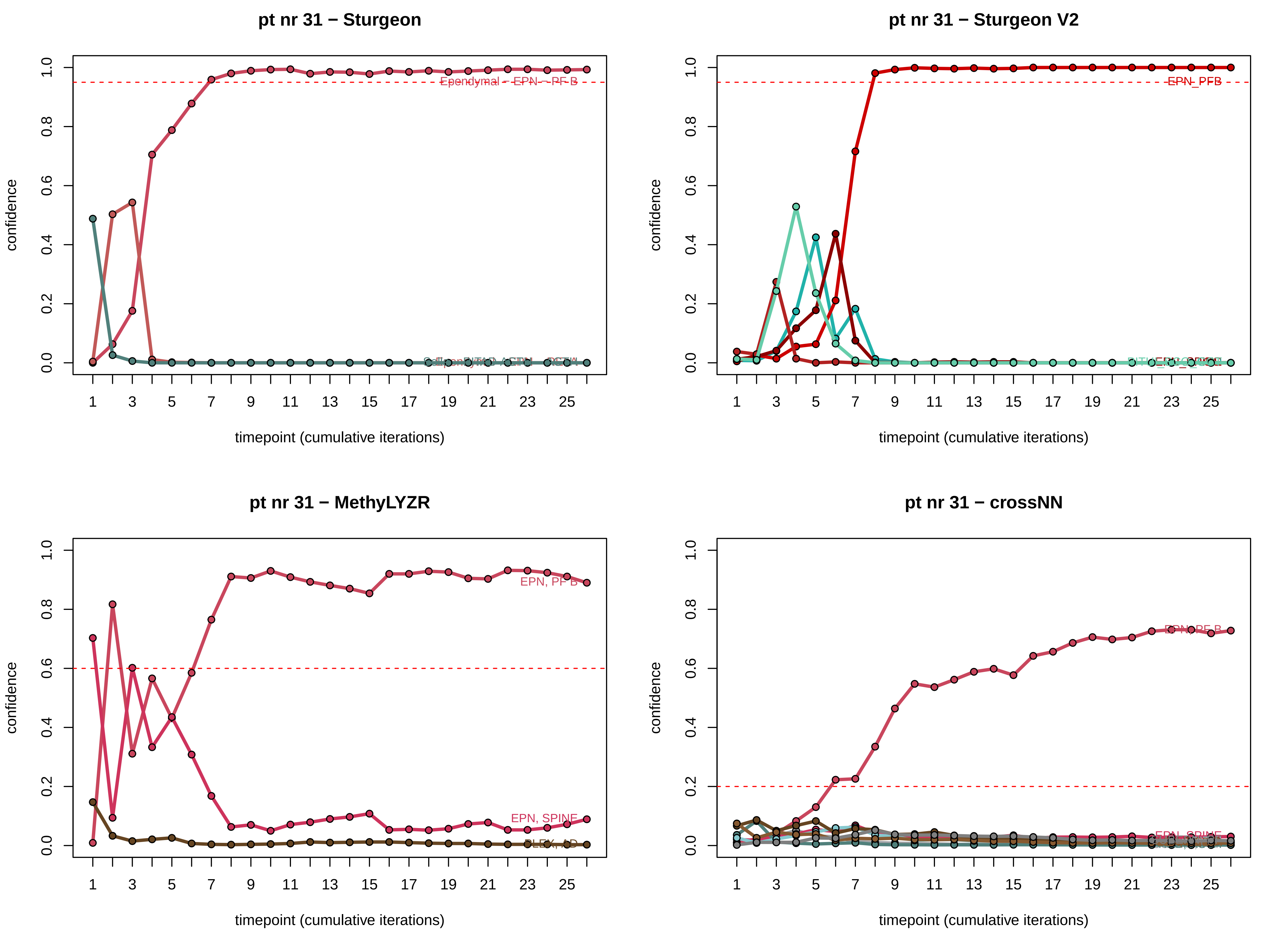

pt nr 31 − Sturgeon
confidence
timepoint (cumulative iterations)
Ependymal - EPN - PF B
pt nr 31 − Sturgeon V2
confidence
timepoint (cumulative iterations)
EPN_PFB
pt nr 31 − MethyLYZR
confidence
timepoint (cumulative iterations)
EPN, PF B
EPN, SPINE
pt nr 31 − crossNN
confidence
timepoint (cumulative iterations)
EPN, PF B
EPN, SPINE

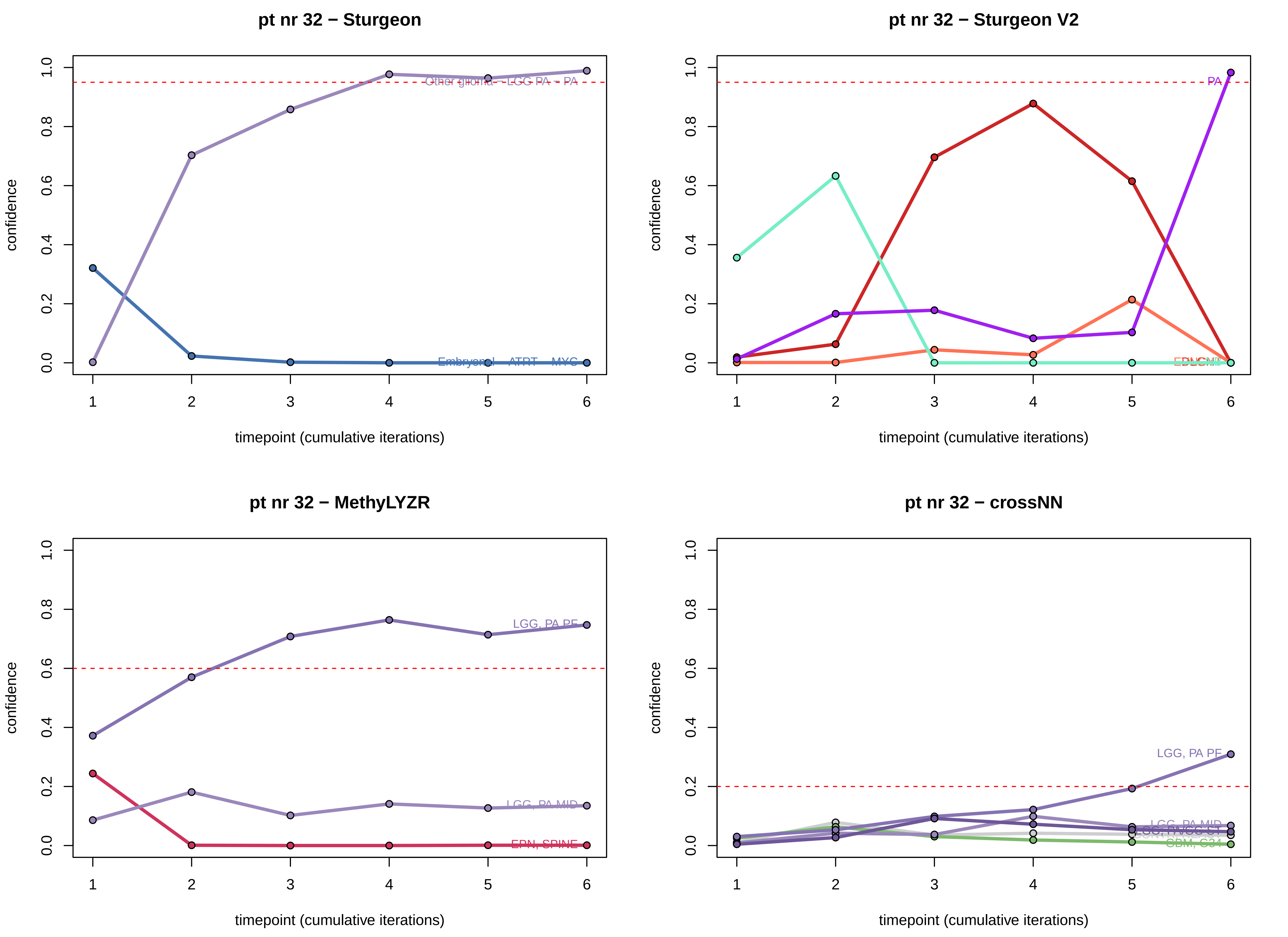

pt nr 32 − Sturgeon
confidence
0.0
0.2
0.4
0.6
0.8
1.0
Other glioma - LGG PA - PA
Embryonal - ATRT - MYC
1
2
3
4
5
6
timepoint (cumulative iterations)
pt nr 32 − Sturgeon V2
confidence
PA
timepoint (cumulative iterations)
pt nr 32 − MethyLYZR
confidence
LGG, PA PF
LGG, PA MID
EPN, SPINE
timepoint (cumulative iterations)
pt nr 32 − crossNN
confidence
LGG, PA PF
LGG, PA MID
GBM, G34
timepoint (cumulative iterations)

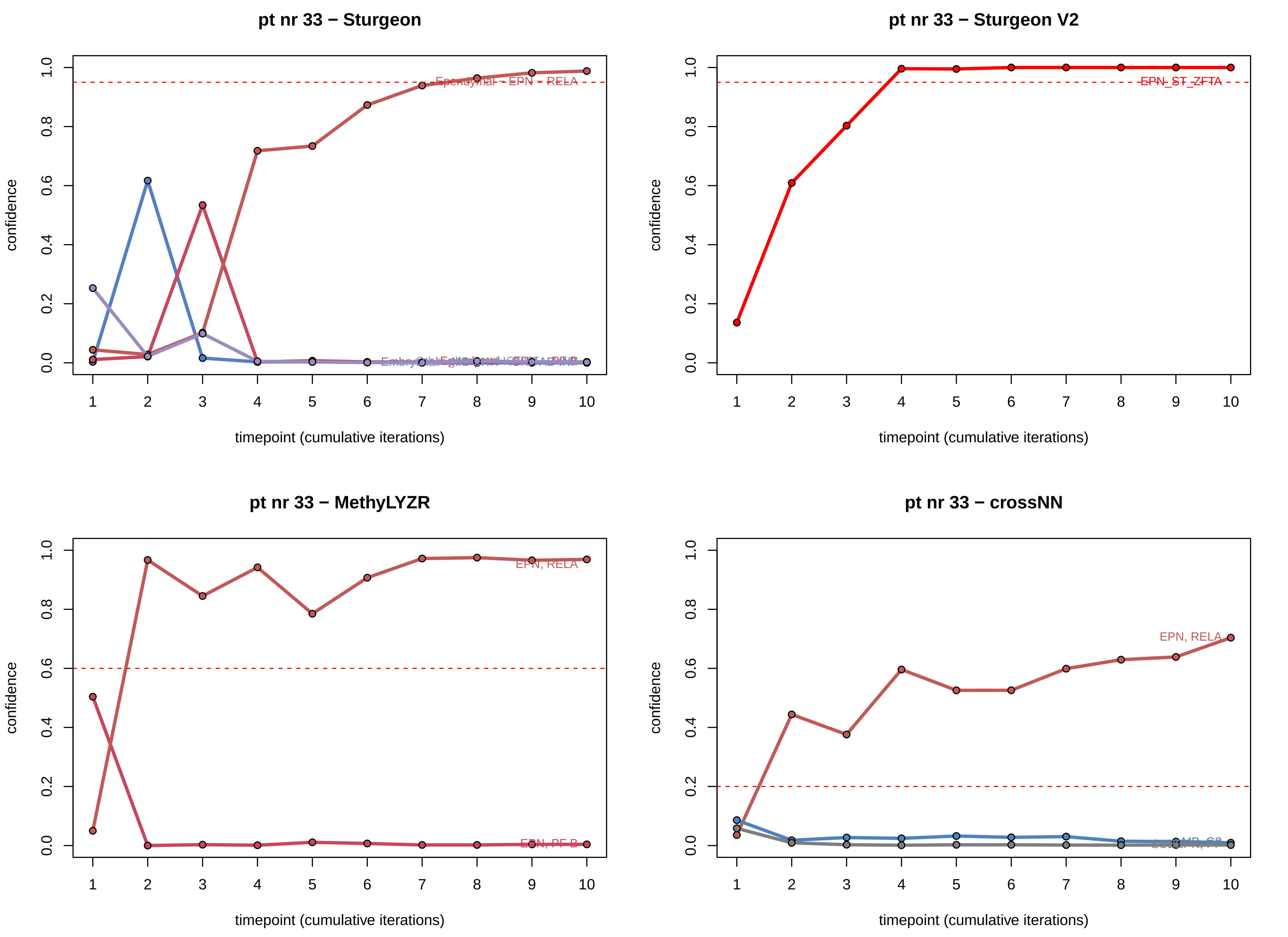
pt nr 33 − Sturgeon
confidence
timepoint (cumulative iterations)
Ependymal − EPN − RELA
pt nr 33 − Sturgeon V2
confidence
timepoint (cumulative iterations)
EPN_ST_ZFTA
pt nr 33 − MethyLYZR
confidence
timepoint (cumulative iterations)
EPN, RELA
EPN, PF-B
pt nr 33 − crossNN
confidence
timepoint (cumulative iterations)
EPN, RELA

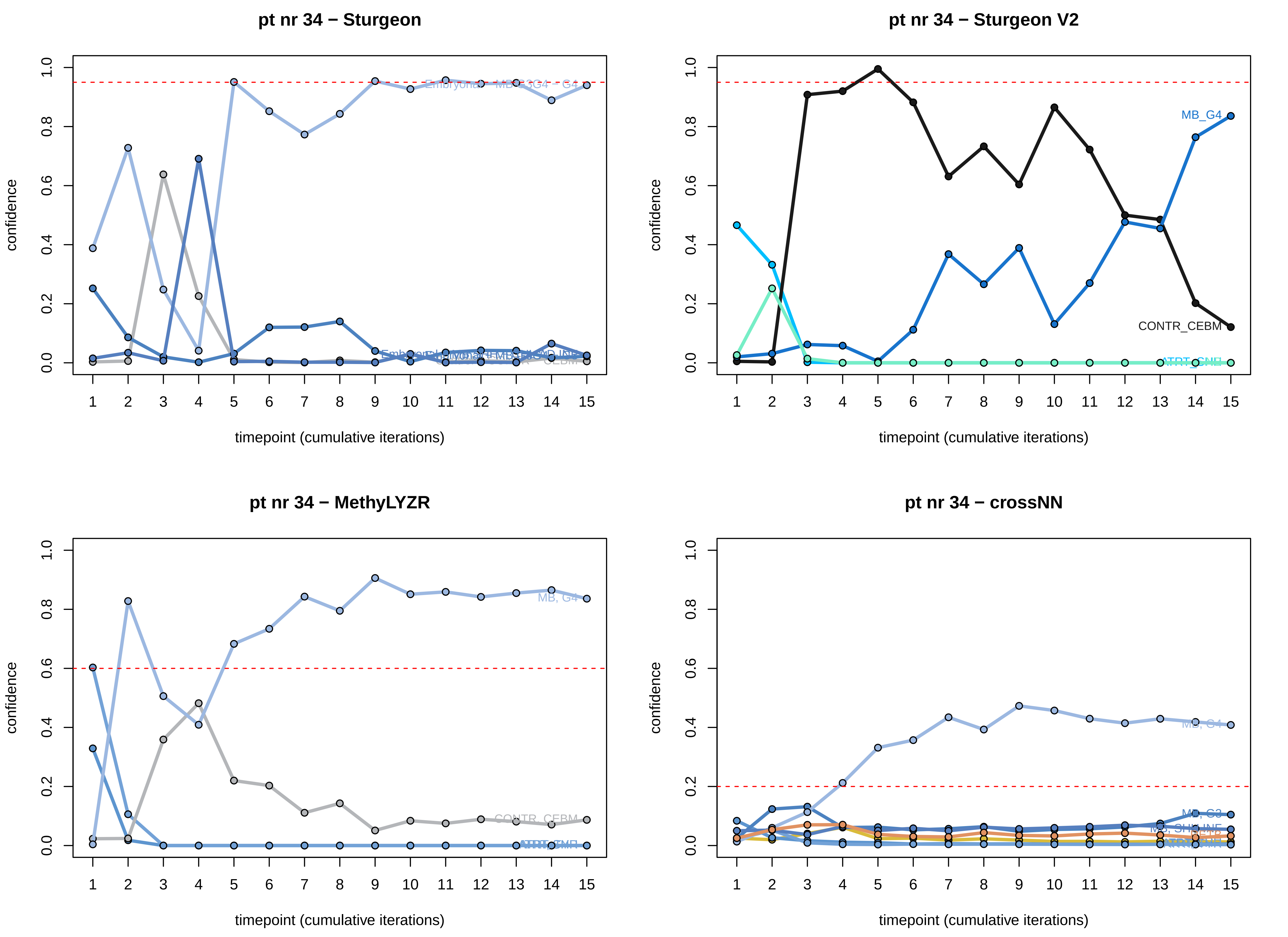

pt nr 34 − Sturgeon
confidence
timepoint (cumulative iterations)
pt nr 34 − Sturgeon V2
MB_G4
CONTR_CEBM
confidence
timepoint (cumulative iterations)
pt nr 34 − MethyLYZR
MB, G4
CONTR_CEBM
confidence
timepoint (cumulative iterations)
pt nr 34 − crossNN
MB, G4
MB, G3
confidence
timepoint (cumulative iterations)

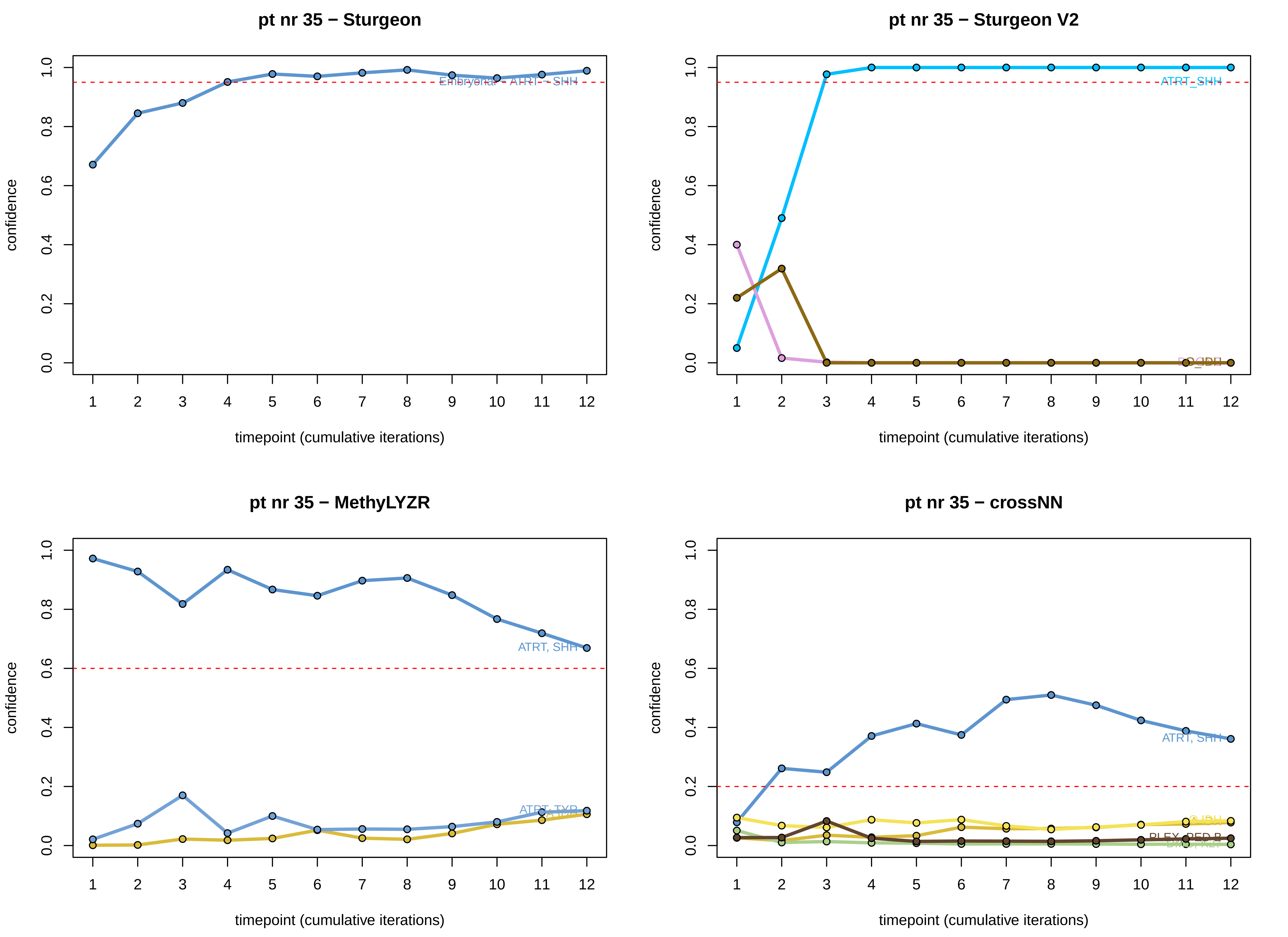

pt nr 35 − Sturgeon
Embryonal - ATRT - SHH
pt nr 35 − Sturgeon V2
ATRT_SHH
pt nr 35 − MethyLYZR
ATRT, SHH
ATRT, TYR
pt nr 35 − crossNN
ATRT, SHH
confidence
timepoint (cumulative iterations)
0.0
0.2
0.4
0.6
0.8
1.0
1
2
3
4
5
6
7
8
9
10
11
12

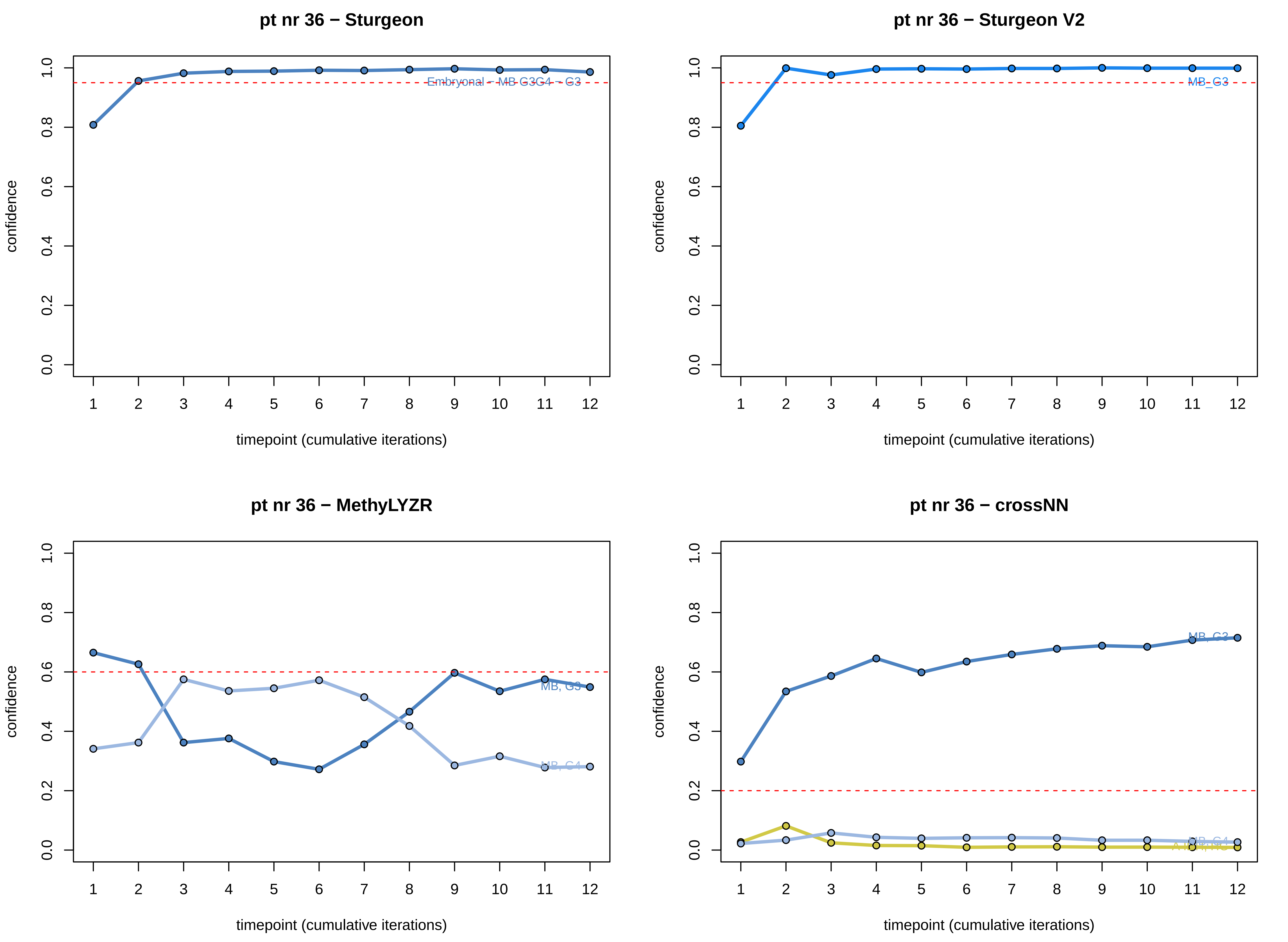

pt nr 36 − Sturgeon
Embryonal - MB G3G4 - G3
pt nr 36 − Sturgeon V2
MB_G3
pt nr 36 − MethyLYZR
MB, G3
MB, G4
pt nr 36 − crossNN
MB, G3
MB, G4
confidence
timepoint (cumulative iterations)
0.0
0.2
0.4
0.6
0.8
1.0
1
2
3
4
5
6
7
8
9
10
11
12

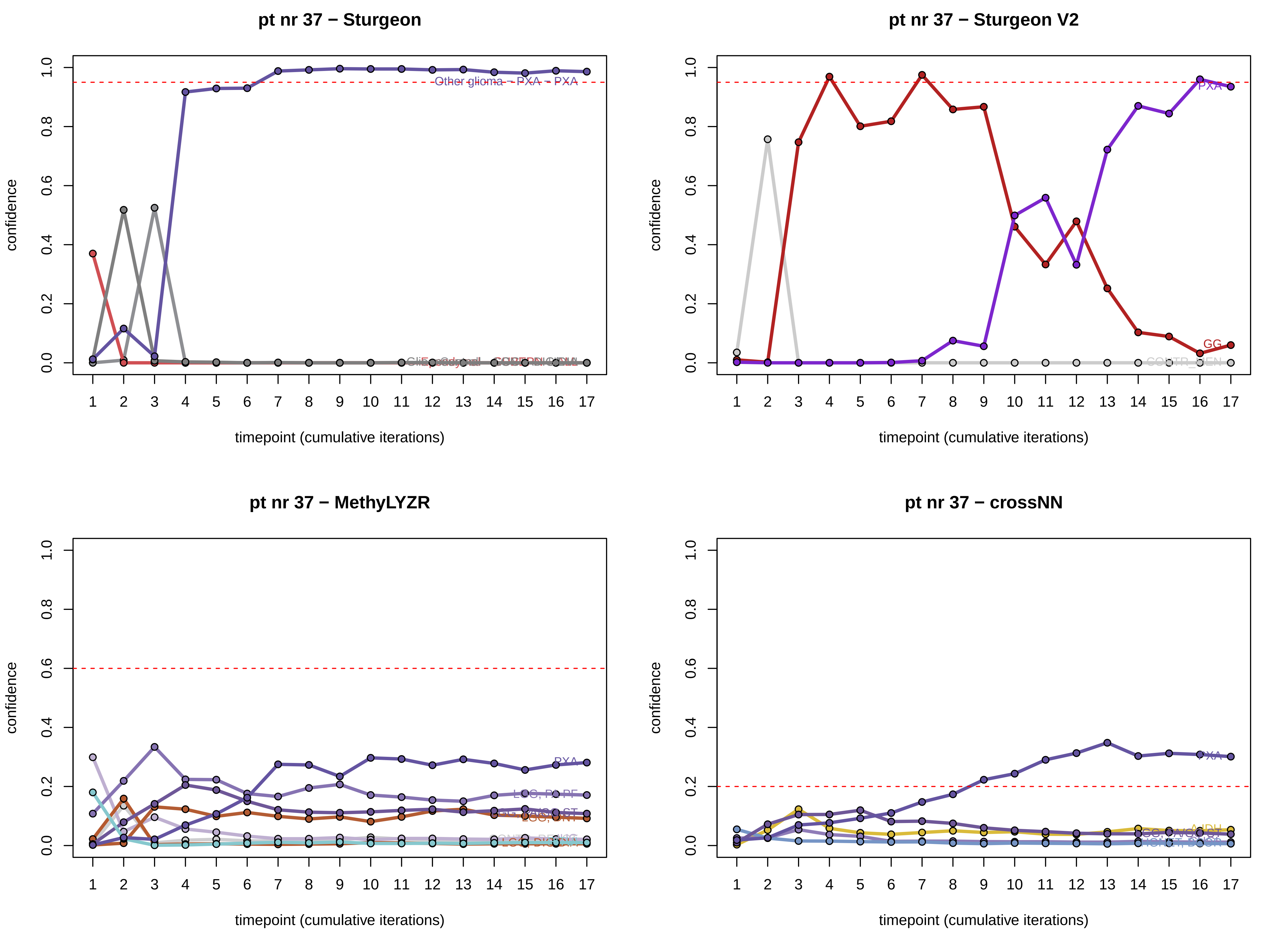

pt nr 37 − Sturgeon
Other glioma − PXA − PXA
confidence
timepoint (cumulative iterations)
pt nr 37 − Sturgeon V2
PXA
GG
confidence
timepoint (cumulative iterations)
pt nr 37 − MethyLYZR
PXA
LGG, PA PF
LGG, DNT
confidence
timepoint (cumulative iterations)
pt nr 37 − crossNN
PXA
A IDH
confidence
timepoint (cumulative iterations)

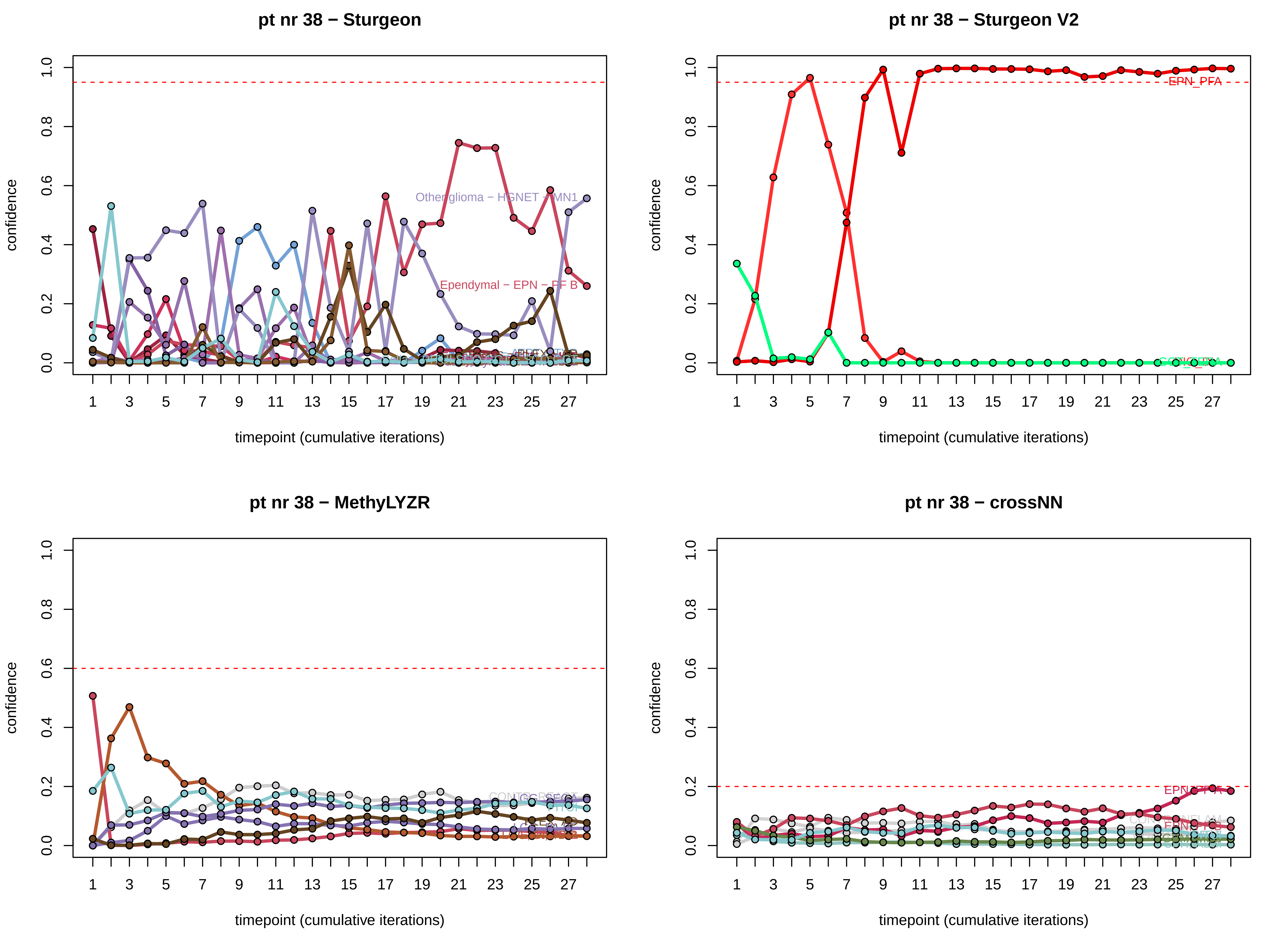
pt nr 38 − Sturgeon
confidence
timepoint (cumulative iterations)
Other glioma − HGNET − MN1
Ependymal − EPN − PF B
pt nr 38 − Sturgeon V2
EPN_PFA
pt nr 38 − MethyLYZR
pt nr 38 − crossNN

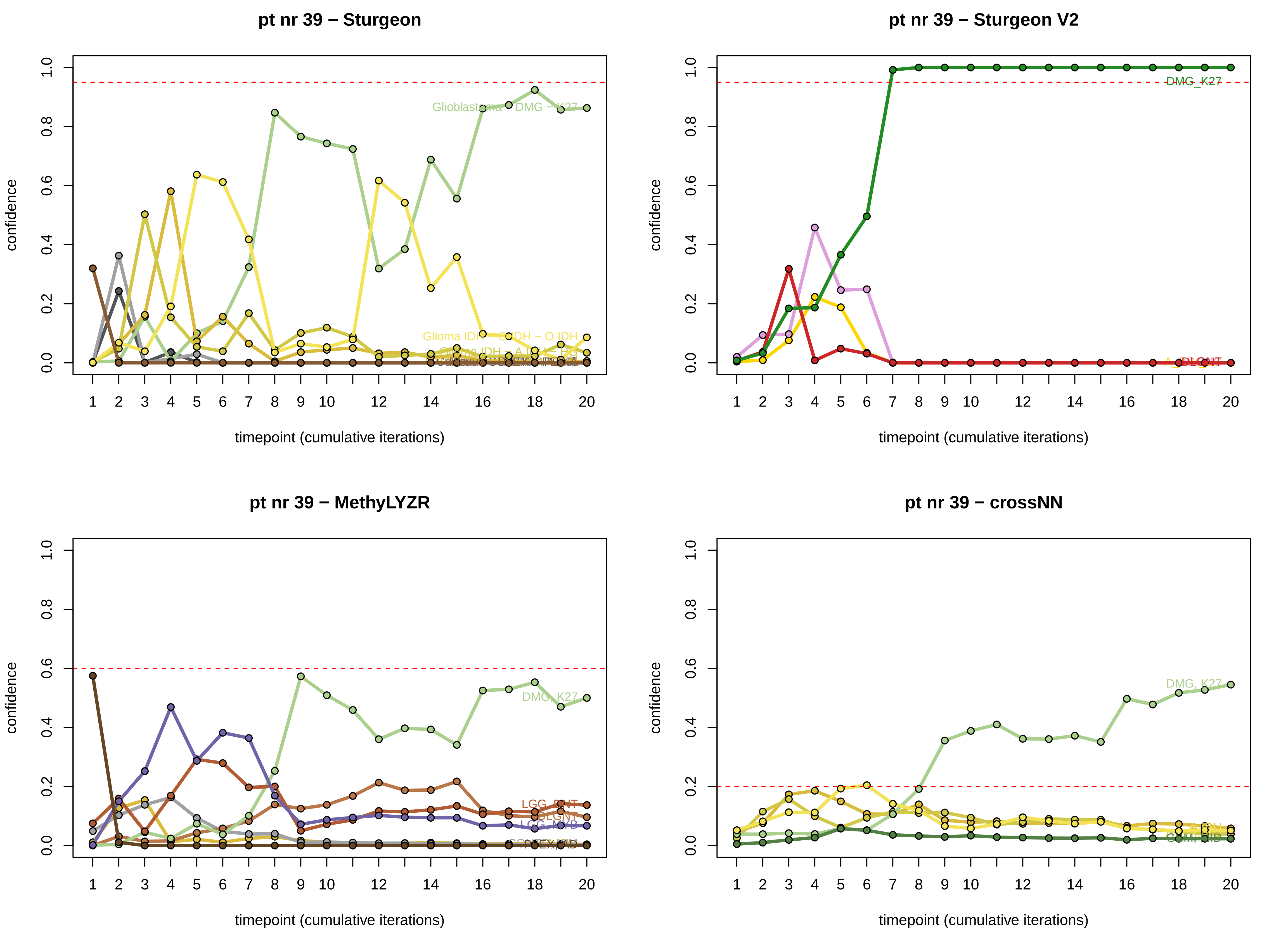
pt nr 39 − Sturgeon
pt nr 39 − Sturgeon V2
pt nr 39 − MethyLYZR
pt nr 39 − crossNN
confidence
timepoint (cumulative iterations)
DMG_K27
DMG_K27
DMG_K27
LGG_DNT
LGG_MYB
GBM_MID

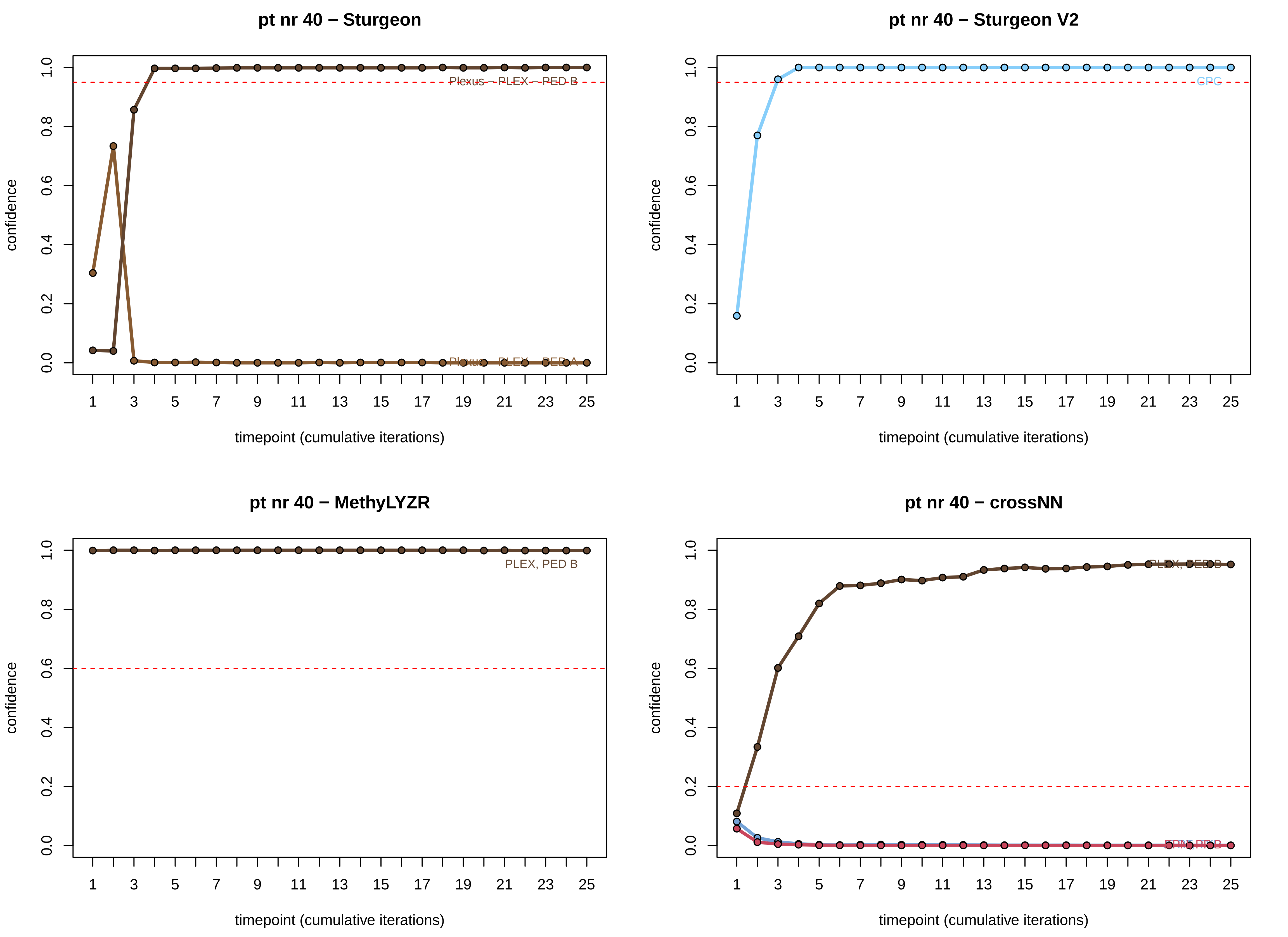
pt nr 40 − Sturgeon
Plexus – PLEX – PED B
Plexus – PLEX – PED A
confidence
timepoint (cumulative iterations)
pt nr 40 − Sturgeon V2
CPG
confidence
timepoint (cumulative iterations)
pt nr 40 − MethyLYZR
PLEX, PED B
confidence
timepoint (cumulative iterations)
pt nr 40 − crossNN
PLEX, PED B
confidence
timepoint (cumulative iterations)

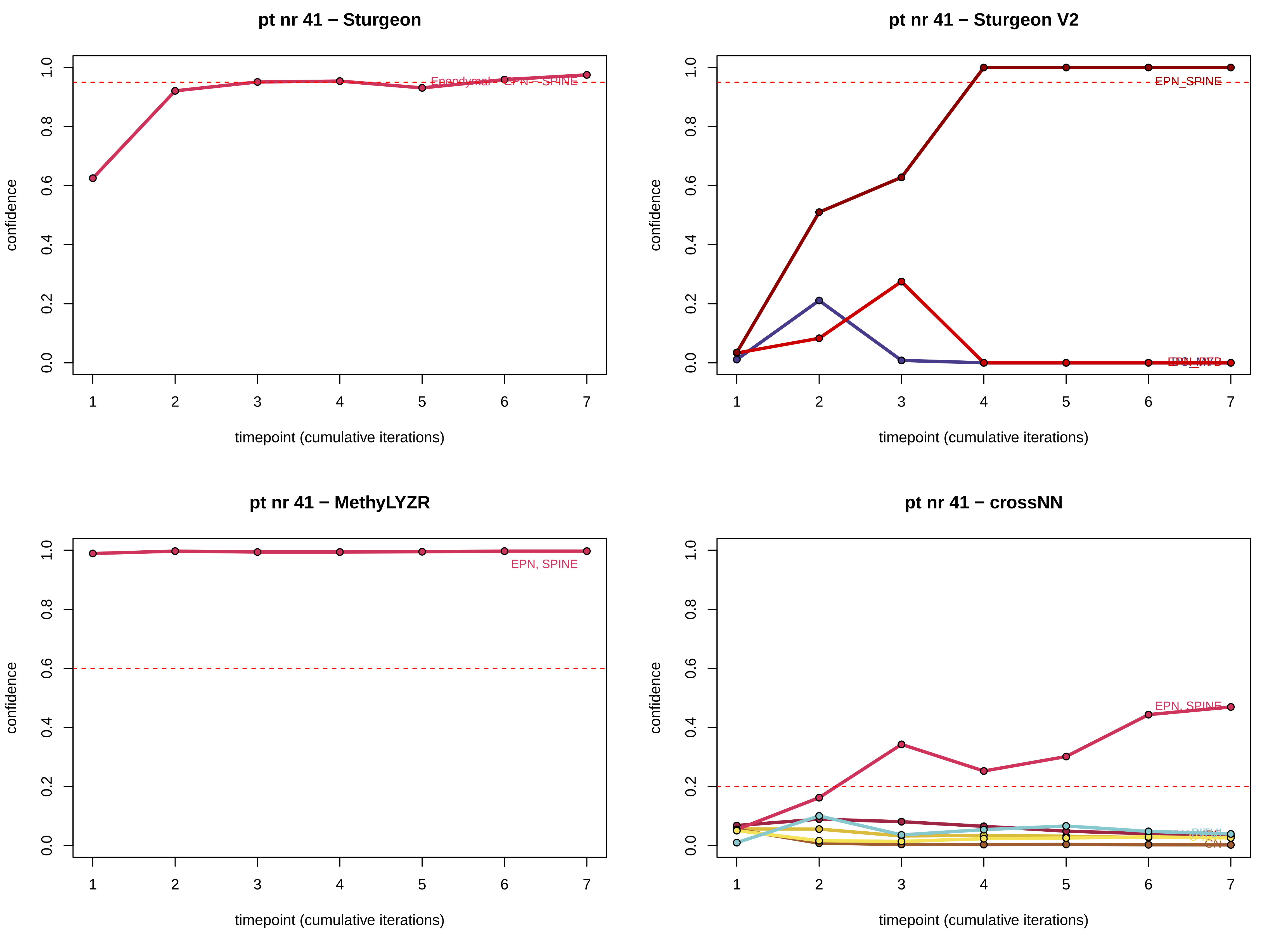
pt nr 41 − Sturgeon
Ependymal
EPN − SPINE
pt nr 41 − Sturgeon V2
EPN_SPINE
pt nr 41 − MethyLYZR
EPN, SPINE
pt nr 41 − crossNN
EPN, SPINE
confidence
timepoint (cumulative iterations)

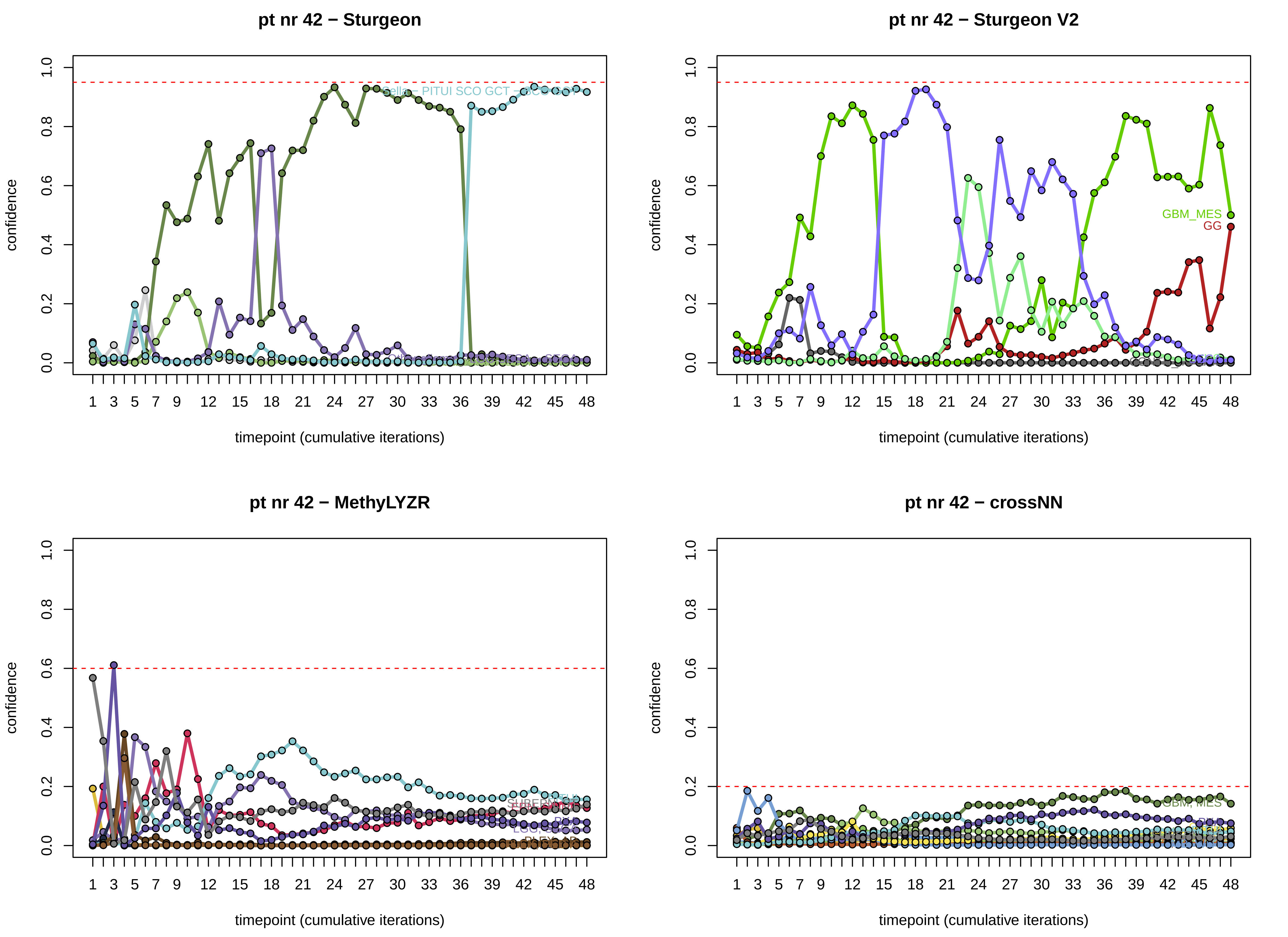
pt nr 42 − Sturgeon
pt nr 42 − Sturgeon V2
pt nr 42 − MethyLYZR
pt nr 42 − crossNN
confidence
timepoint (cumulative iterations)
GBM_MES
GG
0.0
0.2
0.4
0.6
0.8
1.0
1
3
5
7
9
12
15
18
21
24
27
30
33
36
39
42
45
48

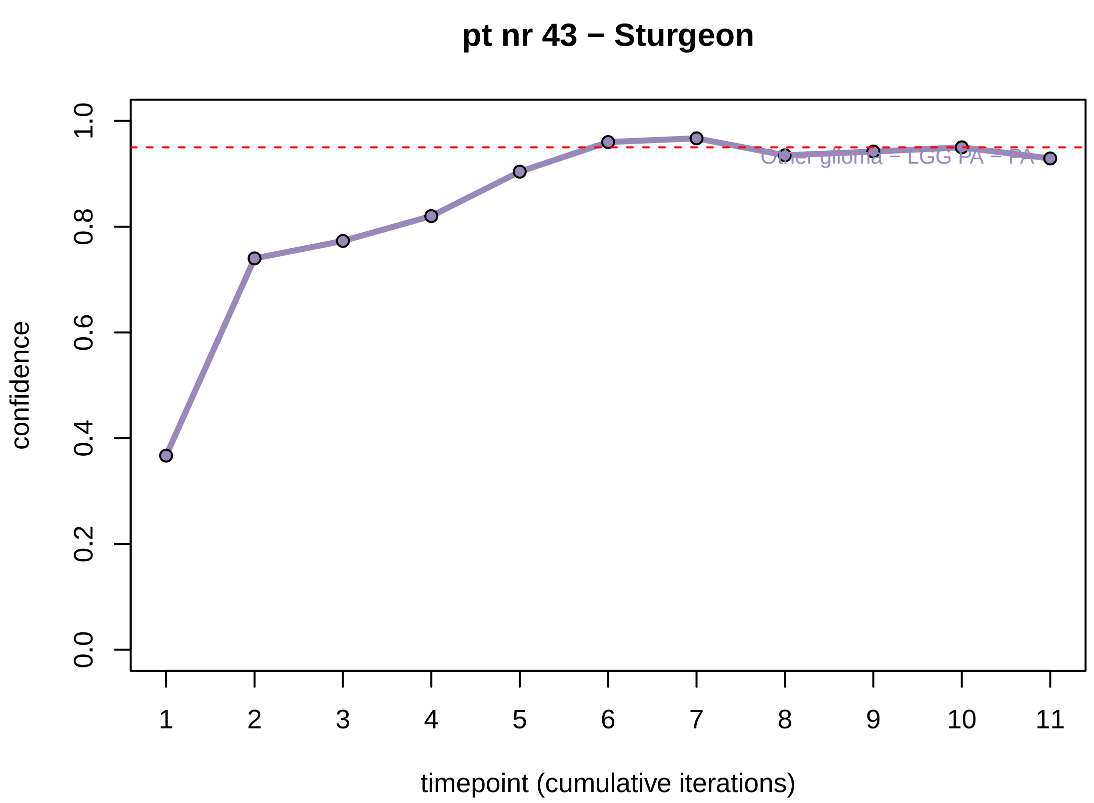
pt nr 43 − Sturgeon
confidence
0.0
0.2
0.4
0.6
0.8
1.0
1 2 3 4 5 6 7 8 9 10 11
timepoint (cumulative iterations)

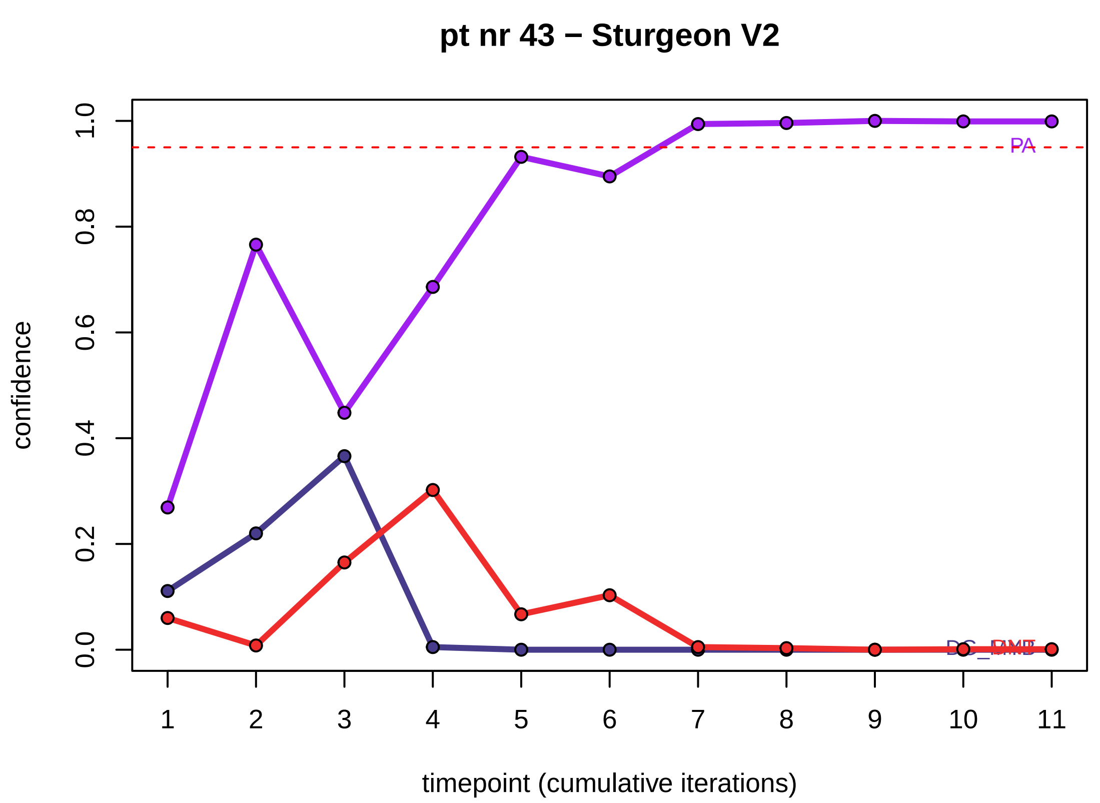
pt nr 43 − Sturgeon V2
PA
confidence
0.0
0.2
0.4
0.6
0.8
1.0
1 2 3 4 5 6 7 8 9 10 11
timepoint (cumulative iterations)

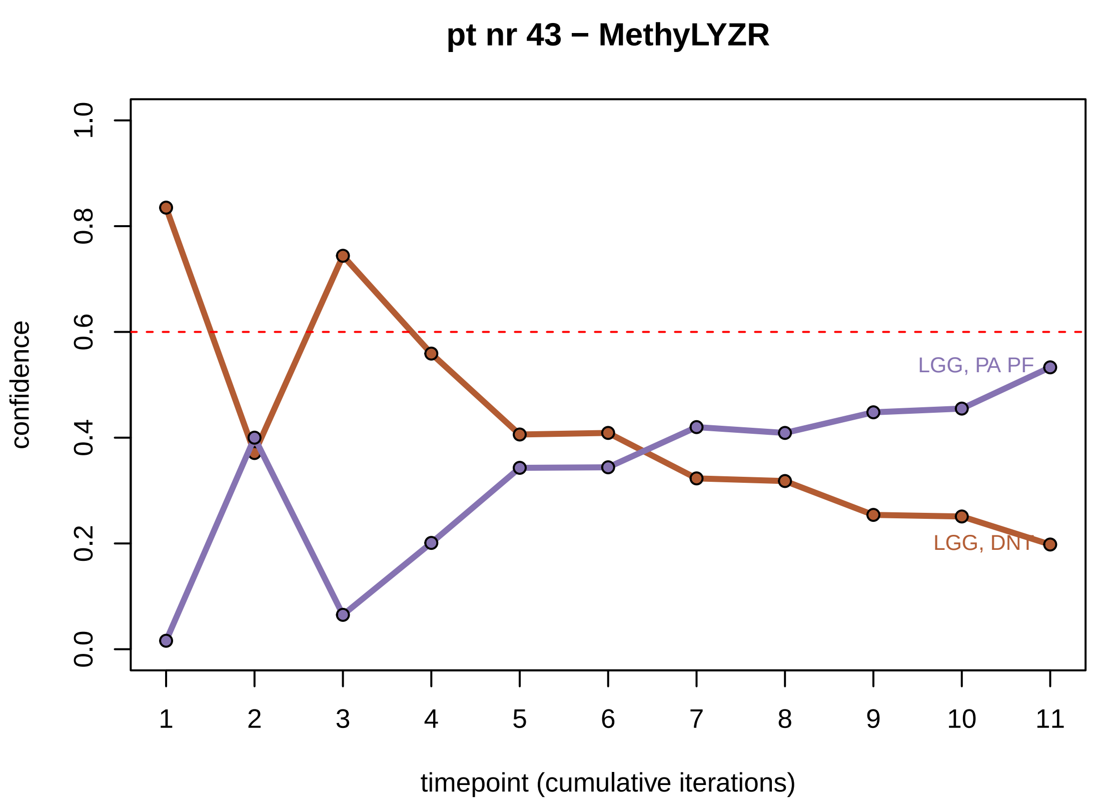
pt nr 43 − MethyLYZR
LGG, PA PF
LGG, DNT
confidence
0.0
0.2
0.4
0.6
0.8
1.0
1 2 3 4 5 6 7 8 9 10 11
timepoint (cumulative iterations)

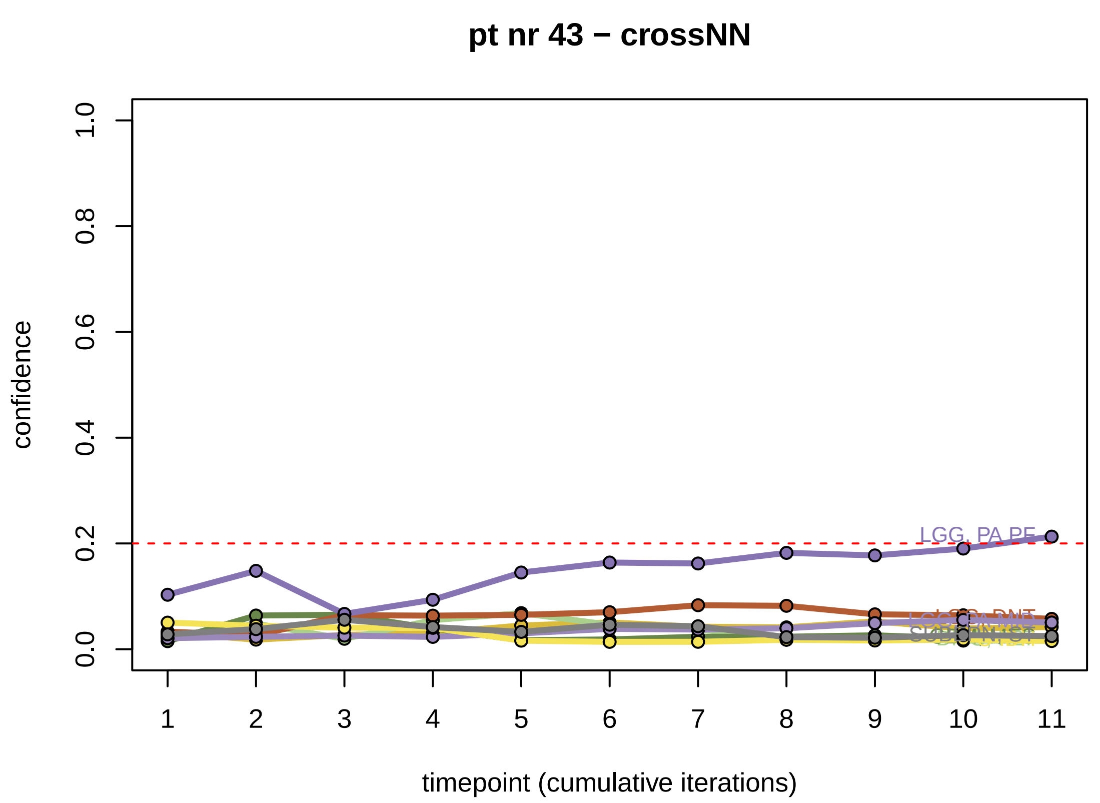
pt nr 43 − crossNN
LGG, PA PF
confidence
0.0
0.2
0.4
0.6
0.8
1.0
1 2 3 4 5 6 7 8 9 10 11
timepoint (cumulative iterations)

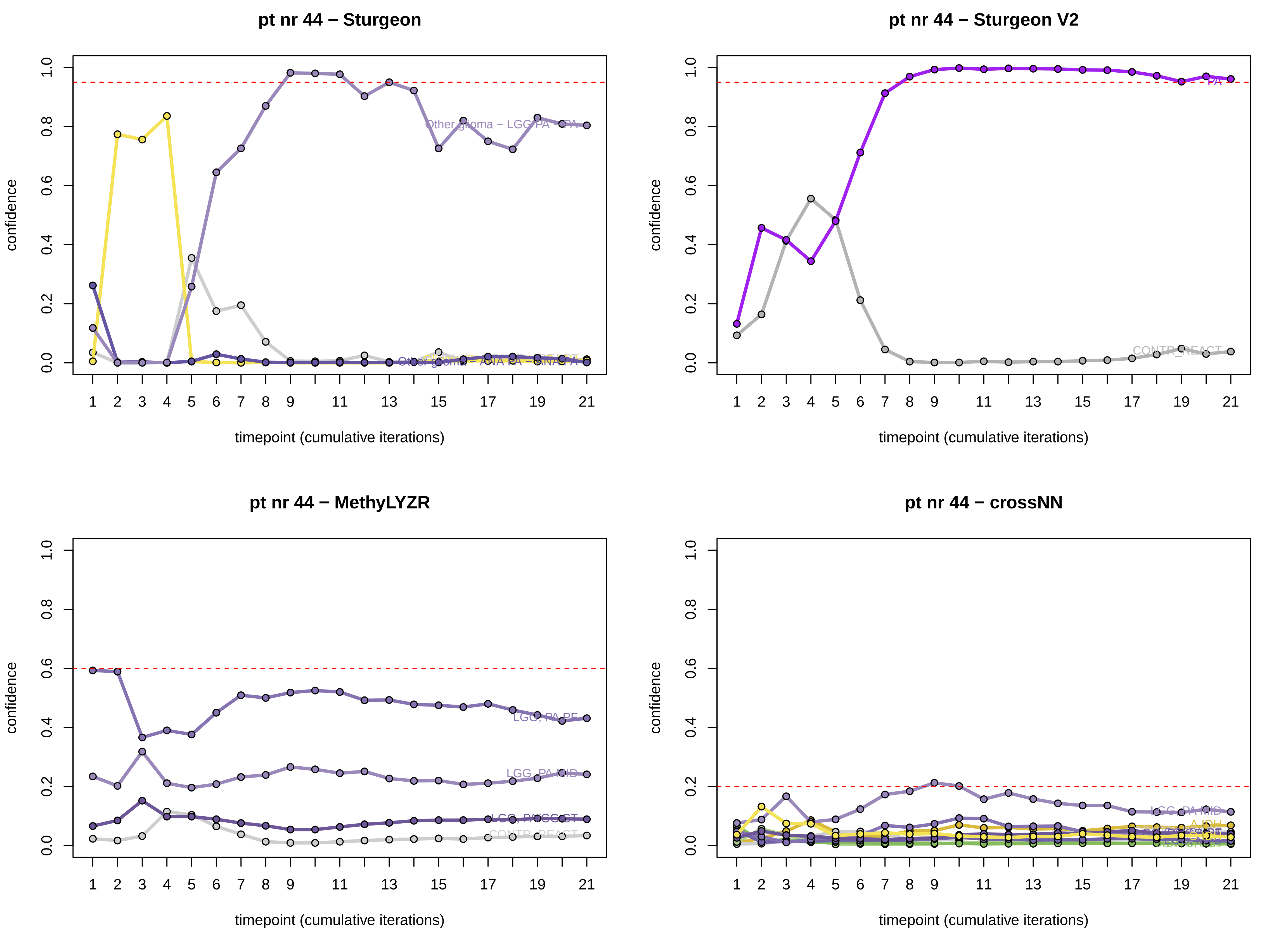
pt nr 44 − Sturgeon
Other glioma − LGG PA − PA
Other glioma − ANA PA − ANA PA
pt nr 44 − Sturgeon V2
PA
CONTR_REACT
pt nr 44 − MethyLYZR
LGG, PA PF
LGG, PA MID
pt nr 44 − crossNN
LGG_PA_MID
A_IDH
confidence
timepoint (cumulative iterations)

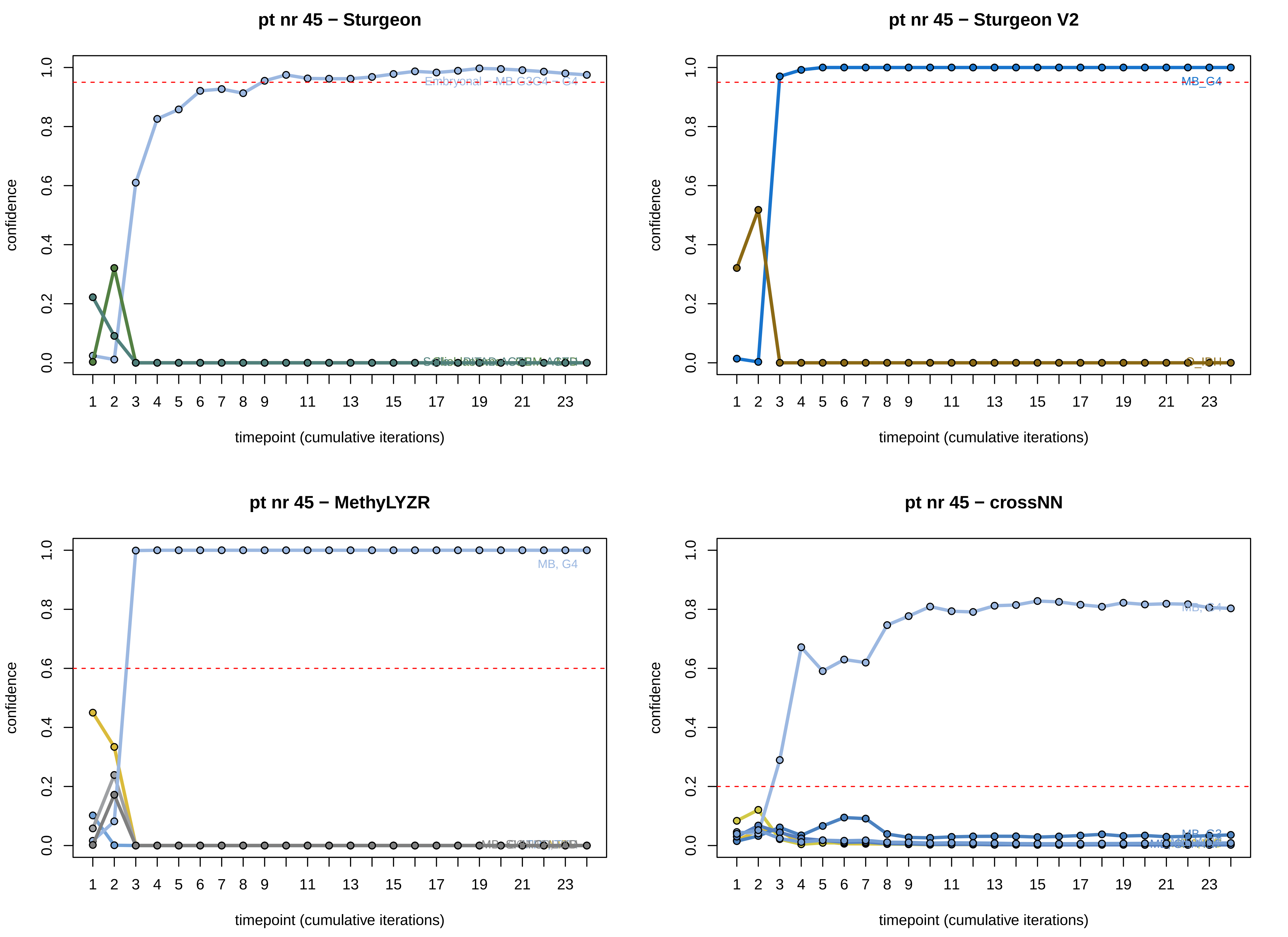

pt nr 45 − Sturgeon
Embryonal - MB G3G4 - G4
pt nr 45 − Sturgeon V2
MB_G4
O_IDH
pt nr 45 − MethyLYZR
MB, G4
pt nr 45 − crossNN
MB, G4
confidence
timepoint (cumulative iterations)

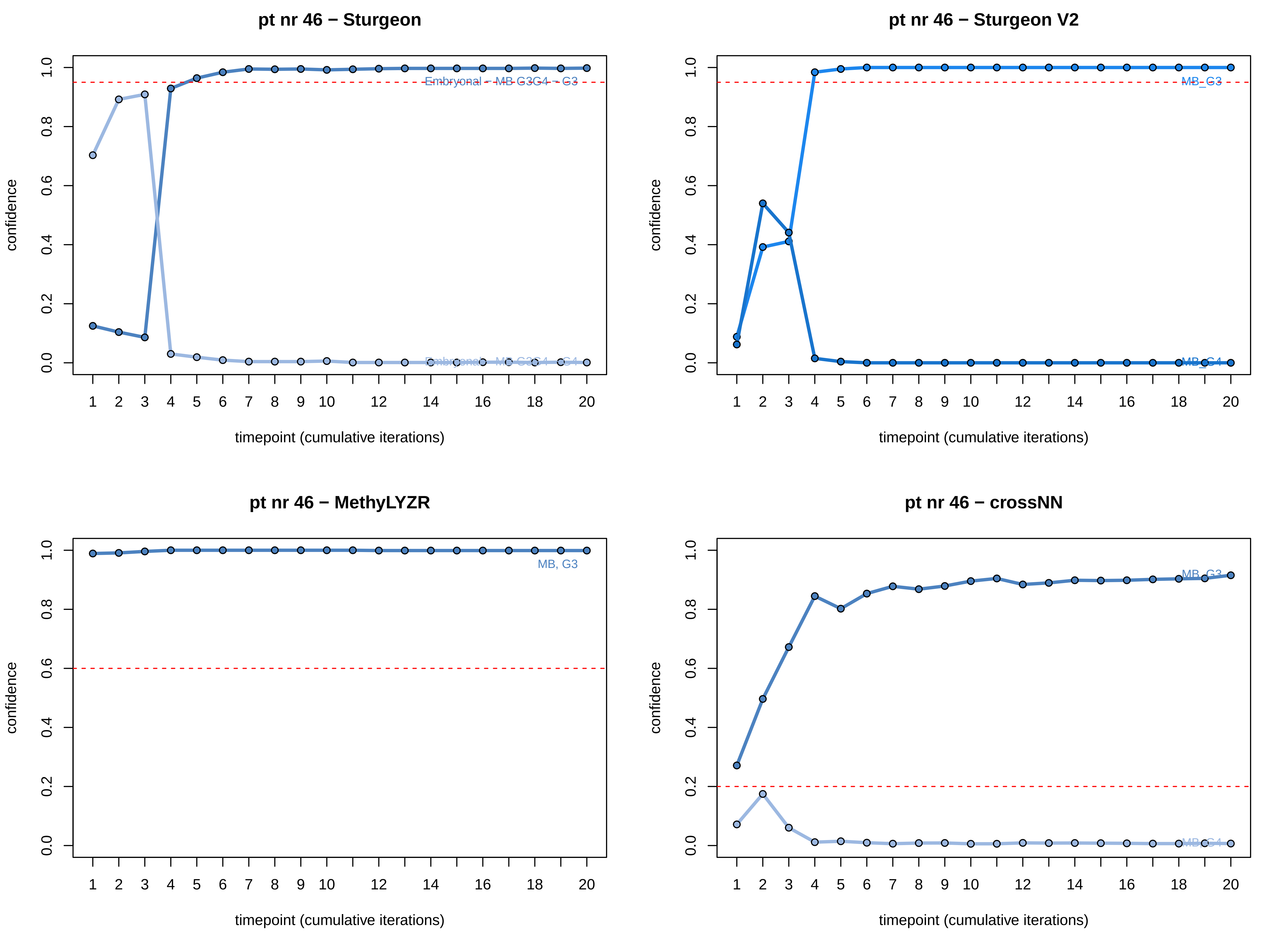

pt nr 46 − Sturgeon
Embryonal − MB G3G4 − G3
Embryonal − MB G3G4 − G4
pt nr 46 − Sturgeon V2
MB_G3
MB_G4
pt nr 46 − MethyLYZR
MB, G3
pt nr 46 − crossNN
MB, G3
MB, G4
confidence
timepoint (cumulative iterations)
0.0
0.2
0.4
0.6
0.8
1.0
1
2
3
4
5
6
7
8
9
10
12
14
16
18
20

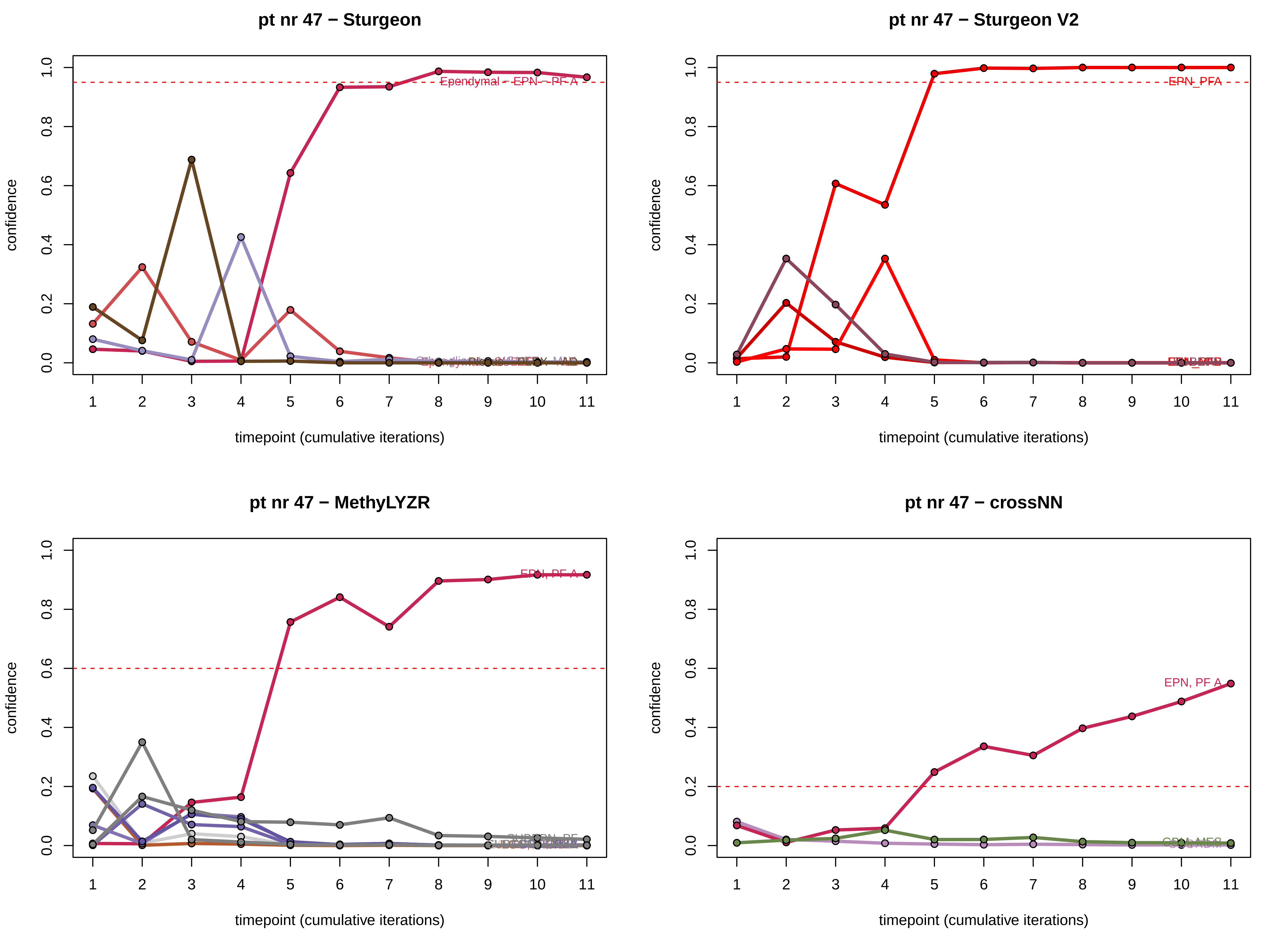
pt nr 47 − Sturgeon
Ependymal − EPN − PF A
pt nr 47 − Sturgeon V2
EPN_PFA
pt nr 47 − MethyLYZR
EPN, PF A
pt nr 47 − crossNN
EPN, PF A
confidence
timepoint (cumulative iterations)
0.0
0.2
0.4
0.6
0.8
1.0
1
2
3
4
5
6
7
8
9
10
11

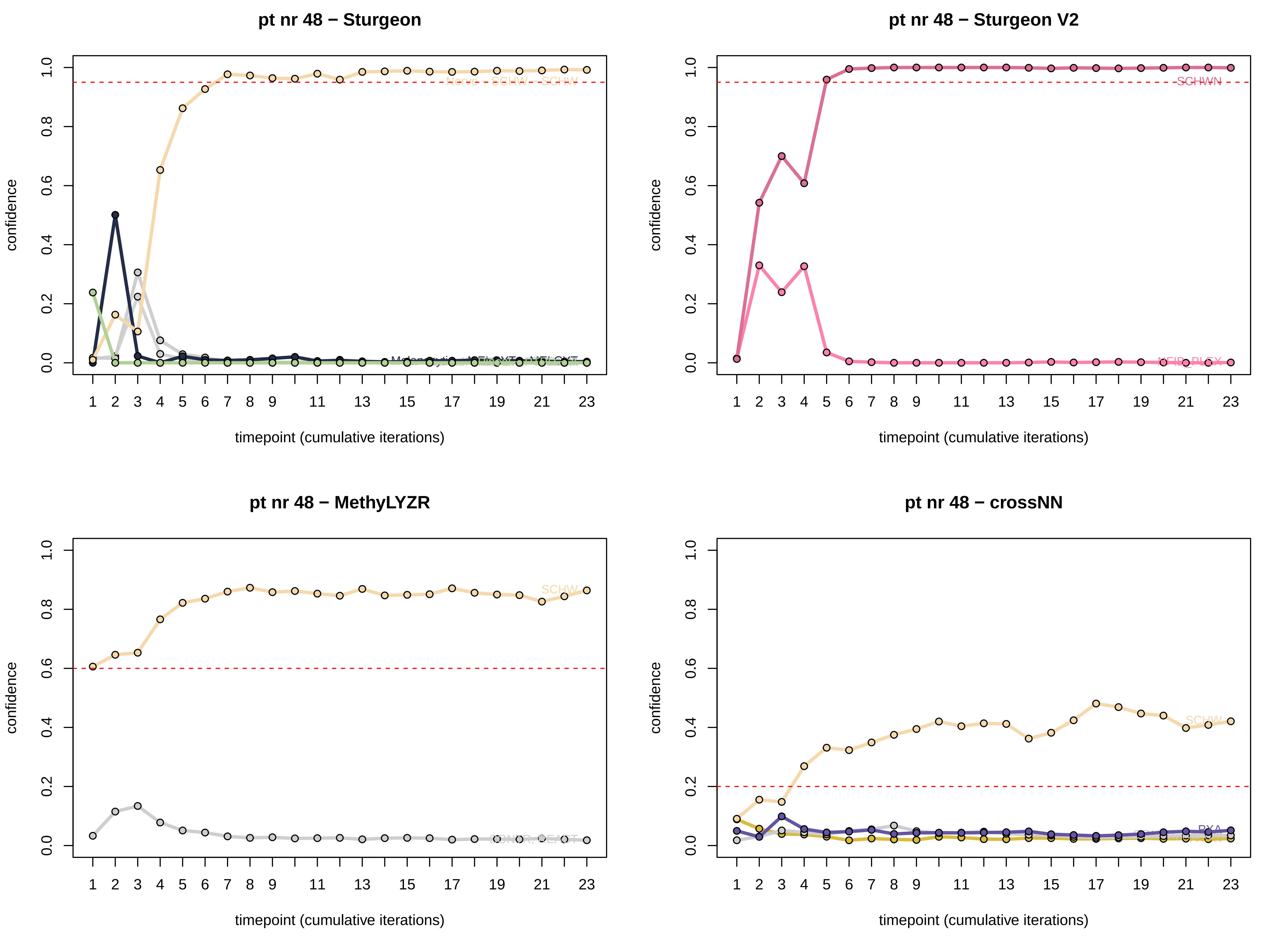

pt nr 48 − Sturgeon
pt nr 48 − Sturgeon V2
pt nr 48 − MethyLYZR
pt nr 48 − crossNN
confidence
timepoint (cumulative iterations)
SCHWN
SCHW
PXA

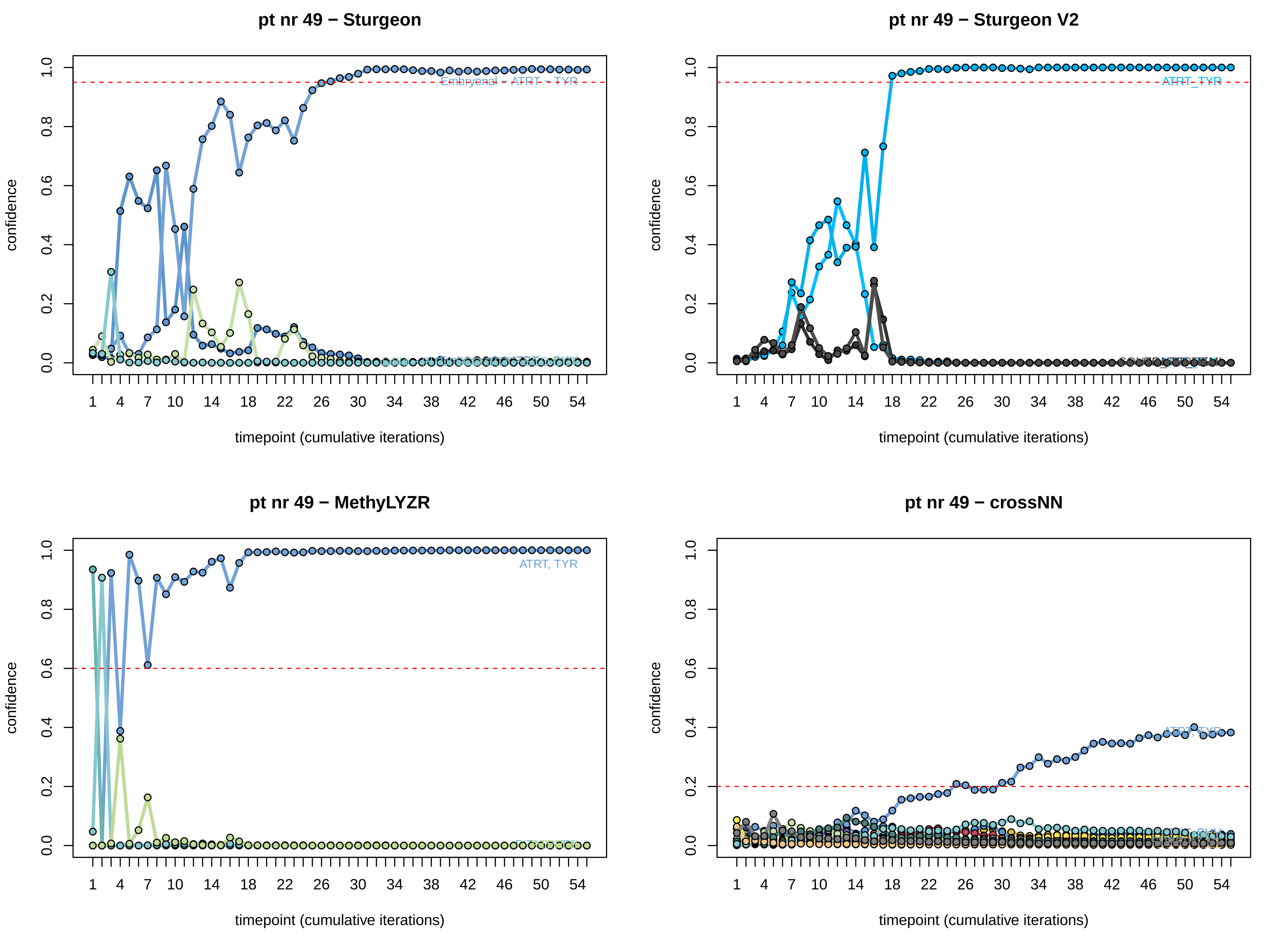
pt nr 49 − Sturgeon
Embryonal − ATRT − TYR
pt nr 49 − Sturgeon V2
ATRT_TYR
pt nr 49 − MethyLYZR
ATRT, TYR
pt nr 49 − crossNN
ATRT, TYR
confidence
timepoint (cumulative iterations)
0.0
0.2
0.4
0.6
0.8
1.0
1 4 7 10 14 18 22 26 30 34 38 42 46 50 54

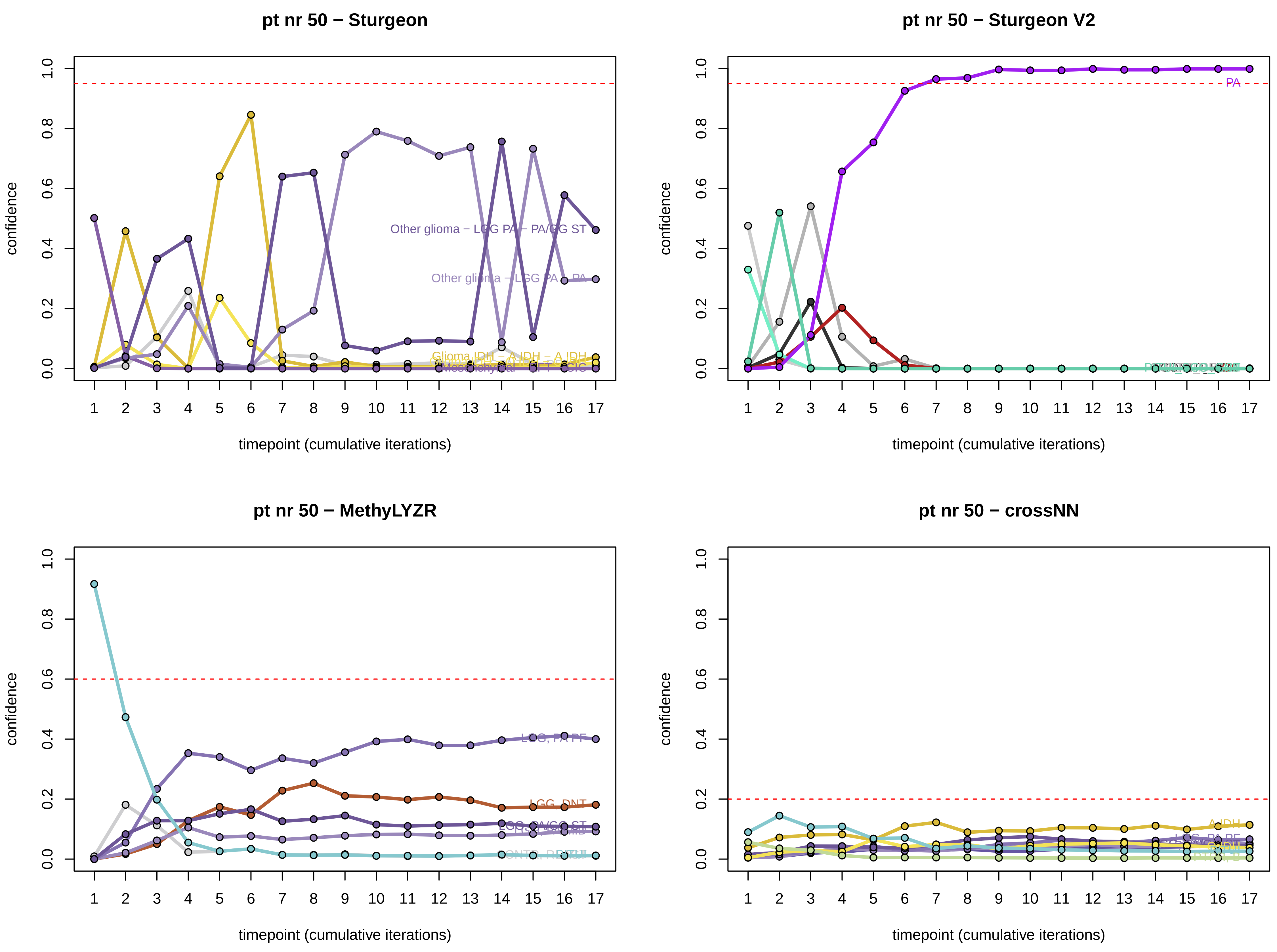
pt nr 50 − Sturgeon
pt nr 50 − Sturgeon V2
pt nr 50 − MethyLYZR
pt nr 50 − crossNN
confidence
timepoint (cumulative iterations)
Other glioma − LGG PA − PA/GG ST
Other glioma − LGG PA − PA
Glioma IDH − A IDH − A IDH
PA
LGG, PA PF
LGG, DNT
LGG, PA/GG ST
A IDH

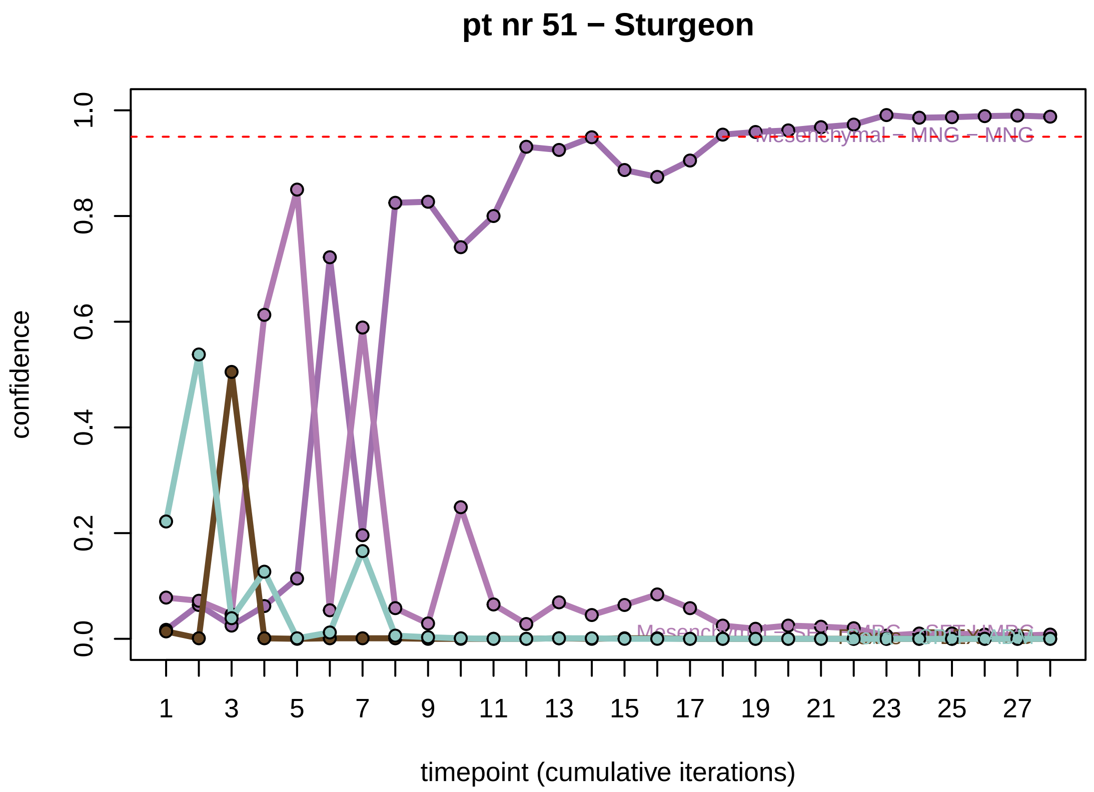
pt nr 51 − Sturgeon
confidence
timepoint (cumulative iterations)

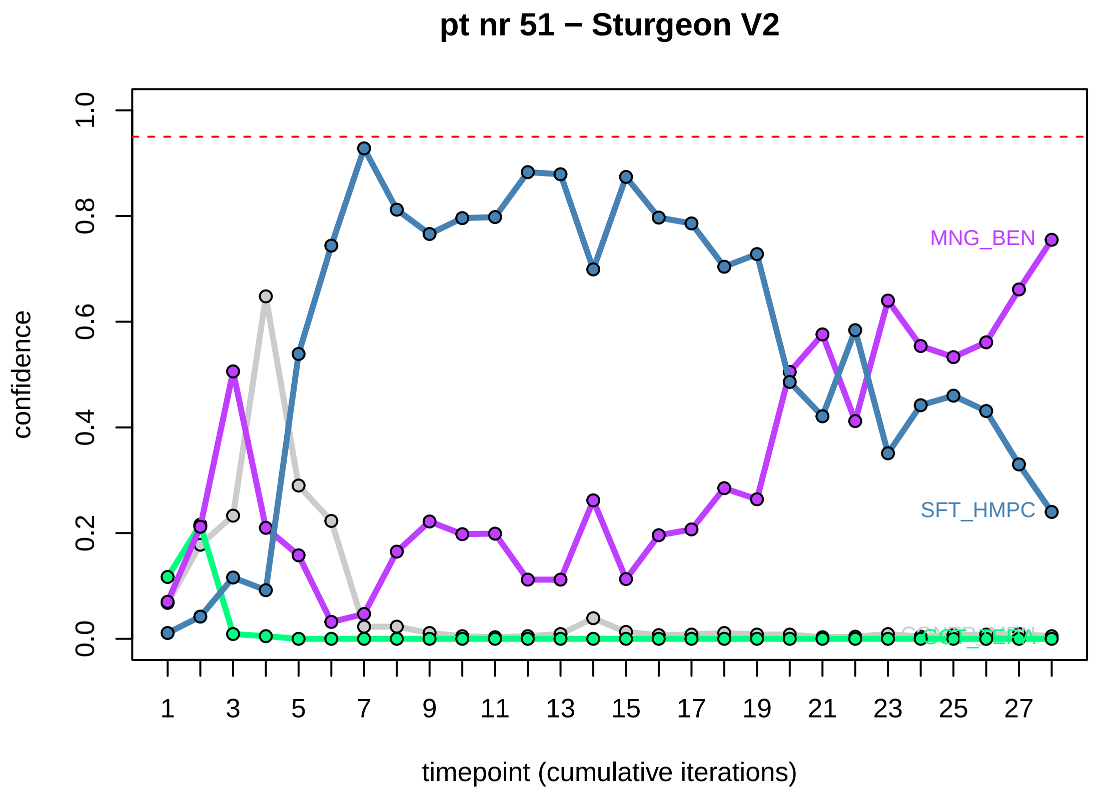
pt nr 51 − Sturgeon V2
confidence
timepoint (cumulative iterations)
MNG_BEN
SFT_HMPC

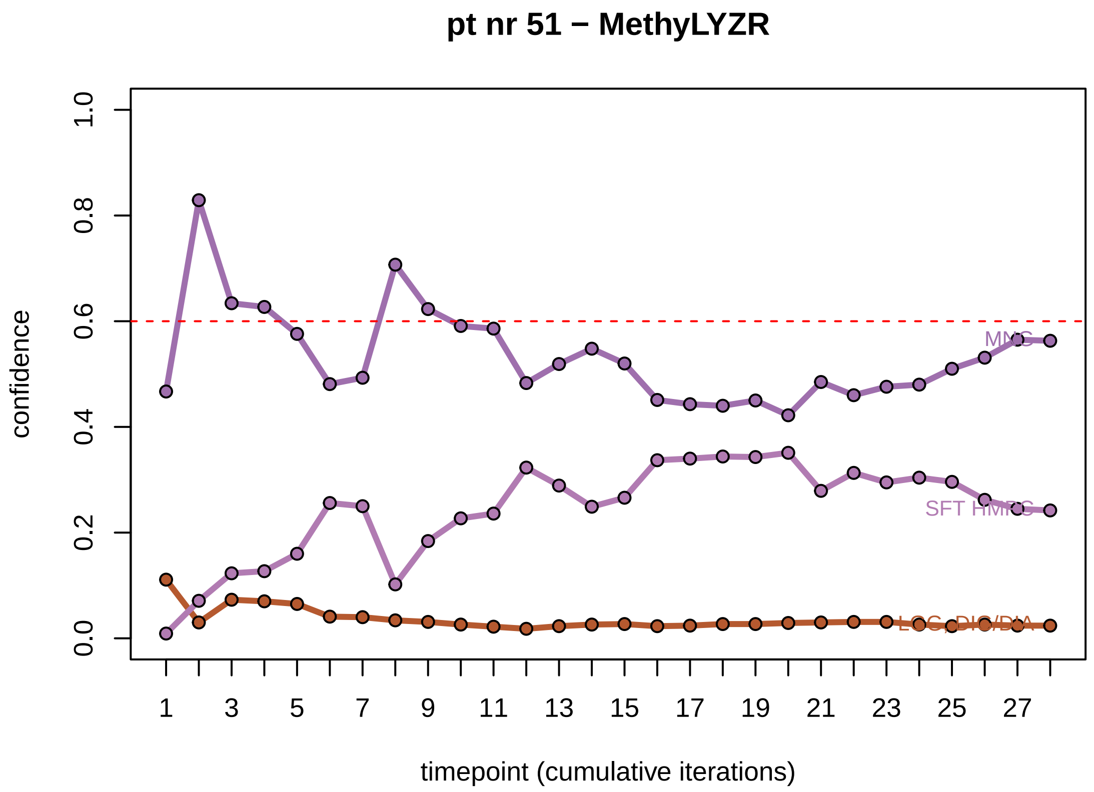
pt nr 51 − MethyLYZR
confidence
timepoint (cumulative iterations)

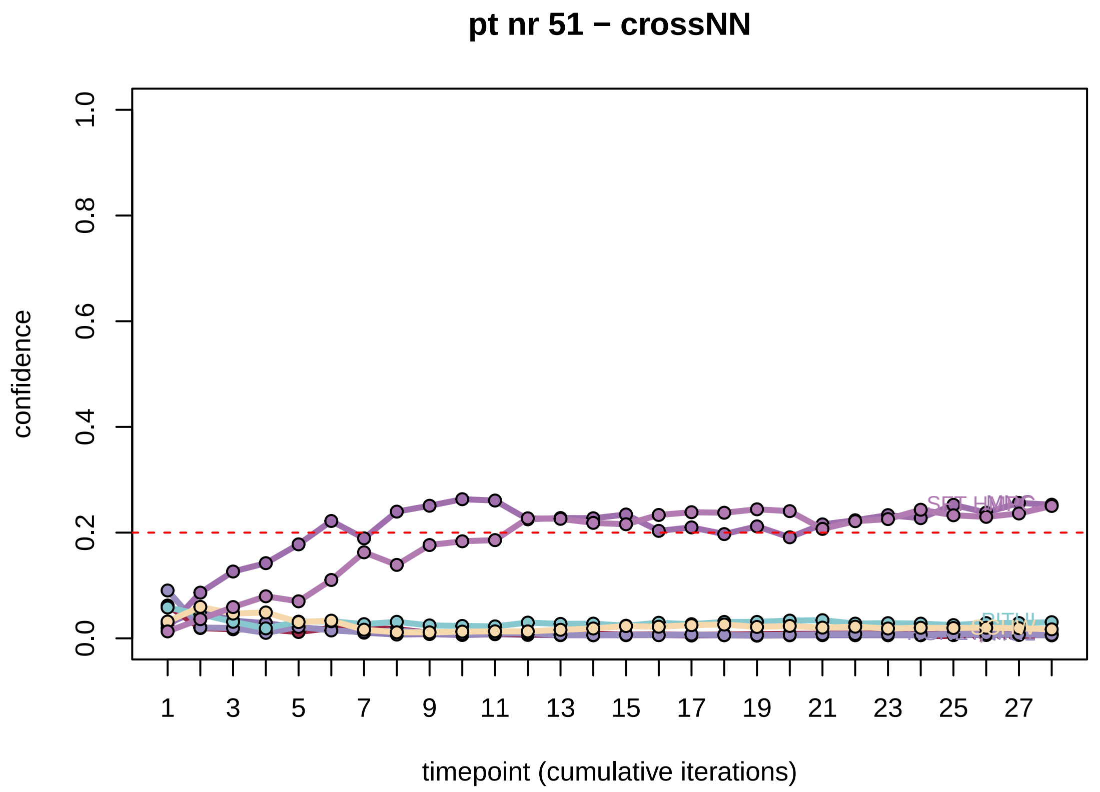
pt nr 51 − crossNN
confidence
timepoint (cumulative iterations)

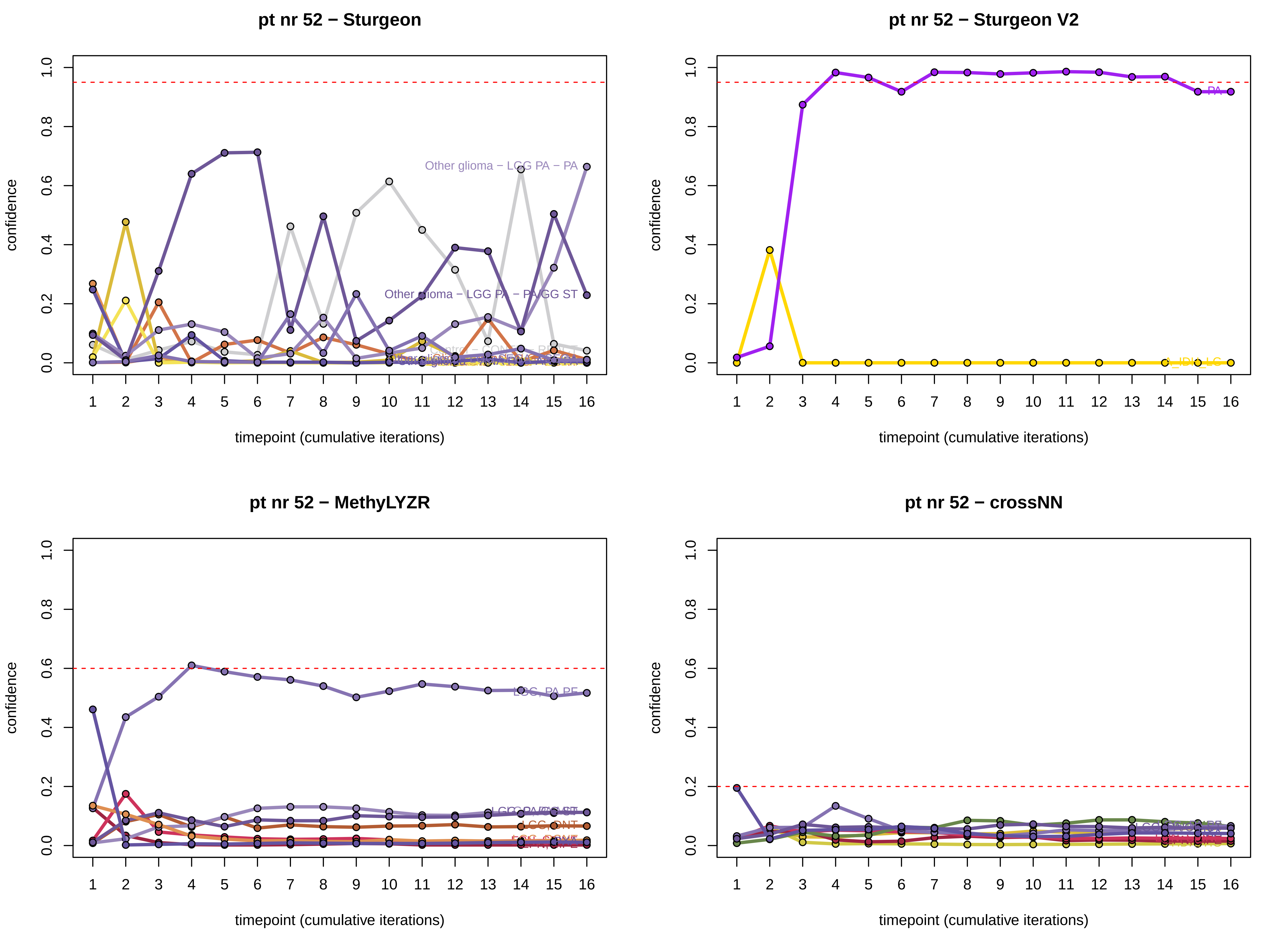

pt nr 52 − Sturgeon
pt nr 52 − Sturgeon V2
pt nr 52 − MethyLYZR
pt nr 52 − crossNN
confidence
timepoint (cumulative iterations)
Other glioma − LGG PA − PA
Other glioma − LGG PA − PA/GG ST
PA
A_IDH_LG
LGG, PA PF
LGG, DNT

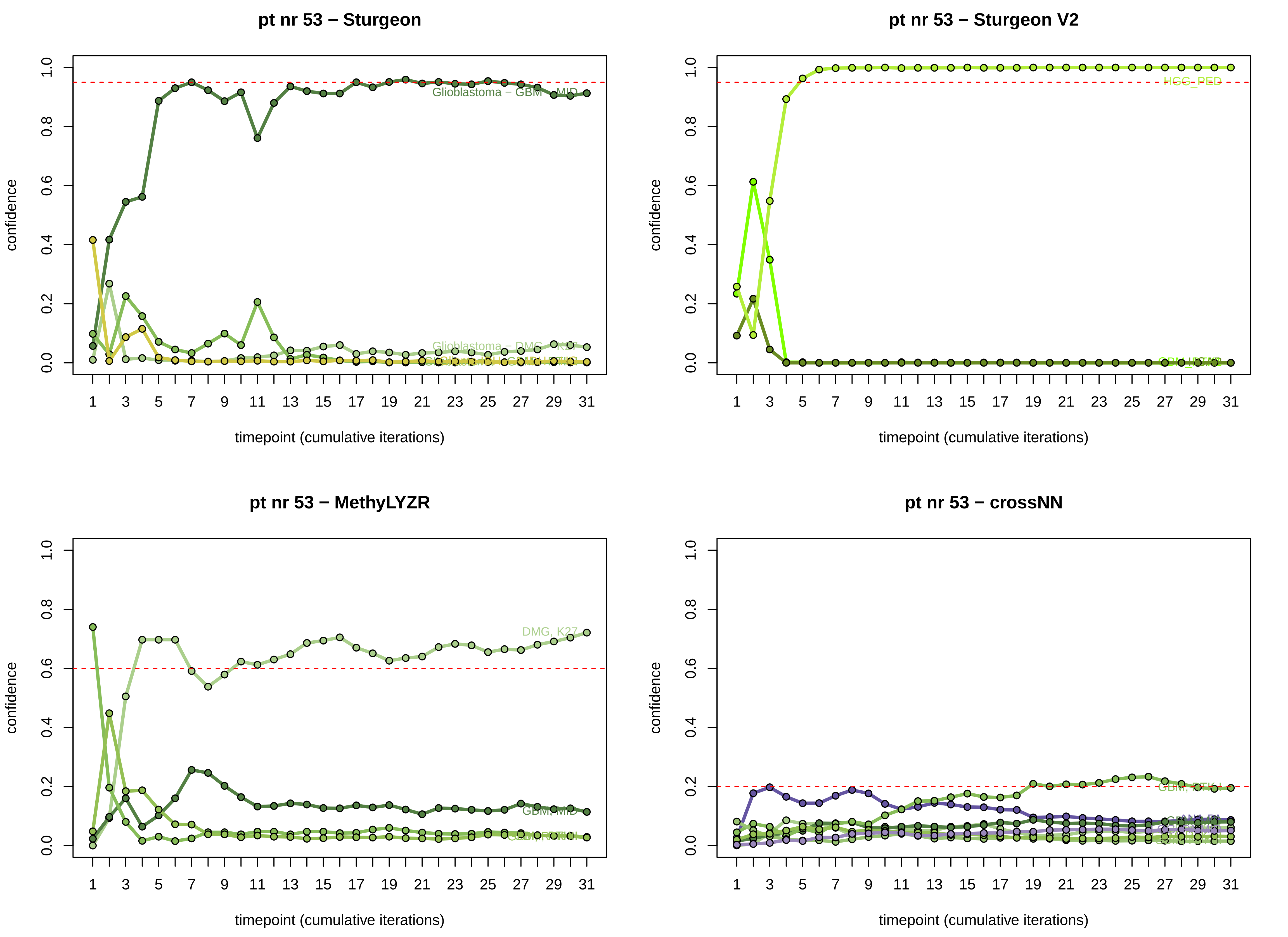

pt nr 53 − Sturgeon
pt nr 53 − Sturgeon V2
pt nr 53 − MethyLYZR
pt nr 53 − crossNN
confidence
timepoint (cumulative iterations)
Glioblastoma − GBM − MID
Glioblastoma − DMG − K27
HGG_PED
DMG, K27
GBM, MID
GBM, RTK I

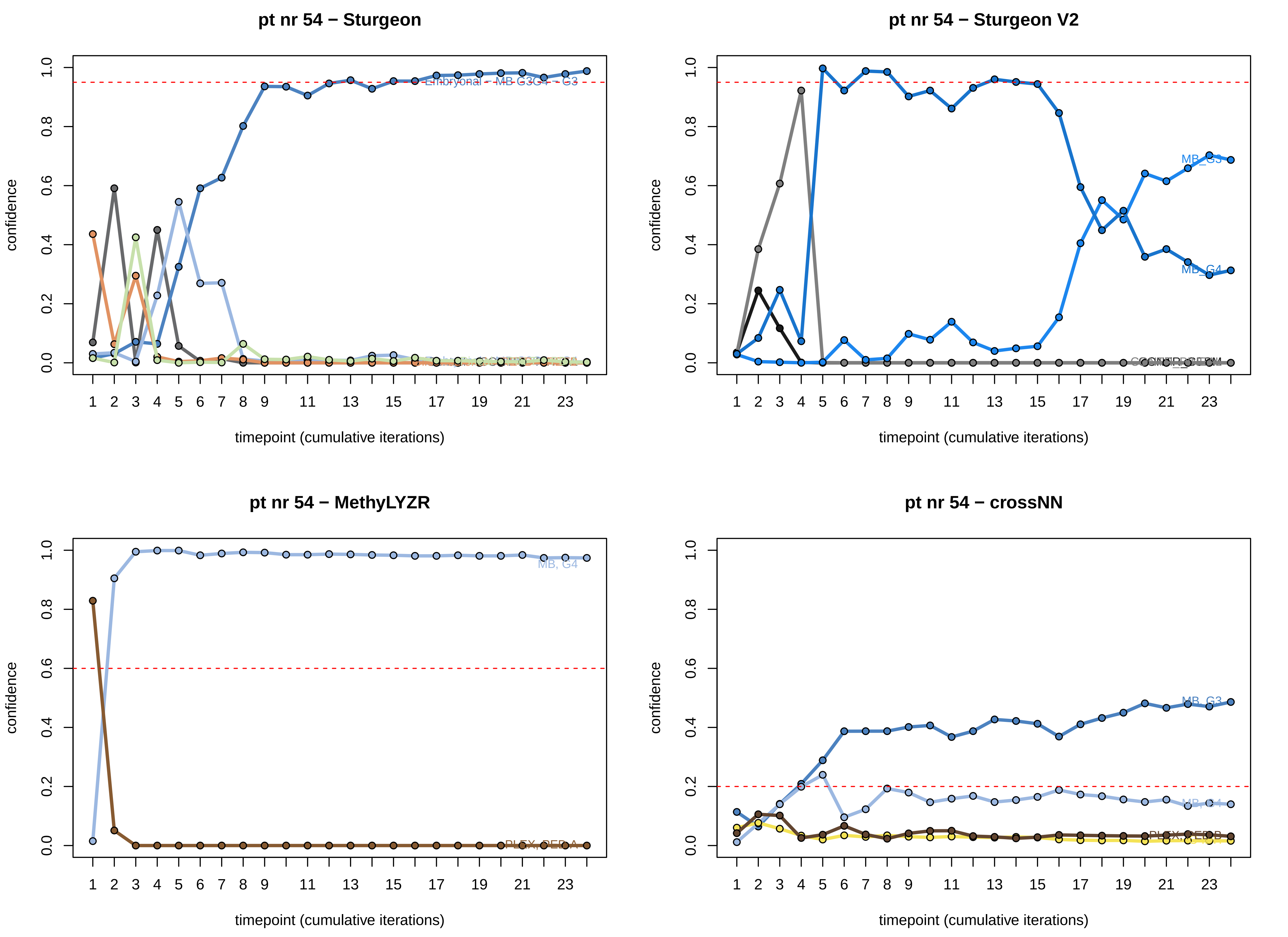

pt nr 54 − Sturgeon
Embryonal − MB G3G4 − G3
pt nr 54 − Sturgeon V2
MB_G3
MB_G4
pt nr 54 − MethyLYZR
MB, G4
PLEX, PED A
pt nr 54 − crossNN
MB, G3
MB, G4
PLEX, PED B
confidence
timepoint (cumulative iterations)

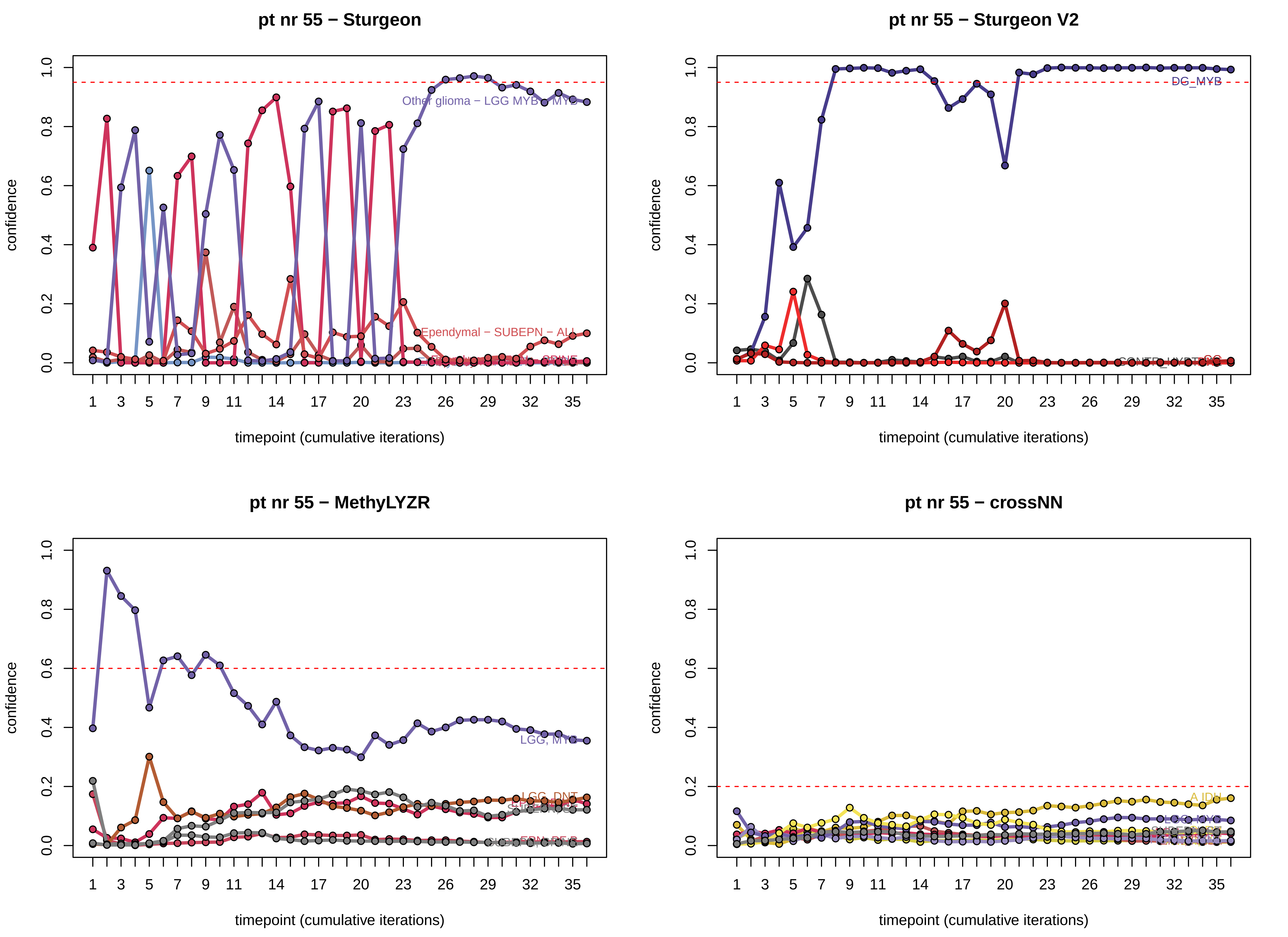
pt nr 55 − Sturgeon
Other glioma − LGG MYB − MYB
Ependymal − SUBEPN − ALL
pt nr 55 − Sturgeon V2
DG_MYB
pt nr 55 − MethyLYZR
LGG, MYB
LGG, DNT
pt nr 55 − crossNN
A IDH
LGG_MYB
confidence
timepoint (cumulative iterations)

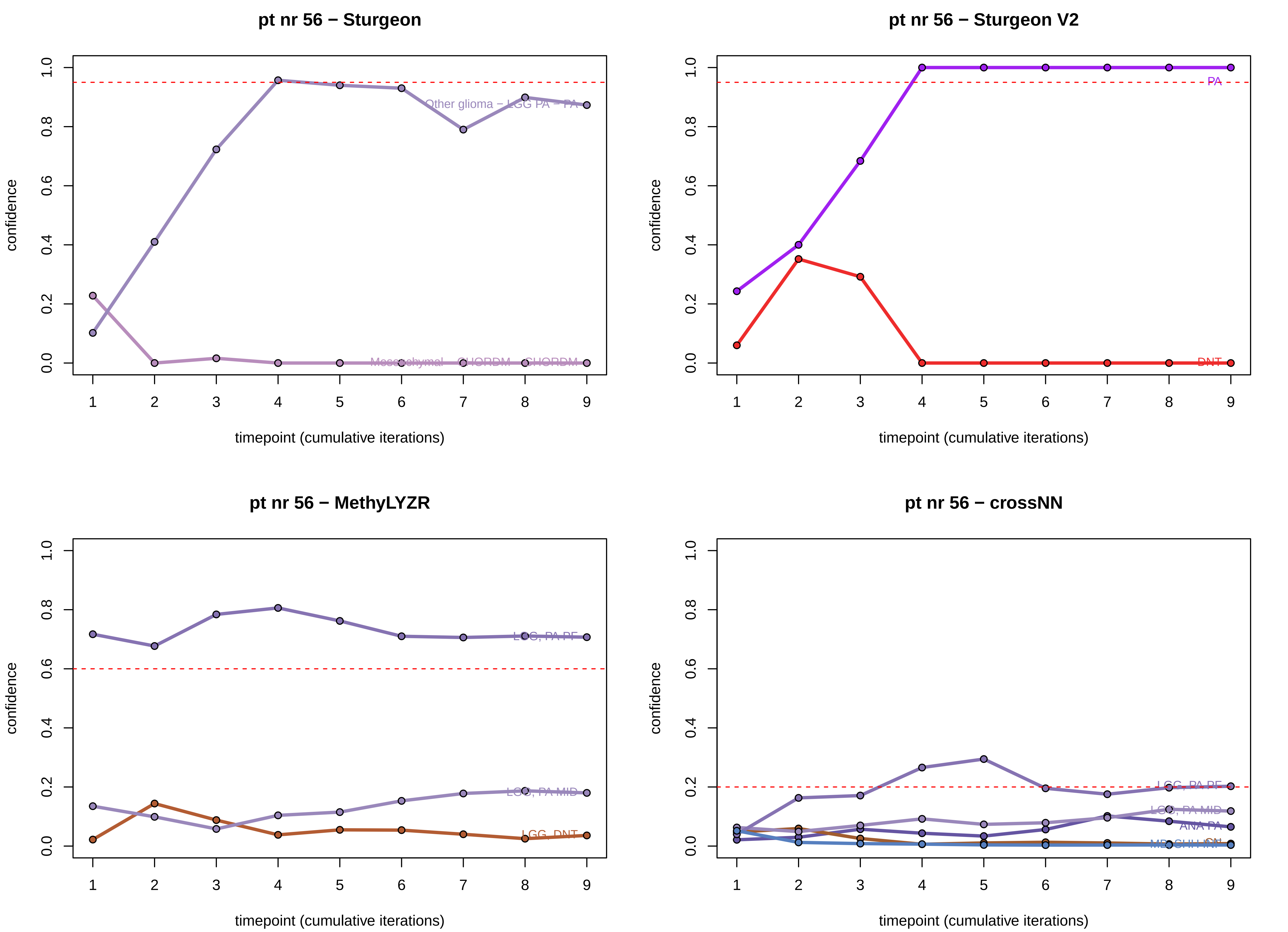

pt nr 56 − Sturgeon
Other glioma − LGG PA − PA
Mesenchymal − CHORDM − CHORDM
pt nr 56 − Sturgeon V2
PA
DNT
pt nr 56 − MethyLYZR
LGG, PA PF
LGG, PA MID
LGG, DNT
pt nr 56 − crossNN
LGG, PA PF
LGG, PA MID
ANA PA
confidence
timepoint (cumulative iterations)

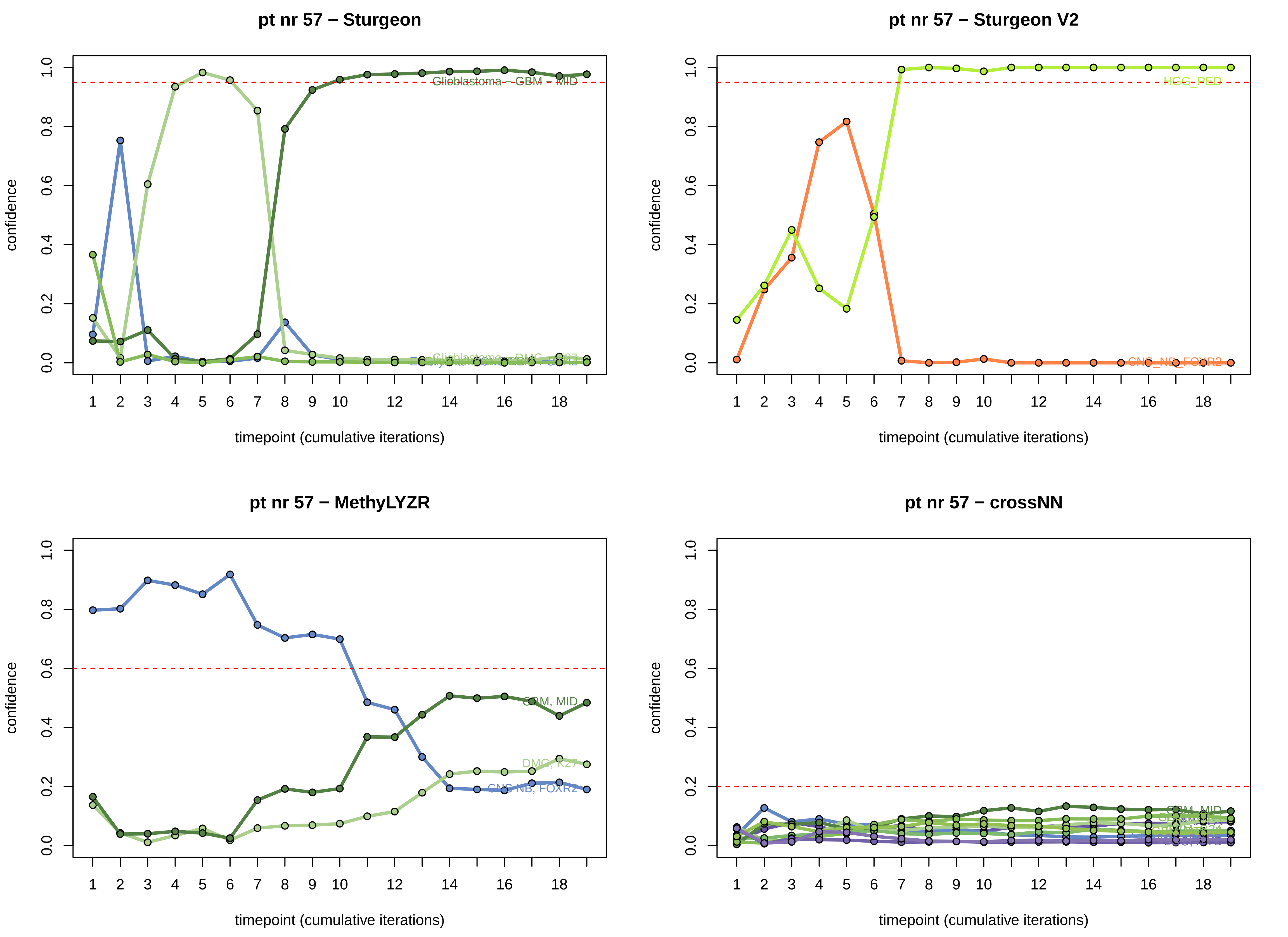
pt nr 57 − Sturgeon
pt nr 57 − Sturgeon V2
pt nr 57 − MethyLYZR
pt nr 57 − crossNN
confidence
timepoint (cumulative iterations)
0.0
0.2
0.4
0.6
0.8
1.0
1
2
3
4
5
6
7
8
9
10
12
14
16
18
Glioblastoma − GBM − MID
HGG_PED
CNS_NB_FOXR2
GBM, MID
DMG, K27
CNS NB, FOXR2
GBM_MID

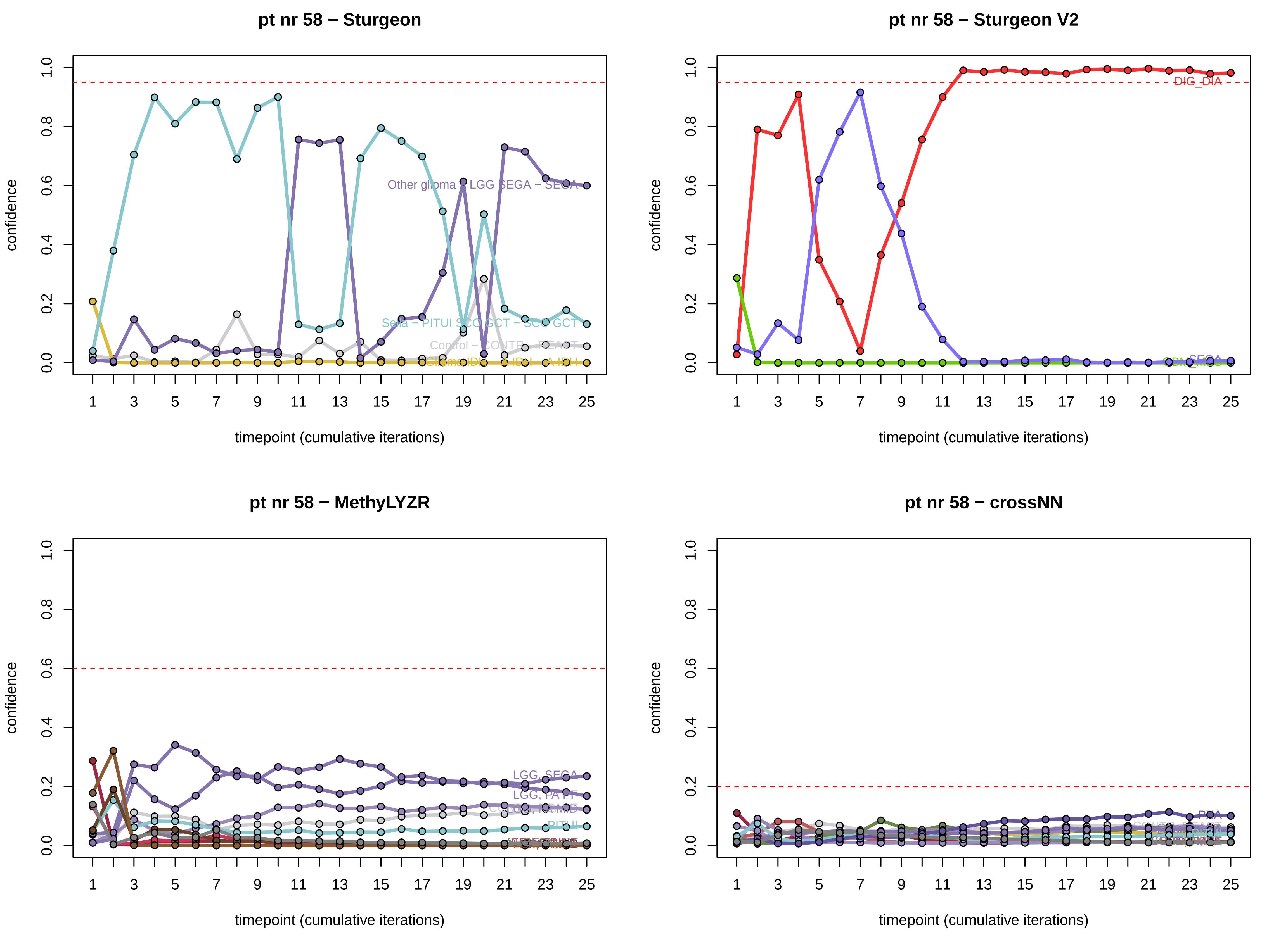

pt nr 58 − Sturgeon
confidence
timepoint (cumulative iterations)
Other glioma − LGG SEGA − SEGA
Sella − PITUI SCO GCT − SCO GCT
Control − CONTR − REACT
pt nr 58 − Sturgeon V2
confidence
timepoint (cumulative iterations)
DIG_DIA
pt nr 58 − MethyLYZR
confidence
timepoint (cumulative iterations)
LGG, SEGA
LGG, PA PT
PITUI
pt nr 58 − crossNN
confidence
timepoint (cumulative iterations)

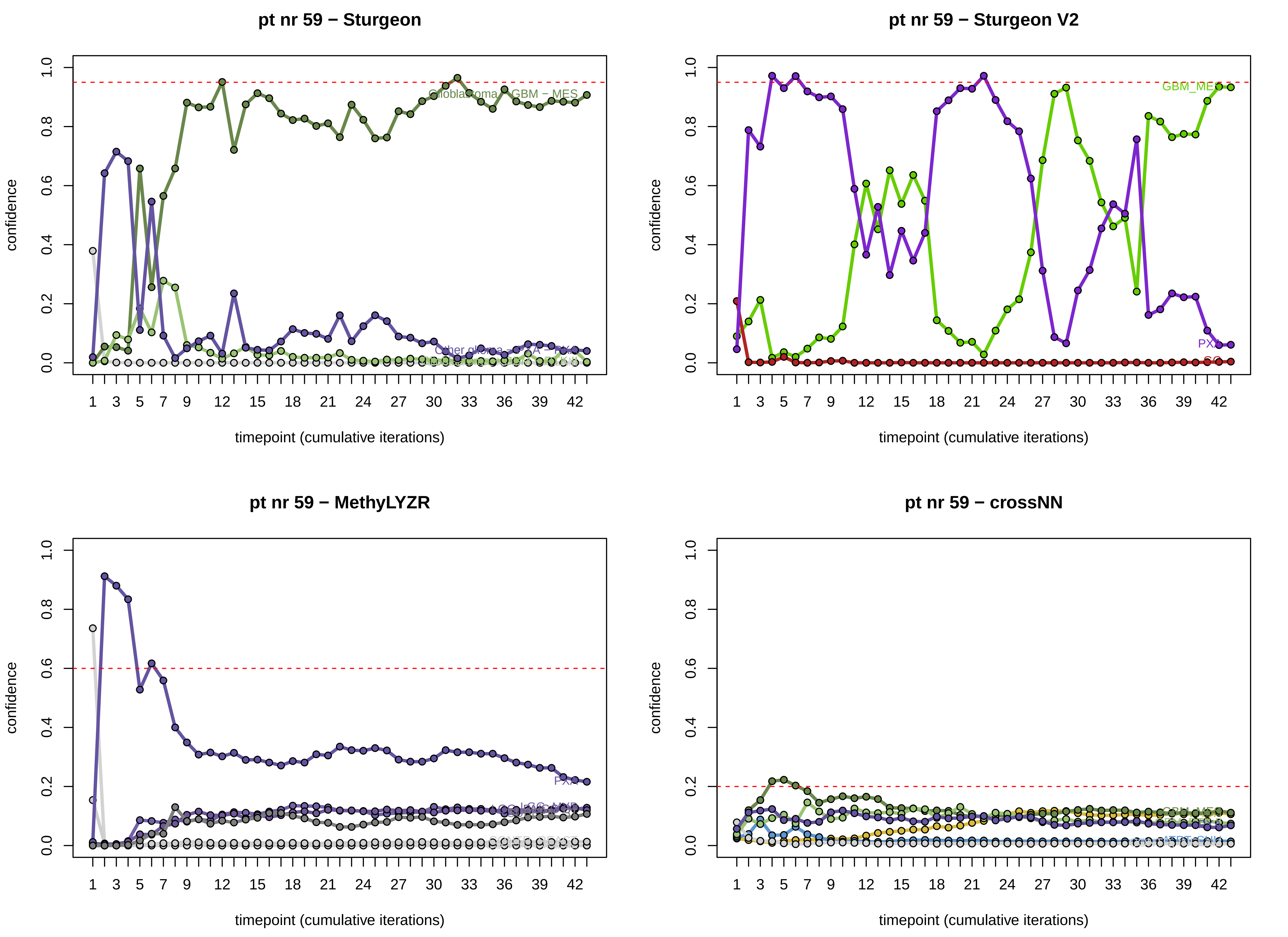

pt nr 59 − Sturgeon
Glioblastoma − GBM − MES
Other glioma − PXA − PXA
pt nr 59 − Sturgeon V2
GBM_MES
PXA
SC
pt nr 59 − MethyLYZR
PXA
pt nr 59 − crossNN
GBM_MES
confidence
timepoint (cumulative iterations)

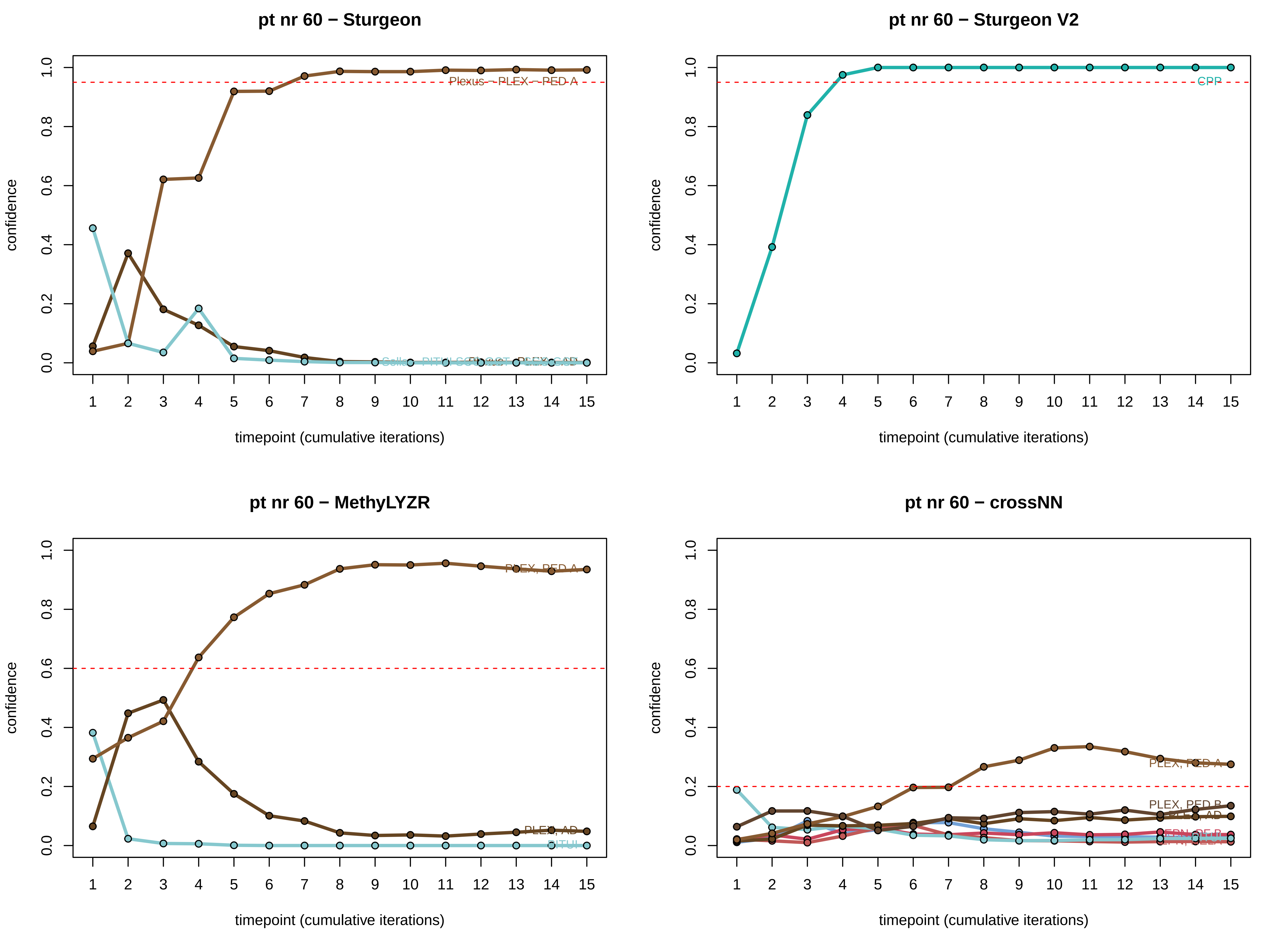

pt nr 60 − Sturgeon
pt nr 60 − Sturgeon V2
pt nr 60 − MethyLYZR
pt nr 60 − crossNN
confidence
timepoint (cumulative iterations)
CPP
PLEX, PED A
PLEX, AD
PITUI
PLEX, PED B

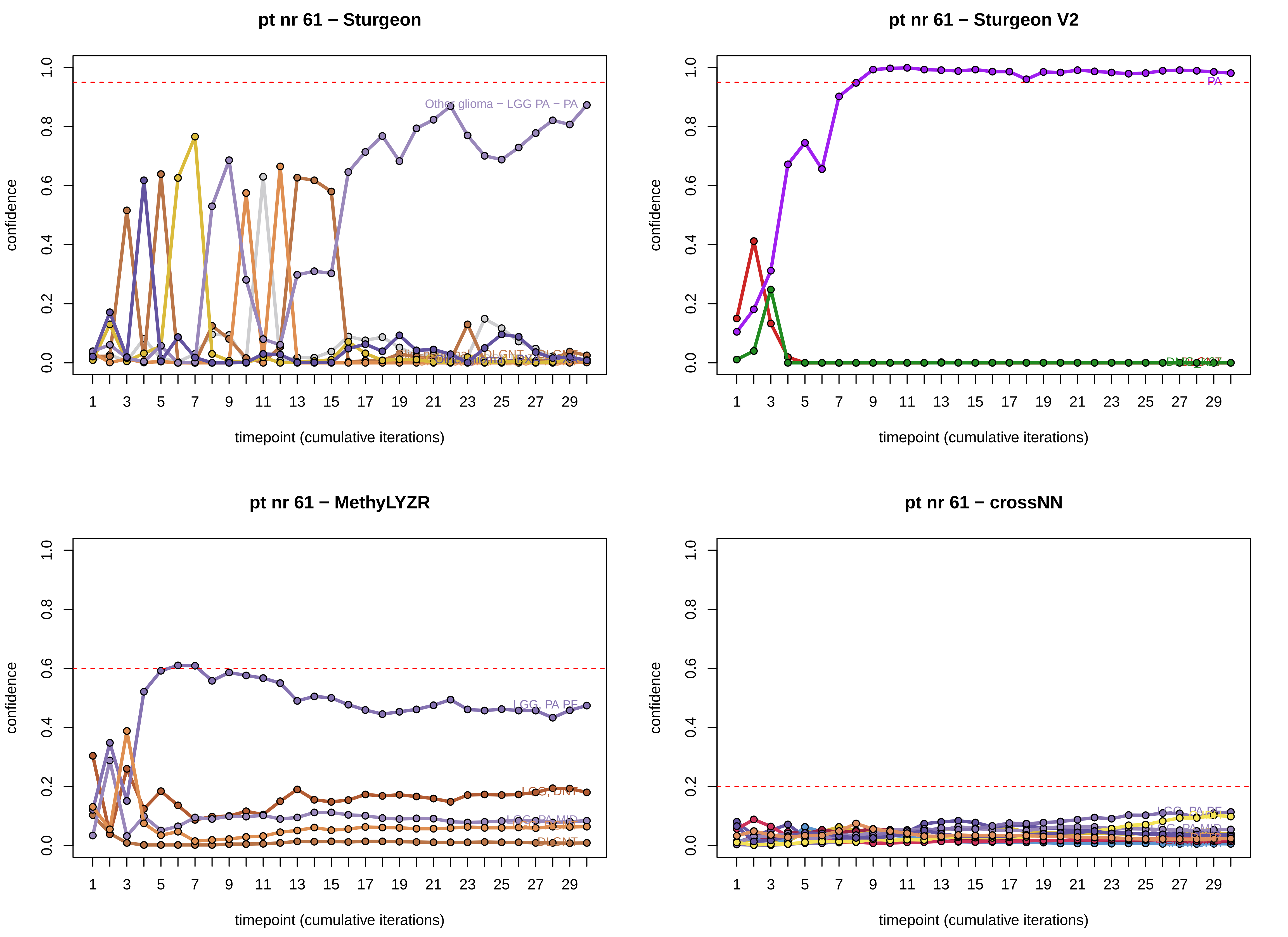

pt nr 61 − Sturgeon
Other glioma − LGG PA − PA
pt nr 61 − Sturgeon V2
PA
pt nr 61 − MethyLYZR
LGG, PA PF
LGG, DNT
LGG, PA MID
DLGNT
pt nr 61 − crossNN
LGG, PA PF
LGG, PA MID
confidence
timepoint (cumulative iterations)

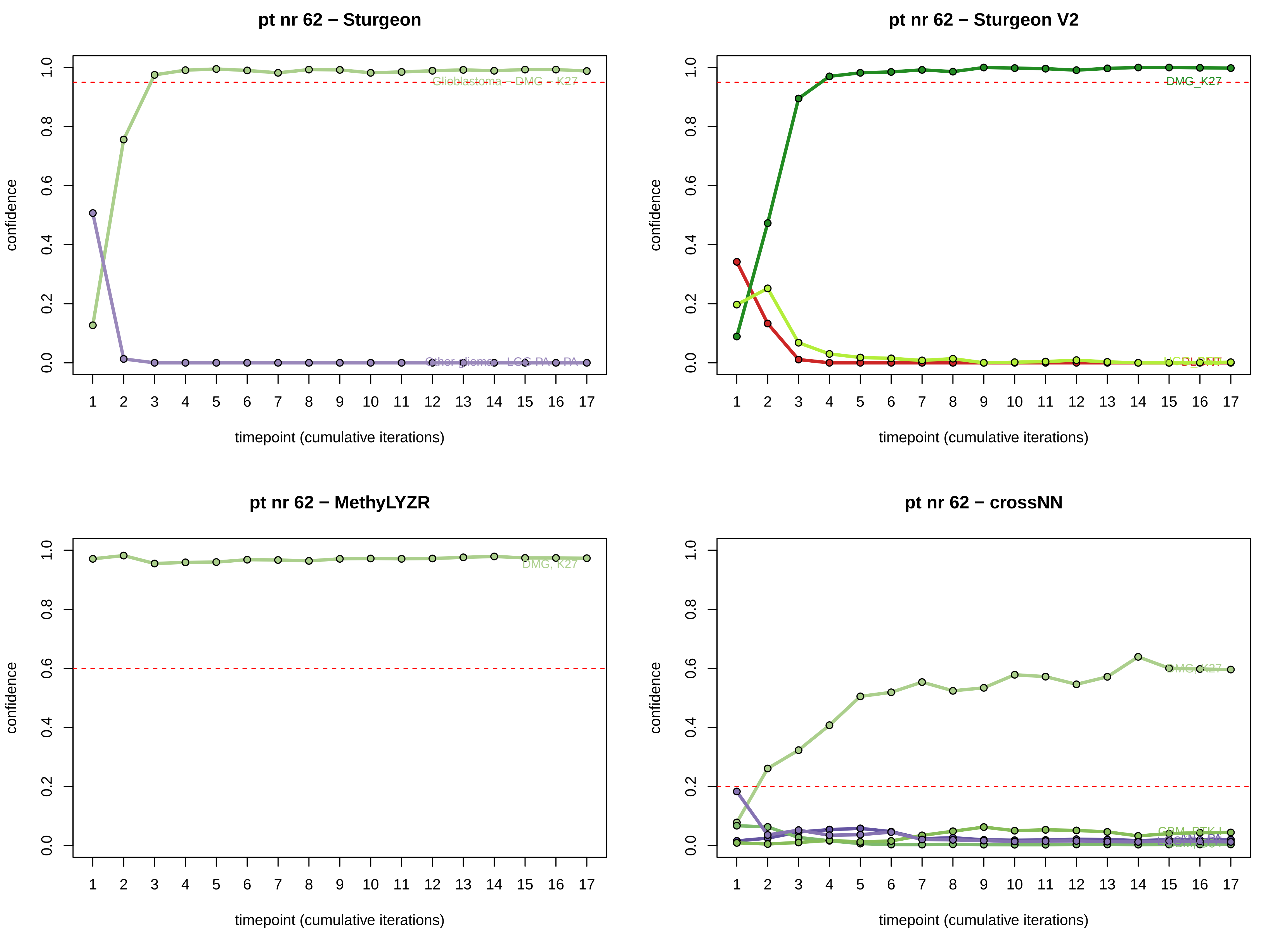

pt nr 62 − Sturgeon
confidence
timepoint (cumulative iterations)
Glioblastoma – DMG – K27
Other glioma – LGG-PA – PA
pt nr 62 − Sturgeon V2
confidence
timepoint (cumulative iterations)
DMG_K27
pt nr 62 − MethyLYZR
confidence
timepoint (cumulative iterations)
DMG, K27
pt nr 62 − crossNN
confidence
timepoint (cumulative iterations)
DMG_K27

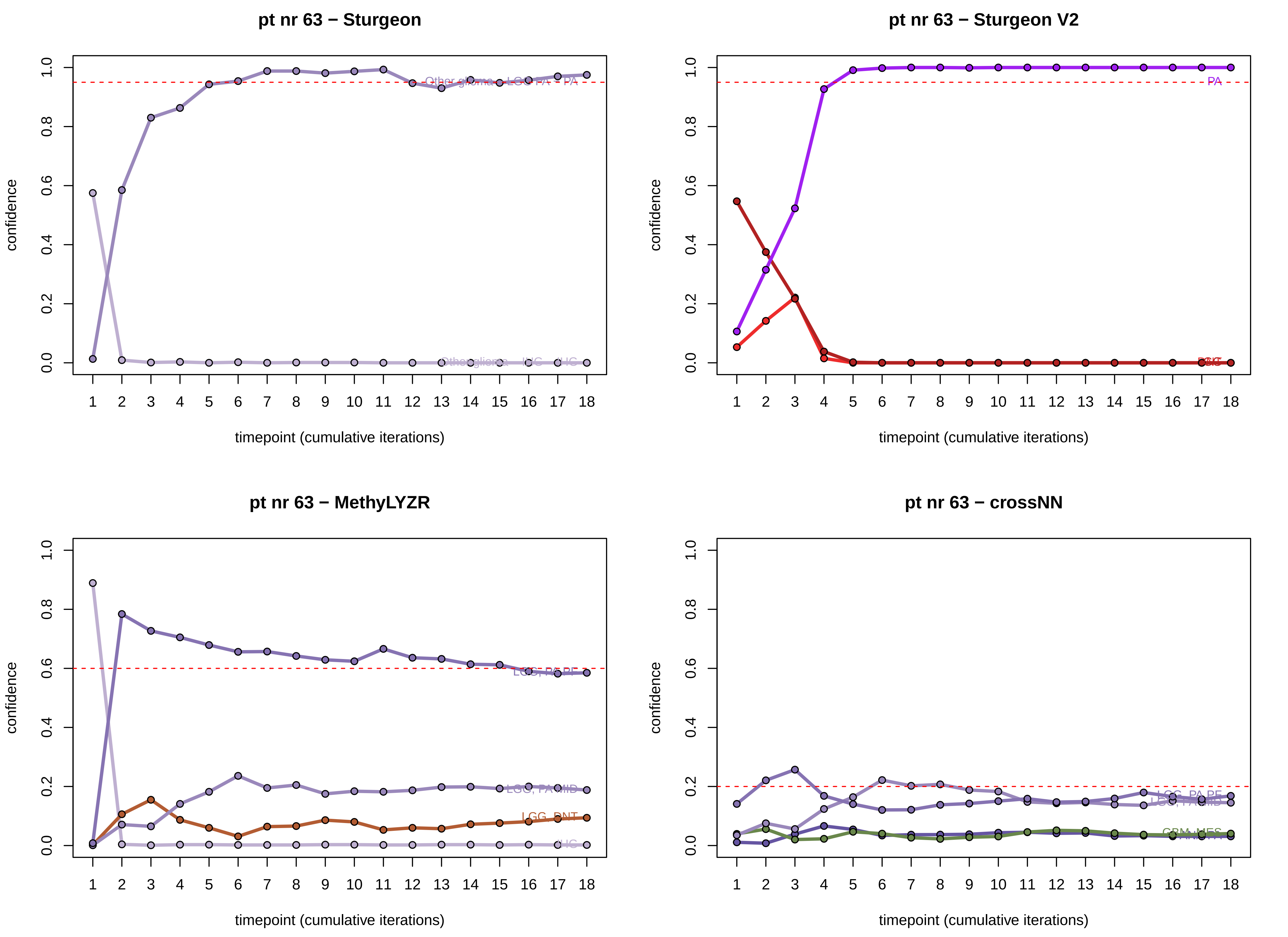
pt nr 63 − Sturgeon
pt nr 63 − Sturgeon V2
pt nr 63 − MethyLYZR
pt nr 63 − crossNN
confidence
timepoint (cumulative iterations)
0.0
0.2
0.4
0.6
0.8
1.0
1 2 3 4 5 6 7 8 9 10 11 12 13 14 15 16 17 18
PA
LGG, PA PF
LGG, PA MID
LGG, DNT
IHG
GBM, MES

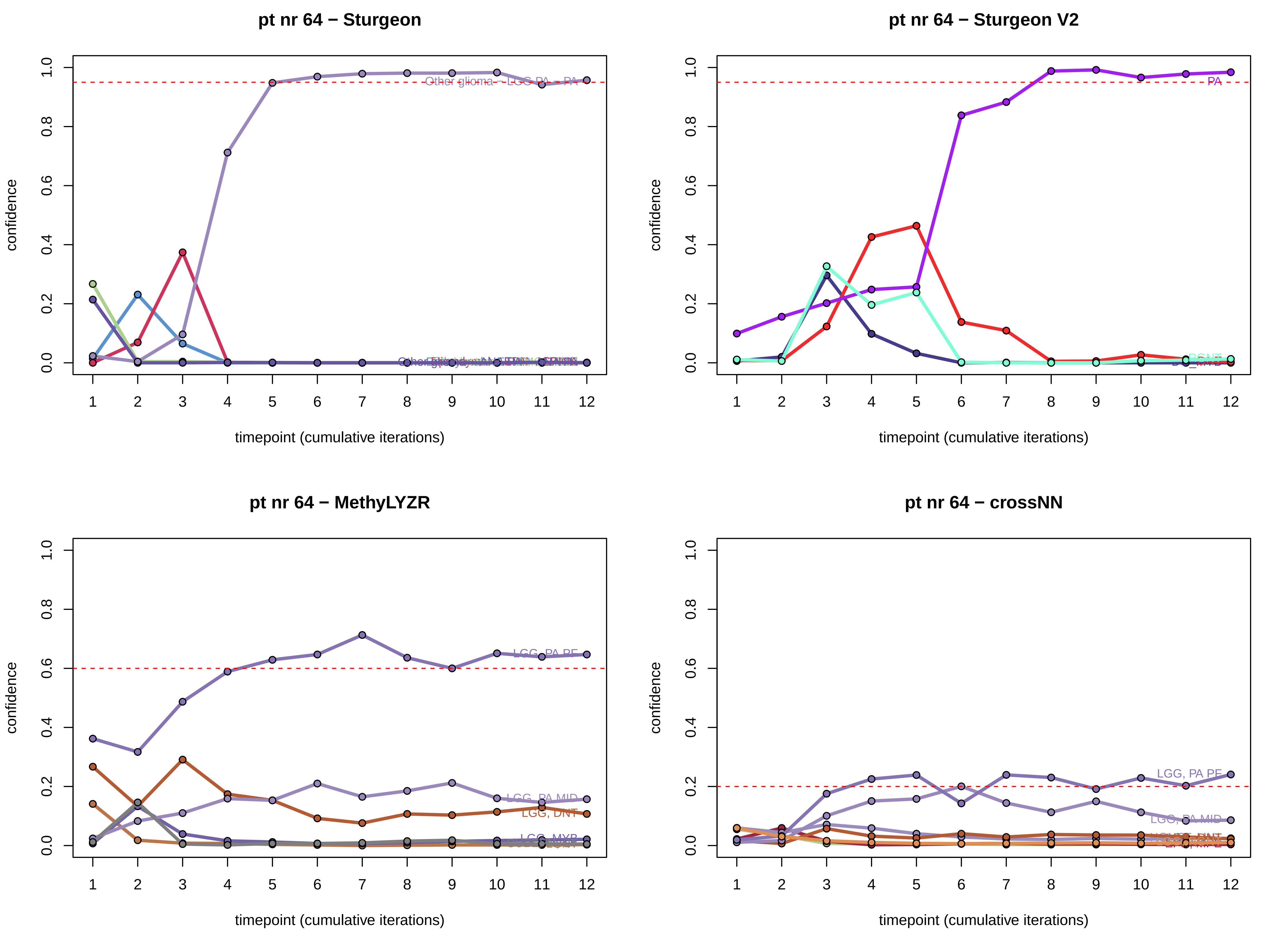

pt nr 64 − Sturgeon
pt nr 64 − Sturgeon V2
pt nr 64 − MethyLYZR
pt nr 64 − crossNN
confidence
timepoint (cumulative iterations)
0.0
0.2
0.4
0.6
0.8
1.0
1
2
3
4
5
6
7
8
9
10
11
12
PA
LGG, PA PF
LGG, PA MID
LGG, DNT
LGG, MYB

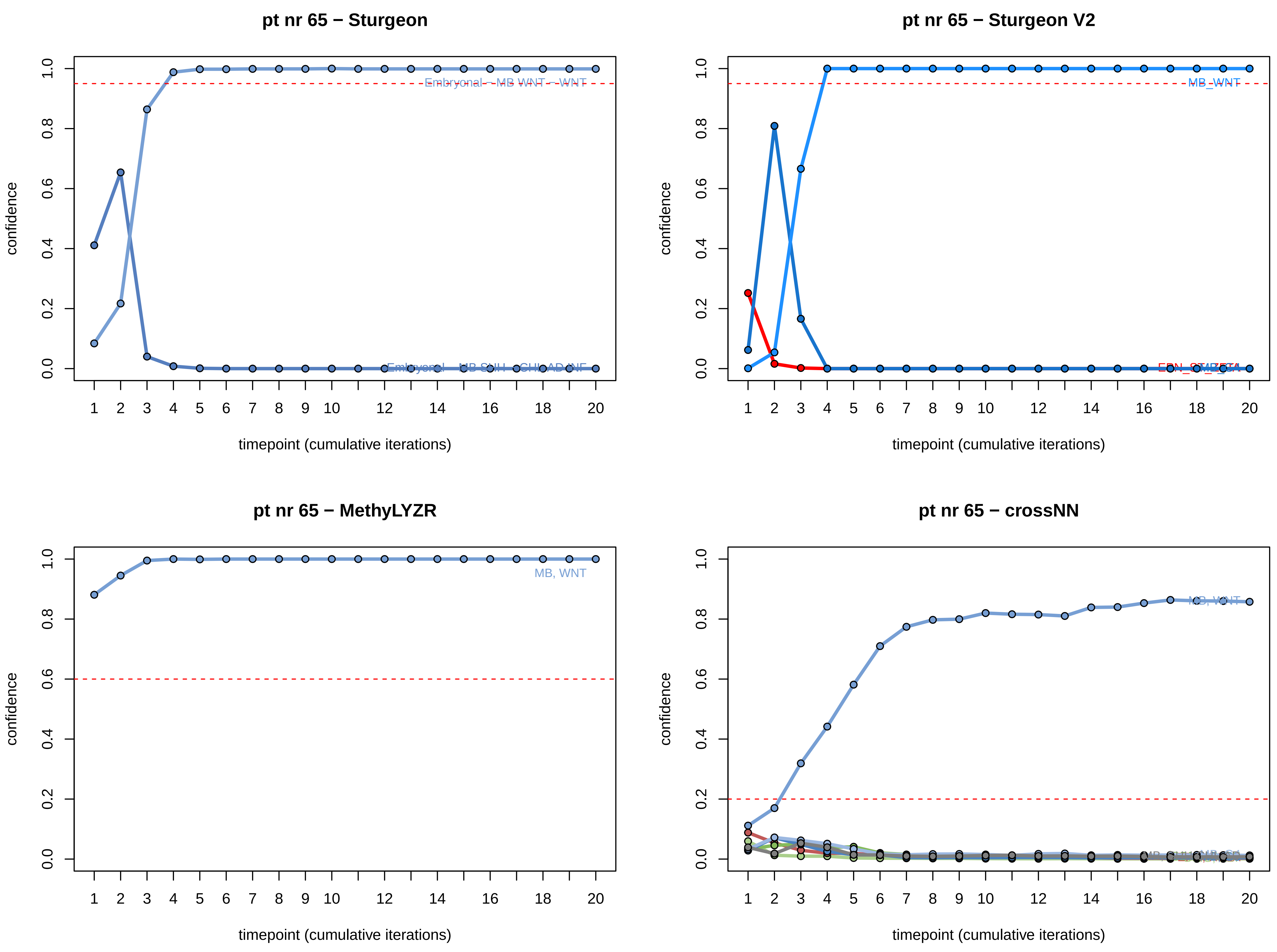
pt nr 65 − Sturgeon
Embryonal − MB WNT − WNT
confidence
timepoint (cumulative iterations)
pt nr 65 − Sturgeon V2
MB_WNT
confidence
timepoint (cumulative iterations)
pt nr 65 − MethyLYZR
MB, WNT
confidence
timepoint (cumulative iterations)
pt nr 65 − crossNN
MB, WNT
confidence
timepoint (cumulative iterations)

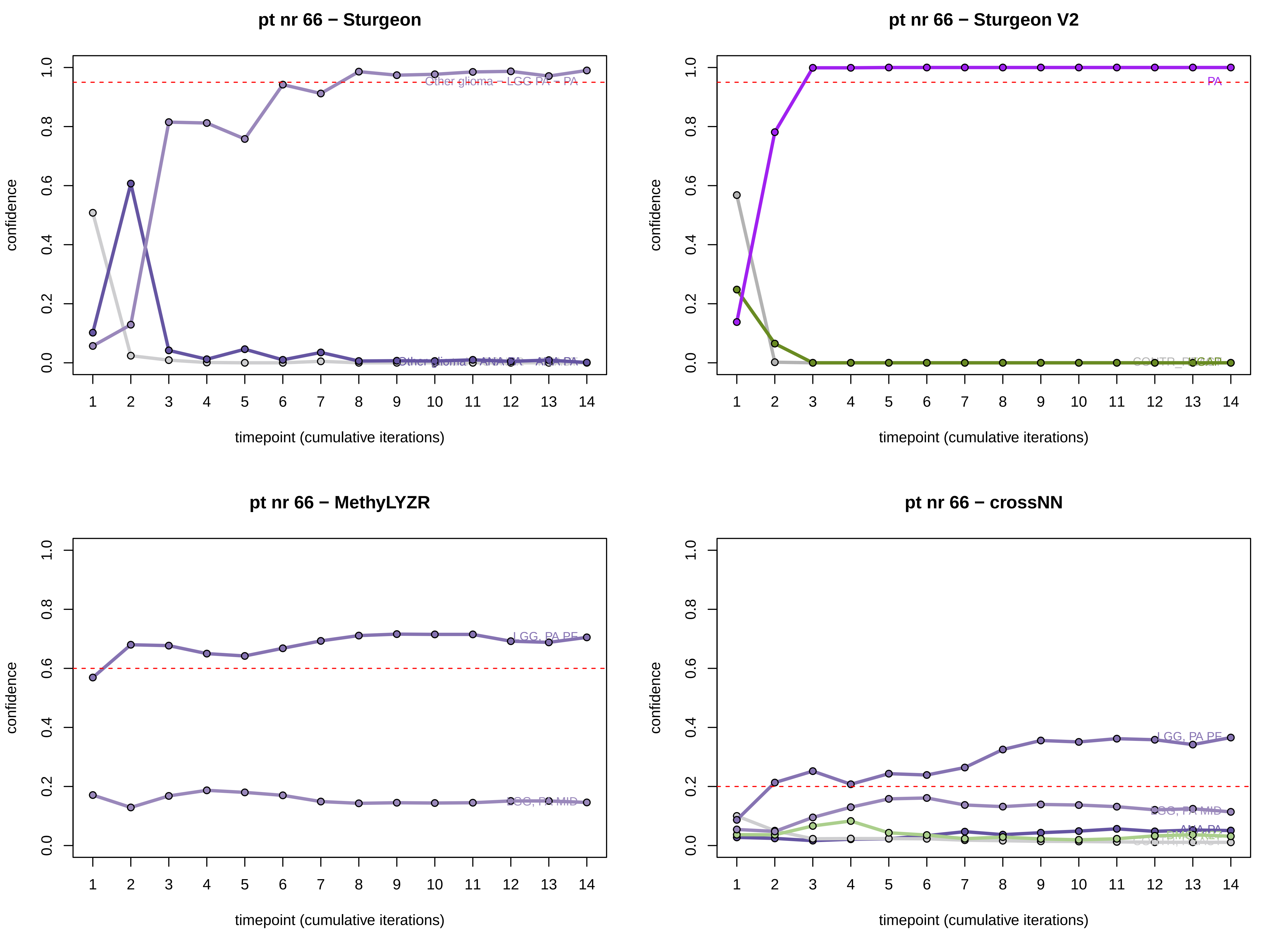
pt nr 66 − Sturgeon
Other glioma – LGG PA – PA
confidence
timepoint (cumulative iterations)
pt nr 66 − Sturgeon V2
PA
confidence
timepoint (cumulative iterations)
pt nr 66 − MethyLYZR
LGG, PA PF
LGG, PA MID
confidence
timepoint (cumulative iterations)
pt nr 66 − crossNN
LGG, PA PF
LGG, PA MID
ANA PA
confidence
timepoint (cumulative iterations)

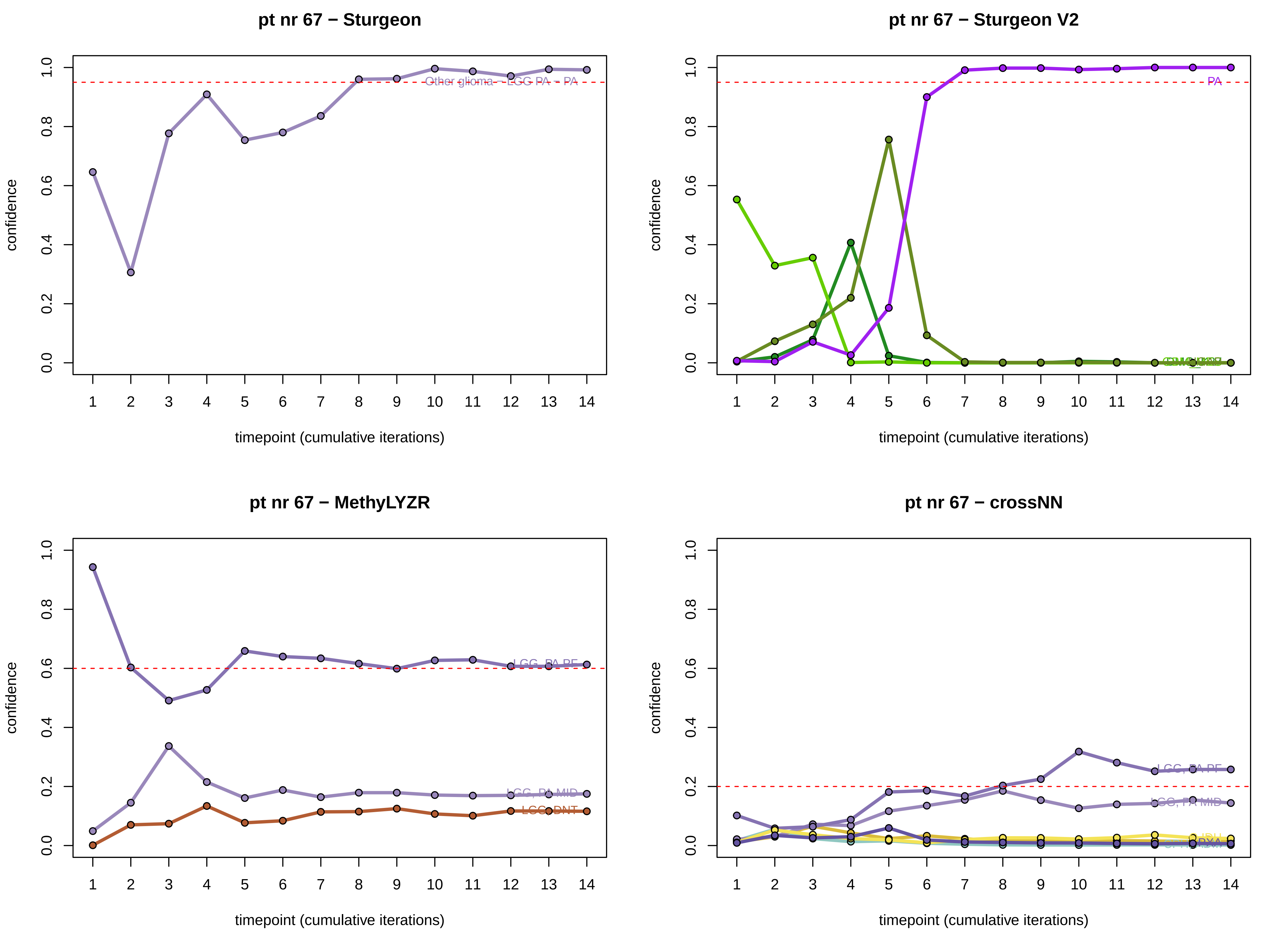
pt nr 67 − Sturgeon
confidence
0.0
0.2
0.4
0.6
0.8
1.0
Other glioma – LGG PA – PA
1 2 3 4 5 6 7 8 9 10 11 12 13 14
timepoint (cumulative iterations)
pt nr 67 − Sturgeon V2
confidence
0.0
0.2
0.4
0.6
0.8
1.0
PA
1 2 3 4 5 6 7 8 9 10 11 12 13 14
timepoint (cumulative iterations)
pt nr 67 − MethyLYZR
confidence
0.0
0.2
0.4
0.6
0.8
1.0
LGG, PA PF
LGG, PA MID
LGG, DNT
1 2 3 4 5 6 7 8 9 10 11 12 13 14
timepoint (cumulative iterations)
pt nr 67 − crossNN
confidence
0.0
0.2
0.4
0.6
0.8
1.0
LGG, PA PF
LGG, PA MID
PXA
1 2 3 4 5 6 7 8 9 10 11 12 13 14
timepoint (cumulative iterations)

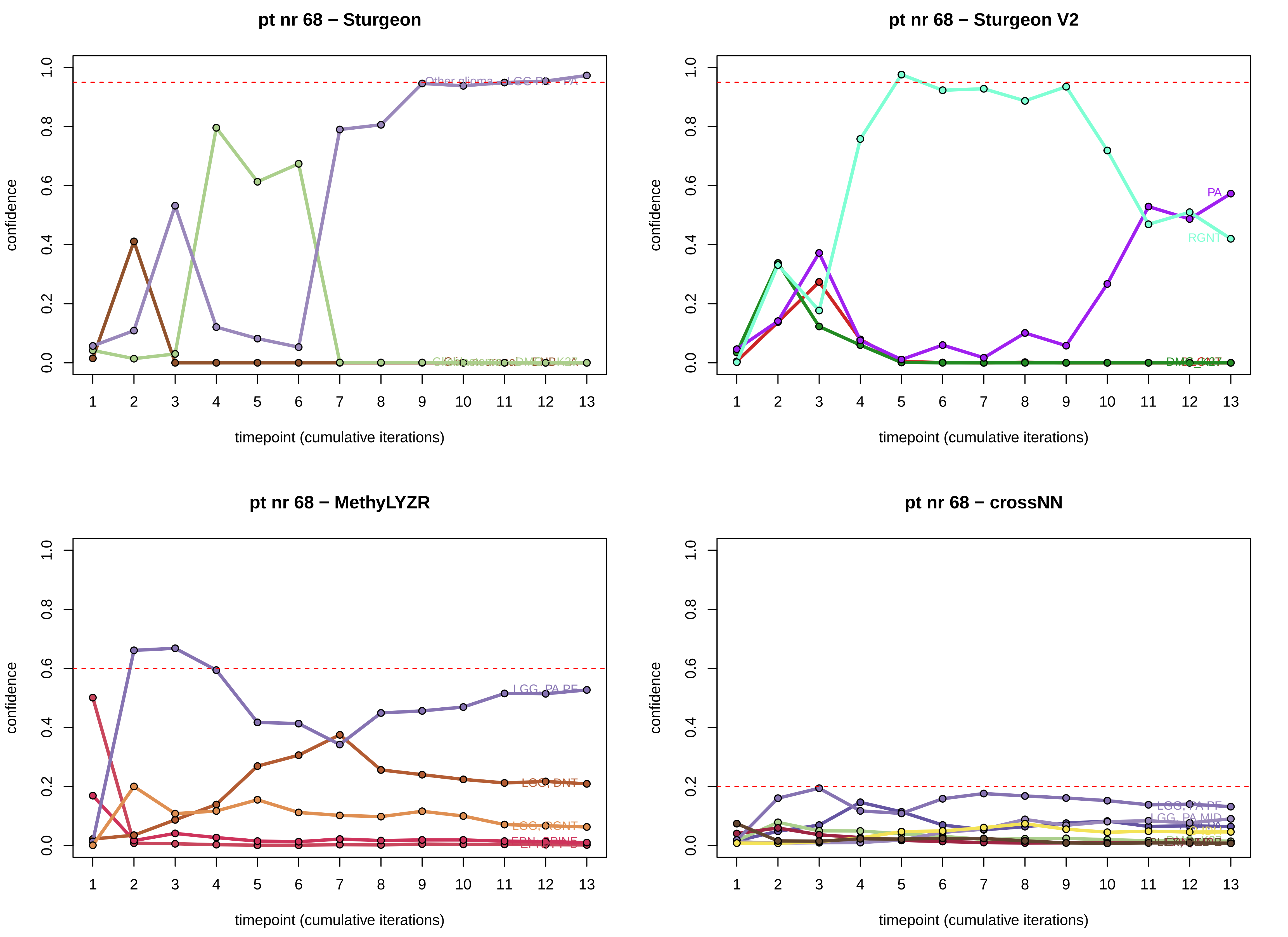
pt nr 68 − Sturgeon
confidence
timepoint (cumulative iterations)
pt nr 68 − Sturgeon V2
PA
RGNT
confidence
timepoint (cumulative iterations)
pt nr 68 − MethyLYZR
LGG, PA PF
LGG, DNT
LGG, RGNT
confidence
timepoint (cumulative iterations)
pt nr 68 − crossNN
LGG, PA PF
LGG, PA MID
confidence
timepoint (cumulative iterations)

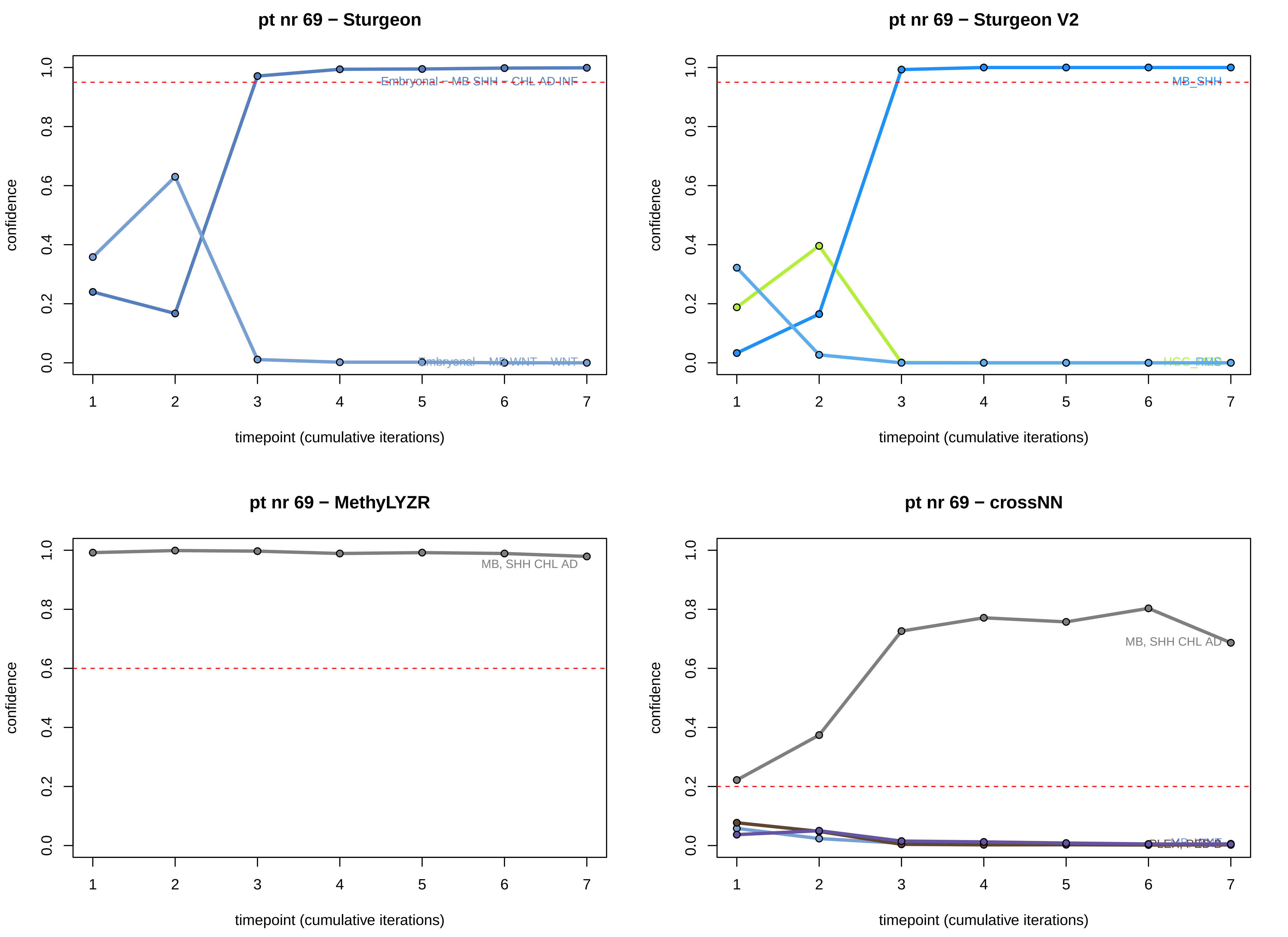
pt nr 69 − Sturgeon
Embryonal - MB SHH - CHL AD INF
Embryonal - MB WNT - WNT
confidence
timepoint (cumulative iterations)
pt nr 69 − Sturgeon V2
MB_SHH
confidence
timepoint (cumulative iterations)
pt nr 69 − MethyLYZR
MB, SHH CHL AD
confidence
timepoint (cumulative iterations)
pt nr 69 − crossNN
MB, SHH CHL AD
confidence
timepoint (cumulative iterations)

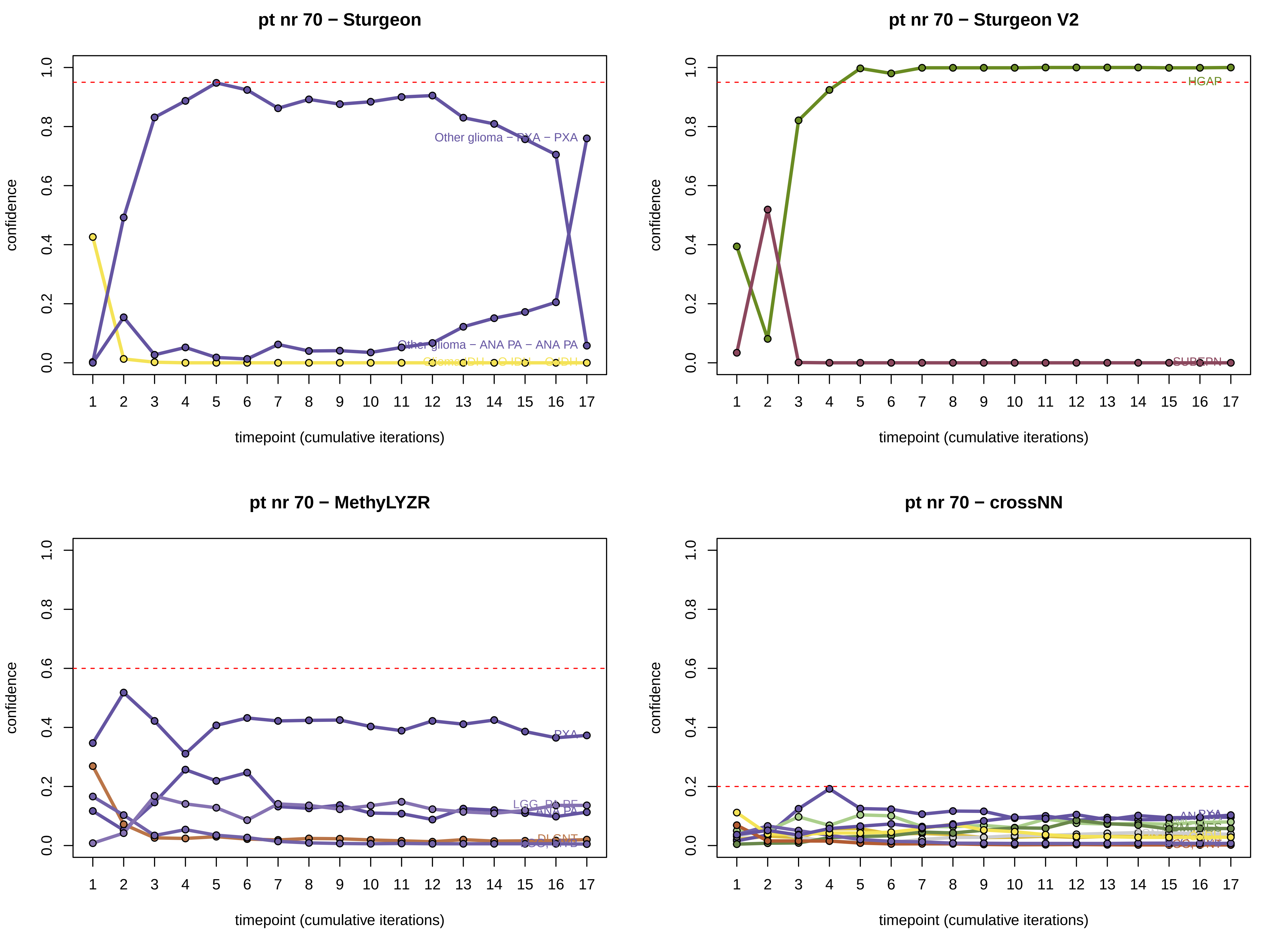
pt nr 70 − Sturgeon
confidence
timepoint (cumulative iterations)
Other glioma − PXA − PXA
Other glioma − ANA PA − ANA PA
pt nr 70 − Sturgeon V2
confidence
timepoint (cumulative iterations)
HGAP
SUBEPN
pt nr 70 − MethyLYZR
confidence
timepoint (cumulative iterations)
PXA
LGG, PA PF
ANA PA
DLGNT
pt nr 70 − crossNN
confidence
timepoint (cumulative iterations)

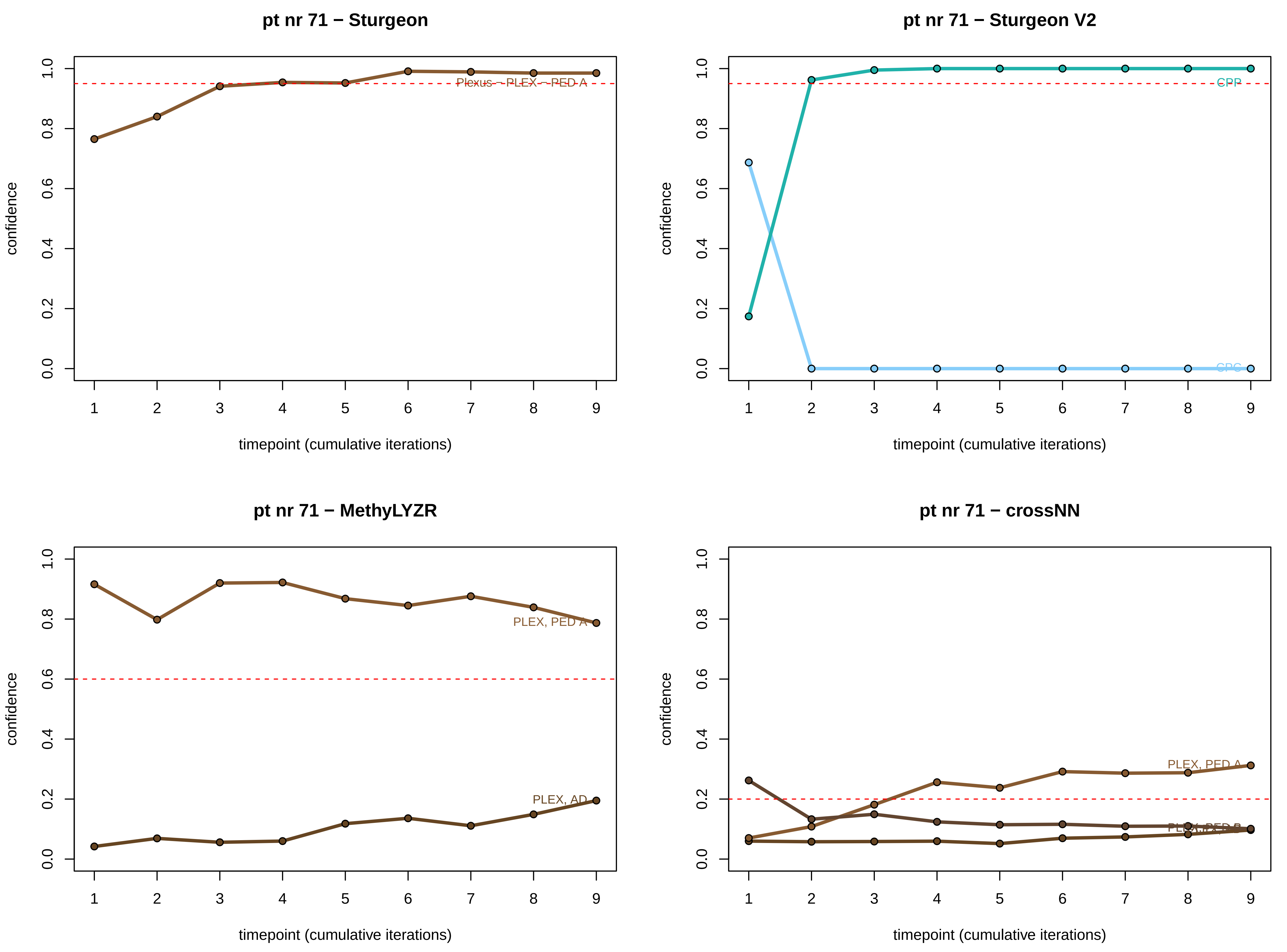
pt nr 71 − Sturgeon
Plexus − PLEX − PED A
pt nr 71 − Sturgeon V2
CPP
pt nr 71 − MethyLYZR
PLEX, PED A
PLEX, AD
pt nr 71 − crossNN
PLEX, PED A
confidence
timepoint (cumulative iterations)
0.0
0.2
0.4
0.6
0.8
1.0
1
2
3
4
5
6
7
8
9

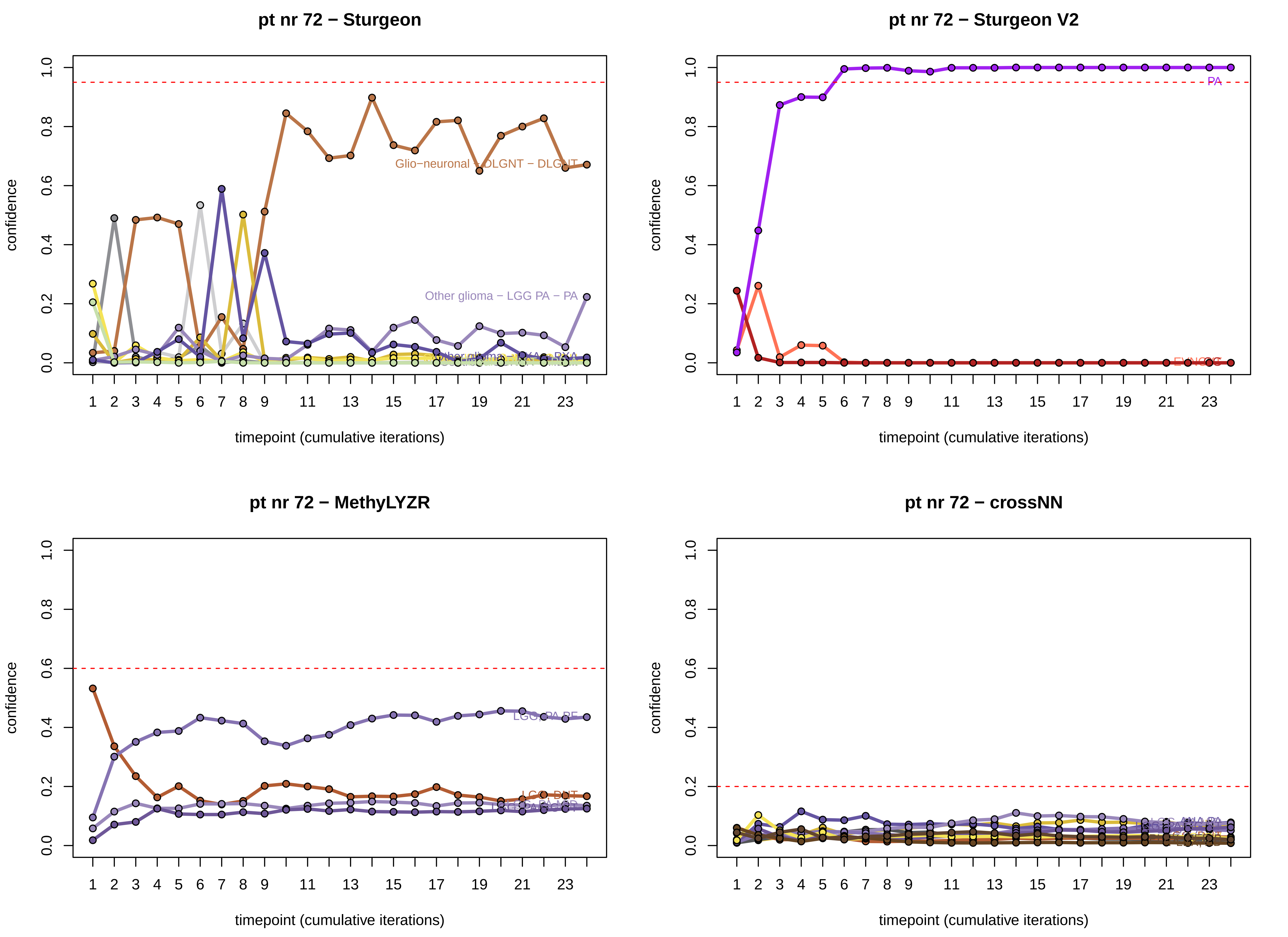
pt nr 72 − Sturgeon
Glio−neuronal − DLGNT − DLGNT
Other glioma − LGG PA − PA
pt nr 72 − Sturgeon V2
PA
pt nr 72 − MethyLYZR
LGG, PA PF
LGG, DNT
pt nr 72 − crossNN
confidence
timepoint (cumulative iterations)

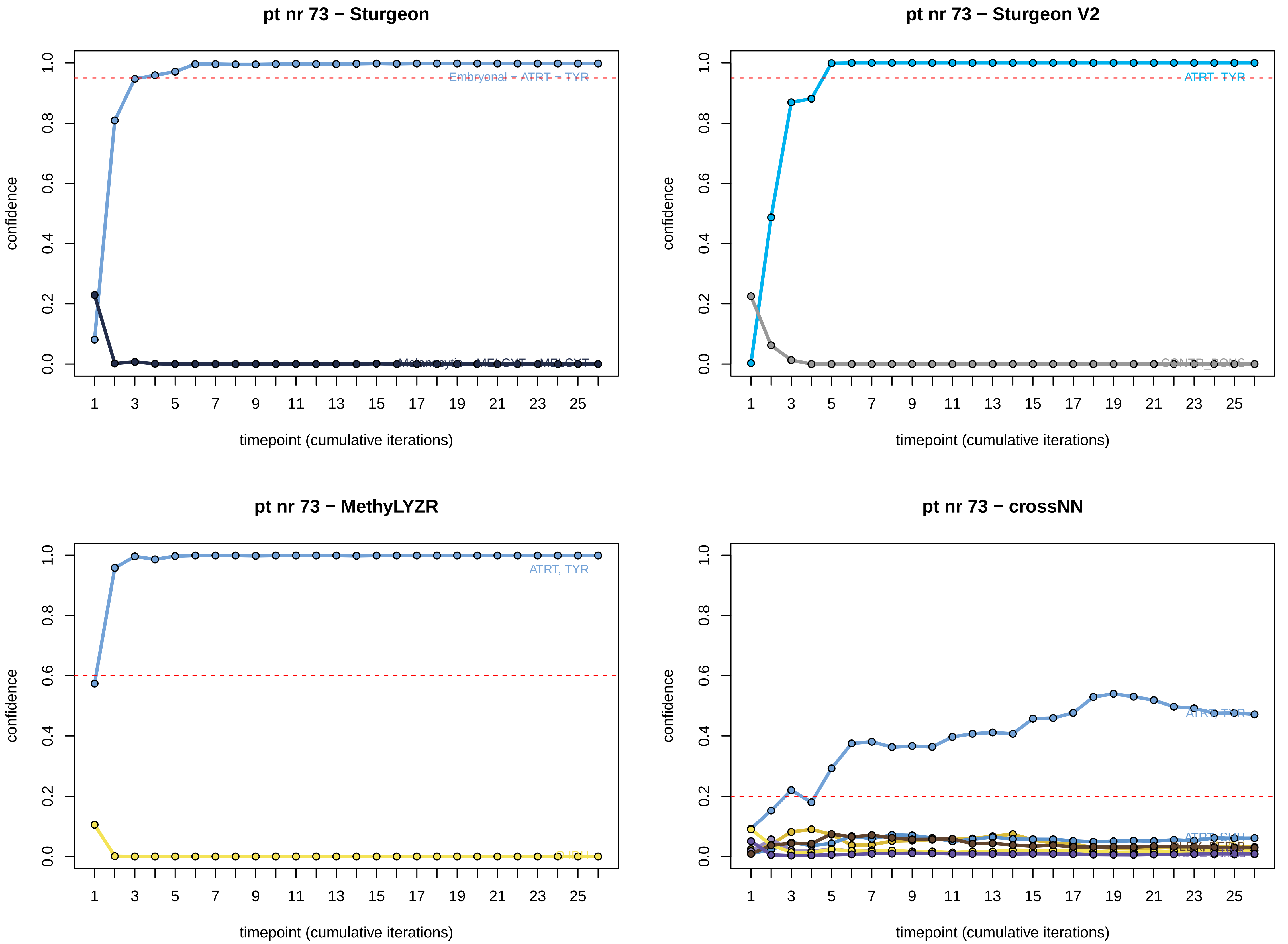
pt nr 73 − Sturgeon
Embryonal - ATRT - TYR
Melanocytic - MELCYT - MELCYT
confidence
timepoint (cumulative iterations)
pt nr 73 − Sturgeon V2
ATRT_TYR
CONTR_PONS
confidence
timepoint (cumulative iterations)
pt nr 73 − MethyLYZR
ATRT, TYR
O IDH
confidence
timepoint (cumulative iterations)
pt nr 73 − crossNN
ATRT, TYR
ATRT, SHH
confidence
timepoint (cumulative iterations)

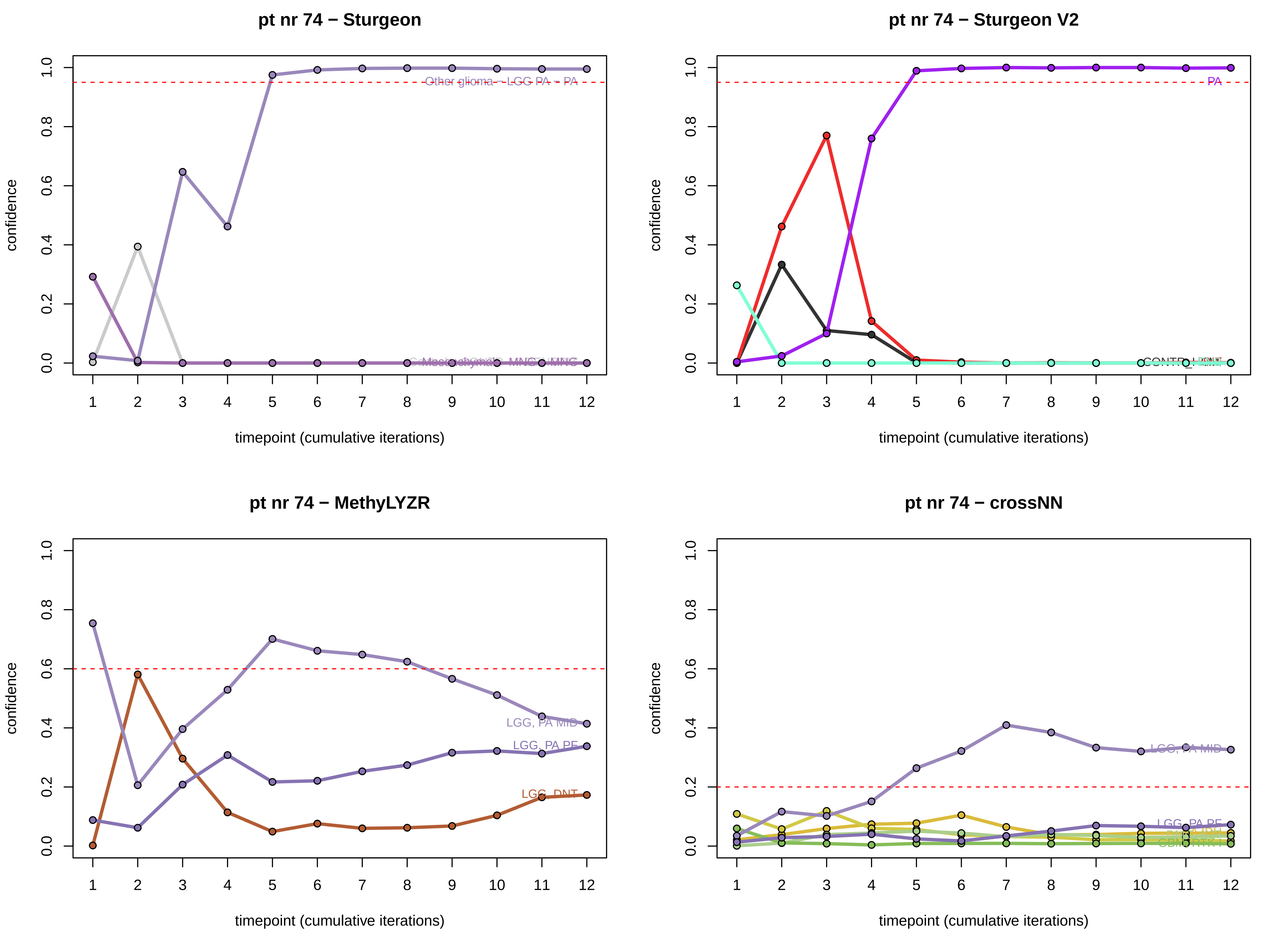
pt nr 74 − Sturgeon
Other glioma − LGG PA − PA
confidence
timepoint (cumulative iterations)
pt nr 74 − Sturgeon V2
PA
confidence
timepoint (cumulative iterations)
pt nr 74 − MethyLYZR
LGG, PA MID
LGG, PA PF
LGG, DNT
confidence
timepoint (cumulative iterations)
pt nr 74 − crossNN
LGG, PA MID
LGG, PA PF
confidence
timepoint (cumulative iterations)

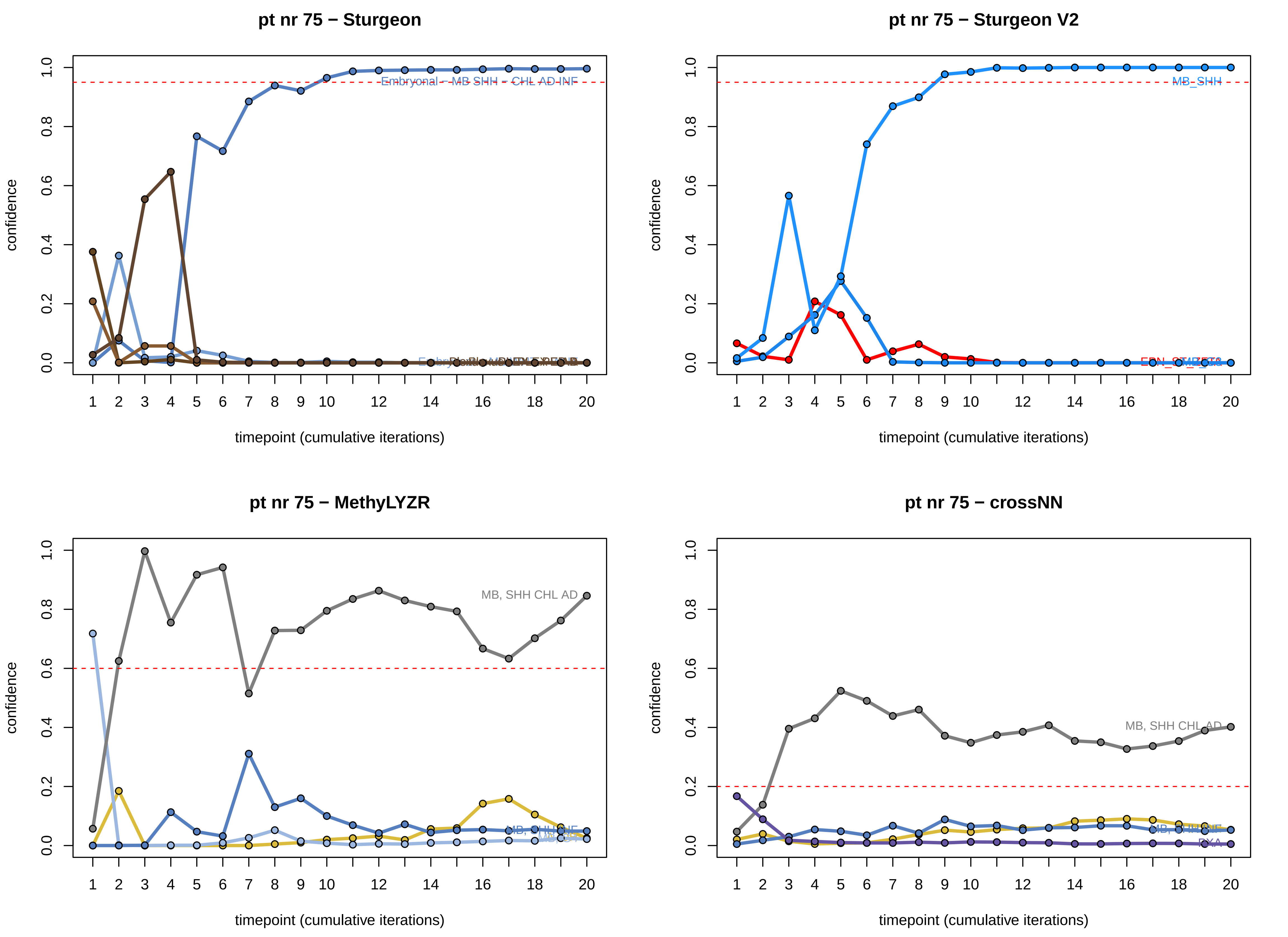

pt nr 75 − Sturgeon
Embryonal - MB SHH - CHL AD INF
confidence
timepoint (cumulative iterations)
pt nr 75 − Sturgeon V2
MB_SHH
confidence
timepoint (cumulative iterations)
pt nr 75 − MethyLYZR
MB, SHH CHL AD
confidence
timepoint (cumulative iterations)
pt nr 75 − crossNN
MB, SHH CHL AD
PXA
confidence
timepoint (cumulative iterations)

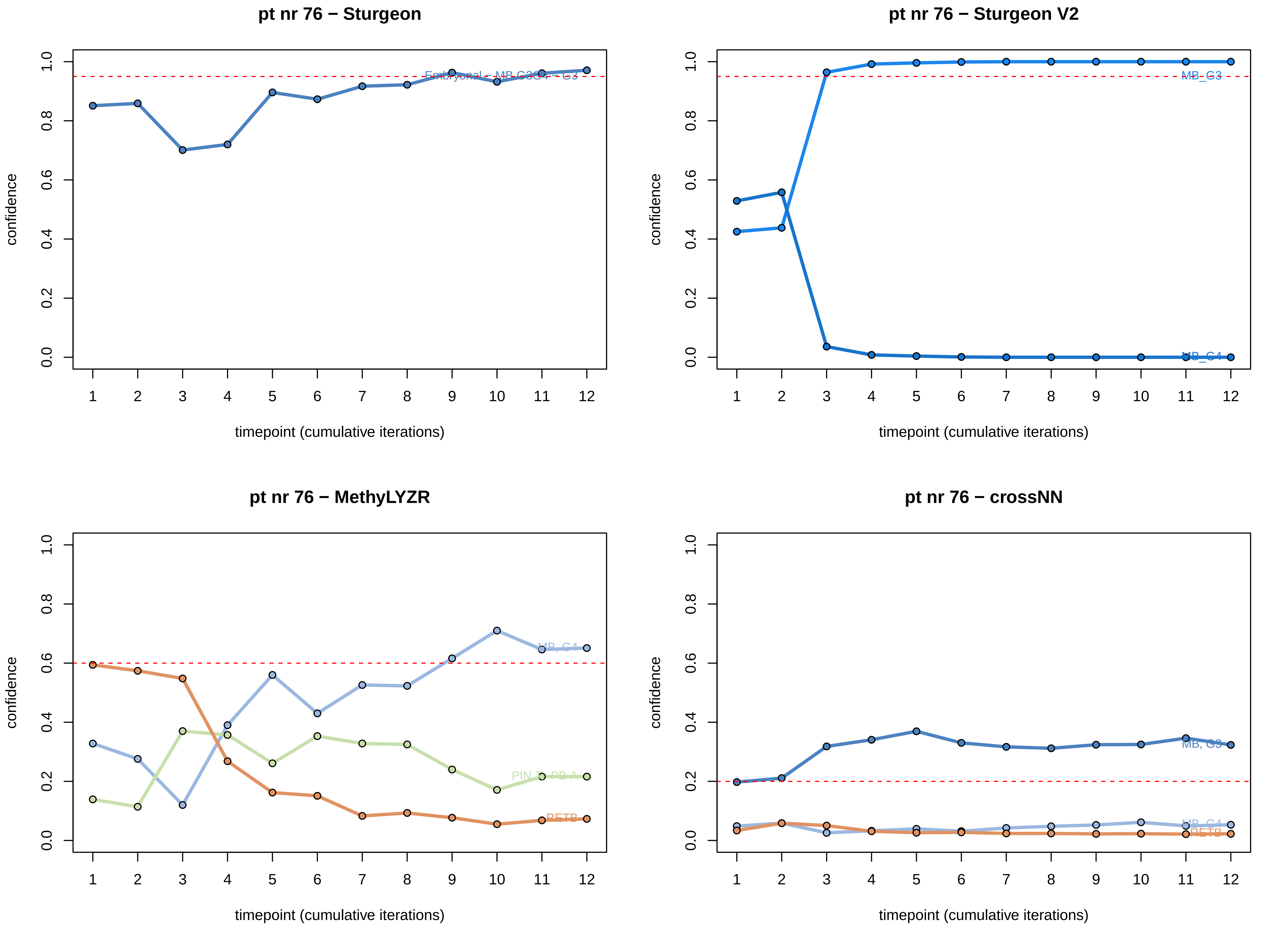
pt nr 76 − Sturgeon
confidence
0.0
0.2
0.4
0.6
0.8
1.0
Embryonal – MB G3G4 – G3
1 2 3 4 5 6 7 8 9 10 11 12
timepoint (cumulative iterations)
pt nr 76 − Sturgeon V2
confidence
0.0
0.2
0.4
0.6
0.8
1.0
MB_G3
MB_G4
1 2 3 4 5 6 7 8 9 10 11 12
timepoint (cumulative iterations)
pt nr 76 − MethyLYZR
confidence
0.0
0.2
0.4
0.6
0.8
1.0
MB, G4
PIN T, PB A
RETB
1 2 3 4 5 6 7 8 9 10 11 12
timepoint (cumulative iterations)
pt nr 76 − crossNN
confidence
0.0
0.2
0.4
0.6
0.8
1.0
MB, G3
MB, G4
RETB
1 2 3 4 5 6 7 8 9 10 11 12
timepoint (cumulative iterations)

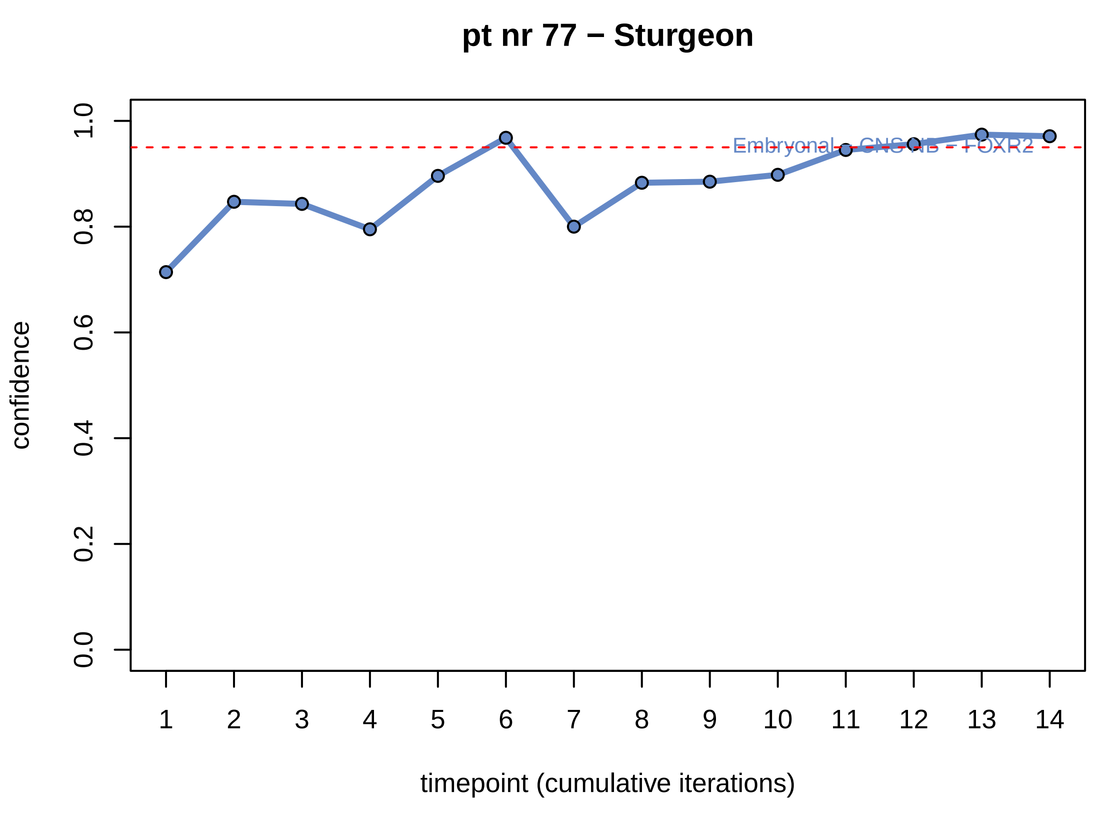
pt nr 77 − Sturgeon
confidence
timepoint (cumulative iterations)
Embryonal - CNS NB - FOXR2

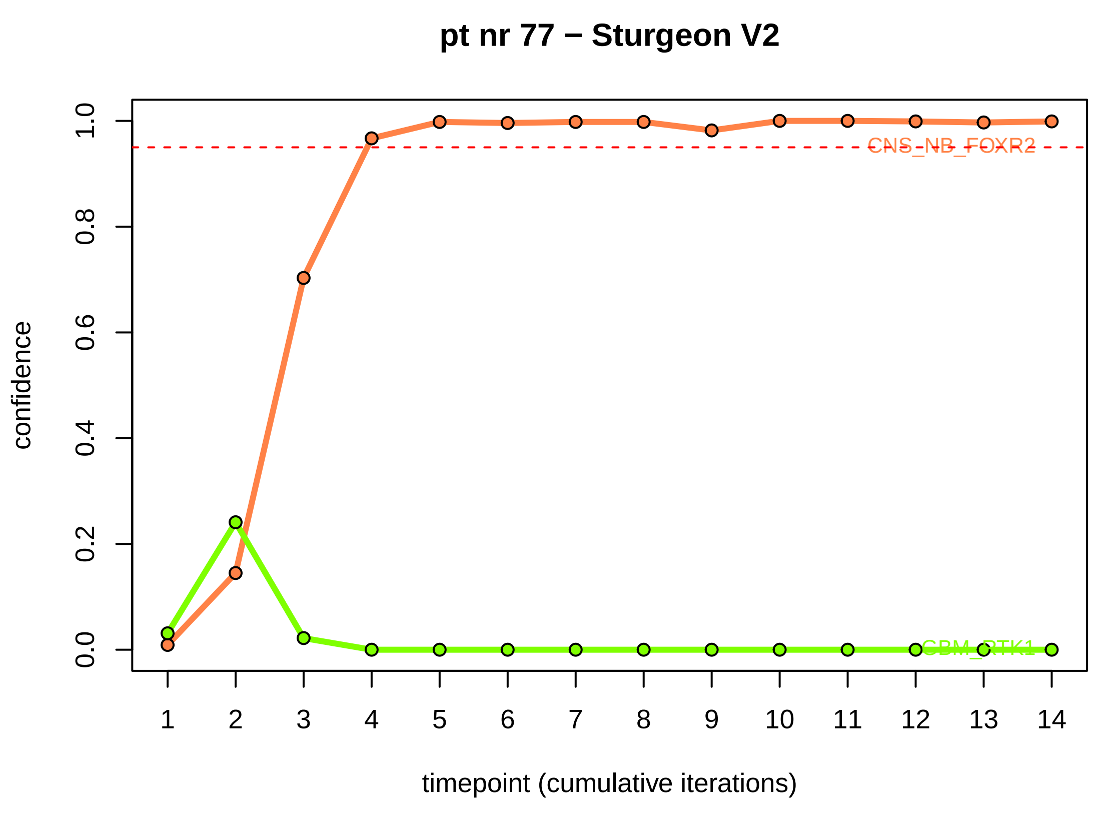
pt nr 77 − Sturgeon V2
confidence
timepoint (cumulative iterations)
CNS_NB_FOXR2
GBM_RTK1

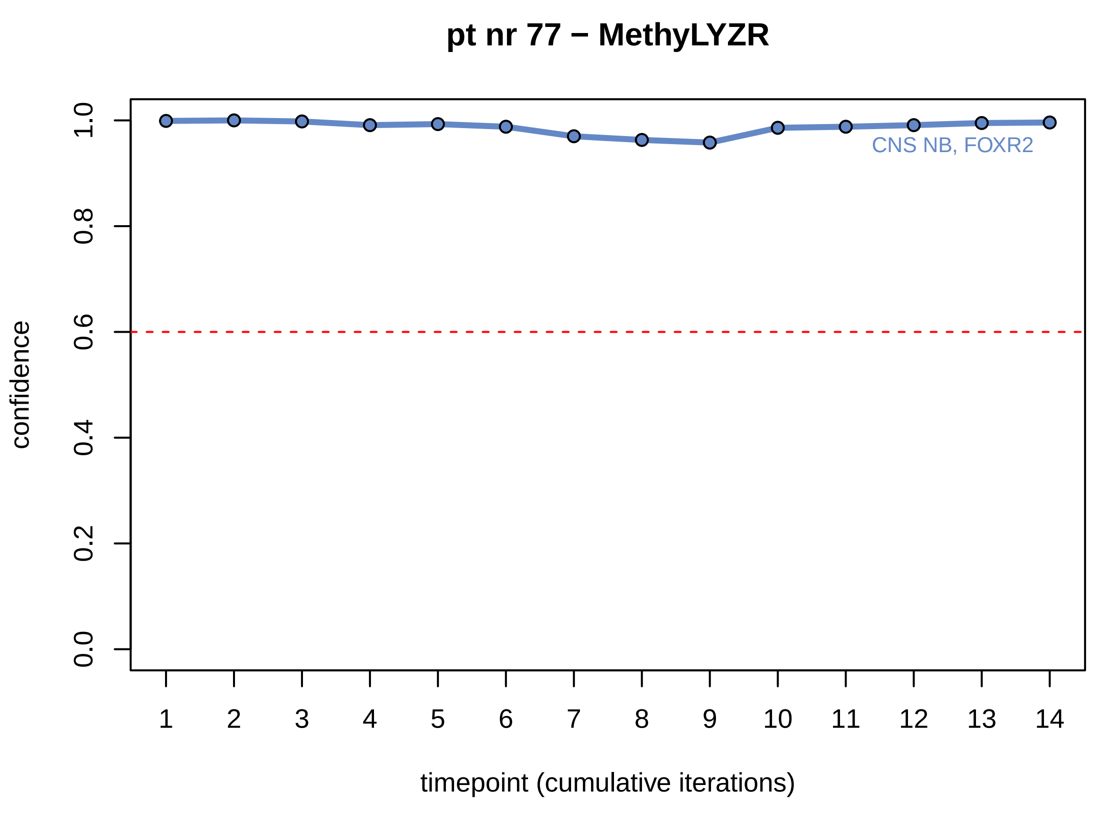
pt nr 77 − MethyLYZR
confidence
timepoint (cumulative iterations)
CNS NB, FOXR2

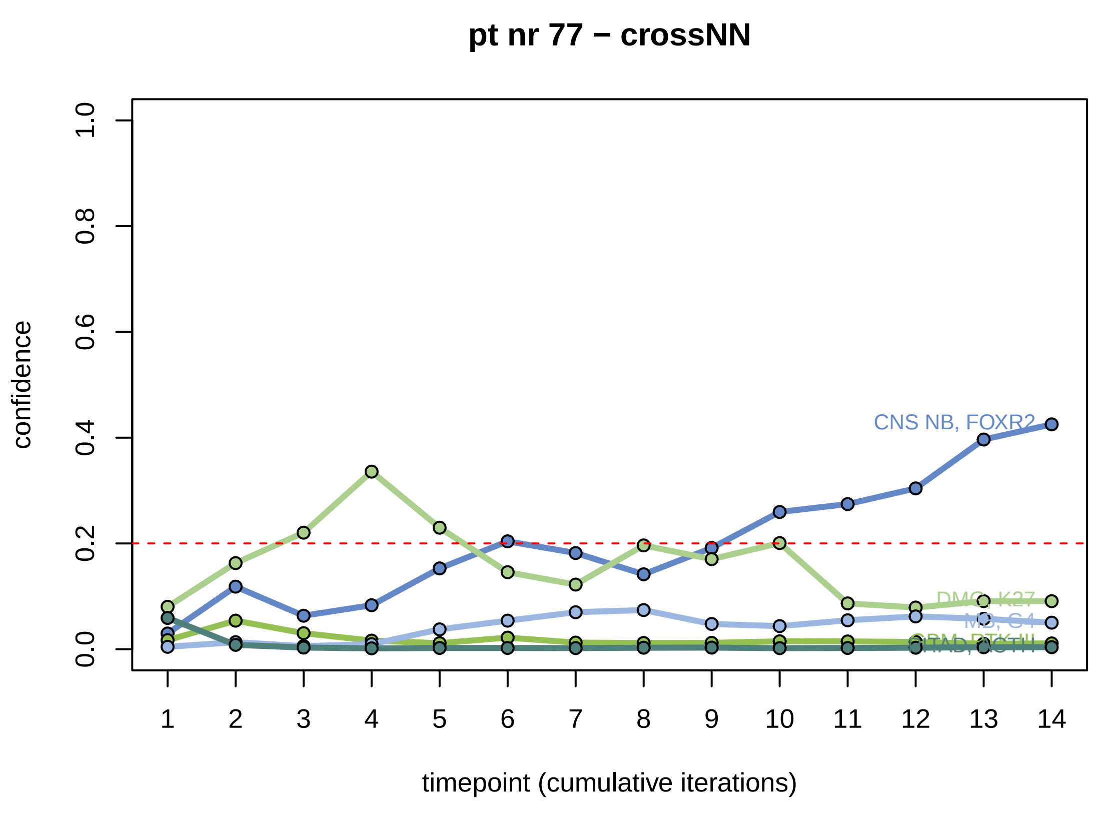
pt nr 77 − crossNN
confidence
timepoint (cumulative iterations)
CNS NB, FOXR2
DMG, K27
MB, G4
PITAD, ACTH

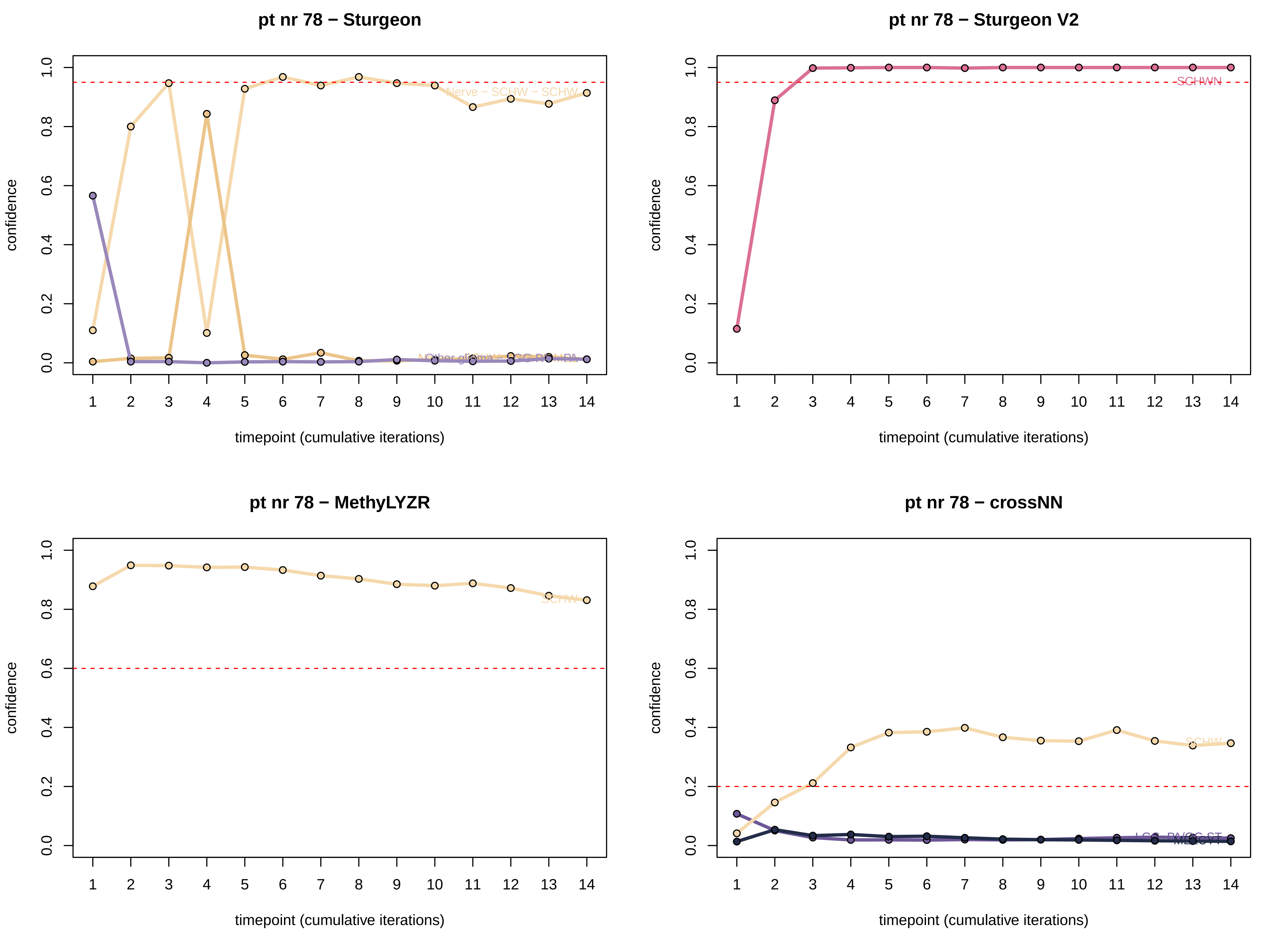
pt nr 78 − Sturgeon
pt nr 78 − Sturgeon V2
pt nr 78 − MethyLYZR
pt nr 78 − crossNN
confidence
timepoint (cumulative iterations)
Nerve − SCHW − SCHW
SCHWN
SCHW
SCHW

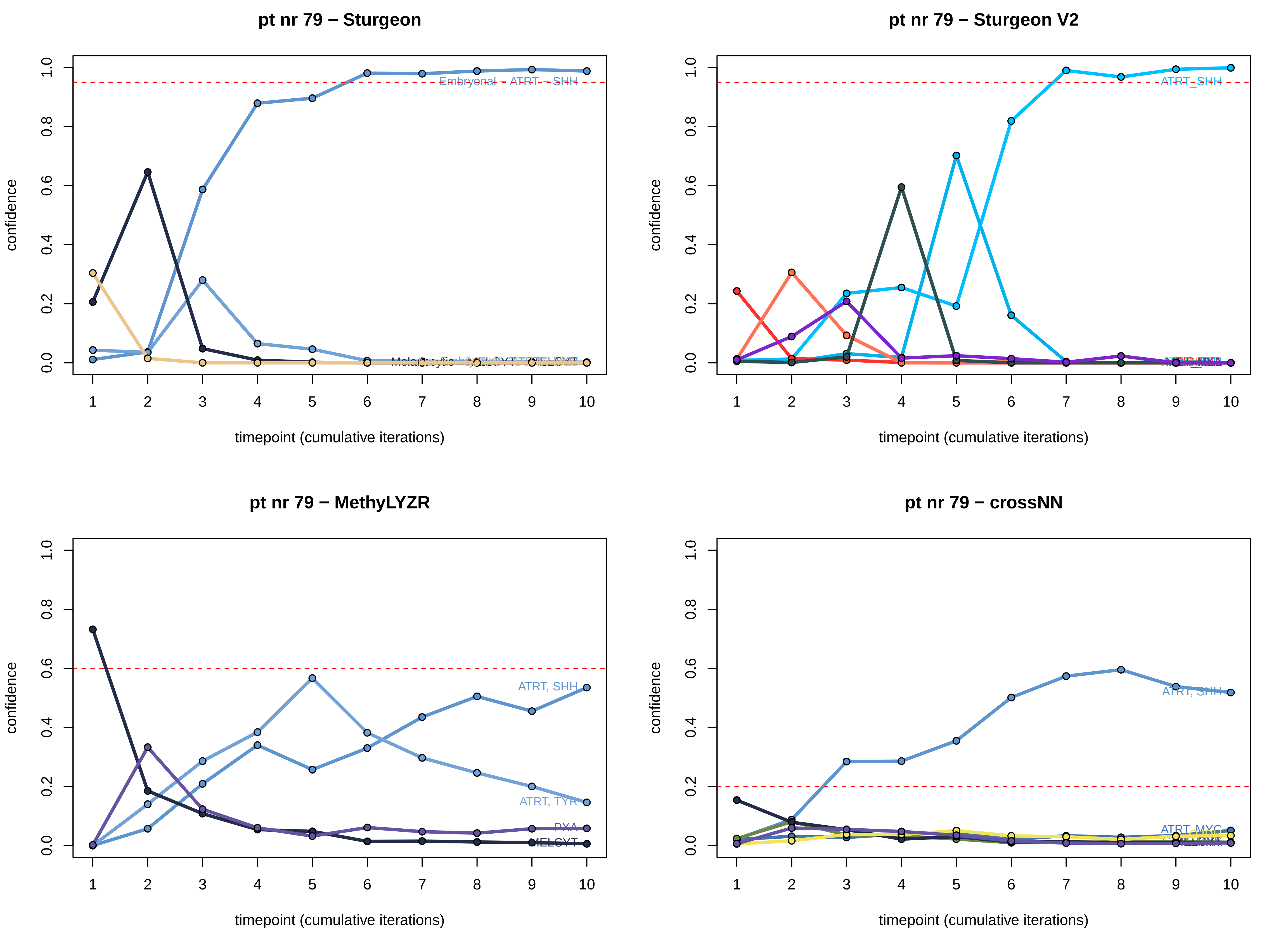
pt nr 79 − Sturgeon
pt nr 79 − Sturgeon V2
pt nr 79 − MethyLYZR
pt nr 79 − crossNN
confidence
timepoint (cumulative iterations)
0.0
0.2
0.4
0.6
0.8
1.0
1
2
3
4
5
6
7
8
9
10
Embryonal − ATRT − SHH
ATRT_SHH
ATRT, SHH
ATRT, TYR
PXA
MELCYT
ATRT, MYC

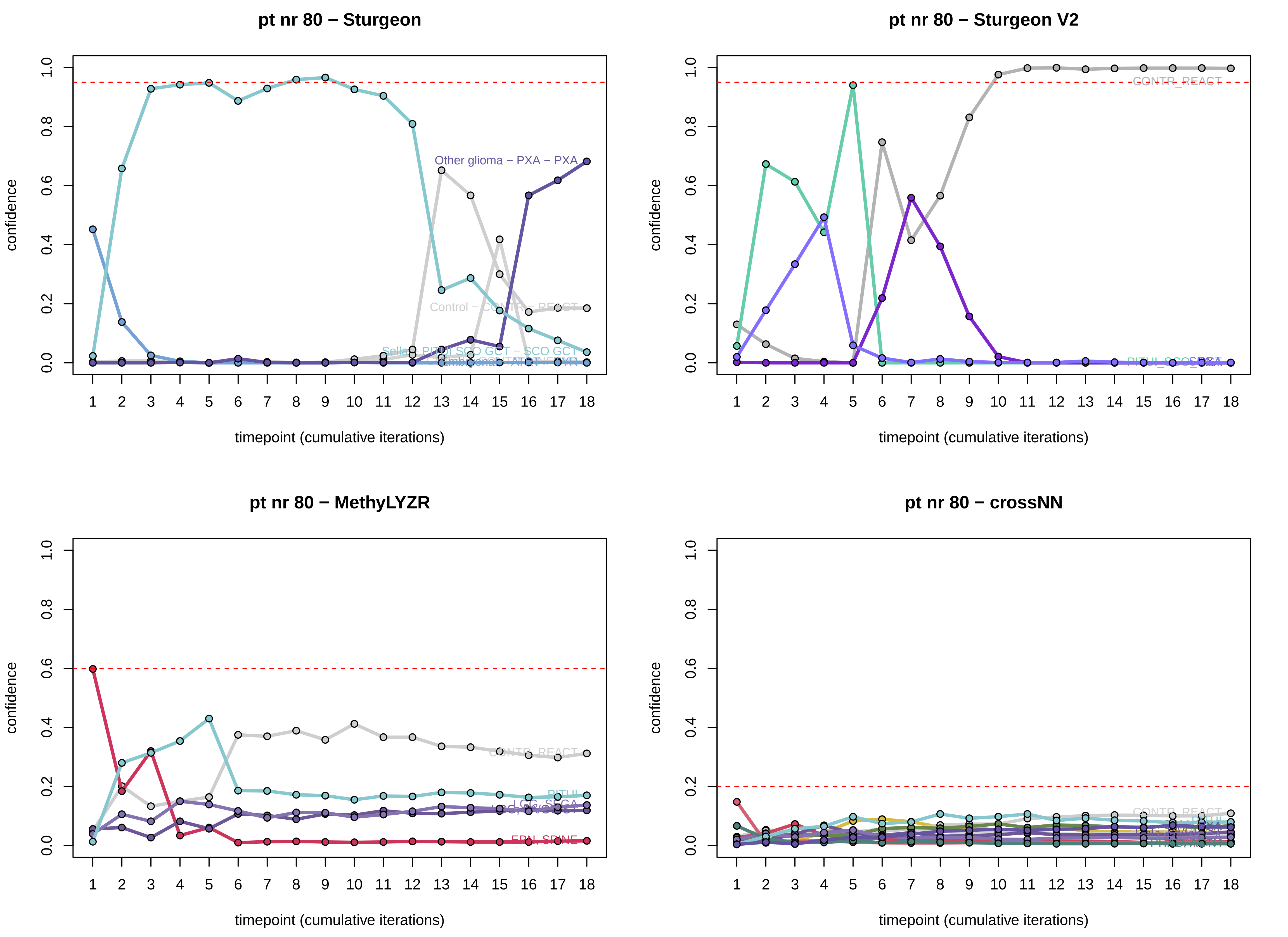
pt nr 80 − Sturgeon
Other glioma − PXA − PXA
Control − CONTR − REACT
timepoint (cumulative iterations)
confidence
pt nr 80 − Sturgeon V2
CONTR_REACT
pt nr 80 − MethyLYZR
CONTR_REACT
PITUI
LGG_SEGA
EPN_SPINE
pt nr 80 − crossNN
CONTR_REACT

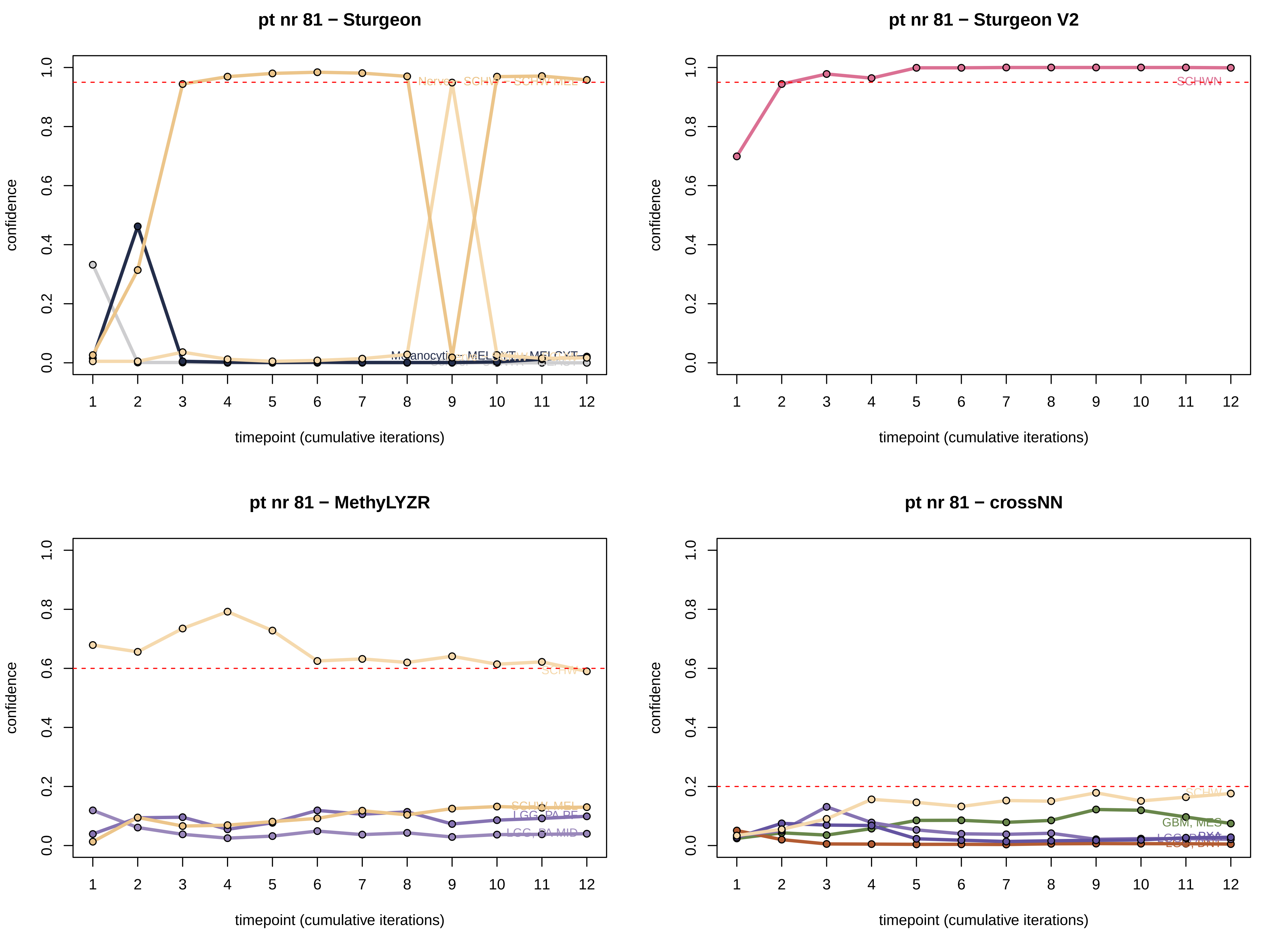
pt nr 81 − Sturgeon
pt nr 81 − Sturgeon V2
pt nr 81 − MethyLYZR
pt nr 81 − crossNN
confidence
timepoint (cumulative iterations)
Nerve - SCHW
SCHW MEL
MELCYT
SCHWN
SCHW
SCHW, MEL
LGG, PA PF
LGG, PA MID
GBM, MES

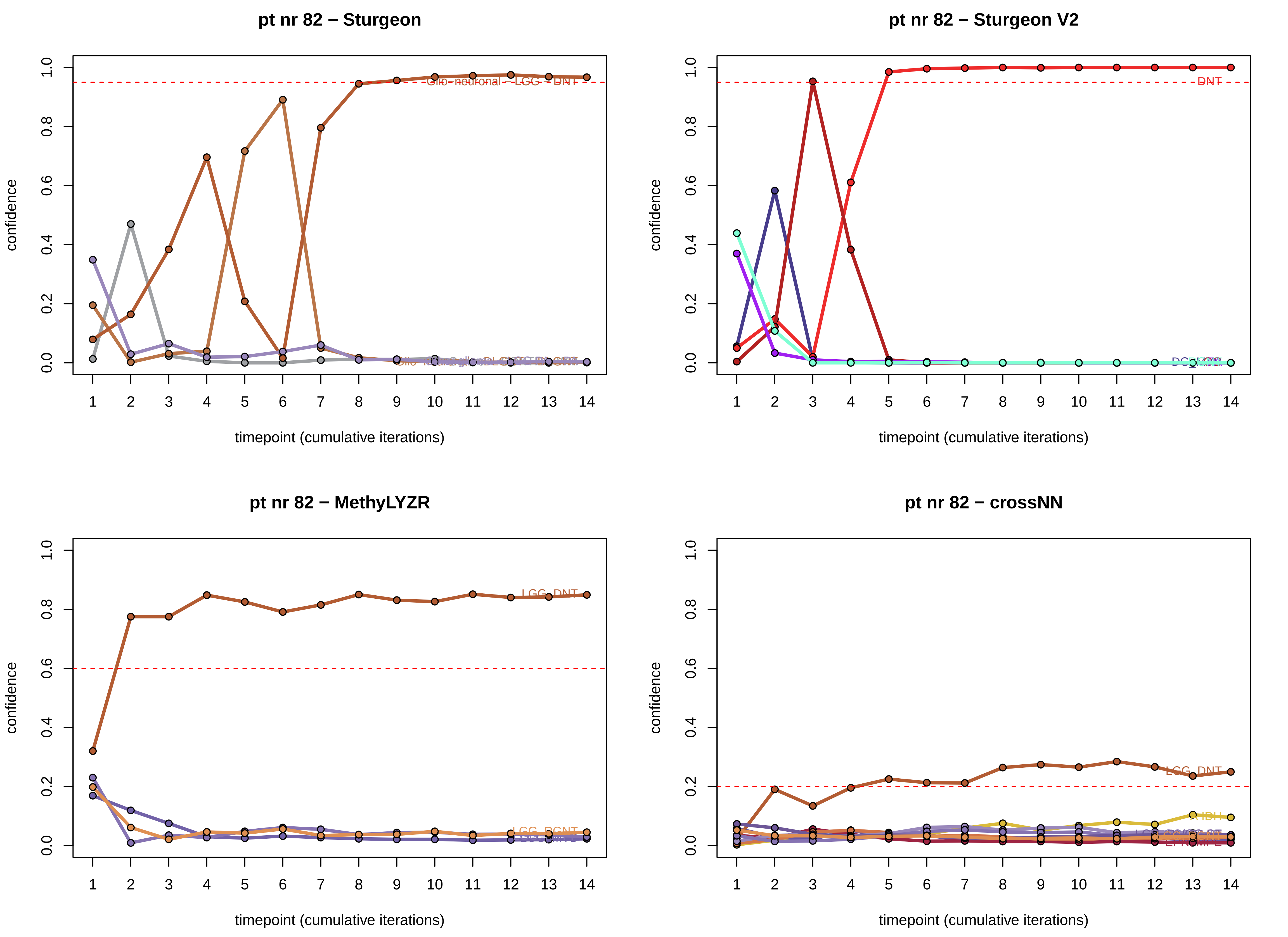
pt nr 82 − Sturgeon
Glio−neuronal − LGG − DNT
confidence
timepoint (cumulative iterations)
pt nr 82 − Sturgeon V2
DNT
confidence
timepoint (cumulative iterations)
pt nr 82 − MethyLYZR
LGG_DNT
LGG_RGNT
confidence
timepoint (cumulative iterations)
pt nr 82 − crossNN
LGG_DNT
A_IDH
confidence
timepoint (cumulative iterations)

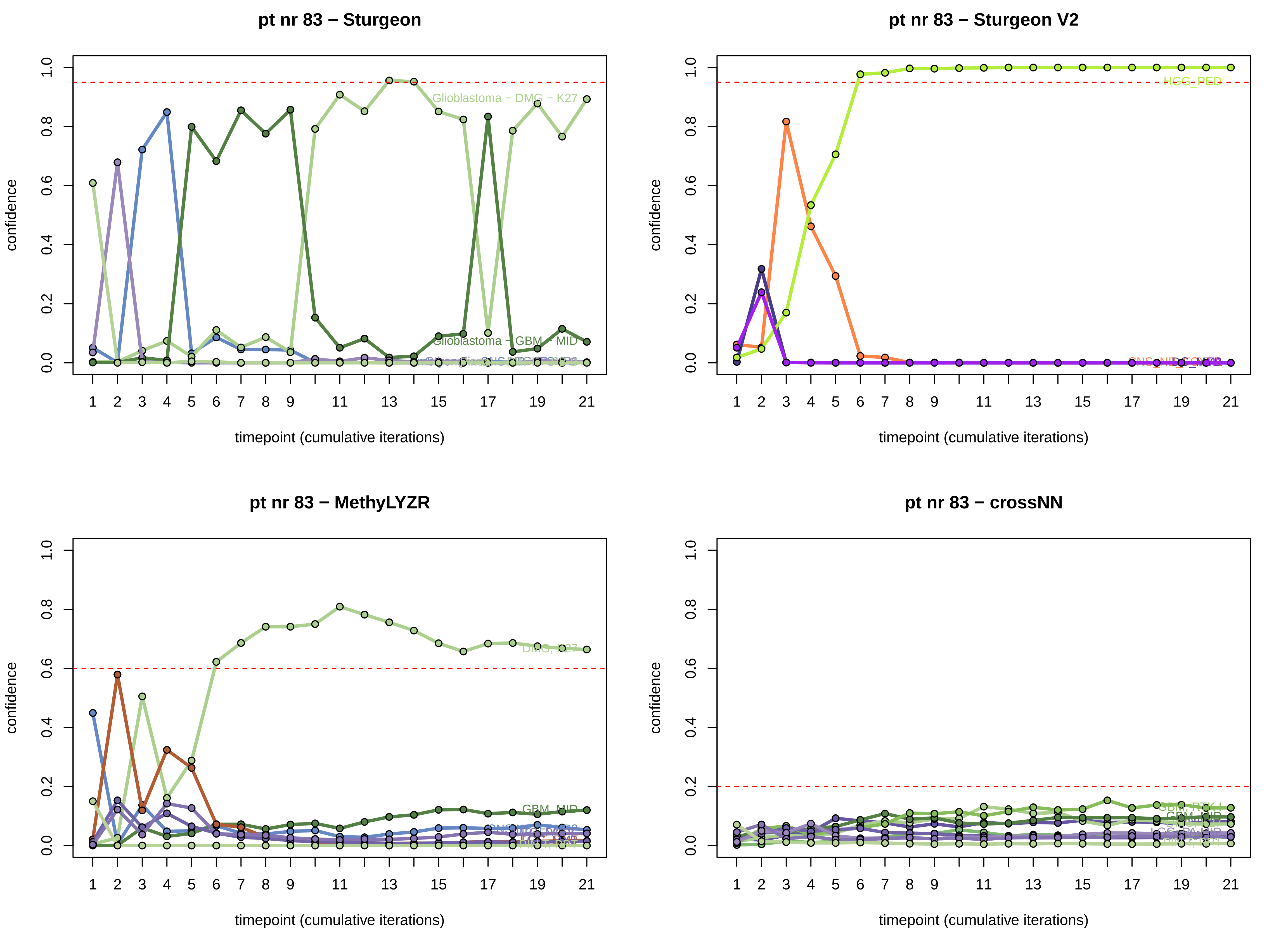
pt nr 83 − Sturgeon
pt nr 83 − Sturgeon V2
pt nr 83 − MethyLYZR
pt nr 83 − crossNN
confidence
timepoint (cumulative iterations)
Glioblastoma − DMG − K27
Glioblastoma − GBM − MID
HGG_PED
DMG, K27
GBM, MID
0.0
0.2
0.4
0.6
0.8
1.0
1
2
3
4
5
6
7
8
9
11
13
15
17
19
21

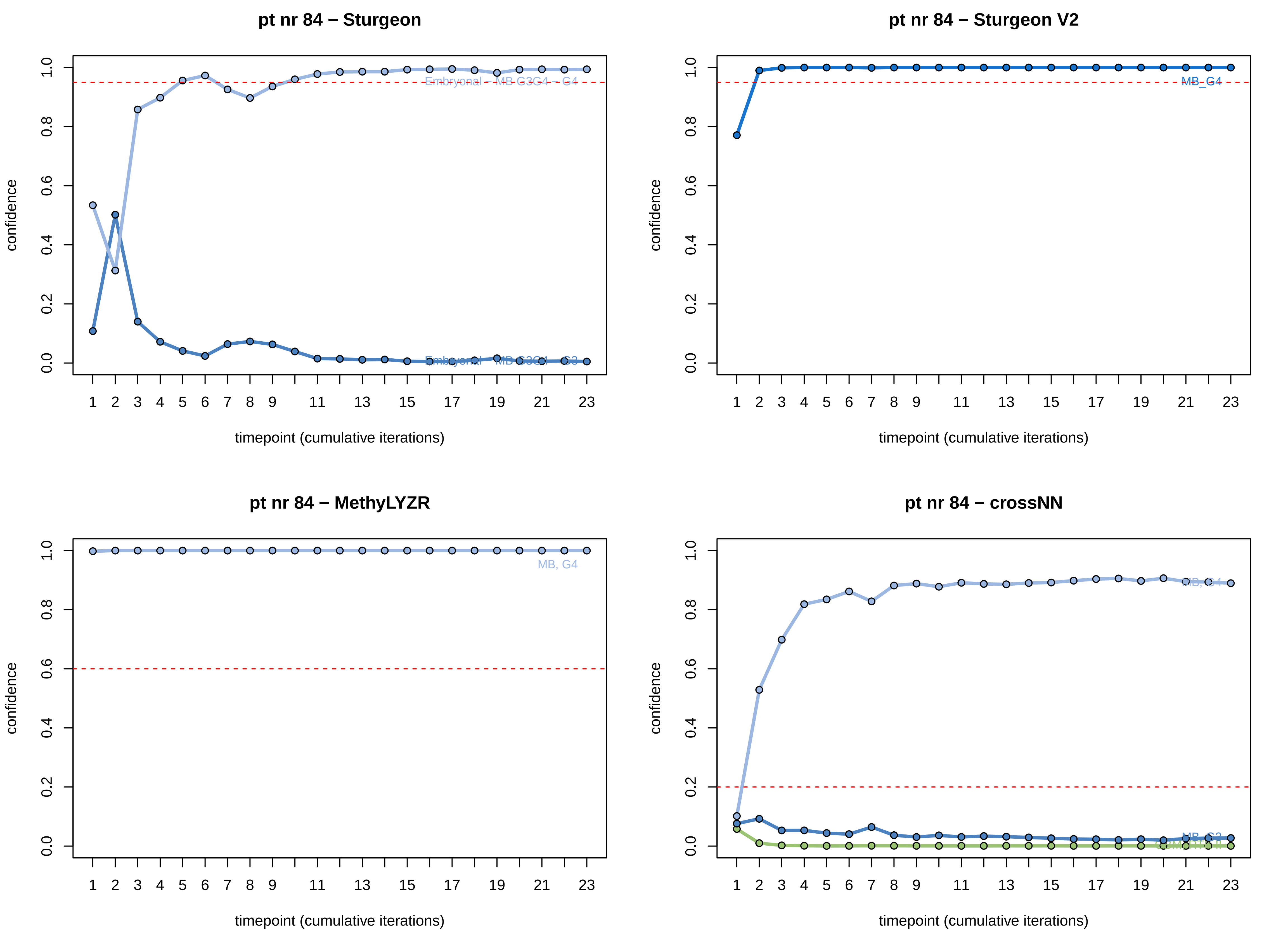
pt nr 84 − Sturgeon
pt nr 84 − Sturgeon V2
pt nr 84 − MethyLYZR
pt nr 84 − crossNN
confidence
0.0
0.2
0.4
0.6
0.8
1.0
timepoint (cumulative iterations)
1 2 3 4 5 6 7 8 9 11 13 15 17 19 21 23
Embryonal − MB G3G4 − G4
MB_G4
MB, G4
MB, G3

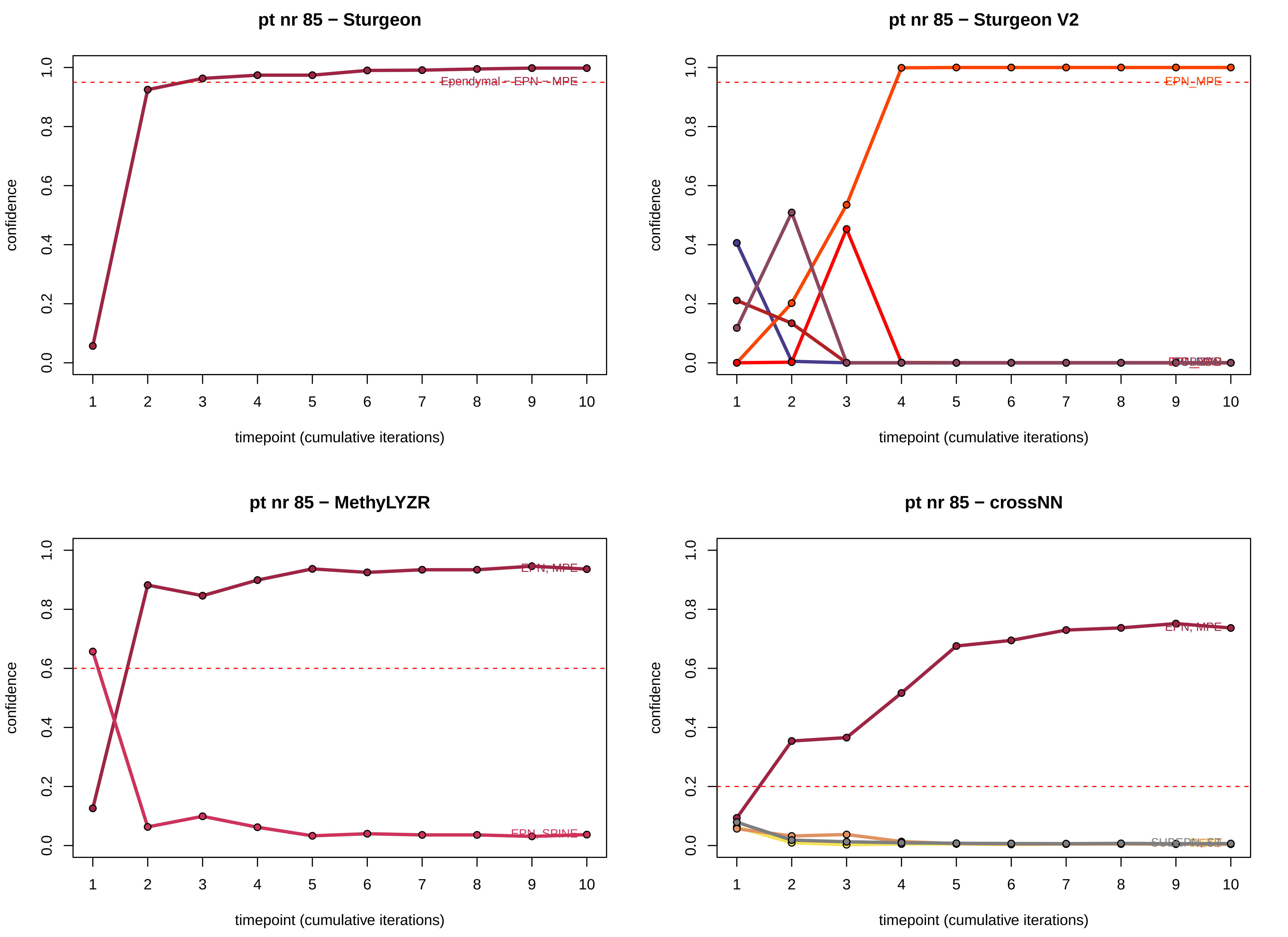
pt nr 85 − Sturgeon
Ependymal − EPN − MPE
pt nr 85 − Sturgeon V2
EPN_MPE
pt nr 85 − MethyLYZR
EPN, MPE
EPN, SPINE
pt nr 85 − crossNN
EPN, MPE
confidence
0.0
0.2
0.4
0.6
0.8
1.0
1
2
3
4
5
6
7
8
9
10
timepoint (cumulative iterations)

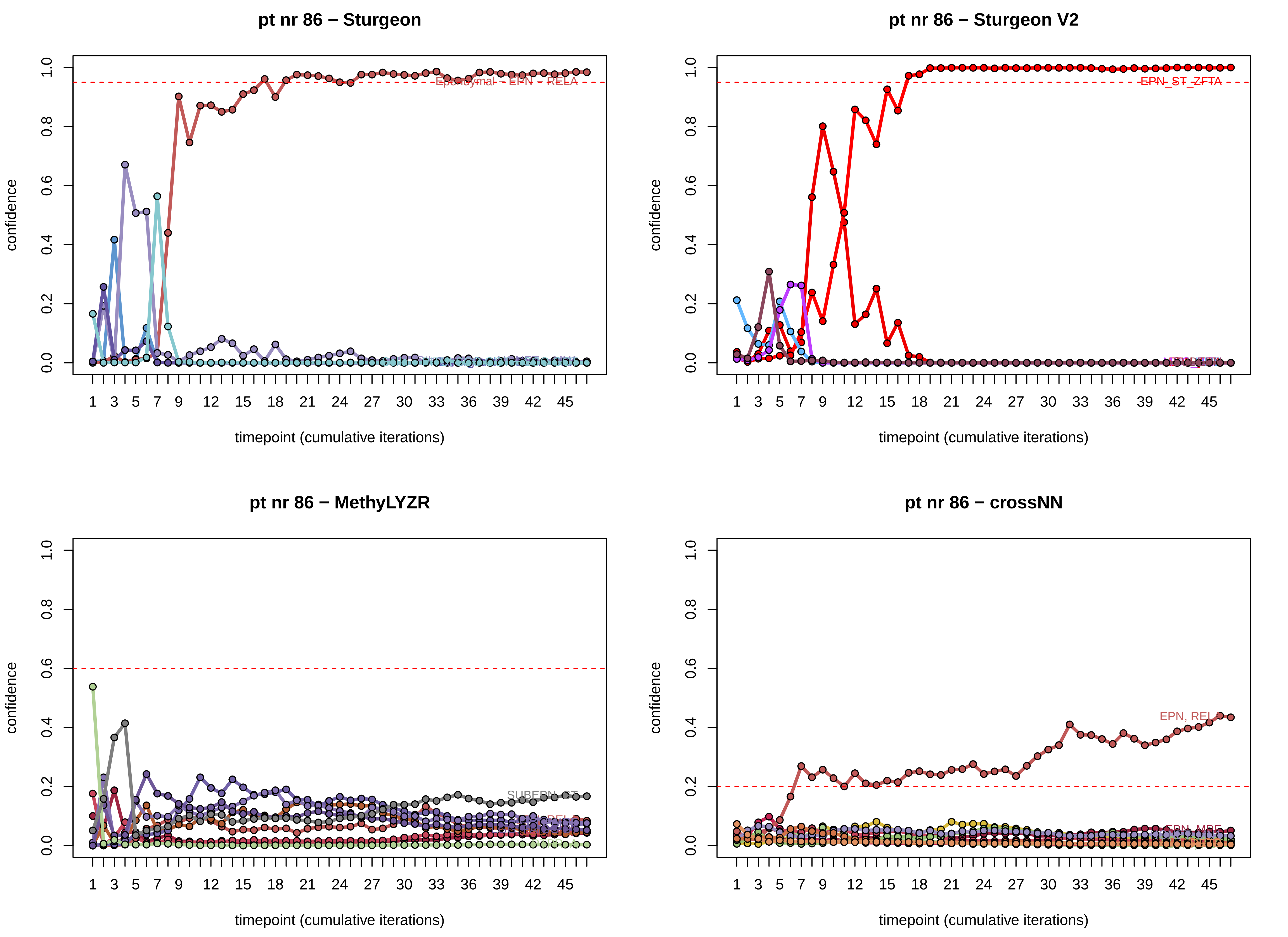
pt nr 86 − Sturgeon
confidence
timepoint (cumulative iterations)
Ependymal - EPN - RELA
pt nr 86 − Sturgeon V2
confidence
timepoint (cumulative iterations)
EPN_ST_ZFTA
pt nr 86 − MethyLYZR
confidence
timepoint (cumulative iterations)
SUBEPN_ST
pt nr 86 − crossNN
confidence
timepoint (cumulative iterations)
EPN, RELA
EPN, MPE

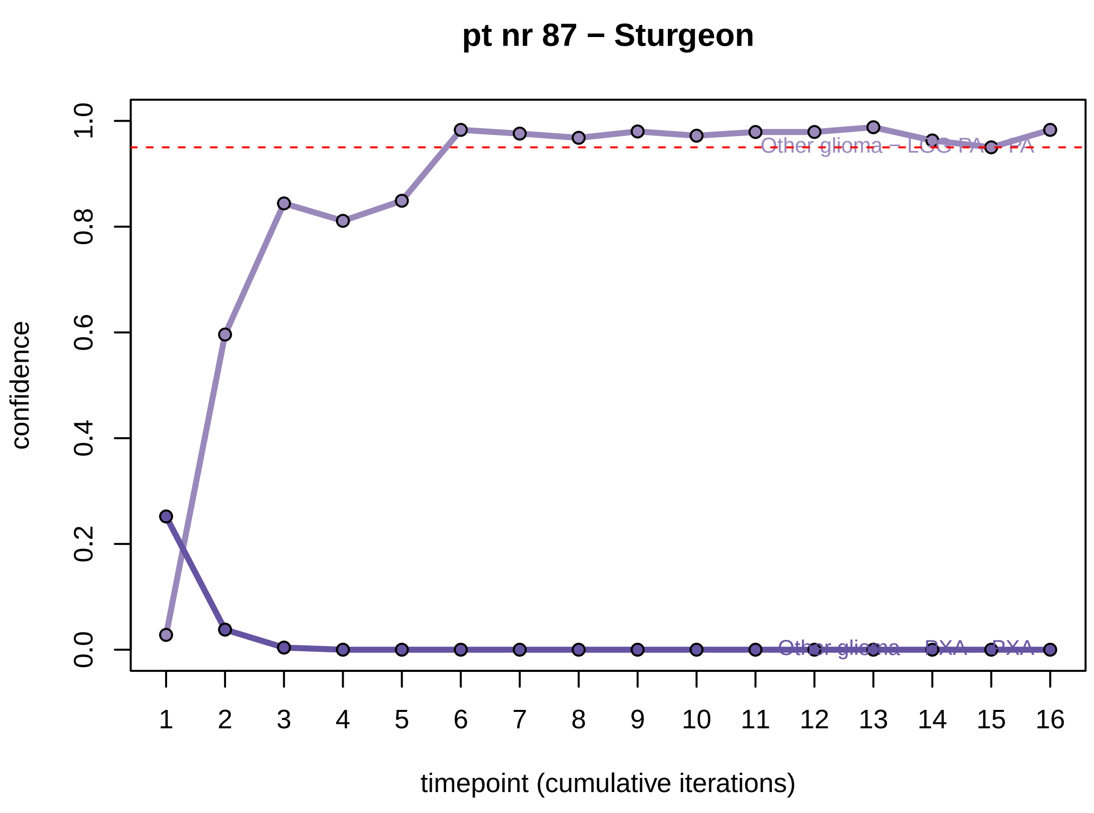
pt nr 87 − Sturgeon
confidence
0.0
0.2
0.4
0.6
0.8
1.0
Other glioma – LGG PA – PA
Other glioma – PXA – PXA
1 2 3 4 5 6 7 8 9 10 11 12 13 14 15 16
timepoint (cumulative iterations)

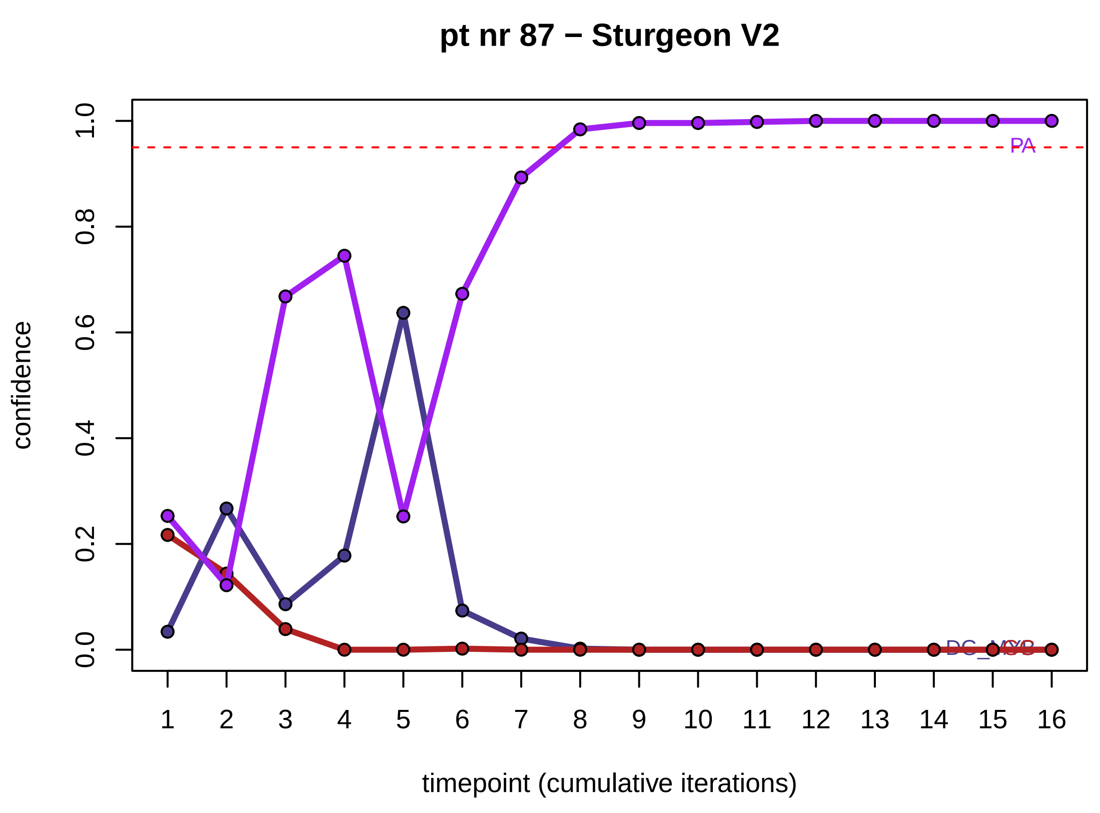
pt nr 87 − Sturgeon V2
confidence
0.0
0.2
0.4
0.6
0.8
1.0
PA
1 2 3 4 5 6 7 8 9 10 11 12 13 14 15 16
timepoint (cumulative iterations)

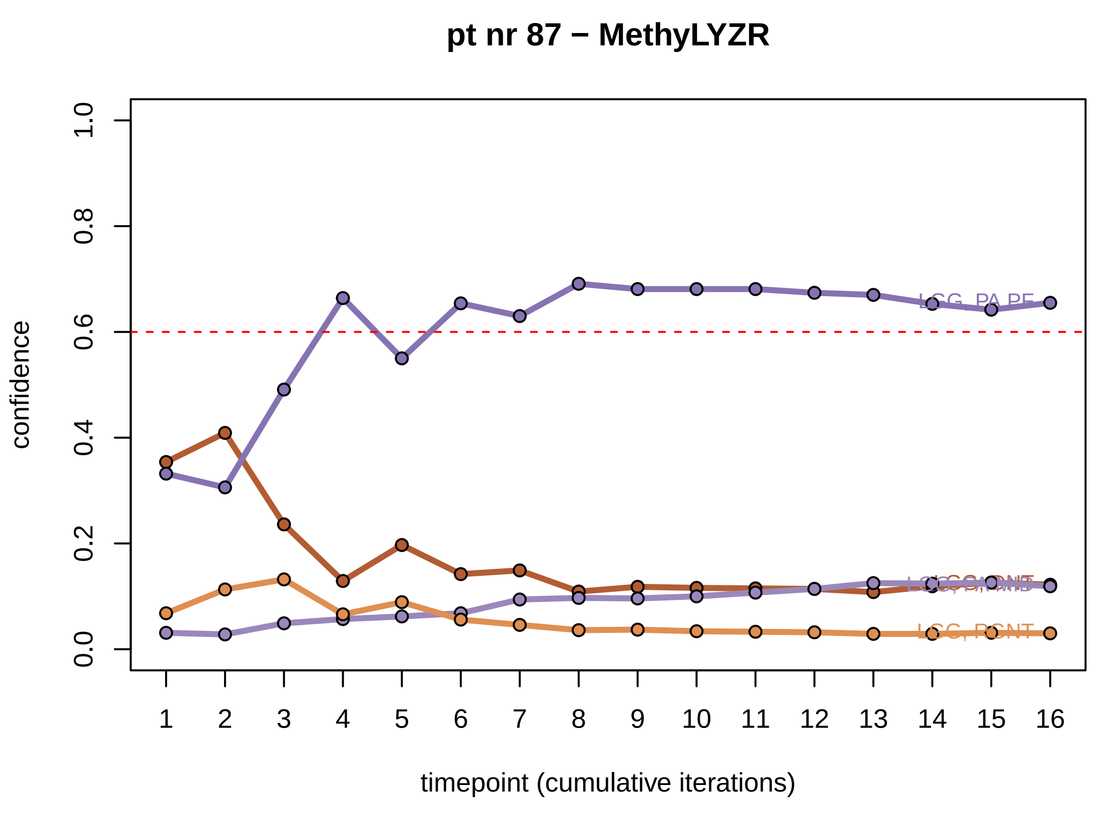
pt nr 87 − MethyLYZR
confidence
0.0
0.2
0.4
0.6
0.8
1.0
LGG, PA PF
LGG, RGNT
1 2 3 4 5 6 7 8 9 10 11 12 13 14 15 16
timepoint (cumulative iterations)

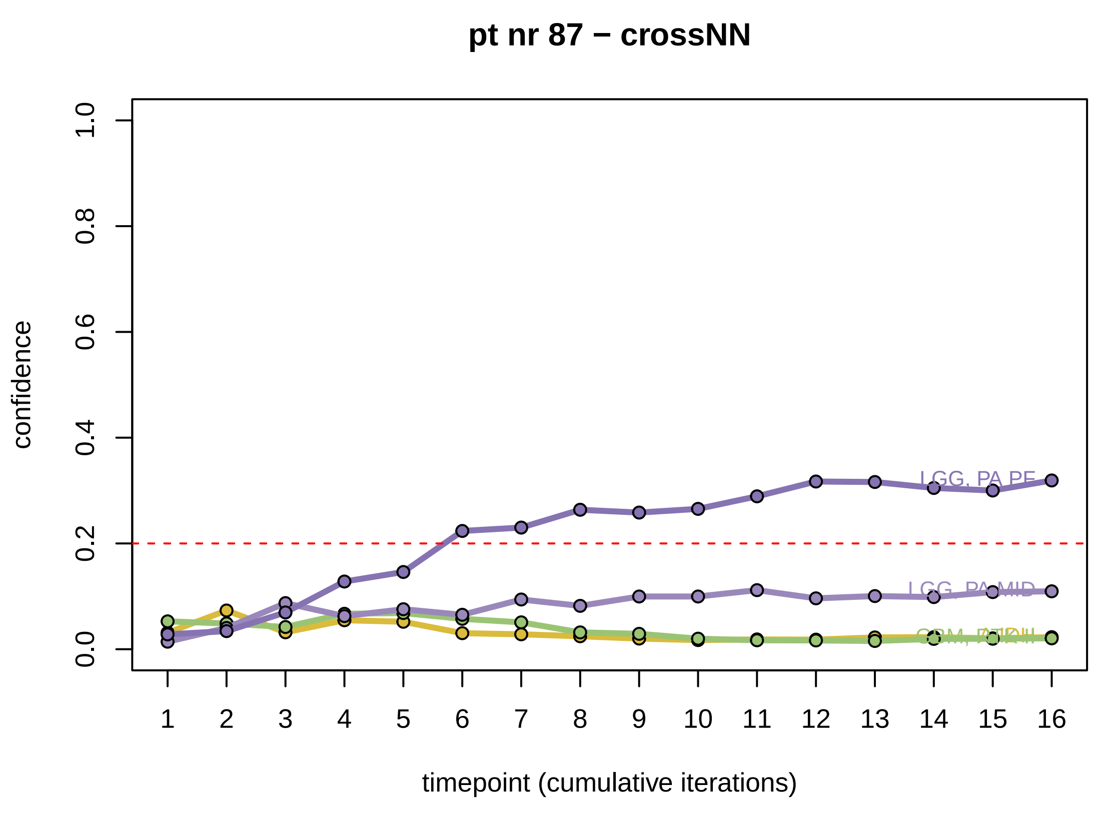
pt nr 87 − crossNN
confidence
0.0
0.2
0.4
0.6
0.8
1.0
LGG, PA PF
LGG, PA MID
1 2 3 4 5 6 7 8 9 10 11 12 13 14 15 16
timepoint (cumulative iterations)

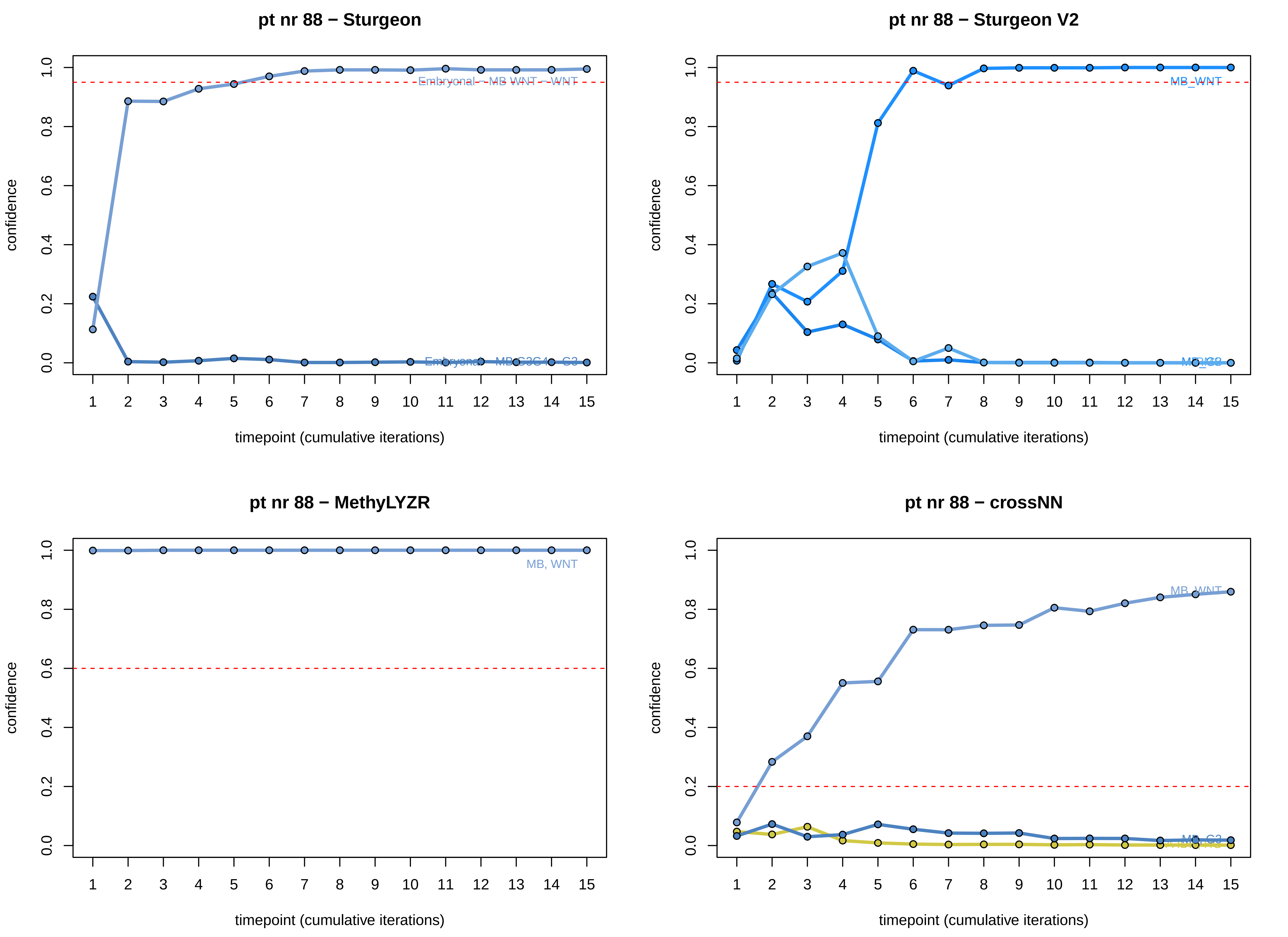
pt nr 88 − Sturgeon
pt nr 88 − Sturgeon V2
pt nr 88 − MethyLYZR
pt nr 88 − crossNN
confidence
timepoint (cumulative iterations)
0.0
0.2
0.4
0.6
0.8
1.0
1 2 3 4 5 6 7 8 9 10 11 12 13 14 15
Embryonal – MB WNT – WNT
Embryonal – MB G3G4 – G3
MB_WNT
MB, WNT
MB, G3

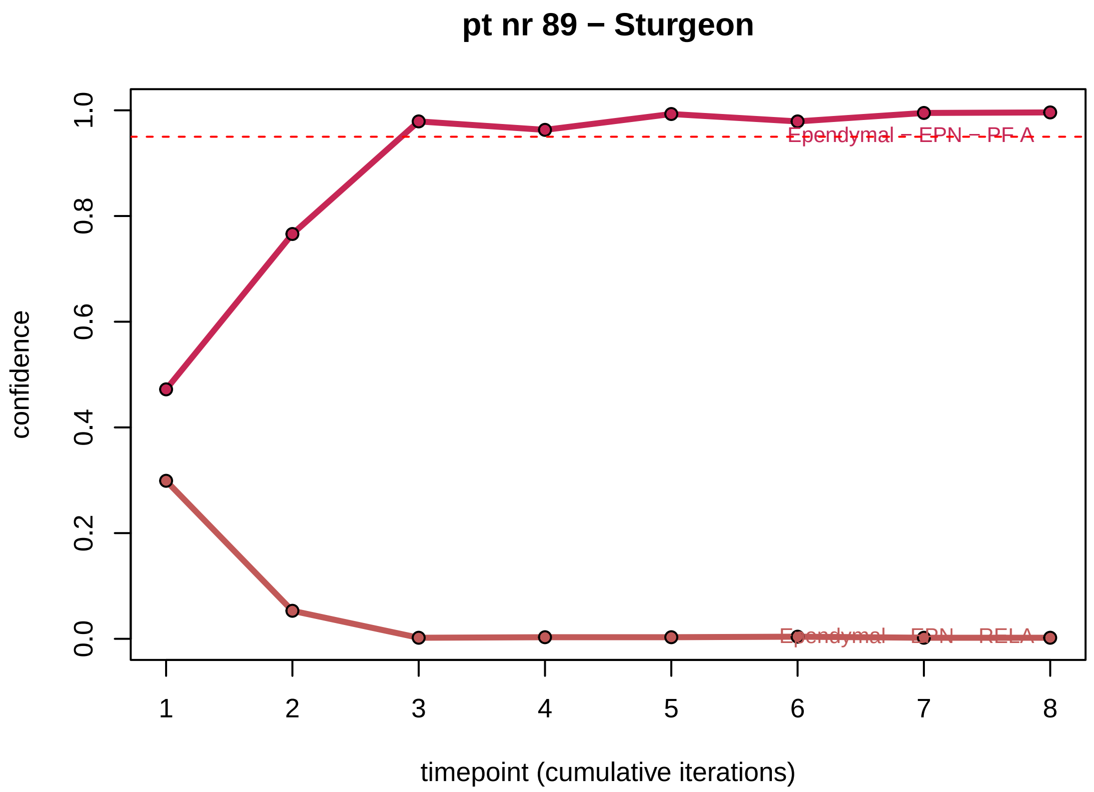
pt nr 89 − Sturgeon
confidence
0.0
0.2
0.4
0.6
0.8
1.0
Ependymal - EPN - PF A
Ependymal - EPN - RELA
1
2
3
4
5
6
7
8
timepoint (cumulative iterations)

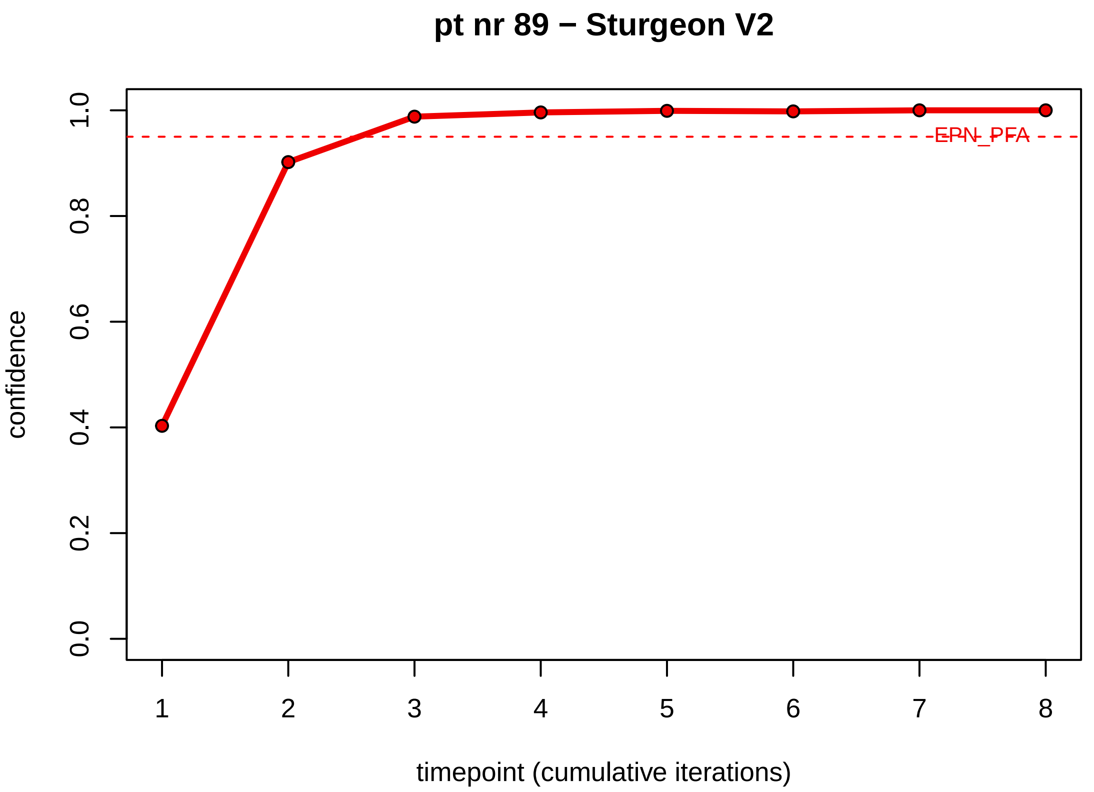
pt nr 89 − Sturgeon V2
confidence
0.0
0.2
0.4
0.6
0.8
1.0
EPN_PFA
1
2
3
4
5
6
7
8
timepoint (cumulative iterations)

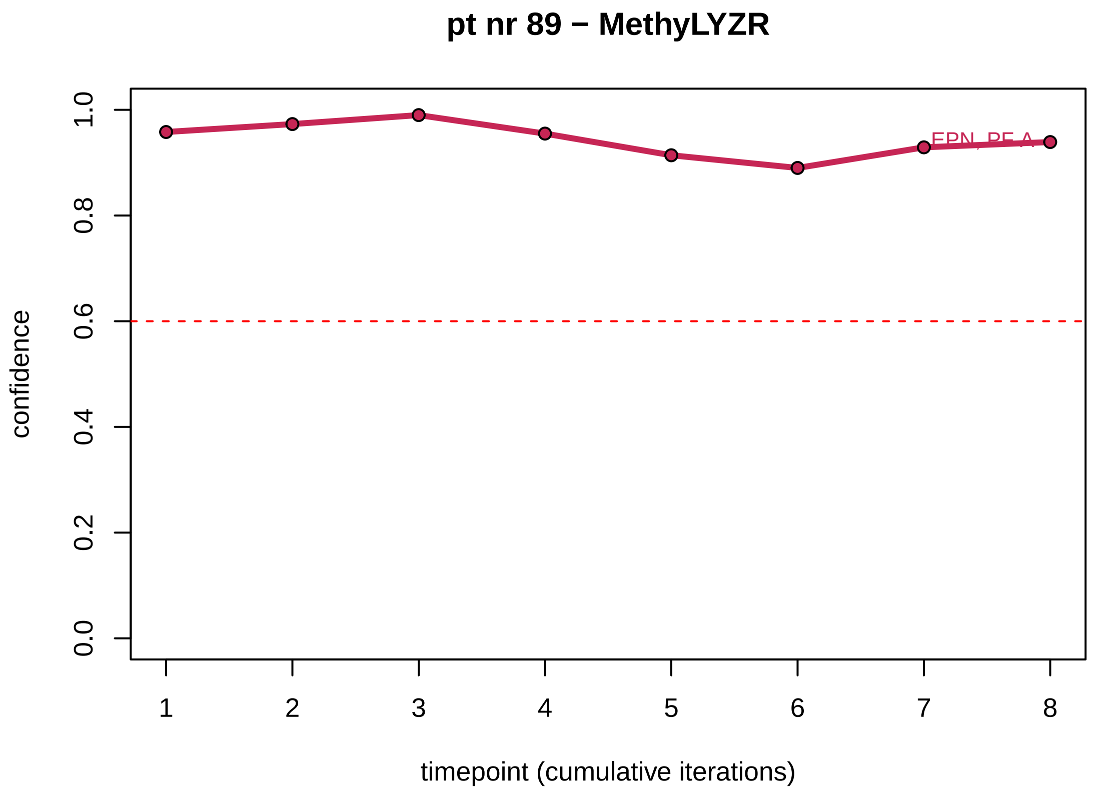
pt nr 89 − MethyLYZR
confidence
0.0
0.2
0.4
0.6
0.8
1.0
EPN_PF A
1
2
3
4
5
6
7
8
timepoint (cumulative iterations)

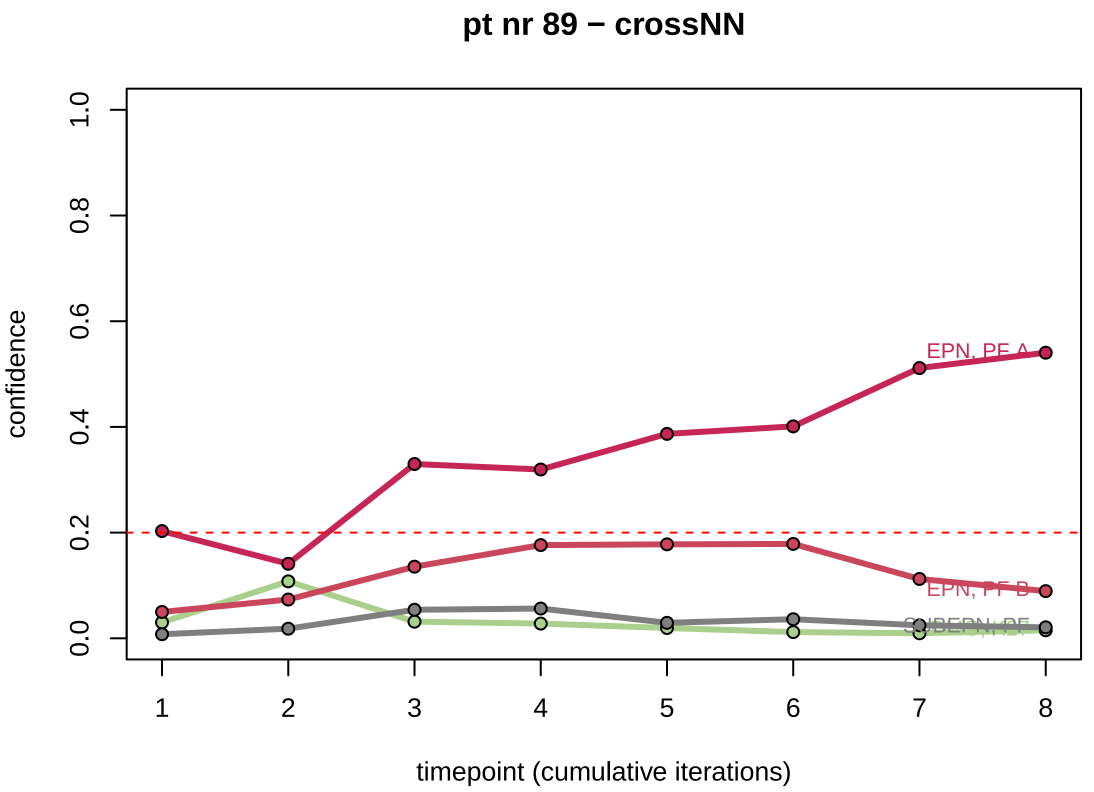
pt nr 89 − crossNN
confidence
0.0
0.2
0.4
0.6
0.8
1.0
EPN, PF A
EPN, PF B
SUBEPN, PF
1
2
3
4
5
6
7
8
timepoint (cumulative iterations)

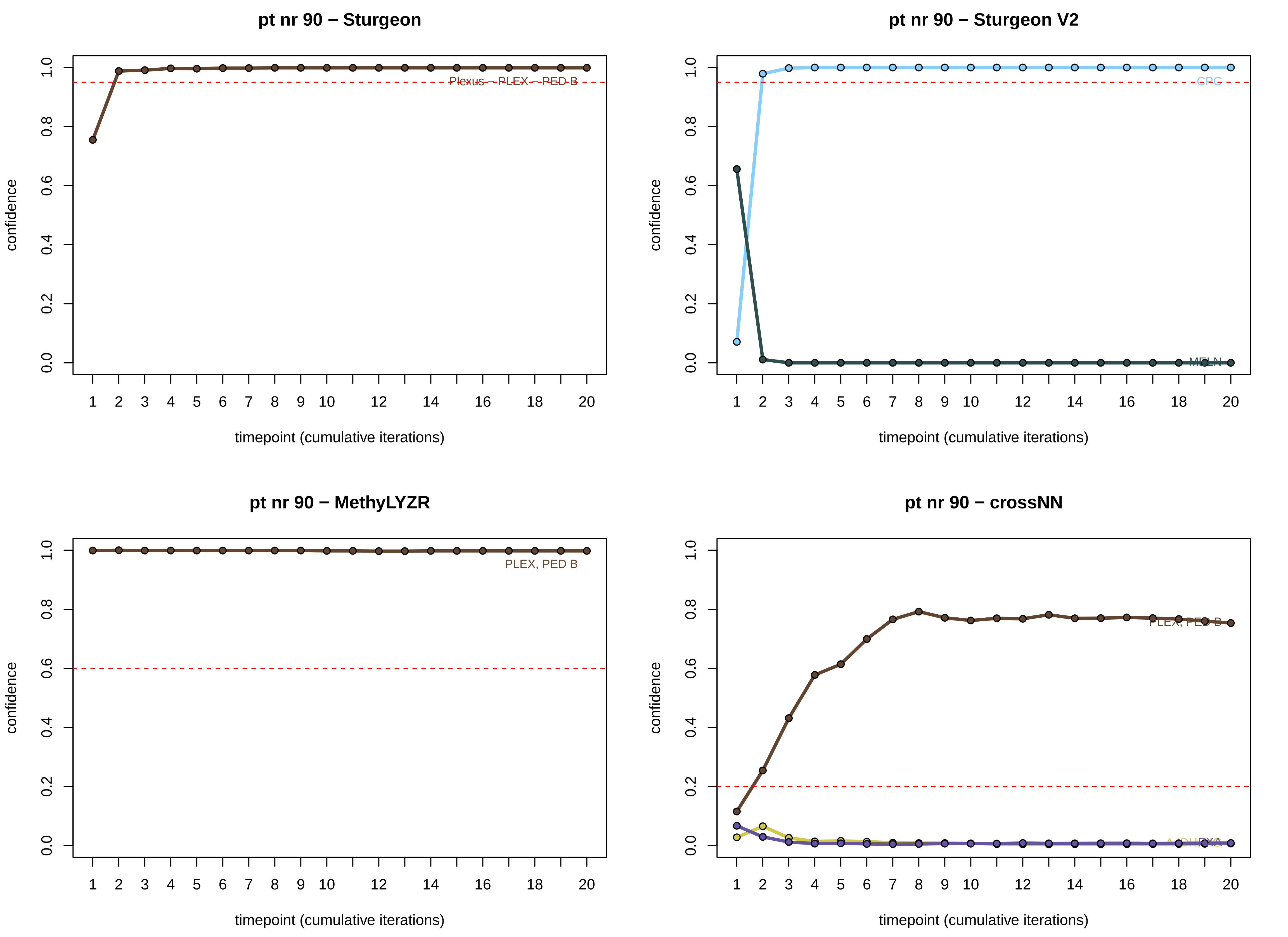

pt nr 90 − Sturgeon
Plexus − PLEX − PED B
pt nr 90 − Sturgeon V2
CPG
MELN
pt nr 90 − MethyLYZR
PLEX, PED B
pt nr 90 − crossNN
confidence
timepoint (cumulative iterations)
0.0
0.2
0.4
0.6
0.8
1.0
1
2
3
4
5
6
7
8
9
10
12
14
16
18
20

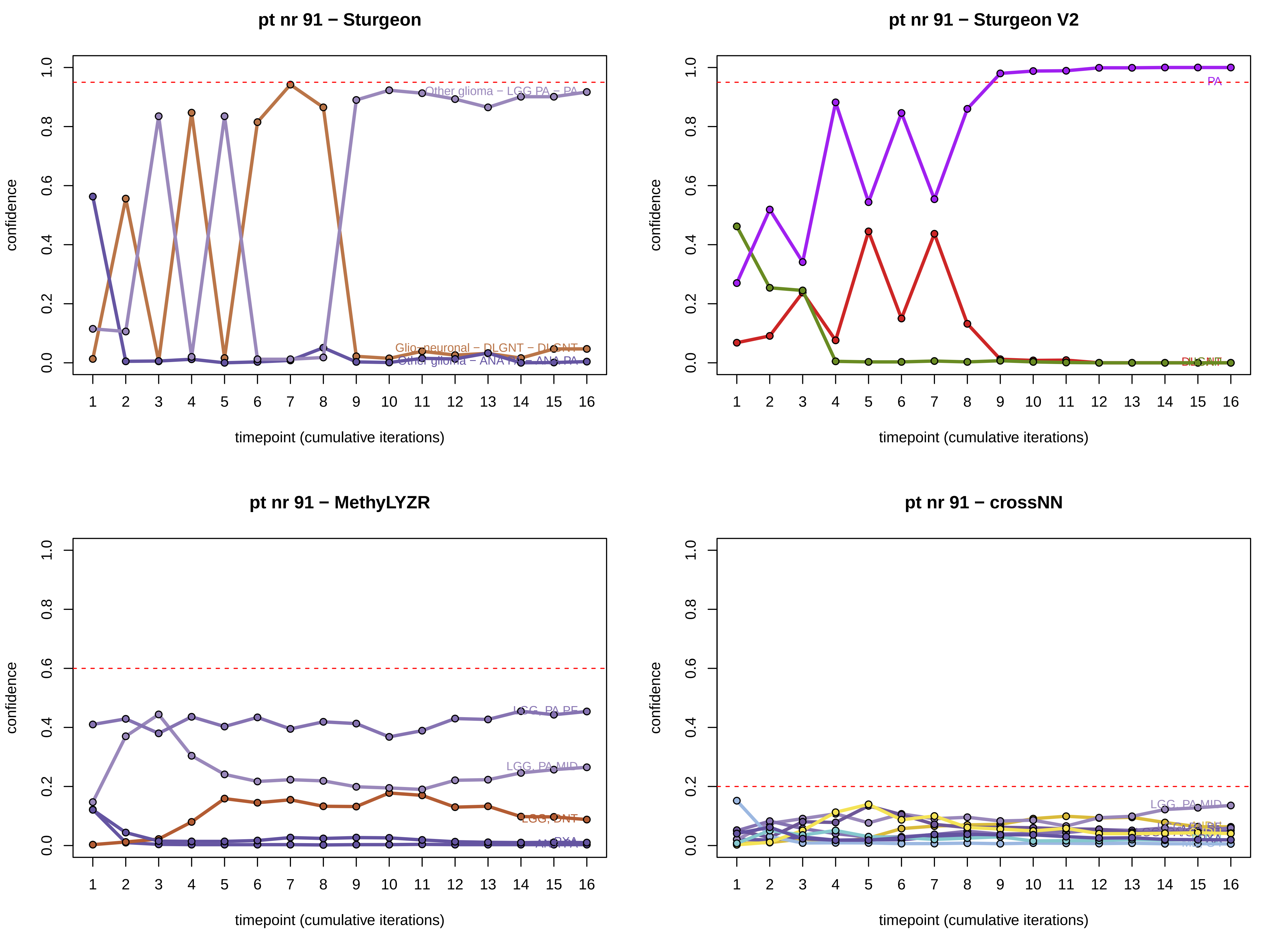
pt nr 91 − Sturgeon
confidence
timepoint (cumulative iterations)
Other glioma − LGG PA − PA
Glio−neuronal − DLGNT − DLGNT
pt nr 91 − Sturgeon V2
confidence
timepoint (cumulative iterations)
PA
pt nr 91 − MethyLYZR
confidence
timepoint (cumulative iterations)
LGG, PA PF
LGG, PA MID
LGG, DNT
pt nr 91 − crossNN
confidence
timepoint (cumulative iterations)
LGG, PA MID

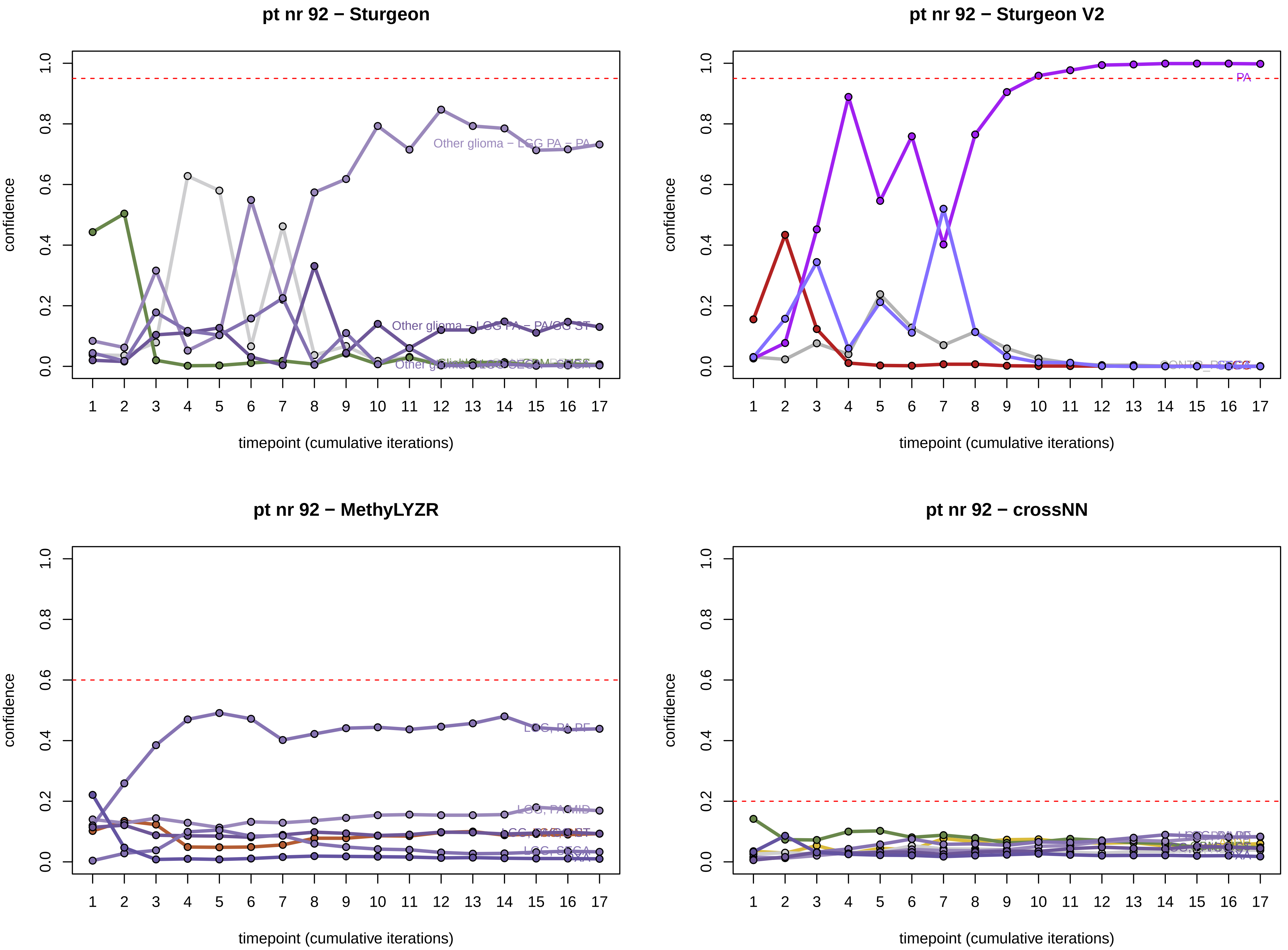
pt nr 92 − Sturgeon
Other glioma − LGG PA − PA
Other glioma − LGG PA − PA/GG ST
confidence
timepoint (cumulative iterations)
pt nr 92 − Sturgeon V2
PA
confidence
timepoint (cumulative iterations)
pt nr 92 − MethyLYZR
LGG, PA PF
LGG, PA MID
confidence
timepoint (cumulative iterations)
pt nr 92 − crossNN
confidence
timepoint (cumulative iterations)

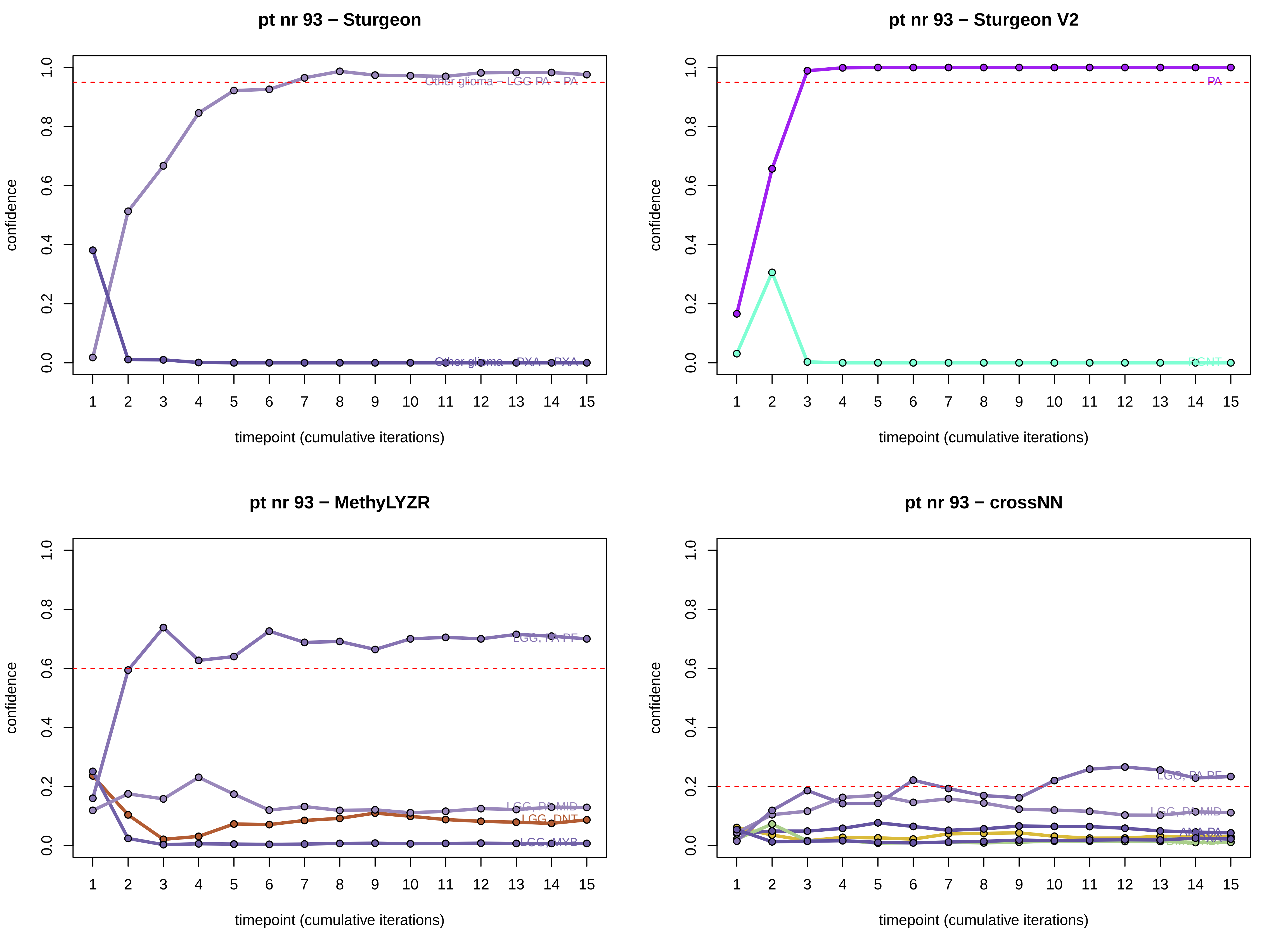
pt nr 93 − Sturgeon
Other glioma − LGG PA − PA
Other glioma − PXA − PXA
pt nr 93 − Sturgeon V2
PA
DGNT
pt nr 93 − MethyLYZR
LGG, PA PF
LGG, PA MID
LGG, DNT
LGG, MYB
pt nr 93 − crossNN
LGG, PA PF
LGG, PA MID
ANA PA
confidence
timepoint (cumulative iterations)

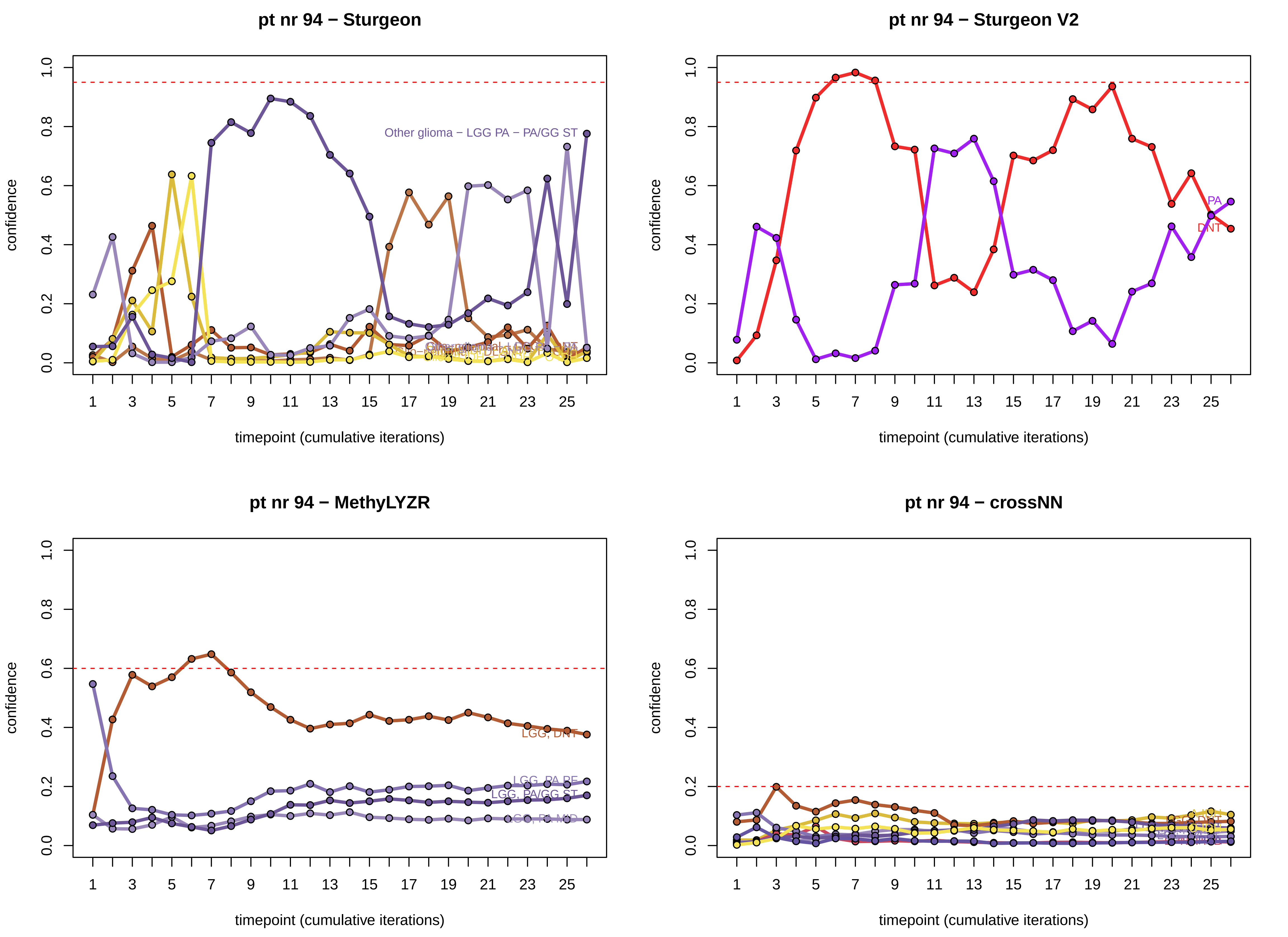
pt nr 94 − Sturgeon
Other glioma − LGG PA − PA/GG ST
confidence
timepoint (cumulative iterations)
pt nr 94 − Sturgeon V2
PA
DNT
confidence
timepoint (cumulative iterations)
pt nr 94 − MethyLYZR
LGG, DNT
LGG, PA PF
LGG, PA/GG ST
LGG, PA MID
confidence
timepoint (cumulative iterations)
pt nr 94 − crossNN
confidence
timepoint (cumulative iterations)
