## Supplementary data 2 for "Sturgeon V2: a clinically applicable CNS tumor classifier trained on relabeled data"

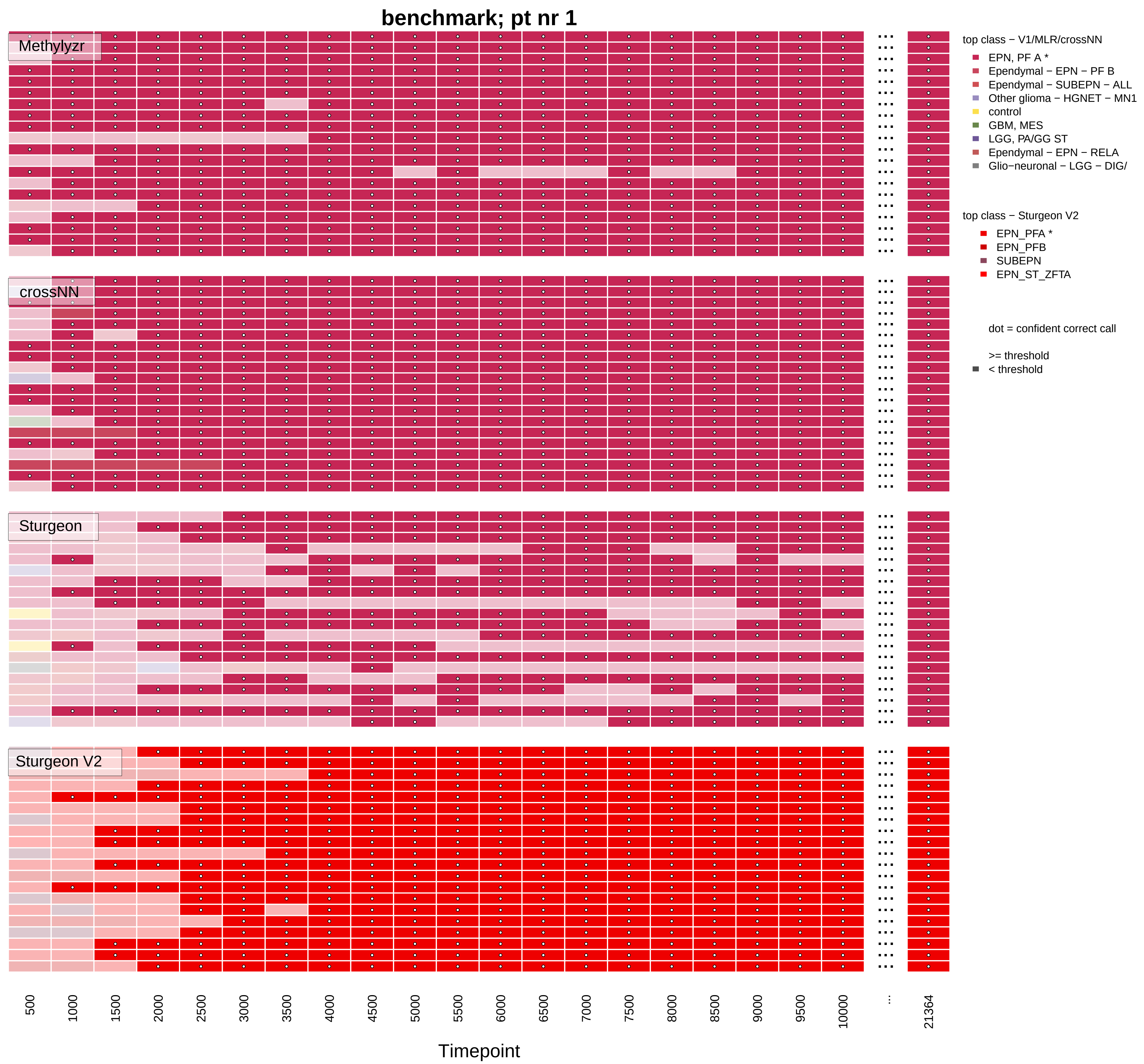

benchmark; pt nr 1
Methylyzr
crossNN
Sturgeon
Sturgeon V2
top class − V1/MLR/crossNN
EPN, PF A *
Ependymal − EPN − PF B
Ependymal − SUBEPN − ALL
Other glioma − HGNET − MN1
control
GBM, MES
LGG, PA/GG ST
Ependymal − EPN − RELA
Glio−neuronal − LGG − DIG/
top class − Sturgeon V2
EPN_PFA *
EPN_PFB
SUBEPN
EPN_ST_ZFTA
dot = confident correct call
>= threshold
< threshold
500
1000
1500
2000
2500
3000
3500
4000
4500
5000
5500
6000
6500
7000
7500
8000
8500
9000
9500
10000
...
21364
Timepoint

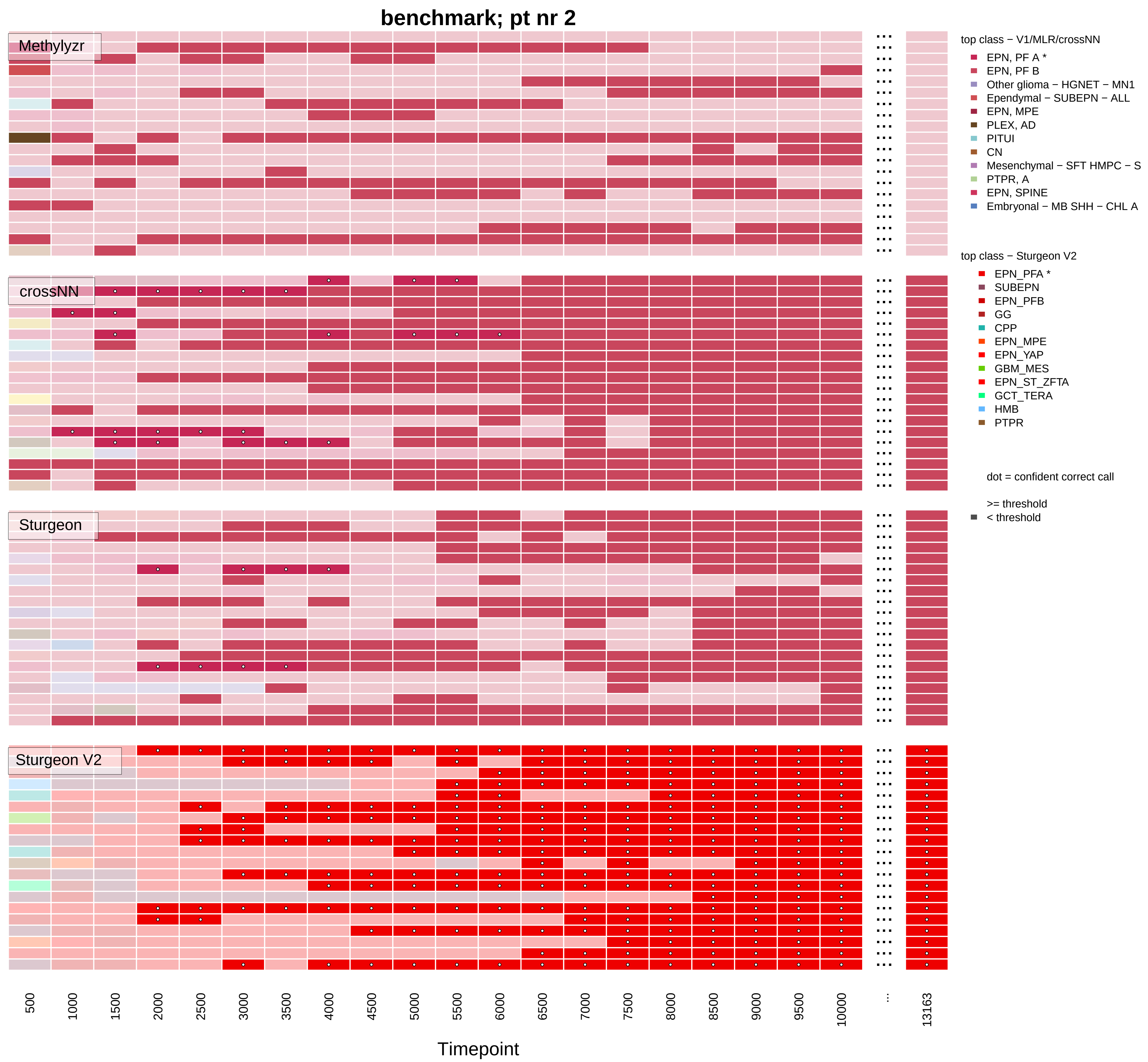

benchmark; pt nr 2
Methylyzr
crossNN
Sturgeon
Sturgeon V2
top class − V1/MLR/crossNN
EPN, PF A *
EPN, PF B
Other glioma − HGNET − MN1
Ependymal − SUBEPN − ALL
EPN, MPE
PLEX, AD
PITUI
CN
Mesenchymal − SFT HMPC − S
PTPR, A
EPN, SPINE
Embryonal − MB SHH − CHL A
top class − Sturgeon V2
EPN_PFA *
SUBEPN
EPN_PFB
GG
CPP
EPN_MPE
EPN_YAP
GBM_MES
EPN_ST_ZFTA
GCT_TERA
HMB
PTPR
dot = confident correct call
>= threshold
< threshold
500
1000
1500
2000
2500
3000
3500
4000
4500
5000
5500
6000
6500
7000
7500
8000
8500
9000
9500
10000
...
13163
Timepoint

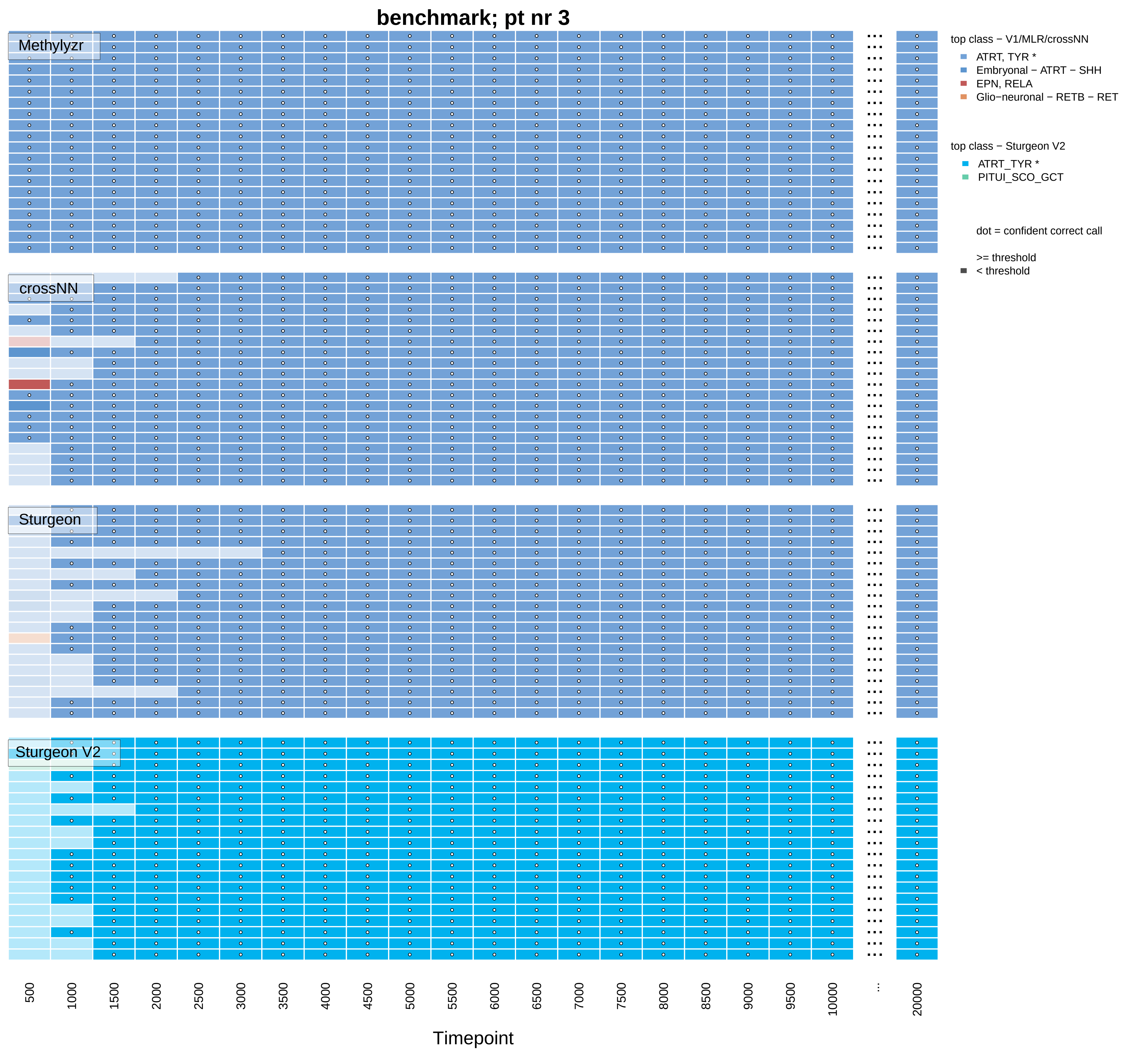

benchmark; pt nr 3
Methylyzr
crossNN
Sturgeon
Sturgeon V2
top class − V1/MLR/crossNN
ATRT, TYR *
Embryonal − ATRT − SHH
EPN, RELA
Glio−neuronal − RETB − RET
top class − Sturgeon V2
ATRT_TYR *
PITUI_SCO_GCT
dot = confident correct call
>= threshold
< threshold
500
1000
1500
2000
2500
3000
3500
4000
4500
5000
5500
6000
6500
7000
7500
8000
8500
9000
9500
10000
...
20000
Timepoint

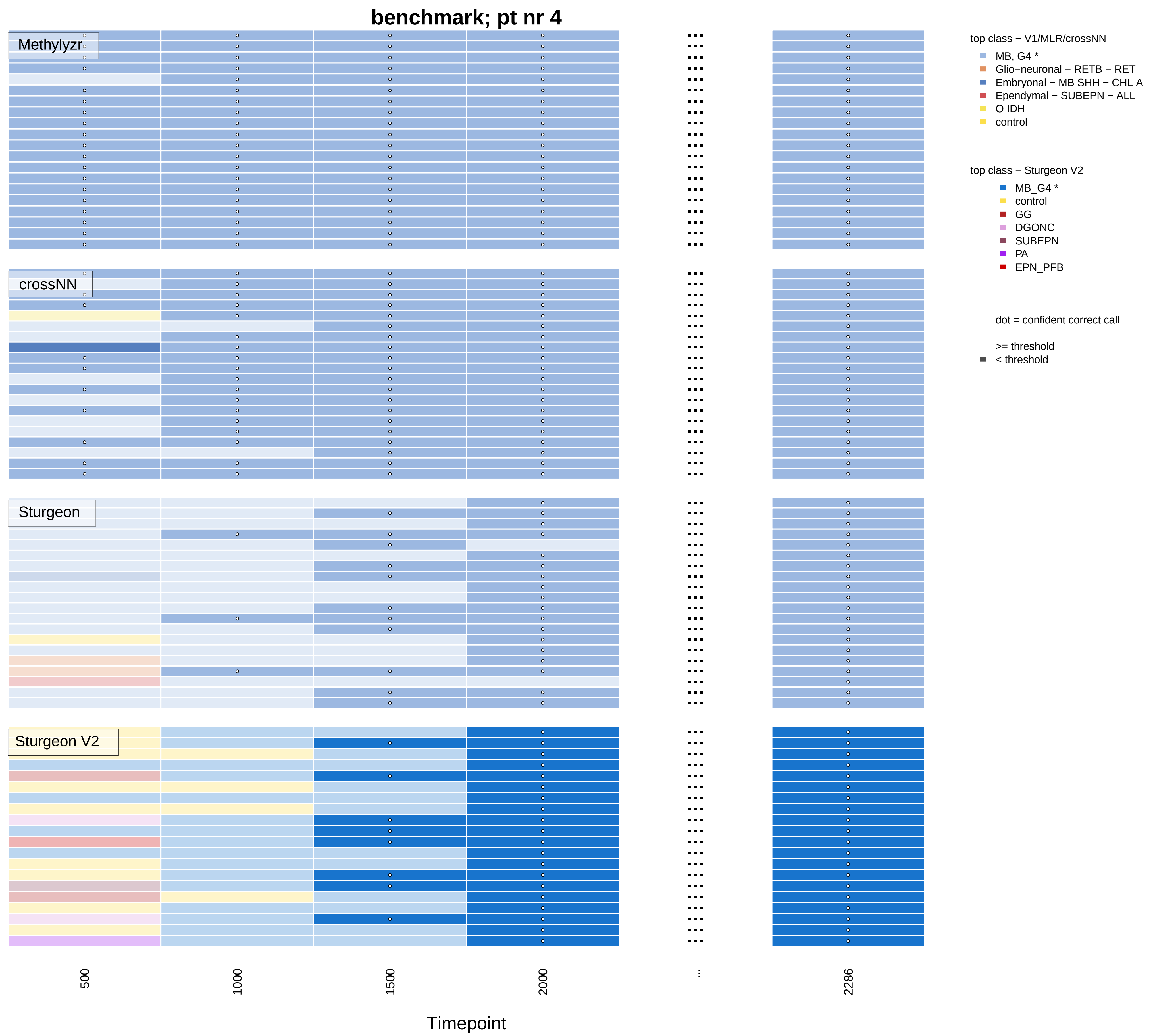

benchmark; pt nr 4
Methylyzr
crossNN
Sturgeon
Sturgeon V2
top class − V1/MLR/crossNN
MB, G4 *
Glio−neuronal − RETB − RET
Embryonal − MB SHH − CHL A
Ependymal − SUBEPN − ALL
O IDH
control
top class − Sturgeon V2
MB_G4 *
control
GG
DGONC
SUBEPN
PA
EPN_PFB
dot = confident correct call
>= threshold
< threshold
500
1000
1500
2000
...
2286
Timepoint

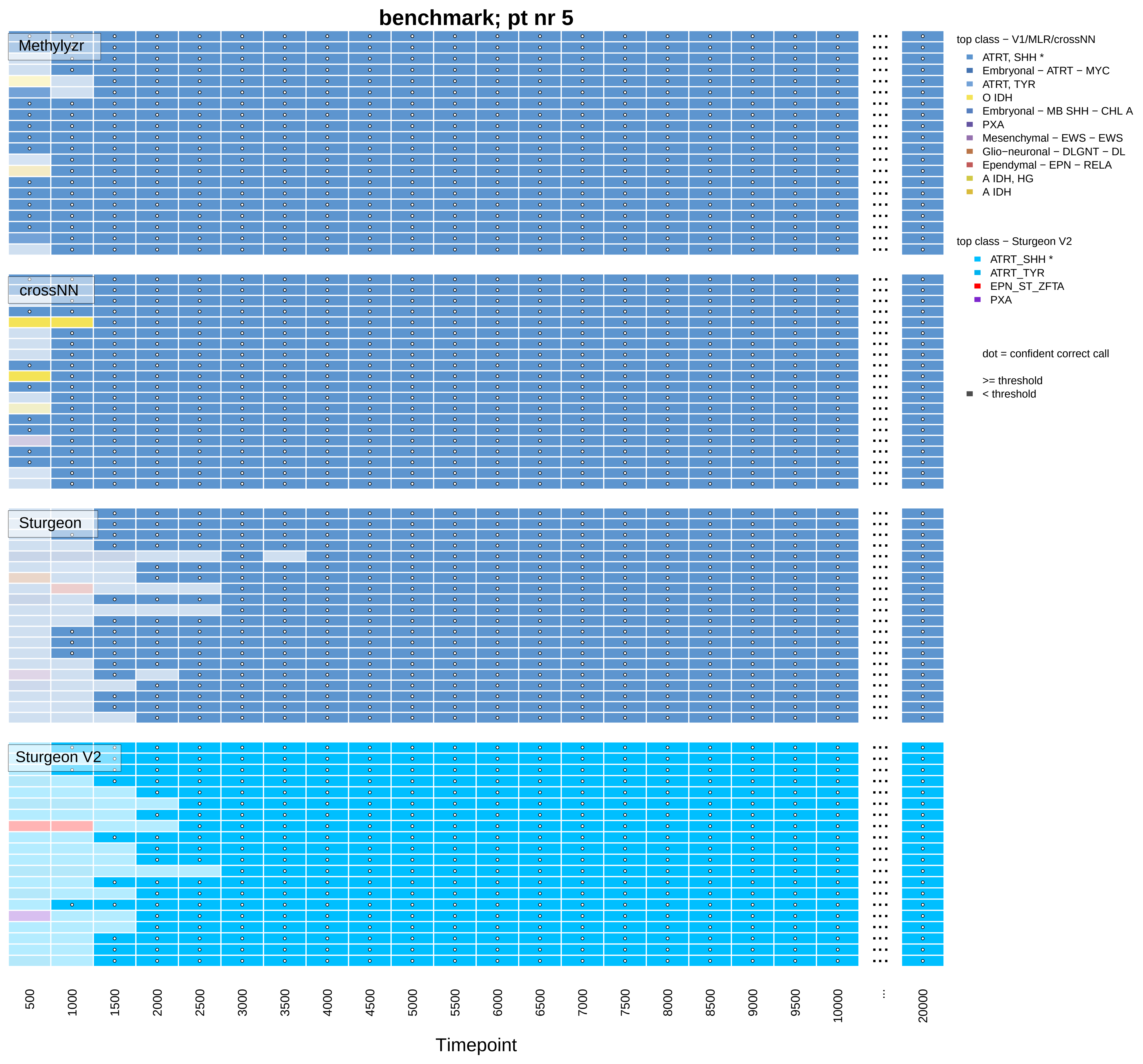
benchmark; pt nr 5
Methylyzr
crossNN
Sturgeon
Sturgeon V2
top class − V1/MLR/crossNN
ATRT, SHH *
Embryonal − ATRT − MYC
ATRT, TYR
O IDH
Embryonal − MB SHH − CHL A
PXA
Mesenchymal − EWS − EWS
Glio−neuronal − DLGNT − DL
Ependymal − EPN − RELA
A IDH, HG
A IDH
top class − Sturgeon V2
ATRT_SHH *
ATRT_TYR
EPN_ST_ZFTA
PXA
dot = confident correct call
>= threshold
< threshold
500
1000
1500
2000
2500
3000
3500
4000
4500
5000
5500
6000
6500
7000
7500
8000
8500
9000
9500
10000
...
20000
Timepoint

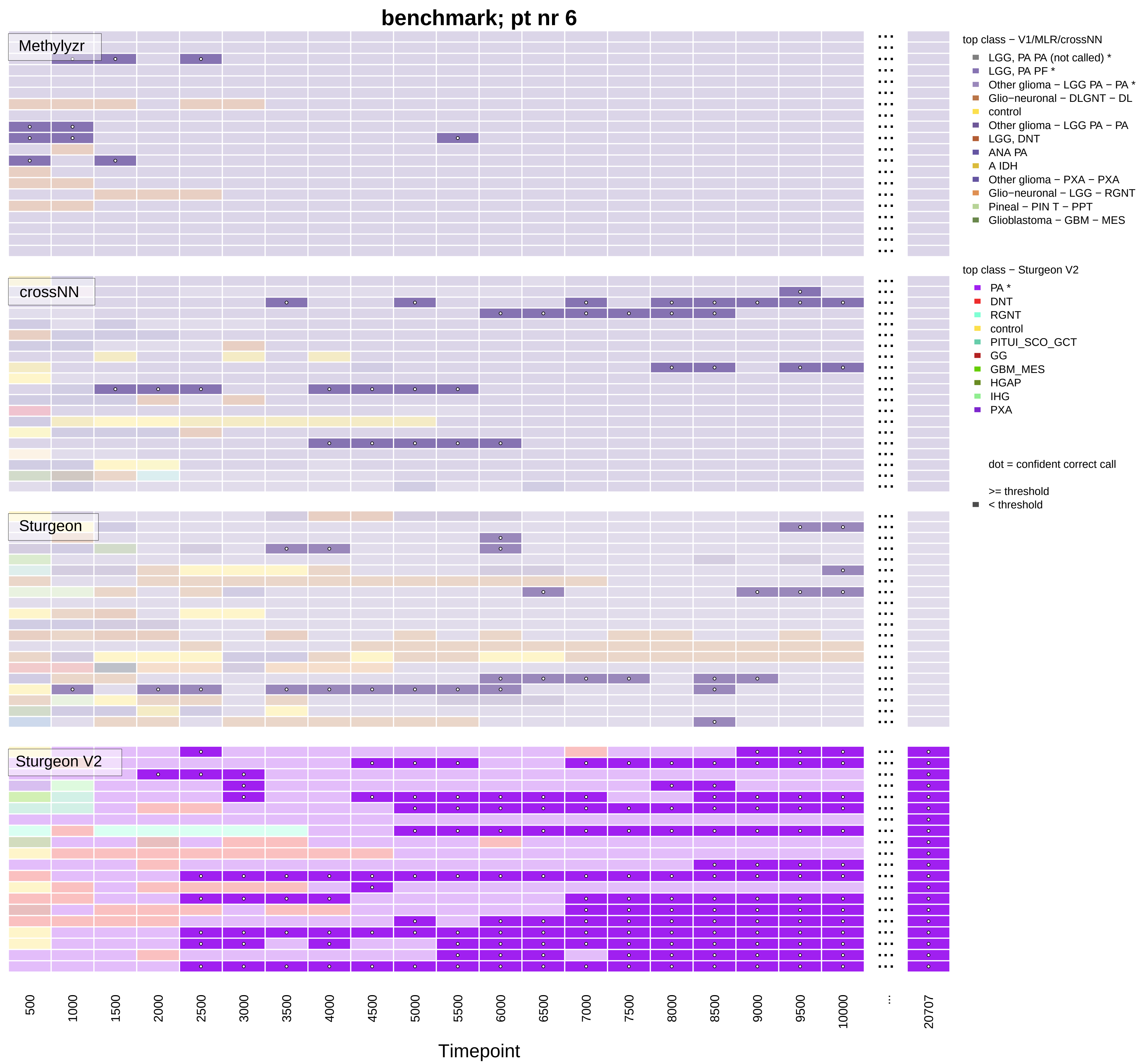

benchmark; pt nr 6
Methylyzr
crossNN
Sturgeon
Sturgeon V2
top class − V1/MLR/crossNN
LGG, PA PA (not called) *
LGG, PA PF *
Other glioma − LGG PA − PA *
Glio−neuronal − DLGNT − DL
control
Other glioma − LGG PA − PA
LGG, DNT
ANA PA
A IDH
Other glioma − PXA − PXA
Glio−neuronal − LGG − RGNT
Pineal − PIN T − PPT
Glioblastoma − GBM − MES
top class − Sturgeon V2
PA *
DNT
RGNT
control
PITUI_SCO_GCT
GG
GBM_MES
HGAP
IHG
PXA
dot = confident correct call
>= threshold
< threshold
500
1000
1500
2000
2500
3000
3500
4000
4500
5000
5500
6000
6500
7000
7500
8000
8500
9000
9500
10000
...
20707
Timepoint

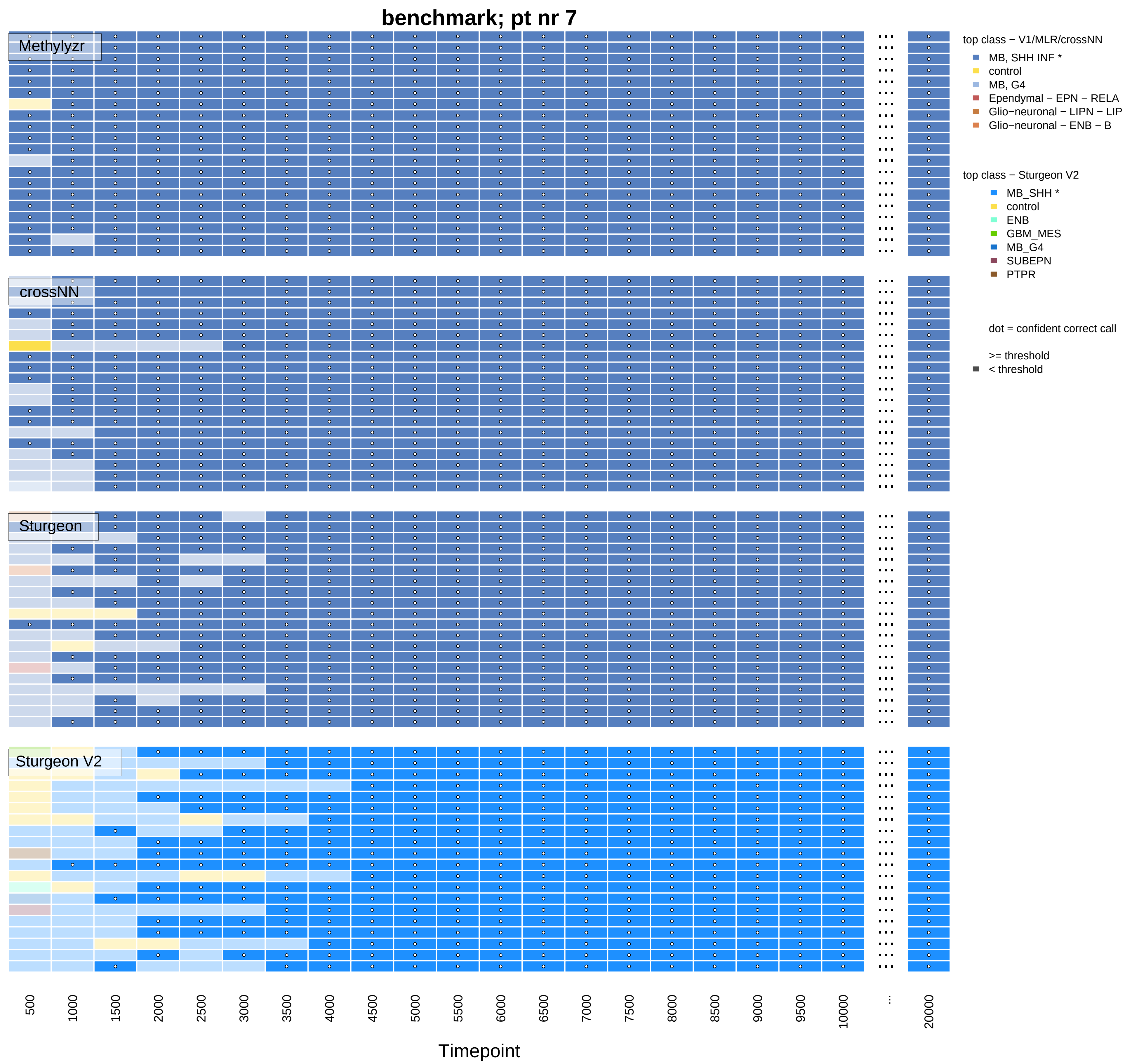
benchmark; pt nr 7
Methylyzr
crossNN
Sturgeon
Sturgeon V2
top class − V1/MLR/crossNN
MB, SHH INF *
control
MB, G4
Ependymal − EPN − RELA
Glio−neuronal − LIPN − LIP
Glio−neuronal − ENB − B
top class − Sturgeon V2
MB_SHH *
control
ENB
GBM_MES
MB_G4
SUBEPN
PTPR
dot = confident correct call
>= threshold
< threshold
500
1000
1500
2000
2500
3000
3500
4000
4500
5000
5500
6000
6500
7000
7500
8000
8500
9000
9500
10000
...
20000
Timepoint

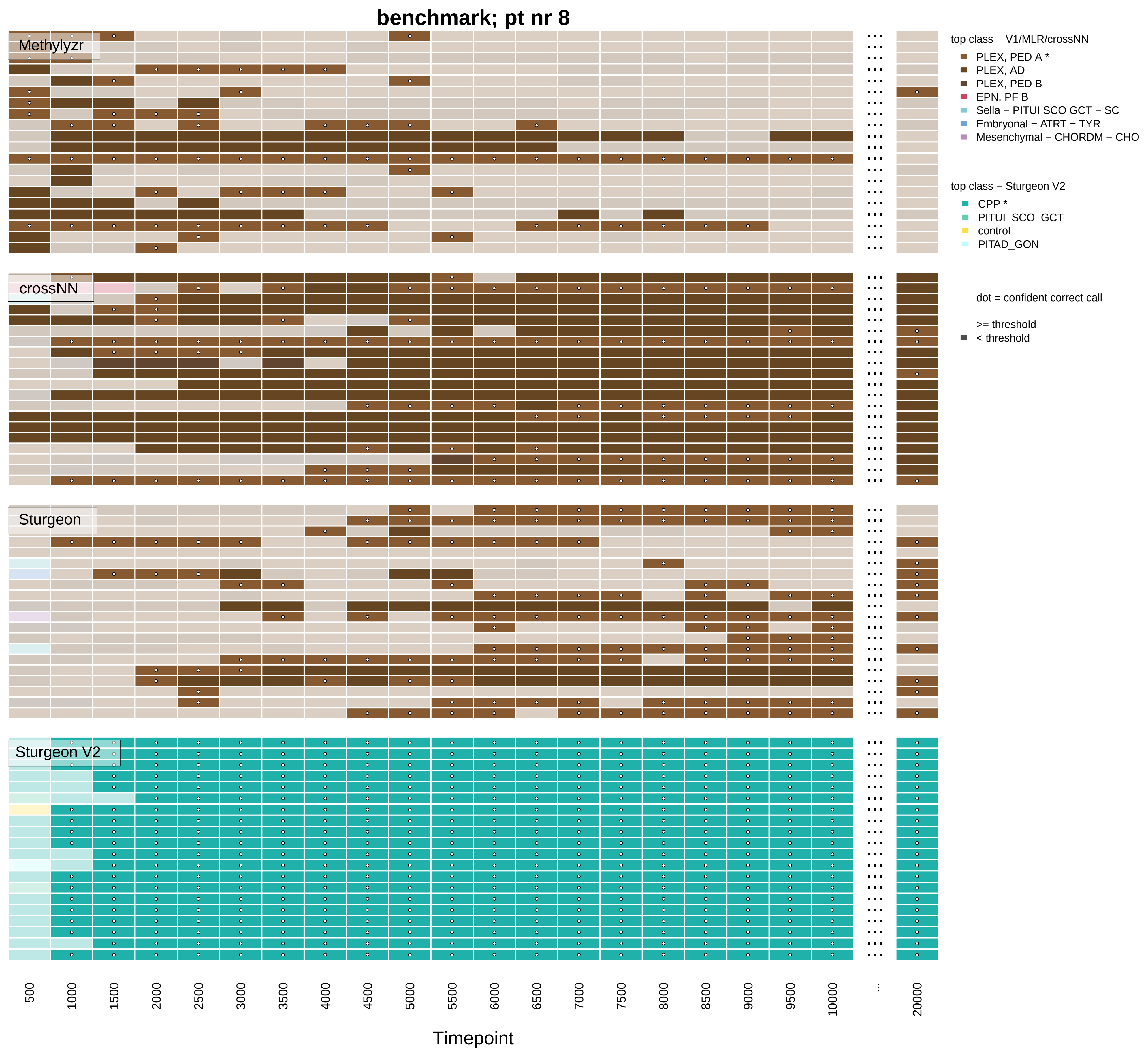

benchmark; pt nr 8
Methylyzr
crossNN
Sturgeon
Sturgeon V2
top class − V1/MLR/crossNN
PLEX, PED A *
PLEX, AD
PLEX, PED B
EPN, PF B
Sella − PITUI SCO GCT − SC
Embryonal − ATRT − TYR
Mesenchymal − CHORDM − CHO
top class − Sturgeon V2
CPP *
PITUI_SCO_GCT
control
PITAD_GON
dot = confident correct call
>= threshold
< threshold
500
1000
1500
2000
2500
3000
3500
4000
4500
5000
5500
6000
6500
7000
7500
8000
8500
9000
9500
10000
...
20000
Timepoint

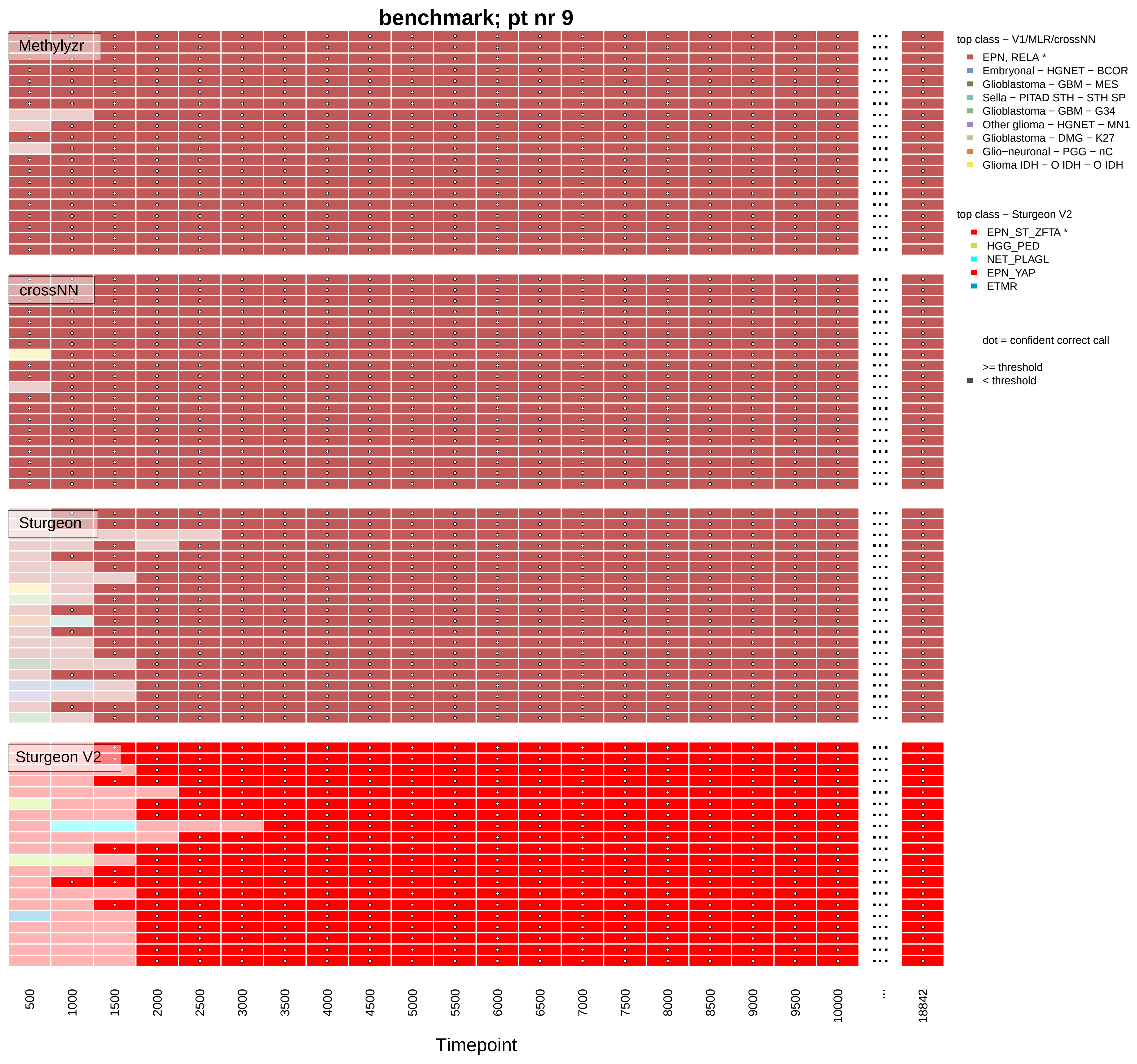
benchmark; pt nr 9
Methylyzr
crossNN
Sturgeon
Sturgeon V2
top class − V1/MLR/crossNN
EPN, RELA *
Embryonal − HGNET − BCOR
Glioblastoma − GBM − MES
Sella − PITAD STH − STH SP
Glioblastoma − GBM − G34
Other glioma − HGNET − MN1
Glioblastoma − DMG − K27
Glio−neuronal − PGG − nC
Glioma IDH − O IDH − O IDH
top class − Sturgeon V2
EPN_ST_ZFTA *
HGG_PED
NET_PLAGL
EPN_YAP
ETMR
dot = confident correct call
>= threshold
< threshold
500
1000
1500
2000
2500
3000
3500
4000
4500
5000
5500
6000
6500
7000
7500
8000
8500
9000
9500
10000
...
18842
Timepoint

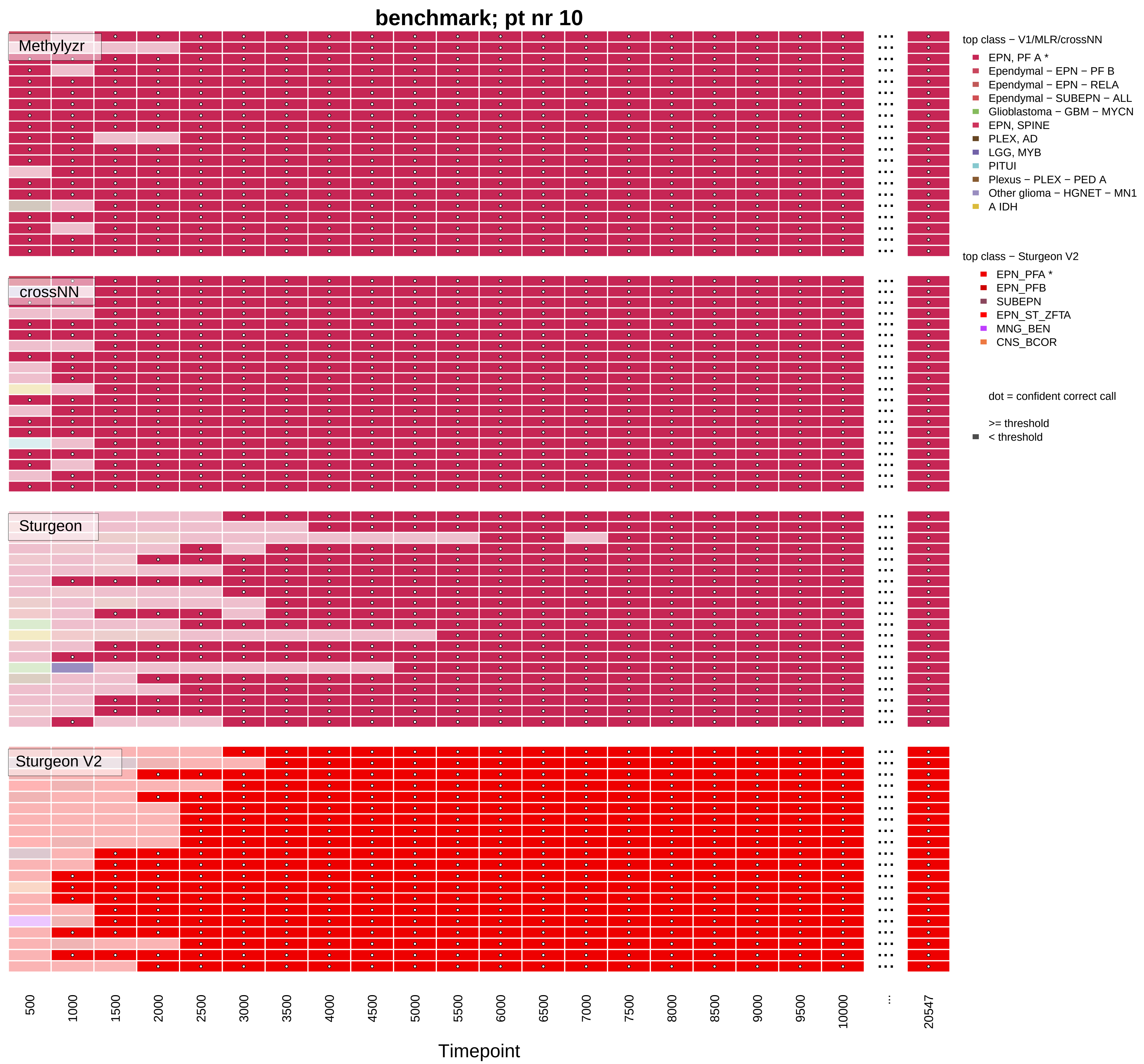
benchmark; pt nr 10
Methylyzr
crossNN
Sturgeon
Sturgeon V2
500
1000
1500
2000
2500
3000
3500
4000
4500
5000
5500
6000
6500
7000
7500
8000
8500
9000
9500
10000
...
20547
Timepoint
top class − V1/MLR/crossNN
EPN, PF A *
Ependymal − EPN − PF B
Ependymal − EPN − RELA
Ependymal − SUBEPN − ALL
Glioblastoma − GBM − MYCN
EPN, SPINE
PLEX, AD
LGG, MYB
PITUI
Plexus − PLEX − PED A
Other glioma − HGNET − MN1
A IDH
top class − Sturgeon V2
EPN_PFA *
EPN_PFB
SUBEPN
EPN_ST_ZFTA
MNG_BEN
CNS_BCOR
dot = confident correct call
>= threshold
< threshold

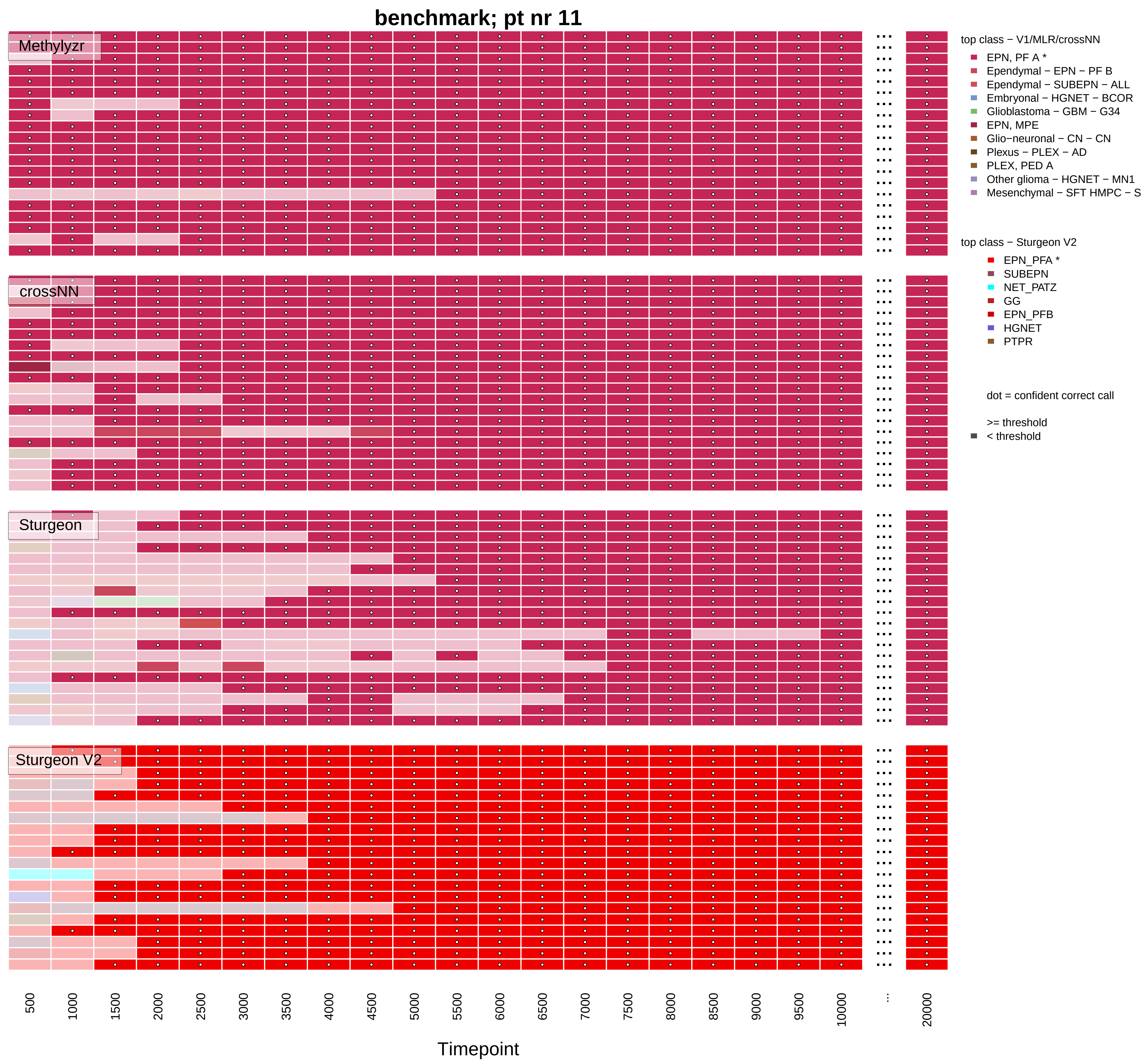

benchmark; pt nr 11
Methylyzr
crossNN
Sturgeon
Sturgeon V2
top class − V1/MLR/crossNN
EPN, PF A *
Ependymal − EPN − PF B
Ependymal − SUBEPN − ALL
Embryonal − HGNET − BCOR
Glioblastoma − GBM − G34
EPN, MPE
Glio−neuronal − CN − CN
Plexus − PLEX − AD
PLEX, PED A
Other glioma − HGNET − MN1
Mesenchymal − SFT HMPC − S
top class − Sturgeon V2
EPN_PFA *
SUBEPN
NET_PATZ
GG
EPN_PFB
HGNET
PTPR
dot = confident correct call
>= threshold
< threshold
500
1000
1500
2000
2500
3000
3500
4000
4500
5000
5500
6000
6500
7000
7500
8000
8500
9000
9500
10000
...
20000
Timepoint

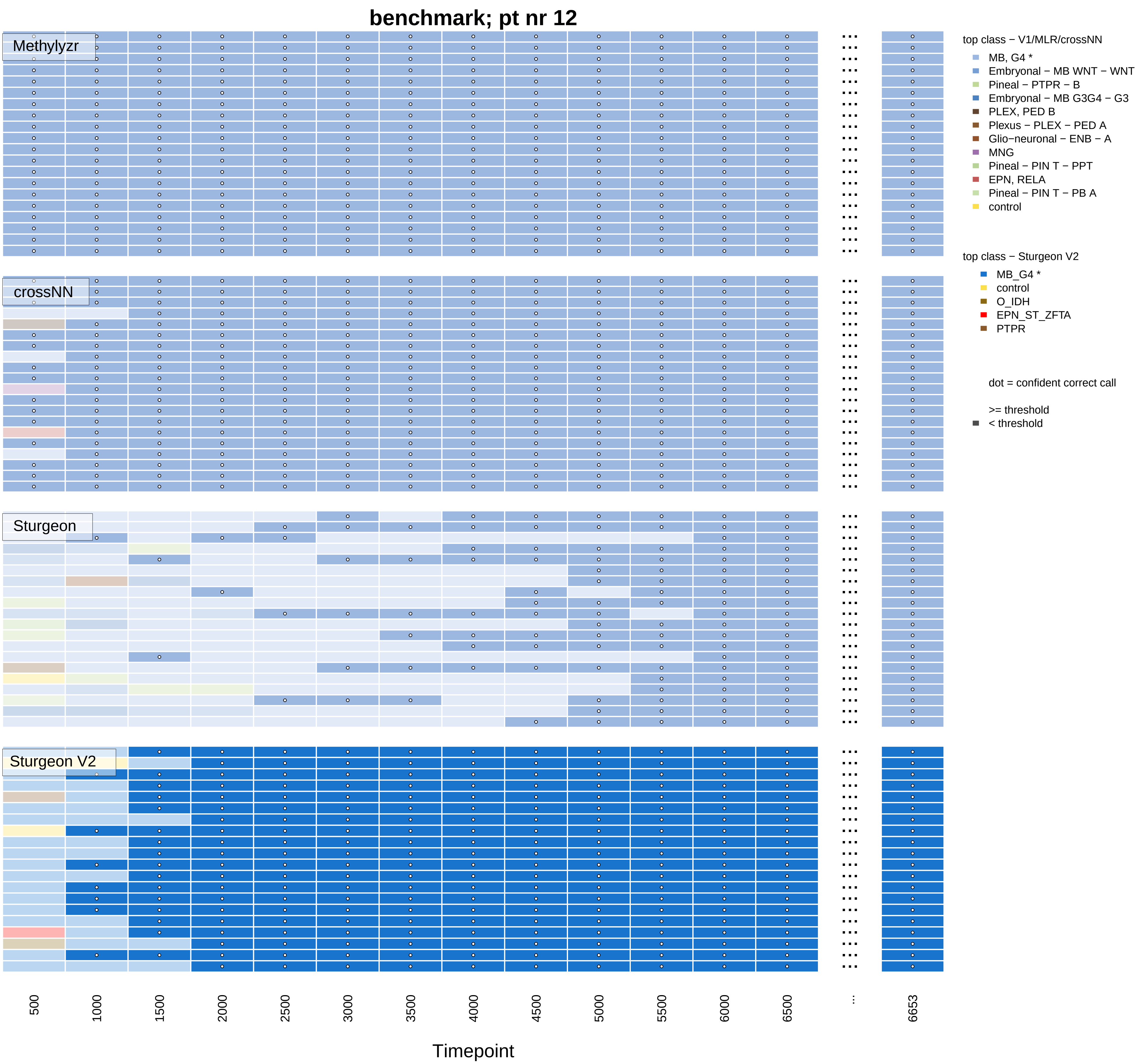

benchmark; pt nr 12
Methylyzr
crossNN
Sturgeon
Sturgeon V2
top class − V1/MLR/crossNN
MB, G4 *
Embryonal − MB WNT − WNT
Pineal − PTPR − B
Embryonal − MB G3G4 − G3
PLEX, PED B
Plexus − PLEX − PED A
Glio−neuronal − ENB − A
MNG
Pineal − PIN T − PPT
EPN, RELA
Pineal − PIN T − PB A
control
top class − Sturgeon V2
MB_G4 *
control
O_IDH
EPN_ST_ZFTA
PTPR
dot = confident correct call
>= threshold
< threshold
500
1000
1500
2000
2500
3000
3500
4000
4500
5000
5500
6000
6500
...
6653
Timepoint

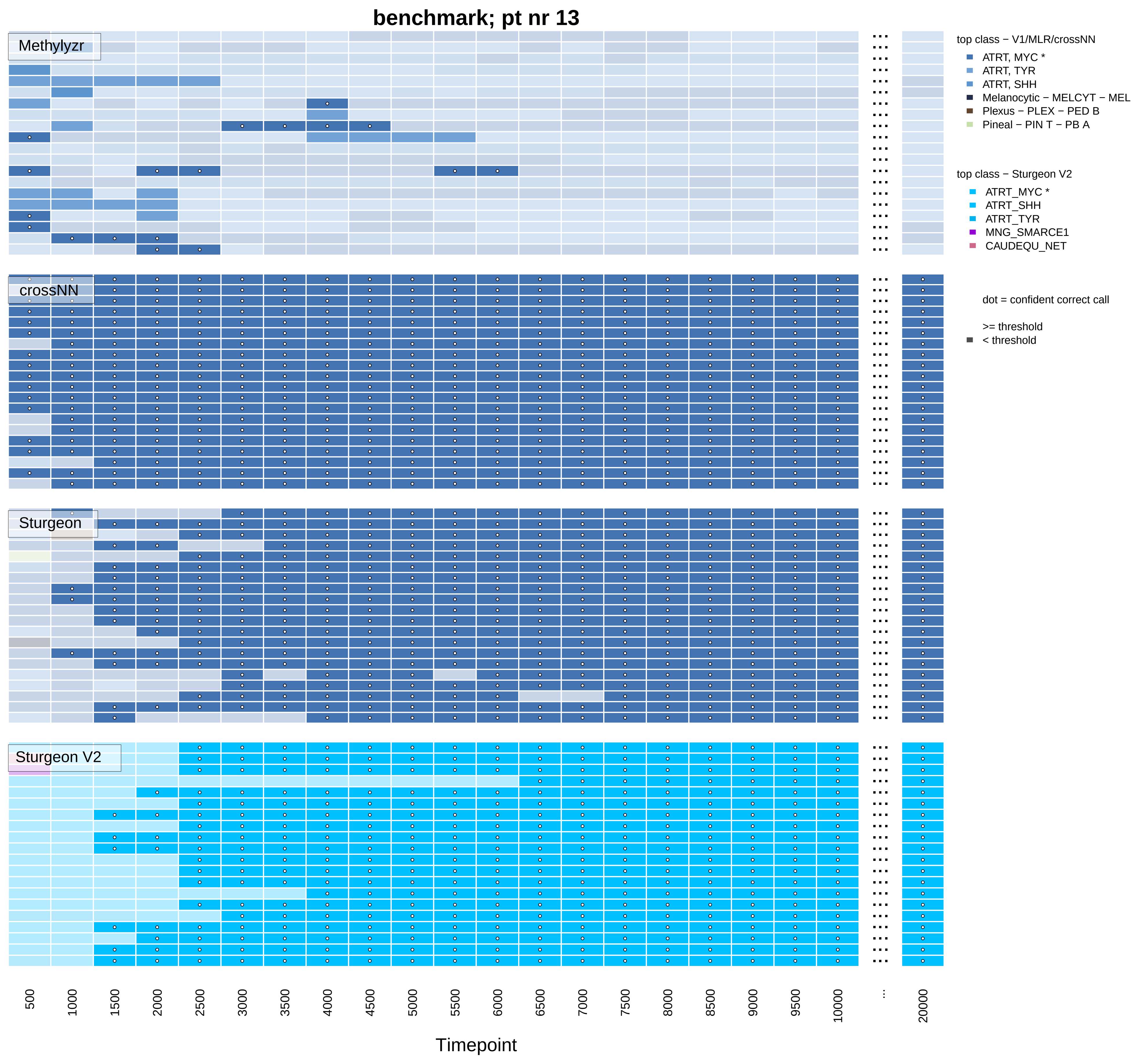
benchmark; pt nr 13
Methylyzr
crossNN
Sturgeon
Sturgeon V2
top class − V1/MLR/crossNN
ATRT, MYC *
ATRT, TYR
ATRT, SHH
Melanocytic − MELCYT − MEL
Plexus − PLEX − PED B
Pineal − PIN T − PB A
top class − Sturgeon V2
ATRT_MYC *
ATRT_SHH
ATRT_TYR
MNG_SMARCE1
CAUDEQU_NET
dot = confident correct call
>= threshold
< threshold
500
1000
1500
2000
2500
3000
3500
4000
4500
5000
5500
6000
6500
7000
7500
8000
8500
9000
9500
10000
20000
Timepoint

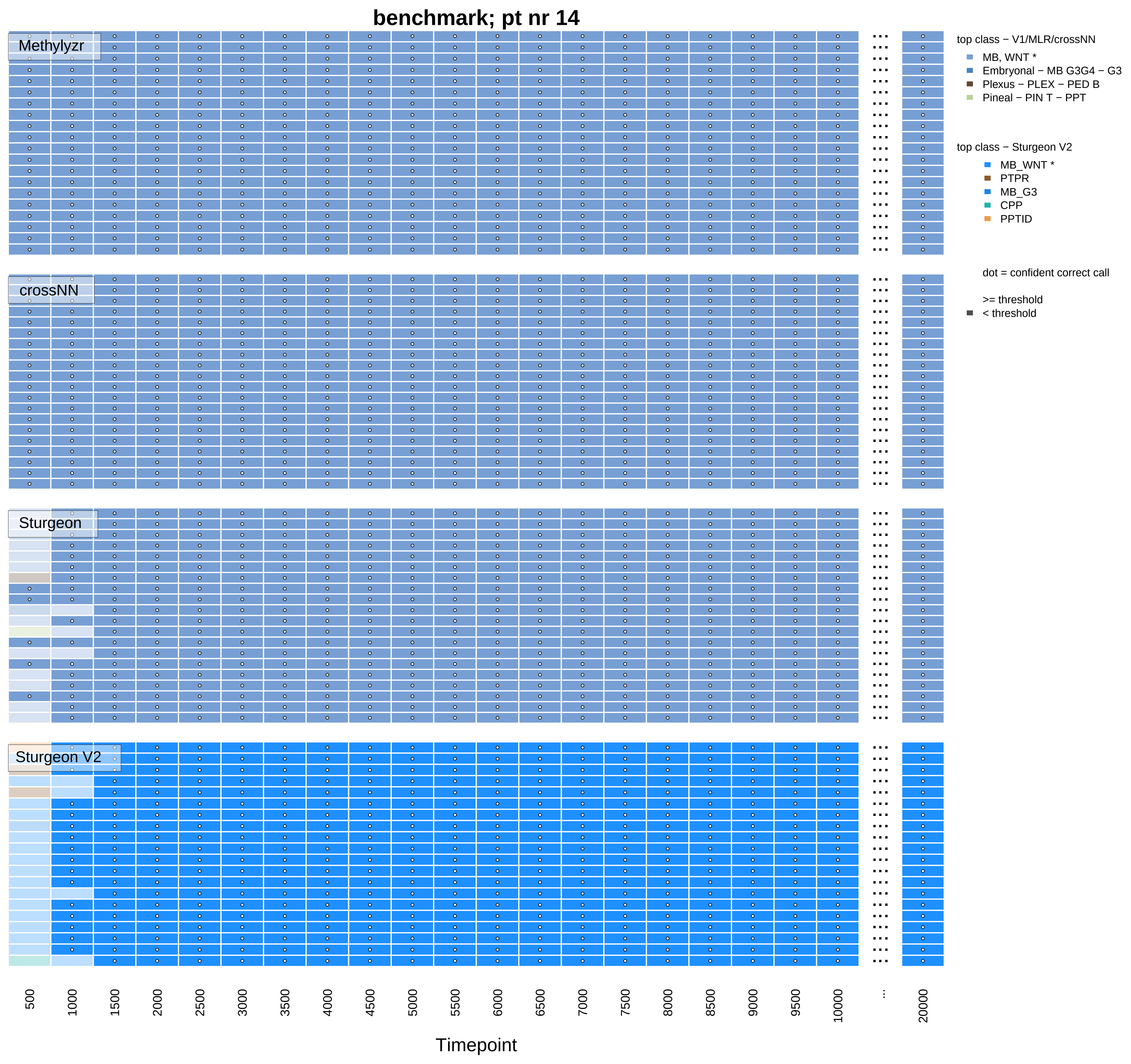
benchmark; pt nr 14
Methylyzr
crossNN
Sturgeon
Sturgeon V2
top class − V1/MLR/crossNN
MB, WNT *
Embryonal − MB G3G4 − G3
Plexus − PLEX − PED B
Pineal − PIN T − PPT
top class − Sturgeon V2
MB_WNT *
PTPR
MB_G3
CPP
PPTID
dot = confident correct call
>= threshold
< threshold
500
1000
1500
2000
2500
3000
3500
4000
4500
5000
5500
6000
6500
7000
7500
8000
8500
9000
9500
10000
...
20000
Timepoint

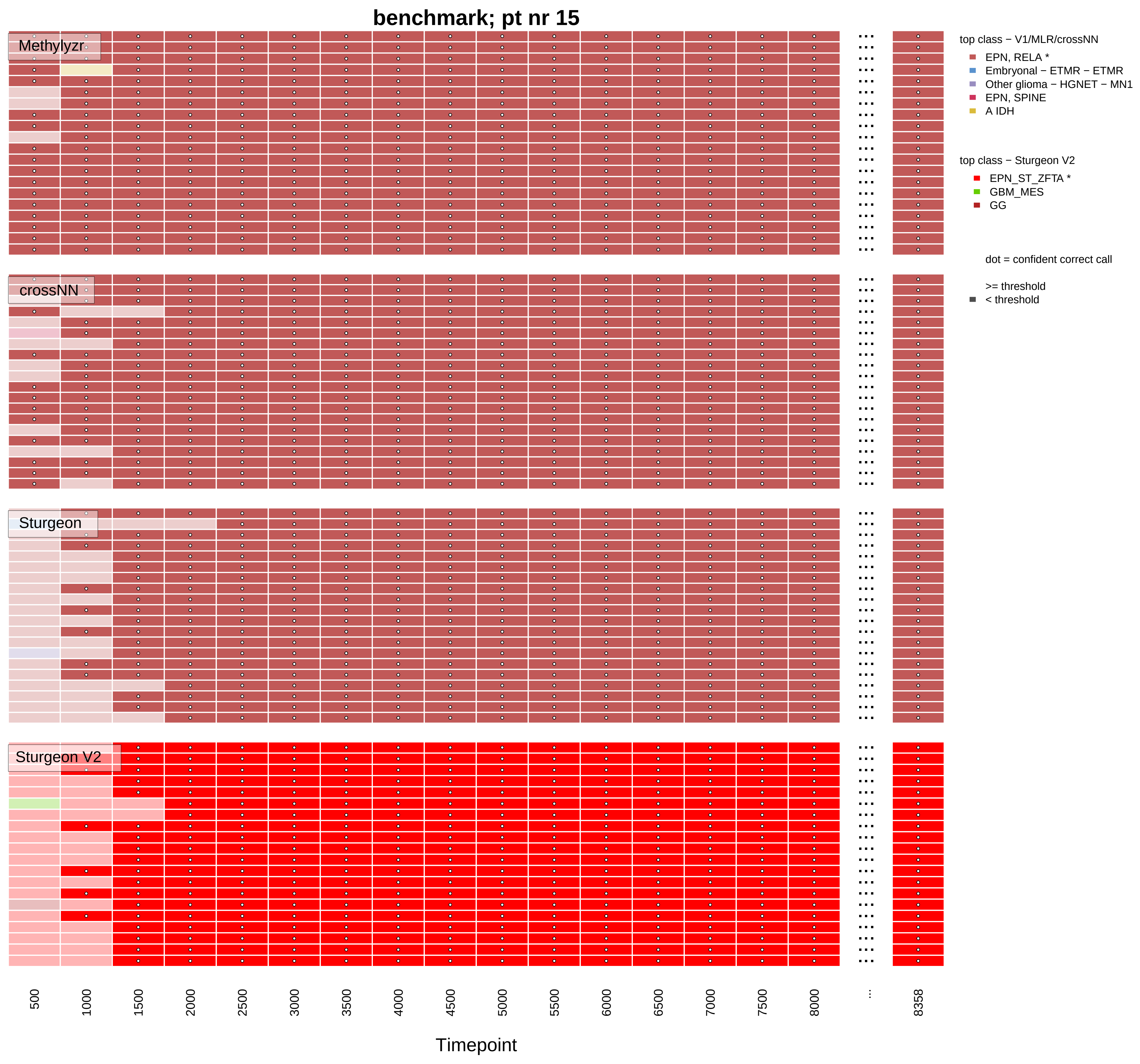
benchmark; pt nr 15
Methylyzr
crossNN
Sturgeon
Sturgeon V2
top class − V1/MLR/crossNN
EPN, RELA *
Embryonal − ETMR − ETMR
Other glioma − HGNET − MN1
EPN, SPINE
A IDH
top class − Sturgeon V2
EPN_ST_ZFTA *
GBM_MES
GG
dot = confident correct call
>= threshold
< threshold
500
1000
1500
2000
2500
3000
3500
4000
4500
5000
5500
6000
6500
7000
7500
8000
...
8358
Timepoint

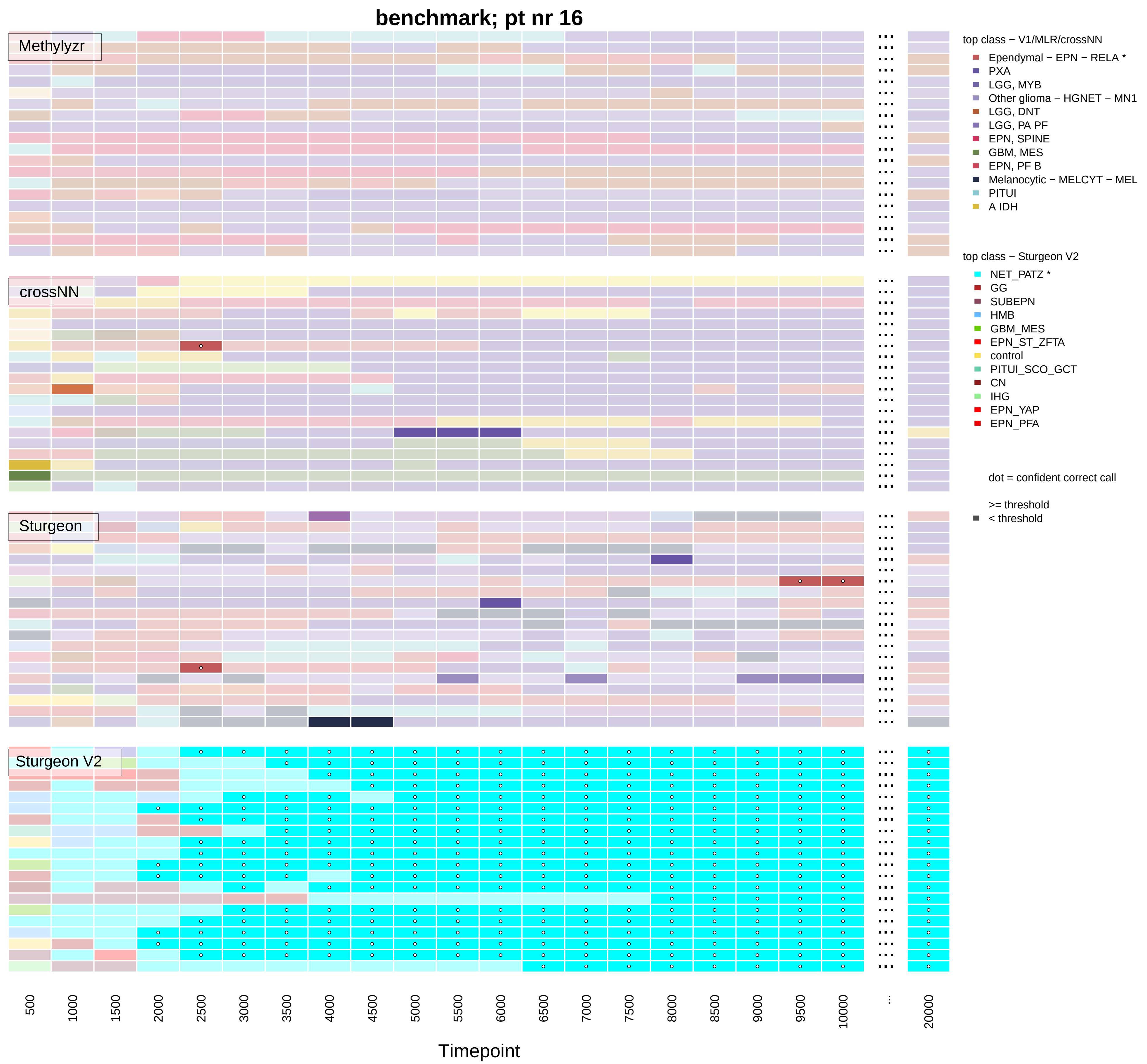
benchmark; pt nr 16
Methylyzr
crossNN
Sturgeon
Sturgeon V2
top class − V1/MLR/crossNN
Ependymal − EPN − RELA *
PXA
LGG, MYB
Other glioma − HGNET − MN1
LGG, DNT
LGG, PA PF
EPN, SPINE
GBM, MES
EPN, PF B
Melanocytic − MELCYT − MEL
PITUI
A IDH
top class − Sturgeon V2
NET_PATZ *
GG
SUBEPN
HMB
GBM_MES
EPN_ST_ZFTA
control
PITUI_SCO_GCT
CN
IHG
EPN_YAP
EPN_PFA
dot = confident correct call
>= threshold
< threshold
500
1000
1500
2000
2500
3000
3500
4000
4500
5000
5500
6000
6500
7000
7500
8000
8500
9000
9500
10000
...
20000
Timepoint

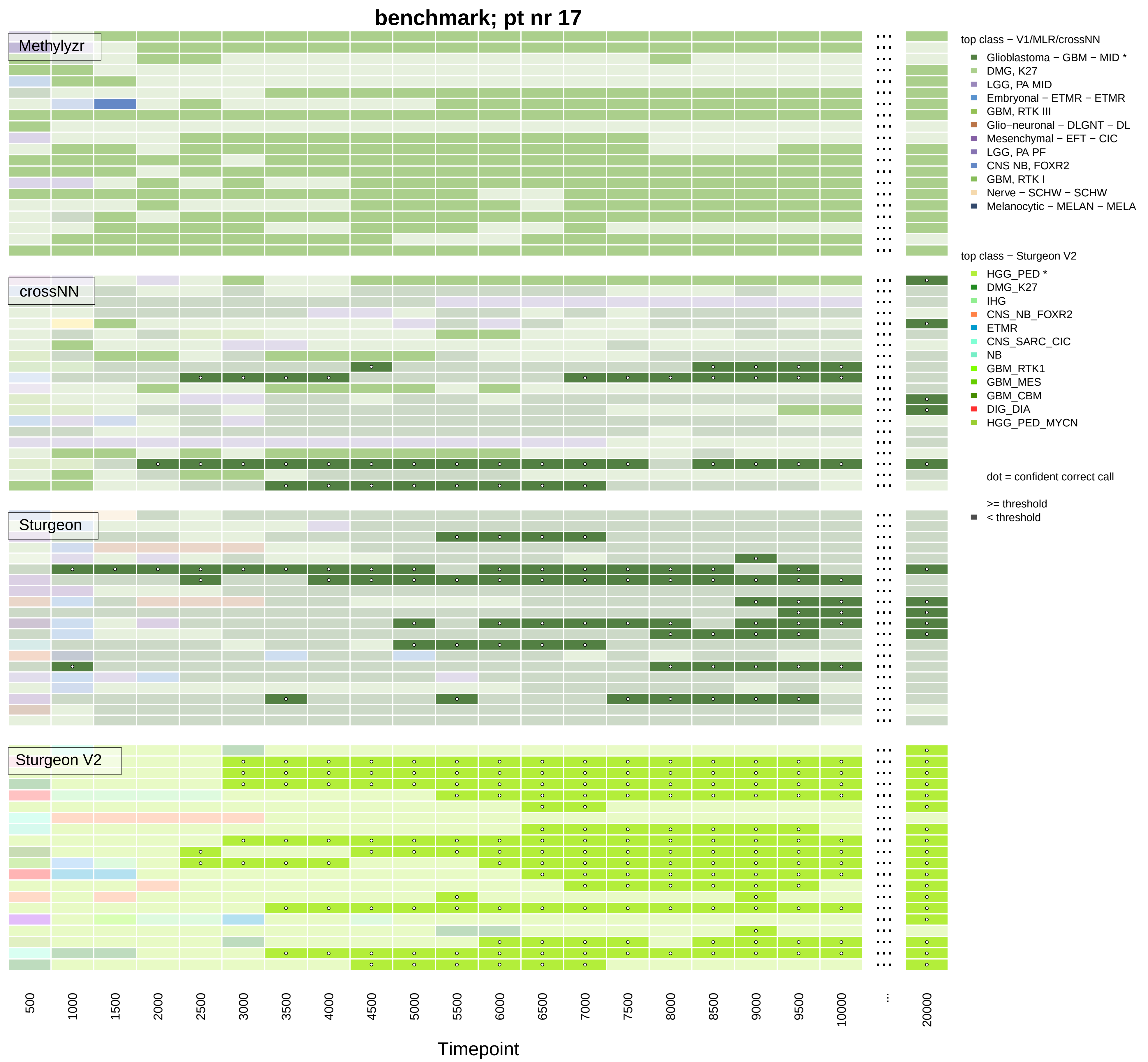

benchmark; pt nr 17
Methylyzr
crossNN
Sturgeon
Sturgeon V2
top class − V1/MLR/crossNN
Glioblastoma − GBM − MID *
DMG, K27
LGG, PA MID
Embryonal − ETMR − ETMR
GBM, RTK III
Glio−neuronal − DLGNT − DL
Mesenchymal − EFT − CIC
LGG, PA PF
CNS NB, FOXR2
GBM, RTK I
Nerve − SCHW − SCHW
Melanocytic − MELAN − MELA
top class − Sturgeon V2
HGG_PED *
DMG_K27
IHG
CNS_NB_FOXR2
ETMR
CNS_SARC_CIC
NB
GBM_RTK1
GBM_MES
GBM_CBM
DIG_DIA
HGG_PED_MYCN
dot = confident correct call
>= threshold
< threshold
500
1000
1500
2000
2500
3000
3500
4000
4500
5000
5500
6000
6500
7000
7500
8000
8500
9000
9500
10000
...
20000
Timepoint

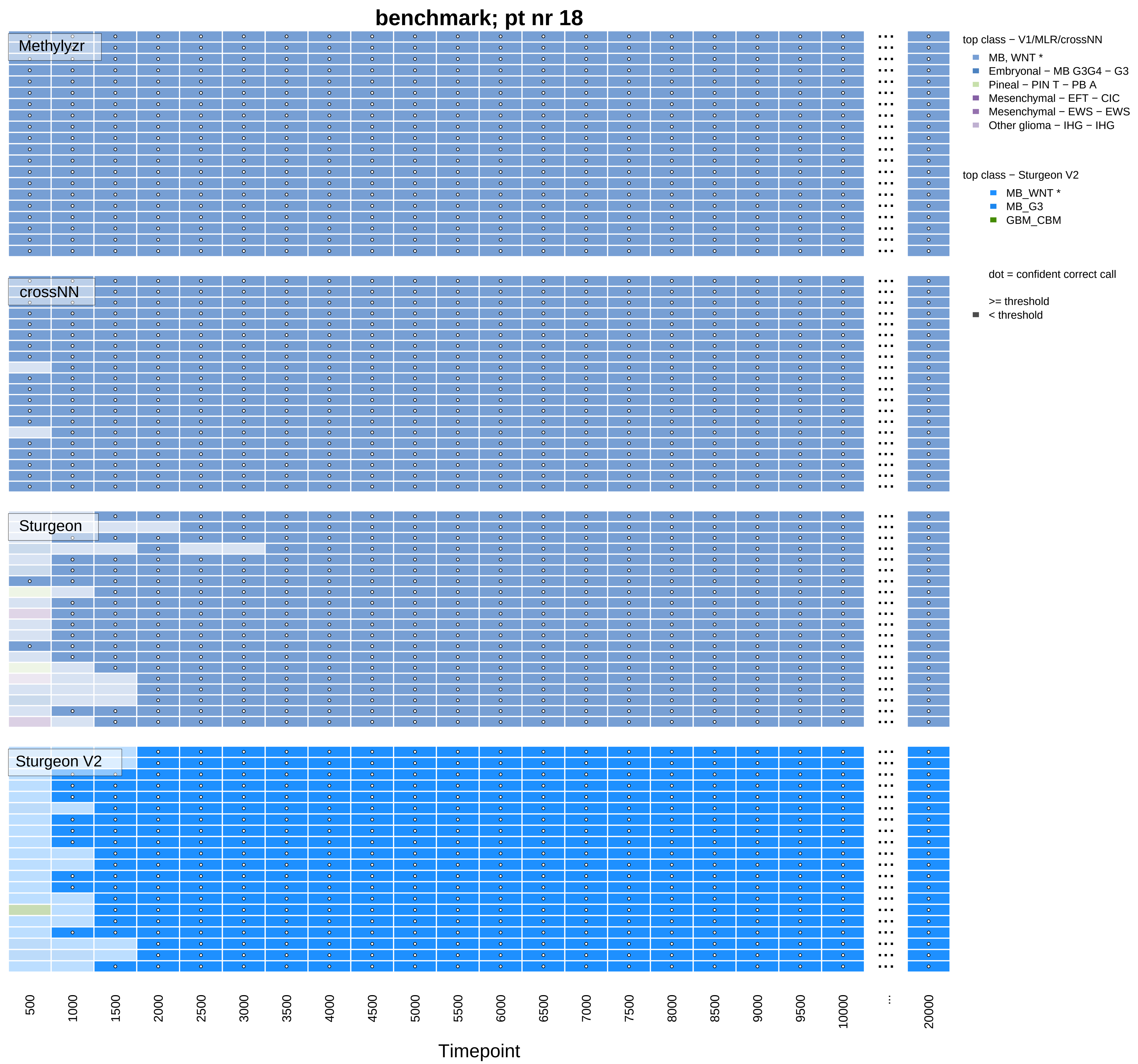

benchmark; pt nr 18
Methylyzr
crossNN
Sturgeon
Sturgeon V2
top class − V1/MLR/crossNN
MB, WNT *
Embryonal − MB G3G4 − G3
Pineal − PIN T − PB A
Mesenchymal − EFT − CIC
Mesenchymal − EWS − EWS
Other glioma − IHG − IHG
top class − Sturgeon V2
MB_WNT *
MB_G3
GBM_CBM
dot = confident correct call
>= threshold
< threshold
500
1000
1500
2000
2500
3000
3500
4000
4500
5000
5500
6000
6500
7000
7500
8000
8500
9000
9500
10000
...
20000
Timepoint

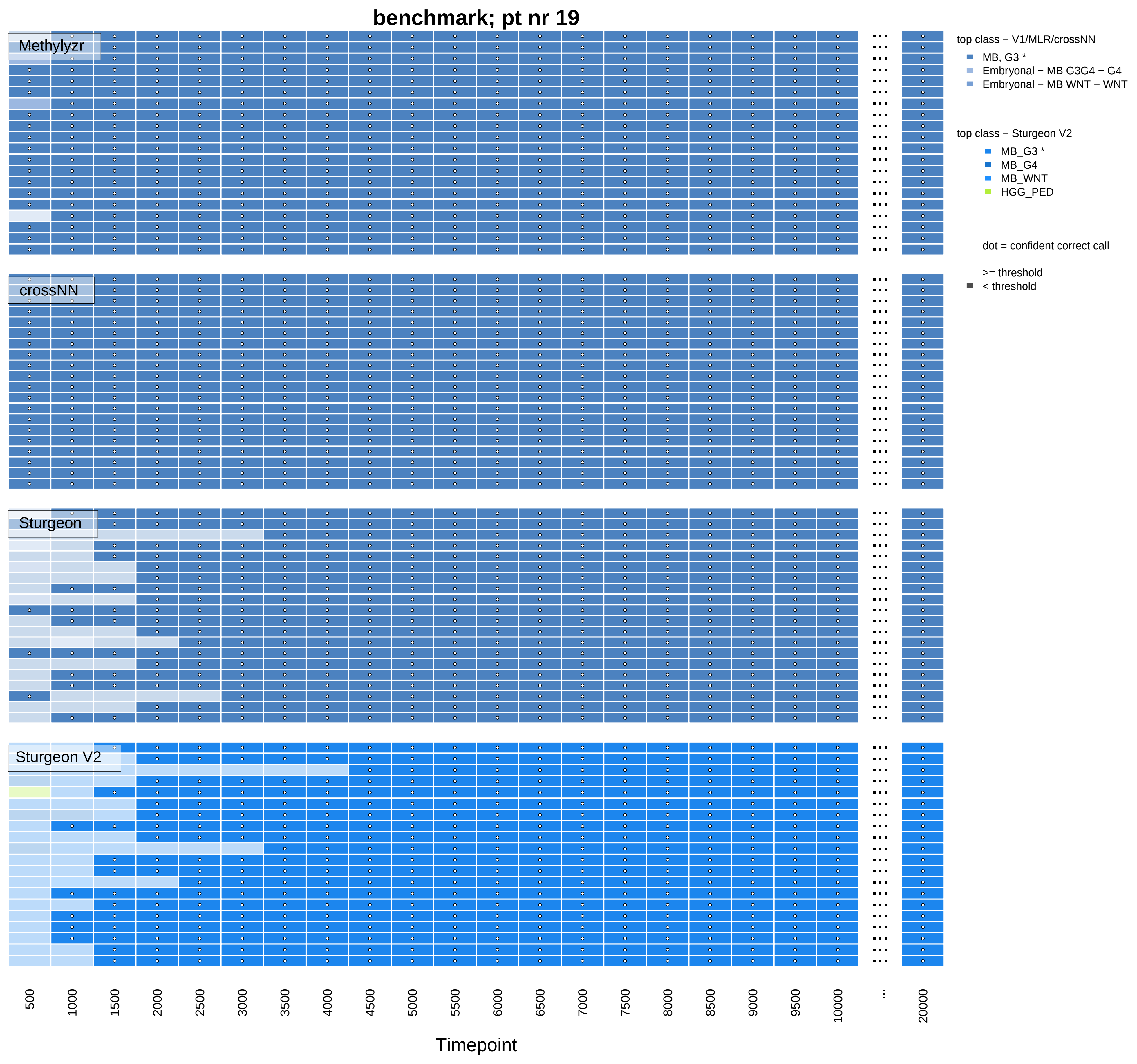
benchmark; pt nr 19
Methylyzr
crossNN
Sturgeon
Sturgeon V2
top class − V1/MLR/crossNN
MB, G3 *
Embryonal − MB G3G4 − G4
Embryonal − MB WNT − WNT
top class − Sturgeon V2
MB_G3 *
MB_G4
MB_WNT
HGG_PED
dot = confident correct call
>= threshold
< threshold
500
1000
1500
2000
2500
3000
3500
4000
4500
5000
5500
6000
6500
7000
7500
8000
8500
9000
9500
10000
...
20000
Timepoint

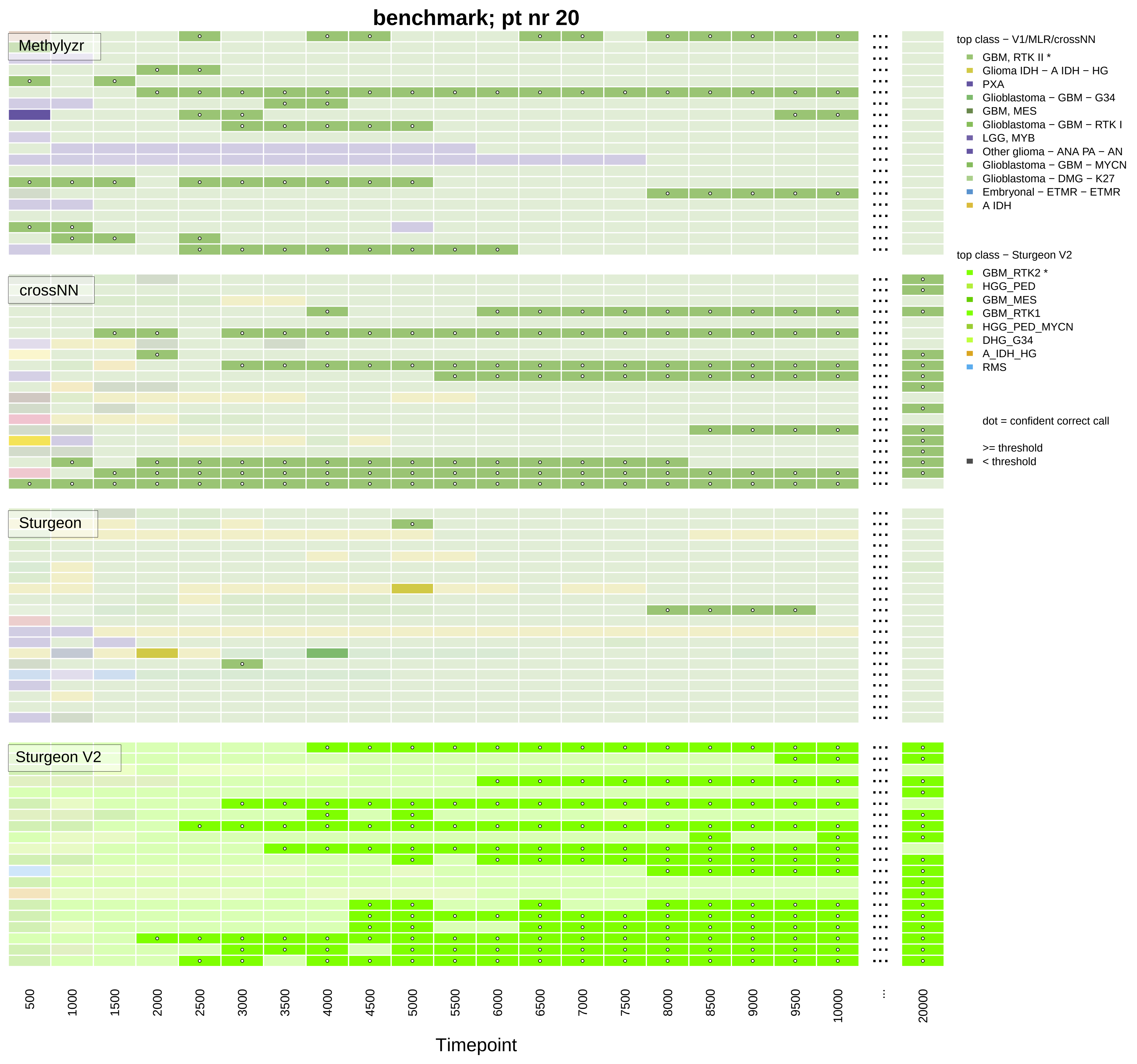

benchmark; pt nr 20
Methylyzr
crossNN
Sturgeon
Sturgeon V2
top class − V1/MLR/crossNN
GBM, RTK II *
Glioma IDH − A IDH − HG
PXA
Glioblastoma − GBM − G34
GBM, MES
Glioblastoma − GBM − RTK I
LGG, MYB
Other glioma − ANA PA − AN
Glioblastoma − GBM − MYCN
Glioblastoma − DMG − K27
Embryonal − ETMR − ETMR
A IDH
top class − Sturgeon V2
GBM_RTK2 *
HGG_PED
GBM_MES
GBM_RTK1
HGG_PED_MYCN
DHG_G34
A_IDH_HG
RMS
dot = confident correct call
>= threshold
< threshold
500
1000
1500
2000
2500
3000
3500
4000
4500
5000
5500
6000
6500
7000
7500
8000
8500
9000
9500
10000
...
20000
Timepoint

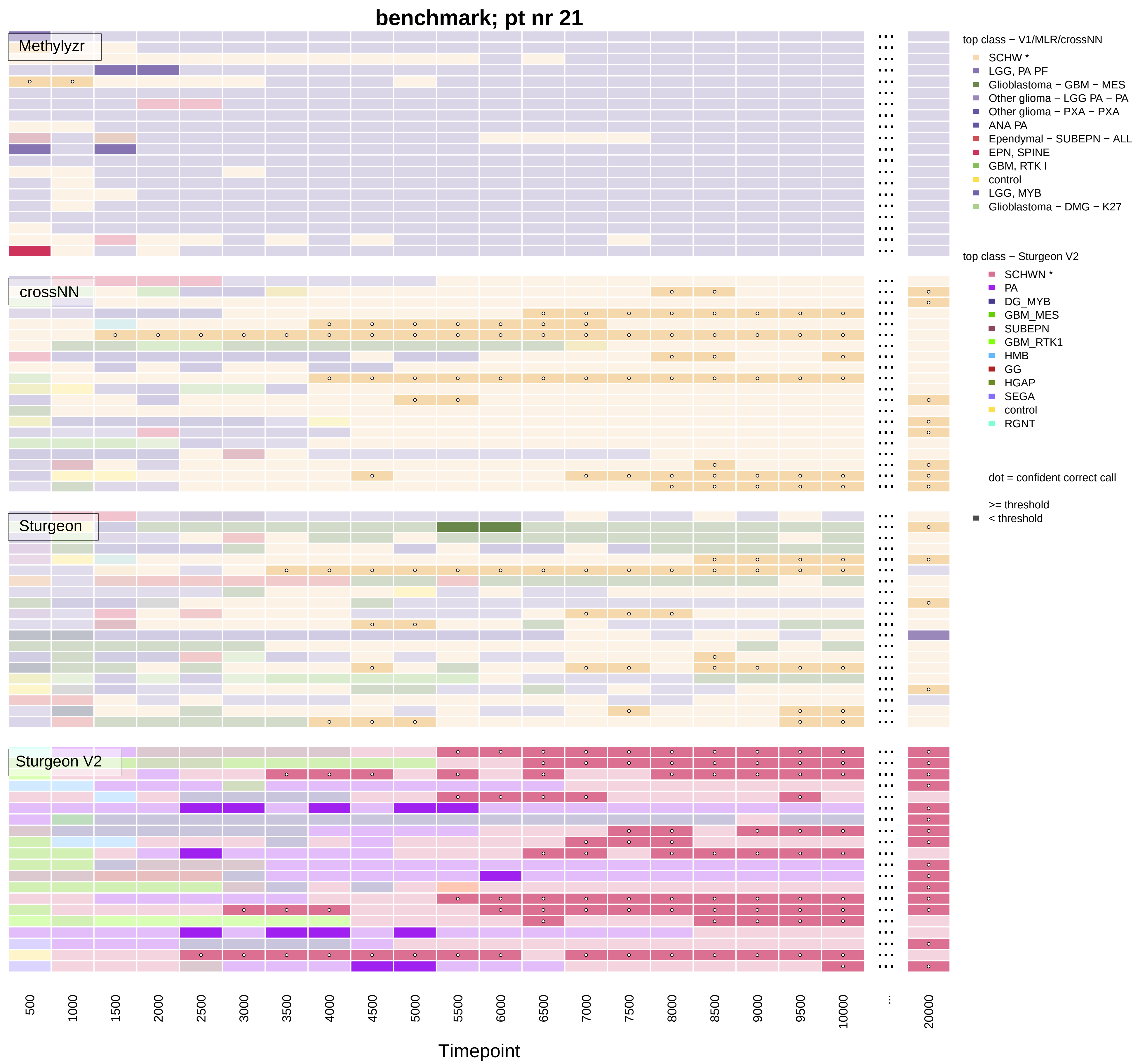
benchmark; pt nr 21
Methylyzr
crossNN
Sturgeon
Sturgeon V2
top class − V1/MLR/crossNN
SCHW *
LGG, PA PF
Glioblastoma − GBM − MES
Other glioma − LGG PA − PA
Other glioma − PXA − PXA
ANA PA
Ependymal − SUBEPN − ALL
EPN, SPINE
GBM, RTK I
control
LGG, MYB
Glioblastoma − DMG − K27
top class − Sturgeon V2
SCHWN *
PA
DG_MYB
GBM_MES
SUBEPN
GBM_RTK1
HMB
GG
HGAP
SEGA
control
RGNT
dot = confident correct call
>= threshold
< threshold
500
1000
1500
2000
2500
3000
3500
4000
4500
5000
5500
6000
6500
7000
7500
8000
8500
9000
9500
10000
...
20000
Timepoint

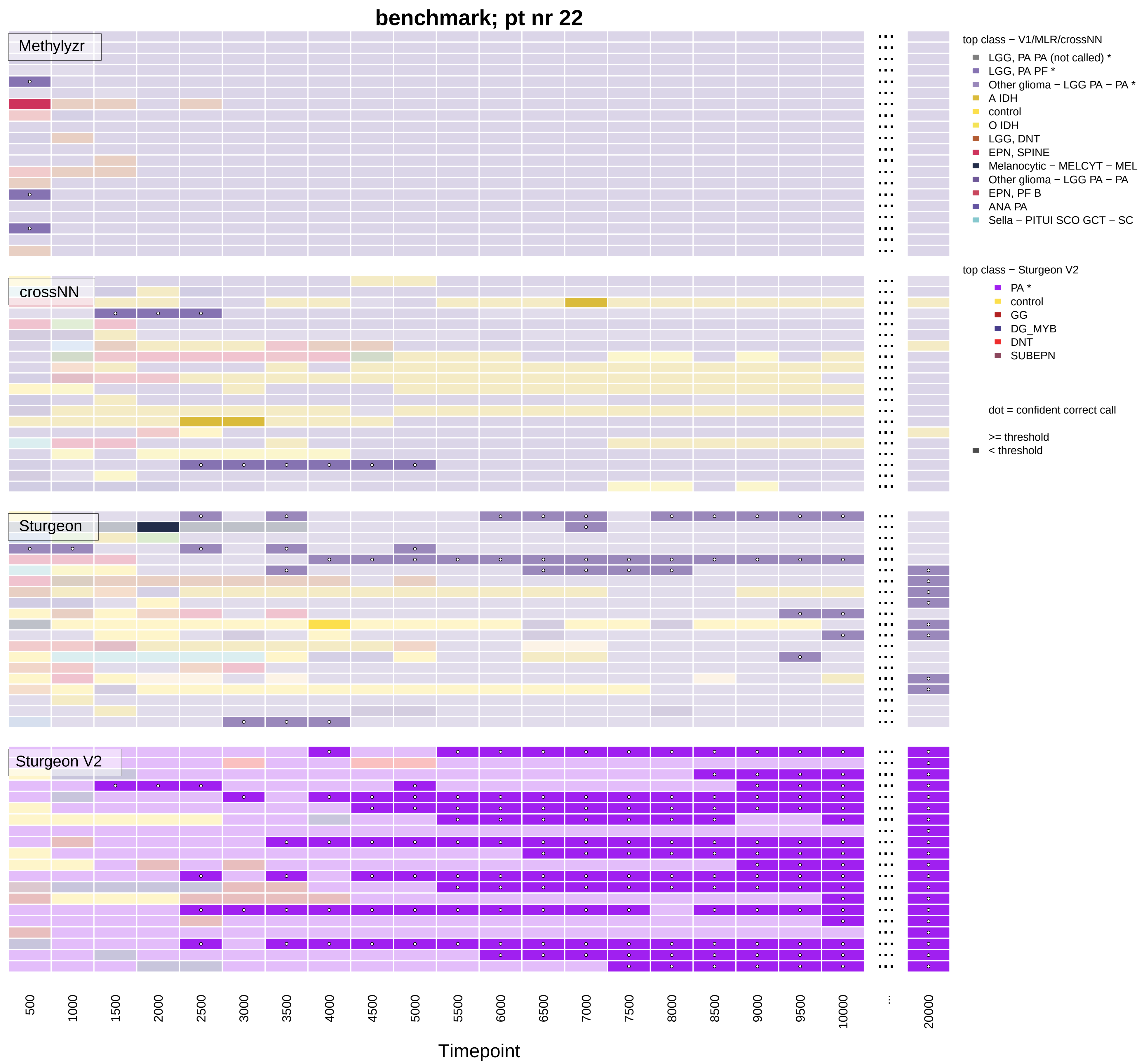

benchmark; pt nr 22
Methylyzr
crossNN
Sturgeon
Sturgeon V2
top class − V1/MLR/crossNN
LGG, PA PA (not called) *
LGG, PA PF *
Other glioma − LGG PA − PA *
A IDH
control
O IDH
LGG, DNT
EPN, SPINE
Melanocytic − MELCYT − MEL
Other glioma − LGG PA − PA
EPN, PF B
ANA PA
Sella − PITUI SCO GCT − SC
top class − Sturgeon V2
PA *
control
GG
DG_MYB
DNT
SUBEPN
dot = confident correct call
>= threshold
< threshold
500
1000
1500
2000
2500
3000
3500
4000
4500
5000
5500
6000
6500
7000
7500
8000
8500
9000
9500
10000
...
20000
Timepoint

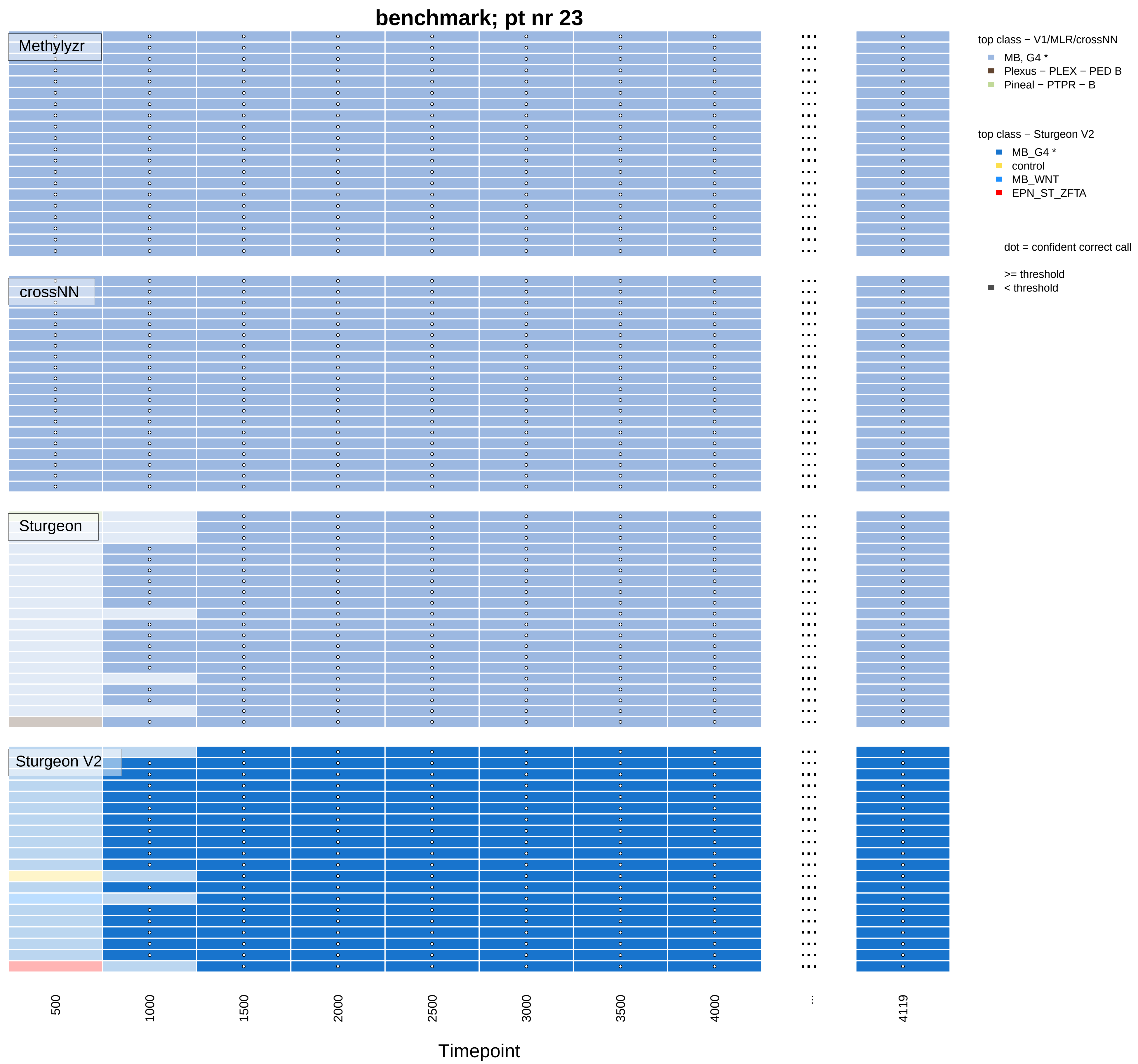

benchmark; pt nr 23
Methylyzr
crossNN
Sturgeon
Sturgeon V2
top class − V1/MLR/crossNN
MB, G4 *
Plexus − PLEX − PED B
Pineal − PTPR − B
top class − Sturgeon V2
MB_G4 *
control
MB_WNT
EPN_ST_ZFTA
dot = confident correct call
>= threshold
< threshold
500
1000
1500
2000
2500
3000
3500
4000
...
4119
Timepoint

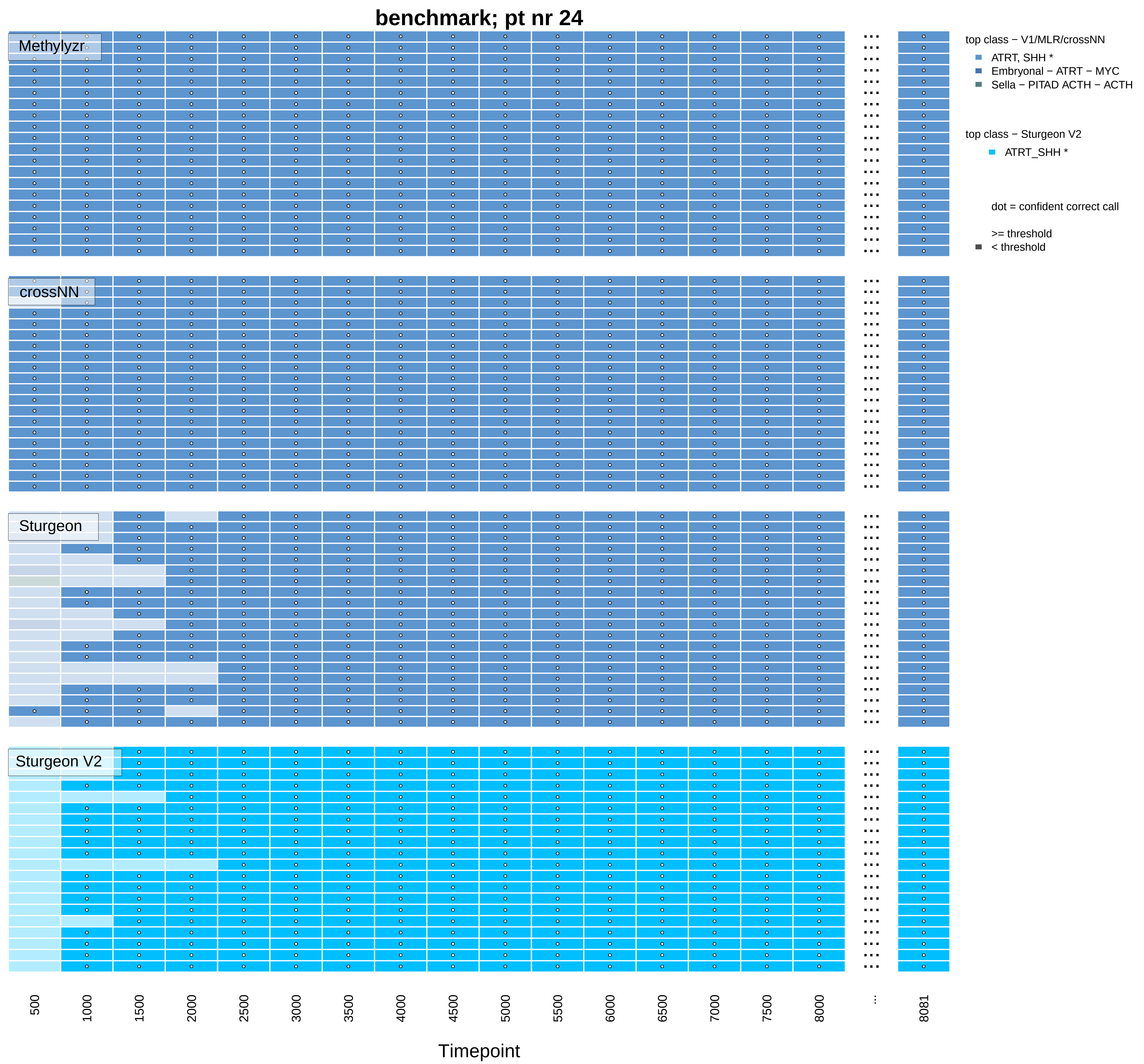

benchmark; pt nr 24
Methylyzr
crossNN
Sturgeon
Sturgeon V2
top class − V1/MLR/crossNN
ATRT, SHH *
Embryonal − ATRT − MYC
Sella − PITAD ACTH − ACTH
top class − Sturgeon V2
ATRT_SHH *
dot = confident correct call
>= threshold
< threshold
500
1000
1500
2000
2500
3000
3500
4000
4500
5000
5500
6000
6500
7000
7500
8000
...
8081
Timepoint

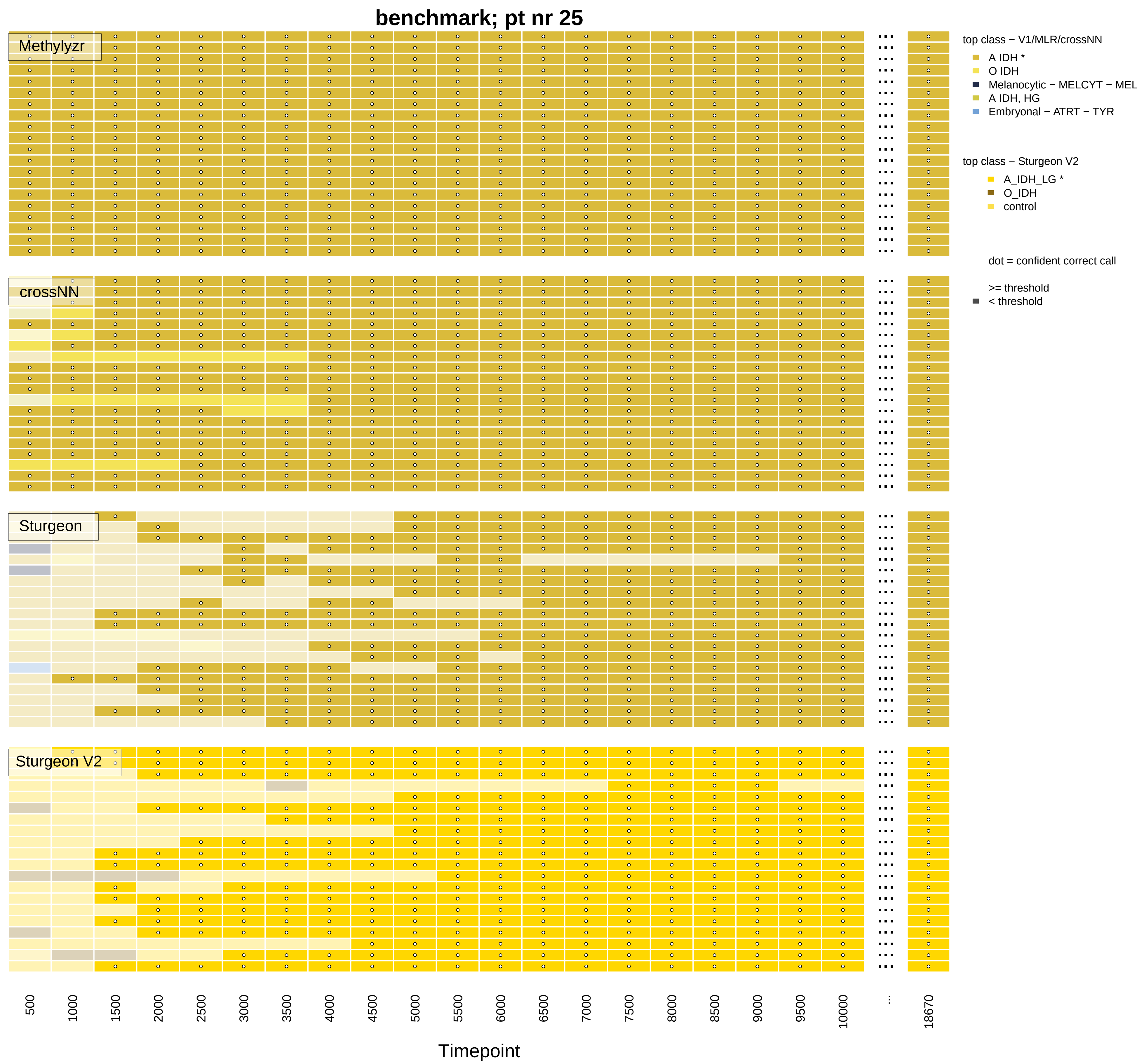
benchmark; pt nr 25
Methylyzr
crossNN
Sturgeon
Sturgeon V2
top class − V1/MLR/crossNN
A IDH *
O IDH
Melanocytic − MELCYT − MEL
A IDH, HG
Embryonal − ATRT − TYR
top class − Sturgeon V2
A_IDH_LG *
O_IDH
control
dot = confident correct call
>= threshold
< threshold
500
1000
1500
2000
2500
3000
3500
4000
4500
5000
5500
6000
6500
7000
7500
8000
8500
9000
9500
10000
...
18670
Timepoint

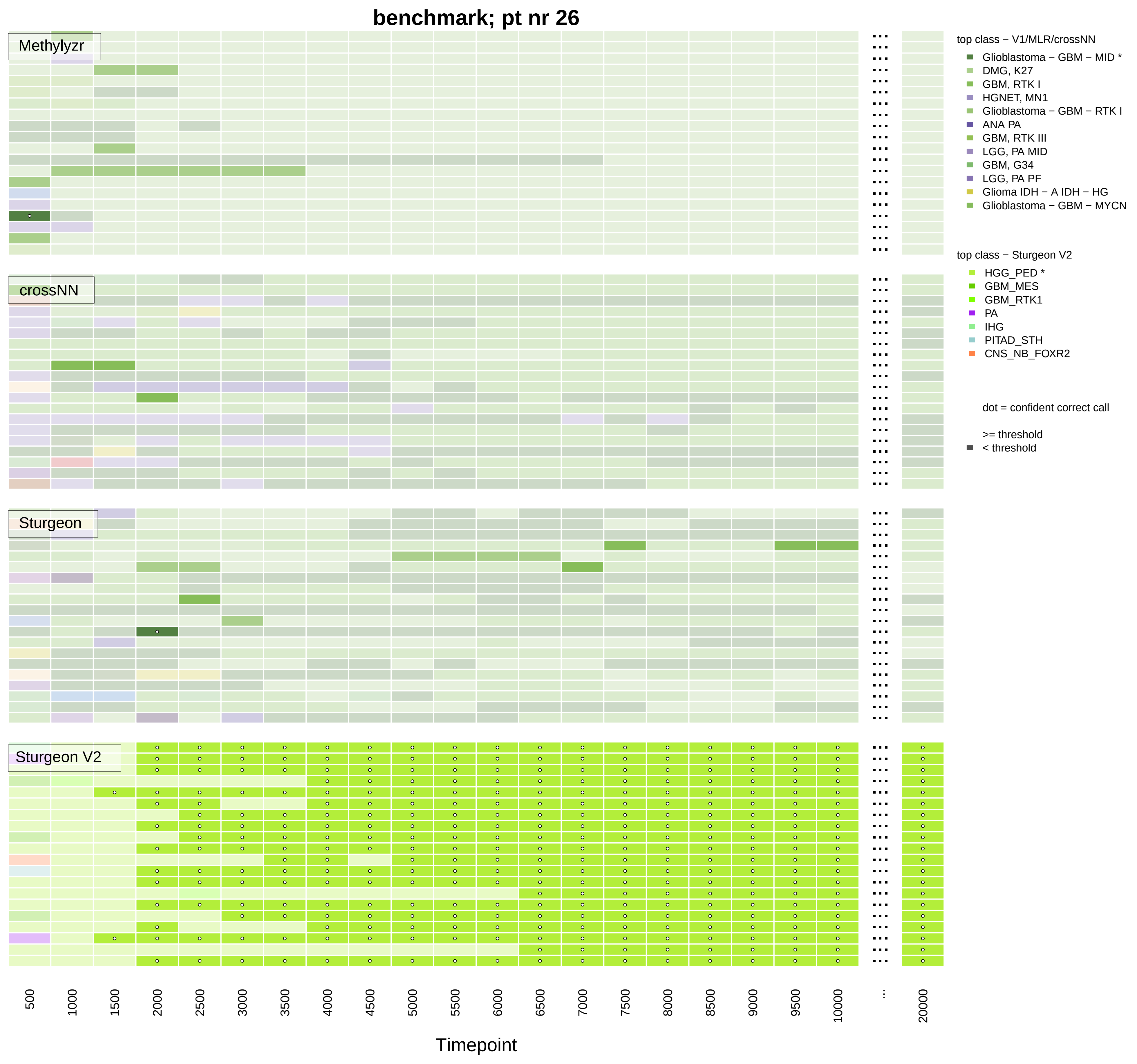
benchmark; pt nr 26
Methylyzr
crossNN
Sturgeon
Sturgeon V2
500
1000
1500
2000
2500
3000
3500
4000
4500
5000
5500
6000
6500
7000
7500
8000
8500
9000
9500
10000
...
20000
Timepoint
top class − V1/MLR/crossNN
Glioblastoma − GBM − MID *
DMG, K27
GBM, RTK I
HGNET, MN1
Glioblastoma − GBM − RTK I
ANA PA
GBM, RTK III
LGG, PA MID
GBM, G34
LGG, PA PF
Glioma IDH − A IDH − HG
Glioblastoma − GBM − MYCN
top class − Sturgeon V2
HGG_PED *
GBM_MES
GBM_RTK1
PA
IHG
PITAD_STH
CNS_NB_FOXR2
dot = confident correct call
>= threshold
< threshold

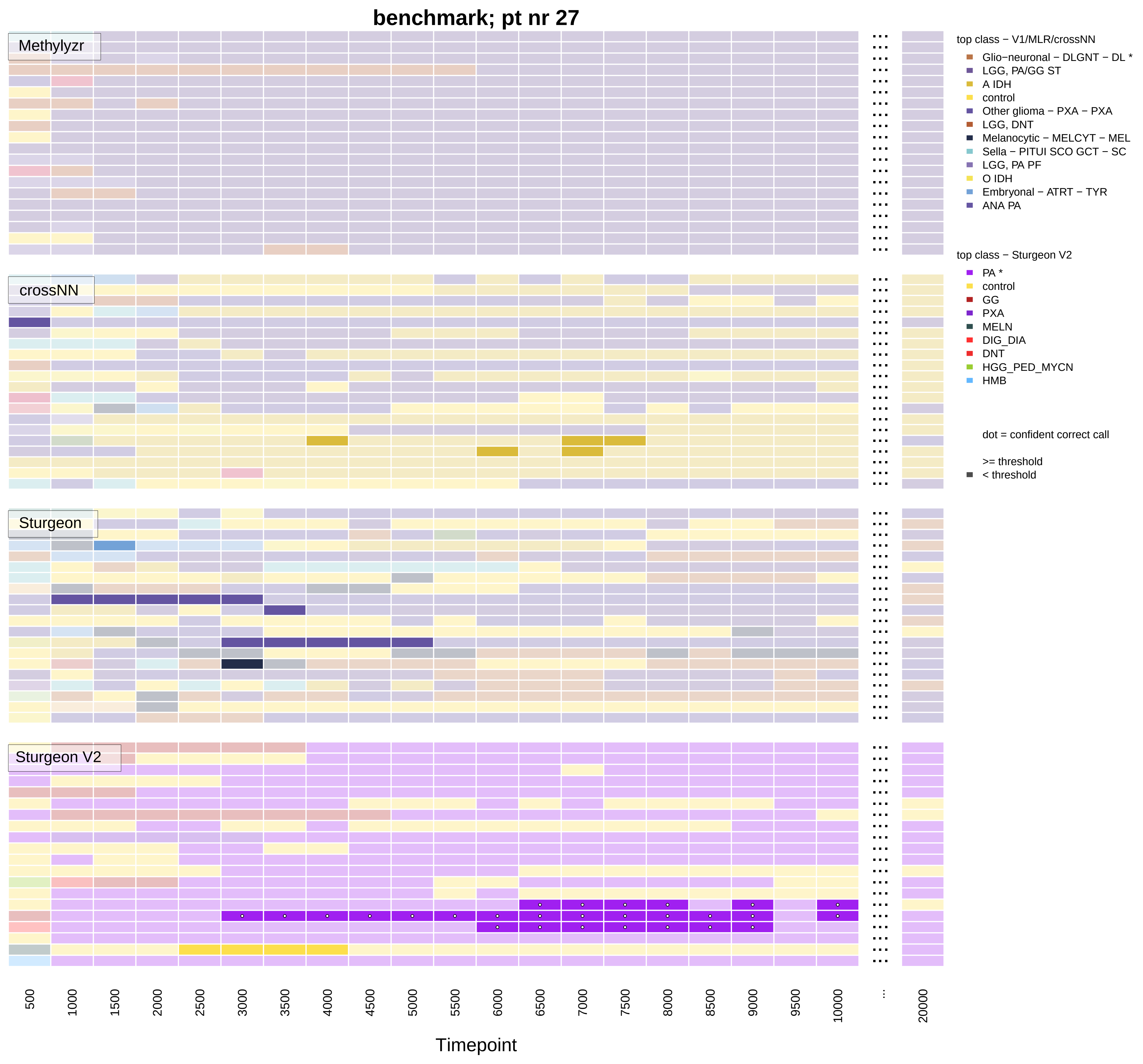
benchmark; pt nr 27
Methylyzr
crossNN
Sturgeon
Sturgeon V2
top class − V1/MLR/crossNN
Glio−neuronal − DLGNT − DL *
LGG, PA/GG ST
A IDH
control
Other glioma − PXA − PXA
LGG, DNT
Melanocytic − MELCYT − MEL
Sella − PITUI SCO GCT − SC
LGG, PA PF
O IDH
Embryonal − ATRT − TYR
ANA PA
top class − Sturgeon V2
PA *
control
GG
PXA
MELN
DIG_DIA
DNT
HGG_PED_MYCN
HMB
dot = confident correct call
>= threshold
< threshold
500
1000
1500
2000
2500
3000
3500
4000
4500
5000
5500
6000
6500
7000
7500
8000
8500
9000
9500
10000
...
20000
Timepoint

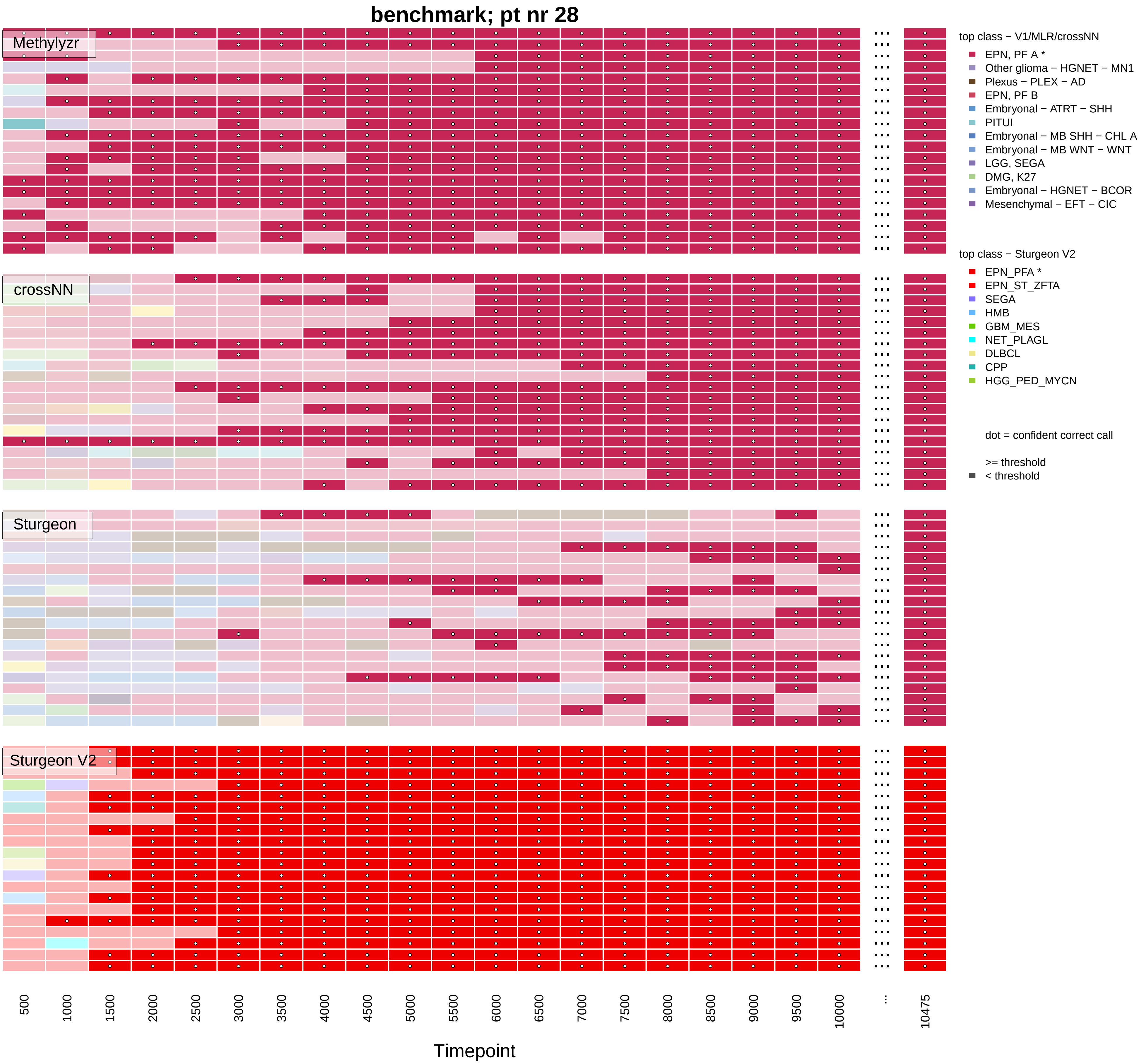

benchmark; pt nr 28
Methylyzr
crossNN
Sturgeon
Sturgeon V2
top class − V1/MLR/crossNN
EPN, PF A *
Other glioma − HGNET − MN1
Plexus − PLEX − AD
EPN, PF B
Embryonal − ATRT − SHH
PITUI
Embryonal − MB SHH − CHL A
Embryonal − MB WNT − WNT
LGG, SEGA
DMG, K27
Embryonal − HGNET − BCOR
Mesenchymal − EFT − CIC
top class − Sturgeon V2
EPN_PFA *
EPN_ST_ZFTA
SEGA
HMB
GBM_MES
NET_PLAGL
DLBCL
CPP
HGG_PED_MYCN
dot = confident correct call
>= threshold
< threshold
500
1000
1500
2000
2500
3000
3500
4000
4500
5000
5500
6000
6500
7000
7500
8000
8500
9000
9500
10000
...
10475
Timepoint

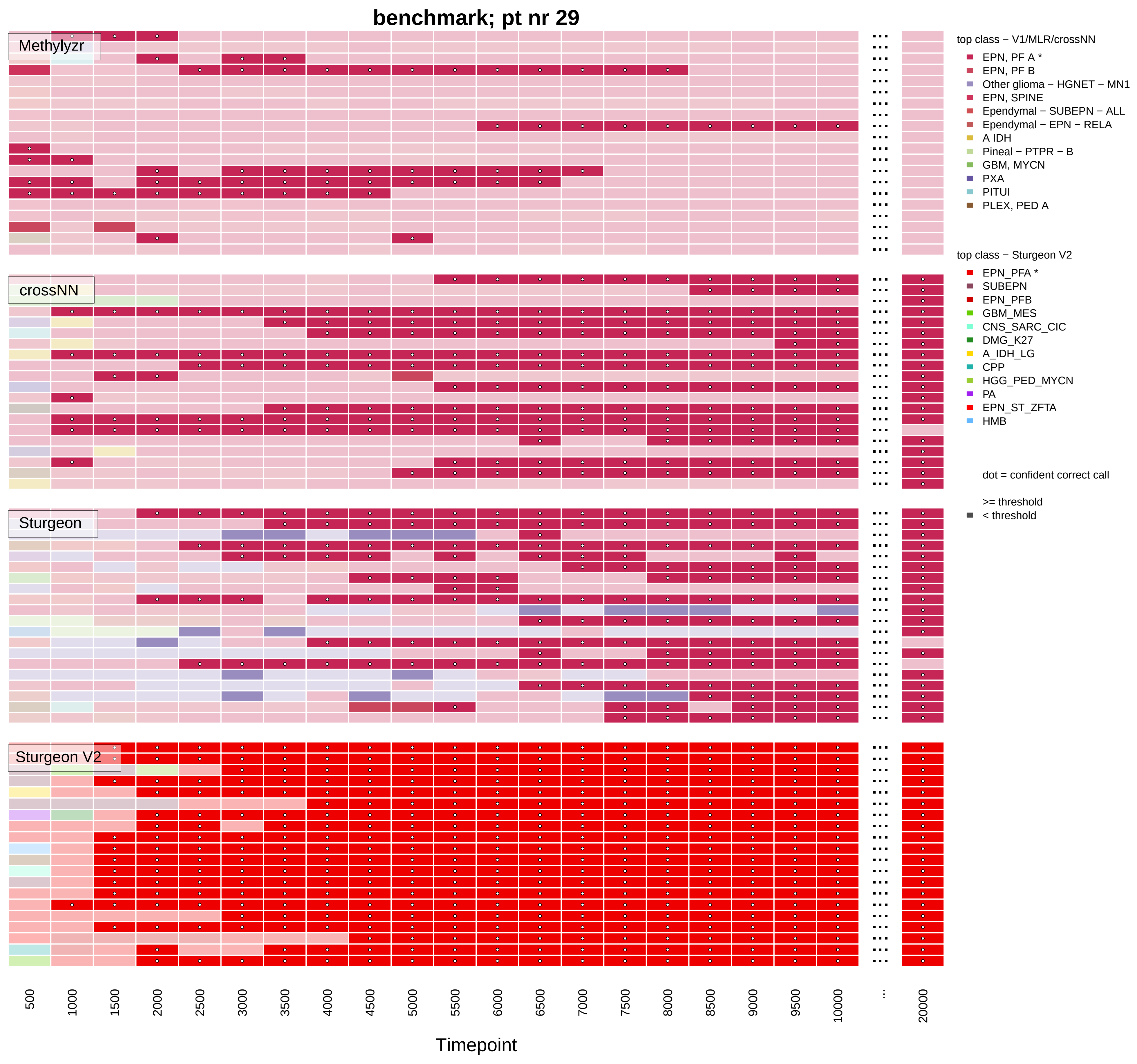
benchmark; pt nr 29
Methylyzr
crossNN
Sturgeon
Sturgeon V2
500
1000
1500
2000
2500
3000
3500
4000
4500
5000
5500
6000
6500
7000
7500
8000
8500
9000
9500
10000
...
20000
Timepoint
top class − V1/MLR/crossNN
EPN, PF A *
EPN, PF B
Other glioma − HGNET − MN1
EPN, SPINE
Ependymal − SUBEPN − ALL
Ependymal − EPN − RELA
A IDH
Pineal − PTPR − B
GBM, MYCN
PXA
PITUI
PLEX, PED A
top class − Sturgeon V2
EPN_PFA *
SUBEPN
EPN_PFB
GBM_MES
CNS_SARC_CIC
DMG_K27
A_IDH_LG
CPP
HGG_PED_MYCN
PA
EPN_ST_ZFTA
HMB
dot = confident correct call
>= threshold
< threshold

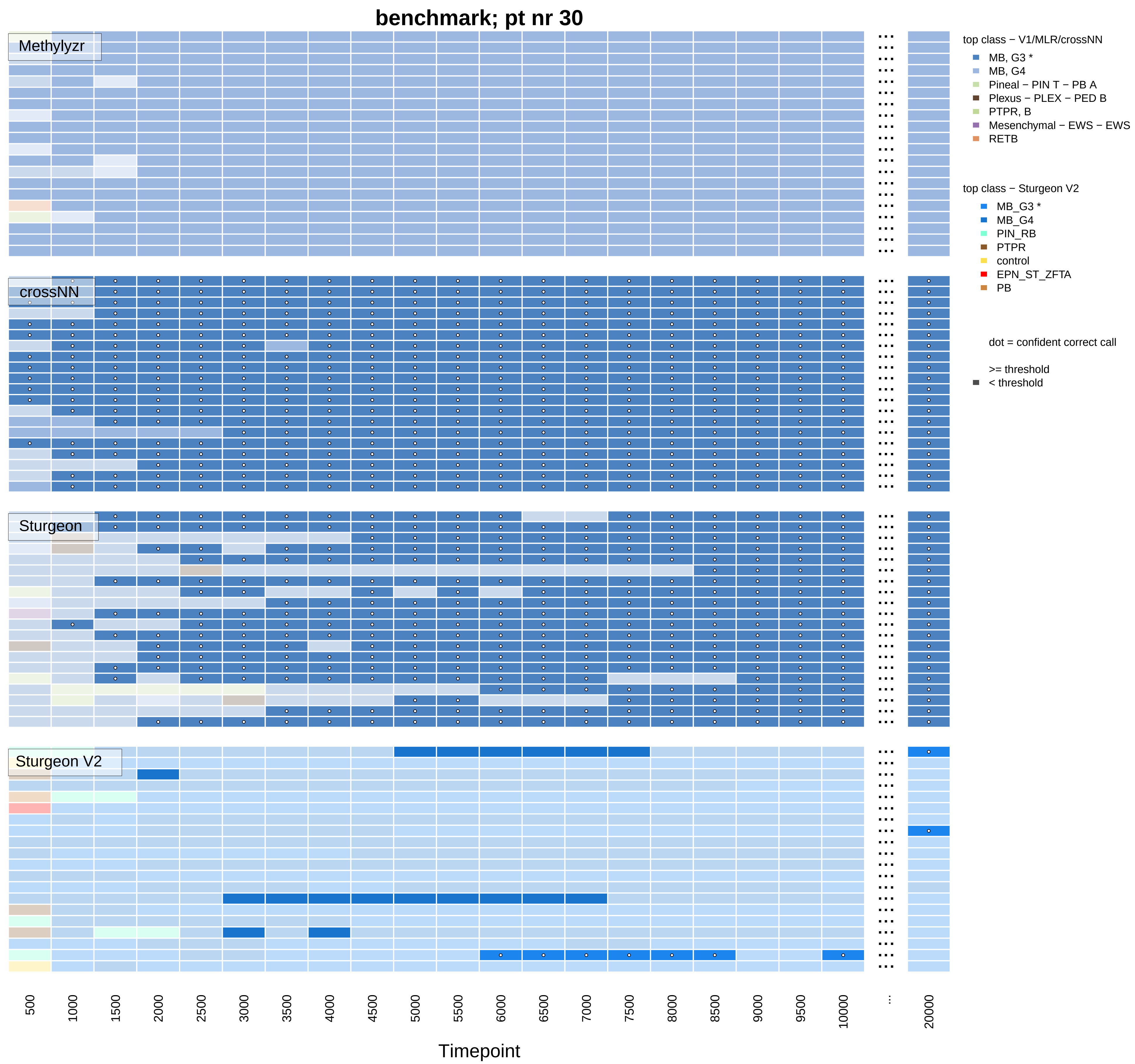

benchmark; pt nr 30
Methylyzr
crossNN
Sturgeon
Sturgeon V2
top class − V1/MLR/crossNN
MB, G3 *
MB, G4
Pineal − PIN T − PB A
Plexus − PLEX − PED B
PTPR, B
Mesenchymal − EWS − EWS
RETB
top class − Sturgeon V2
MB_G3 *
MB_G4
PIN_RB
PTPR
control
EPN_ST_ZFTA
PB
dot = confident correct call
>= threshold
< threshold
500
1000
1500
2000
2500
3000
3500
4000
4500
5000
5500
6000
6500
7000
7500
8000
8500
9000
9500
10000
...
20000
Timepoint

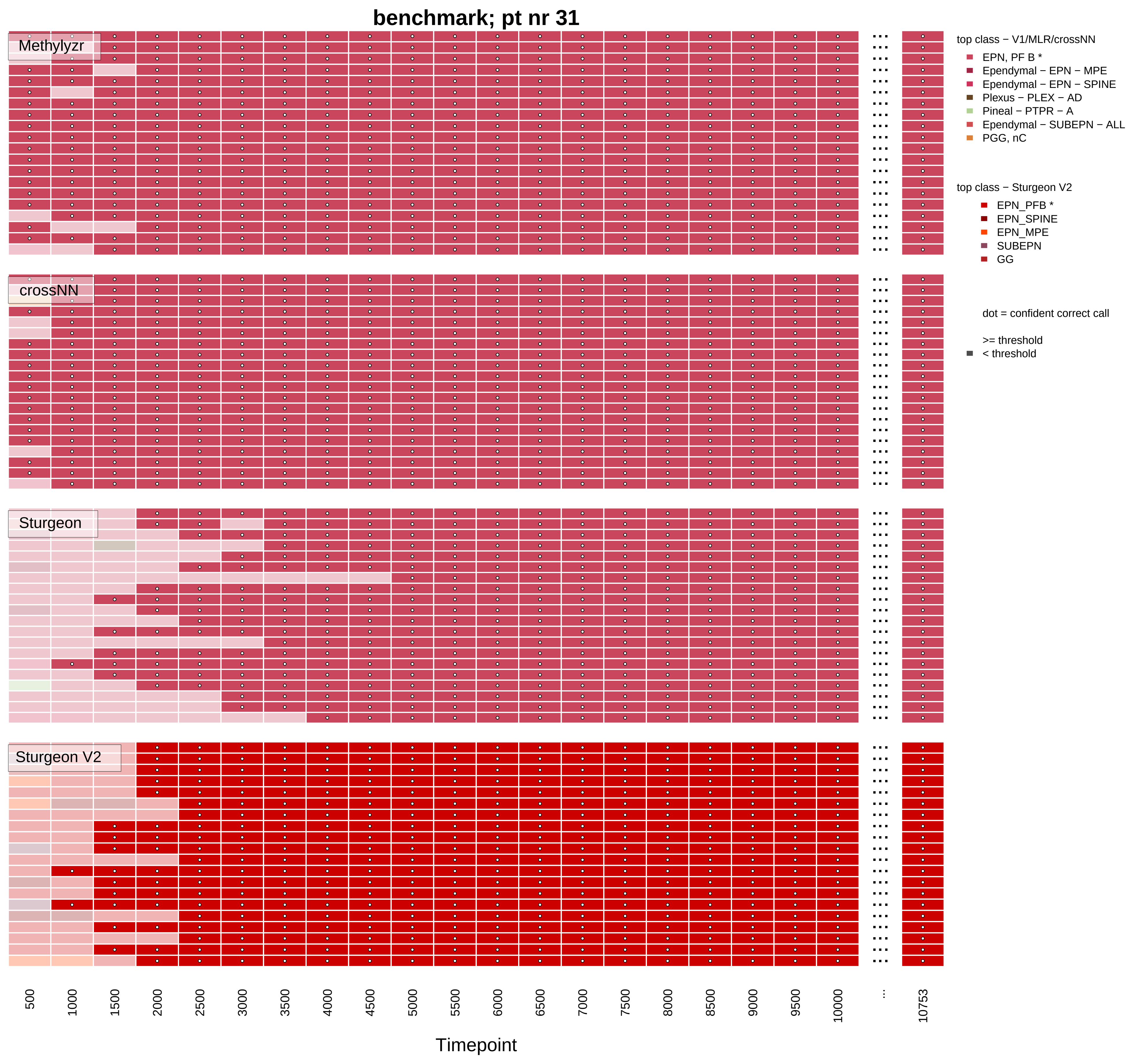

benchmark; pt nr 31
Methylyzr
crossNN
Sturgeon
Sturgeon V2
500
1000
1500
2000
2500
3000
3500
4000
4500
5000
5500
6000
6500
7000
7500
8000
8500
9000
9500
10000
...
10753
Timepoint
top class − V1/MLR/crossNN
EPN, PF B *
Ependymal − EPN − MPE
Ependymal − EPN − SPINE
Plexus − PLEX − AD
Pineal − PTPR − A
Ependymal − SUBEPN − ALL
PGG, nC
top class − Sturgeon V2
EPN_PFB *
EPN_SPINE
EPN_MPE
SUBEPN
GG
dot = confident correct call
>= threshold
< threshold

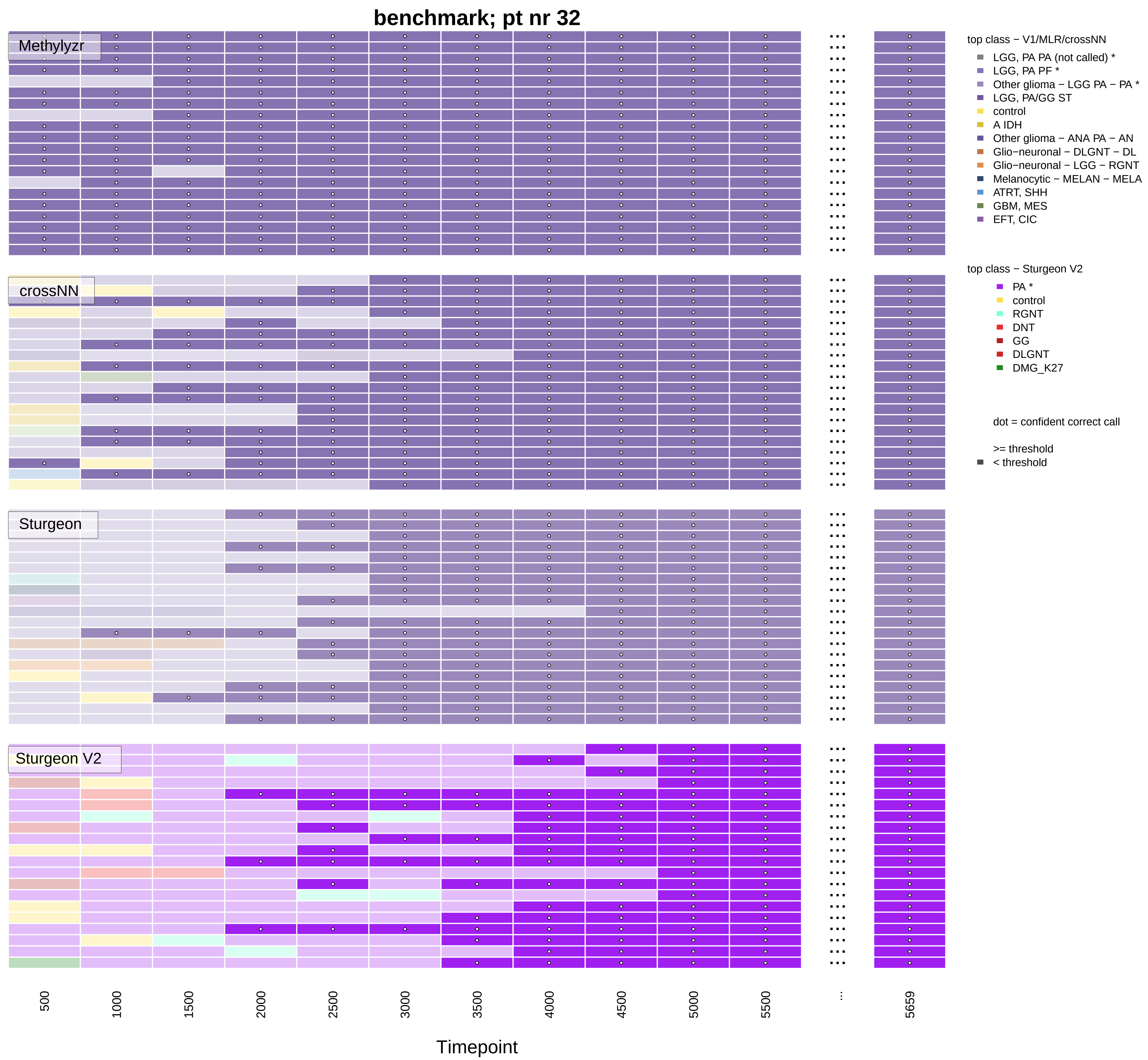
benchmark; pt nr 32
Methylyzr
crossNN
Sturgeon
Sturgeon V2
500
1000
1500
2000
2500
3000
3500
4000
4500
5000
5500
...
5659
Timepoint
top class − V1/MLR/crossNN
LGG, PA PA (not called) *
LGG, PA PF *
Other glioma − LGG PA − PA *
LGG, PA/GG ST
control
A IDH
Other glioma − ANA PA − AN
Glio−neuronal − DLGNT − DL
Glio−neuronal − LGG − RGNT
Melanocytic − MELAN − MELA
ATRT, SHH
GBM, MES
EFT, CIC
top class − Sturgeon V2
PA *
control
RGNT
DNT
GG
DLGNT
DMG_K27
dot = confident correct call
>= threshold
< threshold

### benchmark; pt nr 33

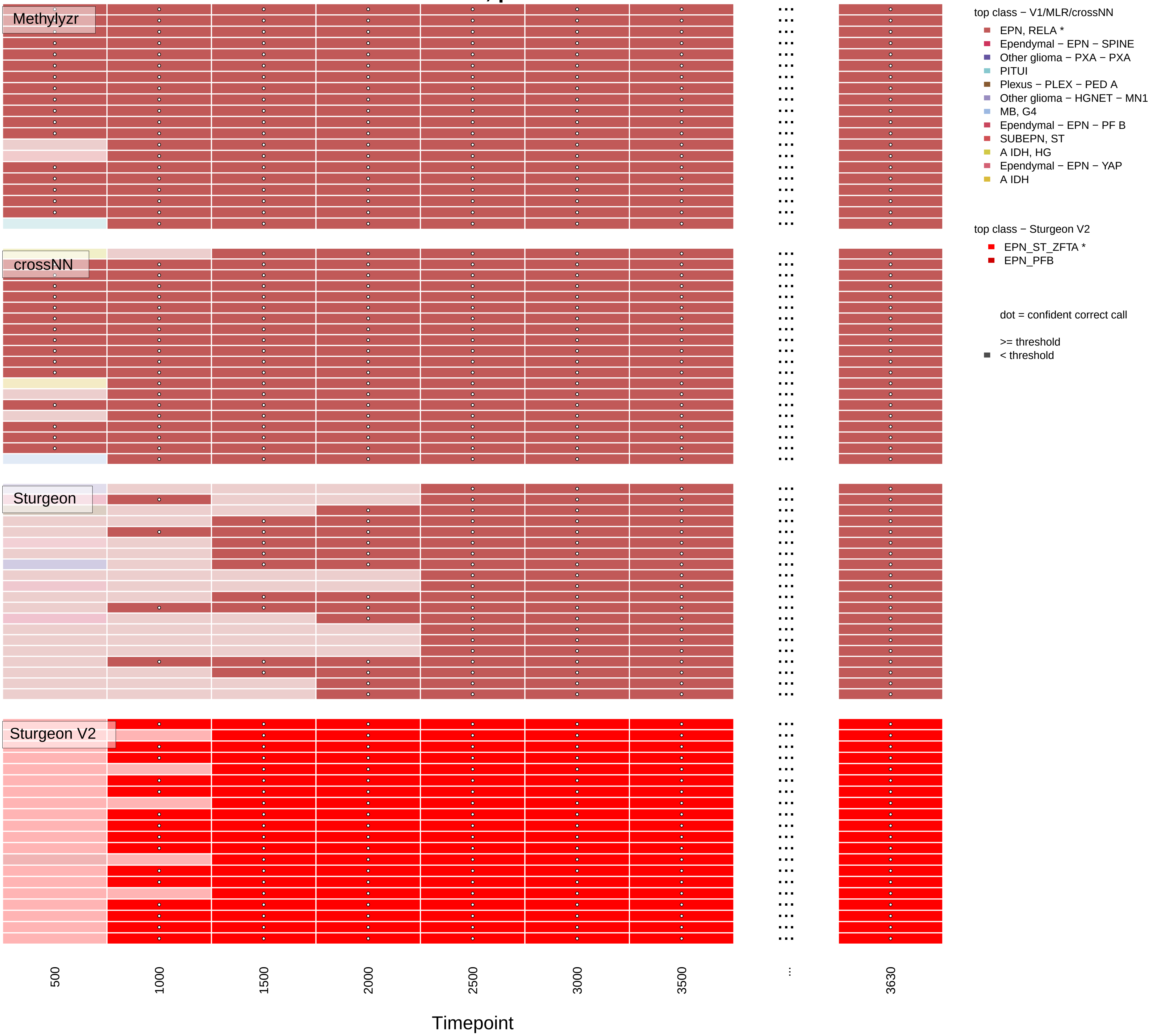

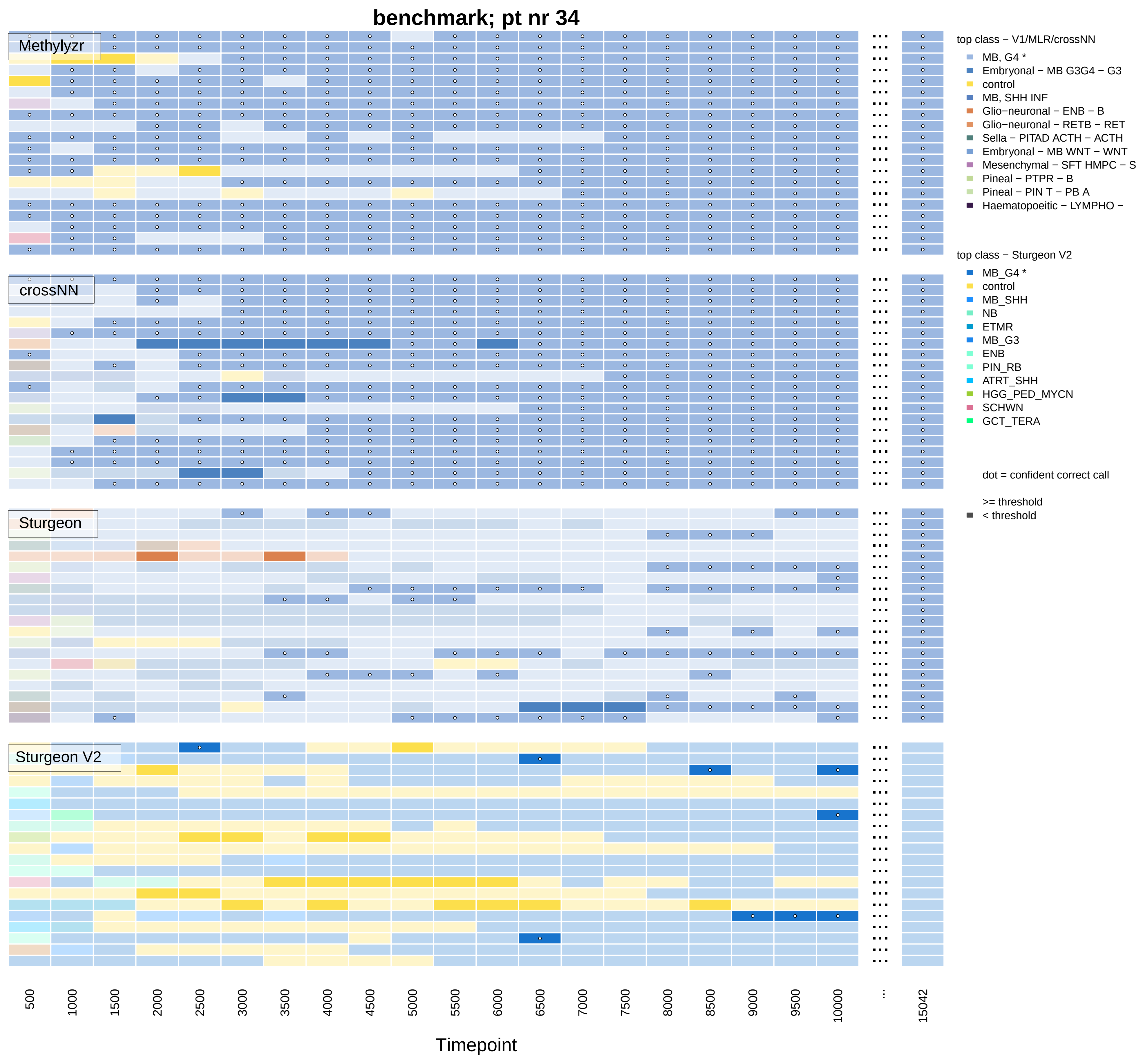
benchmark; pt nr 34
Methylyzr
crossNN
Sturgeon
Sturgeon V2
top class − V1/MLR/crossNN
MB, G4 *
Embryonal − MB G3G4 − G3
control
MB, SHH INF
Glio−neuronal − ENB − B
Glio−neuronal − RETB − RET
Sella − PITAD ACTH − ACTH
Embryonal − MB WNT − WNT
Mesenchymal − SFT HMPC − S
Pineal − PTPR − B
Pineal − PIN T − PB A
Haematopoeitic − LYMPHO −
top class − Sturgeon V2
MB_G4 *
control
MB_SHH
NB
ETMR
MB_G3
ENB
PIN_RB
ATRT_SHH
HGG_PED_MYCN
SCHWN
GCT_TERA
dot = confident correct call
>= threshold
< threshold
500
1000
1500
2000
2500
3000
3500
4000
4500
5000
5500
6000
6500
7000
7500
8000
8500
9000
9500
10000
...
15042
Timepoint

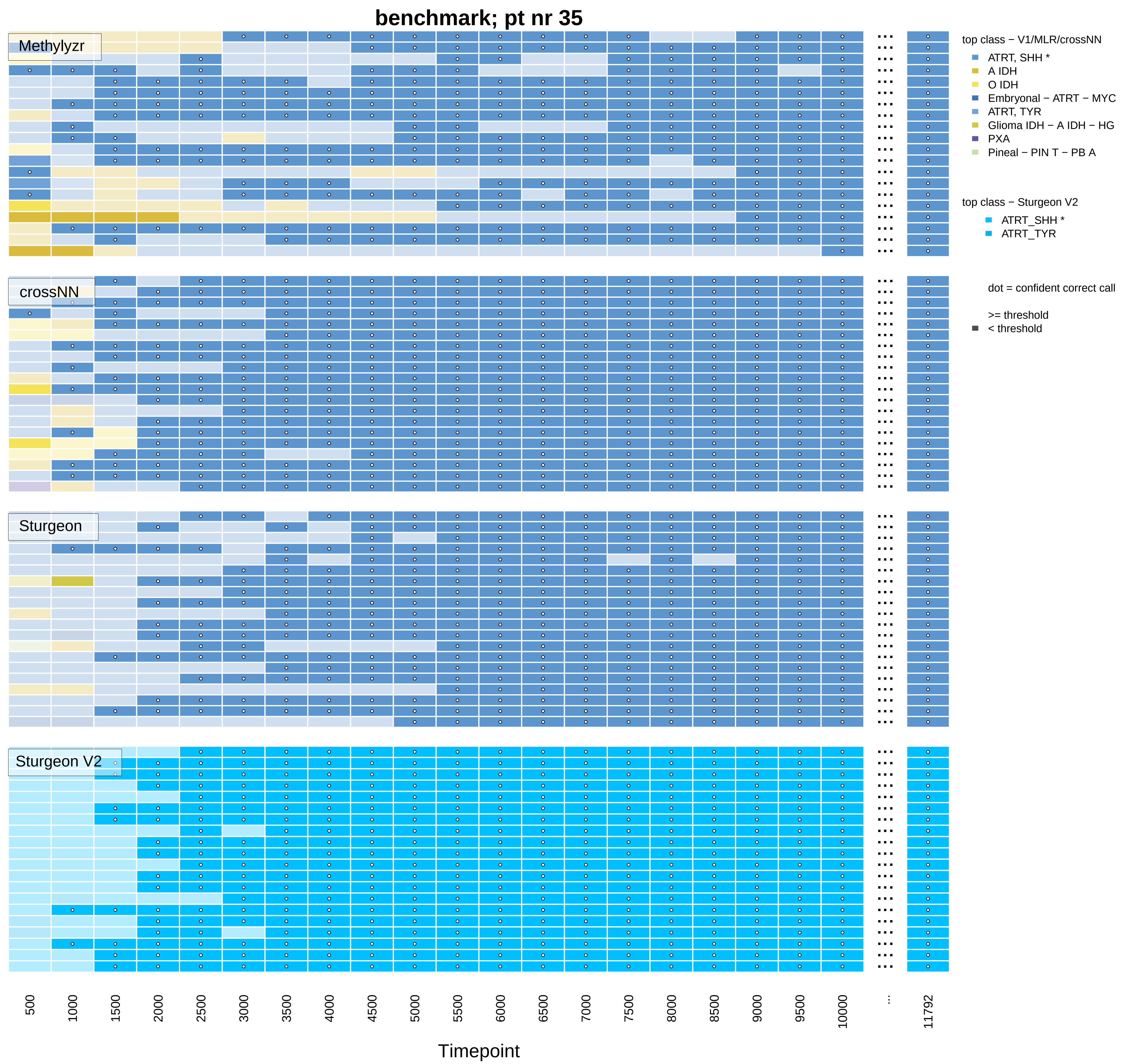

benchmark; pt nr 35
Methylyzr
crossNN
Sturgeon
Sturgeon V2
top class − V1/MLR/crossNN
ATRT, SHH *
A IDH
O IDH
Embryonal − ATRT − MYC
ATRT, TYR
Glioma IDH − A IDH − HG
PXA
Pineal − PIN T − PB A
top class − Sturgeon V2
ATRT_SHH *
ATRT_TYR
dot = confident correct call
>= threshold
< threshold
500
1000
1500
2000
2500
3000
3500
4000
4500
5000
5500
6000
6500
7000
7500
8000
8500
9000
9500
10000
...
11792
Timepoint

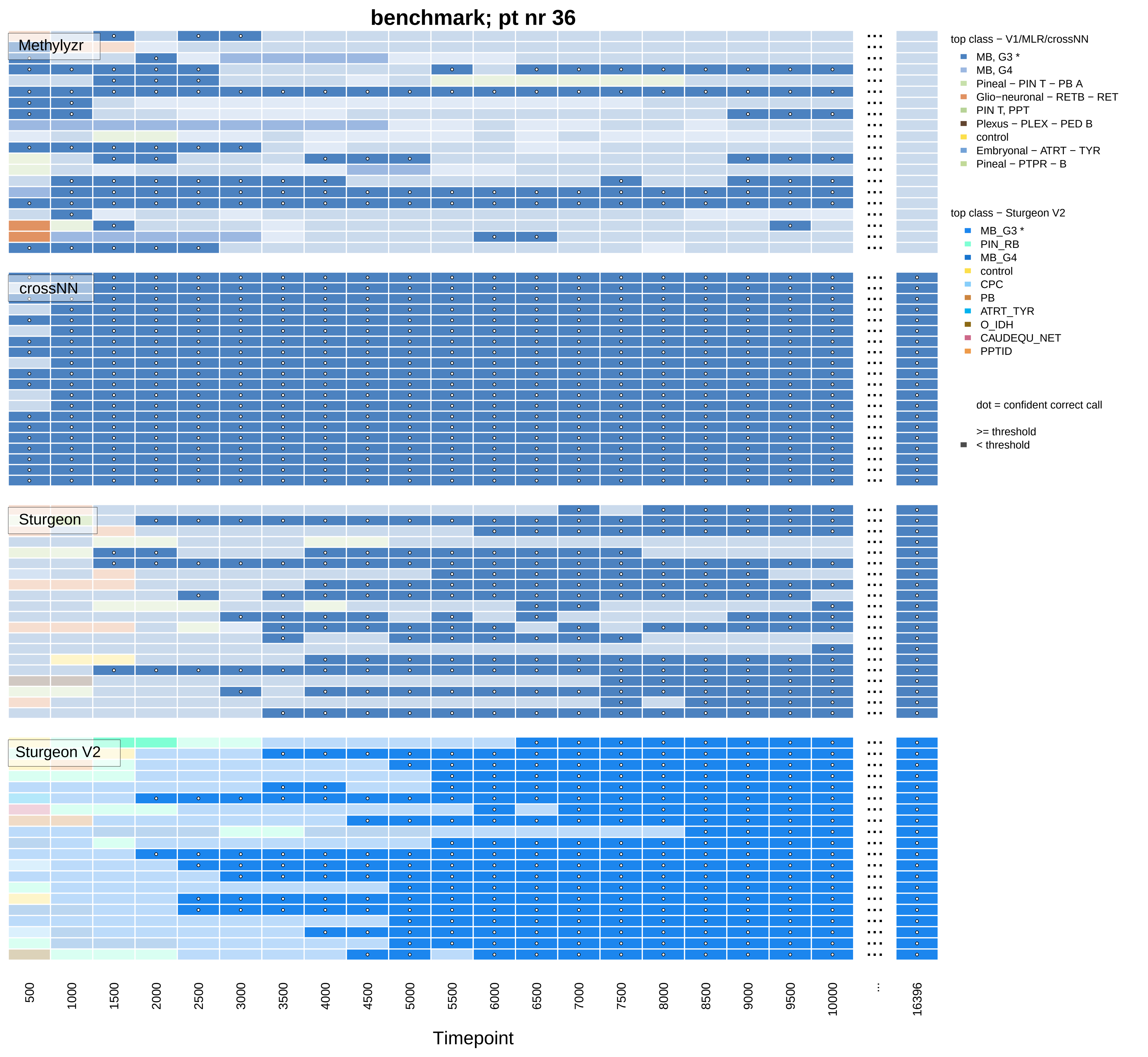

benchmark; pt nr 36
Methylyzr
crossNN
Sturgeon
Sturgeon V2
500
1000
1500
2000
2500
3000
3500
4000
4500
5000
5500
6000
6500
7000
7500
8000
8500
9000
9500
10000
...
16396
Timepoint
top class − V1/MLR/crossNN
MB, G3 *
MB, G4
Pineal − PIN T − PB A
Glio−neuronal − RETB − RET
PIN T, PPT
Plexus − PLEX − PED B
control
Embryonal − ATRT − TYR
Pineal − PTPR − B
top class − Sturgeon V2
MB_G3 *
PIN_RB
MB_G4
control
CPC
PB
ATRT_TYR
O_IDH
CAUDEQU_NET
PPTID
dot = confident correct call
>= threshold
< threshold

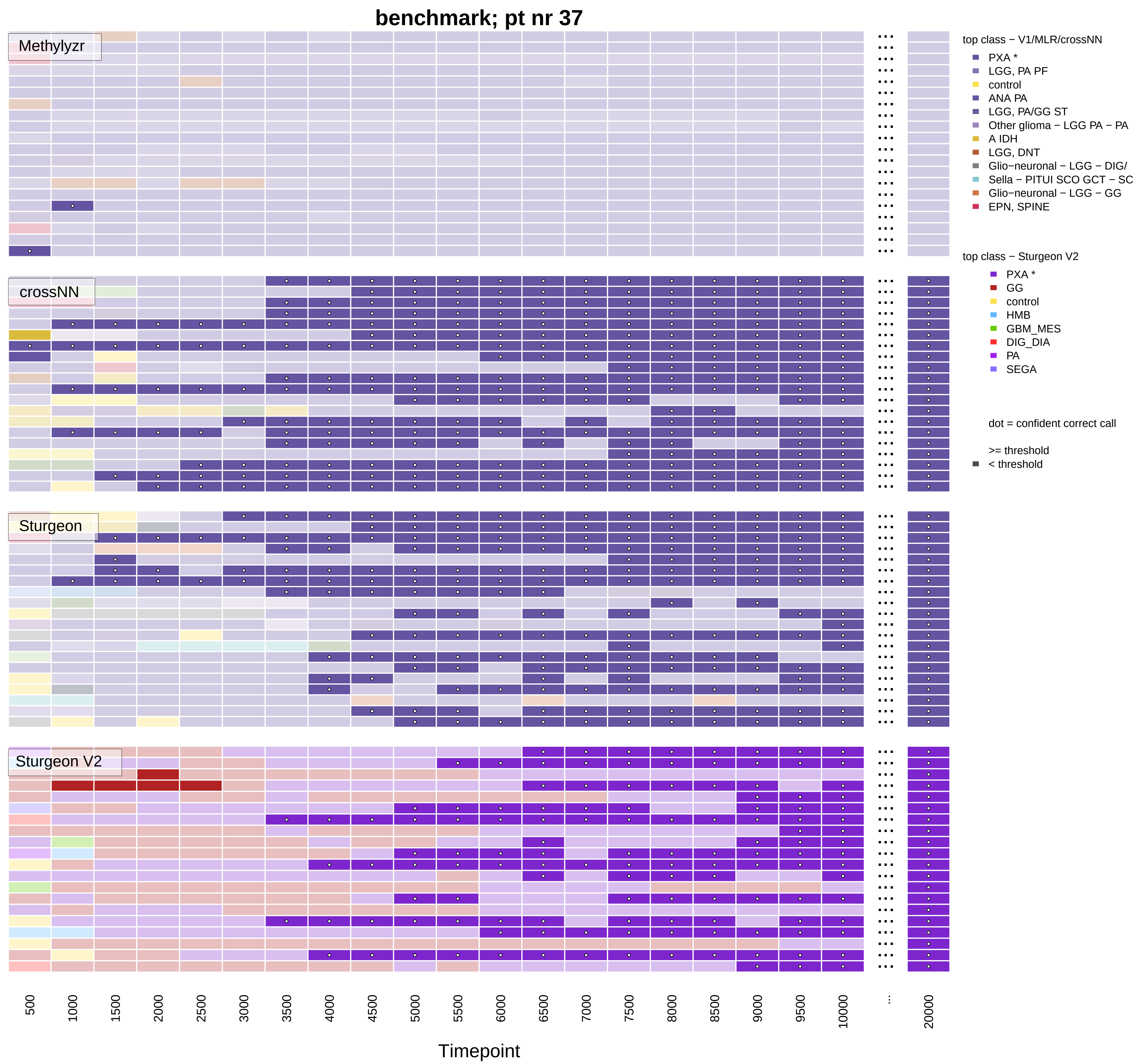

benchmark; pt nr 37
Methylyzr
crossNN
Sturgeon
Sturgeon V2
top class − V1/MLR/crossNN
PXA *
LGG, PA PF
control
ANA PA
LGG, PA/GG ST
Other glioma − LGG PA − PA
A IDH
LGG, DNT
Glio−neuronal − LGG − DIG/
Sella − PITUI SCO GCT − SC
Glio−neuronal − LGG − GG
EPN, SPINE
top class − Sturgeon V2
PXA *
GG
control
HMB
GBM_MES
DIG_DIA
PA
SEGA
dot = confident correct call
>= threshold
< threshold
500 1000 1500 2000 2500 3000 3500 4000 4500 5000 5500 6000 6500 7000 7500 8000 8500 9000 9500 10000 ... 20000
Timepoint

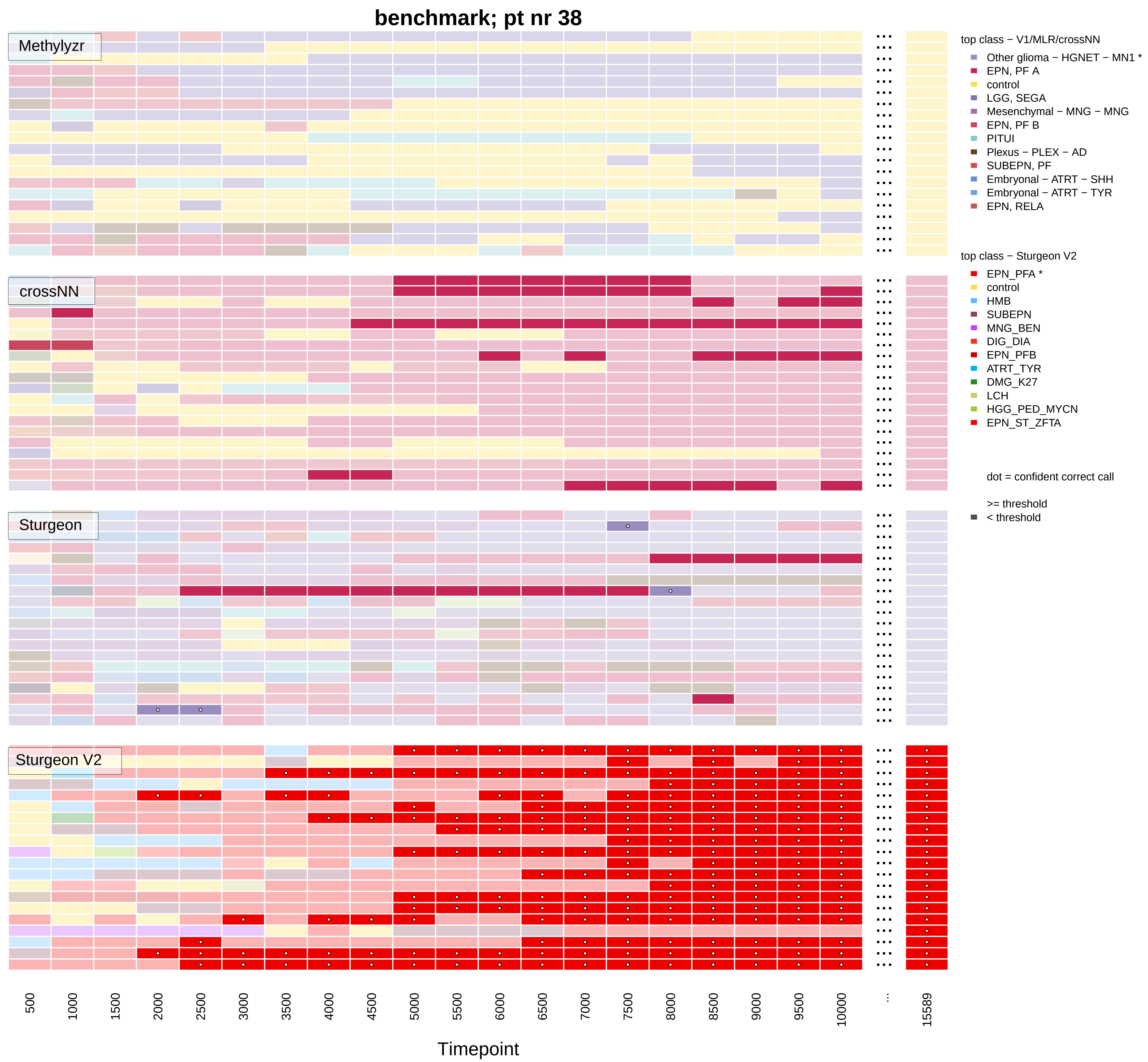

benchmark; pt nr 38
Methylyzr
crossNN
Sturgeon
Sturgeon V2
top class − V1/MLR/crossNN
Other glioma − HGNET − MN1 *
EPN, PF A
control
LGG, SEGA
Mesenchymal − MNG − MNG
EPN, PF B
PITUI
Plexus − PLEX − AD
SUBEPN, PF
Embryonal − ATRT − SHH
Embryonal − ATRT − TYR
EPN, RELA
top class − Sturgeon V2
EPN_PFA *
control
HMB
SUBEPN
MNG_BEN
DIG_DIA
EPN_PFB
ATRT_TYR
DMG_K27
LCH
HGG_PED_MYCN
EPN_ST_ZFTA
dot = confident correct call
>= threshold
< threshold
500
1000
1500
2000
2500
3000
3500
4000
4500
5000
5500
6000
6500
7000
7500
8000
8500
9000
9500
10000
...
15589
Timepoint

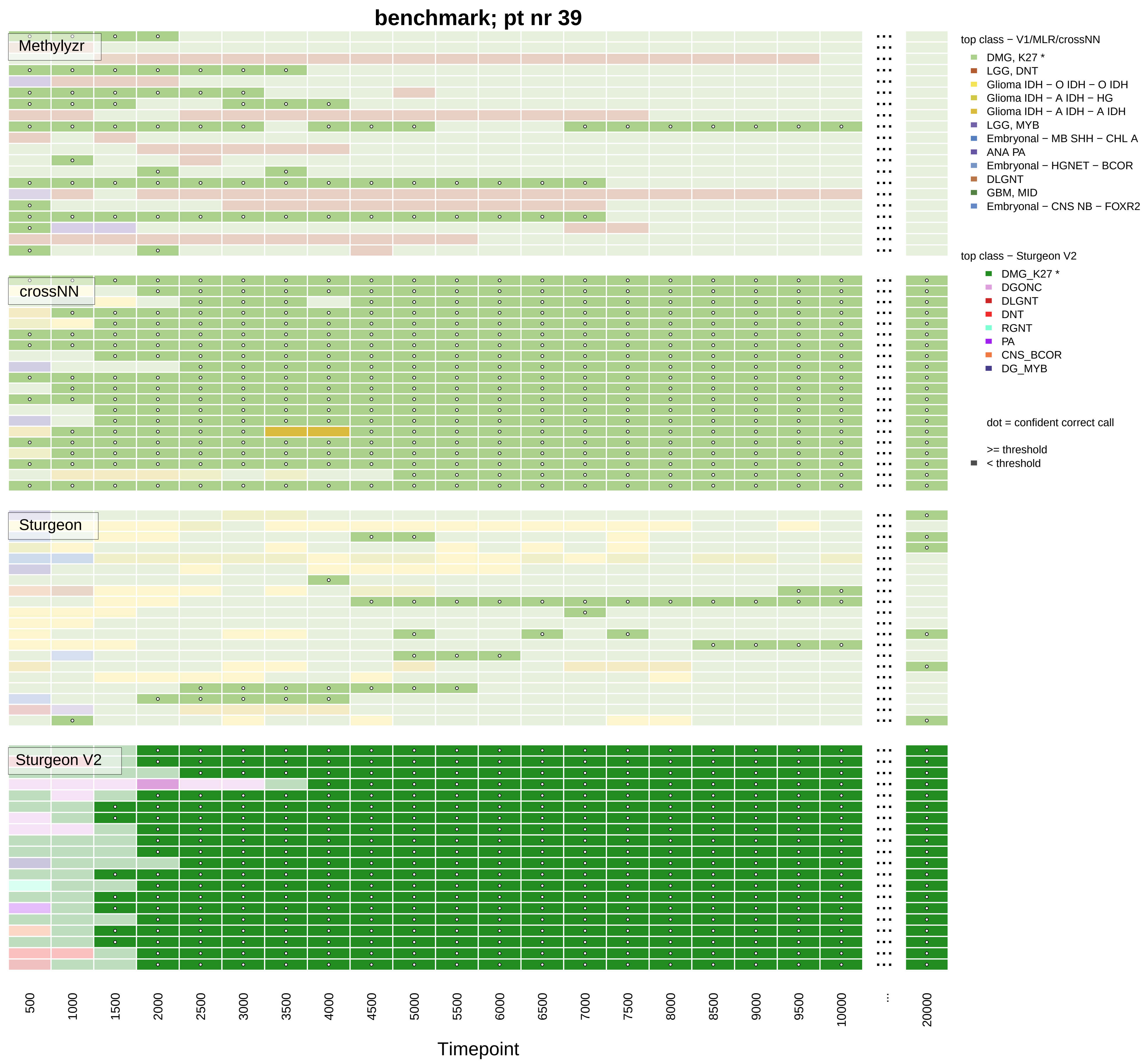

benchmark; pt nr 39
Methylyzr
crossNN
Sturgeon
Sturgeon V2
500
1000
1500
2000
2500
3000
3500
4000
4500
5000
5500
6000
6500
7000
7500
8000
8500
9000
9500
10000
...
20000
Timepoint
top class − V1/MLR/crossNN
DMG, K27 *
LGG, DNT
Glioma IDH − O IDH − O IDH
Glioma IDH − A IDH − HG
Glioma IDH − A IDH − A IDH
LGG, MYB
Embryonal − MB SHH − CHL A
ANA PA
Embryonal − HGNET − BCOR
DLGNT
GBM, MID
Embryonal − CNS NB − FOXR2
top class − Sturgeon V2
DMG_K27 *
DGONC
DLGNT
DNT
RGNT
PA
CNS_BCOR
DG_MYB
dot = confident correct call
>= threshold
< threshold

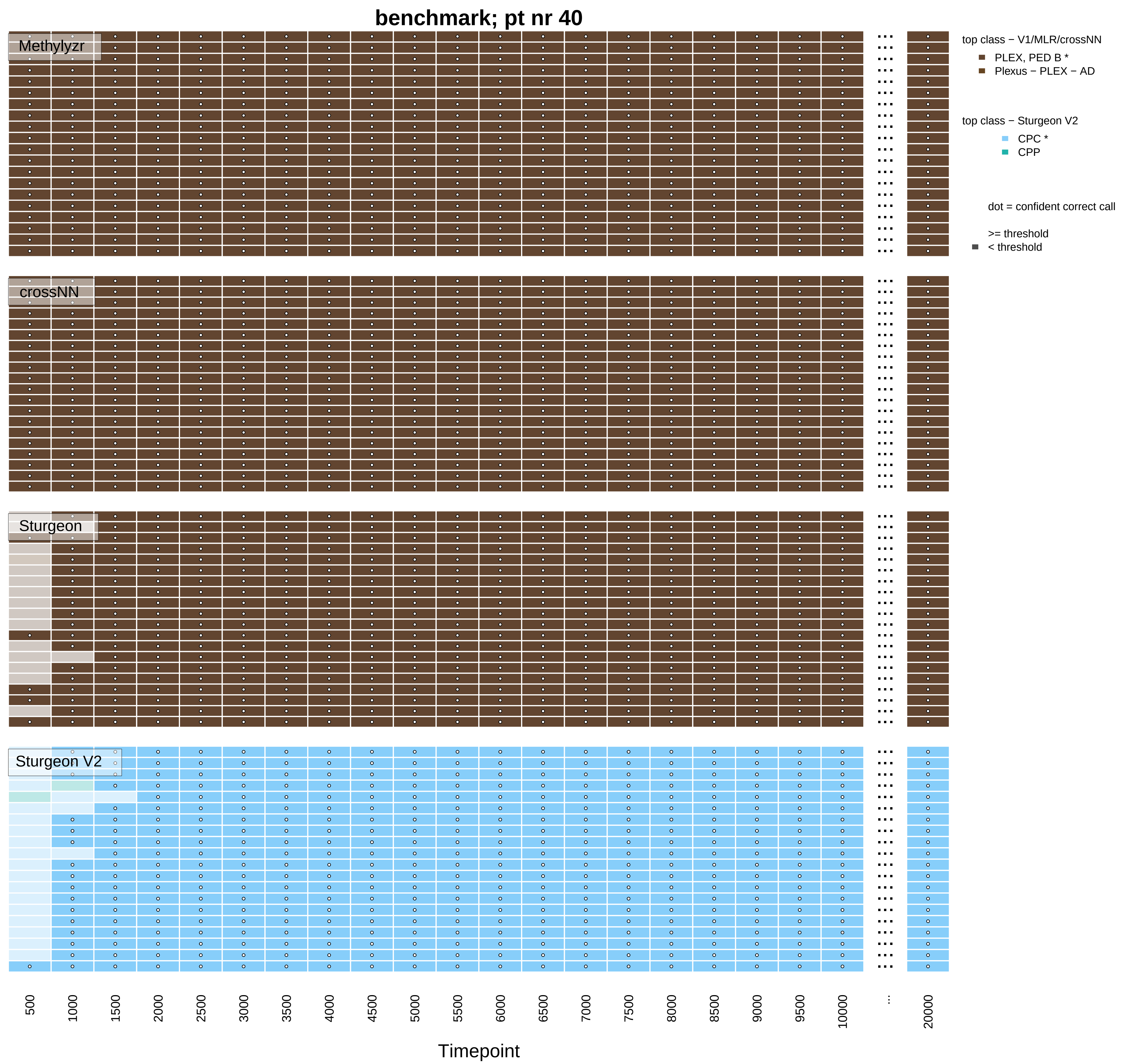
benchmark; pt nr 40
Methylyzr
crossNN
Sturgeon
Sturgeon V2
top class − V1/MLR/crossNN
PLEX, PED B *
Plexus − PLEX − AD
top class − Sturgeon V2
CPC *
CPP
dot = confident correct call
>= threshold
< threshold
500
1000
1500
2000
2500
3000
3500
4000
4500
5000
5500
6000
6500
7000
7500
8000
8500
9000
9500
10000
...
20000
Timepoint

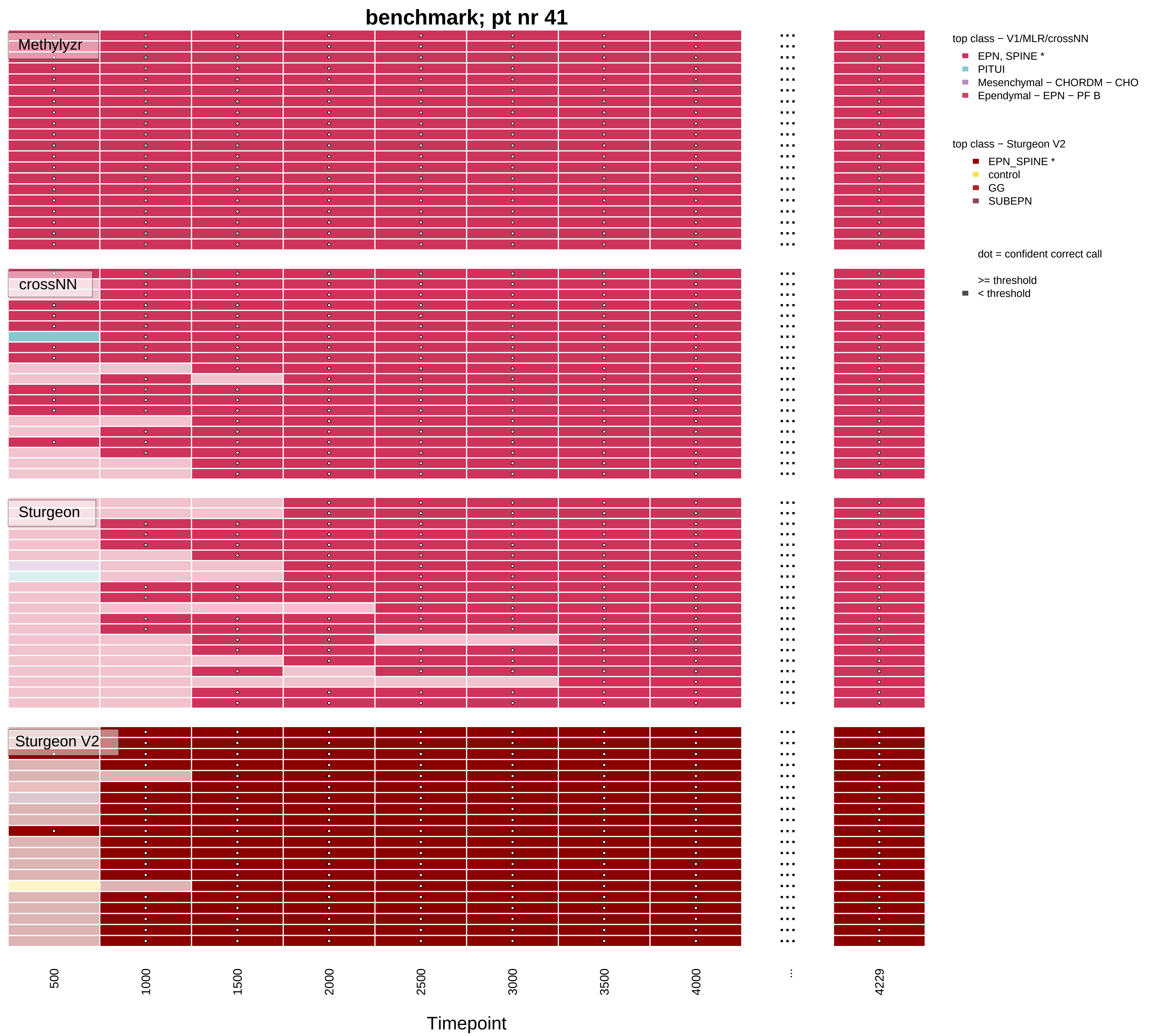

benchmark; pt nr 41
Methylyzr
crossNN
Sturgeon
Sturgeon V2
top class − V1/MLR/crossNN
EPN, SPINE *
PITUI
Mesenchymal − CHORDM − CHO
Ependymal − EPN − PF B
top class − Sturgeon V2
EPN_SPINE *
control
GG
SUBEPN
dot = confident correct call
>= threshold
< threshold
500
1000
1500
2000
2500
3000
3500
4000
...
4229
Timepoint

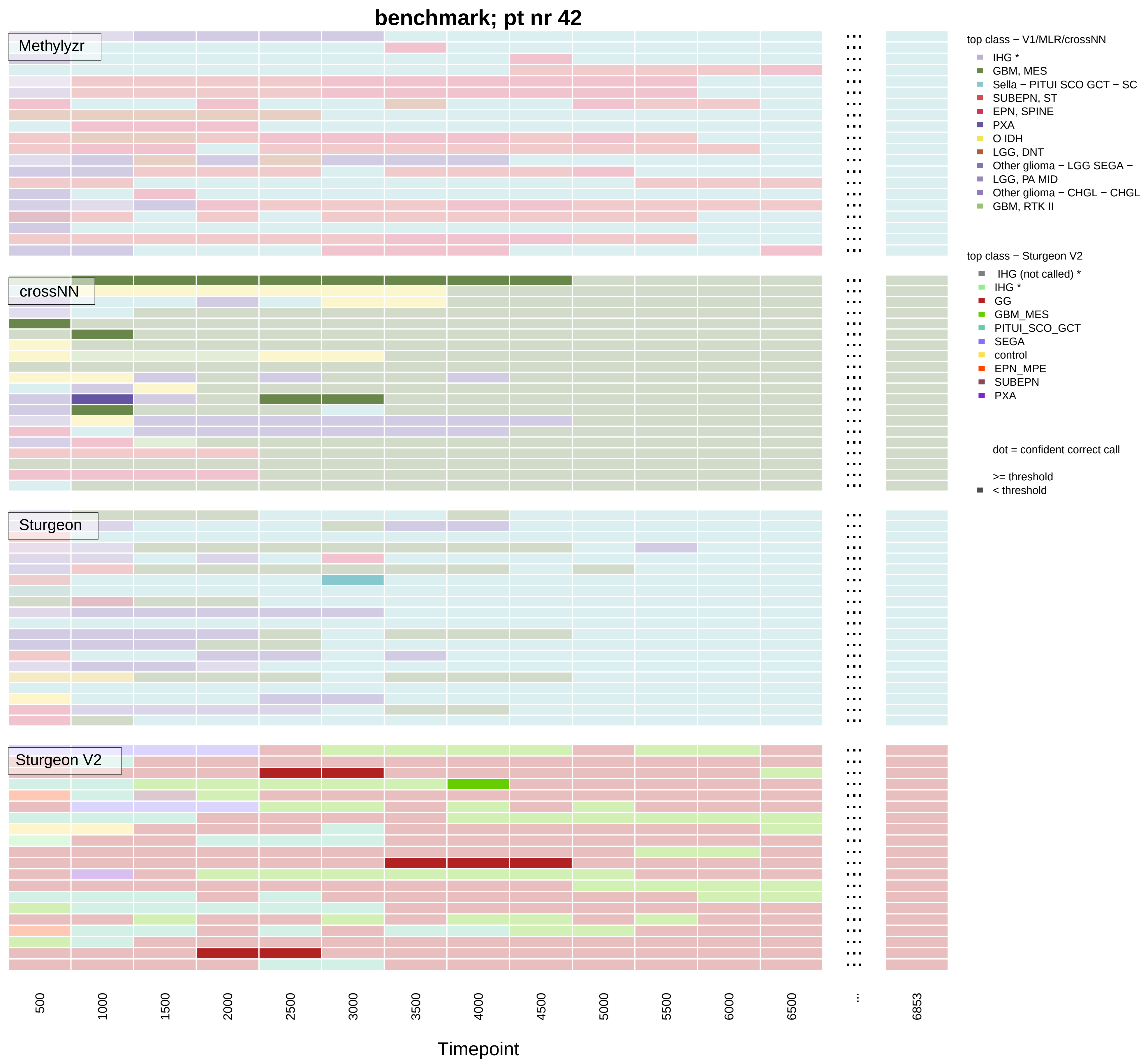

benchmark; pt nr 42
Methylyzr
crossNN
Sturgeon
Sturgeon V2
top class − V1/MLR/crossNN
IHG *
GBM, MES
Sella − PITUI SCO GCT − SC
SUBEPN, ST
EPN, SPINE
PXA
O IDH
LGG, DNT
Other glioma − LGG SEGA −
LGG, PA MID
Other glioma − CHGL − CHGL
GBM, RTK II
top class − Sturgeon V2
IHG (not called) *
IHG *
GG
GBM_MES
PITUI_SCO_GCT
SEGA
control
EPN_MPE
SUBEPN
PXA
dot = confident correct call
>= threshold
< threshold
500
1000
1500
2000
2500
3000
3500
4000
4500
5000
5500
6000
6500
...
6853
Timepoint

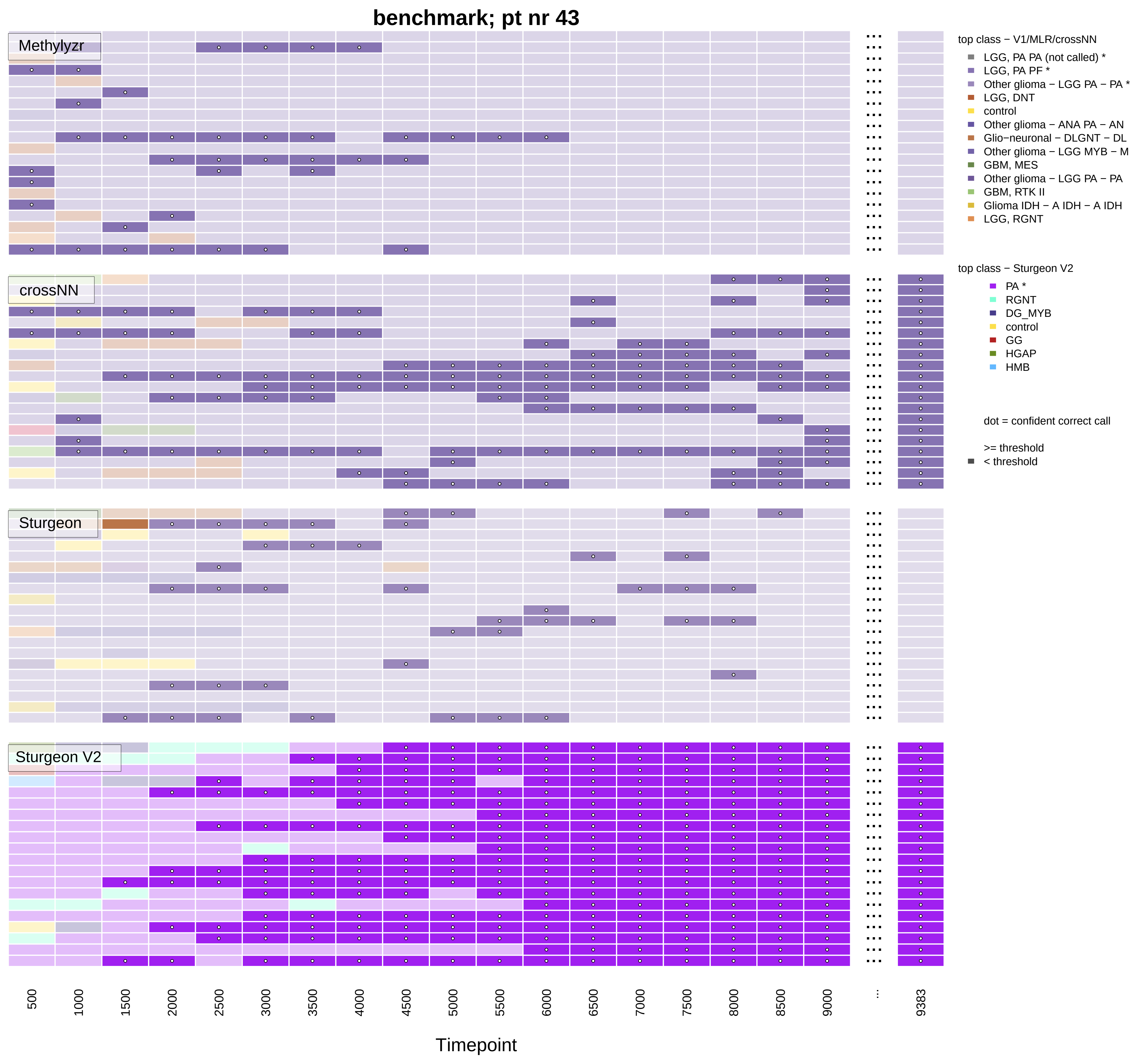

benchmark; pt nr 43
Methylyzr
crossNN
Sturgeon
Sturgeon V2
top class − V1/MLR/crossNN
LGG, PA PA (not called) *
LGG, PA PF *
Other glioma − LGG PA − PA *
LGG, DNT
control
Other glioma − ANA PA − AN
Glio−neuronal − DLGNT − DL
Other glioma − LGG MYB − M
GBM, MES
Other glioma − LGG PA − PA
GBM, RTK II
Glioma IDH − A IDH − A IDH
LGG, RGNT
top class − Sturgeon V2
PA *
RGNT
DG_MYB
control
GG
HGAP
HMB
dot = confident correct call
>= threshold
< threshold
500
1000
1500
2000
2500
3000
3500
4000
4500
5000
5500
6000
6500
7000
7500
8000
8500
9000
...
9383
Timepoint

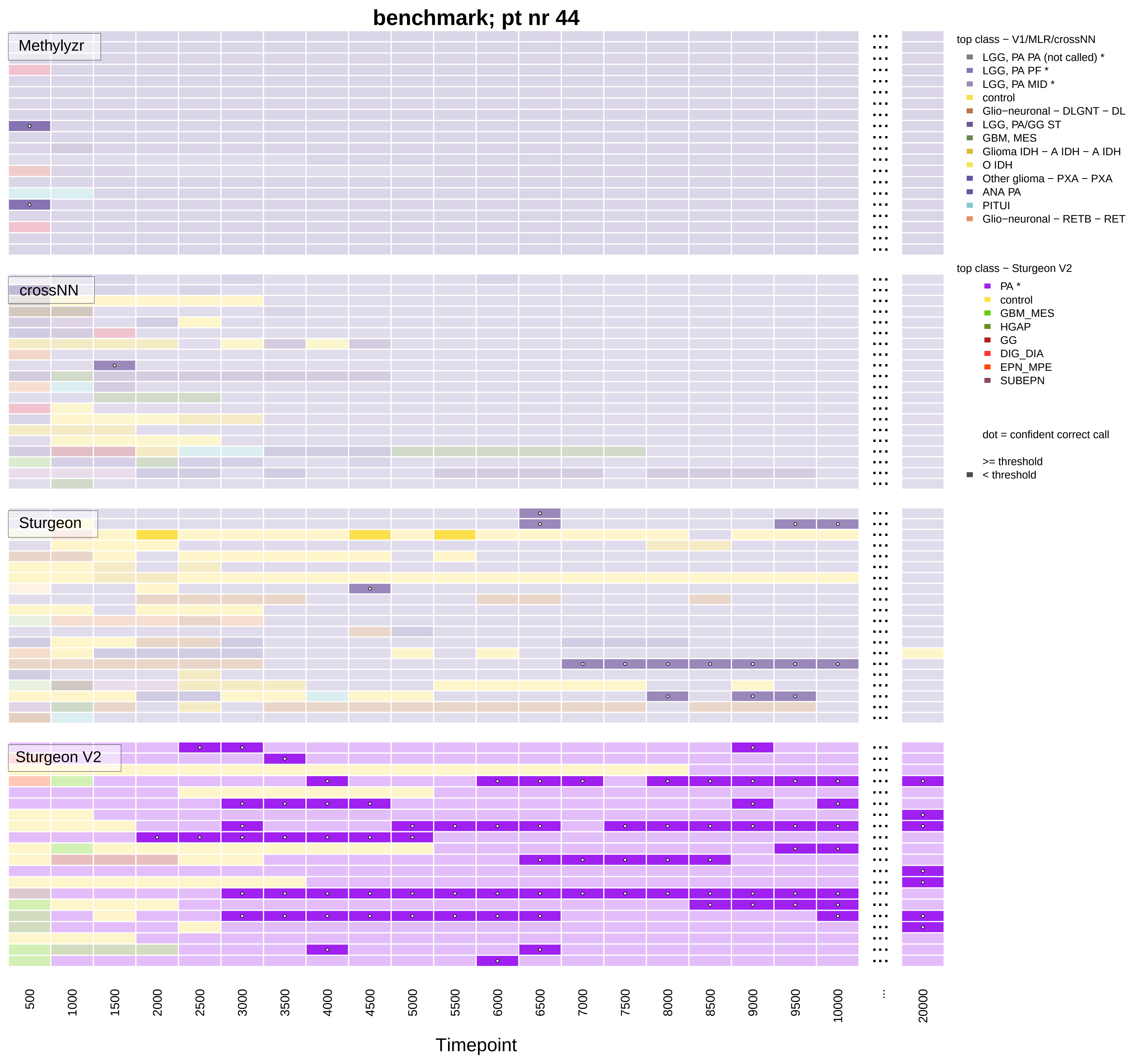

benchmark; pt nr 44
Methylyzr
crossNN
Sturgeon
Sturgeon V2
top class − V1/MLR/crossNN
LGG, PA PA (not called) *
LGG, PA PF *
LGG, PA MID *
control
Glio−neuronal − DLGNT − DL
LGG, PA/GG ST
GBM, MES
Glioma IDH − A IDH − A IDH
O IDH
Other glioma − PXA − PXA
ANA PA
PITUI
Glio−neuronal − RETB − RET
top class − Sturgeon V2
PA *
control
GBM_MES
HGAP
GG
DIG_DIA
EPN_MPE
SUBEPN
dot = confident correct call
>= threshold
< threshold
500
1000
1500
2000
2500
3000
3500
4000
4500
5000
5500
6000
6500
7000
7500
8000
8500
9000
9500
10000
...
20000
Timepoint

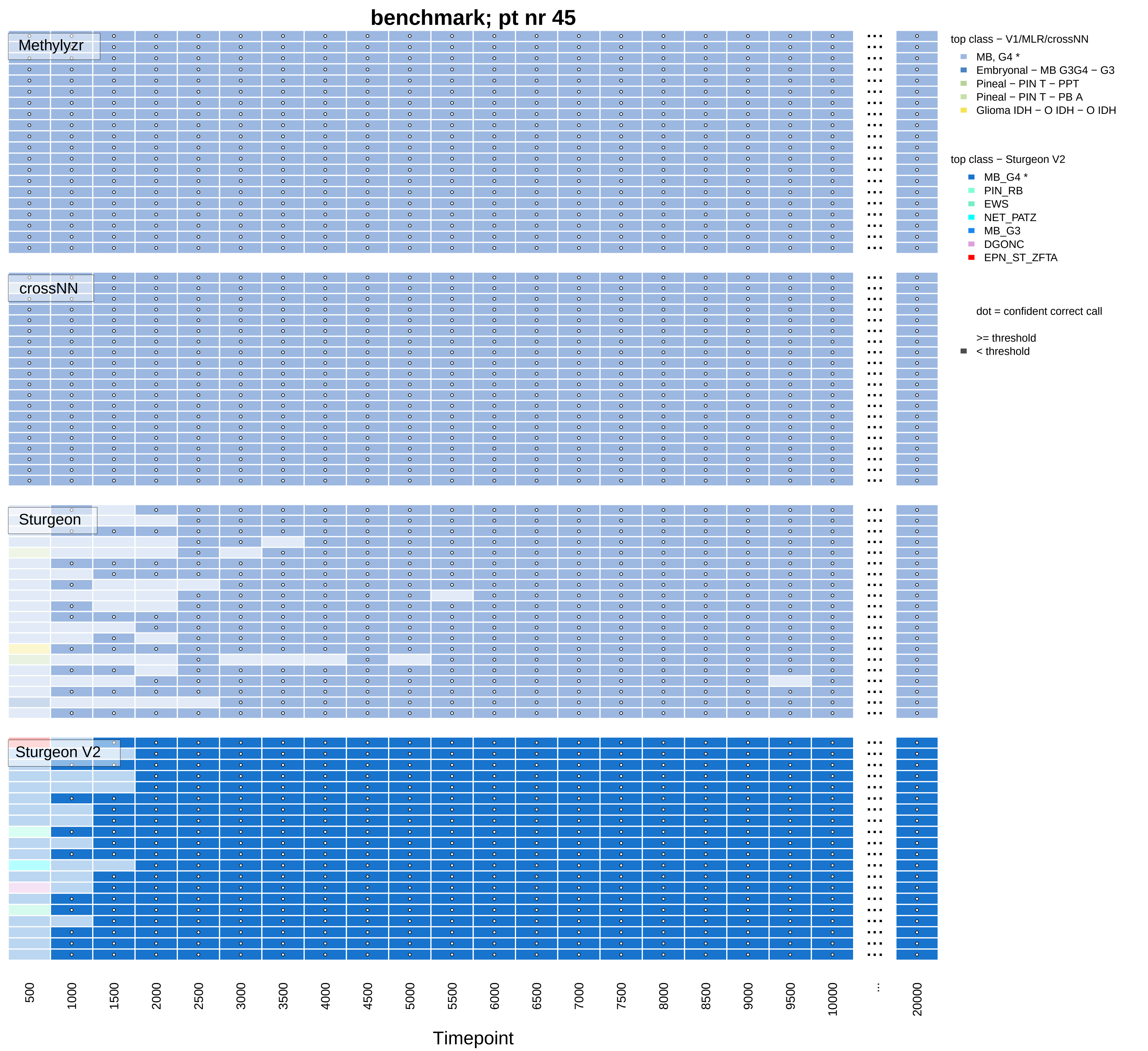

benchmark; pt nr 45
Methylyzr
crossNN
Sturgeon
Sturgeon V2
top class − V1/MLR/crossNN
MB, G4 *
Embryonal − MB G3G4 − G3
Pineal − PIN T − PPT
Pineal − PIN T − PB A
Glioma IDH − O IDH − O IDH
top class − Sturgeon V2
MB_G4 *
PIN_RB
EWS
NET_PATZ
MB_G3
DGONC
EPN_ST_ZFTA
dot = confident correct call
>= threshold
< threshold
500
1000
1500
2000
2500
3000
3500
4000
4500
5000
5500
6000
6500
7000
7500
8000
8500
9000
9500
10000
...
20000
Timepoint

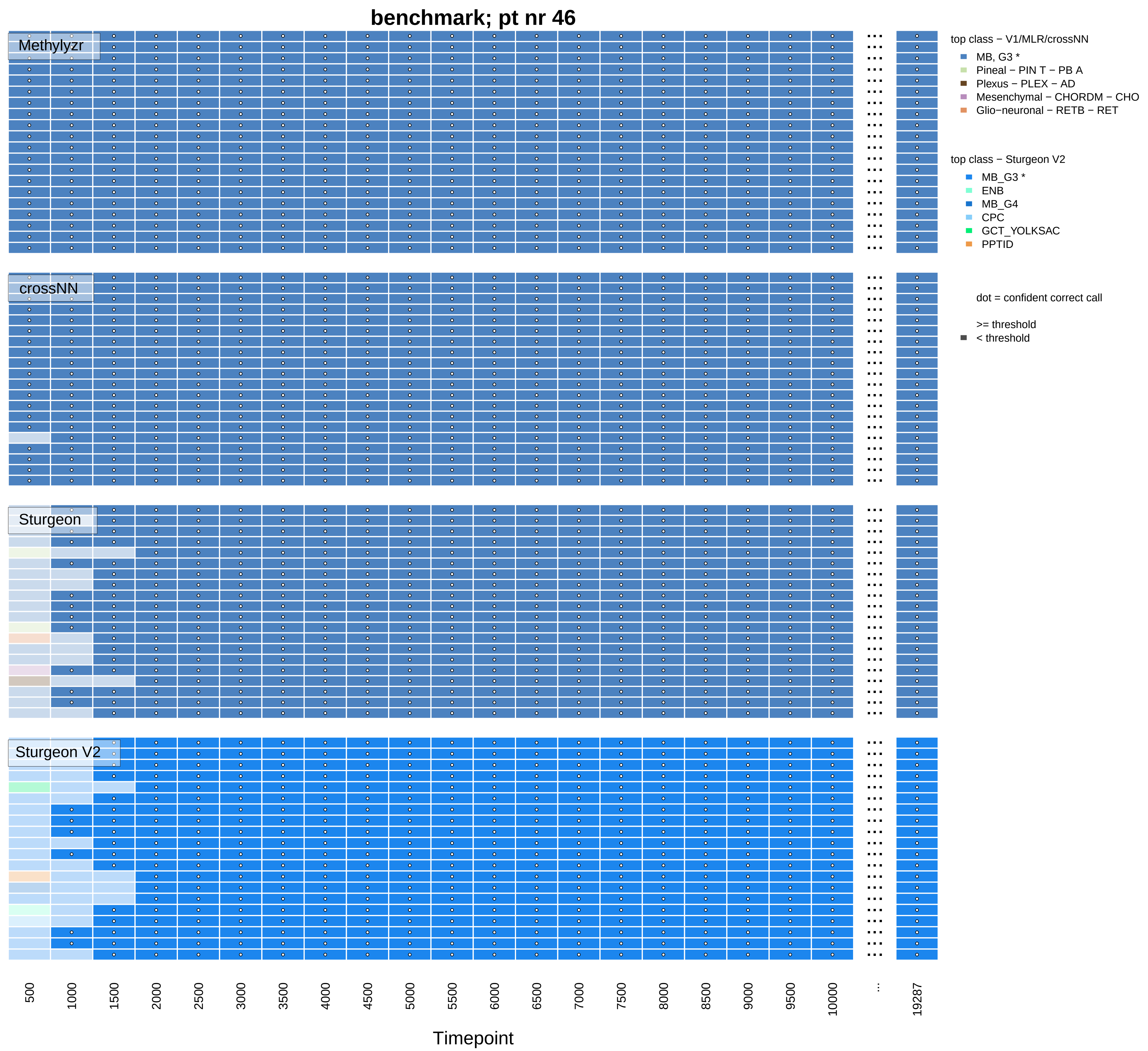
benchmark; pt nr 46
Methylyzr
crossNN
Sturgeon
Sturgeon V2
top class − V1/MLR/crossNN
MB, G3 *
Pineal − PIN T − PB A
Plexus − PLEX − AD
Mesenchymal − CHORDM − CHO
Glio−neuronal − RETB − RET
top class − Sturgeon V2
MB_G3 *
ENB
MB_G4
CPC
GCT_YOLKSAC
PPTID
dot = confident correct call
>= threshold
< threshold
500
1000
1500
2000
2500
3000
3500
4000
4500
5000
5500
6000
6500
7000
7500
8000
8500
9000
9500
10000
...
19287
Timepoint

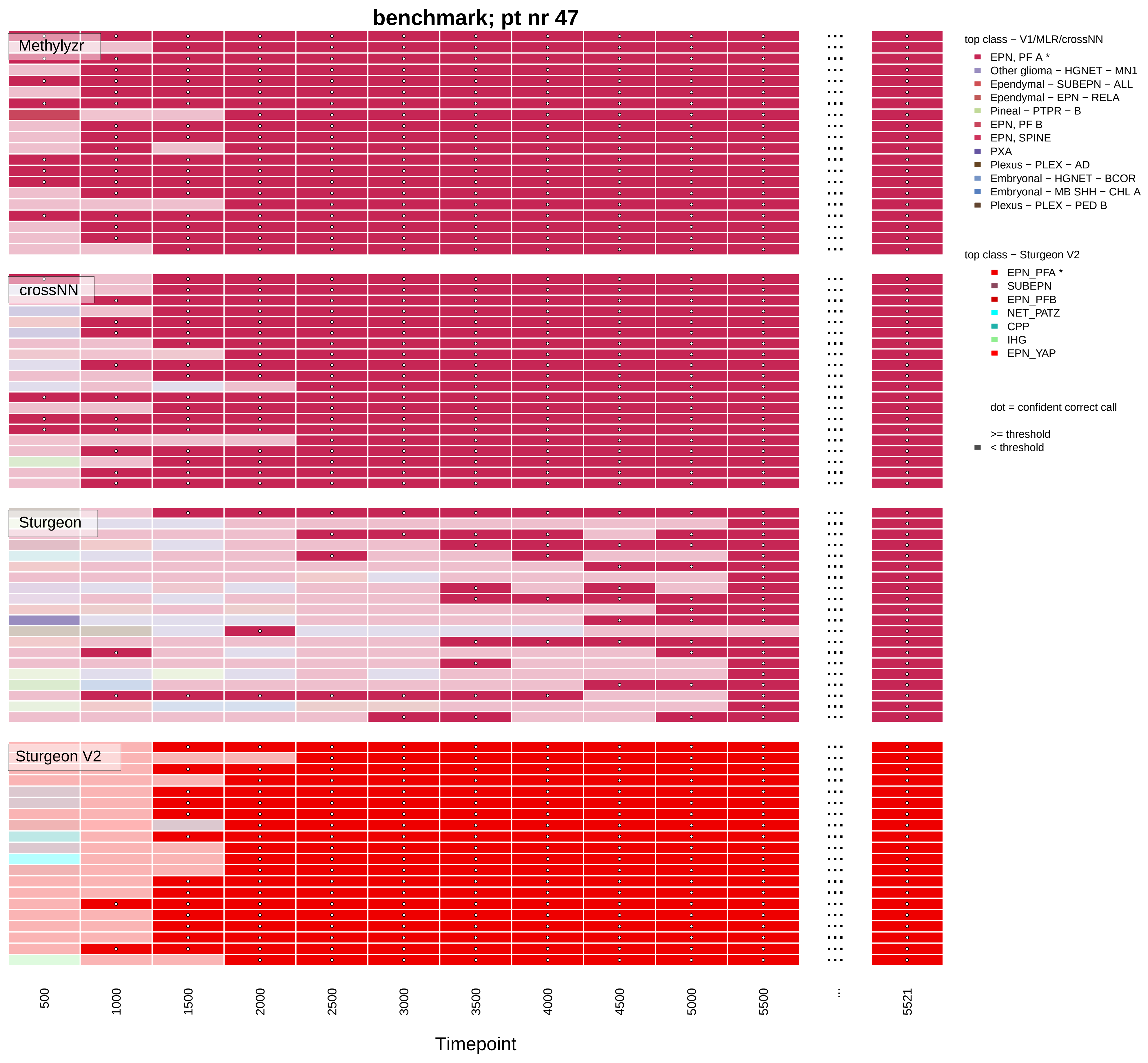

benchmark; pt nr 47
Methylyzr
crossNN
Sturgeon
Sturgeon V2
500
1000
1500
2000
2500
3000
3500
4000
4500
5000
5500
...
5521
Timepoint
top class − V1/MLR/crossNN
EPN, PF A *
Other glioma − HGNET − MN1
Ependymal − SUBEPN − ALL
Ependymal − EPN − RELA
Pineal − PTPR − B
EPN, PF B
EPN, SPINE
PXA
Plexus − PLEX − AD
Embryonal − HGNET − BCOR
Embryonal − MB SHH − CHL A
Plexus − PLEX − PED B
top class − Sturgeon V2
EPN_PFA *
SUBEPN
EPN_PFB
NET_PATZ
CPP
IHG
EPN_YAP
dot = confident correct call
>= threshold
< threshold

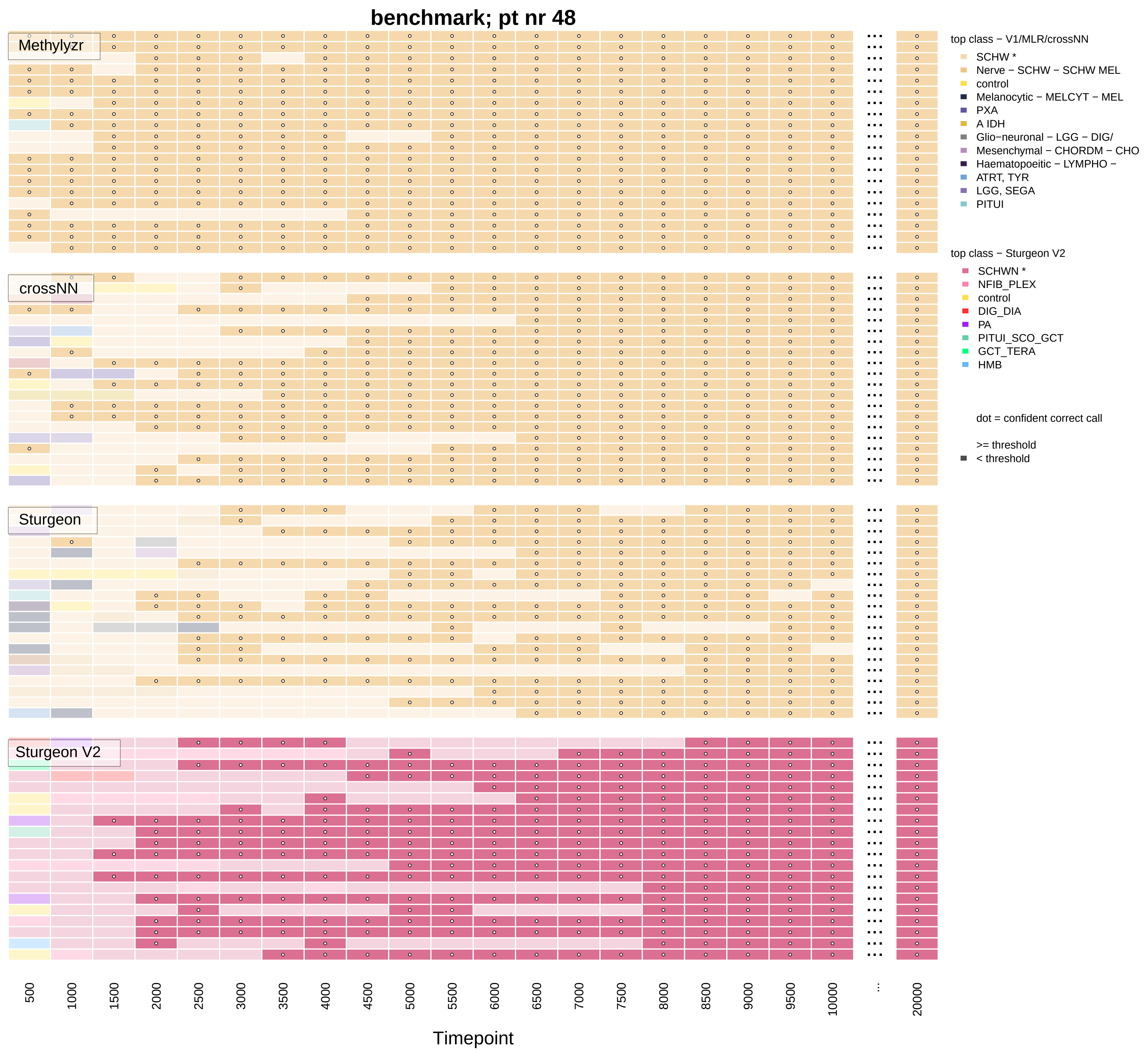

benchmark; pt nr 48
Methylyzr
crossNN
Sturgeon
Sturgeon V2
top class − V1/MLR/crossNN
SCHW *
Nerve − SCHW − SCHW MEL
control
Melanocytic − MELCYT − MEL
PXA
A IDH
Glio−neuronal − LGG − DIG/
Mesenchymal − CHORDM − CHO
Haematopoeitic − LYMPHO −
ATRT, TYR
LGG, SEGA
PITUI
top class − Sturgeon V2
SCHWN *
NFIB_PLEX
control
DIG_DIA
PA
PITUI_SCO_GCT
GCT_TERA
HMB
dot = confident correct call
>= threshold
< threshold
500
1000
1500
2000
2500
3000
3500
4000
4500
5000
5500
6000
6500
7000
7500
8000
8500
9000
9500
10000
...
20000
Timepoint

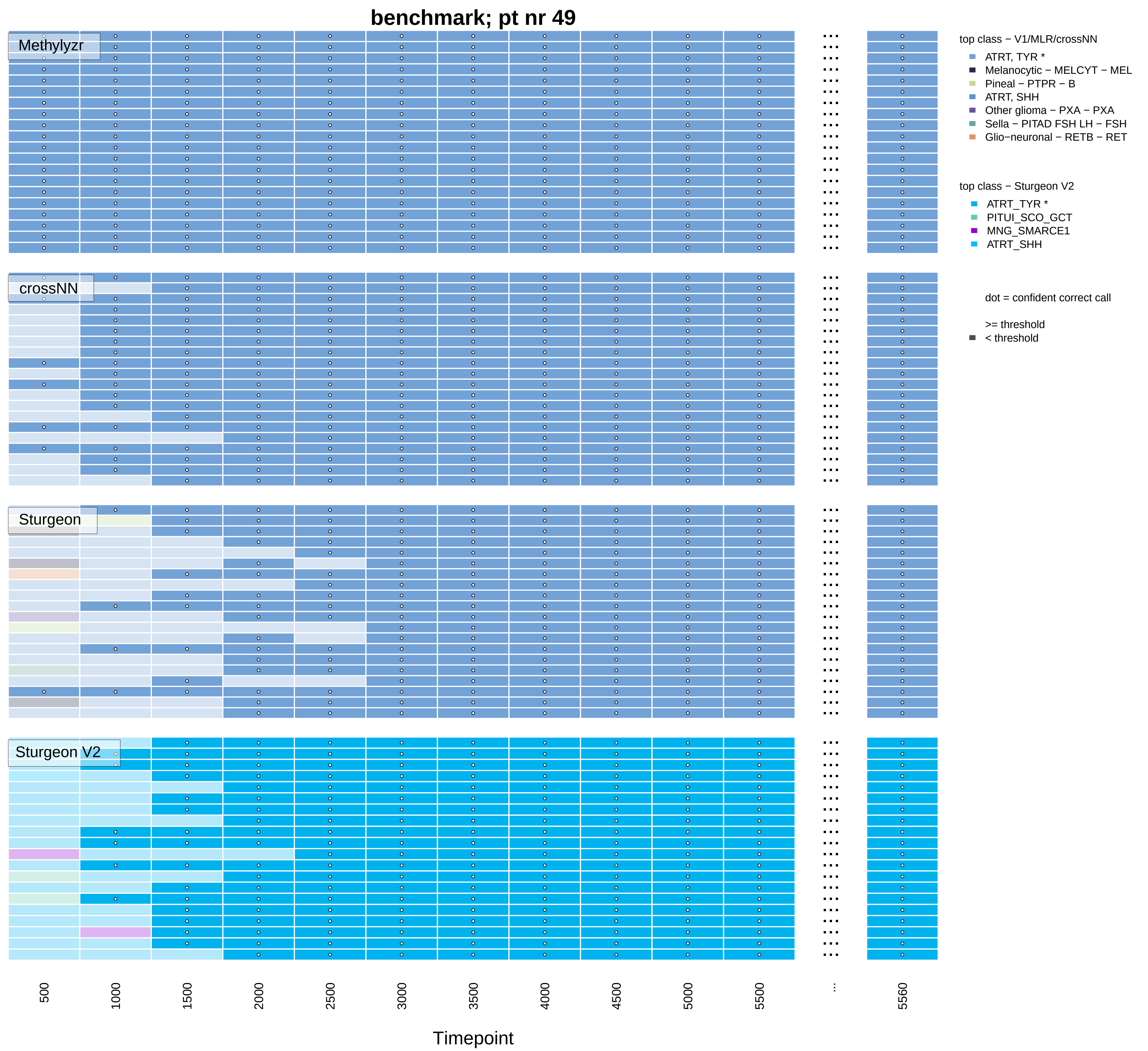
benchmark; pt nr 49
Methylyzr
crossNN
Sturgeon
Sturgeon V2
top class − V1/MLR/crossNN
ATRT, TYR *
Melanocytic − MELCYT − MEL
Pineal − PTPR − B
ATRT, SHH
Other glioma − PXA − PXA
Sella − PITAD FSH LH − FSH
Glio−neuronal − RETB − RET
top class − Sturgeon V2
ATRT_TYR *
PITUI_SCO_GCT
MNG_SMARCE1
ATRT_SHH
dot = confident correct call
>= threshold
< threshold
500
1000
1500
2000
2500
3000
3500
4000
4500
5000
5500
...
5560
Timepoint

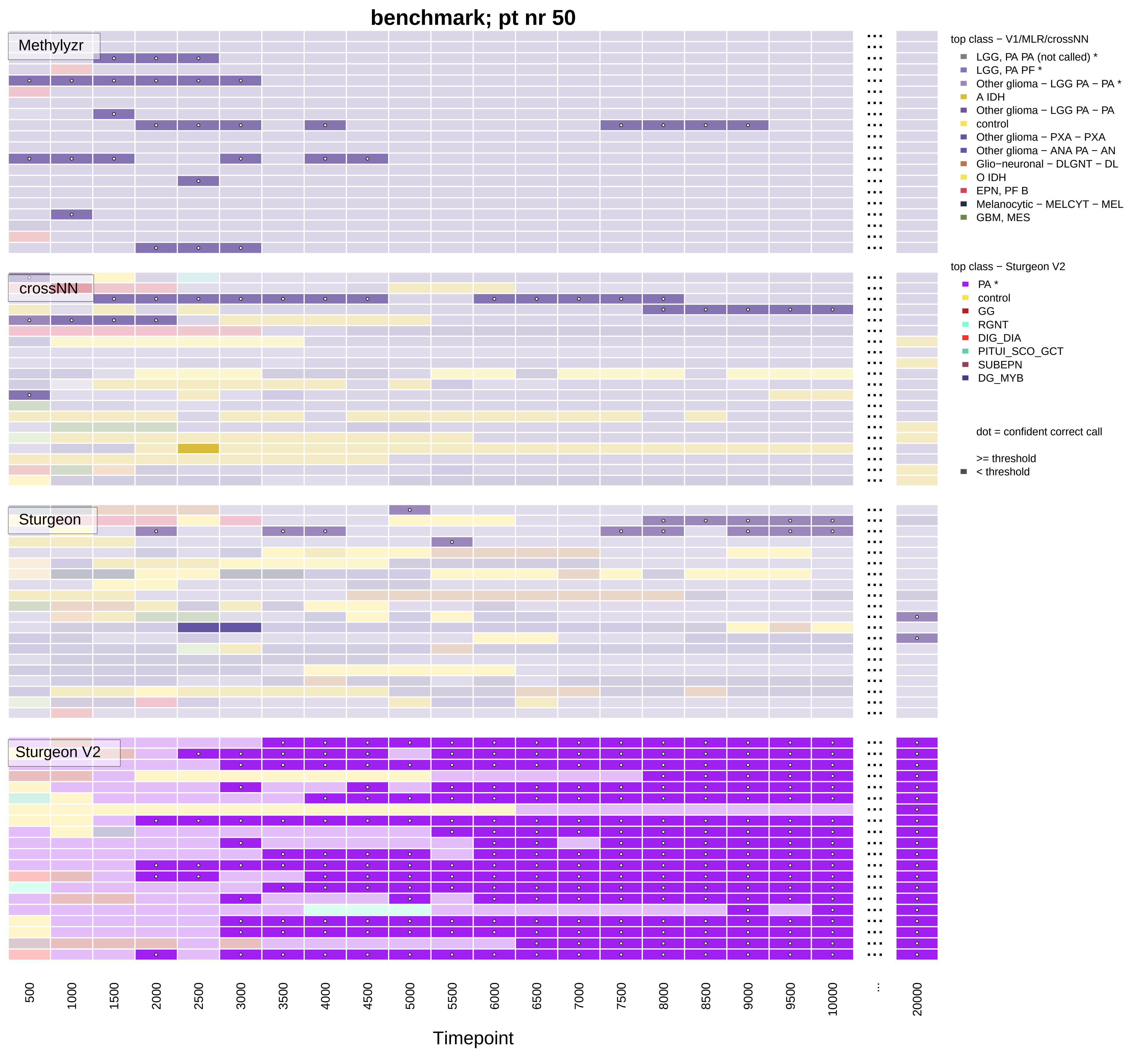

benchmark; pt nr 50
Methylyzr
crossNN
Sturgeon
Sturgeon V2
500
1000
1500
2000
2500
3000
3500
4000
4500
5000
5500
6000
6500
7000
7500
8000
8500
9000
9500
10000
...
20000
Timepoint
top class − V1/MLR/crossNN
LGG, PA PA (not called) *
LGG, PA PF *
Other glioma − LGG PA − PA *
A IDH
Other glioma − LGG PA − PA
control
Other glioma − PXA − PXA
Other glioma − ANA PA − AN
Glio−neuronal − DLGNT − DL
O IDH
EPN, PF B
Melanocytic − MELCYT − MEL
GBM, MES
top class − Sturgeon V2
PA *
control
GG
RGNT
DIG_DIA
PITUI_SCO_GCT
SUBEPN
DG_MYB
dot = confident correct call
>= threshold
< threshold

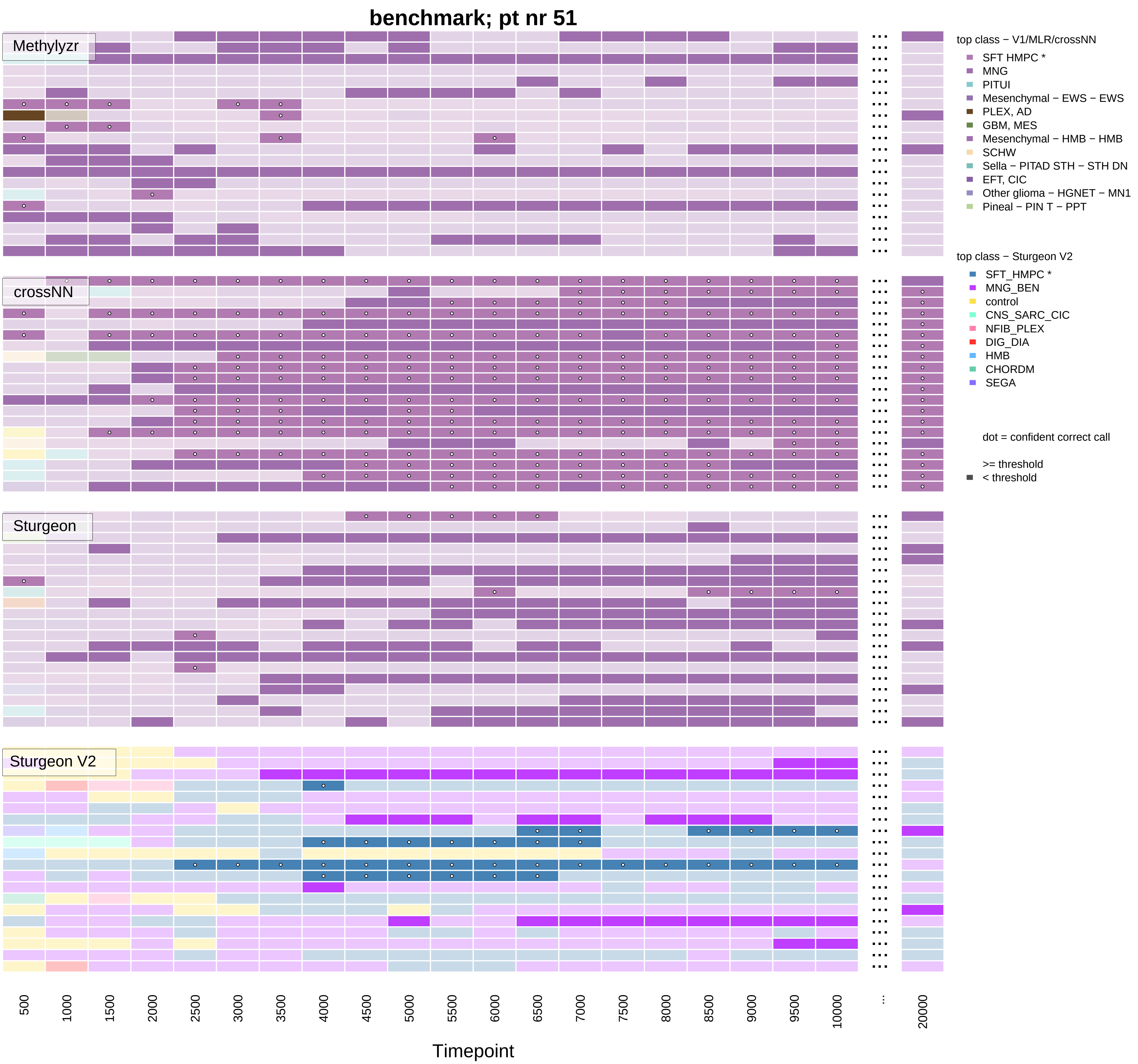

benchmark; pt nr 51
Methylyzr
crossNN
Sturgeon
Sturgeon V2
500
1000
1500
2000
2500
3000
3500
4000
4500
5000
5500
6000
6500
7000
7500
8000
8500
9000
9500
10000
...
20000
Timepoint
top class − V1/MLR/crossNN
SFT HMPC *
MNG
PITUI
Mesenchymal − EWS − EWS
PLEX, AD
GBM, MES
Mesenchymal − HMB − HMB
SCHW
Sella − PITAD STH − STH DN
EFT, CIC
Other glioma − HGNET − MN1
Pineal − PIN T − PPT
top class − Sturgeon V2
SFT_HMPC *
MNG_BEN
control
CNS_SARC_CIC
NFIB_PLEX
DIG_DIA
HMB
CHORDM
SEGA
dot = confident correct call
>= threshold
< threshold

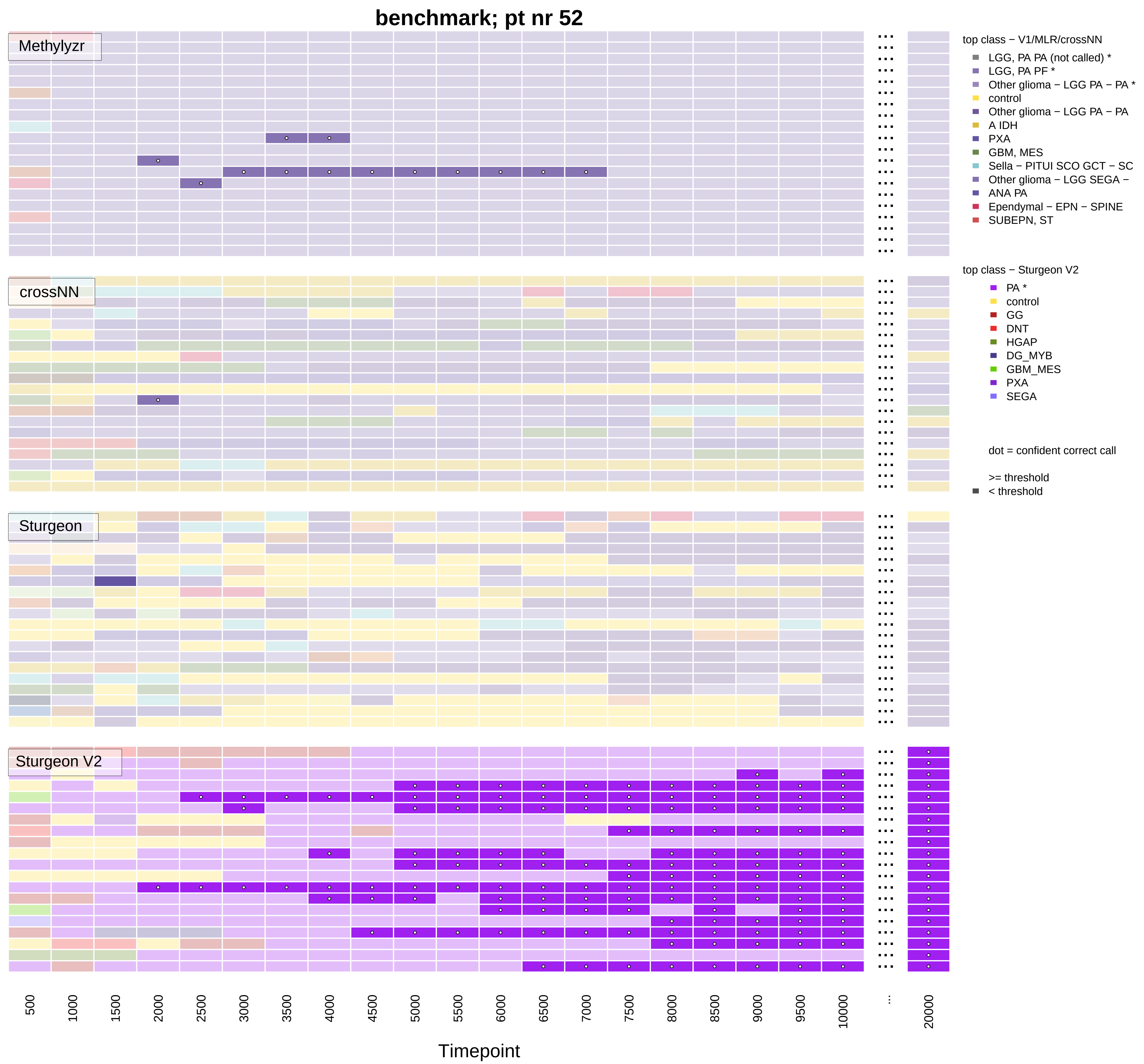

benchmark; pt nr 52
Methylyzr
crossNN
Sturgeon
Sturgeon V2
top class − V1/MLR/crossNN
LGG, PA PA (not called) *
LGG, PA PF *
Other glioma − LGG PA − PA *
control
Other glioma − LGG PA − PA
A IDH
PXA
GBM, MES
Sella − PITUI SCO GCT − SC
Other glioma − LGG SEGA −
ANA PA
Ependymal − EPN − SPINE
SUBEPN, ST
top class − Sturgeon V2
PA *
control
GG
DNT
HGAP
DG_MYB
GBM_MES
PXA
SEGA
dot = confident correct call
>= threshold
< threshold
500
1000
1500
2000
2500
3000
3500
4000
4500
5000
5500
6000
6500
7000
7500
8000
8500
9000
9500
10000
...
20000
Timepoint

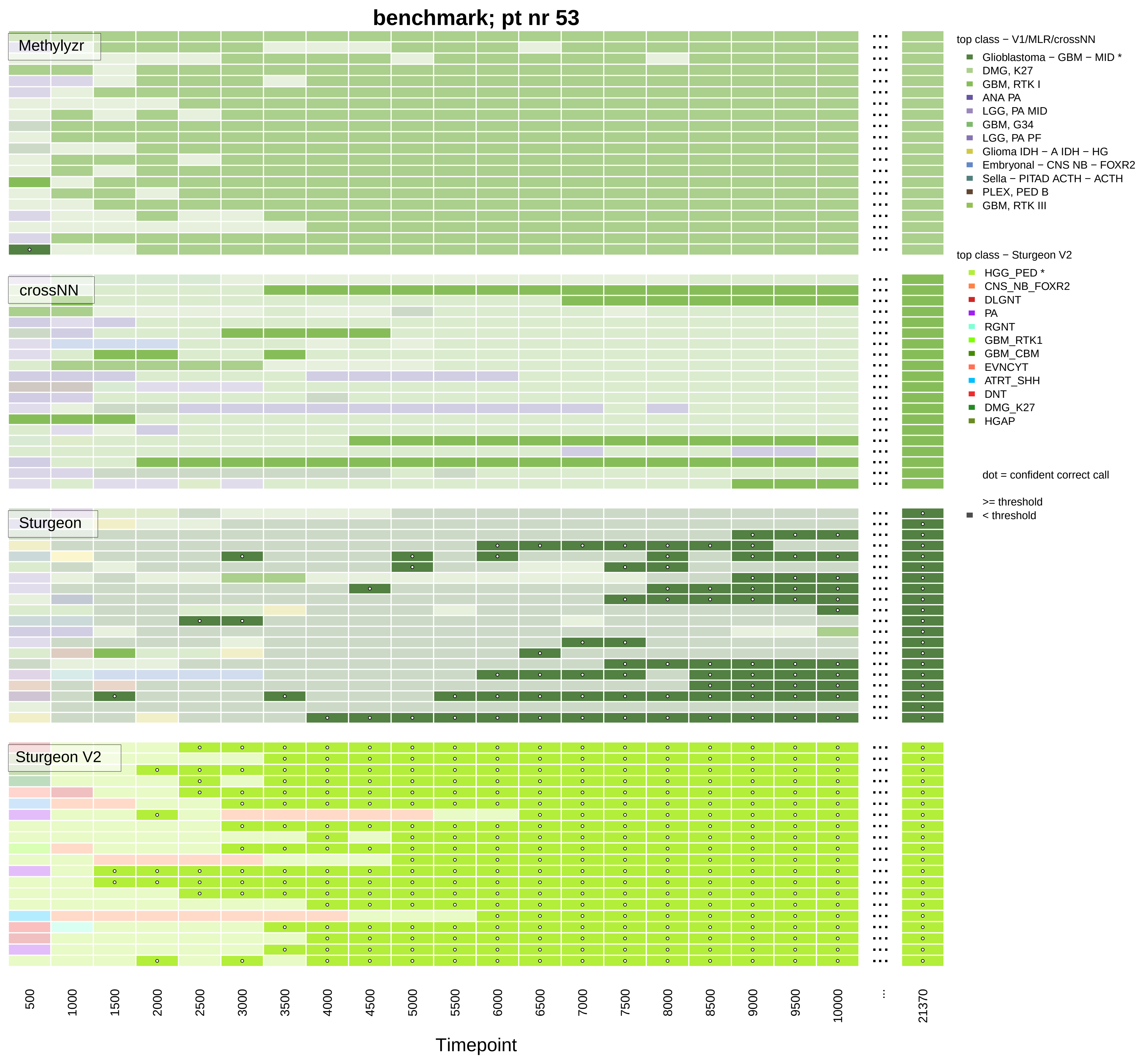

benchmark; pt nr 53
Methylyzr
crossNN
Sturgeon
Sturgeon V2
top class − V1/MLR/crossNN
Glioblastoma − GBM − MID *
DMG, K27
GBM, RTK I
ANA PA
LGG, PA MID
GBM, G34
LGG, PA PF
Glioma IDH − A IDH − HG
Embryonal − CNS NB − FOXR2
Sella − PITAD ACTH − ACTH
PLEX, PED B
GBM, RTK III
top class − Sturgeon V2
HGG_PED *
CNS_NB_FOXR2
DLGNT
PA
RGNT
GBM_RTK1
GBM_CBM
EVNCYT
ATRT_SHH
DNT
DMG_K27
HGAP
dot = confident correct call
>= threshold
< threshold
500
1000
1500
2000
2500
3000
3500
4000
4500
5000
5500
6000
6500
7000
7500
8000
8500
9000
9500
10000
...
21370
Timepoint

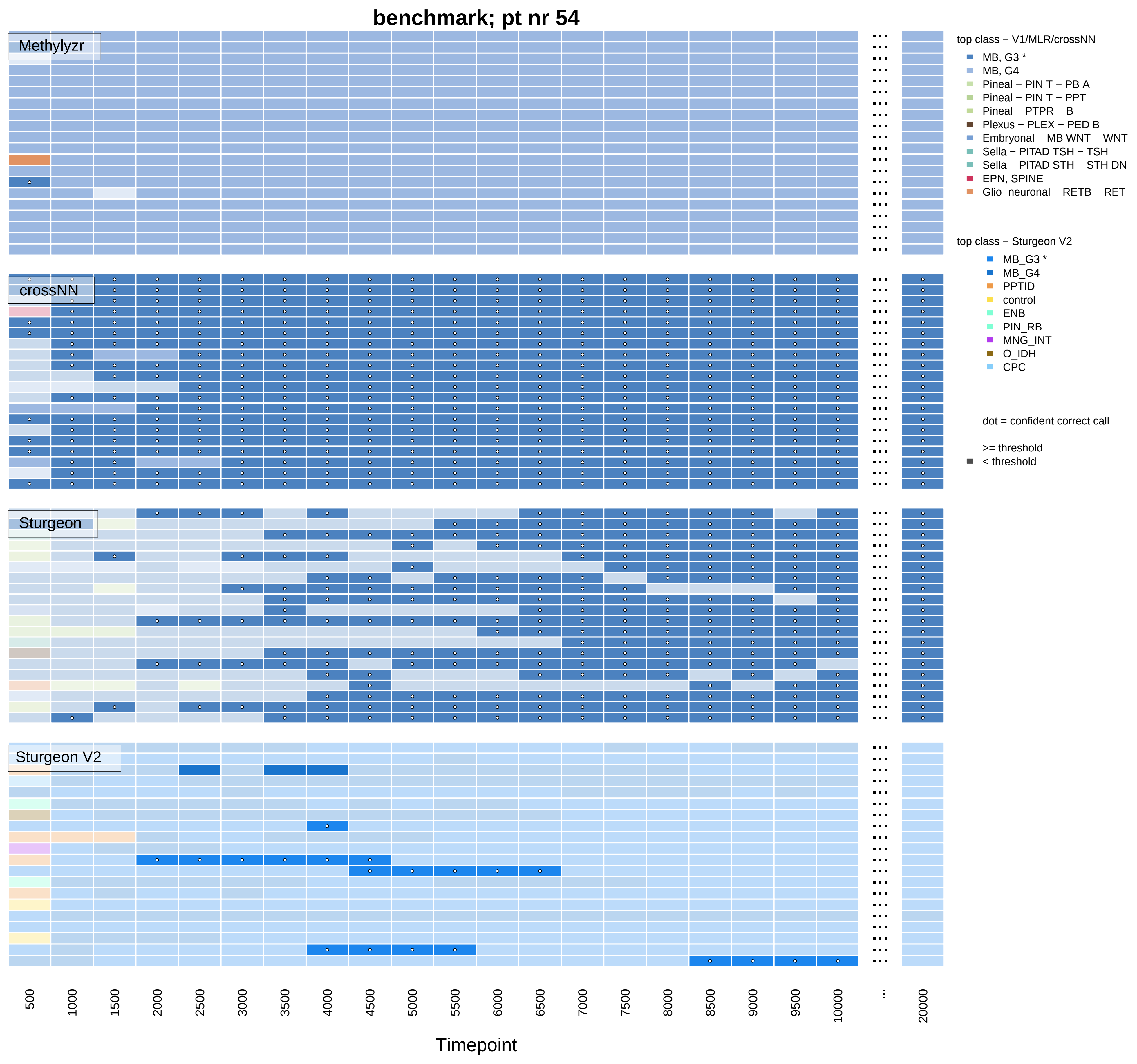
benchmark; pt nr 54
Methylyzr
crossNN
Sturgeon
Sturgeon V2
top class − V1/MLR/crossNN
MB, G3 *
MB, G4
Pineal − PIN T − PB A
Pineal − PIN T − PPT
Pineal − PTPR − B
Plexus − PLEX − PED B
Embryonal − MB WNT − WNT
Sella − PITAD TSH − TSH
Sella − PITAD STH − STH DN
EPN, SPINE
Glio−neuronal − RETB − RET
top class − Sturgeon V2
MB_G3 *
MB_G4
PPTID
control
ENB
PIN_RB
MNG_INT
O_IDH
CPC
dot = confident correct call
>= threshold
< threshold
500
1000
1500
2000
2500
3000
3500
4000
4500
5000
5500
6000
6500
7000
7500
8000
8500
9000
9500
10000
...
20000
Timepoint

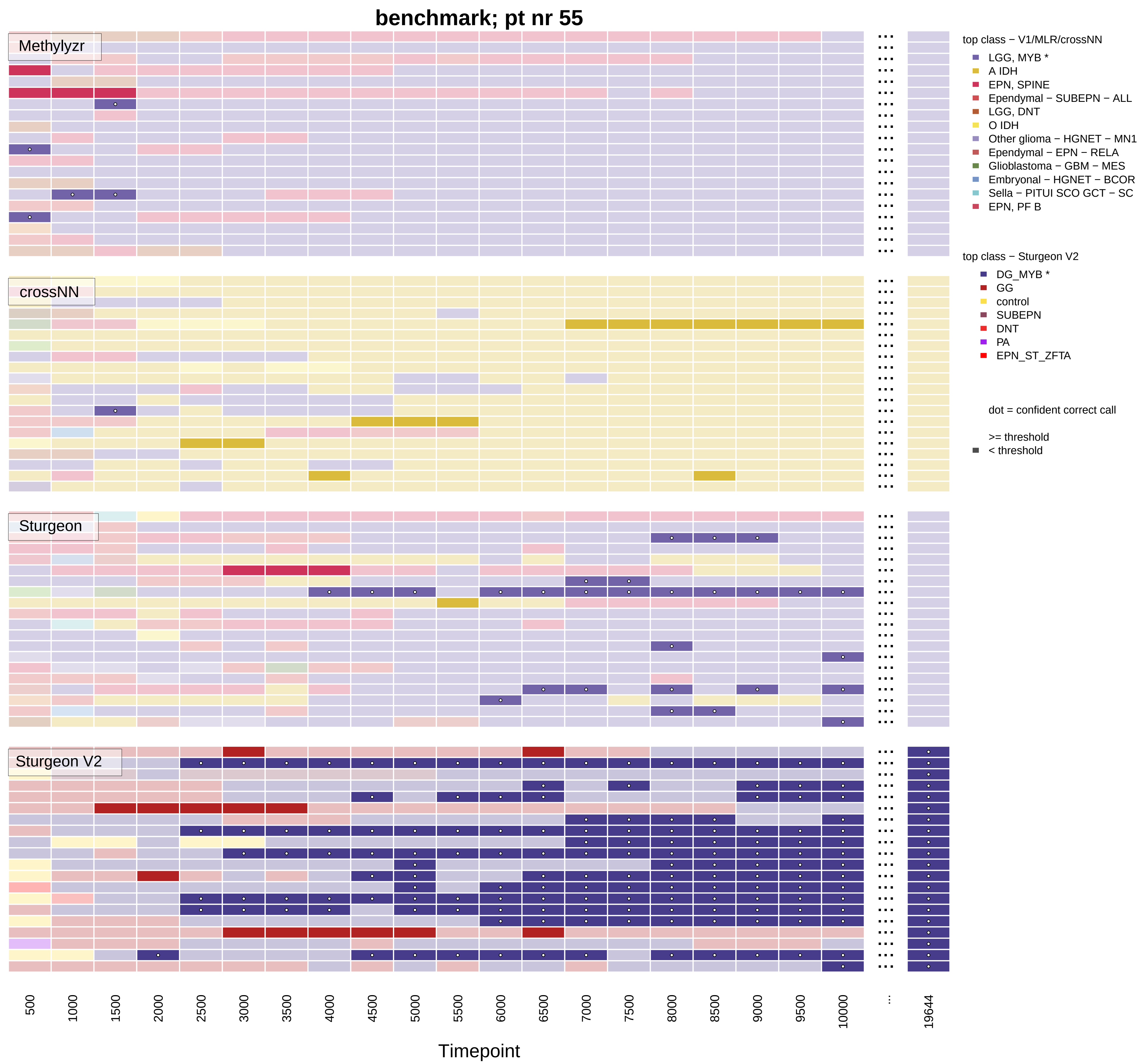

benchmark; pt nr 55
Methylyzr
crossNN
Sturgeon
Sturgeon V2
top class − V1/MLR/crossNN
LGG, MYB *
A IDH
EPN, SPINE
Ependymal − SUBEPN − ALL
LGG, DNT
O IDH
Other glioma − HGNET − MN1
Ependymal − EPN − RELA
Glioblastoma − GBM − MES
Embryonal − HGNET − BCOR
Sella − PITUI SCO GCT − SC
EPN, PF B
top class − Sturgeon V2
DG_MYB *
GG
control
SUBEPN
DNT
PA
EPN_ST_ZFTA
dot = confident correct call
>= threshold
< threshold
500
1000
1500
2000
2500
3000
3500
4000
4500
5000
5500
6000
6500
7000
7500
8000
8500
9000
9500
10000
...
19644
Timepoint

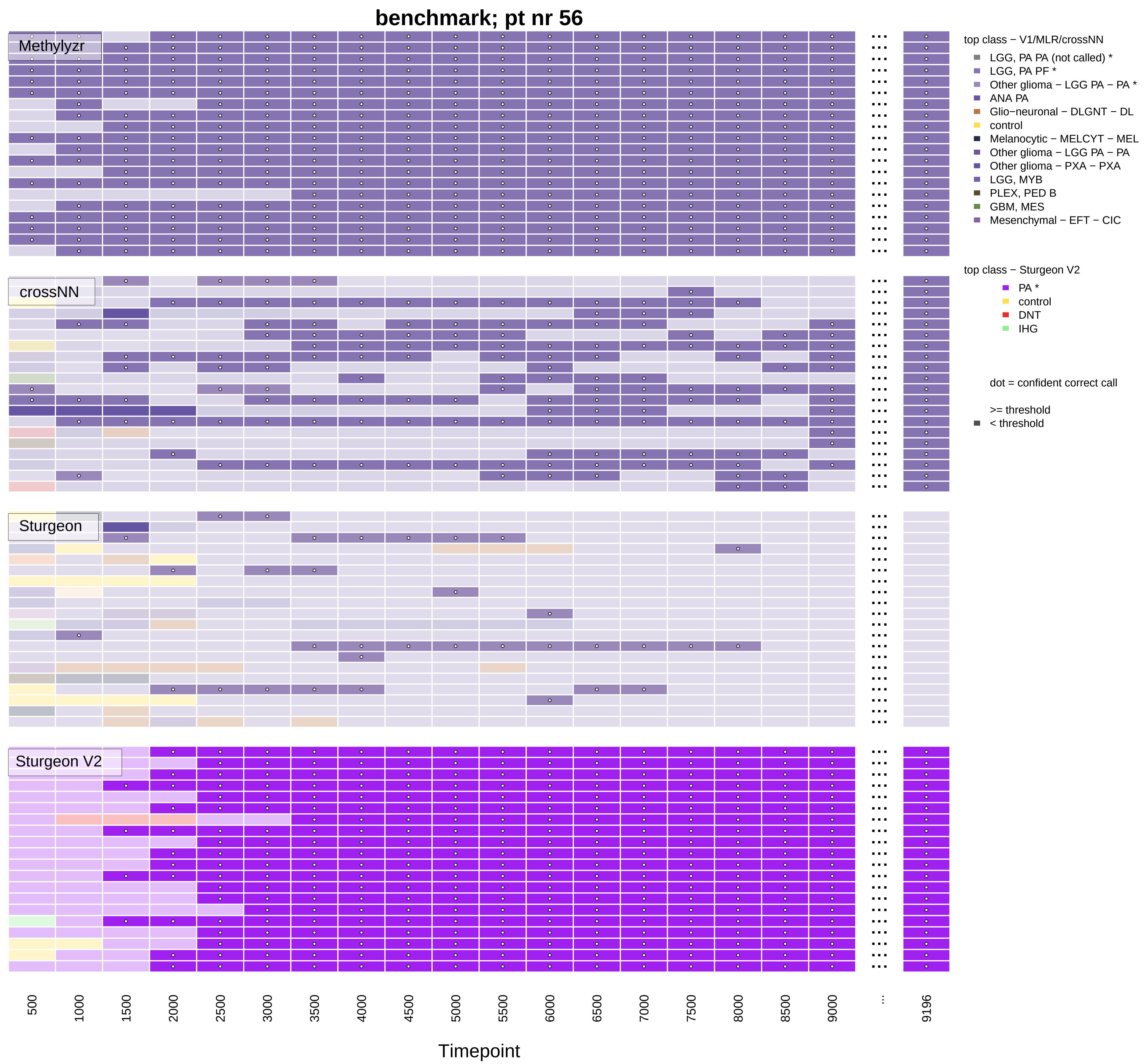
benchmark; pt nr 56
Methylyzr
crossNN
Sturgeon
Sturgeon V2
top class − V1/MLR/crossNN
LGG, PA PA (not called) *
LGG, PA PF *
Other glioma − LGG PA − PA *
ANA PA
Glio−neuronal − DLGNT − DL
control
Melanocytic − MELCYT − MEL
Other glioma − LGG PA − PA
Other glioma − PXA − PXA
LGG, MYB
PLEX, PED B
GBM, MES
Mesenchymal − EFT − CIC
top class − Sturgeon V2
PA *
control
DNT
IHG
dot = confident correct call
>= threshold
< threshold
500
1000
1500
2000
2500
3000
3500
4000
4500
5000
5500
6000
6500
7000
7500
8000
8500
9000
...
9196
Timepoint

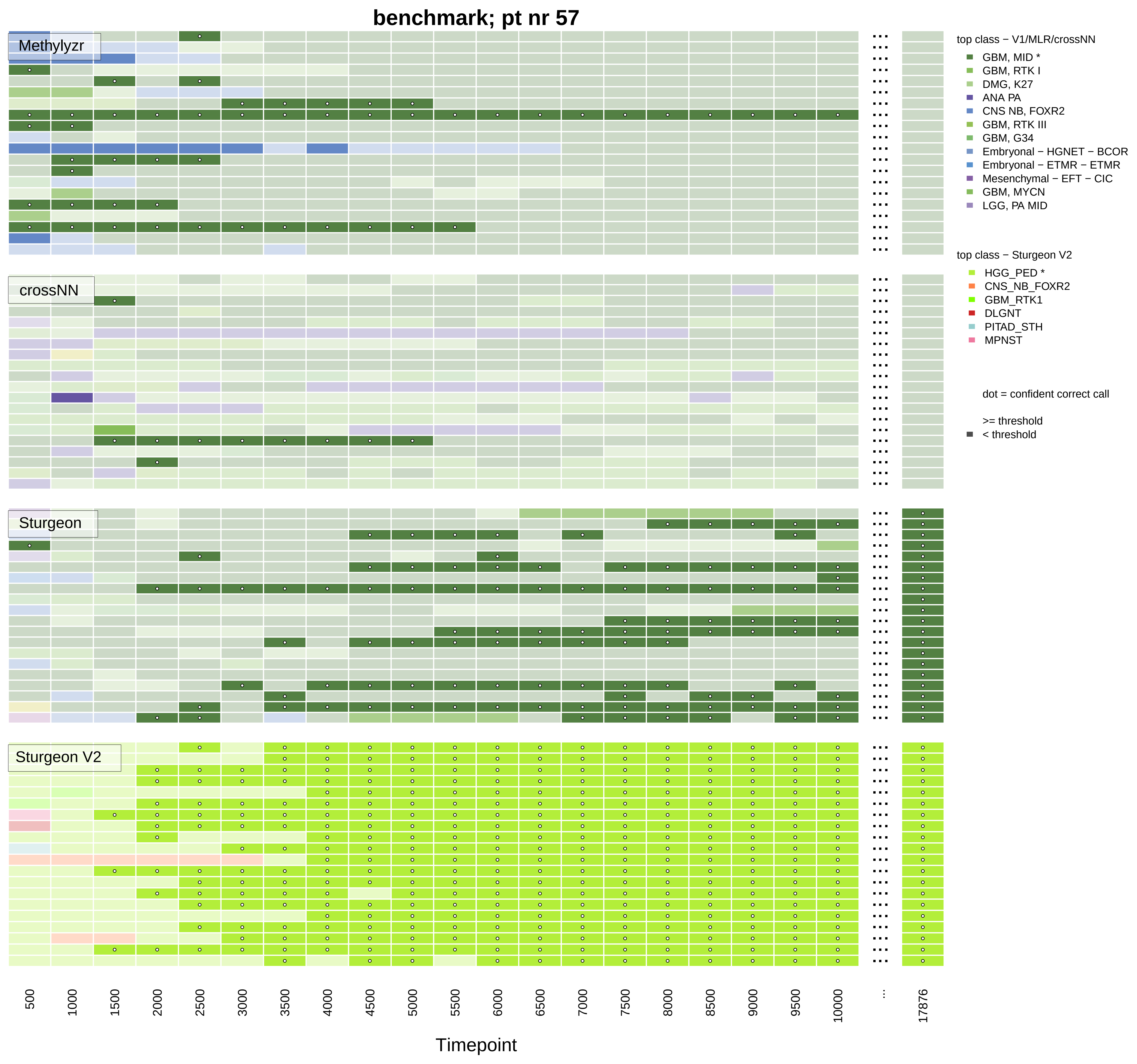

benchmark; pt nr 57
Methylyzr
crossNN
Sturgeon
Sturgeon V2
top class − V1/MLR/crossNN
GBM, MID *
GBM, RTK I
DMG, K27
ANA PA
CNS NB, FOXR2
GBM, RTK III
GBM, G34
Embryonal − HGNET − BCOR
Embryonal − ETMR − ETMR
Mesenchymal − EFT − CIC
GBM, MYCN
LGG, PA MID
top class − Sturgeon V2
HGG_PED *
CNS_NB_FOXR2
GBM_RTK1
DLGNT
PITAD_STH
MPNST
dot = confident correct call
>= threshold
< threshold
500
1000
1500
2000
2500
3000
3500
4000
4500
5000
5500
6000
6500
7000
7500
8000
8500
9000
9500
10000
...
17876
Timepoint

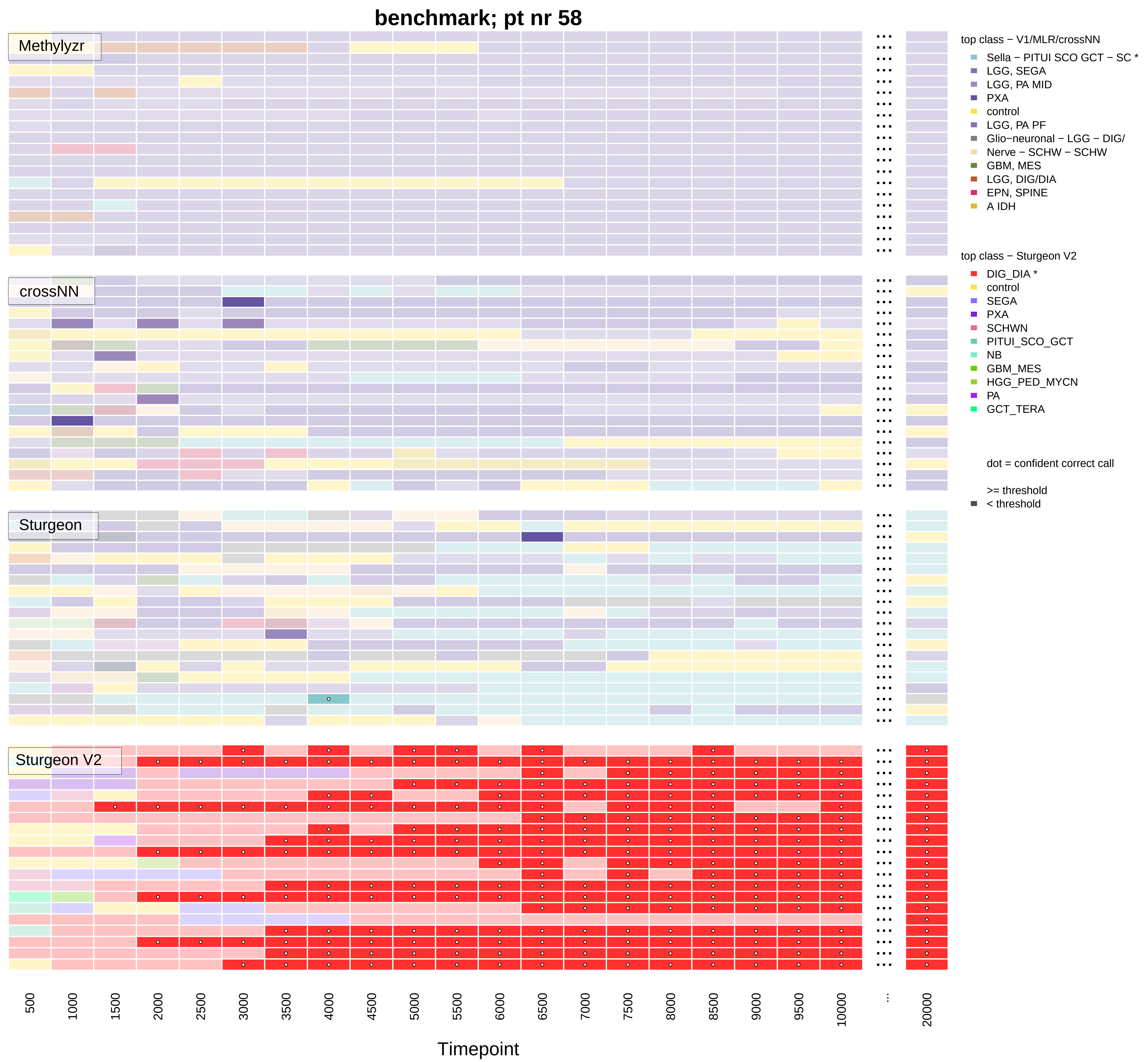

benchmark; pt nr 58
Methylyzr
crossNN
Sturgeon
Sturgeon V2
top class − V1/MLR/crossNN
Sella − PITUI SCO GCT − SC *
LGG, SEGA
LGG, PA MID
PXA
control
LGG, PA PF
Glio−neuronal − LGG − DIG/
Nerve − SCHW − SCHW
GBM, MES
LGG, DIG/DIA
EPN, SPINE
A IDH
top class − Sturgeon V2
DIG_DIA *
control
SEGA
PXA
SCHWN
PITUI_SCO_GCT
NB
GBM_MES
HGG_PED_MYCN
PA
GCT_TERA
dot = confident correct call
>= threshold
< threshold
500
1000
1500
2000
2500
3000
3500
4000
4500
5000
5500
6000
6500
7000
7500
8000
8500
9000
9500
10000
...
20000
Timepoint

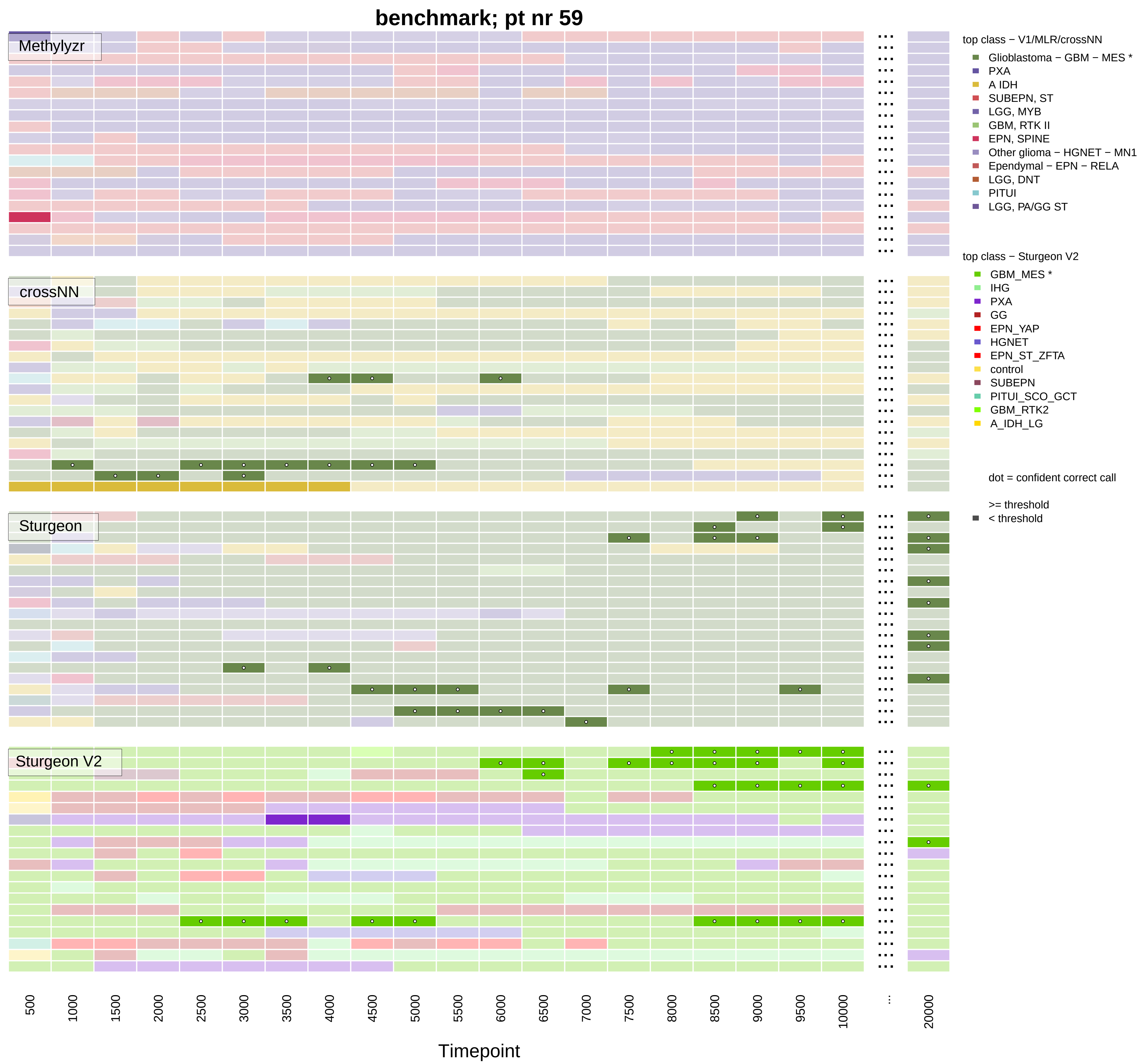
benchmark; pt nr 59
Methylyzr
crossNN
Sturgeon
Sturgeon V2
top class − V1/MLR/crossNN
Glioblastoma − GBM − MES *
PXA
A IDH
SUBEPN, ST
LGG, MYB
GBM, RTK II
EPN, SPINE
Other glioma − HGNET − MN1
Ependymal − EPN − RELA
LGG, DNT
PITUI
LGG, PA/GG ST
top class − Sturgeon V2
GBM_MES *
IHG
PXA
GG
EPN_YAP
HGNET
EPN_ST_ZFTA
control
SUBEPN
PITUI_SCO_GCT
GBM_RTK2
A_IDH_LG
dot = confident correct call
>= threshold
< threshold
500
1000
1500
2000
2500
3000
3500
4000
4500
5000
5500
6000
6500
7000
7500
8000
8500
9000
9500
10000
...
20000
Timepoint

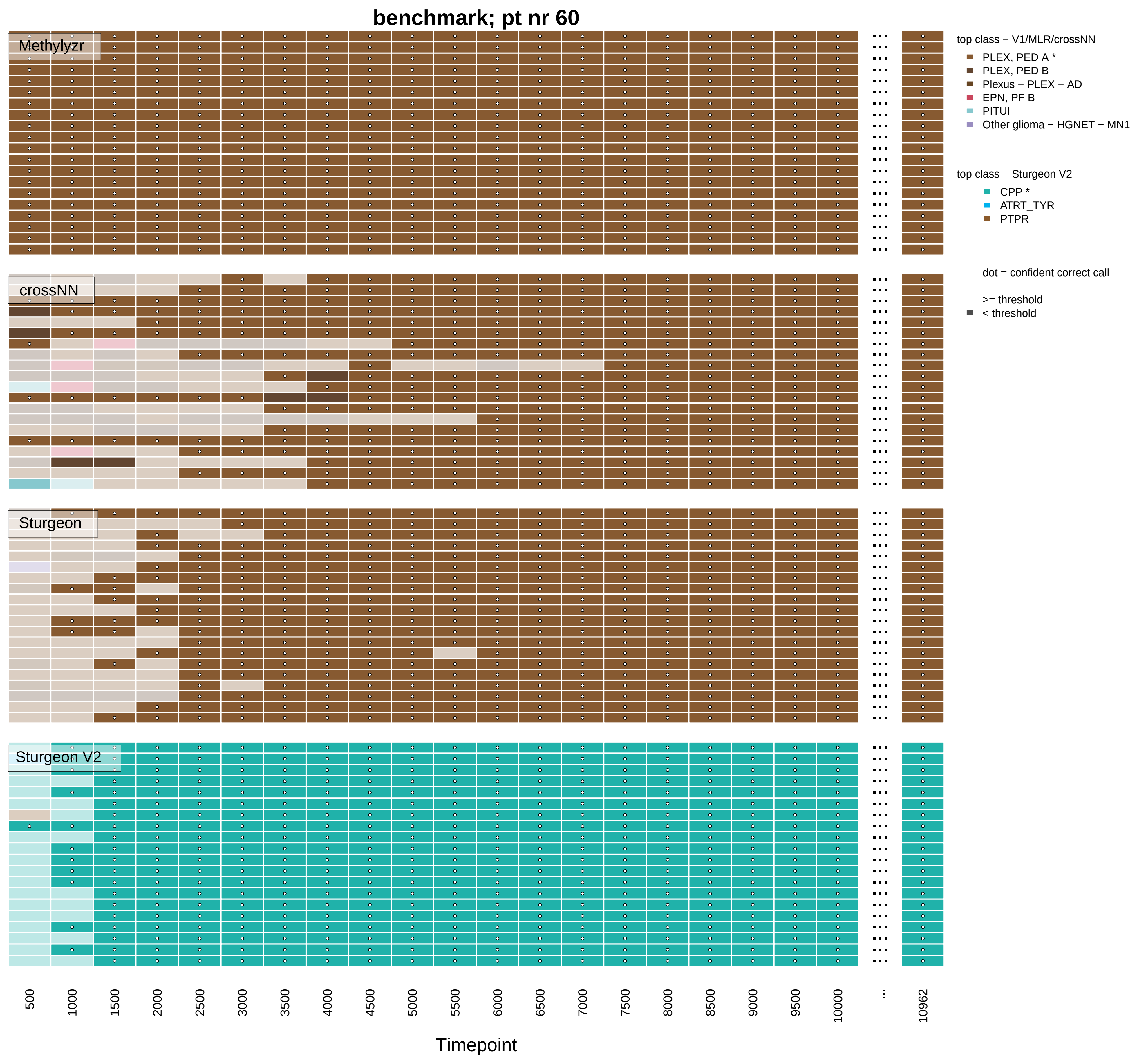
benchmark; pt nr 60
Methylyzr
crossNN
Sturgeon
Sturgeon V2
top class − V1/MLR/crossNN
PLEX, PED A *
PLEX, PED B
Plexus − PLEX − AD
EPN, PF B
PITUI
Other glioma − HGNET − MN1
top class − Sturgeon V2
CPP *
ATRT_TYR
PTPR
dot = confident correct call
>= threshold
< threshold
500
1000
1500
2000
2500
3000
3500
4000
4500
5000
5500
6000
6500
7000
7500
8000
8500
9000
9500
10000
...
10962
Timepoint

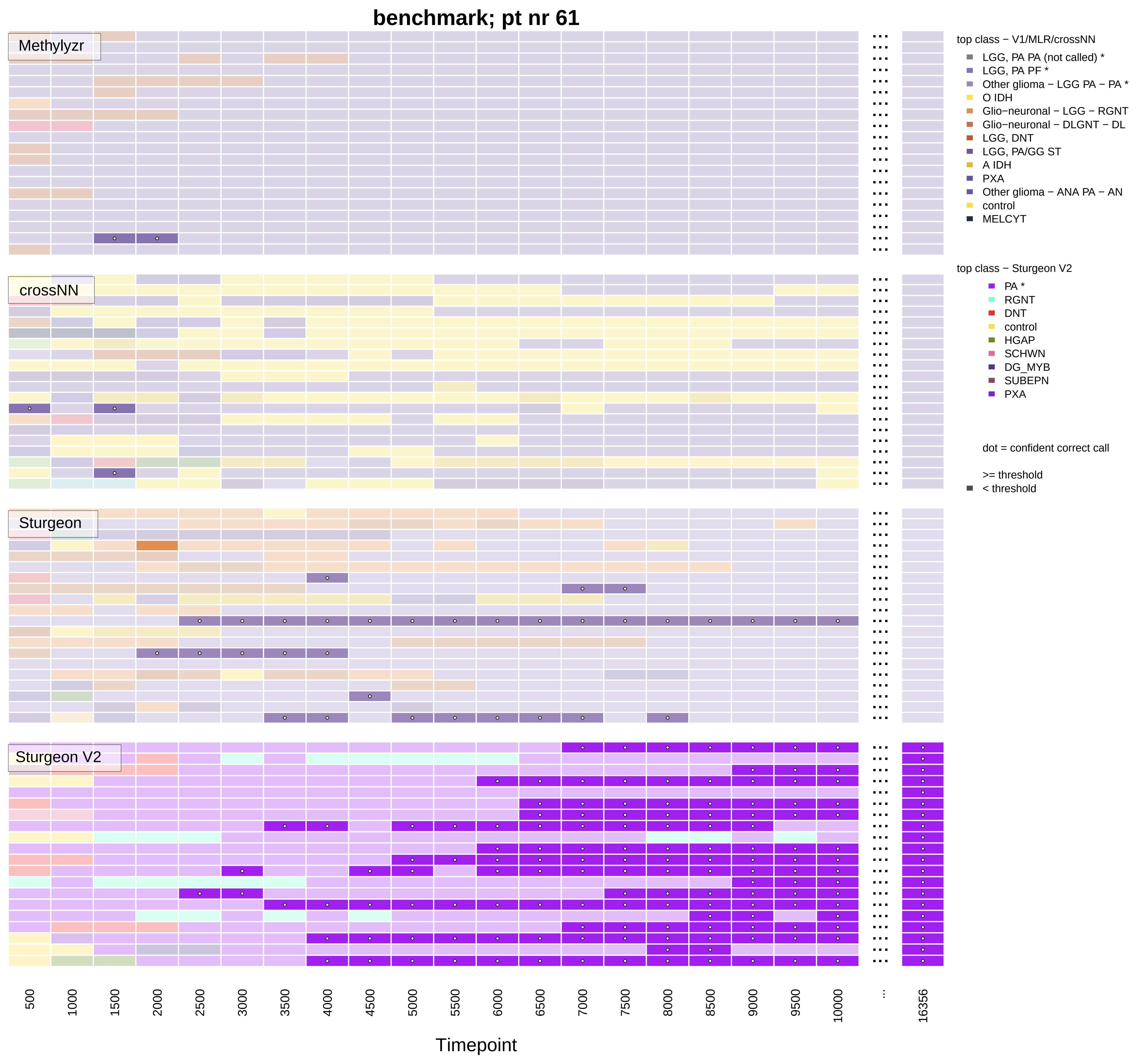
benchmark; pt nr 61
Methylyzr
crossNN
Sturgeon
Sturgeon V2
top class − V1/MLR/crossNN
LGG, PA PA (not called) *
LGG, PA PF *
Other glioma − LGG PA − PA *
O IDH
Glio−neuronal − LGG − RGNT
Glio−neuronal − DLGNT − DL
LGG, DNT
LGG, PA/GG ST
A IDH
PXA
Other glioma − ANA PA − AN
control
MELCYT
top class − Sturgeon V2
PA *
RGNT
DNT
control
HGAP
SCHWN
DG_MYB
SUBEPN
PXA
dot = confident correct call
>= threshold
< threshold
500
1000
1500
2000
2500
3000
3500
4000
4500
5000
5500
6000
6500
7000
7500
8000
8500
9000
9500
10000
...
16356
Timepoint

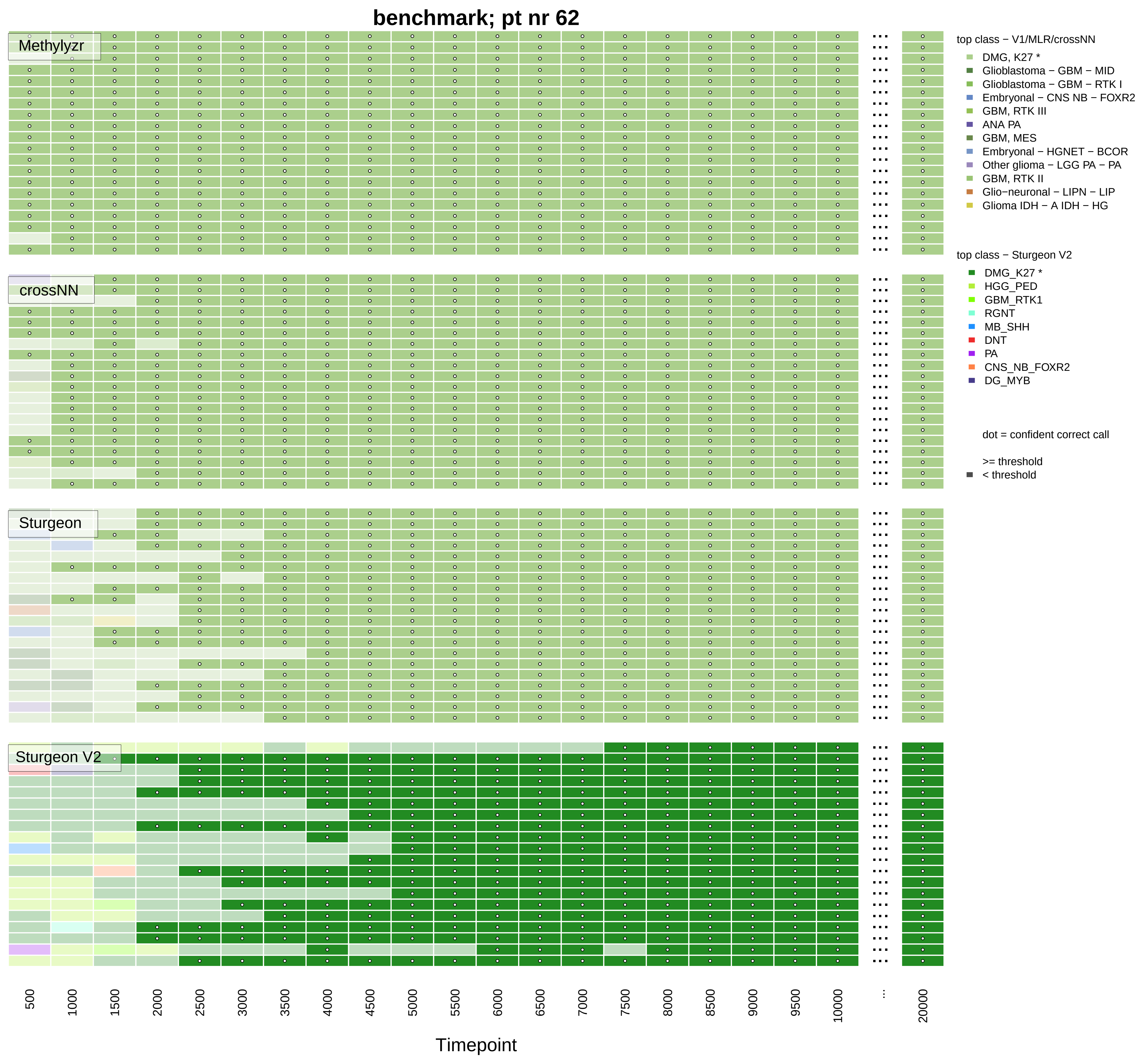
benchmark; pt nr 62
Methylyzr
crossNN
Sturgeon
Sturgeon V2
top class − V1/MLR/crossNN
DMG, K27 *
Glioblastoma − GBM − MID
Glioblastoma − GBM − RTK I
Embryonal − CNS NB − FOXR2
GBM, RTK III
ANA PA
GBM, MES
Embryonal − HGNET − BCOR
Other glioma − LGG PA − PA
GBM, RTK II
Glio−neuronal − LIPN − LIP
Glioma IDH − A IDH − HG
top class − Sturgeon V2
DMG_K27 *
HGG_PED
GBM_RTK1
RGNT
MB_SHH
DNT
PA
CNS_NB_FOXR2
DG_MYB
dot = confident correct call
>= threshold
< threshold
500
1000
1500
2000
2500
3000
3500
4000
4500
5000
5500
6000
6500
7000
7500
8000
8500
9000
9500
10000
...
20000
Timepoint

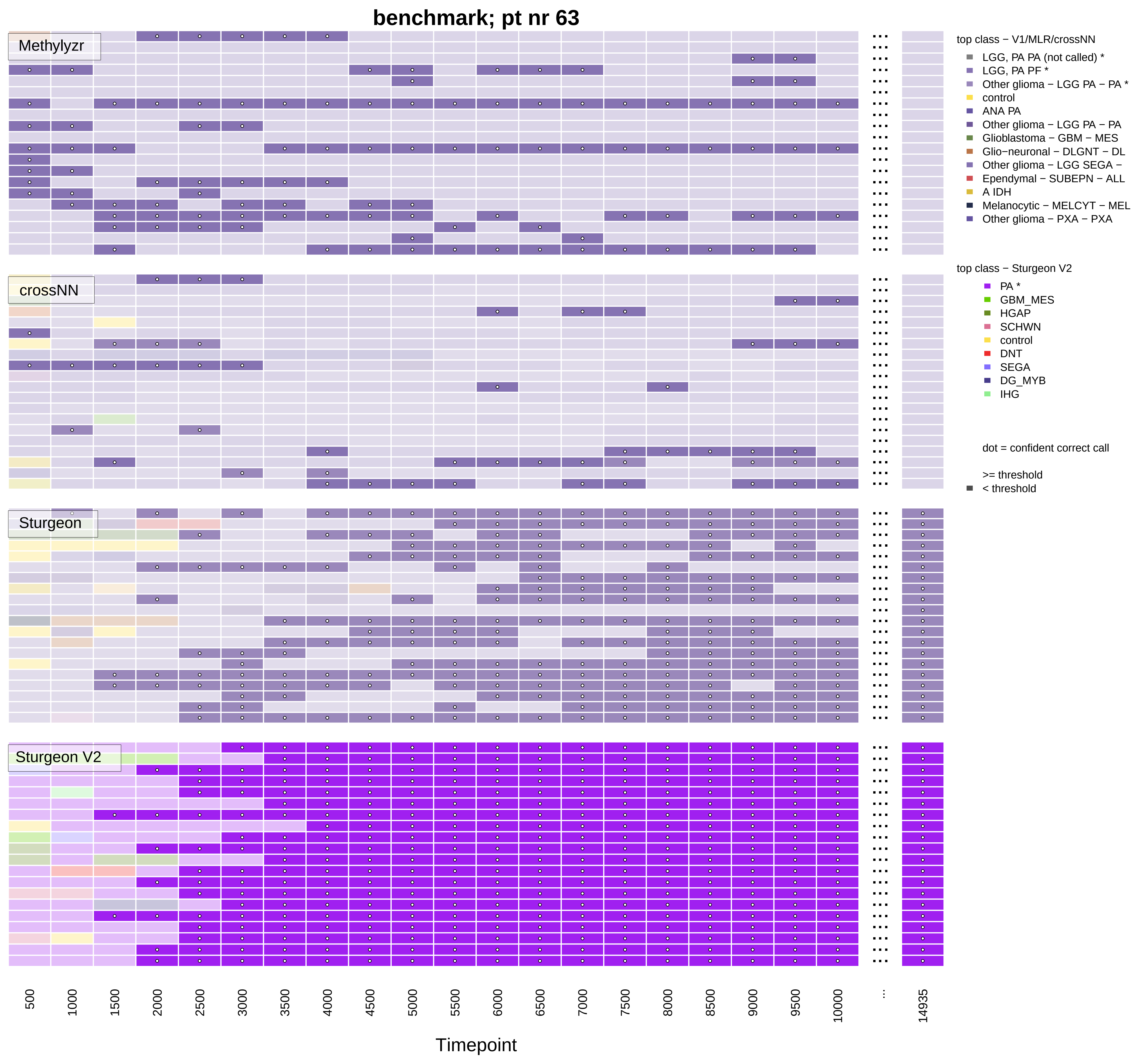
benchmark; pt nr 63
Methylyzr
crossNN
Sturgeon
Sturgeon V2
top class − V1/MLR/crossNN
LGG, PA PA (not called) *
LGG, PA PF *
Other glioma − LGG PA − PA *
control
ANA PA
Other glioma − LGG PA − PA
Glioblastoma − GBM − MES
Glio−neuronal − DLGNT − DL
Other glioma − LGG SEGA −
Ependymal − SUBEPN − ALL
A IDH
Melanocytic − MELCYT − MEL
Other glioma − PXA − PXA
top class − Sturgeon V2
PA *
GBM_MES
HGAP
SCHWN
control
DNT
SEGA
DG_MYB
IHG
dot = confident correct call
>= threshold
< threshold
500
1000
1500
2000
2500
3000
3500
4000
4500
5000
5500
6000
6500
7000
7500
8000
8500
9000
9500
10000
...
14935
Timepoint

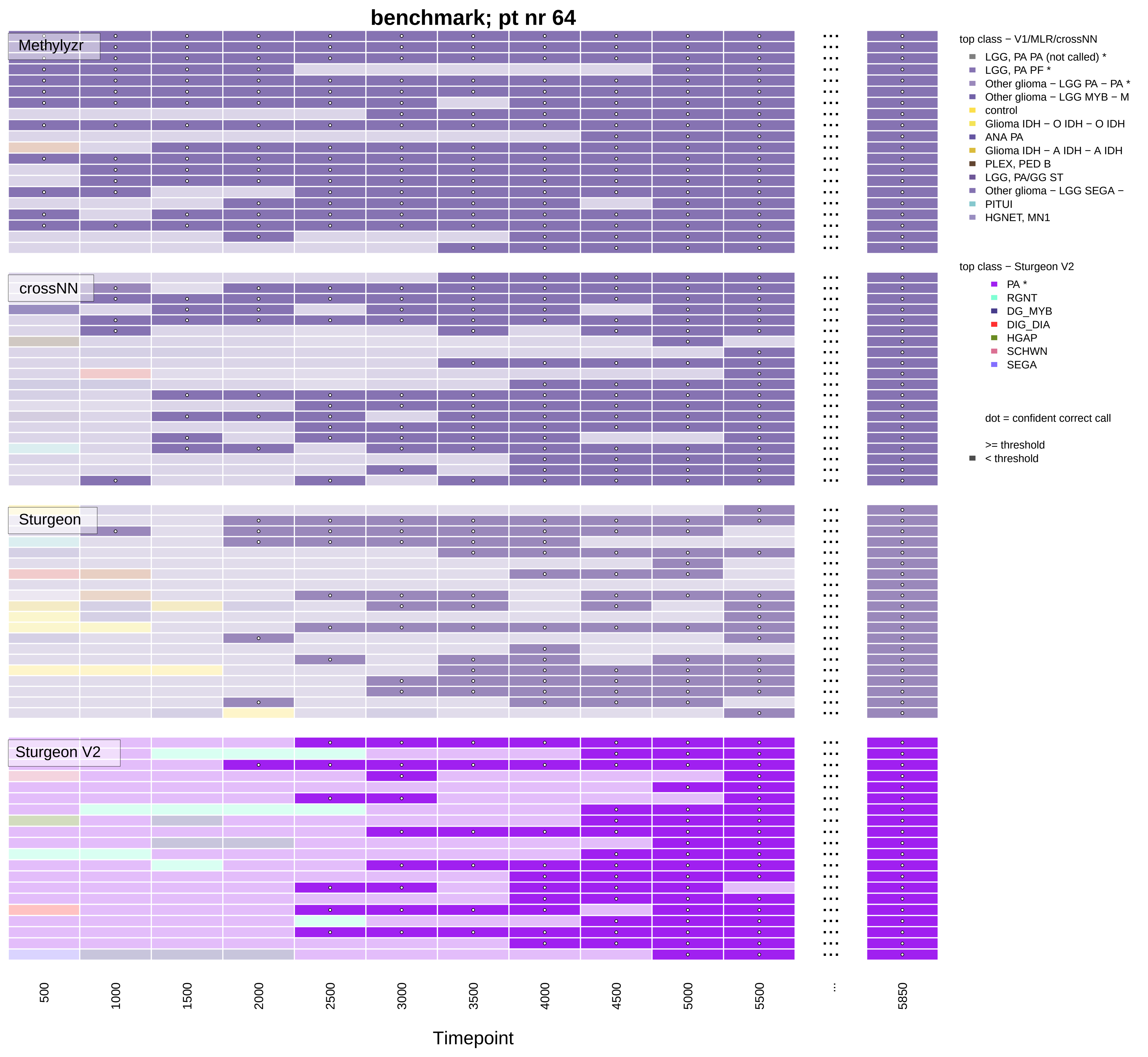

benchmark; pt nr 64
Methylyzr
crossNN
Sturgeon
Sturgeon V2
top class − V1/MLR/crossNN
LGG, PA PA (not called) *
LGG, PA PF *
Other glioma − LGG PA − PA *
Other glioma − LGG MYB − M
control
Glioma IDH − O IDH − O IDH
ANA PA
Glioma IDH − A IDH − A IDH
PLEX, PED B
LGG, PA/GG ST
Other glioma − LGG SEGA −
PITUI
HGNET, MN1
top class − Sturgeon V2
PA *
RGNT
DG_MYB
DIG_DIA
HGAP
SCHWN
SEGA
dot = confident correct call
>= threshold
< threshold
500
1000
1500
2000
2500
3000
3500
4000
4500
5000
5500
...
5850
Timepoint

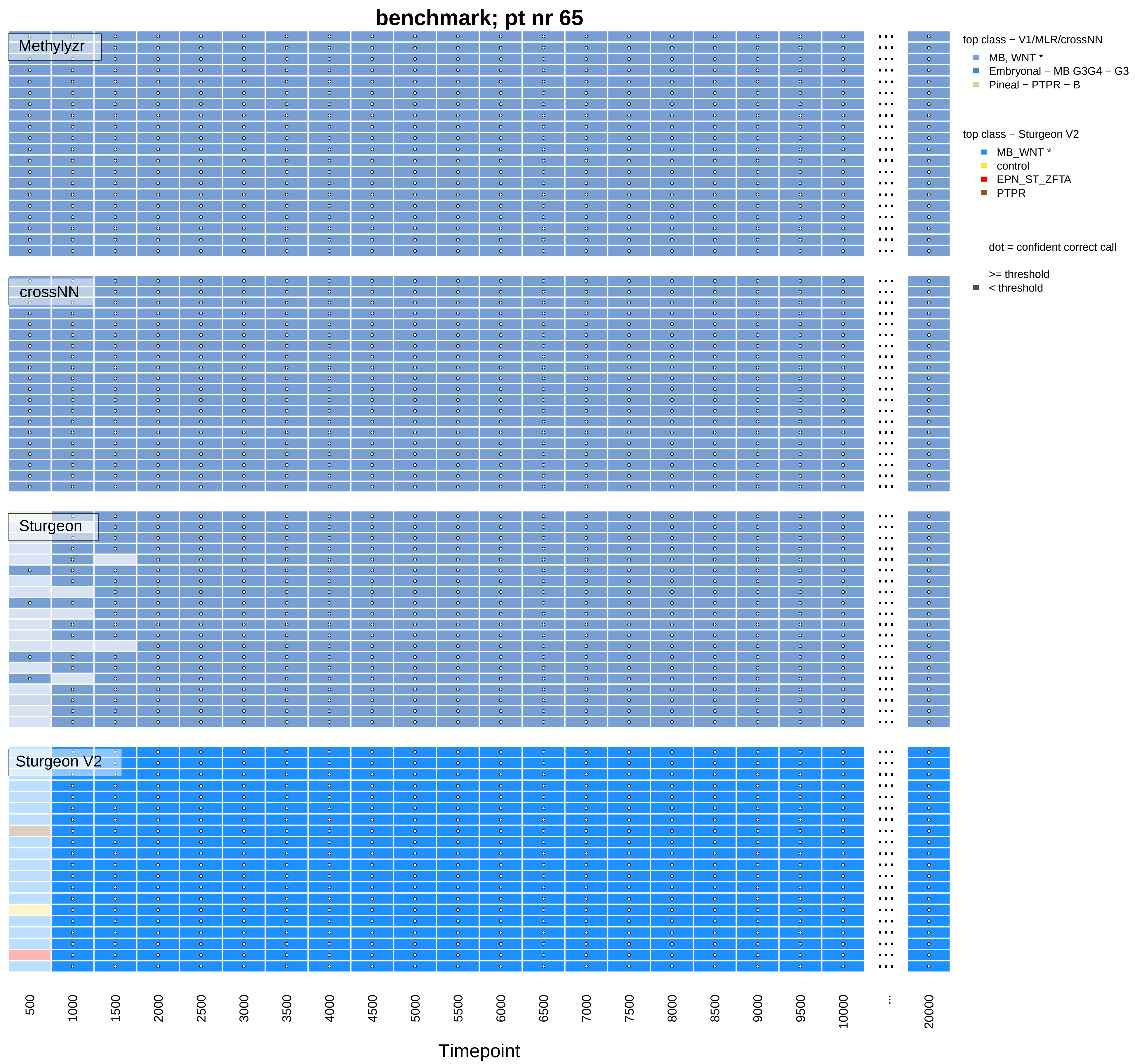
benchmark; pt nr 65
Methylyzr
crossNN
Sturgeon
Sturgeon V2
top class − V1/MLR/crossNN
MB, WNT *
Embryonal − MB G3G4 − G3
Pineal − PTPR − B
top class − Sturgeon V2
MB_WNT *
control
EPN_ST_ZFTA
PTPR
dot = confident correct call
>= threshold
< threshold
500
1000
1500
2000
2500
3000
3500
4000
4500
5000
5500
6000
6500
7000
7500
8000
8500
9000
9500
10000
...
20000
Timepoint

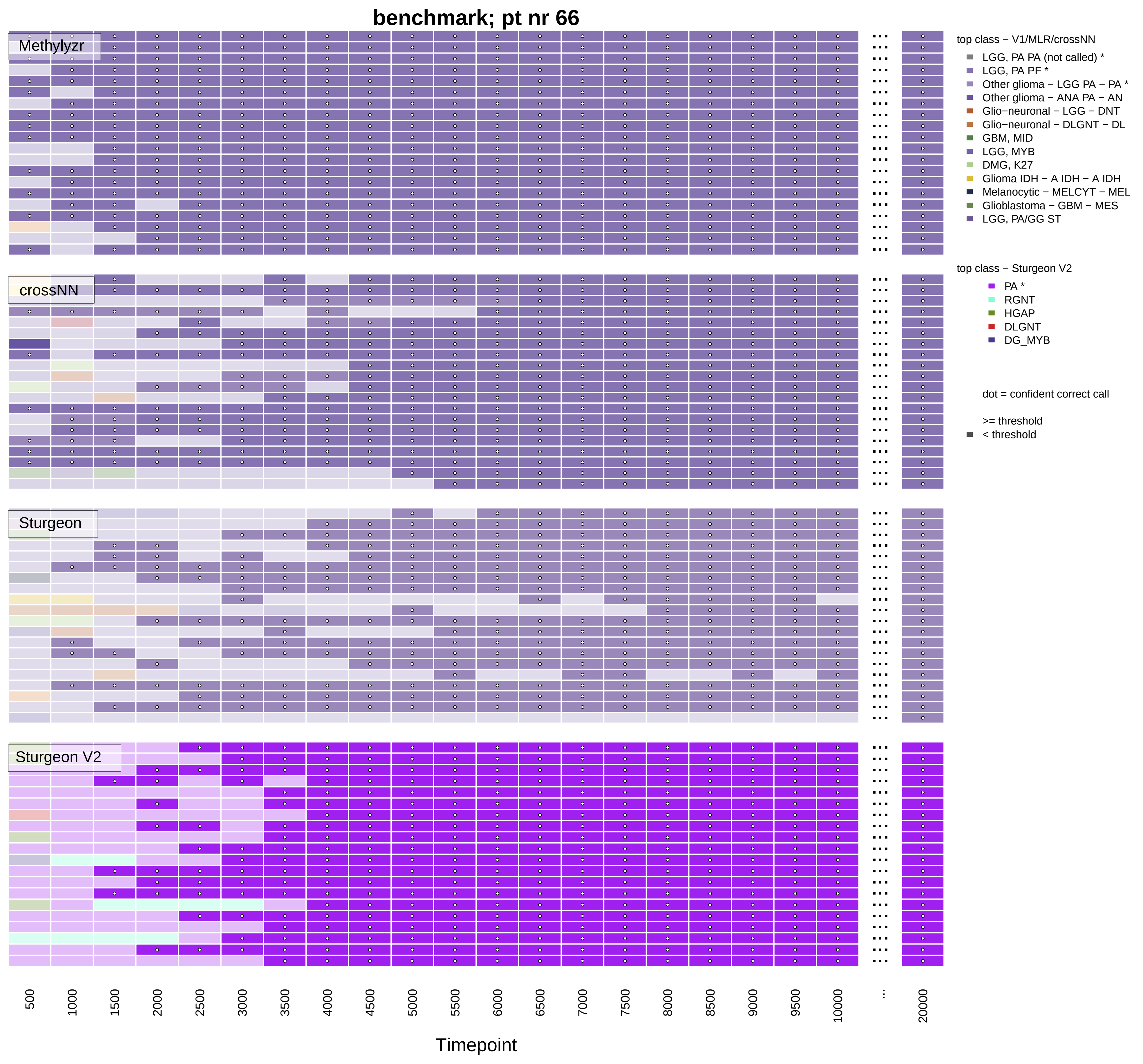

benchmark; pt nr 66
Methylyzr
crossNN
Sturgeon
Sturgeon V2
top class − V1/MLR/crossNN
LGG, PA PA (not called) *
LGG, PA PF *
Other glioma − LGG PA − PA *
Other glioma − ANA PA − AN
Glio−neuronal − LGG − DNT
Glio−neuronal − DLGNT − DL
GBM, MID
LGG, MYB
DMG, K27
Glioma IDH − A IDH − A IDH
Melanocytic − MELCYT − MEL
Glioblastoma − GBM − MES
LGG, PA/GG ST
top class − Sturgeon V2
PA *
RGNT
HGAP
DLGNT
DG_MYB
dot = confident correct call
>= threshold
< threshold
500
1000
1500
2000
2500
3000
3500
4000
4500
5000
5500
6000
6500
7000
7500
8000
8500
9000
9500
10000
...
20000
Timepoint

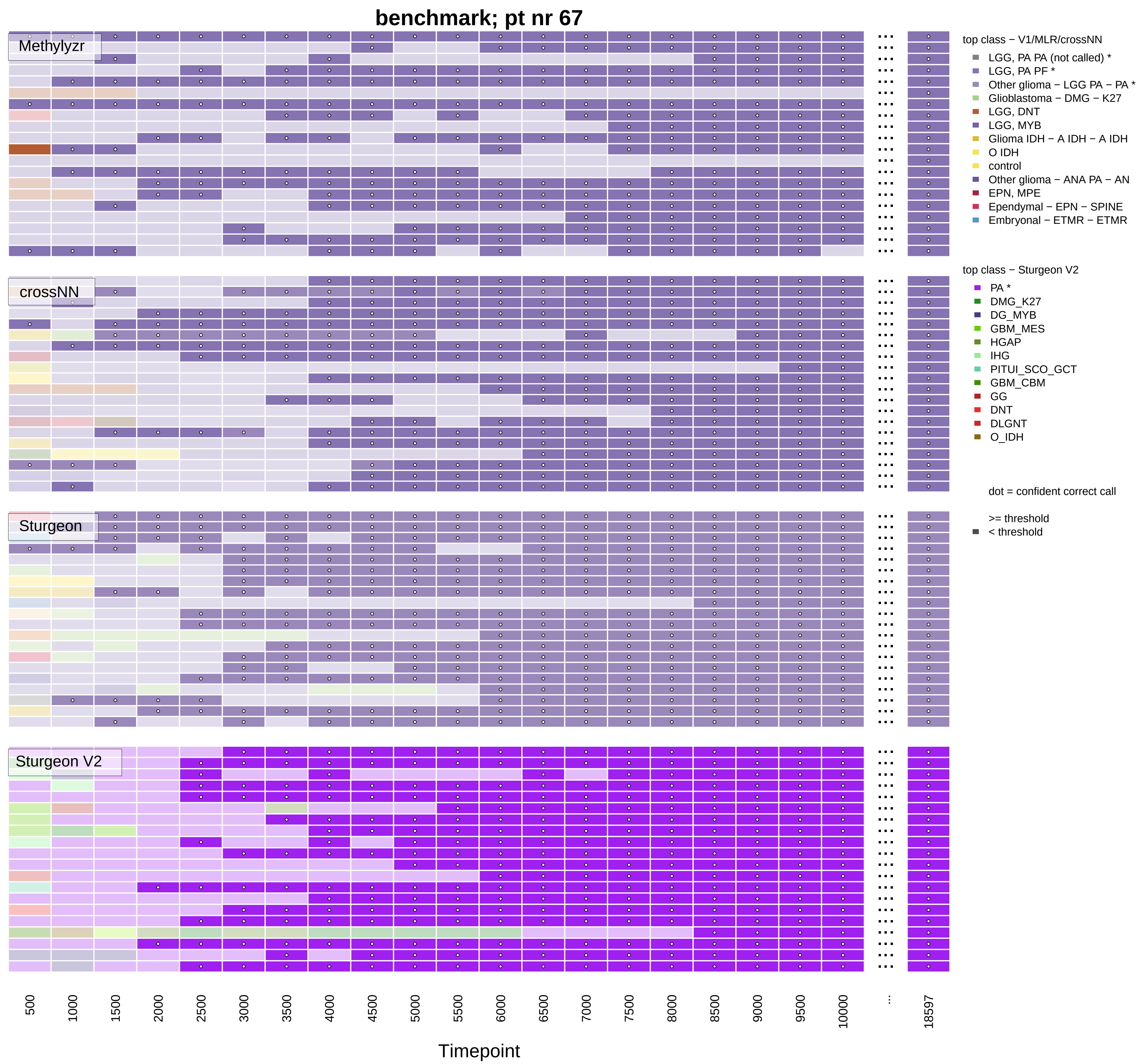
benchmark; pt nr 67
Methylyzr
crossNN
Sturgeon
Sturgeon V2
top class − V1/MLR/crossNN
LGG, PA PA (not called) *
LGG, PA PF *
Other glioma − LGG PA − PA *
Glioblastoma − DMG − K27
LGG, DNT
LGG, MYB
Glioma IDH − A IDH − A IDH
O IDH
control
Other glioma − ANA PA − AN
EPN, MPE
Ependymal − EPN − SPINE
Embryonal − ETMR − ETMR
top class − Sturgeon V2
PA *
DMG_K27
DG_MYB
GBM_MES
HGAP
IHG
PITUI_SCO_GCT
GBM_CBM
GG
DNT
DLGNT
O_IDH
dot = confident correct call
>= threshold
< threshold
500
1000
1500
2000
2500
3000
3500
4000
4500
5000
5500
6000
6500
7000
7500
8000
8500
9000
9500
10000
...
18597
Timepoint

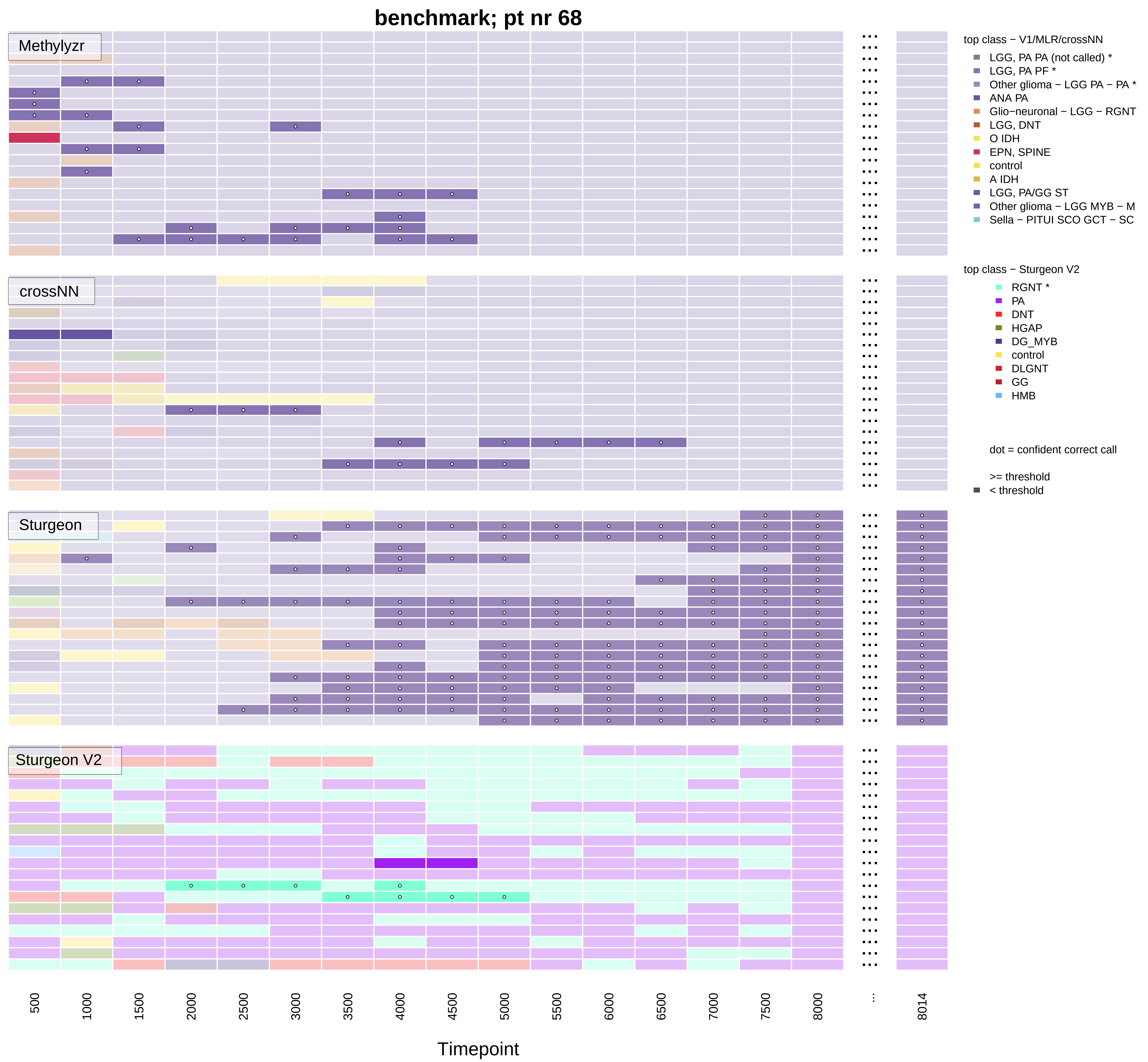
benchmark; pt nr 68
Methylyzr
crossNN
Sturgeon
Sturgeon V2
top class − V1/MLR/crossNN
LGG, PA PA (not called) *
LGG, PA PF *
Other glioma − LGG PA − PA *
ANA PA
Glio−neuronal − LGG − RGNT
LGG, DNT
O IDH
EPN, SPINE
control
A IDH
LGG, PA/GG ST
Other glioma − LGG MYB − M
Sella − PITUI SCO GCT − SC
top class − Sturgeon V2
RGNT *
PA
DNT
HGAP
DG_MYB
control
DLGNT
GG
HMB
dot = confident correct call
>= threshold
< threshold
500
1000
1500
2000
2500
3000
3500
4000
4500
5000
5500
6000
6500
7000
7500
8000
...
8014
Timepoint

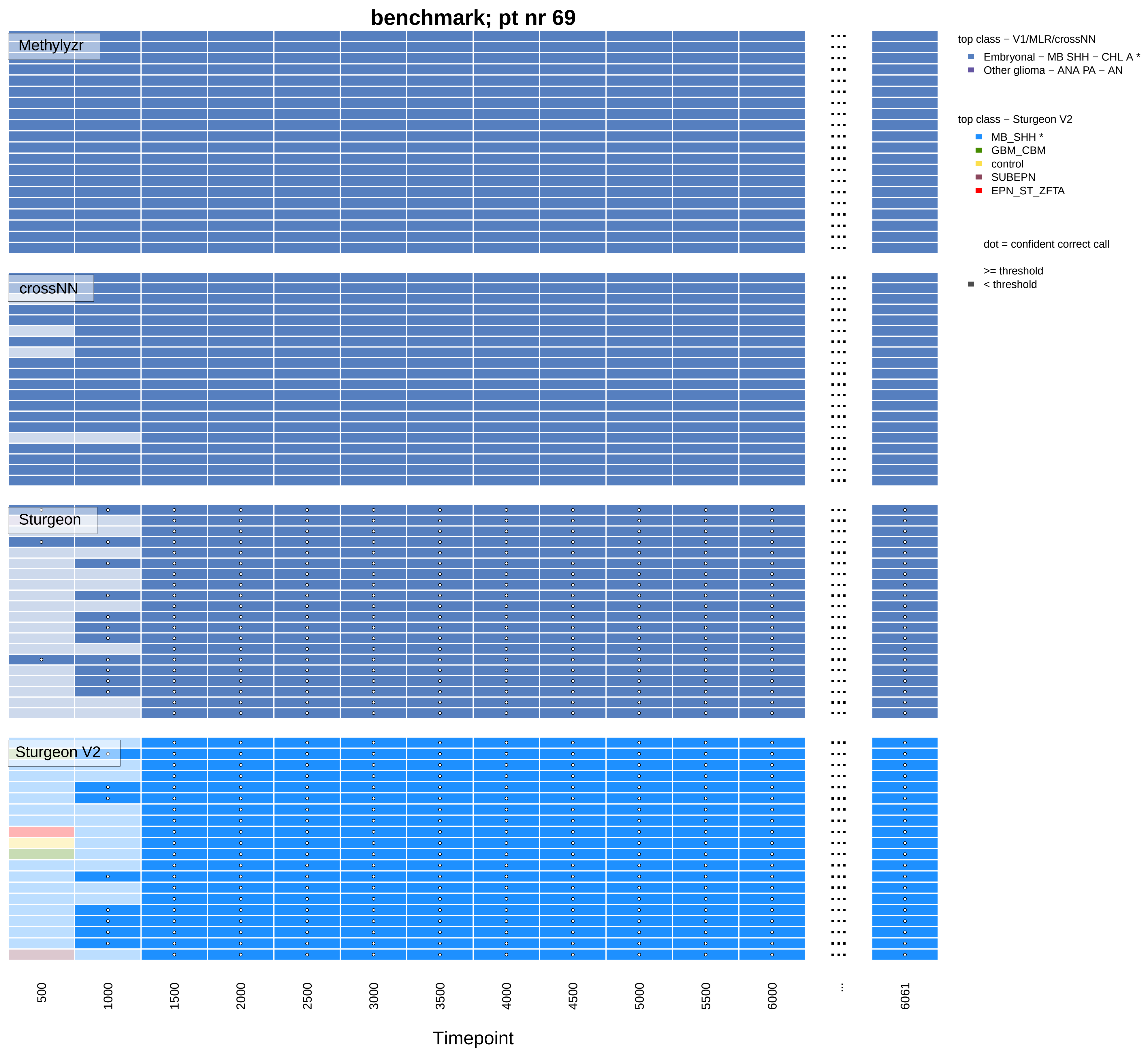
benchmark; pt nr 69
Methylyzr
crossNN
Sturgeon
Sturgeon V2
top class − V1/MLR/crossNN
Embryonal − MB SHH − CHL A *
Other glioma − ANA PA − AN
top class − Sturgeon V2
MB_SHH *
GBM_CBM
control
SUBEPN
EPN_ST_ZFTA
dot = confident correct call
>= threshold
< threshold
500
1000
1500
2000
2500
3000
3500
4000
4500
5000
5500
6000
...
6061
Timepoint

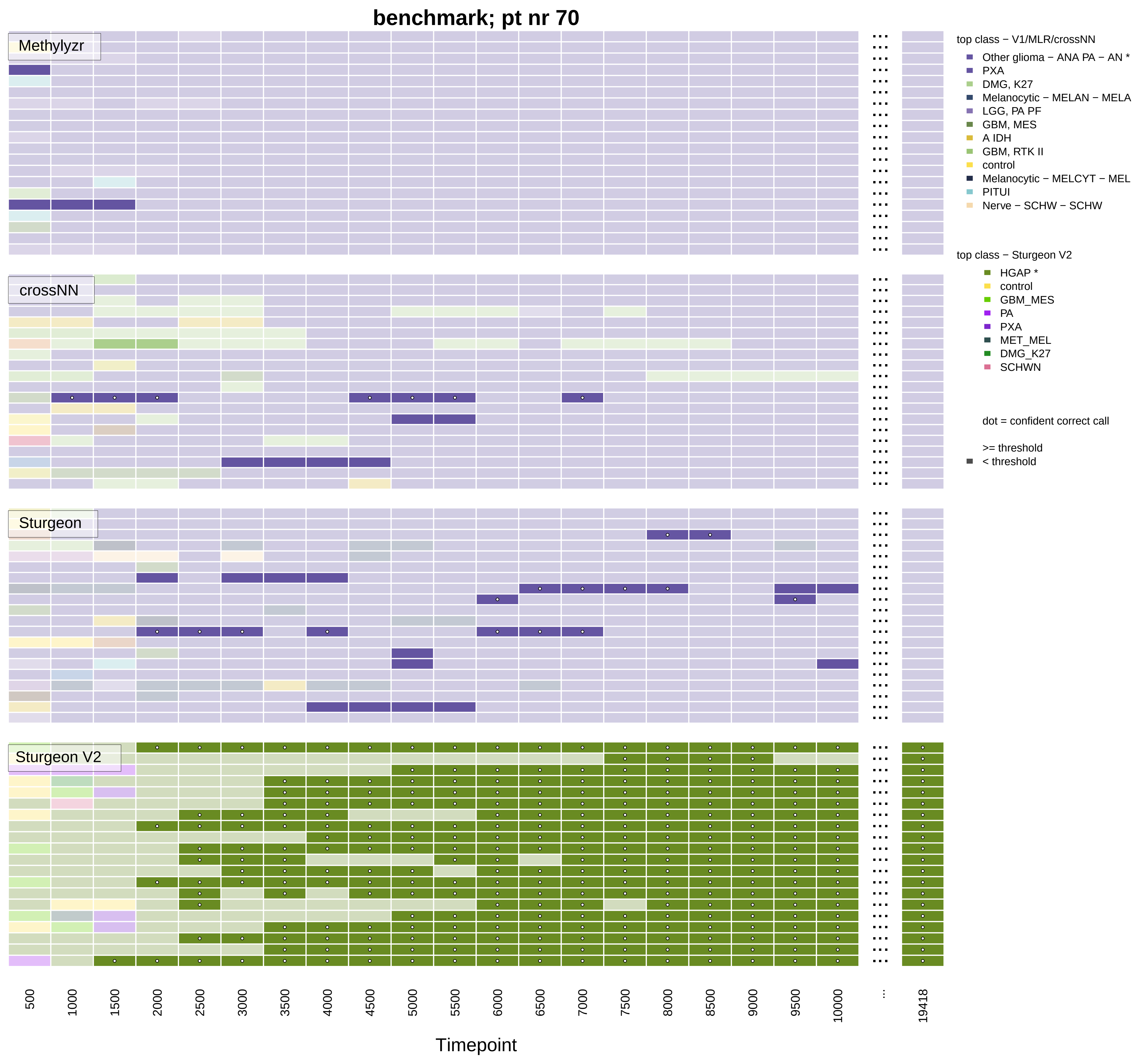

benchmark; pt nr 70
Methylyzr
crossNN
Sturgeon
Sturgeon V2
top class − V1/MLR/crossNN
Other glioma − ANA PA − AN *
PXA
DMG, K27
Melanocytic − MELAN − MELA
LGG, PA PF
GBM, MES
A IDH
GBM, RTK II
control
Melanocytic − MELCYT − MEL
PITUI
Nerve − SCHW − SCHW
top class − Sturgeon V2
HGAP *
control
GBM_MES
PA
PXA
MET_MEL
DMG_K27
SCHWN
dot = confident correct call
>= threshold
< threshold
500
1000
1500
2000
2500
3000
3500
4000
4500
5000
5500
6000
6500
7000
7500
8000
8500
9000
9500
10000
...
19418
Timepoint

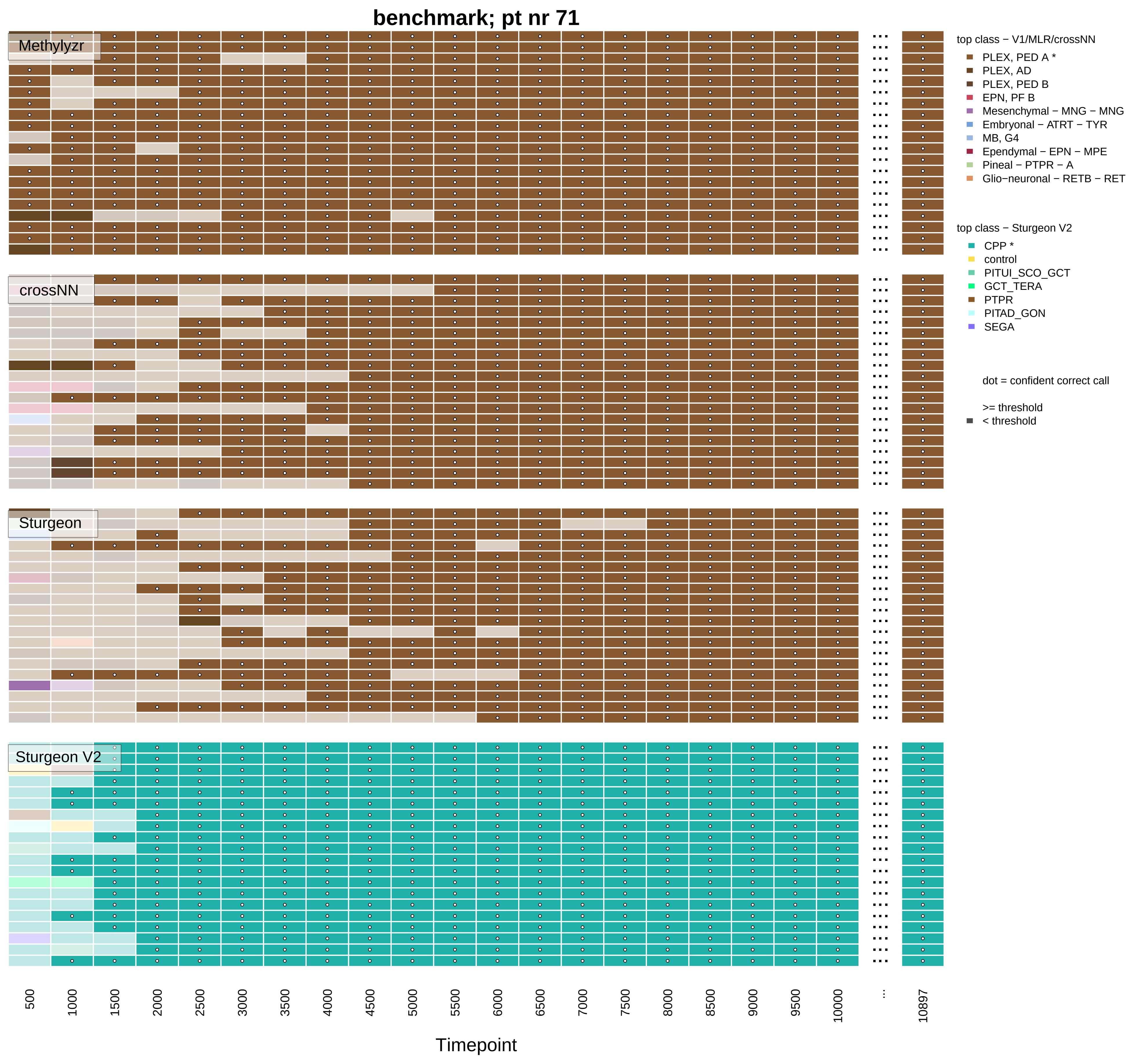

benchmark; pt nr 71
Methylyzr
crossNN
Sturgeon
Sturgeon V2
top class − V1/MLR/crossNN
PLEX, PED A *
PLEX, AD
PLEX, PED B
EPN, PF B
Mesenchymal − MNG − MNG
Embryonal − ATRT − TYR
MB, G4
Ependymal − EPN − MPE
Pineal − PTPR − A
Glio−neuronal − RETB − RET
top class − Sturgeon V2
CPP *
control
PITUI_SCO_GCT
GCT_TERA
PTPR
PITAD_GON
SEGA
dot = confident correct call
>= threshold
< threshold
500
1000
1500
2000
2500
3000
3500
4000
4500
5000
5500
6000
6500
7000
7500
8000
8500
9000
9500
10000
...
10897
Timepoint

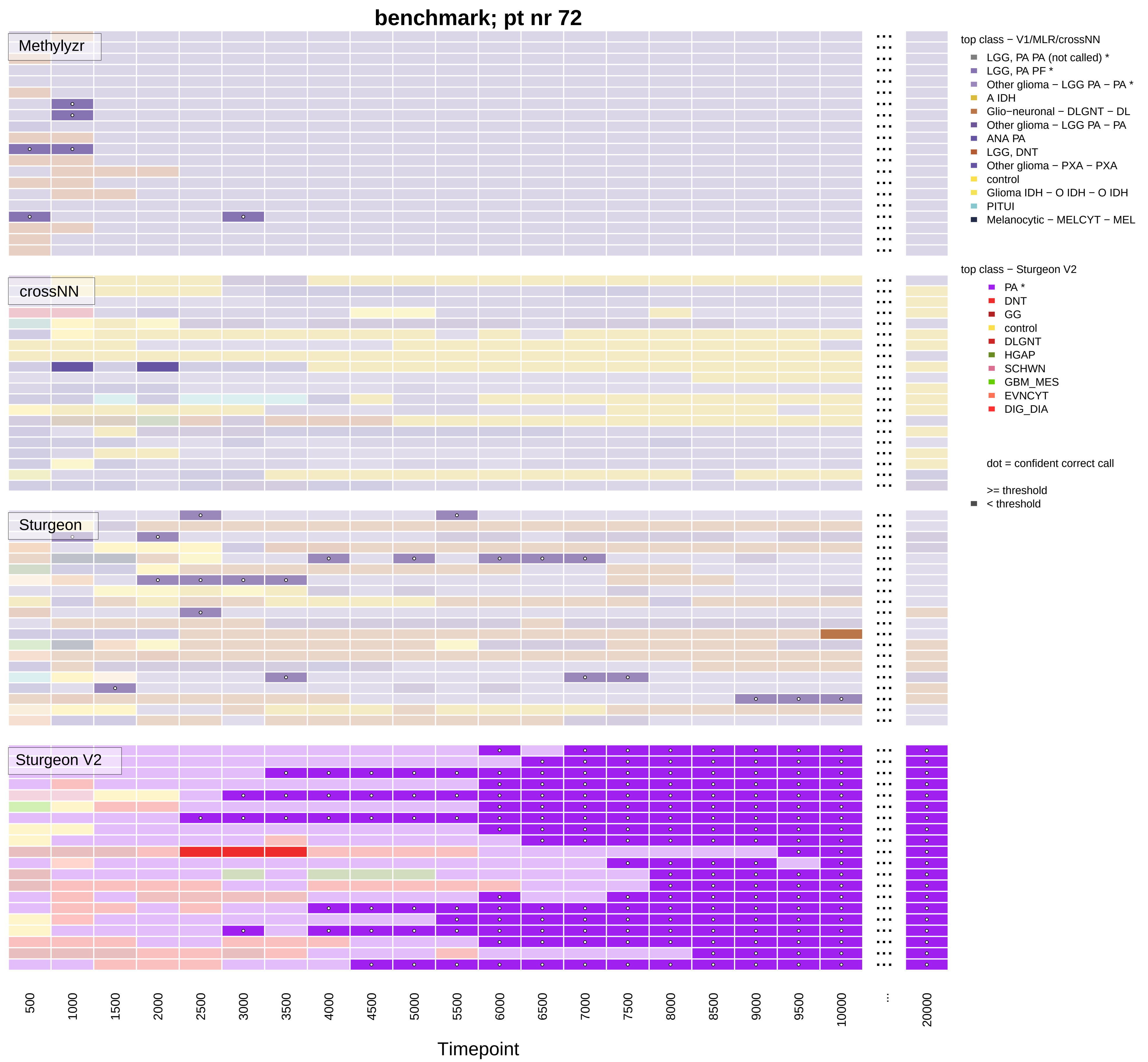

benchmark; pt nr 72
Methylyzr
crossNN
Sturgeon
Sturgeon V2
top class − V1/MLR/crossNN
LGG, PA PA (not called) *
LGG, PA PF *
Other glioma − LGG PA − PA *
A IDH
Glio−neuronal − DLGNT − DL
Other glioma − LGG PA − PA
ANA PA
LGG, DNT
Other glioma − PXA − PXA
control
Glioma IDH − O IDH − O IDH
PITUI
Melanocytic − MELCYT − MEL
top class − Sturgeon V2
PA *
DNT
GG
control
DLGNT
HGAP
SCHWN
GBM_MES
EVNCYT
DIG_DIA
dot = confident correct call
>= threshold
< threshold
500
1000
1500
2000
2500
3000
3500
4000
4500
5000
5500
6000
6500
7000
7500
8000
8500
9000
9500
10000
...
20000
Timepoint

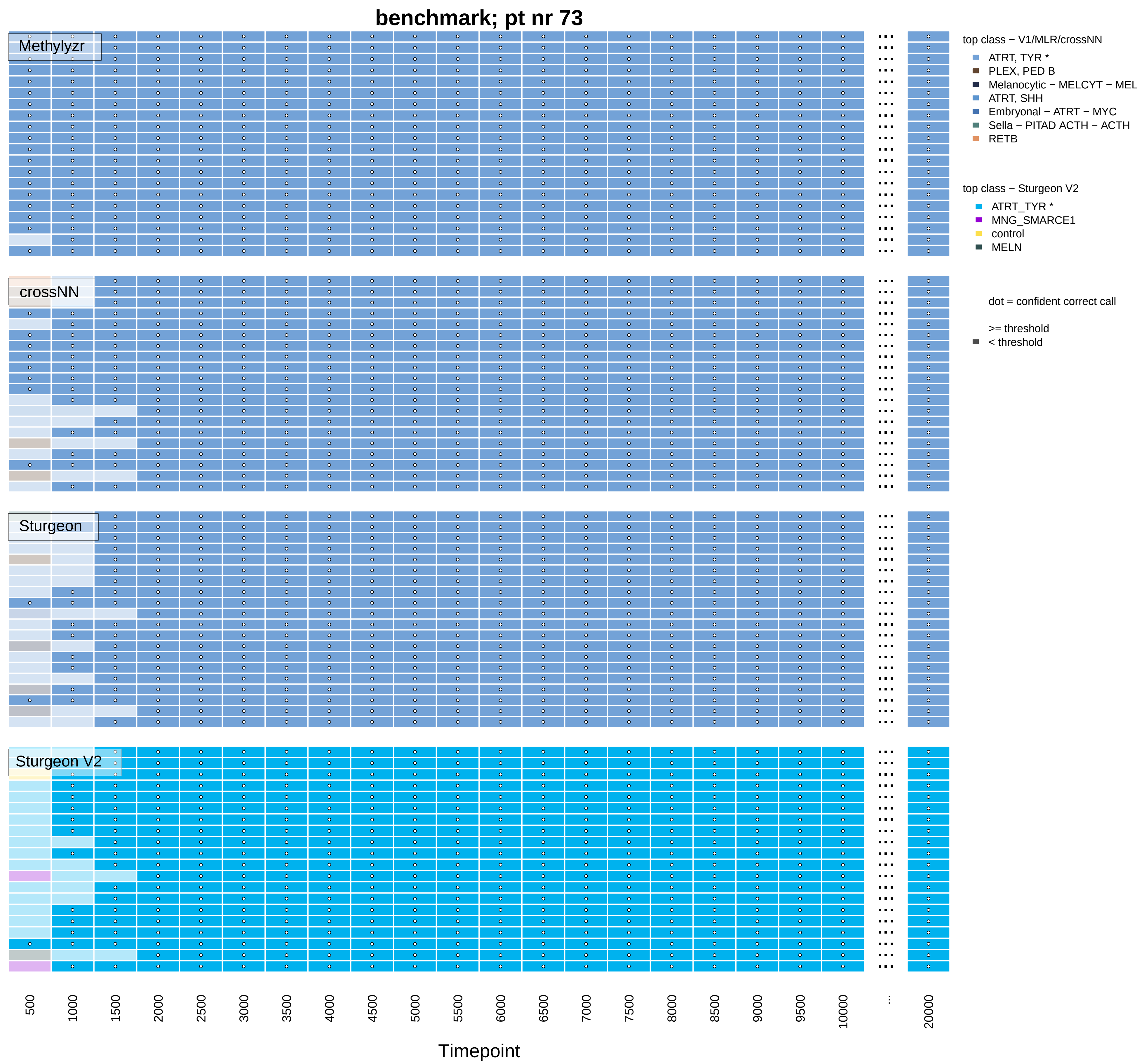
benchmark; pt nr 73
Methylyzr
crossNN
Sturgeon
Sturgeon V2
top class − V1/MLR/crossNN
ATRT, TYR *
PLEX, PED B
Melanocytic − MELCYT − MEL
ATRT, SHH
Embryonal − ATRT − MYC
Sella − PITAD ACTH − ACTH
RETB
top class − Sturgeon V2
ATRT_TYR *
MNG_SMARCE1
control
MELN
dot = confident correct call
>= threshold
< threshold
500
1000
1500
2000
2500
3000
3500
4000
4500
5000
5500
6000
6500
7000
7500
8000
8500
9000
9500
10000
...
20000
Timepoint

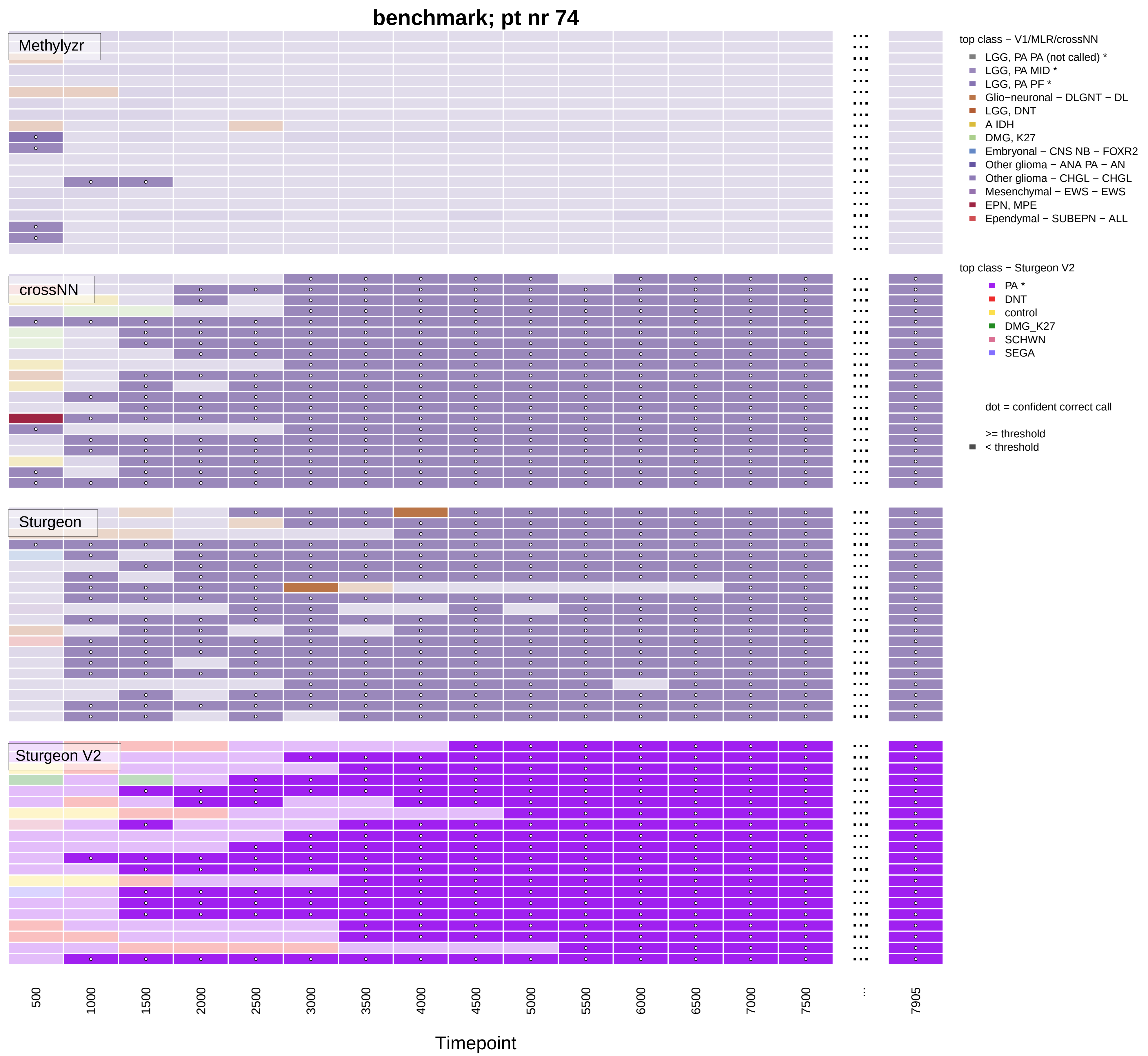

benchmark; pt nr 74
Methylyzr
crossNN
Sturgeon
Sturgeon V2
500
1000
1500
2000
2500
3000
3500
4000
4500
5000
5500
6000
6500
7000
7500
...
7905
Timepoint
top class − V1/MLR/crossNN
LGG, PA PA (not called) *
LGG, PA MID *
LGG, PA PF *
Glio−neuronal − DLGNT − DL
LGG, DNT
A IDH
DMG, K27
Embryonal − CNS NB − FOXR2
Other glioma − ANA PA − AN
Other glioma − CHGL − CHGL
Mesenchymal − EWS − EWS
EPN, MPE
Ependymal − SUBEPN − ALL
top class − Sturgeon V2
PA *
DNT
control
DMG_K27
SCHWN
SEGA
dot = confident correct call
>= threshold
< threshold

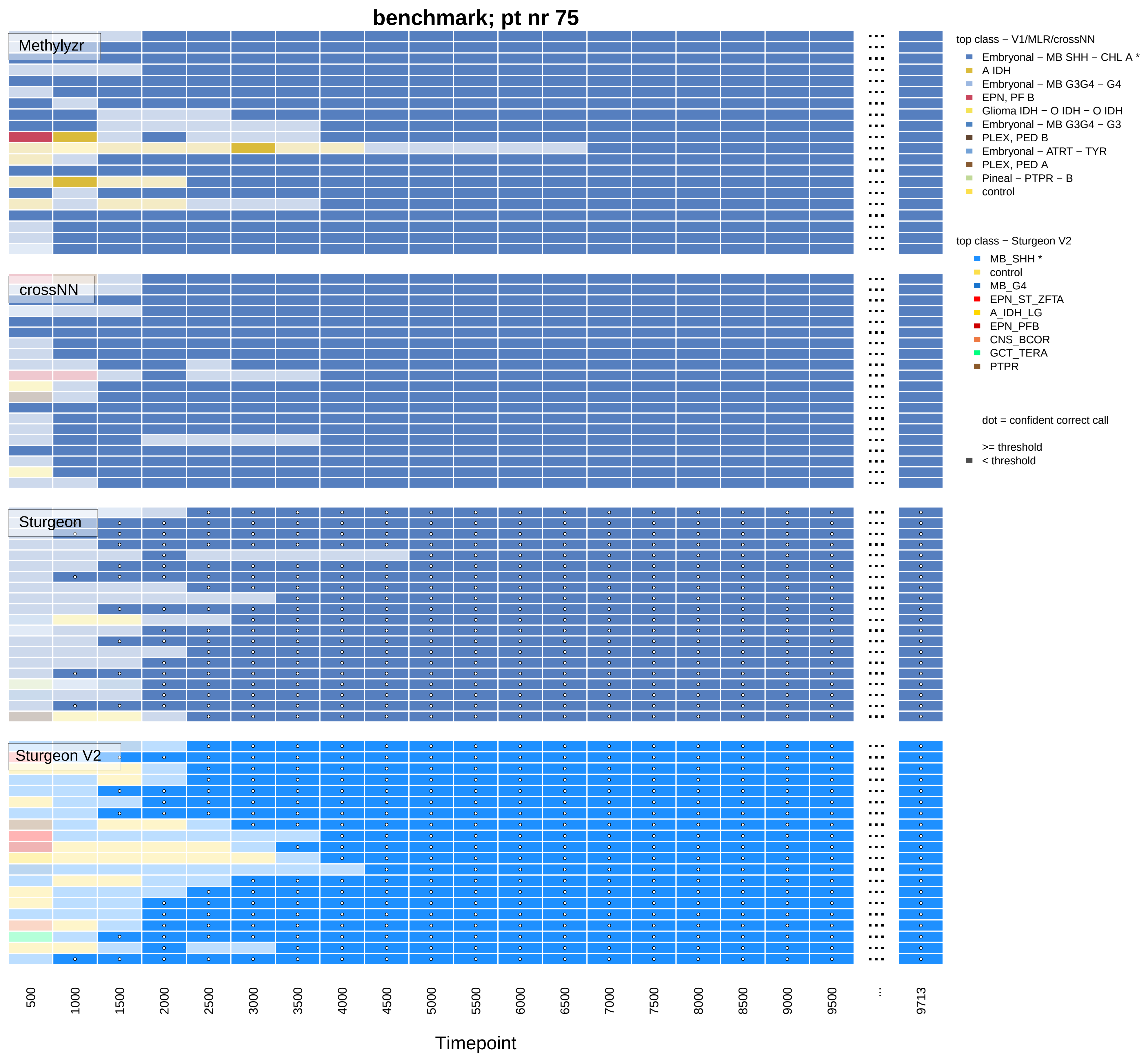

benchmark; pt nr 75
Methylyzr
crossNN
Sturgeon
Sturgeon V2
top class − V1/MLR/crossNN
Embryonal − MB SHH − CHL A *
A IDH
Embryonal − MB G3G4 − G4
EPN, PF B
Glioma IDH − O IDH − O IDH
Embryonal − MB G3G4 − G3
PLEX, PED B
Embryonal − ATRT − TYR
PLEX, PED A
Pineal − PTPR − B
control
top class − Sturgeon V2
MB_SHH *
control
MB_G4
EPN_ST_ZFTA
A_IDH_LG
EPN_PFB
CNS_BCOR
GCT_TERA
PTPR
dot = confident correct call
>= threshold
< threshold
500
1000
1500
2000
2500
3000
3500
4000
4500
5000
5500
6000
6500
7000
7500
8000
8500
9000
9500
...
9713
Timepoint

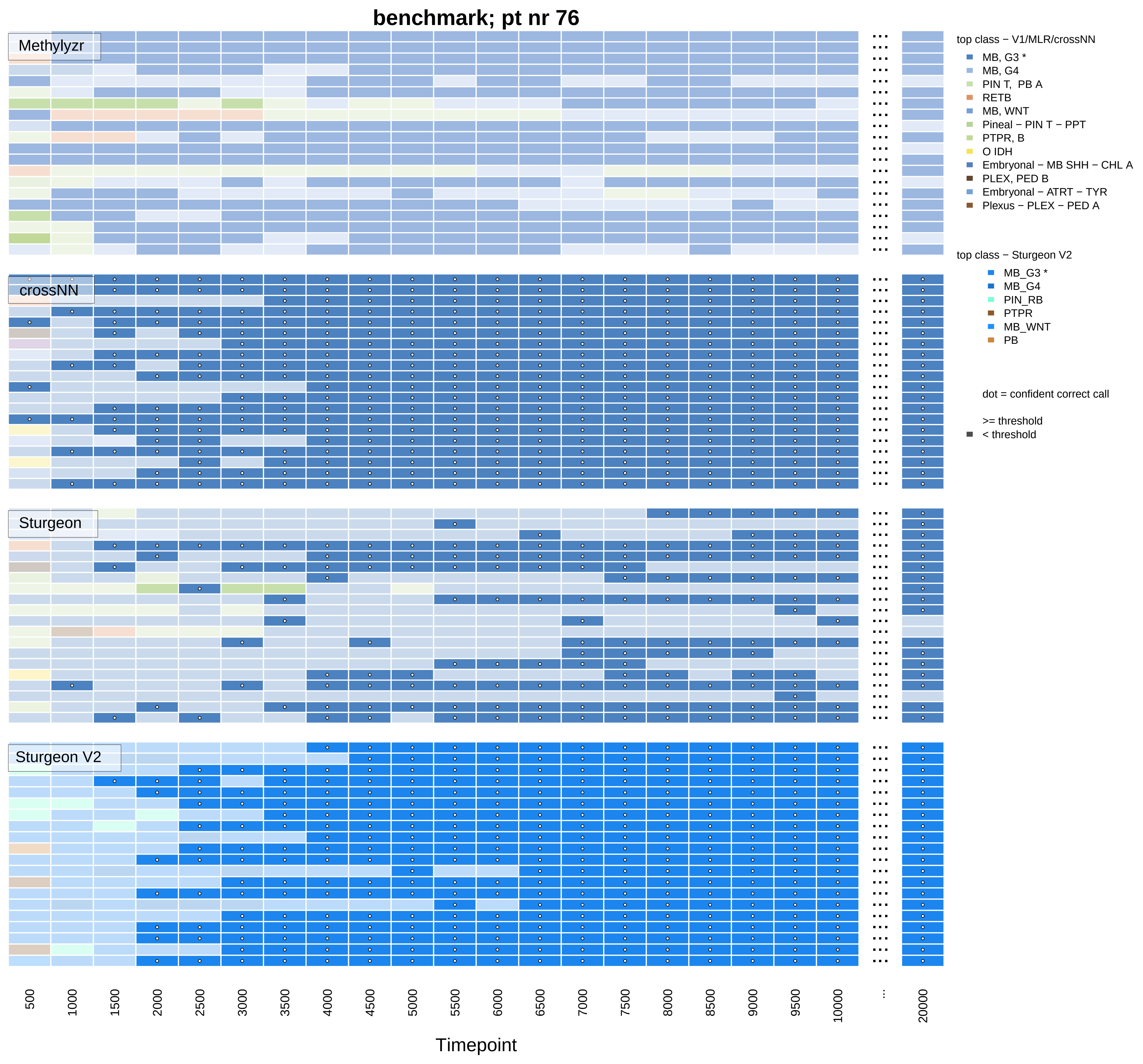
benchmark; pt nr 76
Methylyzr
crossNN
Sturgeon
Sturgeon V2
500
1000
1500
2000
2500
3000
3500
4000
4500
5000
5500
6000
6500
7000
7500
8000
8500
9000
9500
10000
...
20000
Timepoint
top class − V1/MLR/crossNN
MB, G3 *
MB, G4
PIN T, PB A
RETB
MB, WNT
Pineal − PIN T − PPT
PTPR, B
O IDH
Embryonal − MB SHH − CHL A
PLEX, PED B
Embryonal − ATRT − TYR
Plexus − PLEX − PED A
top class − Sturgeon V2
MB_G3 *
MB_G4
PIN_RB
PTPR
MB_WNT
PB
dot = confident correct call
>= threshold
< threshold

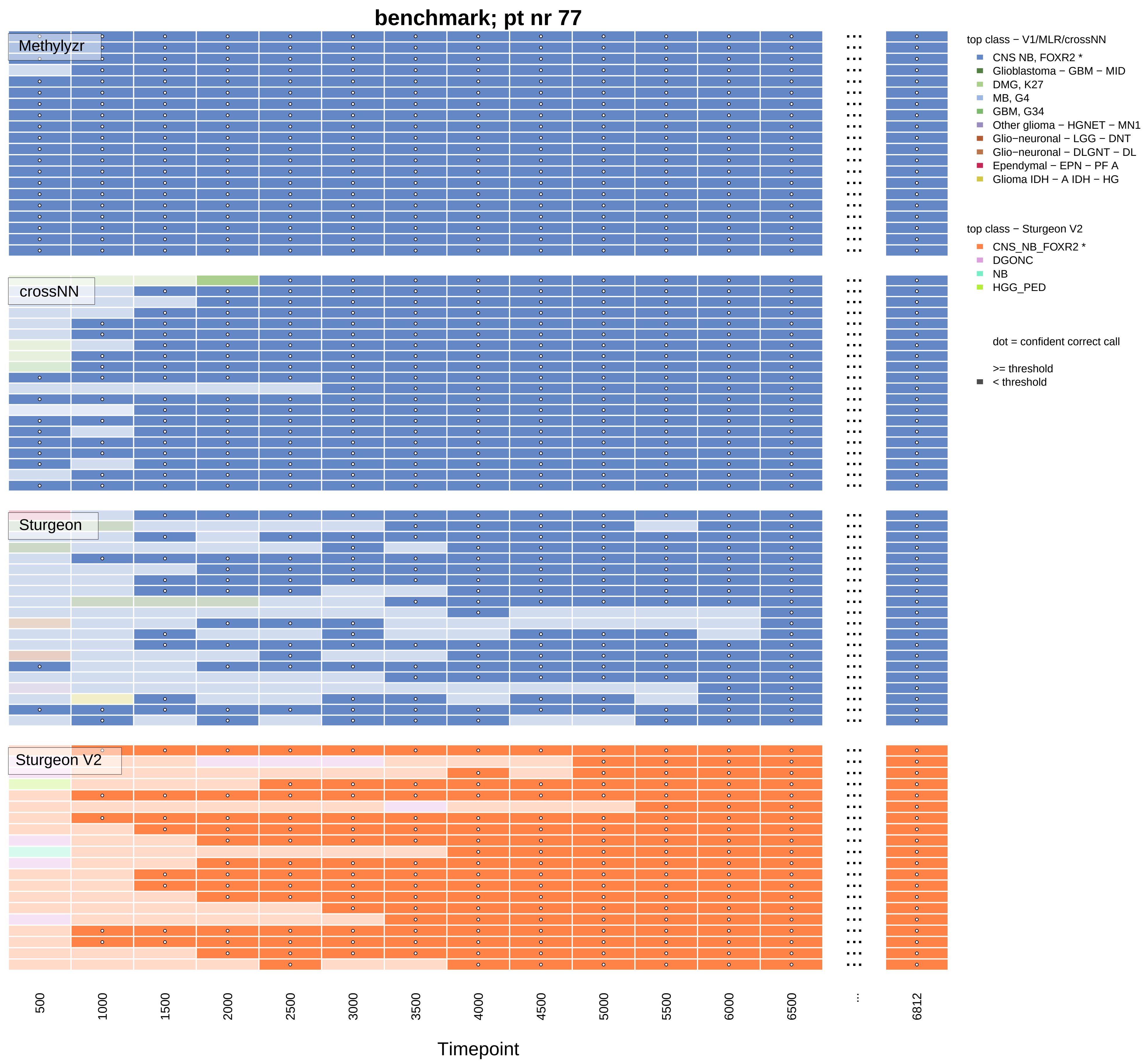
benchmark; pt nr 77
Methylyzr
crossNN
Sturgeon
Sturgeon V2
500
1000
1500
2000
2500
3000
3500
4000
4500
5000
5500
6000
6500
...
6812
Timepoint
top class − V1/MLR/crossNN
CNS NB, FOXR2 *
Glioblastoma − GBM − MID
DMG, K27
MB, G4
GBM, G34
Other glioma − HGNET − MN1
Glio−neuronal − LGG − DNT
Glio−neuronal − DLGNT − DL
Ependymal − EPN − PF A
Glioma IDH − A IDH − HG
top class − Sturgeon V2
CNS_NB_FOXR2 *
DGONC
NB
HGG_PED
dot = confident correct call
>= threshold
< threshold

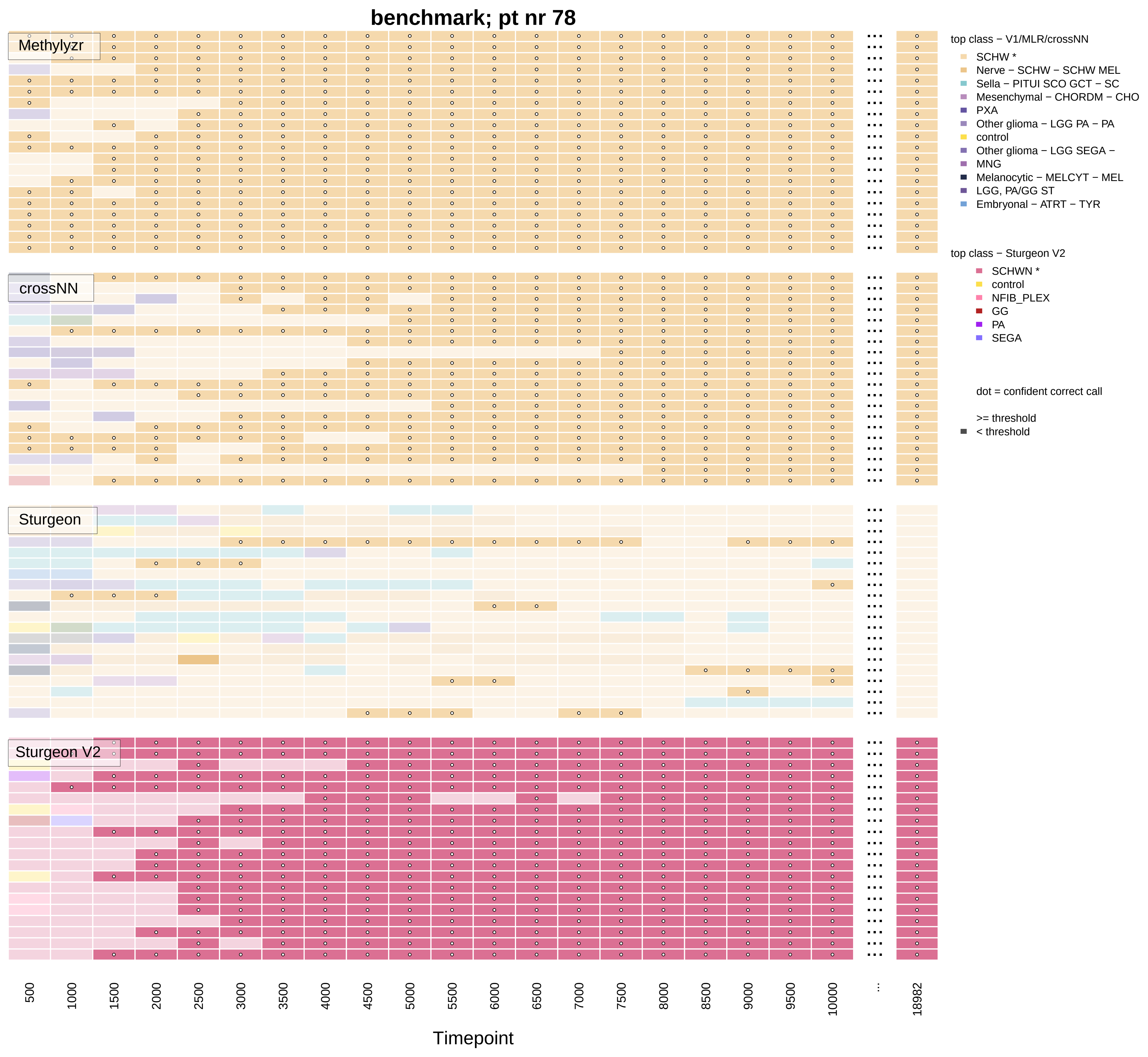

benchmark; pt nr 78
Methylyzr
crossNN
Sturgeon
Sturgeon V2
top class − V1/MLR/crossNN
SCHW *
Nerve − SCHW − SCHW MEL
Sella − PITUI SCO GCT − SC
Mesenchymal − CHORDM − CHO
PXA
Other glioma − LGG PA − PA
control
Other glioma − LGG SEGA −
MNG
Melanocytic − MELCYT − MEL
LGG, PA/GG ST
Embryonal − ATRT − TYR
top class − Sturgeon V2
SCHWN *
control
NFIB_PLEX
GG
PA
SEGA
dot = confident correct call
>= threshold
< threshold
500
1000
1500
2000
2500
3000
3500
4000
4500
5000
5500
6000
6500
7000
7500
8000
8500
9000
9500
10000
...
18982
Timepoint

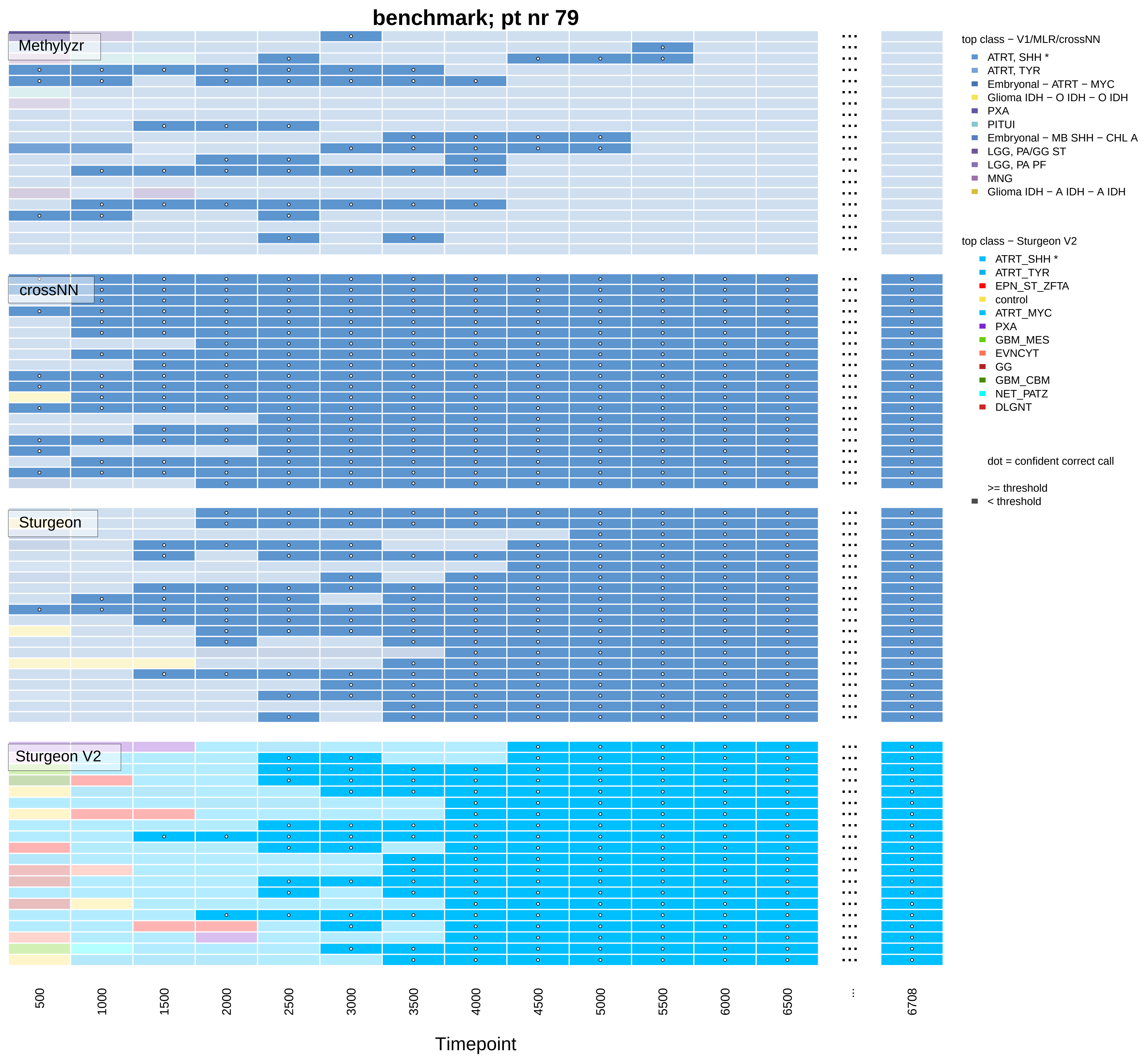
benchmark; pt nr 79
Methylyzr
crossNN
Sturgeon
Sturgeon V2
500
1000
1500
2000
2500
3000
3500
4000
4500
5000
5500
6000
6500
...
6708
Timepoint
top class − V1/MLR/crossNN
ATRT, SHH *
ATRT, TYR
Embryonal − ATRT − MYC
Glioma IDH − O IDH − O IDH
PXA
PITUI
Embryonal − MB SHH − CHL A
LGG, PA/GG ST
LGG, PA PF
MNG
Glioma IDH − A IDH − A IDH
top class − Sturgeon V2
ATRT_SHH *
ATRT_TYR
EPN_ST_ZFTA
control
ATRT_MYC
PXA
GBM_MES
EVNCYT
GG
GBM_CBM
NET_PATZ
DLGNT
dot = confident correct call
>= threshold
< threshold

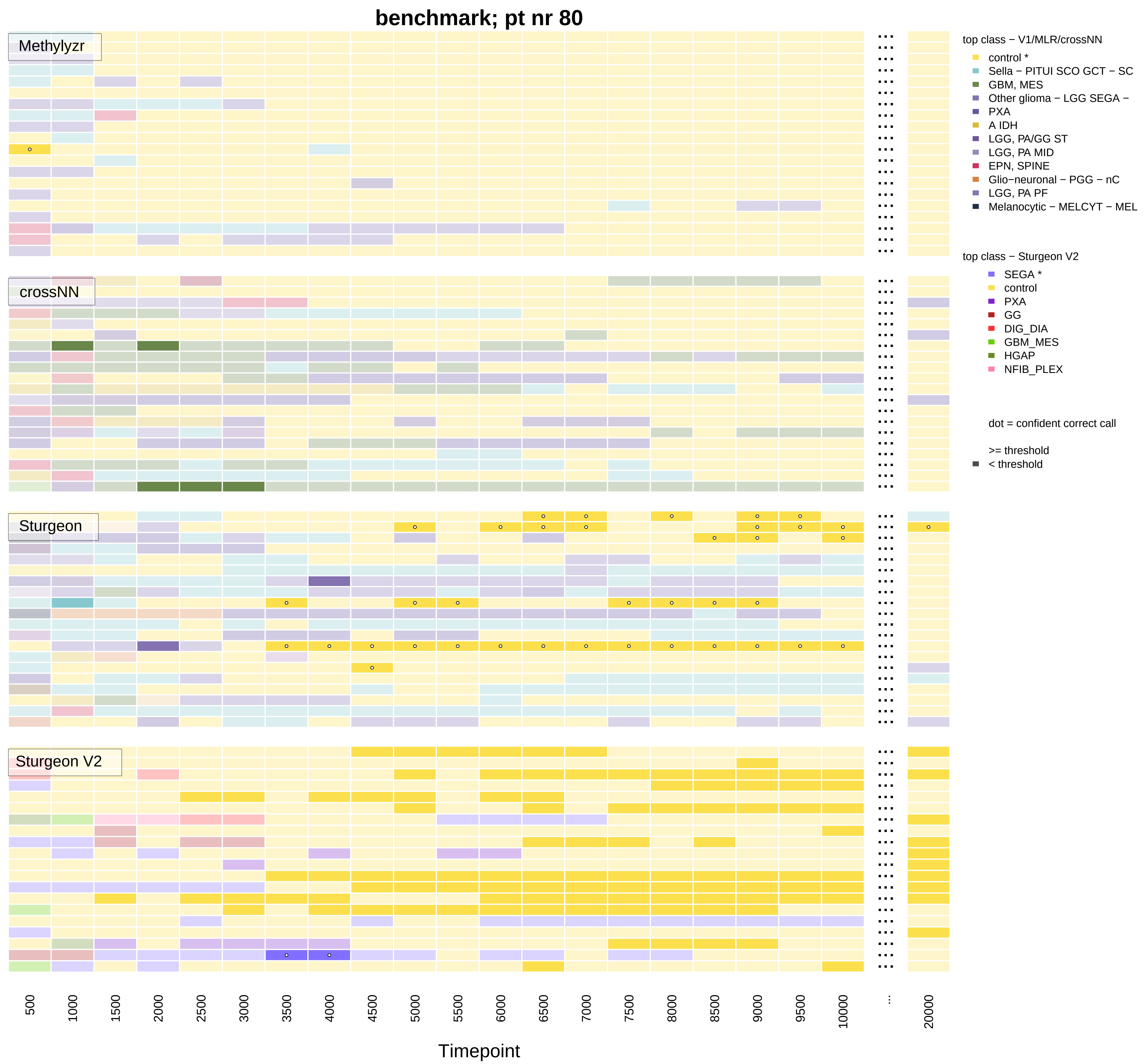
benchmark; pt nr 80
Methylyzr
crossNN
Sturgeon
Sturgeon V2
top class − V1/MLR/crossNN
control *
Sella − PITUI SCO GCT − SC
GBM, MES
Other glioma − LGG SEGA −
PXA
A IDH
LGG, PA/GG ST
LGG, PA MID
EPN, SPINE
Glio−neuronal − PGG − nC
LGG, PA PF
Melanocytic − MELCYT − MEL
top class − Sturgeon V2
SEGA *
control
PXA
GG
DIG_DIA
GBM_MES
HGAP
NFIB_PLEX
dot = confident correct call
>= threshold
< threshold
500
1000
1500
2000
2500
3000
3500
4000
4500
5000
5500
6000
6500
7000
7500
8000
8500
9000
9500
10000
...
20000
Timepoint

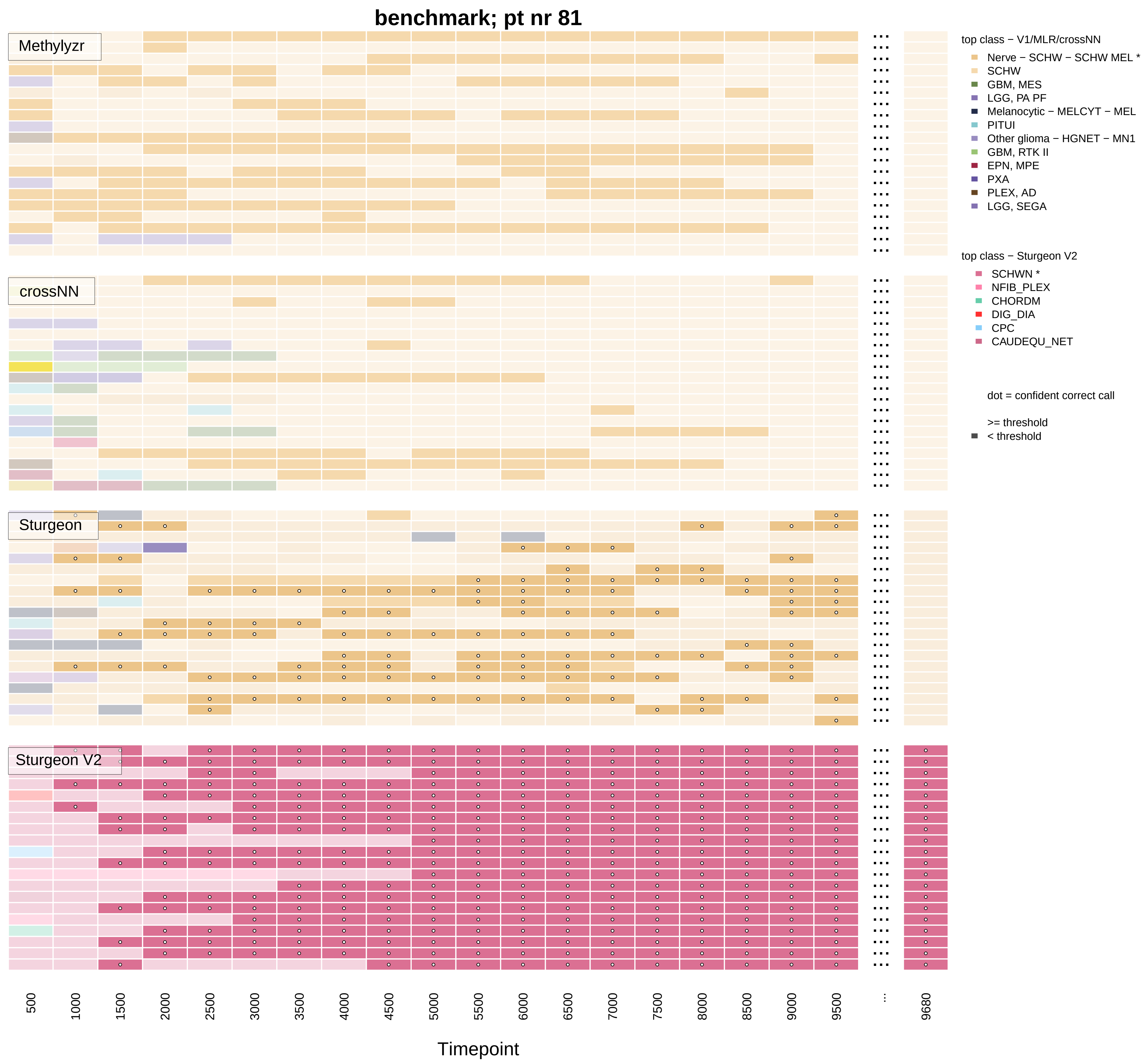

benchmark; pt nr 81
Methylyzr
crossNN
Sturgeon
Sturgeon V2
top class − V1/MLR/crossNN
Nerve − SCHW − SCHW MEL *
SCHW
GBM, MES
LGG, PA PF
Melanocytic − MELCYT − MEL
PITUI
Other glioma − HGNET − MN1
GBM, RTK II
EPN, MPE
PXA
PLEX, AD
LGG, SEGA
top class − Sturgeon V2
SCHWN *
NFIB_PLEX
CHORDM
DIG_DIA
CPC
CAUDEQU_NET
dot = confident correct call
>= threshold
< threshold
500
1000
1500
2000
2500
3000
3500
4000
4500
5000
5500
6000
6500
7000
7500
8000
8500
9000
9500
...
9680
Timepoint

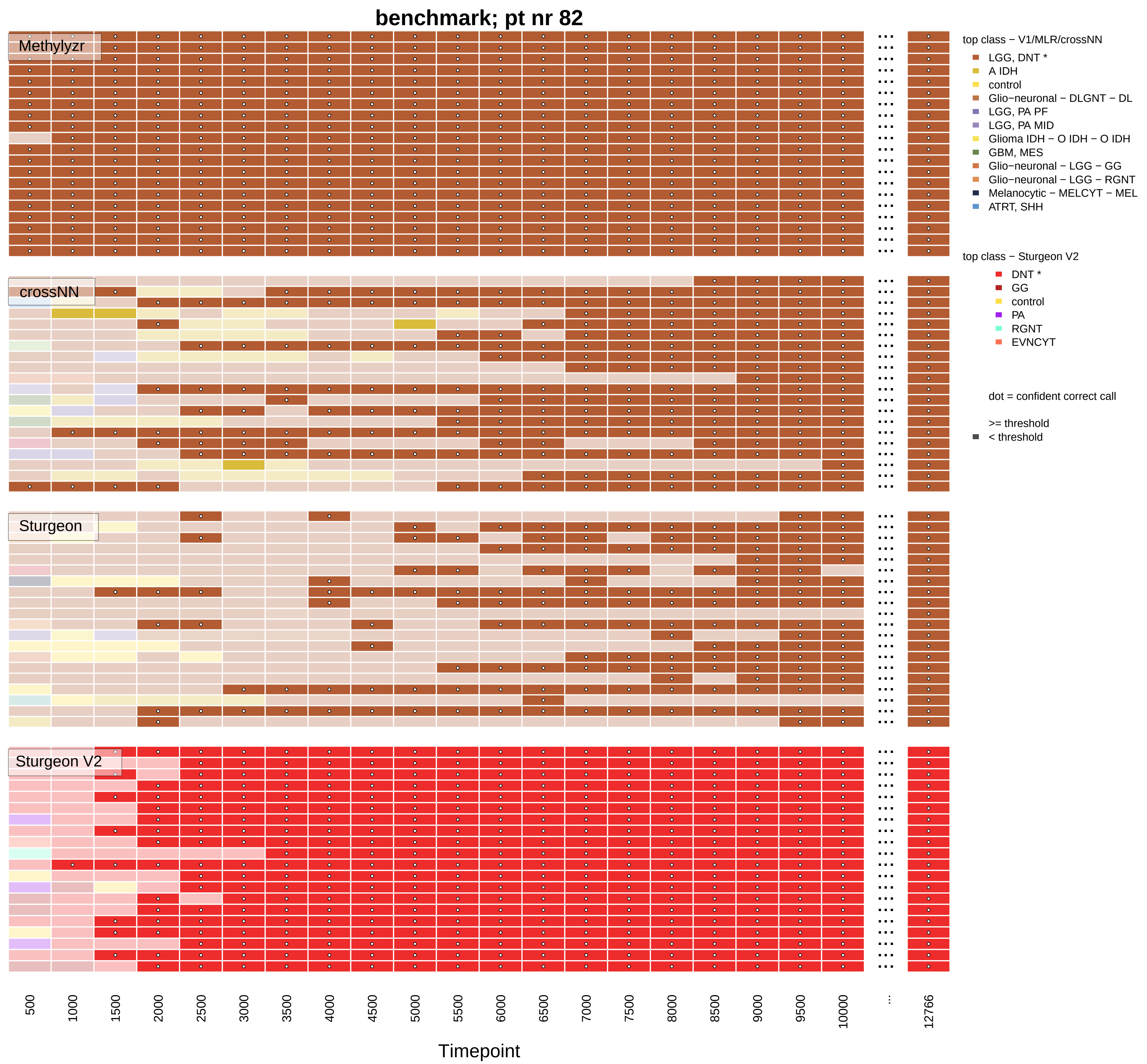
benchmark; pt nr 82
Methylyzr
crossNN
Sturgeon
Sturgeon V2
top class − V1/MLR/crossNN
LGG, DNT *
A IDH
control
Glio−neuronal − DLGNT − DL
LGG, PA PF
LGG, PA MID
Glioma IDH − O IDH − O IDH
GBM, MES
Glio−neuronal − LGG − GG
Glio−neuronal − LGG − RGNT
Melanocytic − MELCYT − MEL
ATRT, SHH
top class − Sturgeon V2
DNT *
GG
control
PA
RGNT
EVNCYT
dot = confident correct call
>= threshold
< threshold
500
1000
1500
2000
2500
3000
3500
4000
4500
5000
5500
6000
6500
7000
7500
8000
8500
9000
9500
10000
...
12766
Timepoint

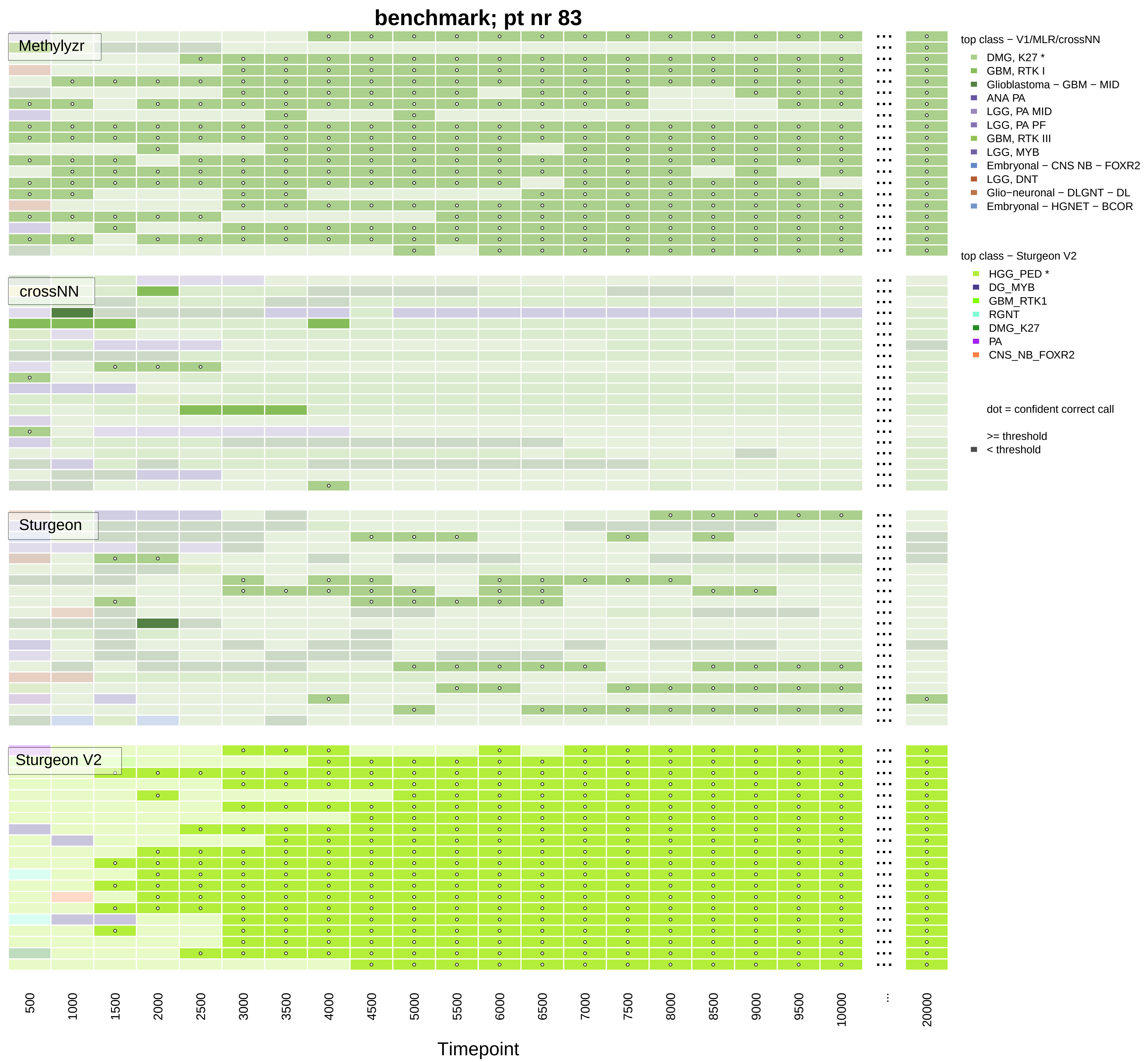
benchmark; pt nr 83
Methylyzr
crossNN
Sturgeon
Sturgeon V2
top class − V1/MLR/crossNN
DMG, K27 *
GBM, RTK I
Glioblastoma − GBM − MID
ANA PA
LGG, PA MID
LGG, PA PF
GBM, RTK III
LGG, MYB
Embryonal − CNS NB − FOXR2
LGG, DNT
Glio−neuronal − DLGNT − DL
Embryonal − HGNET − BCOR
top class − Sturgeon V2
HGG_PED *
DG_MYB
GBM_RTK1
RGNT
DMG_K27
PA
CNS_NB_FOXR2
dot = confident correct call
>= threshold
< threshold
500
1000
1500
2000
2500
3000
3500
4000
4500
5000
5500
6000
6500
7000
7500
8000
8500
9000
9500
10000
...
20000
Timepoint

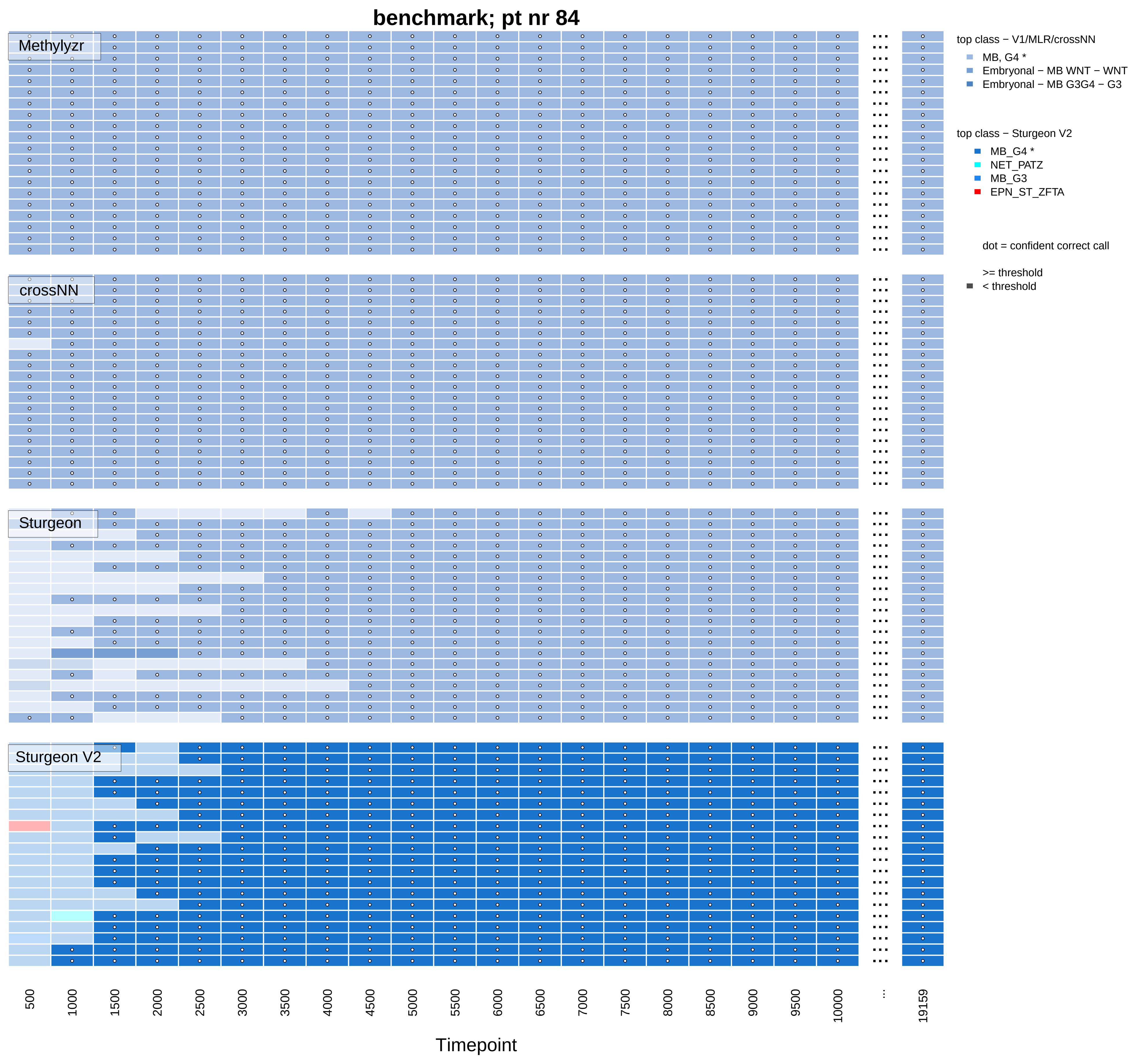

benchmark; pt nr 84
Methylyzr
crossNN
Sturgeon
Sturgeon V2
top class − V1/MLR/crossNN
MB, G4 *
Embryonal − MB WNT − WNT
Embryonal − MB G3G4 − G3
top class − Sturgeon V2
MB_G4 *
NET_PATZ
MB_G3
EPN_ST_ZFTA
dot = confident correct call
>= threshold
< threshold
500
1000
1500
2000
2500
3000
3500
4000
4500
5000
5500
6000
6500
7000
7500
8000
8500
9000
9500
10000
...
19159
Timepoint

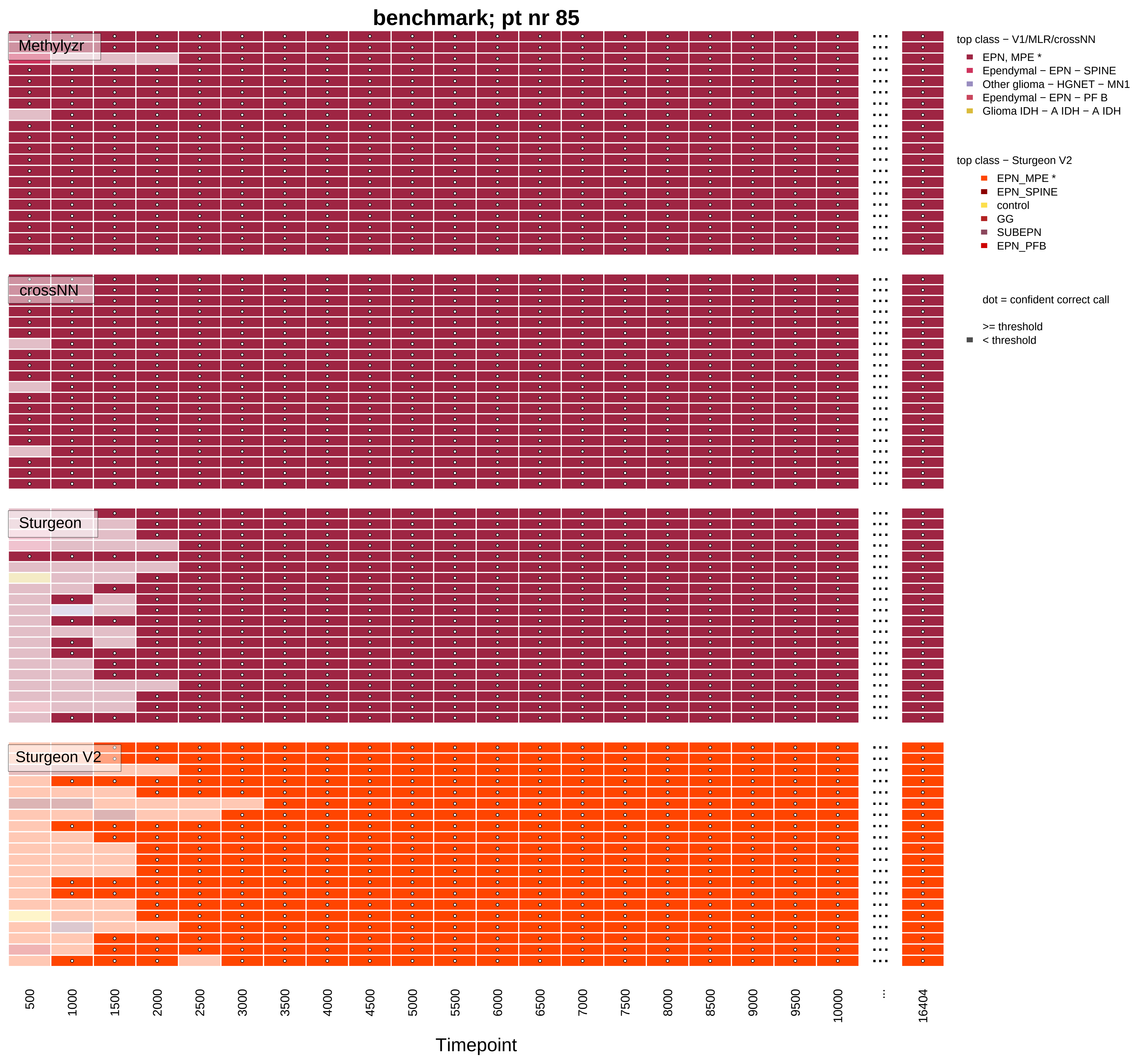

benchmark; pt nr 85
Methylyzr
crossNN
Sturgeon
Sturgeon V2
top class − V1/MLR/crossNN
EPN, MPE *
Ependymal − EPN − SPINE
Other glioma − HGNET − MN1
Ependymal − EPN − PF B
Glioma IDH − A IDH − A IDH
top class − Sturgeon V2
EPN_MPE *
EPN_SPINE
control
GG
SUBEPN
EPN_PFB
dot = confident correct call
>= threshold
< threshold
500 1000 1500 2000 2500 3000 3500 4000 4500 5000 5500 6000 6500 7000 7500 8000 8500 9000 9500 10000 ... 16404
Timepoint

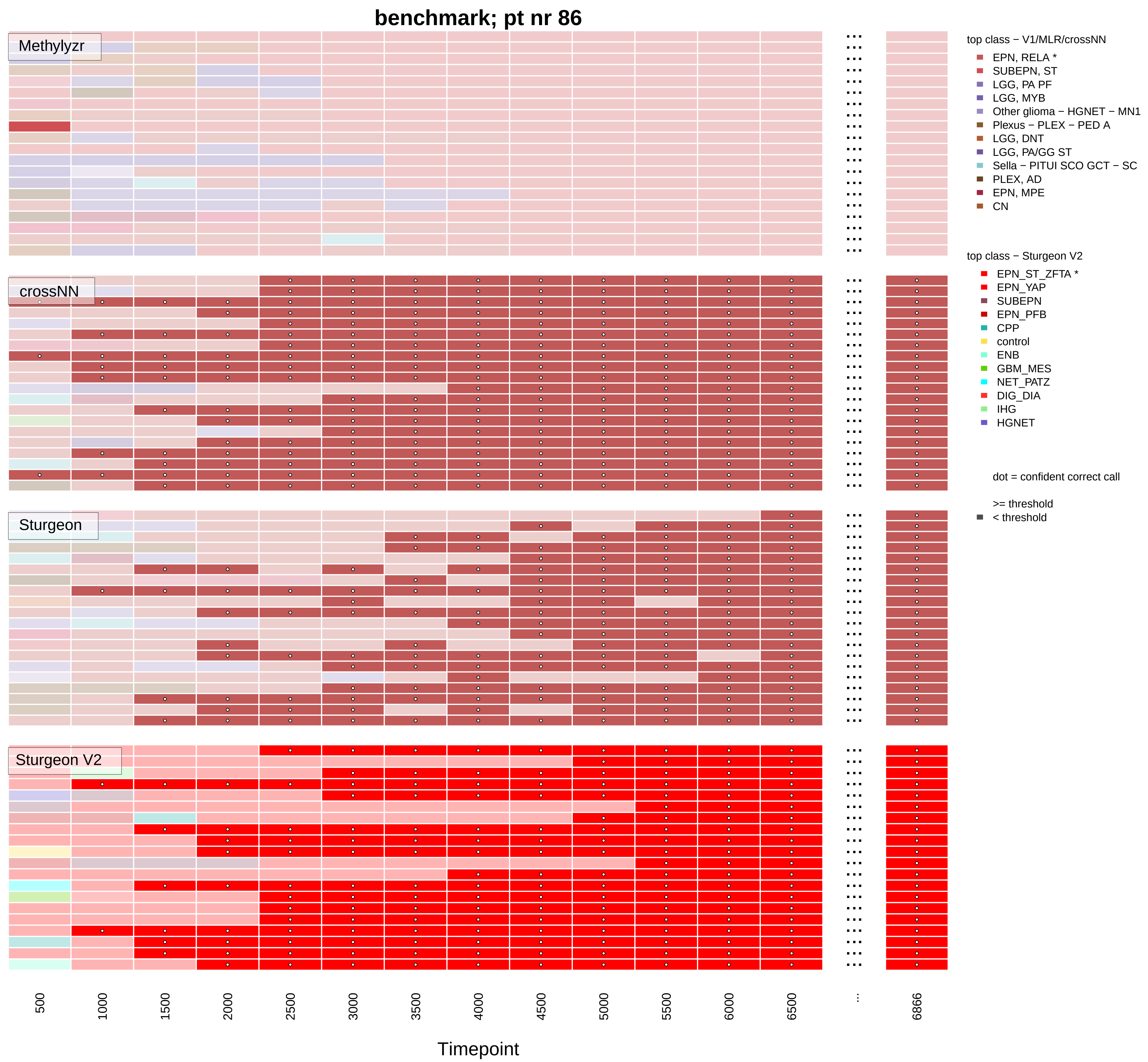

benchmark; pt nr 86
Methylyzr
crossNN
Sturgeon
Sturgeon V2
500
1000
1500
2000
2500
3000
3500
4000
4500
5000
5500
6000
6500
...
6866
Timepoint
top class − V1/MLR/crossNN
EPN, RELA *
SUBEPN, ST
LGG, PA PF
LGG, MYB
Other glioma − HGNET − MN1
Plexus − PLEX − PED A
LGG, DNT
LGG, PA/GG ST
Sella − PITUI SCO GCT − SC
PLEX, AD
EPN, MPE
CN
top class − Sturgeon V2
EPN_ST_ZFTA *
EPN_YAP
SUBEPN
EPN_PFB
CPP
control
ENB
GBM_MES
NET_PATZ
DIG_DIA
IHG
HGNET
dot = confident correct call
>= threshold
< threshold

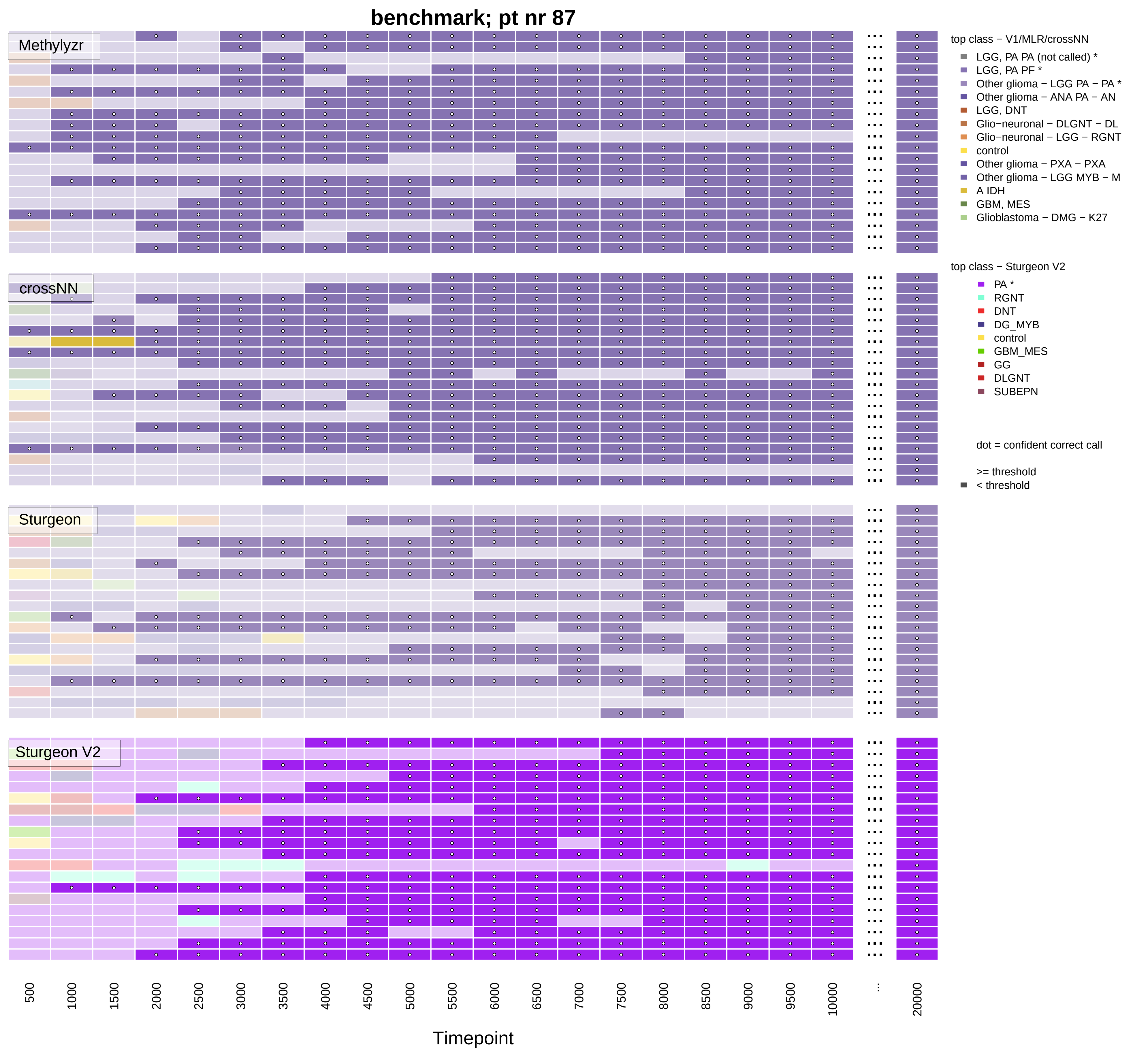

benchmark; pt nr 87
Methylyzr
crossNN
Sturgeon
Sturgeon V2
top class − V1/MLR/crossNN
LGG, PA PA (not called) *
LGG, PA PF *
Other glioma − LGG PA − PA *
Other glioma − ANA PA − AN
LGG, DNT
Glio−neuronal − DLGNT − DL
Glio−neuronal − LGG − RGNT
control
Other glioma − PXA − PXA
Other glioma − LGG MYB − M
A IDH
GBM, MES
Glioblastoma − DMG − K27
top class − Sturgeon V2
PA *
RGNT
DNT
DG_MYB
control
GBM_MES
GG
DLGNT
SUBEPN
dot = confident correct call
>= threshold
< threshold
500
1000
1500
2000
2500
3000
3500
4000
4500
5000
5500
6000
6500
7000
7500
8000
8500
9000
9500
10000
...
20000
Timepoint

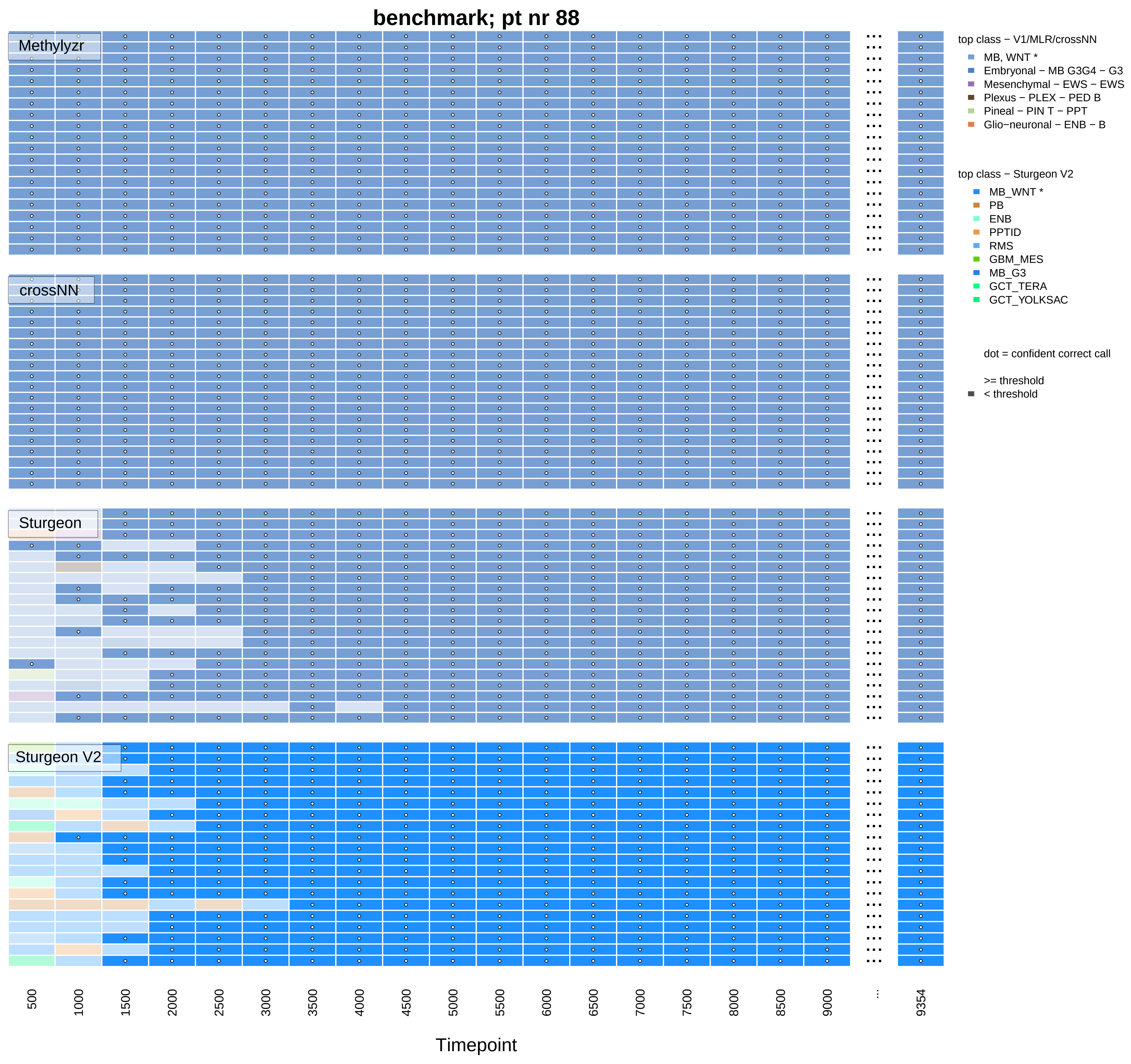
benchmark; pt nr 88
Methylyzr
crossNN
Sturgeon
Sturgeon V2
top class − V1/MLR/crossNN
MB, WNT *
Embryonal − MB G3G4 − G3
Mesenchymal − EWS − EWS
Plexus − PLEX − PED B
Pineal − PIN T − PPT
Glio−neuronal − ENB − B
top class − Sturgeon V2
MB_WNT *
PB
ENB
PPTID
RMS
GBM_MES
MB_G3
GCT_TERA
GCT_YOLKSAC
dot = confident correct call
>= threshold
< threshold
500
1000
1500
2000
2500
3000
3500
4000
4500
5000
5500
6000
6500
7000
7500
8000
8500
9000
...
9354
Timepoint

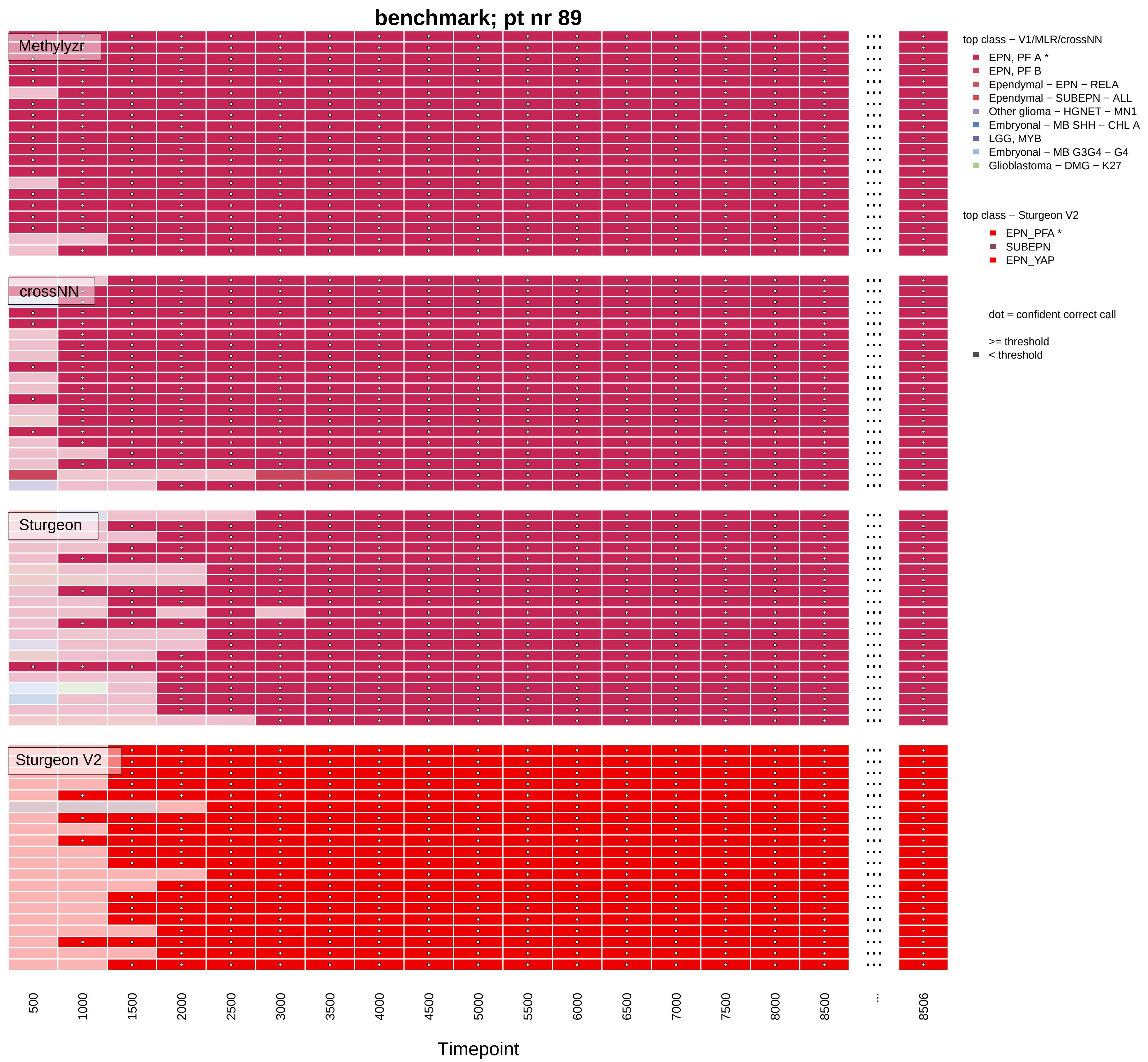
benchmark; pt nr 89
Methylyzr
crossNN
Sturgeon
Sturgeon V2
500
1000
1500
2000
2500
3000
3500
4000
4500
5000
5500
6000
6500
7000
7500
8000
8500
...
8506
Timepoint
top class − V1/MLR/crossNN
EPN, PF A *
EPN, PF B
Ependymal − EPN − RELA
Ependymal − SUBEPN − ALL
Other glioma − HGNET − MN1
Embryonal − MB SHH − CHL A
LGG, MYB
Embryonal − MB G3G4 − G4
Glioblastoma − DMG − K27
top class − Sturgeon V2
EPN_PFA *
SUBEPN
EPN_YAP
dot = confident correct call
>= threshold
< threshold

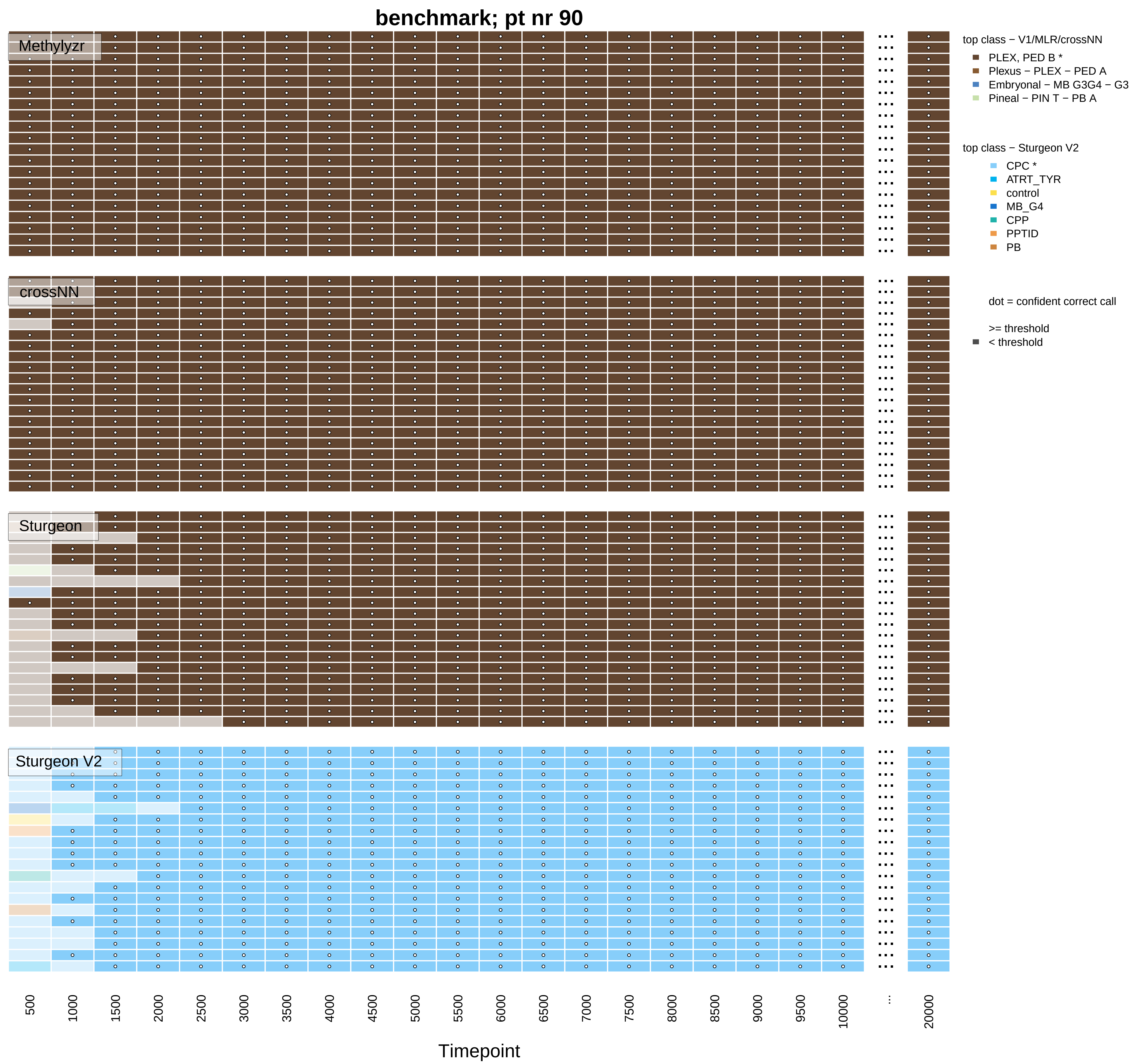

benchmark; pt nr 90
Methylyzr
crossNN
Sturgeon
Sturgeon V2
top class − V1/MLR/crossNN
PLEX, PED B *
Plexus − PLEX − PED A
Embryonal − MB G3G4 − G3
Pineal − PIN T − PB A
top class − Sturgeon V2
CPC *
ATRT_TYR
control
MB_G4
CPP
PPTID
PB
dot = confident correct call
>= threshold
< threshold
500
1000
1500
2000
2500
3000
3500
4000
4500
5000
5500
6000
6500
7000
7500
8000
8500
9000
9500
10000
...
20000
Timepoint

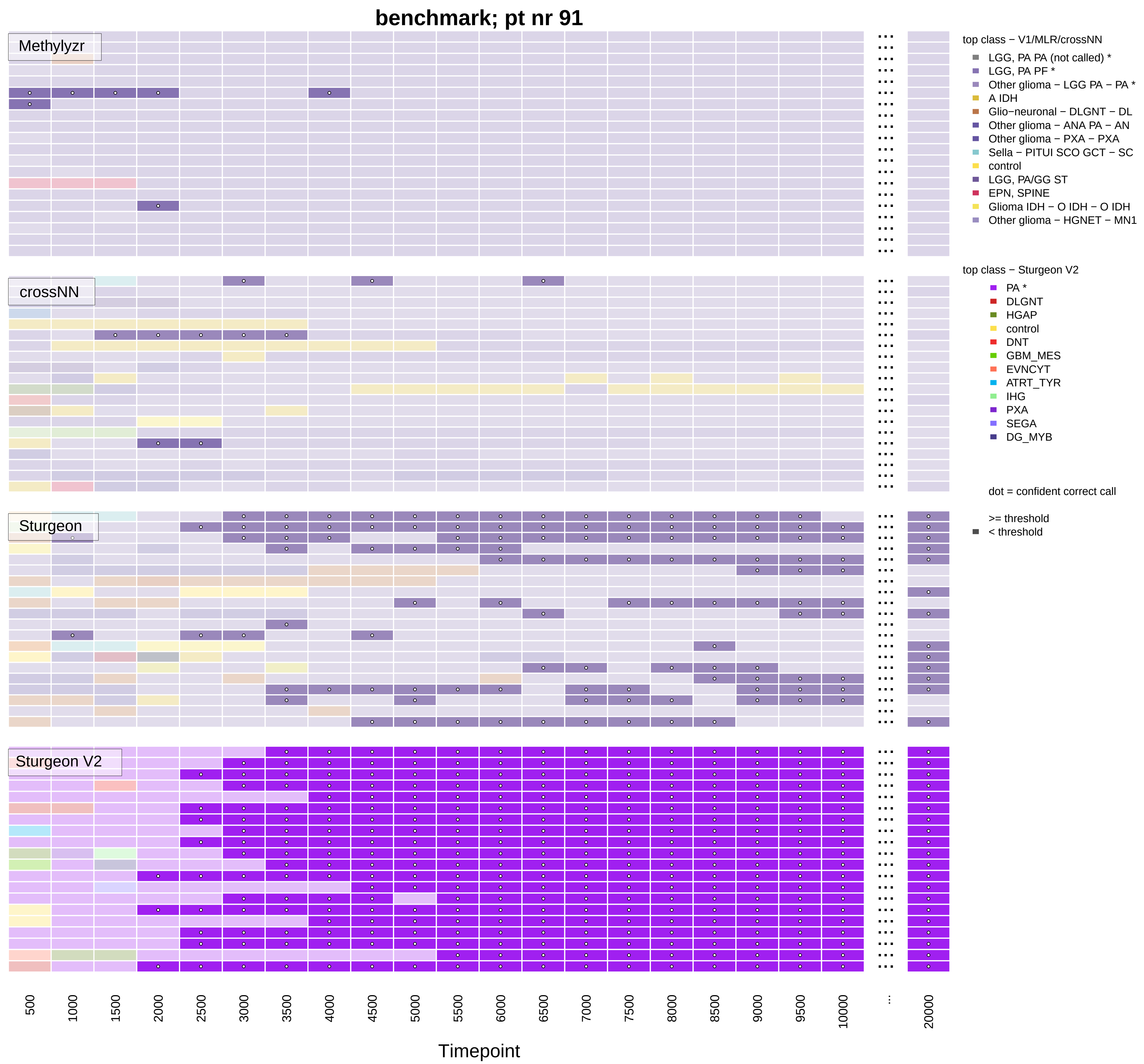
benchmark; pt nr 91
Methylyzr
crossNN
Sturgeon
Sturgeon V2
top class − V1/MLR/crossNN
LGG, PA PA (not called) *
LGG, PA PF *
Other glioma − LGG PA − PA *
A IDH
Glio−neuronal − DLGNT − DL
Other glioma − ANA PA − AN
Other glioma − PXA − PXA
Sella − PITUI SCO GCT − SC
control
LGG, PA/GG ST
EPN, SPINE
Glioma IDH − O IDH − O IDH
Other glioma − HGNET − MN1
top class − Sturgeon V2
PA *
DLGNT
HGAP
control
DNT
GBM_MES
EVNCYT
ATRT_TYR
IHG
PXA
SEGA
DG_MYB
dot = confident correct call
>= threshold
< threshold
500
1000
1500
2000
2500
3000
3500
4000
4500
5000
5500
6000
6500
7000
7500
8000
8500
9000
9500
10000
...
20000
Timepoint

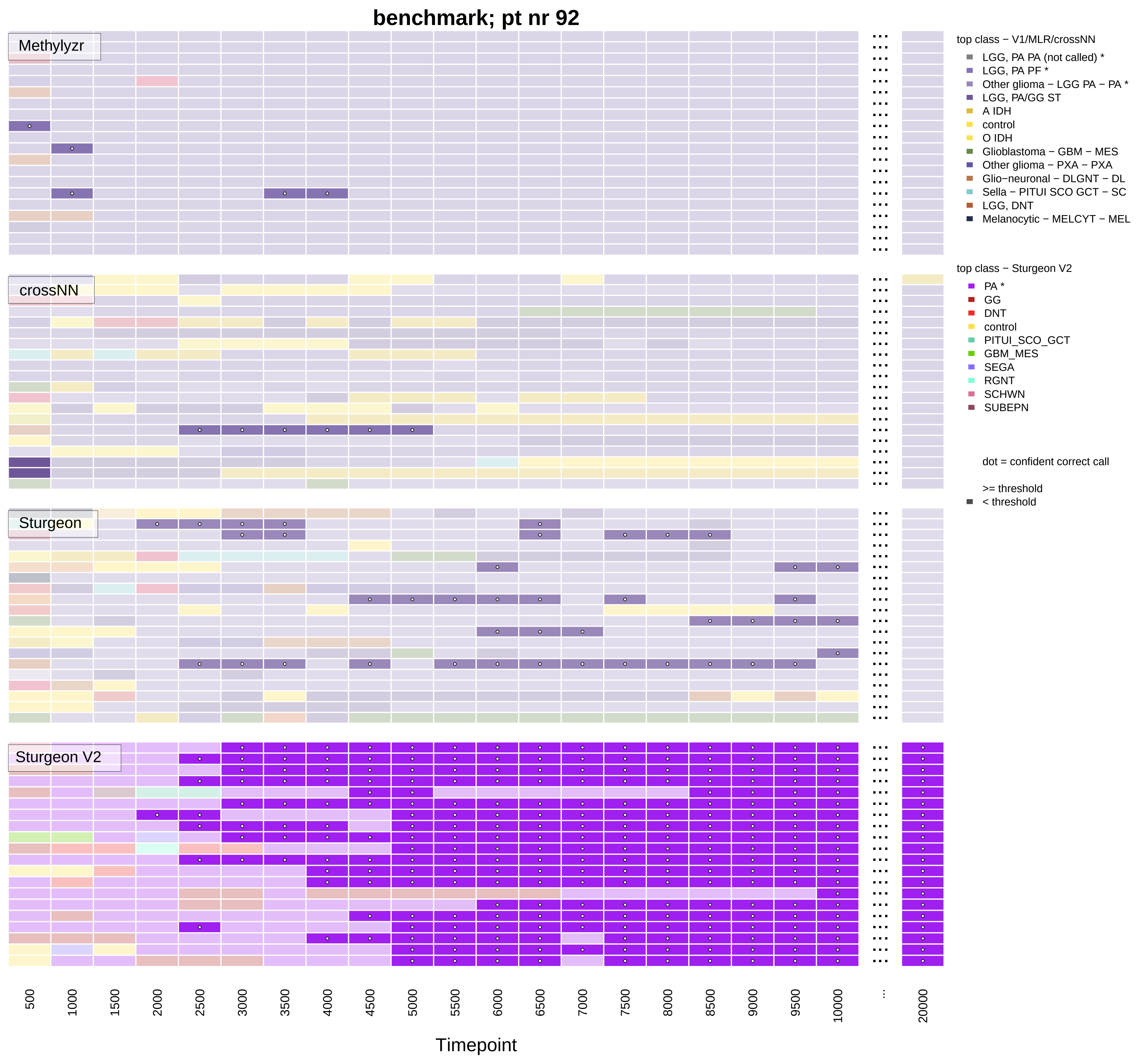

benchmark; pt nr 92
Methylyzr
crossNN
Sturgeon
Sturgeon V2
top class − V1/MLR/crossNN
LGG, PA PA (not called) *
LGG, PA PF *
Other glioma − LGG PA − PA *
LGG, PA/GG ST
A IDH
control
O IDH
Glioblastoma − GBM − MES
Other glioma − PXA − PXA
Glio−neuronal − DLGNT − DL
Sella − PITUI SCO GCT − SC
LGG, DNT
Melanocytic − MELCYT − MEL
top class − Sturgeon V2
PA *
GG
DNT
control
PITUI_SCO_GCT
GBM_MES
SEGA
RGNT
SCHWN
SUBEPN
dot = confident correct call
>= threshold
< threshold
500
1000
1500
2000
2500
3000
3500
4000
4500
5000
5500
6000
6500
7000
7500
8000
8500
9000
9500
10000
...
20000
Timepoint

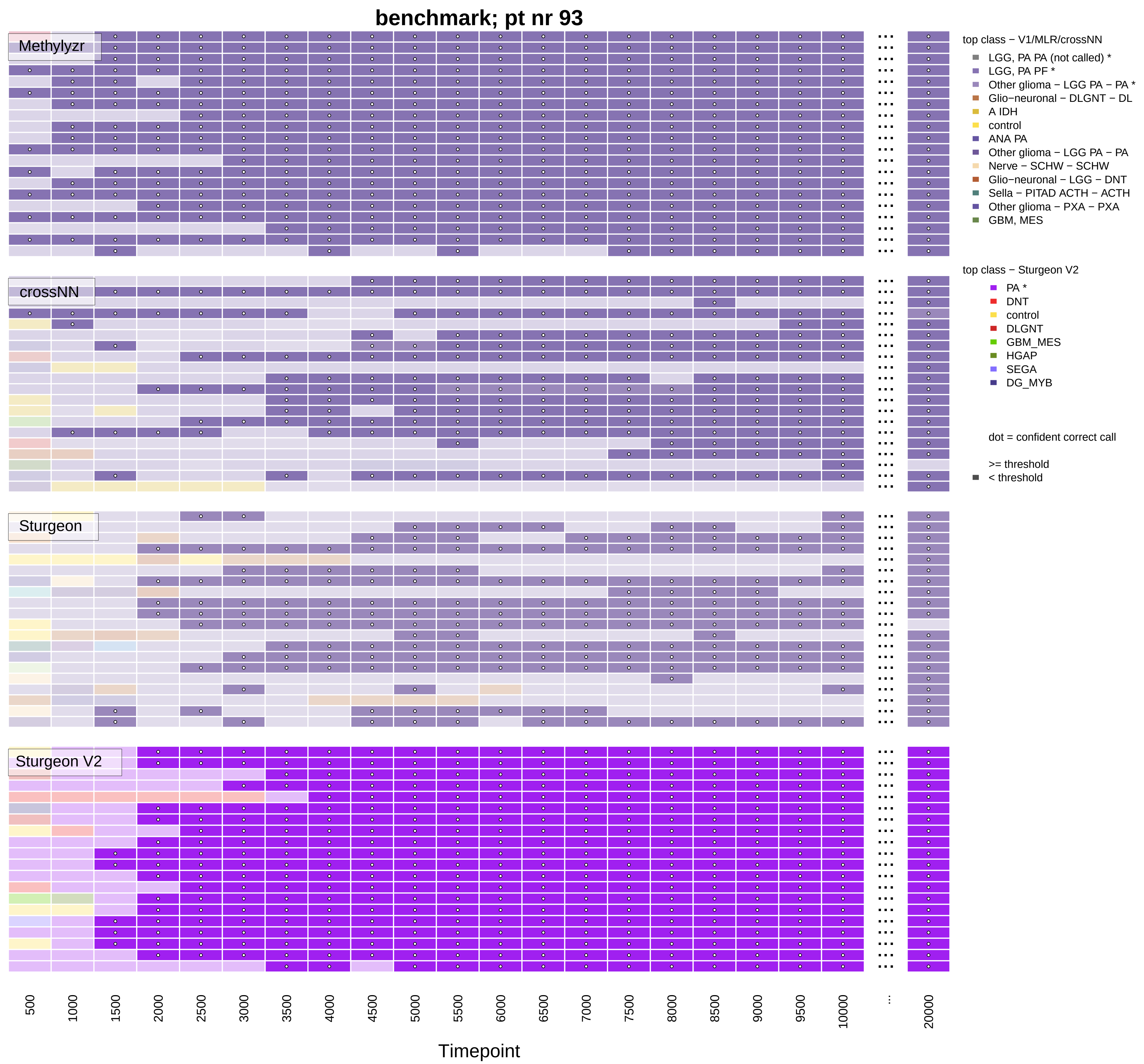

benchmark; pt nr 93
Methylyzr
crossNN
Sturgeon
Sturgeon V2
500
1000
1500
2000
2500
3000
3500
4000
4500
5000
5500
6000
6500
7000
7500
8000
8500
9000
9500
10000
...
20000
Timepoint
top class − V1/MLR/crossNN
LGG, PA PA (not called) *
LGG, PA PF *
Other glioma − LGG PA − PA *
Glio−neuronal − DLGNT − DL
A IDH
control
ANA PA
Other glioma − LGG PA − PA
Nerve − SCHW − SCHW
Glio−neuronal − LGG − DNT
Sella − PITAD ACTH − ACTH
Other glioma − PXA − PXA
GBM, MES
top class − Sturgeon V2
PA *
DNT
control
DLGNT
GBM_MES
HGAP
SEGA
DG_MYB
dot = confident correct call
>= threshold
< threshold

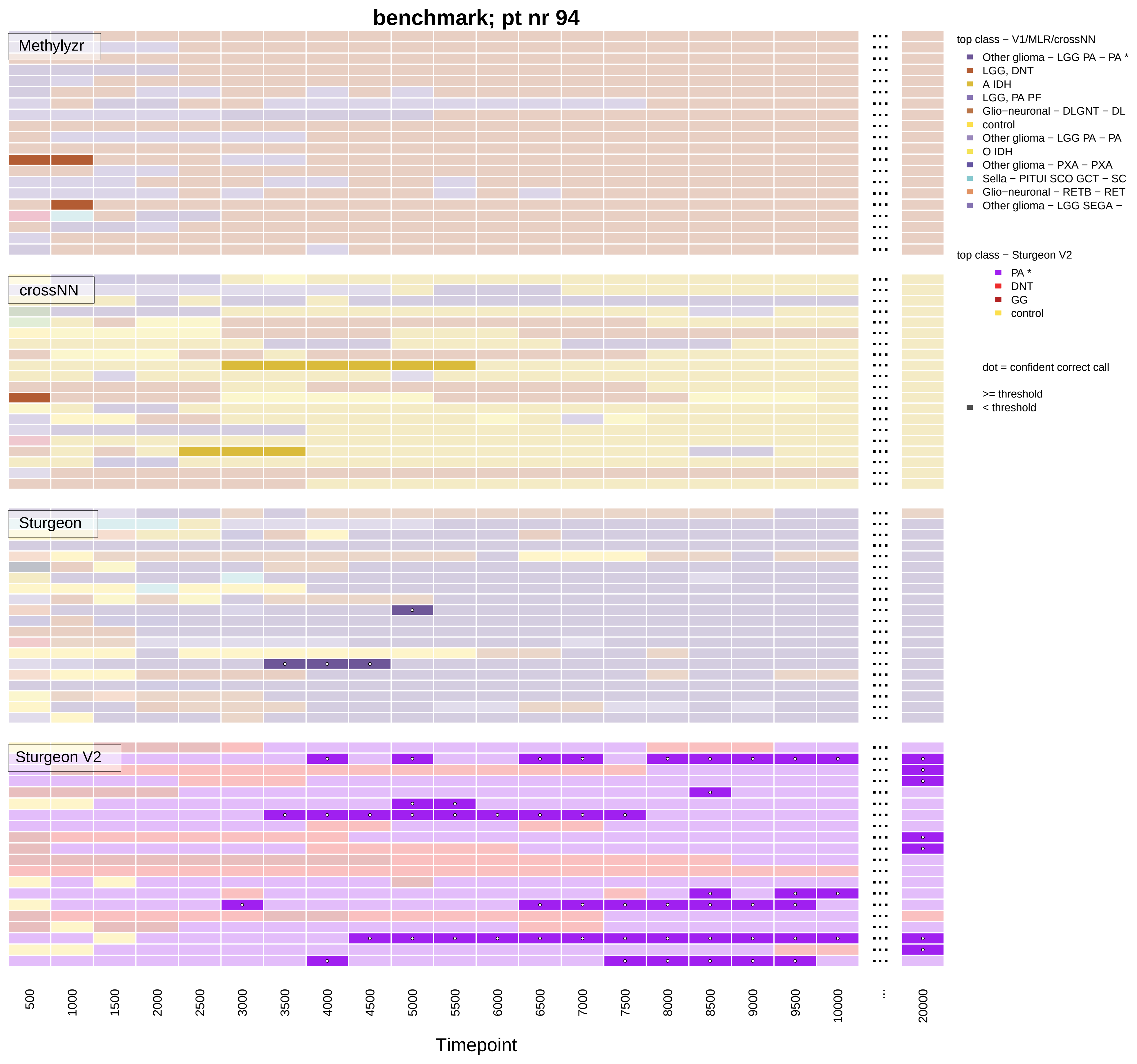
benchmark; pt nr 94
Methylyzr
crossNN
Sturgeon
Sturgeon V2
top class − V1/MLR/crossNN
Other glioma − LGG PA − PA *
LGG, DNT
A IDH
LGG, PA PF
Glio−neuronal − DLGNT − DL
control
Other glioma − LGG PA − PA
O IDH
Other glioma − PXA − PXA
Sella − PITUI SCO GCT − SC
Glio−neuronal − RETB − RET
Other glioma − LGG SEGA −
top class − Sturgeon V2
PA *
DNT
GG
control
dot = confident correct call
>= threshold
< threshold
500
1000
1500
2000
2500
3000
3500
4000
4500
5000
5500
6000
6500
7000
7500
8000
8500
9000
9500
10000
...
20000
Timepoint
